# Cross-LLM AI platform meta-research: Non-inferiority of bovine milk-based fortifiers to human milk-based fortifiers

**DOI:** 10.64898/2026.06.24.26356426

**Authors:** Duan Ni, Anjie Ge, Archita Mishra, Ju Lee Oei, Ralph Nanan

## Abstract

Necrotizing enterocolitis (NEC), frequently resulting in sepsis, is among the leading causes of morbidity and mortality of pre-term newborns. However, diagnostic and therapeutic strategies for NEC and sepsis are still limited and controversial. In this context, there are ongoing debates regarding the application of human milk-based fortifiers (HMF) versus bovine milk-based fortifiers (BMF), but robust evidence is lacking. Systematic reviews and meta-analyses are expected to provide the highest level of evidence, but they are time-consuming and resource-intensive and are at risk of potential bias and subjectivity. The rapidly progressing large language model (LLM) artificial intelligence (AI) tools thus emerge as a promising complementary methodology for systematic review and meta-analysis.

We conceptualized a cross-LLM AI platform meta-research and evidence synthesis workflow, leveraging 3 representative state-of-the-art platforms, ChatGPT, Claude and Manus AI. We analyzed 3371 PubMed-indexed publications. 3 platforms reported highly concordant findings. We found that prior systematic reviews and meta-analyses generally reported mixed findings comparing HMF versus BMF. Our LLM AI-assisted meta-research and evidence synthesis found non-inferiority of BMF to HMF for NEC and sepsis outcomes.

Here, we present an unbiased direct head-to-head comparison between HMF and BMF in the context of NEC and sepsis. Our analyses also represent a proof-of-concept example for LLM AI-assisted meta-research and evidence synthesis, supporting the integration of LLM AI methodologies into evidence-based medicine and digital health.

## Introduction

Substantial advances in neonatal care have significantly reduced morbidity and mortality of high-risk term and pre-term newborns. In this context, necrotizing enterocolitis (NEC), frequently resulting in sepsis, is one of the leading causes of death^1,2^, affecting 5% of premature neonates^2,3^. Neither the diagnostic nor therapeutic strategies for NEC have improved over the past 30 years^2^. Current prevention strategies for NEC are still limited and controversial^2^. The most important and accepted NEC-preventive approach is the use of maternal breastmilk^2^. However, for premature infants, satisfactory weigh gain is seen as a major challenge when fed with breastmilk alone. Hence, fortifiers aiming to increase the caloric density and/or modify macronutrient compositions are widely applied^4,5^. Pivoting on the benefits of breastmilk, human milk-based fortifiers (HMF) are claimed to effectively protect against NEC and sepsis, whilst providing adequate energy for growth. Specifically, HMF are deemed to be superior to conventional bovine milk-based fortifiers (BMF). However, implementation of HMF has been controversial, because robust evidence supporting the use of HMF appears to be lacking. This is a result of suboptimal study designs, heterogeneous feeding protocols, and mixed dietary interventions, which preclude a reliable comparison between HMF versus BMF versus unfortified breastmilk. In addition, HMF are among the costliest interventions in neonatology, and therefore cause significant financial burdens to health systems. This constellation thus represents a dilemma for clinicians and policymakers.

Systematic review and meta-analysis apply defined methodologies and provide a tiered level of evidence to inform clinical practice by supporting the development of clinical guidelines with diagnostic or therapeutic recommendations. Nevertheless, systematic review and meta-analysis have several inherent limitations^6–8^. For example, they are generally time-consuming and labour-and resource-intensive^9^. A prior study reported that, on average, completion of systematic reviews registered in the International Prospective Register of Systematic Reviews (PROSPERO) took approximately 1.3 years^10^. Additionally, systematic reviews are prone towards bias, resulting either from intentionality to yield desired outcomes, or from vulnerability of subjectivity in literature screening, reviewing and study evaluation^6–8^. Therefore, more unbiased and stringent improvements are needed for these approaches to further support evidence-based medicine.

The rapid advances of large language model (LLM) and artificial intelligence (AI) tools are likely to transform systematic review and meta-analysis workflows. Multiple studies have reported that LLM AI platforms such as ChatGPT and Claude could reliably – comparable to human reviewers – retrieve data for systematic review and meta-analysis across multiple fields^9,11–14^. Some latest papers also demonstrated that, even for evidence synthesis, performances of LLM AI platforms were similar to traditional human-based processes^9,13,14^. Hence, LLM AI tools might represent an unprecedented opportunity for faster and more effective meta-research and evidence synthesis, with reduced risk of bias and subjectivity.

Here, we aim to leverage various representative state-of-the-art LLM AI platforms to carry out meta-research to synthesize and cross-validate evidence comparing the effects of HMF versus BMF on NEC and sepsis. This will address a critical question in neonatology regarding the use of HMF versus BMF, as well as inform future broader implications for incorporating LLM AI-assisted methodologies into evidence-based medicine and digital health.

## Materials and methods

### Literature curation

Literature search was carried out on PubMed dated 7^th^ May 2026. Below list of keywords were used to search for HMF-related publications:

“*((((((((((((((human milk fortifier) OR (Fortified Human Milk)) OR (human-derived fortifier)) OR (human milk-derived fortifier)) OR (human milk fortification)) OR (human milk-based fortification)) OR (human milk-based fortifier)) OR (Donor Milk-Derived Fortifier)) OR (Donor-Derived Fortifier)) OR (human’s milk fortifier)) OR (human’s milk fortification)) OR (human milk-based human milk fortifier)) OR (human’s milk-based human milk fortifier)) OR (human milk-based human’s milk fortifier)) OR (human’s milk-based human’s milk fortifier)*”

3371 publications were found (Table S1 and Supplementary Information File 1) and based on the *Article Type* category in PubMed, 130 of them are meta-analysis, network meta-analysis or systematic review (Table S2 and Supplementary Information File 2). Detailed information for these papers including publication titles, publication details in journals, author names, author affiliations, DOI, PMID, and conflicts of interest statement were downloaded from PubMed as in Supplementary Information Files 1-2. This information was provided to LLM AI platforms for analysis detailed below.

### Cross-LLM AI platform meta-research

We here aim to present a proof-of-concept example of using LLM AI platforms for meta-research and evidence synthesis. We harness 3 presentative state-of-the-art platforms, namely, ChatGPT (GPT-5.5), Claude (Opus 4.7) and Manus AI (Manus Pro, Manus 1.6). LLM AI-driven analyses were conducted across all 3 platforms within a 1-week period from 7^th^ May 2026 to 14^th^ May 2026. It is worth noting that after our analyses, LLM AI platforms undergo continuous updates, such as the release of Opus 4.8 for Claude on 28^th^ May 2026. Notably, our work does not indicate the superiority of these platforms over others, but rather, propose a workflow that could be applicable for all other LLM AI tools, including DeepSeek and Gemini.

To improve the transparency and reproducibility of our analyses and avoid potential risk of hallucinations or fabricated outputs, we designed a series of standardized prompts for all 3 platforms for analyses, as suggested by prior publication^15^.

For analyses of previously published 130 meta-analysis, network meta-analysis and systematic review, following prompts were used:

“*Here are 130 meta-analysis, network meta-analysis or systematic review from pubmed, and their titles, abstracts, DOIs, etc. info from Pubmed that I searched using key words “((((((((((((((human milk fortifier) OR (Fortified Human Milk)) OR (human-derived fortifier)) OR (human milk-derived fortifier)) OR (human milk fortification)) OR (human milk-based fortification)) OR (human milk-based fortifier)) OR (Donor Milk-Derived Fortifier)) OR (Donor-Derived Fortifier)) OR (human’s milk fortifier)) OR (human’s milk fortification)) OR (human milk-based human milk fortifier)) OR (human’s milk-based human milk fortifier)) OR (human milk-based human’s milk fortifier)) OR (human’s milk-based human’s milk fortifier)”;*

*Please go through all their related information very carefully in details!*

*I want to find the meta-analysis and systematic review in them that directly compare the effects of human-based milk fortifier versus bovine/cow milk-based fortifier on Necrotizing Enterocolitis (NEC) (any stages) and sepsis*.

*Please be careful with all the synonym for the keywords related to human-based milk fortifier and bovine/cow milk-based fortifier*.

*Please extract their related information into an excel spreadsheet*.

*Thank you!*”

Based on such instruction, Claude identified 10 publications (Table S3), including 7 that were consistently found by both ChatGPT (Table S4) and Manus AI (Table S5).

For our own standalone meta-research and evidence synthesis, a stepwise approach was applied:

(1) We first ask the LLM AI platforms to identify RCTs directly comparing HMF versus BMF:

“*Here are 3371 papers and their titles, abstracts, DOIs, etc. info from Pubmed that I searched using key words “((((((((((((((human milk fortifier) OR (Fortified Human Milk)) OR (human-derived fortifier)) OR (human milk-derived fortifier)) OR (human milk fortification)) OR (human milk-based fortification)) OR (human milk-based fortifier)) OR (Donor Milk-Derived Fortifier)) OR (Donor-Derived Fortifier)) OR (human’s milk fortifier)) OR (human’s milk fortification)) OR (human milk-based human milk fortifier)) OR (human’s milk-based human milk fortifier)) OR (human milk-based human’s milk fortifier)) OR (human’s milk-based human’s milk fortifier)”;*

*Please go through all their related information very carefully in details!*

*I want to do a meta-analysis and systematic review directly comparing the effects of human-based milk fortifier versus bovine/cow milk-based fortifier on Necrotizing Enterocolitis (NEC) (any stages) and sepsis*.

*Please only find me the Randomized Controlled Trial (RCT) in these 3371 papers/abstracts that could be used for aforementioned comparison and meta-analysis and systematic review*.

*Please be careful with all the synonym for the keywords related to human-based milk fortifier and bovine/cow milk-based fortifier*.

*Please extract their related information into an excel spreadsheet. Thank you!*”

Based on this prompt, Claude identified 10 papers (Table S6), while Manus AI (Table S7) and ChatGPT (Table S8) identified the same 9 papers. The relevance of these papers was reviewed and approved by the authors. There were 7 papers consistently selected by all 3 LLM AI platforms. Among these 12 papers, the work by Polberger *et al.* had no full-text publication available online and was thus excluded, resulting in a total of 11 papers for further analyses.

Full-text manuscripts, including supplementary information files were downloaded for all these 11 papers for analyses.

(2) Before further analysis, we provided LLM AI platforms with the pdf file of “*Revised Cochrane risk-of-bias tool for randomized trials (RoB 2)*” (Supplementary Information Files 3), the excel spreadsheet for data extraction (Table S9) and the following prompt to inform them about their tasks for data extraction and analysis:

“*Please read attached PDF and excel. The Excel contains a list of questions for you to answer, and the PDF is the guideline for you to read the paper and answer those questions. Do you understand? If you understand, let me know and I would provide you the papers for analysis.*”

(3) After (2), 11 publications selected in (1) were uploaded to each LLM AI platform, which was asked to perform the specific tasks in (2) with the following prompt:

“*Here is a zip file containing all the papers and their related supplementary information of interests. The file names all contain the PMIDs of the papers correspondingly, and some have been labelled as supplementary information*.

*Please read through all these papers and supplementary information carefully and based on the PDF guidelines I just sent you, answer the questions in the excel I just sent you. After finishing analyzing all the papers, please combine all the results into 1 excel*.

*Please be careful and match the papers and their corresponding supplementary information correctly when doing analysis.*”

(4) Based on results from (3), each LLM AI platform was asked to perform further analyses with the prompt below:

“*Ok, now based on all these papers, the related information and analysis and the results, please see if it is possible to perform a meta-analysis based on them directly comparing the effects of human-based milk fortifier versus bovine/cow milk-based fortifier on Necrotizing Enterocolitis (NEC) (any stages) and sepsis*.

*Please be careful with all the synonym for the keywords related to human-based milk fortifier and bovine/cow milk-based fortifier*.

*If that’s feasible, please proceed and do it.*”

Results from above workflow were compared across platforms and reviewed by authors.

## Results

### Overview of cross-LLM AI platform meta-research workflow

We aimed to harness representative state-of-the-art LLM AI platforms to perform meta-research, identify potential new findings and synthesize updated evidence comparing the effects of HMF versus BMF on NEC and sepsis outcomes.

We conceptualized a cross-LLM AI platform meta-research workflow using 3 representative independent platforms: ChatGPT, Claude and Manus AI (Materials and methods). We first asked these LLM AI platforms to screen a collection of publications related to HMF manually curated from PubMed and to identify: 1) existing systematic reviews and meta-analyses related to HMF versus BMF comparisons (Figure 1); 2) existing RCTs directly comparing HMF versus BMF for outcomes of interests (i.e., NEC and sepsis) (Figure 2). Next, selected publications were analyzed by LLM AI platforms in parallel: publications from 1) were analyzed based on study outcomes, while publications from 2) were analyzed using a defined meta-analysis methodology for evidence synthesis. Final results were compared across platforms.

**Figure 1.**
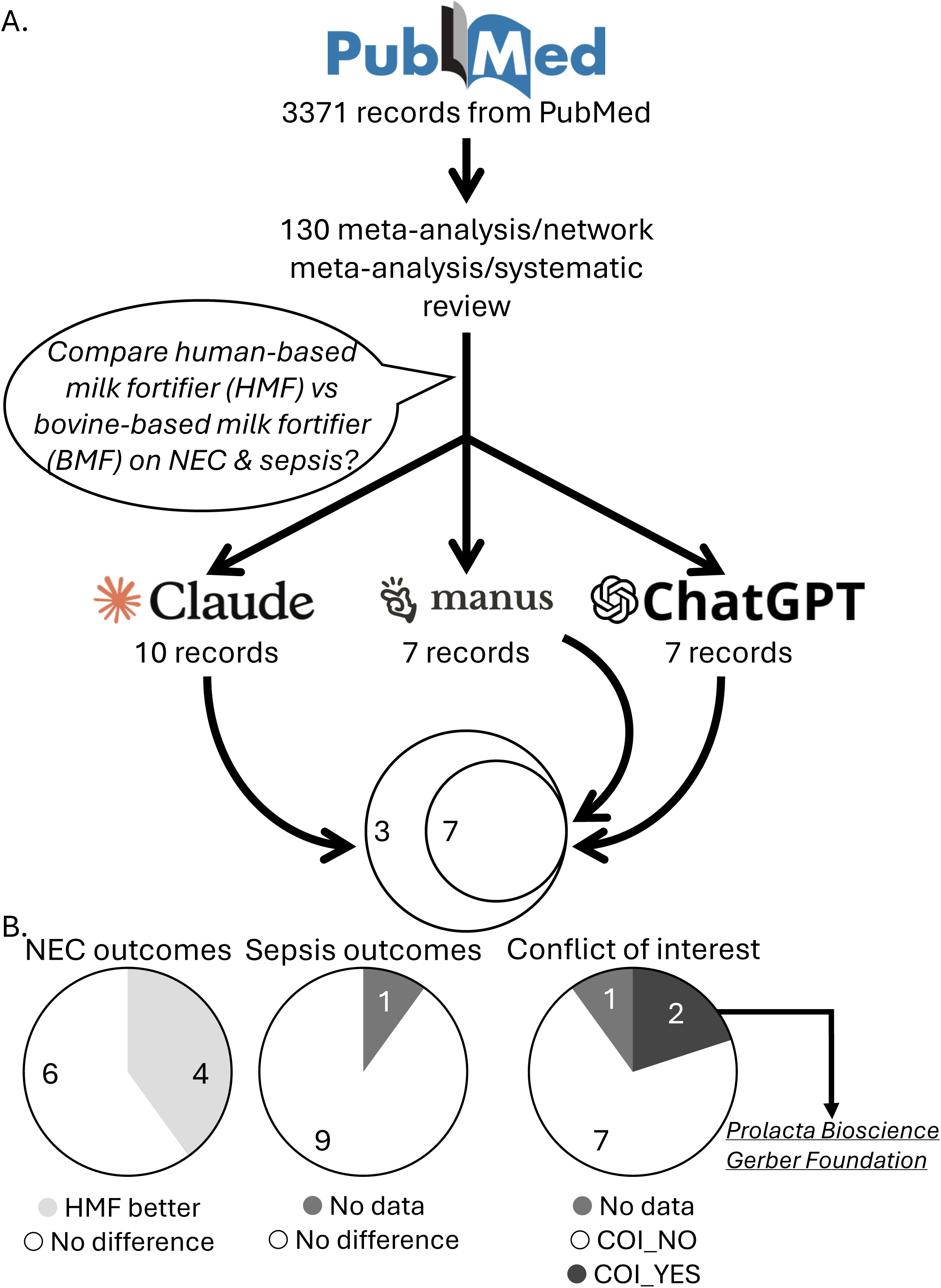
**A.** Overview of the cross-large language model (LLM) artificial intelligence (AI) platform meta-research workflow for re-analyzing existing systematic reviews and meta-analyses comparing human milk-based fortifiers (HMF) versus bovine milk-based fortifier (BMF) for necrotizing enterocolitis (NEC) and sepsis outcomes. **B.** NEC and sepsis outcomes and conflict of interest (COI) statuses reported in prior systematic reviews and meta-analyses.

**Figure 2.**
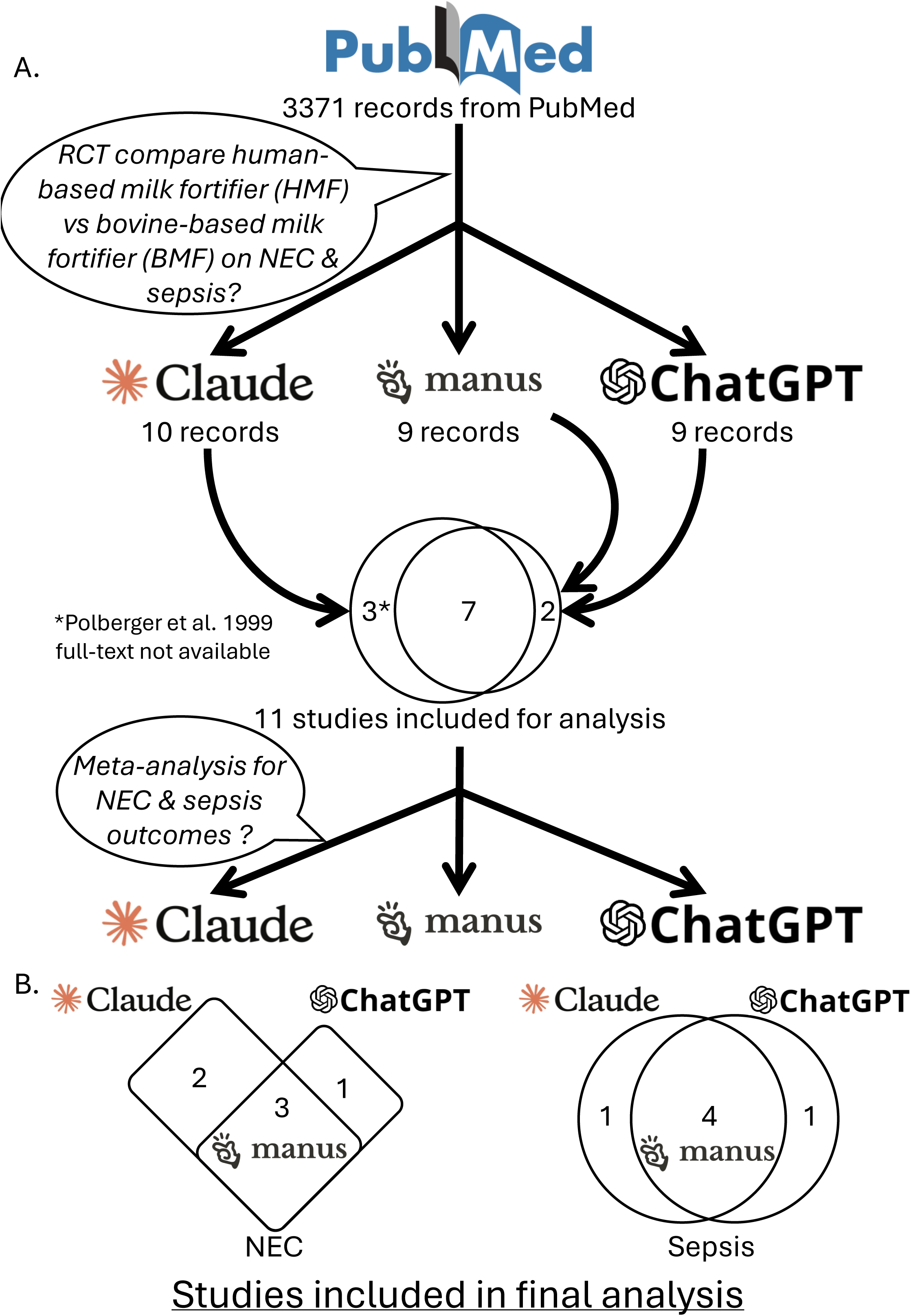
**A.** Overview of the cross-large language model (LLM) artificial intelligence (AI) platform meta-research workflow for independent meta-research and evidence synthesis from randomized controlled trials (RCTs) comparing human milk-based fortifiers (HMF) versus bovine milk-based fortifier (BMF) for necrotizing enterocolitis (NEC) and sepsis outcomes. **B.** Numbers of studies included for meta-research and evidence synthesis for NEC (left) and sepsis (right) outcomes by Claude, ChatGPT and Manus AI.

### Overview of current systematic reviews and meta-analyses comparing HMF versus BMF

3371 papers were found on PubMed dated 7^th^ May 2026 based on keyword search related to HMF (Table S1, Supplementary Information File 1, and Materials and methods). Among them, there were 130 *meta-analyses* or *network meta-analyses* or *systematic reviews* based on the PubMed *Article Type* categories (Table S2, Supplementary Information File 2, and Materials and methods). ChatGPT, Claude and Manus AI were provided with the detailed information of these publications including titles and abstracts and were asked to identify studies that compare the effects of HMF versus BMF on NEC and/or sepsis (Figure 1A). Claude identified 10 publications (Table S3)^1,16–24^, including 7 that were consistently found by both ChatGPT (Table S4) and Manus AI (Table S5)^1,16,17,19–22^. These 10 publications were further reviewed for their findings by all LLM AI platforms and human reviewers. 4/10 publications reported that HMF performed superior to BMF in improving NEC, while in 6/10 publications there was no difference (Figure 1B). For sepsis, for all but one publications identified non-inferiority of BMF versus HMF (Figure 1B). Notably, 2/10 studies reported conflicts of interests, such as authors directly receiving financial support from manufacturers of HMF (Figure 1B & Table S10).

### Workflow for cross-LLM AI platform meta-research and evidence synthesis

Mixed results from previous systematic reviews and meta-analyses prompted us to harness the state-of-the-art LLM AI platforms to perform additional stand-alone meta-research and evidence synthesis, in an attempt to address concerns with current systematic reviews and meta-analyses for HMF versus BMF comparisons.

To achieve this, we started with 3371 HMF-related PubMed-indexed publications and asked Claude, Manus AI and ChatGPT to identify papers reporting randomized controlled trials (RCTs) directly comparing the impacts of HMF versus BMF on NEC and/or sepsis, aiming to perform the cleanest and most direct head-to-head comparison of HMF versus BMF (Figure 2A).

Claude identified 10 papers (Table S6)^25–34^, while Manus AI (Table S7) and ChatGPT (Table S8) identified the same 9 papers^27–29,31–36^. The relevance of these papers was reviewed and approved by human reviewers (i.e., authors). There were 7 papers consistently selected by all 3 LLM AI platforms. Among these 12 papers, the work by Polberger *et al.*^26^ had no full-text publication available online and was thus excluded from further analyses (Figure 2A).

Notably, among remaining 11 papers, all except one – published in 1990, predating routine conflicts of interest declaration – reported conflicts of interest with considerable involvements of industry funding supports. Specifically, 2 studies^30,34^ disclosed that industry funding bodies were directly involved in study design and/or result analysis. Such observations suggest the vulnerability to bias and/or subjectivity for studies involving HMF and BMF (Table S11).

For the 11 selected studies, their full-text publications were downloaded for following meta-research and evidence synthesis using LLM AI platforms in a similar manner to meta-analysis, questing the effects of HMF versus BMF on NEC and sepsis respectively.

### Effects of HMF and BMF on NEC

LLM AI platforms were provided with the full-text manuscripts including their supplementary information files of 11 selected studies to perform literature screening, data extraction, and meta-research evidence synthesis. After reviewing full-text manuscript, 3 out of these 11 publications were consistently included by Manus AI (Table S12-13), ChatGPT (Table S14-15) and Claude (Table S16-17) for analyses of NEC as an outcome. Claude included 2 additional studies and ChatGPT included another 1 study (Figure 2B). Additional publications selected by Claude and ChatGPT were excluded by Manus AI because 1) study was outdated and might not reflective of modern HMF/BMF^25^; 2) studies reported 0 NEC incidence^25,30^; 3) study with potential confounders like mixed feeding regimens, where infants were on an exclusive human milk diet with pasteurized human milk for any shortfall in mothers’ own milk and HMF versus bovine formula and BMF^31^ (Table S18). Despite differences in inclusions, random-effects meta-analyses from all 3 LLM AI platforms consistently found non-inferiority of BMF versus HMF on NEC outcomes.

Analysis by Manus AI covered 3 publications^29,32,33^ (Figure 3A), with 193 and 188 infants in HMF versus BMF groups respectively. They had comparable NEC incidences (HMF: 12/193; BMF: 15/188) and HMF was not associated with statistically significant reduction in NEC relative to BMF (risk ratio (RR): 0.78, 95% confidence interval: (0.37, 1.63)). No evidence of statistical heterogeneity was found (I^2^ = 0.0%, τ^2^ = 0.000).

**Figure 3.**
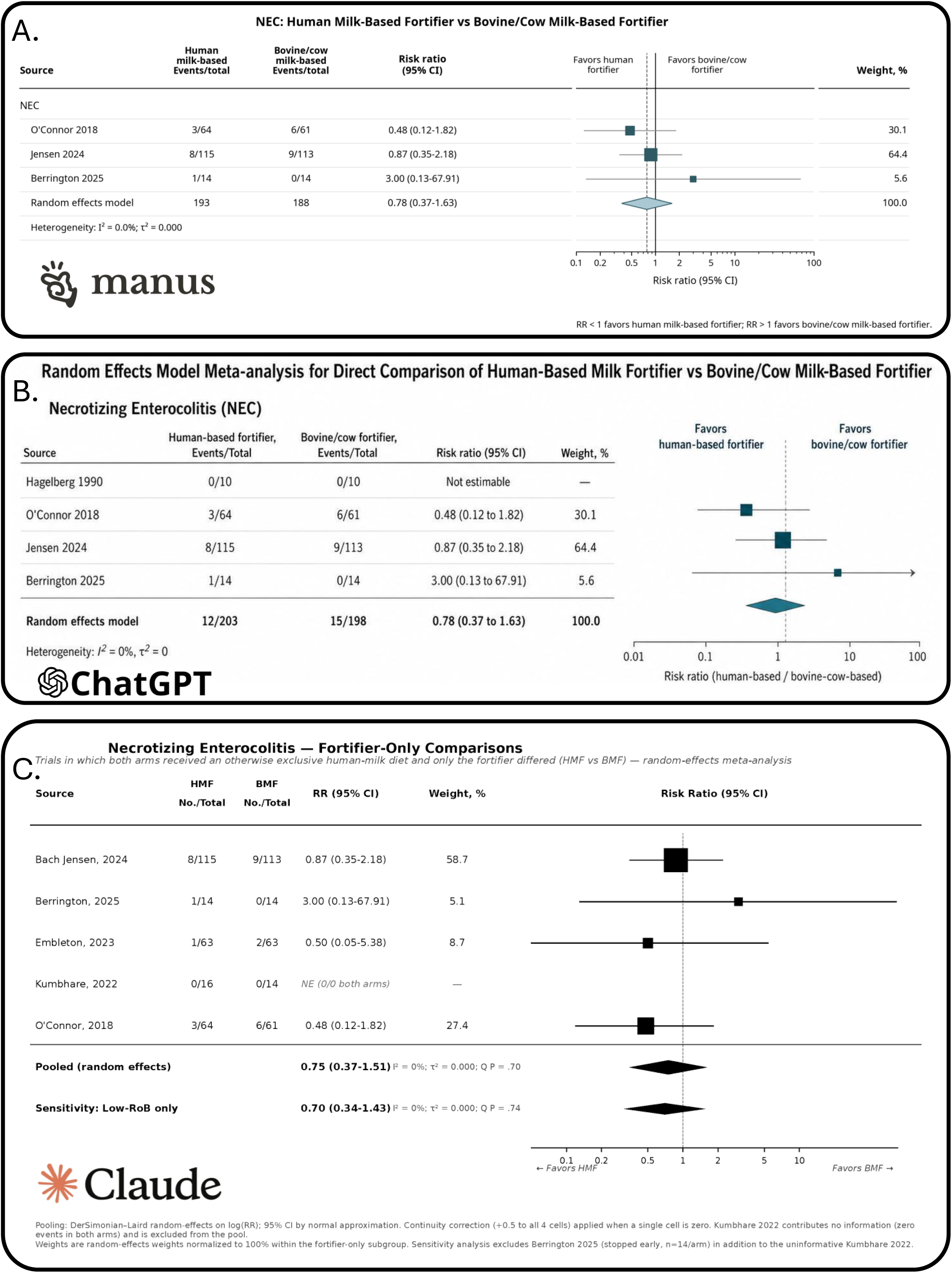
Random effects model meta-analysis for changes in necrotizing enterocolitis (NEC) outcomes comparing human milk-based fortifiers (HMF) versus bovine milk-based fortifier (BMF) by Manus AI (**A**), ChatGPT (**B**) and Claude (**C**).

Analysis by ChatGPT covered 4 publications^25,29,32,33^ (Figure 3B), with 203 and 198 infants in HMF versus BMF groups and 12/203 (HMF) versus 15/198 (BMF) of them reported NEC respectively. No association was found between HMF and reduction in NEC (RR: 0.78 (0.37, 1.63)), and no evidence of statistical heterogeneity was detected (I^2^ = 0.0%, τ^2^ = 0.000).

Analysis by Claude covered 5 publications^29–33^ (Figure 3C), and there are 272 and 265 individuals in HMF versus BMF respectively. Among them, 13/272 (HMF) and 17/265 (BMF) reported NEC, without statistically significant difference. HMF is not linked to reduction in NEC (RR: 0.75 (0.37, 1.51)) and no evidence of statistical heterogeneity was found (I^2^ = 0.0%, τ^2^ = 0.000).

Overall, there is no evidence supporting the superiority of HMF over BMF in NEC outcomes.

### Effects of HMF and BMF on sepsis

A similar workflow was applied for analysis for sepsis. For sepsis outcomes, 4 studies were consistently included by Manus AI (Table S12-13), ChatGPT (Table S14-15) and Claude (Table S16-17) for analyses. Claude and ChatGPT each included 1 additional study (Figure 2B), both of which were excluded by Manus AI based on similar reasons as described above for NEC-related analyses (Table S18). Nevertheless, analyses by 3 LLM AI platforms also yielded similar results.

Manus AI covered 4 publications^29,30,32,33^ and 209 HMF and 202 BMF individuals for analysis (Figure 4A). Sepsis incidences were similar in HMF (49/209) and BMF (44/202) groups, with a RR = 1.11 (0.57, 2.14). Moderate heterogeneity was found, with I^2^ = 45.1%, τ^2^ = 0.190.

**Figure 4.**
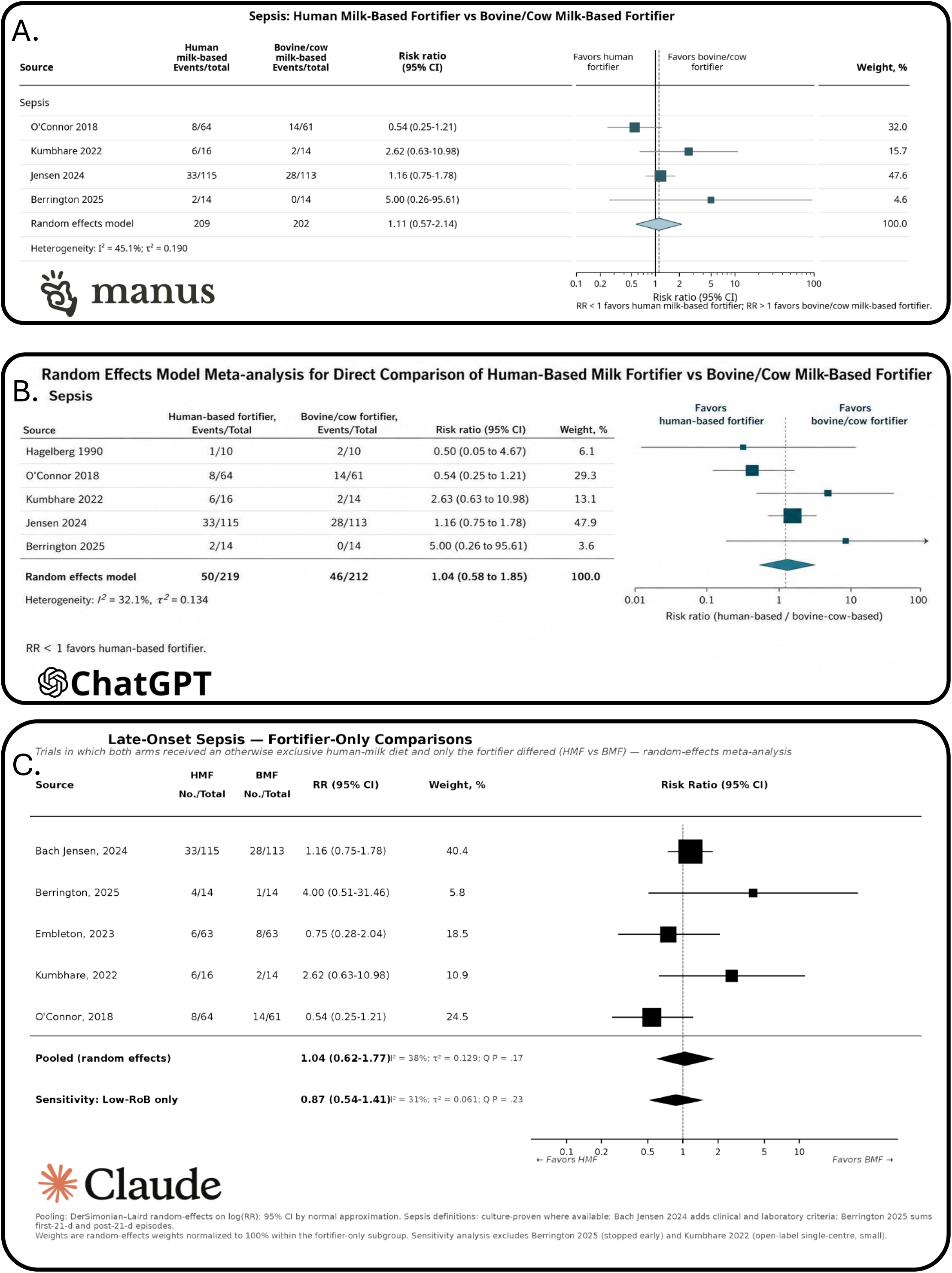
Random effects model meta-analysis for changes in sepsis outcomes comparing human milk-based fortifiers (HMF) versus bovine milk-based fortifier (BMF) by Manus AI (**A**), ChatGPT (**B**) and Claude (**C**).

ChatGPT included 5 publications for analysis^25,29,30,32,33^ (Figure 4B), involving 219 HMF and 212 BMF individuals respectively. HMF (50/219) and BMF (46/212) groups had comparable sepsis incidences (RR: 1.04, (0.58, 1.85)), with moderate between-study heterogeneity (I^2^ = 32.1%, τ^2^ = 0.134).

Claude used 5 publications for analysis^29–33^ (Figure 4C), with 272 HMF and 265 BMG individuals. They reported 57/272 (HMF) and 40/265 (BMF) sepsis cases respectively, with no statistically significant difference (RR: 1.04, (0.62, 1.77)). Moderate heterogeneity was found among studies, with I^2^ = 38.0%, τ^2^ = 0.129.

Together, there is also no evidence suggesting that HMF is superior to BMF in improving sepsis outcomes.

## Discussion

We present a cross-LLM AI platform meta-research workflow to address one of the pressing controversies in neonatology regarding the use of HMF versus BMF for NEC and sepsis outcomes. Thorough review of prior systematic reviews and meta-analyses found that they generally yielded inconsistent findings. These mixed results might result from intentional or unintentional bias. Our meta-research and evidence synthesis using 3 representative cutting-edge LLM AI platforms, namely, ChatGPT, Claude and Manus AI, consistently found non-inferiority of BMF versus HMF for NEC and sepsis outcomes. This presents independent evidence for a controversial nutritional intervention and provides an exemplary proof-of-concept incorporating LLM AI platforms into evidence-based medicine as a complementary methodology.

Despite advances in neonatology, NEC still represents one of the critical challenges in the field, lacking effective interventions. HMF seems to emerge as a new intervention option in this context but face significant barriers in implementation. Concrete evidence supporting the superiority of HMF over BMF towards NEC and associated conditions like sepsis is lacking, although it is generally acknowledged that human milk would confer benefits in these contexts. There are very limited stringently controlled clinical studies or systematic reviews and meta-analyses directly comparing HMF versus BMF, and they generally reported inconsistent findings. These comparisons are frequently subject to confounders such as mixed dietary interventions, where HMF and BMF were mixed with human milk and preterm formula respectively, and heterogeneous feeding regimen, where dosages of HMF versus BMF might not be properly controlled^1^. These factors might all interfere with the actual comparison between HMF and BMF. To address these issues, our current analyses specifically focused on RCTs where HMF/BMF fortifier types were the only variable between two arms, providing the cleanest head-to-head comparisons. Overall, with modest sample sizes (∼400 individuals), we found no evidence indicating that HMF is superior to BMF in NEC and sepsis outcomes.

The applications of our cross-LLM AI platform pipeline in such analysis present several advantages. Firstly, it is acknowledged that traditional human-driven systematic review and meta-analysis are inevitably vulnerable to bias and/or subjectivity in different steps of the process such as during literature screening and eligibility evaluation^6–8^. This might be particularly critical in fields at risk of conflicts of interest like studies involving HMF and BMF, as our analyses showed. Thus, LLM AI platforms might provide a more unbiased and neutral alternative. Our cross-validations with 3 independent state-of-the-art LLM AI tools, as well as human reviewing, further support the robustness and reliability of findings from our workflow. Another limitation of traditional systematic review and meta-analysis is their transparency and reproducibility^6–9,14^. Here, built upon standardized prompts, our LLM AI-based analyses generated clear logs and documentations, facilitating the tracing and tracking of the entire analytical process, which is expected to considerably improve the transparency and reproducibility. Finally, with their reasonably optimal performance, LLM AI-based workflows present a simpler and faster streamlined process for meta-research and evidence synthesis. As demonstrated in our study here, our analyses merely require literature lists and/or PDFs and standardized prompts as inputs, supplying significantly enhanced simplicity and reducing the barriers for meta-research and evidence synthesis.

Prior attempts of using LLM AI platforms for meta-research raised concerns such as risks of hallucinations or fabricated outputs, and omission of eligible studies^9^. These issues seemed to be minimized in our case, likely because we adopted standardized detailed prompts for instructions, as previously suggested^15^. The continuous advances in LLM AI models might also contribute to such improvements. Previous studies also highlighted the necessity of human supervision and reviewing to improve the robustness and reduce the risk of error in LLM AI-based analyses^9,14,15^. In this regard, it must be noted that the rapid developments in LLM and AI fields are likely to progress towards resolutions of these issues, with updated and newer models and algorithms. On the other hand, integrative comparative analyses with various platforms/models, as in our present work, might provide another plausible solution.

We here focused on 3 representative LLM AI platforms, ChatGPT, Claude and Manus AI, but our workflow, applications and findings could be easily extended to other platforms such as Gemini, Llama and DeepSeek, and finetuned for other research questions and scientific areas.

Overall, we here showcased a proof-of-concept example for cross-LLM AI platform meta-research and evidence synthesis. Harnessing such workflow, we addressed a critical question in neonatological nutrition, demonstrating the non-inferiority of BMF compared with HMF for NEC and sepsis outcomes. Our analyses also highlight the unprecedented possibility of leveraging LLM AI tools as a novel methodological paradigm for evidence-based medicine with enhanced efficiency, transparency, and reproducibility.

## List of supplementary files

**Supplementary File 1.** Abstracts and detailed information of the 3371 PubMed-indexed papers included in analysis.

**Supplementary File 2.** Abstracts and detailed information of the 130 PubMed-indexed meta-analysis, network meta-analysis and systematic review included in analysis.

**Supplementary File 3.** Revised Cochrane risk-of-bias tool for randomized trials (RoB 2, 22^nd^ August 2019)

**Table S1.** Detailed information of the 3371 PubMed-indexed papers included in analysis.

**Table S2.** Detailed information of the 130 PubMed-indexed meta-analysis, network meta-analysis, and systematic review included in analysis.

**Table S3.** Current systematic reviews and meta-analyses identified by Claude for direct comparisons of HMF vs BMF (Direct output from Claude).

**Table S4.** Current systematic reviews and meta-analyses identified by ChatGPT for direct comparisons of HMF vs BMF (Direct output from ChatGPT).

**Table S5.** Current systematic reviews and meta-analyses identified by Manus AI for direct comparisons of HMF vs BMF (Direct output from Manus AI).

**Table S6.** Papers reporting randomized controlled trials (RCTs) comparing HMF versus BMF identified by Claude (Direct output from Claude).

**Table S7.** Papers reporting randomized controlled trials (RCTs) comparing HMF versus BMF identified by Manus AI (Direct output from Manus AI).

**Table S8.** Papers reporting randomized controlled trials (RCTs) comparing HMF versus BMF identified by ChatGPT (Direct output from ChatGPT).

**Table S9.** Questionnaire for literature analysis and data extraction used by LLM AI platforms for meta-research and evidence synthesis.

**Table S10.** Conflict of interest statuses from current systematic reviews and meta-analyses comparing HMF versus BMF.

**Table S11.** Conflict of interest statements for 11 papers selected for meta-research and evidence synthesis by LLM AI platforms, with details from publications where industry funding bodies were directly implicated in studies.

**Table S12.** Manus AI data extraction and risk of bias analysis results (direct output from Manus AI).

**Table S13.** Manus AI meta-research results (direct output from Manus AI).

**Table S14.** ChatGPT data extraction and risk of bias analysis results (direct output from ChatGPT).

**Table S15.** ChatGPT meta-research results (direct output from ChatGPT).

**Table S16.** Claude data extraction and risk of bias analysis results (direct output from Claude).

**Table S17.** Claude meta-research results (direct output from Claude).

**Table S18.** Details for paper inclusion/exclusion by Manus AI.

## Author contribution

D.N.: conceptualization, analysis, editing and writing original draft. A.G.: analysis. A.M.: analysis. J.L.O.: analysis. R.N.: conceptualization, analysis, editing and writing original draft, supervision.

All authors have read and approved the final version of the manuscript.

## Data Availability

All data produced in the present study are available upon reasonable request to the authors

## Acknowledgement

The study is supported by the Norman Ernest Bequest Fund and The University of Sydney Charles Perkins Centre EMCR Seed Funding.

