## Supplementary material for "Cross-LLM AI platform meta-research: Non-inferiority of bovine milk-based fortifiers to human milk-based fortifiers": SI1_3371 papers abstracts.txt

1. Semin Perinatol. 2026 May 4:152238. doi: 10.1016/j.semperi.2026.152238. Online 
ahead of print.

Preface.

Moya F(1), Poindexter BB(2).

Author information:
(1)Pediatrix Medical Group of Austin; Texas A & M College of Medicine. 
Electronic address:.
(2)Division of Neonatology, Emory University School of Medicine and Children's 
Healthcare of Atlanta.

DOI: 10.1016/j.semperi.2026.152238
PMID: 42086449

Conflict of interest statement: Disclosures FM: Part of the Speaker Bureau of 
Mead Johnson Nutrition; has received grant funding from Mead Johnson Nutrition 
for studies of human milk fortification; has received educational grant funding 
from Abbott Laboratories. BP: none.


2. Nutrients. 2026 Apr 17;18(8):1271. doi: 10.3390/nu18081271.

Selenium in the Preterm Infant: Are We Supplementing Enough in This Vulnerable 
Population?

Eckert JV(1), Rogers LK(1), Tipple TE(1), Mathias M(1).

Author information:
(1)Neonatal-Perinatal Medicine, Department of Pediatrics, University of Oklahoma 
Health, Oklahoma City, OK 73104, USA.

Selenium (Se) is an essential trace element. The bioactivity of Se arises from 
its incorporation into the 21st amino acid, selenocysteine (Sec). Twenty-five 
human genes have been identified that encode selenoproteins, each of which 
contains at least one Sec residue. Selenoprotein functions include antioxidant 
responses, thyroid hormone synthesis, and maintenance of cellular redox 
homeostasis. Due to its role in critical cellular functions, Se deficiency is 
associated with morbidities of the cardiovascular system and connective tissue 
in regions of countries with low soil Se content. While these morbidities are 
geography-specific and have been mitigated in adults through public health 
interventions, preterm infants remain susceptible to Se deficiency worldwide. 
Infants born preterm are deprived of fetal Se accrual in the 3rd trimester of 
pregnancy, a deficiency compounded by higher Se needs than term infants and 
older infants and dependence on parenteral nutrition (PN) and fortification. In 
addition, the composition of selenoproteins and selenometabolites in human milk 
is different from that in formula and PN, yet little is known about the 
biological impact of these differences. The knowledge gap in optimal Se 
supplementation is reflected in discrepant guidelines between North American and 
European/Chinese nutrition societies, whose recommended Se supplementation in 
preterm infants differs by more than 2-fold. In this review, we describe the 
biosynthesis, metabolism, and maternal-fetal transfer of Se. In addition, we 
address how developmentally regulated aspects of metabolism may impact how 
preterm infants respond to supplementation with different forms of Se. Lastly, 
we highlight current challenges and recommendations for optimizing Se levels in 
neonates based on available data.

DOI: 10.3390/nu18081271
PMCID: PMC13118472
PMID: 42075084 [Indexed for MEDLINE]

Conflict of interest statement: The authors declare no conflicts of interest.


3. Nutrients. 2026 Apr 10;18(8):1194. doi: 10.3390/nu18081194.

Vitamin D Fortification Strategies and Policy Landscape in Selected European 
Countries.

Czyżniewski B(1), Chmielowiec J(1), Chmielowiec K(1), Gibas-Dorna M(1).

Author information:
(1)Institute of Health Sciences, University of Zielona Góra, Zyty Str. 28, 
65-046 Zielona Góra, Poland.

BACKGROUND: Vitamin D deficiency remains a widespread public health issue in 
Europe, despite the availability of sunlight, dietary sources, supplements, and 
food fortification. National fortification strategies differ substantially in 
their regulatory approaches, food vehicles, and fortification levels, 
influencing the population's vitamin D intake and status.
OBJECTIVE: The primary objective of this study was to map vitamin D food 
fortification policies across European Union (EU) Member States, European Free 
Trade Association (EFTA) countries, and the United Kingdom (UK), focusing on 
regulatory frameworks, eligible food categories, and implementation models.
METHODS: A structured review of national legislation and official guidance on 
vitamin D food fortification was conducted between December 2025 and March 2026 
across EU Member States (n = 27), EFTA countries (n = 4), and the UK. For EU 
Member States, the framework established by Regulation (EC) No 1925/2006 was 
examined alongside national implementation measures. For EFTA countries and the 
UK, corresponding national legislation and official regulatory guidance were 
reviewed. Data were extracted on fortification policy status, eligible food 
categories, legal basis, and fortification levels. Targeted searches of PubMed 
and Scopus were performed to identify modeling studies and policy analyses 
supporting the interpretation of the findings.
RESULTS: Fortification policies show marked heterogeneity. Mandatory 
fortification is limited to a few countries and specific foods: Finland 
(homogenized skim milk), Sweden (low-fat milk, fermented dairy, plant-based 
alternatives, and fat spreads), Belgium (margarine and selected fats), and 
Poland (margarine and fat spreads). In most other European countries, vitamin D 
fortification is voluntary under EU legislation or equivalent national 
legislation, depending on market uptake. Food vehicles vary regionally, with 
Northern Europe extending fortification beyond fats to include fluid milk and 
plant-based drinks, whereas other regions mainly fortify margarines, cereals, 
dairy products, and plant-based beverages. Fortification levels also differ, 
with some countries specifying maximal or exact levels, while others lack 
national standards. Data on fortified foods are limited in several Central and 
Southern European countries. Modeling indicates that multi-vehicle fortification 
is more effective than single-vehicle approaches, safely increasing population 
intakes while reducing deficiency prevalence.
CONCLUSIONS: Vitamin D fortification policies across Europe are highly 
heterogeneous. Most countries rely on voluntary approaches, which provide 
limited coverage. Strengthening policy through mandatory and well-coordinated 
multi-vehicle strategies, informed by modeling and population-based studies, can 
improve vitamin D intake, reduce deficiency prevalence, and enhance health 
equity.

DOI: 10.3390/nu18081194
PMCID: PMC13119029
PMID: 42075008 [Indexed for MEDLINE]

Conflict of interest statement: The authors declare no conflicts of interest.


4. J Nutr Sci Vitaminol (Tokyo). 2026;72(2):189-195. doi: 10.3177/jnsv.72.189.

Effects of Consuming a Docosahexaenoic Acid-Enriched Milk Beverage on the 
Erythrocyte Omega-3 Index and Stress Index in Healthy Japanese Schoolchildren 
Aged 12 to 15 Years.

Ichinose T(1), Tachibana N(1), Matsuzaki K(2), Hashimoto A(3), Iwaoka E(3), Kato 
S(4), Ohata S(4), Ohno M(4), Nagai A(5), Hashimoto M(6).

Author information:
(1)Research Institute for Creating the Future, Fuji Oil Co., Ltd.
(2)Department of Environmental Physiology, Faculty of Medicine, Shimane 
University.
(3)Oils & Fats R&D Department, Fuji Oil Co., Ltd.
(4)Kato Hospital, Jinjukai Healthcare Corporation.
(5)Department of Internal Medicine III, Faculty of Medicine, Shimane University.
(6)Shimane Rehabilitation Gakuin.

Docosahexaenoic acid (DHA) and eicosapentaenoic acid (EPA) are polyunsaturated 
fatty acids that are essential to human health. DHA in particular plays an 
important role in the development and maintenance of the brain. As seafood 
consumption has decreased sharply in Japan, concerns have grown about brain 
growth and development in infants and school-aged children. It is therefore 
critical to investigate the effects of DHA/EPA intake on the brain functions of 
school-aged children. The International Society for the Study of Fatty Acids and 
Lipids has proposed the erythrocyte omega-3 index as the standard measure of 
omega-3 fatty acid uptake. In this pilot study, we investigated the effects of 
consuming a DHA-enriched milk beverage fortified with 297 mg DHA and 137 mg EPA 
on the erythrocyte omega-3 and stress indices in schoolchildren. Intake of the 
DHA-enriched milk beverage for 12 wk led to an increase in the erythrocyte 
omega-3 index from 6.46% to 7.67% in children aged 12 to 15 y. Moreover, it led 
to a significant decrease in the "helplessness" score, which is an item on the 
stress response scale. These results suggest that daily intake of DHA, even in 
low amounts, could alleviate stress in school-aged children, potentially 
preventing various problematic behaviors.

DOI: 10.3177/jnsv.72.189
PMID: 42055724 [Indexed for MEDLINE]


5. Nutr Clin Pract. 2026 Apr 29. doi: 10.1002/ncp.70127. Online ahead of print.

Growth and human milk intake outcomes from postdischarge feedings among 
very-low-birth-weight preterm infants: A systematic review.

Rebele T(1), Gomes A(1), Newkirk M(1), Ghajar M(1), Byham-Gray L(1), Ziegler 
J(1).

Author information:
(1)Department of Clinical and Preventive Nutrition Sciences, Rutgers, The State 
University of New Jersey, New Brunswick, New Jersey, USA.

BACKGROUND: Preterm infants of <34 weeks gestation or <1500 g are at risk for 
postnatal growth failure and often require nutrition support after discharge. 
Despite expert recommendations for postdischarge human milk (HM) fortification 
or nutrient-enriched formula feeding, the effects of current feeding strategies 
on growth and HM intake remain unclear. This systematic review evaluated the 
impact of postdischarge fortified expressed HM or direct breastfeeding (BF) with 
postdischarge formula (PDF) bottle feeds compared to unfortified HM or BF with 
standard term formula bottle feeds on growth and HM intake outcomes through 6 
and 12 months corrected age (CA).
METHODS: A systematic search of eight databases and grey literature sources was 
conducted for studies published in English since 2012 identifying 3146 studies. 
Risk of bias was assessed using standardized Cochrane RoB 2 and ROBINS-I tools. 
Meta-analysis was not conducted due to heterogeneity in interventions, outcome 
timing, and outcome metrics; results were narratively synthesized.
RESULTS: Four studies (n = 2 randomized controlled trials, n = 2 cohorts), 
including 548 preterm infants, met eligibility criteria. Two studies showed 
significantly greater weight gain, and one showed significantly greater head 
circumference in the fortified groups at 6 months CA compared to the unfortified 
groups. Length differences were not significant. Three studies reporting HM 
intake found comparable or greater HM intake in the fortified groups.
CONCLUSION: Current evidence suggests that postdischarge fortified HM or direct 
BF with supplemental PDF bottle feeds may support improved weight and head 
circumference outcomes without compromising HM intake. However, methodological 
variability highlights the need for standardized, prospective studies.

© 2026 The Author(s). Nutrition in Clinical Practice published by Wiley 
Periodicals LLC on behalf of American Society for Parenteral and Enteral 
Nutrition.

DOI: 10.1002/ncp.70127
PMID: 42051106


6. J Perinatol. 2026 Apr 27. doi: 10.1038/s41372-026-02695-w. Online ahead of 
print.

Growth and safety evaluation in very low birth weight infants receiving an 
exclusive human milk diet: a phase III randomized control trial in Japan.

Mizuno K(1), Miyazawa T(2), Kondo U(3), Nishikubo T(4), Yamamoto Y(5), Nakano 
Y(2), Hiroma T(6), Ikeda K(7), Murase M(2), Jimi H(8), Hokuto I(9), Miyata 
M(10).

Author information:
(1)Department of Pediatrics, Showa Medical University, Tokyo, Japan. 
.
(2)Department of Pediatrics, Showa Medical University, Tokyo, Japan.
(3)Department of Neonatology, Tokyo Metropolitan Bokutoh Hospital, Tokyo, Japan.
(4)Division of Neonatal Intensive Care, Center of Maternal-Fetal Medicine, Nara 
Medical University, Kashihara, Japan.
(5)Department of Neonatology, Gifu Prefectural General Medical Center, 
Noisshiki, Japan.
(6)Division of Neonatology, Nagano Children's Hospital, Azumino, Japan.
(7)Division of Neonatology, Department of Pediatrics, Saitama City Hospital, 
Saitama, Japan.
(8)Department of Neonatology, Takatsuki General Hospital, Osaka, Japan.
(9)Department of Pediatrics, St. Marianna University, Kawasaki, Japan.
(10)Department of Pediatrics, Fujita Health University School of Medicine, 
Toyoake City, Japan.

OBJECTIVE: To determine safety and efficacy of exclusive human milk diet (EHMD) 
for very low birth weight (VLBW, <1500 g) infants.
DESIGN: Prospective, controlled, open trial of VLBW infants randomized to EHMD 
(mother's own milk [MOM]/donor milk [DM] with human milk-based fortifiers) or 
Standard Diet (MOM/DM with cow milk-based fortifier/formula). Outcomes included 
superiority of weight gain velocity from birth to 34 weeks gestational age, time 
to full feeds, and safety.
RESULT: In total, 147 infants received EHMD (n = 77) or Standard Diet (n = 70). 
The EHMD group had superior mean weight gain velocity versus the Standard Diet 
(13.44 vs 11.96 g/kg/day, p = 0.006) and earlier mean time to full feeds (20.0 
vs 25.9 days, p = 0.03); adverse events were not significantly different (32.5% 
vs 21.4%, p = 0.13).
CONCLUSION: EHMD is viable for management of VLBW infants, providing improved 
growth without reduced safety.
TRIAL REGISTRATION AND ID: Japan Registry of Clinical Trials: jRCT2031210384.

© 2026. The Author(s).

DOI: 10.1038/s41372-026-02695-w
PMID: 42045666

Conflict of interest statement: Competing interests: The authors declare no 
competing interests. Ethics approval: This study was conducted with strict 
adherence to the ethical principles outlined in Good Clinical Practice and the 
standards set by the International Council for Harmonisation (ICH) Tripartite 
Guideline, as well as the Japan Good Clinical Practice regulations. Moreover, 
this trial upholds the ethical considerations and patient rights stipulated in 
the Declaration of Helsinki and was performed in accordance with the ethical 
standards of the Independent Ethics Committee at each participating site, which 
approved all related materials and informed consent forms, as follows: 1. Showa 
Medical University Hospital Institutional Review Board. 2. Showa Medical 
University Koto Toyosu Hospital Institutional Review Board. 3. Showa Medical 
University Northern Yokohama Hospital Institutional Review Board. 4. Nara 
Medical University Hospital Institutional Review Board. 5. Saitama City Hospital 
Institutional Review Board. 6. The Central Institutional Review Board for the 
Fujita Health University Hospitals. 7. Bokutoh Hospital Clinical Trial Review 
Committee. 8. Review Board of Human Rights and Ethics for Clinical Studies. 9. 
Social Medical Corporation Aijinkai Group Takatsuki General Hospital 
Institutional Review Board. 10. Gifu Prefectural General Medical Center 
Institutional Review Board. 11. St. Marianna University Group Institutional 
Review Board.


7. Int J Biol Macromol. 2026 May;362:152175. doi: 10.1016/j.ijbiomac.2026.152175. 
Epub 2026 Apr 23.

Digestive properties and bioactive peptide release of lactoferrin-fortified 
elderly milk powder under different processes: An in vitro study based on the 
dynamic gastrointestinal digestive system of the elderly.

Li C(1), Zhang W(1), Wang M(1), Li H(1), Li H(1), Yu J(2).

Author information:
(1)College of Food Science and Engineering, Tianjin University of Science & 
Technology, Tianjin, 300457, China.
(2)College of Food Science and Engineering, Tianjin University of Science & 
Technology, Tianjin, 300457, China. Electronic address:.

In this study, an in vitro dynamic digestion model simulating the 
gastrointestinal conditions of the elderly was used to compare the effects of 
dry mixing (F1) and wet mixing (F2) on the digestive characteristics of 
lactoferrin (Lf) fortified elderly milk powder. During the gastric digestion 
phase, F2 exhibited higher protein digestibility and lower gastric retention, 
and the molecular weight distribution indicated a more complete degree of 
protein hydrolysis. The addition of Lf enhanced the antioxidant activity of the 
digestion products and promoted peptide release during the late digestion phase. 
Notably, F1_Lfmax released more Lf fortified peptides than F2_Lfmax. A total of 
six peptides with bioactivity scores >0.4 were identified, and they were 
predicted to possess antioxidant, antibacterial, anticancer, and 
immunomodulatory properties. These findings suggest that the wet-dry blending 
process can improve the physicochemical properties and digestibility of the 
elderly milk powder, while the dry blending process may be more conducive to the 
release of bioactive peptides during protein digestion. However, these results 
are based solely on in vitro simulation analyses and require further in vivo 
validation to confirm their physiological relevance. This study provides 
preliminary insights into the effects of processing methods on the digestibility 
of elderly milk powder and offers a reference for product formulation 
optimization.

Copyright © 2026 Elsevier B.V. All rights reserved.

DOI: 10.1016/j.ijbiomac.2026.152175
PMID: 42034144 [Indexed for MEDLINE]

Conflict of interest statement: Declaration of competing interest No conflict of 
interest exits in the submission of this manuscript.


8. Clin Exp Pediatr. 2026 May;69(5):428-436. doi: 10.3345/cep.2025.02509. Epub 2026 
Apr 22.

Human milk macronutrient composition and intake in relation to preterm infant 
growth: a cohort study.

Mekangkul E(1)(2), Visuthranukul C(2), Sirisabya A(3), Chitsinchayakul T(3), 
Punnahitananda S(3), Chomtho S(2).

Author information:
(1)Division of Nutrition, Department of Pediatrics, King Chulalongkorn Memorial 
Hospital, The Thai Red Cross Society, Bangkok, Thailand.
(2)Center of Excellence in Pediatric Nutrition, Division of Nutrition, 
Department of Pediatrics, Faculty of Medicine, Chulalongkorn University, 
Bangkok, Thailand.
(3)Division of Neonatology, Department of Pediatrics, Faculty of Medicine, 
Chulalongkorn University, Bangkok, Thailand.

BACKGROUND: Individualized targeted fortification based on human milk (HM) 
analysis reportedly achieves optimal outcomes in preterm infants. Therefore, 
understanding the effects of macronutrients in HM on preterm infant growth is 
essential.
PURPOSE: This study aimed to determine the association between HM macronutrients 
and the growth of preterm infants. We also compared macronutrient intake data 
obtained from an HM analyzer (HMA)-based calculation with those derived from a 
reference-based calculation.
METHODS: This prospective-retrospective cohort study included infants born at 34 
weeks' gestation or less. HM samples were collected weekly for up to 4 weeks or 
until discharge, whichever occurred first. Clinical outcomes were recorded. The 
macronutrient composition was analyzed using midinfrared HMA. Associations were 
determined using a linear regression model.
RESULTS: Of 121 preterm infants, 65 (51.2%) were male. A total of 200 HM samples 
were analyzed. Fat composition showed a significant positive association with 
weight gain velocity, with an adjusted unstandardized coefficient (aB) of 3.07 
(95% confidence interval [CI], 0.22-5.93). Total protein and fat intakes were 
positively associated with weight gain (aB, 3.41; 95% CI, 0.83-5.98; and aB, 
7.07; 95% CI, 1.73-12.42, respectively). When using the HMA-based calculation, 
protein intake was lower and carbohydrate intake was higher throughout the 
4-week period compared with those obtained based on the reference-based 
calculation.
CONCLUSION: Higher protein and fat intakes could potentially enhance weight gain 
in preterm infants. These findings provide further evidence to support the 
concept of individualized HM fortification. Our findings underscore the 
importance of using HMA-based methods to calculate macronutrient intakes among 
preterm infants.

DOI: 10.3345/cep.2025.02509
PMID: 42019927


9. J Perinatol. 2026 Apr 21. doi: 10.1038/s41372-026-02679-w. Online ahead of 
print.

Macronutrient and bioactive profiles of donor milk differ in commercial vs 
non-profit milk banks.

Tarkleson T(1), Ackley D(2), Cole L(3), Srivastava M(2), Scholer L(2), Gale 
C(2), Meyers J(2), D' Angio C(2), Young B(3).

Author information:
(1)University of Rochester School of Medicine and Dentistry, Rochester, NY, USA. 
.
(2)Department of Pediatrics - Neonatology, University of Rochester School of 
Medicine and Dentistry, Rochester, NY, USA.
(3)Department of Pediatrics - Breastfeeding and Lactation Medicine, University 
of Rochester School of Medicine and Dentistry, Rochester, NY, USA.

Update of
    Res Sq. 2025 Oct 07:rs.3.rs-7652186. doi: 10.21203/rs.3.rs-7652186/v1.

OBJECTIVE: Characterize variation of macronutrient and bioactive composition 
between levels of human milk-derived human milk fortifier (HM-HMF), and between 
donor human milk (DHM) from commercial vs non-profit banks.
STUDY DESIGN: Macronutrients, Immunoglobulin A (IgA), and cortisol levels were 
measured in 10 lots of each HM-HMF product (20, 24, 26, 28, and 30 (n = 5) 
kcal/oz), including both "ready to feed" products and fortifiers reconstituted 
in donor milk.
RESULTS: Caloric density, protein, fat, carbohydrate, and total IgA 
concentration generally increased with the level of HM-HMF labeled caloric 
density, while cortisol concentration did not. Protein concentration of DHM did 
not differ between commercial and non-profit milk banks. Carbohydrates, fat, 
calories, and cortisol were higher, and IgA was lower in DHM from a commercial 
milk bank. Lot-to-lot variability of all components, besides cortisol, was lower 
in commercial DHM.
CONCLUSION: Macronutrient composition and variability differ in DHM from 
commercial vs non-profit milk banks.

© 2026. The Author(s), under exclusive licence to Springer Nature America, Inc.

DOI: 10.1038/s41372-026-02679-w
PMID: 42014842

Conflict of interest statement: Competing interests: The authors declare no 
competing interests.


10. Semin Fetal Neonatal Med. 2026 Apr 2:101711. doi: 10.1016/j.siny.2026.101711. 
Online ahead of print.

Human milk fortification in preterm infants: Navigating evidence, practice, and 
controversy.

Hair AB(1), Poindexter BB(2).

Author information:
(1)Division of Neonatology, Department of Pediatrics, Baylor College of 
Medicine, Texas Children's Hospital, One Baylor Plaza, Houston, TX, 77030, USA. 
Electronic address:.
(2)Division of Neonatology, Department of Pediatrics, Children's Healthcare of 
Atlanta and Emory University School of Medicine, 2220 N. Druid Hills Rd. NE, 
Atlanta, GA, 30329, USA.

Human milk is universally recognized as the optimal source of nutrition for 
preterm infants, providing immunologic, bioactive, and enzymatic components that 
provide health benefits and are associated with decreased morbidities of 
prematurity. However, fortification of both maternal breast milk (MBM) and donor 
breast milk (DBM) is needed to meet the elevated metabolic needs and nutrient 
requirements of preterm infants. This narrative review, reflecting the content 
presented, audience questions, and the roundtable discussion from the 2025 
Neonatal Insights Conference, synthesizes current evidence and literature to 
evaluate the rationale, composition, and outcomes associated with differing 
fortification approaches. Randomized controlled trials evaluating fortifier type 
remain limited to small studies with high MBM usage. Future research must focus 
on trials evaluating fortifier type in infants who receive MBM and those that 
receive primarily DBM. Advancing fortification practices requires individualized 
nutrition planning and maternal breastfeeding support to achieve optimal growth 
and developmental trajectories for preterm infants.

Copyright © 2026 Elsevier Ltd. All rights reserved.

DOI: 10.1016/j.siny.2026.101711
PMID: 42002446

Conflict of interest statement: Declaration of competing interest None.


11. Semin Perinatol. 2026 Apr 17:152236. doi: 10.1016/j.semperi.2026.152236. Online 
ahead of print.

Enteral nutrition in neonates: current evidence from clinical trials and 
evolving strategies.

Romero-Lopez M(1), Salas AA(2).

Author information:
(1)Division of Perinatal-Neonatal Medicine, Department of Pediatrics, McGovern 
Medical School at UTHealth, Houston, TX, USA; Institute for Clinical Research 
and Learning Health Care, McGovern Medical School at UTHealth, Houston, TX, USA.
(2)Division of Neonatology, Department of Pediatrics, Heersink School of 
Medicine, University of Alabama at Birmingham, Birmingham, AL, USA. Electronic 
address:.

Despite advances in neonatal intensive care and increased availability of 
randomized clinical trial data, optimal enteral nutrition strategies remain 
debated, with significant gaps between evidence and practice. We reviewed 
current evidence for enteral feeding in neonates. Recent clinical trials support 
the early progression of enteral feeding within 96 h of birth and faster 
advancement rates of 30-40 mL/kg/day in preterm infants. Evidence demonstrates 
that these strategies reduce the time to full feeds and decrease the risk of 
sepsis without increasing the risk of necrotizing enterocolitis. A recent 
multicenter trial confirmed that, when maternal milk supply is limited, donor 
human milk reduces necrotizing enterocolitis incidence compared to formula, 
however, without neurodevelopmental advantages. Multiple trials found no 
superiority of human milk-based over bovine-based fortifiers. Abandoning routine 
gastric residual assessment reduces time to full feeds without increasing 
complications, but a large confirmatory trial is underway. Evidence now supports 
low-volume feeding during therapeutic hypothermia for hypoxic-ischemic 
encephalopathy, with fed infants achieving full feeds earlier and having a 
shorter hospital stay. For congenital heart disease, hemodynamically stable 
infants can safely receive minimal human milk feeding pre and post-operatively 
within 5-7 days. Future trials need to focus on precision nutrition, optimal 
fortification strategies, and long-term effects.

Copyright © 2026 Elsevier Inc. All rights reserved.

DOI: 10.1016/j.semperi.2026.152236
PMID: 41997817

Conflict of interest statement: Disclosure AAS has a patent on an instrumented 
feeding bottle. AAS has received honoraria for participation in Mead Johnson 
Nutrition advisory board meetings and honoraria for participation in symposiums 
sponsored by Mead Johnson Nutrition and Abbott. MR-L has no conflicts of 
interest relevant to this article to disclose.


12. Ethiop J Health Sci. 2025 Jul;35(4):227-236. doi: 10.4314/ejhs.v35i4.2.

Consumption of Vitamin A Rich Foods and Their Primary Dietary Sources among 
Rural Preschoolers in the Gambella Region, Southwest Ethiopia.

Tamiru D(1), Dak GK(1), Wondafrash B(1).

Author information:
(1)Department of Nutrition and Dietetics, Faculty of Public Health, Jimma 
University, Ethiopia.

BACKGROUND: Globally, the consumption of vitamin A-rich foods remains a 
significant public health challenge, particularly in sub-Saharan Africa, where 
access to these foods is limited. Preschool-aged children in rural areas are 
especially vulnerable to vitamin A deficiency due to poor dietary diversity and 
limited access to healthcare services. In Ethiopia, there is a scarcity of 
research on vitamin A intake among rural preschoolers, particularly in the 
Gambella Region. The objective of this study was to assess the consumption 
patterns and primary dietary sources of vitamin A-rich foods among rural 
preschool children in the Gambella Region of Ethiopia.
METHODS: A community-based cross-sectional study was conducted among 381 
preschool children in the Gambella Region. Data were collected using the Helen 
Keller International (HKI) food frequency questionnaire and a 24-hour dietary 
recall. Statistical analysis was performed using SPSS version 26.0, with 
findings presented through frequency tables and graphs to illustrate dietary 
trends.
RESULTS: Of the 381 participants, 380 (99.7%) completed the study. The majority 
of children (74.3%) ate two meals per day, predominantly cereals and legumes. 
Only 32.4% consumed vitamin A-rich foods, with particularly low intake reported 
for milk (2.9%), dark green leafy vegetables (6.1%), fish (5%), and fortified 
margarine (0.5%). Over 75% of children had inadequate vitamin A intake (P < 
0.01), and 78.4% of caregivers lacked awareness of vitamin A-rich food sources. 
Nearly half of households (48.1%) relied on personal production for these foods, 
and 54.9% indicated that availability was seasonal.
CONCLUSION: The study highlights critically low consumption of vitamin A-rich 
foods among rural preschool children in the Gambella Region, placing them at 
increased risk of vitamin A deficiency and related health complications such as 
impaired vision and weakened immunity. To address this issue, targeted nutrition 
education and community awareness programs are essential, alongside policy 
interventions to improve food access and dietary diversity.

© 2025 Dessalegn Tamiru, et al.

DOI: 10.4314/ejhs.v35i4.2
PMCID: PMC13077171
PMID: 41987863 [Indexed for MEDLINE]

Conflict of interest statement: Competing Interests: The authors declare that 
this manuscript was approved by all authors in its form and that no competing 
interest exists.


13. Cell Rep Med. 2026 Apr 21;7(4):102755. doi: 10.1016/j.xcrm.2026.102755. Epub 
2026 Apr 13.

Breastfeeding may lessen socioeconomic disparities in child health through 
differences in the infant gut microbiome.

Dai DLY(1), Manus MB(2), Hoskinson C(3), Jiang J(4), Sbihi H(5), Miliku K(6), 
Campisi SC(7), Korczak DJ(8), Duan Q(9), Moraes TJ(10), Mandhane PJ(11), Finlay 
BB(12), Simons E(13), Lishman H(14), Patrick DM(5), Subbarao P(15), Azad MB(16), 
Chawes B(17), Bønnelykke K(17), Sørensen SJ(18), Thorsen J(17), Stokholm J(19), 
Petersen C(20), Turvey SE(21).

Author information:
(1)Department of Pediatrics, BC Children's Hospital, University of British 
Columbia, Vancouver, BC, Canada.
(2)Department of Anthropology, University of Texas at San Antonio, San Antonio, 
TX, USA.
(3)Department of Pediatrics, BC Children's Hospital, University of British 
Columbia, Vancouver, BC, Canada; Department of Microbiology and Immunology, 
University of British Columbia, Vancouver, BC, Canada.
(4)COPSAC, Copenhagen Prospective Studies on Asthma in Childhood, Copenhagen 
University Hospital - Herlev and Gentofte, Copenhagen, Denmark; Department of 
Food Science, Faculty of Science, University of Copenhagen, Frederiksberg C, 
Denmark.
(5)BC Centre for Disease Control, Vancouver, BC, Canada; School of Population 
and Public Health, University of British Columbia, Vancouver, BC, Canada.
(6)Department of Nutritional Sciences, Temerty Faculty of Medicine, University 
of Toronto, Toronto, ON, Canada; Department of Medicine, Faculty of Health 
Sciences, McMaster University, Hamilton, ON, Canada.
(7)Nutrition and Dietetics Program, Clinical Public Health Division, Dalla Lana 
School of Public Health, University of Toronto, Toronto, ON, Canada; Department 
of Psychiatry, The Hospital for Sick Children, Toronto, ON, Canada.
(8)Department of Psychiatry, The Hospital for Sick Children, Toronto, ON, 
Canada; Department of Psychiatry, University of Toronto, Toronto, ON, Canada.
(9)Department of Biomedical and Molecular Sciences, Queen's University, 
Kingston, ON, Canada; School of Computing, Queen's University, Kingston, ON, 
Canada.
(10)Department of Pediatrics, The Hospital for Sick Children, Toronto, ON, 
Canada.
(11)Department of Pediatrics, University of Alberta, Edmonton, AB, Canada; 
Faculty of Medicine and Health Sciences, UCSI University, Kuala Lumpur, 
Malaysia.
(12)Department of Microbiology and Immunology, University of British Columbia, 
Vancouver, BC, Canada; Michael Smith Laboratories, UBC, Vancouver, BC, Canada; 
Department of Biochemistry and Molecular Biology, University of British 
Columbia, Vancouver, BC, Canada.
(13)Section of Allergy and Immunology, Department of Pediatrics and Child 
Health, University of Manitoba, Winnipeg, MB, Canada.
(14)BC Centre for Disease Control, Vancouver, BC, Canada.
(15)Department of Medicine, Faculty of Health Sciences, McMaster University, 
Hamilton, ON, Canada; Department of Pediatrics, The Hospital for Sick Children, 
Toronto, ON, Canada; Dalla Lana School of Public Health, University of Toronto, 
Toronto, ON, Canada.
(16)Section of Allergy and Immunology, Department of Pediatrics and Child 
Health, University of Manitoba, Winnipeg, MB, Canada; Department of Food and 
Human Nutritional Sciences, University of Manitoba, Winnipeg, MB, Canada; 
Manitoba Interdisciplinary Lactation Centre (MILC), Children's Hospital Research 
Institute of Manitoba, Winnipeg, MB, Canada.
(17)COPSAC, Copenhagen Prospective Studies on Asthma in Childhood, Copenhagen 
University Hospital - Herlev and Gentofte, Copenhagen, Denmark; Department of 
Clinical Medicine, Faculty of Health and Medical Sciences, University of 
Copenhagen, Copenhagen, Denmark.
(18)Section of Microbiology, University of Copenhagen, Copenhagen, Denmark.
(19)COPSAC, Copenhagen Prospective Studies on Asthma in Childhood, Copenhagen 
University Hospital - Herlev and Gentofte, Copenhagen, Denmark; Department of 
Food Science, Faculty of Science, University of Copenhagen, Frederiksberg C, 
Denmark; Department of Pediatrics, Slagelse Hospital, Slagelse, Denmark.
(20)Department of Pediatrics, BC Children's Hospital, University of British 
Columbia, Vancouver, BC, Canada. Electronic address:.
(21)Department of Pediatrics, BC Children's Hospital, University of British 
Columbia, Vancouver, BC, Canada. Electronic address:.

Lower familial socioeconomic status (SES) is linked to increased childhood 
disease risk. Since SES has no inherent biological basis, identifying how it 
becomes physiologically embedded is essential for equitable intervention. Using 
data from the Canadian CHILD birth cohort (n = 2,752) with replication in the 
Danish Copenhagen Prospective Studies on Asthma in Childhood 2010 (COPSAC2010) 
cohort (n = 681), we analyze modifiable pathways linking SES to child health and 
find that the infant gut microbiota plays a key mediating role. Breastfeeding is 
associated with a stabilized infant microbiota, buffering against environmental 
impacts and reducing health risks in lower SES contexts. The presence of 
Bifidobacterium infantis, enriched through breastfeeding, is linked to 
protection against adverse outcomes from SES. Together, these results suggest 
that improving breastfeeding rates and restoring breastfeeding-enriched 
microbes, like B. infantis, may help buffer early biological impacts of social 
inequality and support healthier trajectories for children growing up in 
industrialized settings.

Copyright © 2026 The Author(s). Published by Elsevier Inc. All rights reserved.

DOI: 10.1016/j.xcrm.2026.102755
PMCID: PMC13130633
PMID: 41980780 [Indexed for MEDLINE]

Conflict of interest statement: Declaration of interests M.B.A. has received 
speaker honoraria from Prolacta Biosciences (a human milk fortifier company), 
consulted for DSM Nutritional Products (a food ingredient company), and serves 
as an advisor for Tiny Health (an infant microbiome testing company).


14. Nutrients. 2026 Apr 7;18(7):1167. doi: 10.3390/nu18071167.

Effects of Fortified Formula Milk Supplementation on Neurocognitive Development 
and the Microbiota-Gut-Brain Axis in Preschool Children: A Cluster-Randomized, 
Double-Blind, Controlled Trial.

Gong Y(1)(2), Zhao X(3), Zhang Q(2), Yan X(2), Sun B(4), Li X(2), Han Q(2), Guan 
Y(2), Chen H(2), Li M(2), Guo J(2), Liu B(5), Wang R(2), Zhao B(1), Zhang Y(1), 
He J(2).

Author information:
(1)College of Food Science and Engineering, Gansu Agricultural University, 
Lanzhou 730070, China.
(2)Key Laboratory of Precision Nutrition and Food Quality, Department of 
Nutrition and Health, China Agricultural University, Beijing 100193, China.
(3)Dali Vocational and Technical College of Agriculture and Forestry, Dali 
671003, China.
(4)National Technology Innovation Center for Dairy, Hohhot 010110, China.
(5)Milk Powder BU, Inner Mongolia Yili Industrial Group Co., Ltd., Hohhot 
010110, China.

BACKGROUND/OBJECTIVES: The preschool period is critical for neurodevelopment, 
yet evidence investigating fortified formula's effect and potential 
microbiota-gut-brain axis mechanisms in this age group is limited. To evaluate 
fortified formula milk's effect on neurodevelopment and explore potential 
microbiota-gut-brain axis mechanisms in preschool children.
METHODS: In this 9-month cluster-randomized, double-blind, controlled trial, 120 
healthy children aged 3-6 years from four kindergarten classes were stratified 
by grade and randomly allocated (1:1) to receive either multi-nutrient fortified 
formula (intervention, n = 60) or standard control milk (n = 60). Neurocognitive 
function was assessed using the Wechsler Preschool and Primary Scale of 
Intelligence, Fourth Edition (WPPSI-IV). Safety was evaluated through 
anthropometry and blood biochemistry. Gut microbiota (16S rRNA sequencing) and 
fecal metabolomes (untargeted LC-MS) were analyzed at baseline and 9 months.
RESULTS: The intention-to-treat (ITT) analysis showed no significant difference 
in Full Scale Intelligence Quotient (adjusted mean difference: 1.05 points; 95% 
CI: -1.42, 3.52; p = 0.400). However, the intervention group significantly 
improved the Processing Speed Index (adjusted mean difference: 5.91 points; 95% 
CI: 1.88, 9.93; p = 0.004), increased gut microbial alpha diversity (Shannon 
index) and Bifidobacterium abundance. Metabolomic analysis revealed elevated 
fecal 2-hydroxybutyric acid (2-HB), a marker of propanoate metabolism. Increases 
in both Bifidobacterium and 2-HB levels showed a positive association with PSI 
improvement (both p < 0.05). All children maintained normal growth and safety 
parameters.
CONCLUSIONS: Fortified formula milk improved processing speed in preschoolers, a 
benefit associated with gut ecosystem modulation characterized by 
Bifidobacterium enrichment and upregulated microbial propanoate metabolism. 
These results offer preliminary evidence for the role of the 
microbiota-gut-brain axis in nutritional cognitive programming during early 
childhood. (Clinical Trial Registry: ChiCTR2400084211).

DOI: 10.3390/nu18071167
PMCID: PMC13074376
PMID: 41978217 [Indexed for MEDLINE]

Conflict of interest statement: Biao Liu is an employee of Inner Mongolia Yili 
Industrial Group Co., Ltd., which provided the intervention and control milk 
products used in this study. As a co-author, B.L. participated in the research 
as detailed in the Author Contributions section. The authors declare that this 
commercial affiliation did not inappropriately influence the interpretation of 
the study results. All other authors declare no conflicts of interest.


15. Nutrients. 2026 Mar 24;18(7):1026. doi: 10.3390/nu18071026.

Rationale, Design, and Participant Baseline Characteristics of a Parallel 
Randomized Trial of the Effect of Replacing SSBs with Cow's Milk Versus Soymilk 
on Intrahepatocellular Lipid and Other Cardiometabolic Risk Factors in Adults 
with Obesity Who Consume Sugar-Sweetened Beverages: The Soy Treatment Evaluation 
for Metabolic health (STEM) Trial.

Erlich MN(1)(2), Ghidanac D(1)(2), Blanco Mejia S(1)(2), Ayoub-Charette S(1)(2), 
Vittes Combe C(1)(2), Khan TA(1)(2), Ramdath D(1)(2), Crewson H(2)(3), Beck 
A(2)(4)(5), Silva C(1)(2), Ramdath DD(6), Metherel AH(1), Leiter 
LA(1)(2)(7)(8)(9), Bazinet RP(1), Kendall CWC(1)(2)(6), Jenkins 
DJA(1)(2)(7)(8)(9), Chiavaroli L(1)(2)(7), Sievenpiper JL(1)(2)(7)(8)(9).

Author information:
(1)Department of Nutritional Sciences, Temerty Faculty of Medicine, University 
of Toronto, Toronto, ON M5S 1A8, Canada.
(2)Toronto 3D Knowledge Synthesis and Clinical Trials Unit, Clinical Nutrition 
and Risk Factor Modification Centre, St. Michael's Hospital, Toronto, ON M5B 
1W8, Canada.
(3)Department of Family Relations and Applied Nutrition, University of Guelph, 
Guelph, ON N1G 2W1, Canada.
(4)Clinical Nutrition Department, Civic Campus, The Ottawa Hospital, Ottawa, ON 
K1Y 4E9, Canada.
(5)Division of Cardiac Prevention and Rehabilitation, University of Ottawa Heart 
Institute, Ottawa, ON K1Y 4W7, Canada.
(6)College of Pharmacy and Nutrition, University of Saskatchewan, Saskatoon, SK 
S7N 5E5, Canada.
(7)Li Ka Shing Knowledge Institute, St. Michael's Hospital, Toronto, ON M5B 1A6, 
Canada.
(8)Department of Medicine, Temerty Faculty of Medicine, University of Toronto, 
Toronto, ON M5S 1A8, Canada.
(9)Division of Endocrinology and Metabolism, Department of Medicine, St. 
Michael's Hospital, Toronto, ON M5B 1W8, Canada.

Background/Objectives: Liver fat represents an early metabolic lesion in the 
development of diabetes and its cardiometabolic complications. Diets high in 
free sugars, particularly from sugar-sweetened beverages (SSBs), are associated 
with abdominal obesity and increased cardiometabolic risk, prompting global 
guidelines to limit SSBs as a major public health strategy. Low-fat cow's milk 
is promoted as the preferred caloric replacement strategy for SSBs due to its 
high nutritional value and cardiometabolic advantages. Fortified soymilk is a 
plant-based alternative with approved health claims for cholesterol and coronary 
heart disease risk reduction that offers an equivalent nutritional value to 
cow's milk. However, given concerns about its classification as an 
ultra-processed food (UPF), it is unclear whether soymilk offers comparable 
metabolic health benefits to milk as part of clinical and public health 
strategies to reduce SSB intake. The Soy Treatment Evaluation for Metabolic 
(STEM) health trial seeks to evaluate the impact of replacing SSBs with either 
2% soymilk or 2% cow's milk on liver fat and other cardiometabolic risk factors 
in habitual adult consumers of SSBs with obesity. Methods: The STEM trial is a 
24-week, pragmatic, 3-arm, parallel, randomized trial. We recruited adults with 
obesity (high BMI plus high waist circumference based on ethnic specific 
cut-offs) consuming ≥1 SSB/day. Participants were randomized to one of three 
groups based on their usual SSB intake at baseline (servings/day): continued SSB 
(355 mL can) intake; replacement with fortified, sweetened 2% soymilk (250 mL); 
or replacement with 2% cow's milk (250 mL). The primary outcome is the change in 
intrahepatocellular lipid (IHCL) measured by 1H-MRS at 24 weeks. Hierarchical 
testing will be done to reduce the familywise error rate. The superiority of 
cow's milk to SSBs will be assessed first to establish assay sensitivity. If 
superiority is established, then the non-inferiority of soymilk to cow's milk 
will be assessed using a pre-specified non-inferiority margin of 1.5% IHCL units 
(assessed by difference of means using a 90% confidence interval [CI]). Analyses 
will be conducted according to the intention-to-treat (ITT) principle using 
inverse probability weighting (IPW) for superiority testing and per-protocol 
analyses for non-inferiority testing, using ANCOVA adjusted for age, sex, 
metabolic dysfunction-associated steatotic liver disease (MASLD) status, 
medication use, intervention dose, and baseline levels. We hypothesize that 
soymilk will be non-inferior to cow's milk (Clinicaltrials.gov NCT05191160). 
Results: Recruitment began in November 2021. A total of 3050 individuals were 
screened. We randomized 186 participants (62 per group) between 19 April 2022 
and 16 April 2024. Participants are 57% male; with a mean [SD] age of 39.9 
[11.8] years; BMI of 34.6 [6.1] kg/m2, waist circumference of 112.6 [13.8] cm; 
IHCL of 10.0 [8.2] % with 64.1% meeting the criteria for MASLD; and SSBs intake 
of 2.3 [1.3] servings/day. Conclusions: Baseline characteristics were balanced 
across the study arms, with participants representing adults with a high-risk 
metabolic phenotype, and 64.1% meeting the criteria for MASLD. Findings will 
contribute to evidence on the cardiometabolic benefits of soymilk, informing 
clinical practice guidelines and public health policy.

DOI: 10.3390/nu18071026
PMCID: PMC13074930
PMID: 41978078 [Indexed for MEDLINE]

Conflict of interest statement: M.N.E. has received funding from the United 
Soybean Board (the United States Department of Agriculture [USDA] Soy 
“Check-off” Program), the Toronto 3D PhD Scholarship, the CIHR Canadian Graduate 
Scholarship Master’s Award, and the CIHR Graduate Scholarship Doctoral Award. 
D.G. has received funding from the Ontario Graduate Scholarship and the Toronto 
3D PhD Scholarship. S.A.-C. has received an honorarium from the International 
Food Information Council (IFIC) (for a talk on NNS and the microbiome) and from 
the Arkansas Children’s Hospital for participation in a “lumping vs. splitting” 
event. T.A.K. discloses receiving research grants from Diabetes Canada, the 
CIHR, the Institute for the Advancement of Food and Nutrition Sciences (IAFNS, 
formerly ILSI North America), the National Honey Board (USDA Checkoff Program), 
and the Toronto 3D Knowledge Synthesis and Clinical Trials Foundation. T.A.K. 
has received honoraria for speaking engagements or advisory roles from IAFNS, 
IFIC, the Calorie Control Council (CCC), the International Sweeteners 
Association (ISA), AmCham Dubai, and the Ontario Maple Syrup Producers’ 
Association (OMSPA). Additionally, T.A.K. has served as a consultant for 
Heartland Foods Product Group. H.C. has received funding from the Undergraduate 
Summer Research Opportunity Program (UROP) award and the Toronto 3D Summer 
Scholarship Top-up Award. A.B. has received funding from the Toronto 3D Summer 
Scholarship Award. CS has received funding from the CIHR Canadian Graduate 
Scholarship Master’s Award. D.D.R. has received research support from Pulse 
Canada, the Saskatchewan Pulse Growers Association, and the Ontario Bean Growers 
Association. A.H.M. has received research support from CIHR, CFI, NSERC, and was 
a co-applicant on a joint government/industry funder research grant with Natures 
Crops International. He is a Scientific Advisor for Natures Crops International 
and Benexia and is on the Board of Directors of the International Society for 
the Study of Fatty Acids and Lipids. R.P.B. has received industrial grants, 
including those matched by the Canadian government, and/or travel support or 
consulting fees largely related to work on brain fatty acid metabolism from 
Arctic Nutrition (Ørsta, Norway), Bunge Ltd. (St. Louis, MO, USA), DSM (Heerlen, 
The Netherlands), Fonterra Inc. (Auckland, New Zealand), Mead Johnson (Chicago, 
IL, USA), Natures Crops International (Raleigh, NC, USA), Nestec Inc. (Vevey, 
Switzerland), Pharmavite (West Hills, CA, USA), and Sancero Inc. (Fort Worth, 
TX, USA). RPB is on the executive of the International Society for the Study of 
Fatty Acids and Lipids and held a meeting on behalf of Fatty Acids and Cell 
Signaling, both of which rely on corporate sponsorship, and has given expert 
testimony in relation to supplements and the brain. C.W.C.K. has received grants 
or research support from the Advanced Food Materials Network, Agriculture and 
Agri-Foods Canada, Almond Board of California, American Pistachio Growers, 
Barilla (Parma, Italy), Calorie Control Council, Canadian Institutes of Health 
Research, Canola Council of Canada, International Nut and Dried Fruit Council, 
International Tree Nut Council Research and Education Foundation, Loblaw Brands, 
Pulse Canada, Saskatchewan Pulse Growers and Unilever; has received in-kind 
research support from the Almond Board of California, American Peanut Council, 
Barilla, California Walnut Commission, Kellogg Canada, Loblaw Companies 
(Brampton, ON, Canada), Quaker (PepsiCo) (New York, NY, USA), Primo (Toronto, 
ON, Canada), Unico (Concord, ON, USA), Unilever (London, UK), WhiteWave Foods 
(Denver, CO, USA); has received travel support or honorariums from the American 
Peanut Council, American Pistachio Growers, Barilla, California Walnut 
Commission, Canola Council of Canada, General Mills (Minneapolis, MN, USA), 
International Nut and Dried Fruit Council, International Pasta Organization, 
Loblaw Brands Ltd. (Brampton, ON, Canada), Nutrition Foundation of Italy, 
Oldways Preservation Trust, Paramount Farms (Lost Hills, CA, USA), Peanut 
Institute, Pulse Canada, Sabra Dipping (White Plains, NY, USA), Saskatchewan 
Pulse Growers, Sun-Maid (Kingsburg, CA, USA), Tate & Lyle (London, UK), Unilever 
and White Wave Foods; has served on the scientific advisory board for the 
International Tree Nut Council, International Pasta Organization, Lantmannen, 
McCormick Science Institute, Oldways Preservation Trust, Paramount Farms and 
Pulse Canada; is a member of the International Carbohydrate Quality Consortium, 
executive board member of the Diabetes and Nutrition Study Group of the European 
Association for the Study of Diabetes; is on the Clinical Practice Guidelines 
Expert Committee for Nutrition Therapy of the European Association for the Study 
of Diabetes; and is a director of the Toronto 3D Knowledge Synthesis and 
Clinical Trials Foundation. D.J.A.J. has received research grants from Heart and 
Stroke Foundation Grant-in-Aid, Diabetes Canada, Loblaw Companies Ltd. 
(Brampton, ON, Canada), the Almond Board of California, Soy Nutrition Institute 
(SNI), and the Canadian Institutes of Health Research (CIHR). He has received 
in-kind supplies for trials as research support from the Almond Board of 
California, Walnut Council of California, American Peanut Council, Barilla, 
Unilever, Unico, Primo, Loblaw Companies, Quaker (Pepsico), Pristine Gourmet, 
Bunge Limited, Kellogg Canada, and WhiteWave Foods. He has been on the speaker’s 
panel, served on the scientific advisory board and/or received travel support 
and/or honoraria for lectures/presentations from Diabetes and Nutrition Study 
Group (DNSG), Lawson Centre Nutrition Digital Series, 19th Annual Stare-Hegsted 
Lecture, 2024 Diabetes Canada Conference, Nutritional Fundamentals for Health 
(NFH)-Nutramedica, Saint Barnabas Medical Center, The University of Chicago, 
2020 China Glycemic Index (GI) International Conference, Atlantic Pain 
Conference, Academy of Life Long Learning, the Almond Board of California, 
Canadian Agriculture Policy Institute, the Loblaw Companies Ltd., the Griffin 
Hospital (for the development of the NuVal scoring system), the Coca-Cola 
Company, Epicure, Danone, Diet Quality Photo Navigation (DQPN), Better 
Therapeutics (FareWell), Verywell, True Health Initiative (THI), Heali AI Corp, 
Institute of Food Technologists (IFT), Soy Nutrition Institute (SNI), Herbalife 
Nutrition Institute (HNI), Herbalife International, Pacific Health Laboratories, 
Nutritional Fundamentals for Health (NFH), the Soy Foods Association of North 
America, the Nutrition Foundation of Italy (NFI), the Toronto Knowledge 
Translation Group (St. Michael’s Hospital), the Canadian College of Naturopathic 
Medicine, The Hospital for Sick Children, the Canadian Nutrition Society (CNS), 
and the American Society of Nutrition (ASN). He is a member of the International 
Carbohydrate Quality Consortium (ICQC). He is invited by the International 
Diabetes Federation (IDF) to join the committee on diabetes treatment and to 
take the lead in writing the dietary guidelines for the treatment of diabetes. 
His wife, Alexandra L Jenkins, is senior scientist of INQUIS Clinical Research 
Inc. (Clinical Research Organization), his 2 daughters, Wendy Jenkins and Amy 
Jenkins, have published a vegetarian book that promotes the use of the low 
glycemic index plant foods advocated here, The Portfolio Diet for Cardiovascular 
Risk Reduction (Academic Press/Elsevier 2020 ISBN:978-0-12-810510-8) and his 
sister, Caroline Brydson, received funding through a grant from the St. 
Michael’s Hospital Foundation to develop a cookbook for one of his studies. L.C. 
has received research support from the Canadian Institutes of Health Research 
(CIHR), Protein Industries Canada (a Government of Canada Global Innovation 
Clusters), The United Soybean Board (USDA soy “Checkoff” program), and the 
Alberta Pulse Growers Association and honoraria for lectures/presentations from 
the Arkansas Children’s Hospital and the Physicians Committee for Responsible 
Medicine. J.L.S. has received research support from the Canadian Foundation for 
Innovation, Ontario Research Fund, Province of Ontario Ministry of Research and 
Innovation and Science, Canadian Institutes of Health Research (CIHR), Diabetes 
Canada, American Society for Nutrition (ASN), Institute for the Advancement of 
Food and Nutrition Sciences (IAFNS), The United Soybean Board (USDA soy 
“Checkoff” program), Protein Industries Canada (a Government of Canada Global 
Innovation Cluster), Almond Board of California, European Fruit Juice 
Association, The Glycemic Control and Cardiovascular Disease in Type 2 Diabetes 
Fund at the University of Toronto (a fund established by the Alberta Pulse 
Growers), The Plant Protein Fund at the University of Toronto (a fund which has 
received contributions from IFF among other donors), The Plant Milk Fund at the 
University of Toronto (a fund established by the Karuna Foundation through Vegan 
Grants), and The Nutrition Trialists Network Fund at the University of Toronto 
(a fund established by donations from the Calorie Control Council, Physicians 
Committee for Responsible Medicine, and Login5 Foundation). He has received food 
donations to support randomized controlled trials from the Almond Board of 
California, California Walnut Commission, Danone, Mantra Pharma, Terra Delyssa, 
House Foods, General Mills, WK Kellogg Company, Quaker, Procter & Gamble (P&G), 
Abbott, and Dairy Farmers of Canada. He has received travel support, speaker 
fees and/or honoraria from Nestlé (Vevey, Switzerland), Abbott (Abbott Park, IL, 
USA), General Mills (Minneapolis, MN, USA), International Food Information 
Council (IFIC), International Sweeteners Association, Calorie Control Council, 
International Stevia Council, Mantra Pharma Inc. (Laval, QC, Canada), Chinese 
Institute of Food Science and Technology (CIFST), and Collaborative CME and 
Research Network (CCRN). He has or has had ad hoc consulting arrangements with 
Almond Board of California, Perkins Coie LLP (Seattle, WA, USA), Tate & Lyle, 
Ingredion (Westchester, IL, USA), and Brightseed (San Francisco, CA, USA). He is 
on the Clinical Practice Guidelines Expert Committees of Diabetes Canada, 
European Association for the study of Diabetes (EASD), Canadian Cardiovascular 
Society (CCS), and Obesity Canada/Canadian Association of Bariatric Physicians 
and Surgeons. He serves as an unpaid member of the Board of Trustees of IAFNS. 
He is Vice President—Clinical of the Canadian Nutrition Society (CNS), founding 
member of the International Carbohydrate Quality Consortium (ICQC), Executive 
Board Member of the Diabetes and Nutrition Study Group (DNSG) of the EASD, and 
Director of the Toronto 3D Knowledge Synthesis and Clinical Trials foundation. 
His spouse is an employee of AB InBev (Labatt Breweries of Canada, Toronto, ON, 
Canada). All other authors report no conflicts of interest.


16. Food Res Int. 2026 Jun 1;233(Pt 2):118943. doi: 10.1016/j.foodres.2026.118943. 
Epub 2026 Mar 15.

Global evidence on vitamins in human milk: A 2002-2022 systematic review with a 
focus on China and implications for maternal diet and infant formula.

Yang Y(1), Ni X(1), Liu N(1), Li Q(1), Fang Q(1), Szeto IM(2), Duan S(2), Wei 
T(1), Rao T(1), Deng Z(3), Li T(4), Li J(5).

Author information:
(1)State Key Laboratory of Food Science and Resources, Nanchang University, 
Nanchang, Jiangxi 330047, China.
(2)National Center of Technology Innovation for Dairy, Hohhot 010110, China; 
Inner Mongolia Yili Industrial Group Co., Ltd, Hohhot, PR China.
(3)State Key Laboratory of Food Science and Resources, Nanchang University, 
Nanchang, Jiangxi 330047, China; International Institute of Food Innovation, 
Nanchang University, Nanchang, Jiangxi 330051, China. Electronic address: 
.
(4)Inner Mongolia Dairy Technology Research Institute Co. Ltd., Hohhot 010110, 
China; National Center of Technology Innovation for Dairy, Hohhot 010110, China. 
Electronic address:.
(5)State Key Laboratory of Food Science and Resources, Nanchang University, 
Nanchang, Jiangxi 330047, China; International Institute of Food Innovation, 
Nanchang University, Nanchang, Jiangxi 330051, China. Electronic address: 
.

Current knowledge on vitamin composition of human milk in China remains 
fragmented, with most evidence derived from small-scale studies focusing on 
individual vitamins. To provide a comprehensive perspective relevant to 
nutrition science and infant formula design, this study systematically reviewed 
global trends in human milk vitamins over the past two decades (2002-2022), with 
attention to regional disparities, maternal dietary patterns, and local 
standards. Literature was retrieved from PubMed, Web of Science, ScienceDirect, 
Chongqing VIP Information, China National Knowledge Infrastructure (CNKI), and 
Wanfang databases following predefined inclusion criteria. Fifty-seven studies 
encompassing 9833 human milk samples from 33 countries were included. Across 
lactation, concentrations of water-soluble vitamins generally increased, whereas 
fat-soluble vitamins A (VA) and E (VE) exhibited a declining tendency. Compared 
with Chinese samples, non-Chinese human milk contained higher levels of vitamin 
B1 (VB1), vitamin B6 (VB6), VA, and VE; while vitamin B5 (VB5) was consistently 
higher in Chinese human milk. Temporal analysis within China revealed elevated 
levels of VA (2016-2017), B vitamins (2014), and VE (2022), likely reflecting 
shifts in maternal dietary intake and nutritional awareness. Concentrations of 
VA, VE, VB5, and vitamin VB7 (VB7) in Chinese human milk met national adequate 
intake (AI) recommendations for infants aged 0-6 months. However, these 
concentrations remained substantially lower than the levels specified in China's 
national standard for infant formula. Notably, vitamin D (VD) emerged as the 
nutrient most vulnerable to inadequacy among exclusively breastfed infants, 
indicating a persistent gap between physiological supply from human milk and 
recommended intake. These findings emphasize the need for nationally 
representative, lactation-stage-specific reference ranges for human milk 
vitamins in China. Such evidence is essential for optimizing maternal dietary 
strategies, for guiding rational vitamin fortification, and formulation of 
next-generation infant formulas that better reflect physiological nutrition.

Copyright © 2026 Elsevier Ltd. All rights reserved.

DOI: 10.1016/j.foodres.2026.118943
PMID: 41956646 [Indexed for MEDLINE]

Conflict of interest statement: Declaration of competing interest The authors 
declare that they have no known competing financial interests or personal 
relationships that could have appeared to influence the work reported in this 
paper.


17. Allergy. 2026 Apr 6. doi: 10.1111/all.70325. Online ahead of print.

Iron Physiology and Its Impact on Atopic Diseases: An EAACI Taskforce Report.

Roth-Walter F(1), Agache I(2), Cabanillas B(3), Canani RB(4)(5)(6), Comberiati 
P(7), Garn H(8), Gomez-Casado C(9), Hufnagl K(10), Khaleva E(11)(12), Milani 
GP(13)(14), Munblit D(15)(16), Douladiris N(17), O'Mahony L(18)(19)(20), 
Redegeld F(21), Riggioni C(22)(23), Smith PK(24), van Esch B(21), Vassilopoulou 
E(13)(14)(25)(26), Venter C(27), Peroni DG(7).

Author information:
(1)Centre for Animal Nutrition and Welfare, University of Veterinary Medicine, 
Vienna, Austria.
(2)Faculty of Medicine, Transylvania University, Brasov, Romania.
(3)Department of Allergy, Instituto de Investigación Biosanitaria Hospital 12 de 
Octubre (imas12), Madrid, Spain.
(4)Department of Translational Medical Science, University Federico II, Naples, 
Italy.
(5)Engineering and Advanced Biotechnology Center (CEINGE) Advanced 
Biotechnologies Research Center, University Federico II, Naples, Italy.
(6)Task Force for Microbiome Studies, University Federico II, Naples, Italy.
(7)Department of Clinical and Experimental Medicine, Section of Paediatrics, 
University of Pisa, Pisa, Italy.
(8)Translational Inflammation Research Division and Core Facility for Single 
Cell Multiomics, Philipps University Marburg, Member of the German Center for 
Lung Research (DZL), Universities of Giessen and Marburg Lung Center, Marburg, 
Germany.
(9)Scientific Advisory Unit, Spanish Council for Scientific Research, Madrid, 
Spain.
(10)Messerli Research Institute, Department of Interdisciplinary Life Sciences, 
University of Veterinary Medicine, Vienna, Austria.
(11)Human Development and Health, Faculty of Medicine, University of 
Southampton, Southampton, UK.
(12)NIHR Southampton Biomedical Research Centre, University Hospital Southampton 
NHS Foundation Trust, Southampton, UK.
(13)Department of Clinical Sciences and Community Health, Università Degli Studi 
di Milano, Milan, Italy.
(14)Pediatric Unit, Fondazione IRCCS Ca' Granda-Ospedale Maggiore Policlinico, 
Milan, Italy.
(15)Care for Long Term Conditions Division, Florence Nightingale Faculty of 
Nursing, Midwifery and Palliative Care, King's College London, London, UK.
(16)Department of Paediatrics and Paediatric Infectious Diseases, Institute of 
Child's Health, I.M. Sechenov First Moscow State Medical University, Sechenov 
University, Moscow, Russia.
(17)Allergy Department, 2nd Pediatric Clinic, University of Athens, Athens, 
Greece.
(18)APC Microbiome Ireland, University College Cork, Cork, Ireland.
(19)School of Microbiology, University College Cork, Cork, Ireland.
(20)Department of Medicine, University College Cork, Cork, Ireland.
(21)Division of Pharmacology, Utrecht Institute of Pharmaceutical Sciences, 
Faculty of Science, Utrecht University, Utrecht, the Netherlands.
(22)Division of Immunology and Allergy, The Hospital for Sick Children and the 
SickKids Food Allergy and Anaphylaxis Program, Toronto, Ontario, Canada.
(23)Department of Paediatrics, Temerty Faculty of Medicine, University of 
Toronto, Toronto, Ontario, Canada.
(24)School of Medicine and Dentistry, Griffith University, Parklands Drive, 
Queensland, Australia.
(25)Department of Nutritional Sciences and Dietetics, School of Health Sciences, 
International Hellenic University, Thessaloniki, Greece.
(26)Department of Life Sciences, School of Life and Health Sciences, Universiy 
of Nicosia, Nicosia, Cyprus.
(27)Department of Pediatrics, Section of Allergy and Immunology, University of 
Colorado School of Medicine/Children's Hospital Colorado, Aurora, Colorado, USA.

Iron is essential for oxygen transport, energy metabolism, and immune 
regulation. Yet iron deficiency is the most common micronutrient disorder across 
all age groups, affecting nearly one quarter of the global population. Iron 
deficiency triggers nutritional immunity, a host defense mechanism that 
withholds and redistributes iron, contributing to increased morbidity and 
mortality. This review outlines normal iron physiology, distribution and 
absorption pathways and on the consequences of deficiency across body 
compartments, with particular attention to type 2-driven diseases. Beyond 
anemia, insufficient iron availability disrupts immune homeostasis by promoting 
type 2 inflammation, elevating IgE, and activating mast cells and eosinophils. 
Regulatory macrophages, the central hub of iron cycling, adopt an inflammatory, 
iron-sequestering state that reinforces malabsorption and redistribution. 
Epidemiology studies show higher iron-deficiency risk in allergic individuals; 
low maternal iron or early-life iron predisposes to eczema, wheeze, and asthma, 
while food-allergen elimination (notably cow's milk) further worsens anemia 
risk. Clinical evidence indicates that restoring iron status through diet, 
supplementation, or fortification lowers IgE levels, improves lung function, and 
alleviates symptoms of rhinitis, urticaria, and asthma. Iron may therefore 
represent a modifiable determinant of allergic disease development and severity. 
Integrating iron assessment and nutritional care into allergy management may 
reduce disease burden and slow the progression of allergic march.

© 2026 The Author(s). Allergy published by European Academy of Allergy and 
Clinical Immunology and John Wiley & Sons Ltd.

DOI: 10.1111/all.70325
PMID: 41943501


18. JPEN J Parenter Enteral Nutr. 2026 Apr 6. doi: 10.1002/jpen.70089. Online ahead 
of print.

Impact of fortifiers on donor milk nutrient composition: An experimental study.

Mansen K(1), Sanders H(1), Taylor SN(2), Dunstan R(1), Israel-Ballard K(1), 
Perrin MT(3).

Author information:
(1)Integrated Maternal and Child Health and Development, PATH, Seattle, 
Washington, USA.
(2)Department of Pediatrics, Yale School of Medicine, New Haven, Connecticut, 
USA.
(3)Department of Nutrition, University of North Carolina Greensboro, Greensboro, 
North Carolina, USA.

BACKGROUND: Preterm infants frequently require human milk fortification to meet 
their unique nutrients requirements. How commercial fortifiers change essential 
macronutrients and micronutrients in donor human milk has not been well-studied.
METHODS: Our sampling frame included milk from approved United States milk bank 
donors (n = 400), measured for macronutrients, vitamins, and minerals, and used 
to produce 1000 pools of donor milk. Fortification ratios were computed to 
describe the extent that fortifiers changed donor milk composition (<100% 
indicates decreased; >100% indicates increased). Intake ratios were computed to 
evaluate how unfortified and fortified donor milk at 160 mL/kg/day compared to 
upper and lower preterm nutrient recommendations.
RESULTS: Fortifiers varied greatly in their impact on individual donor milk 
nutrients, with final macronutrient concentrations between 75% and 399% of 
unfortified values, while vitamin and mineral concentrations were between 50% 
and 800+% of unfortified values. Unfortified donor milk was able to meet the 
lower intake recommendations for three nutrients-lactose, fat, and pantothenic 
acid-highlighting the importance of utilizing multi-nutrient fortifiers. 
Bovine-derived fortifiers frequently did not meet lower targets for protein, 
choline, and several minerals, while human-derived fortifiers did not meet lower 
targets for most B-vitamins and choline, and exceeded some targets including 
protein, fat, calcium, phosphorus, and magnesium.
CONCLUSION: Fortification products available in the United States, when used 
with donor milk, frequently lead to both deficient and excessive nutrient intake 
compared to preterm enteral nutrition recommendations. Development of fortifier 
products based on robust nutritional reference values for donor milk is 
warranted to improve the nutritional care of preterm infants.

© 2026 The Author(s). Journal of Parenteral and Enteral Nutrition published by 
Wiley Periodicals LLC on behalf of American Society for Parenteral and Enteral 
Nutrition.

DOI: 10.1002/jpen.70089
PMID: 41942687


19. Front Plant Sci. 2026 Mar 18;17:1783391. doi: 10.3389/fpls.2026.1783391. 
eCollection 2026.

Effects of iodinated copper-based foliar fertilizers on iodine content, yield 
and quality of lettuce (Lactuca sativa L.).

Liu A(1)(2), Sun G(3), Ma G(4), Zhang M(1), Liu Z(1), Tang L(1)(2).

Author information:
(1)Shandong Agricultural University, Taian, China.
(2)Institute of Agricultural Resources and Regional Planning, Chinese Academy of 
Agricultural Sciences, Beijing, China.
(3)Taishan Polytechnic, Taian, China.
(4)Pingdu Branch of Qingdao Ecological Environment Bureau, Qingdao, 
Shandong, China.

Milk, dairy products, and certain marine foods are important dietary sources of 
iodine for humans. However, iodine intake is strongly influenced by dietary 
habits, local food availability, and iodine fortification policies, and ensuring 
adequate iodine intake remains a challenge in populations with low consumption 
of dairy products or seafood or in regions where salt iodization is limited. 
Under these circumstances, iodine-biofortified vegetables may serve as an 
important dietary source of iodine, and fertilization is an effective strategy 
to enhance their iodine content. In this study, using lettuce as a test crop, we 
conducted experiments over two growing seasons to evaluate an improved iodinated 
copper-based foliar fertilizer derived from traditional Bordeaux mixture (BDM), 
with treatments differing in iodine form and concentration. The results showed 
that the iodinated copper-based foliar fertilizers reduced copper concentrations 
in shoots and roots by 57.6%-91.57% and 61.56%-91.24%, respectively, compared 
with BDM, thereby reducing copper accumulation in plants and the potential risk 
of soil contamination. Compared with deionized water (CK), iodine application 
with copper-based fertilizers significantly increased iodine levels in lettuce 
tissues. Application of 1 g/L and 2 g/L KI copper-based foliar fertilizer 
increased shoot iodine content by 99.91% and 191.95%, respectively, and root 
iodine content by 180.66% and 211.41%, respectively. In another growing season, 
shoot iodine content increased by 24.71- to 66.20-fold, while root iodine 
content increased by 1.52- to 2.50-fold. In terms of quality, 1 g/L KI 
copper-based foliar fertilization increased vitamin C content by 33.19%, whereas 
2 g/L reduced nitrate content by 39.08%; soluble sugar content increased by 
34.41% and 39.27%. However, high iodine concentrations exerted a suppressive 
effect on yield, with high-concentration iodine treatments consistently reducing 
yield by approximately 13.85%-35.84% relative to CK across the two growing 
seasons. Overall, iodinated copper-based foliar fertilization effectively 
enhanced iodine enrichment in lettuce while simultaneously reducing copper 
accumulation, suggesting its potential as a safer alternative to traditional BDM 
for iodine biofortification. Among the tested treatments, 2 g/L KI copper-based 
foliar fertilization produced the greatest improvements in iodine enrichment and 
quality traits, although this was accompanied by a reduction in yield, 
indicating that the trade-off between nutritional enhancement and yield penalty 
should be carefully considered when determining the optimal application rate.

Copyright © 2026 Liu, Sun, Ma, Zhang, Liu and Tang.

DOI: 10.3389/fpls.2026.1783391
PMCID: PMC13041569
PMID: 41929839

Conflict of interest statement: The author(s) declared that this work was 
conducted in the absence of any commercial or financial relationships that could 
be construed as a potential conflict of interest. The author ZL declared that 
they were an editorial board member of Frontiers, at the time of submission. 
This had no impact on the peer review process and the final decision.


20. Indian Pediatr. 2026 Apr 1. doi: 10.1007/s13312-026-00320-9. Online ahead of 
print.

Outbreak of Gastrointestinal Sepsis Associated with Bacillus cereus-Contaminated 
Human Milk Fortifier in Preterm Neonates.

Laha W(1), Paswal CM(1), Satpathy SK(1), Gupta N(2), Tak V(3).

Author information:
(1)Department of Neonatology, All India Institute of Medical Sciences, Jodhpur, 
Rajasthan, 342005, India.
(2)Department of Neonatology, All India Institute of Medical Sciences, Jodhpur, 
Rajasthan, 342005, India..
(3)Department of Microbiology, All India Institute of Medical Sciences, Jodhpur, 
Rajasthan, India.

We report a cluster of acute gastrointestinal illness in seven preterm neonates 
temporally associated with a single batch of powdered human milk fortifier 
(HMF). Symptoms developed within 3-4 h of ingestion and included profuse 
diarrhea, dehydration, hypoglycemia, and shock. Blood and stool cultures were 
sterile. Microbiological analysis of unopened HMF sachets revealed contamination 
with Bacillus cereus and other spore-forming organisms. Withdrawal of the 
implicated batch and reinforcement of infection-control measures resulted in no 
further cases. This outbreak highlights the risk of pathogenic contamination of 
powdered HMF in neonatal intensive care units.

© 2026. The Author(s), under exclusive licence to Indian Academy of Pediatrics.

DOI: 10.1007/s13312-026-00320-9
PMID: 41920518

Conflict of interest statement: Conflict of interest: None. Ethics approval: The 
study was approved by the Institutional Ethics Committee; Ref No: 
AIIMS/JDH/NEO/2025/07/359 dated July 31, 2025. Consent to publish: Consent for 
publication of anonymized data and results was obtained from the parents or 
legal guardians of the participants. Consent to Participate: Informed consent 
was obtained from the parents or legal guardians of all participating neonates 
prior to inclusion in the study.


21. Int J Biol Macromol. 2026 Apr;358:151694. doi: 10.1016/j.ijbiomac.2026.151694. 
Epub 2026 Mar 27.

Structural interactions and technological and sensory effects of Mauritia 
flexuosa (buriti) oil microparticles microencapsulated with the polysaccharides 
gum arabic and inulin in the yogurt protein matrix.

de Oliveira JP(1), Silva FN(2), Soares JF(3), da Silva MAN(4), Felix PHC(3), de 
Oliveira Ferreira Rocha L(2), Ramos CL(2), Sosa FHB(5), da Costa JMG(2).

Author information:
(1)Graduate Program in Food Engineering and Science, State University of Bahia, 
Itapetinga, Bahia, 45700-000, Brazil. Electronic address: 
.
(2)Department of Food Engineering, Federal University of the Jequitinhonha and 
Mucuri Valleys, Diamantina, Minas Gerais, 39100-000, Brazil.
(3)Department Chemistry, Federal University of Minas Gerais, Belo Horizonte, 
Minas Gerais, 31270-901, Brazil.
(4)Faculty of Agrarian Science, Federal University of Amazonas, Manaus, 
Amazonas, 69080-900, Brazil.
(5)CICECO - Aveiro Institute of Materials, Department of Chemistry, University 
of Aveiro, 3810-193, Aveiro, Portugal.

Buriti oil, a rich source of β-carotene and antioxidants, presents limitations 
for direct incorporation into yogurt due to oxidative instability and 
hydrophobicity. In this study, buriti oil was microencapsulated into lyophilized 
microparticles using gum arabic and inulin and incorporated into yogurt 
formulations at gum arabic:inulin ratios of 3:1, 1:1, and 1:3 (w/w; coded as 
AG75IN25, AG50IN50, and AG25IN75, respectively). A control yogurt without 
microparticles was also prepared. All enriched formulations provided 1437.13 μg 
of β-carotene per 200 g of yogurt (40% of the recommended daily intake of 
vitamin A), with variations only in the wall material ratio. Structural, 
physicochemical, microbiological, and sensory analyses were conducted during 
refrigerated storage (4 °C). Scanning electron microscopy revealed 
microstructural changes after microparticle incorporation, while FTIR 
spectroscopy suggested modifications in molecular organization and 
protein-polysaccharide associations. AG50IN50 showed the most balanced 
performance, retaining 97% of β-carotene after 15 days and increasing firmness 
and elasticity by ∼20% compared to the control. This formulation-maintained 
pH 4.1-4.3, titratable acidity 1.11-1.28 g·100 g-1 lactic acid, syneresis 
22.9-35.6%, and microbial viability above 8.4 log₁₀ CFU·g-1. AG25IN75 retained 
92% of β-carotene but had lower sensory acceptance, while AG75IN25 retained 89% 
with intermediate scores. Lyophilized microparticles improved β-carotene 
retention without compromising yogurt quality. Freeze-drying microencapsulation 
effectively preserved β-carotene and ensured homogeneous oil dispersion, 
mitigating sensory impacts, guiding future research on lipophilic compound 
delivery and supporting the dairy industry in producing stable, nutritious, and 
sensorially acceptable fortified yogurts.

Copyright © 2026 The Authors. Published by Elsevier B.V. All rights reserved.

DOI: 10.1016/j.ijbiomac.2026.151694
PMID: 41903638 [Indexed for MEDLINE]

Conflict of interest statement: Declaration of competing interest The authors 
declare that they have no known competing financial interests or personal 
relationships that could have appeared to influence the work reported in this 
paper.


22. AJP Rep. 2026 Mar 24;16(1):e63-e66. doi: 10.1055/a-2832-1849. eCollection 2026 
Jan.

Freeze-Dried Mother's Own Milk for Novel Fortification in a Late Preterm Infant 
with Complicated Intestinal Atresia and Congenital Shortened Bowel: A Case 
Report.

Ward A(1)(2), Reyes SM(2)(3), Luck B(2), Serke L(2), Figard L(2), Kimball 
A(4)(5).

Author information:
(1)UChicago Medicine AdventHealth, Division of Women and Children's, Hinsdale, 
Illinois, United States.
(2)Milkify, Inc, Houston, Texas, United States.
(3)Rev Bioscience, LLC, Boise, Idaho, United States.
(4)Division of Neonatology, Rady Children's Hospital, San Diego, United States.
(5)Department of Pediatrics, University of California San Diego School of 
Medicine, San Diego, California, United States.

OBJECTIVE: Feeding intolerance and growth failure commonly complicate recovery 
in infants with complicated intestinal atresia, often requiring prolonged human 
milk fortification after hospital discharge. Our objective was to describe a 
novel fortification strategy that enabled an exclusive mother's own milk (MOM) 
diet during postdischarge fortification in a medically complex infant with 
feeding intolerance.
STUDY DESIGN: This case report details the use of freeze-dried mother's own milk 
(FDMOM) to fortify expressed MOM in a late preterm infant with complicated 
atresia and congenital shortened bowel to resolve feeding intolerance and weight 
faltering. Freeze-drying of MOM took place at a commercial facility using 
SafeDry, a patented contact-free process. FDMOM was used to increase the caloric 
density of expressed MOM under medical supervision using a targeted 
fortification approach.
RESULTS: The patient tolerated unfortified MOM but developed severe fussiness, 
abdominal distention, and increased stooling upon fortification with 
hypoallergenic formulas. These symptoms resolved within 24 hours of 
transitioning to FDMOM fortification. Remarkably, the infant went from the 24th 
percentile for weight-for-age to the 66th percentile within 86 days.
CONCLUSION: FDMOM fortification may represent a novel, well-tolerated strategy 
to support growth while maintaining an exclusive MOM diet in infants after 
complex gastrointestinal surgery and hospital discharge.

The Author(s). This is an open access article published by Thieme under the 
terms of the Creative Commons Attribution License, permitting unrestricted use, 
distribution, and reproduction so long as the original work is properly cited. ( 
https://creativecommons.org/licenses/by/4.0/ ).

DOI: 10.1055/a-2832-1849
PMCID: PMC13012845
PMID: 41884655

Conflict of interest statement: Conflict of Interest A.W., S.M.R., B.L., L.S., 
and L.F. are employed by or are consultants for Milkify. A.K. has no conflict of 
interest to declare.


23. Rev Prat. 2026 Mar;76(3):269-275.

[Early management of preterm infants in neonatal intensive care units (excluding 
respiratory diseases and ulcerative necrotizing enterocolitis)].

[Article in French; Abstract available in French from the publisher]

Frérot A(1), Biran V(1).

Author information:
(1)Service de néonatologie et réanimation néonatale, hôpital Robert-Debré, 
AP-HP, Université Paris Cité, Paris France.

Preterm neonates present multiple challenges due to organ 
immaturity.Cardiovascular:Patent ductus arteriosus (PDA) is common in lower 
gestational ages and may cause respiratory and circulatory complications. 
Treatment includes medical (ibuprofen, para-cetamol), surgical, or percutaneous 
approaches. Bradycardia is frequent and requires continuous 
monitoring.Digestive/Metabolic:Early enteral nutrition with fortified breast 
milk is preferred; parenteral nutrition is often required. Gastroesophageal 
reflux is usually benign. Preventing metabolic bone disease necessitates 
adequate calcium and phosphate intake. Jaundice is common; cholestasis often 
relates to parenteral nutrition.Infectious:Immune immaturity and invasive 
devices increase risks of early-onset (E.coli), nosocomial (CoNS), and fungal 
(Candida) infections. Diagnosis relies on blood cultures; treatment is empirical 
then targeted.Neurological:Common lesions include periventricular leukomalacia 
and intraventricular hemorrhage, with potential sequelae. Monitoring includes 
cranial ultrasound, EEG, MRI, and routine screening for hearing loss and 
retinopathy of prematurity.

Publisher: Le nouveau-né prématuré présente de nombreuses particularités liées à 
son immaturité.Sur le plan cardiovasculaire, la persistance du canal artériel 
est d’autant plus fréquente que l’âge gestationnel est bas, avec des 
conséquences respiratoires et circulatoires. Le traitement peut être médical 
(ibuprofène, paracétamol), percutané ou chirurgical. Les bradycardies sont 
également fréquentes et nécessitent une surveillance continue.Sur le plan 
digestif et métabolique une nutrition entérale précoce avec lait maternel 
enrichi est recommandée ; la nutrition parentérale est souvent nécessaire. Le 
reflux gastro-œsophagien est fréquent mais le plus souvent bénin, traité par des 
mesures hygiénodiététiques. La prévention de la maladie osseuse métabolique 
nécessite un apport phosphocalcique adapté. L’ictère est fréquent, la cholestase 
souvent liée à la nutrition parentérale.Sur le plan infectieux, l’immaturité 
immunitaire et l’utilisation de dispositifs invasifs exposent à des infections 
bactériennes précoces (E. coli), nosocomiales (SCN) et fongiques (Candida). Le 
diagnostic est difficile et repose sur les hémocultures. Le traitement est 
empirique, puis adapté aux résultats.Sur le plan neurologique, les lésions 
cérébrales les plus fréquentes sont la leucomalacie et les hémorragies 
intraventriculaires, à risque de séquelles. Le suivi repose sur l’échographie, 
l’électroencéphalo­gramme et l’IRM. Le dépistage de la surdité et de la 
rétinopathie du prématuré est essentiel.

PMID: 41878969 [Indexed for MEDLINE]

Conflict of interest statement: Les auteures déclarent n’avoir aucun lien 
d’intérêts.


24. Food Funct. 2026 Apr 14;17(7):3091-3098. doi: 10.1039/d5fo04757a.

Effects of infant formula supplied with medium- and long-chain triacylglycerols 
and sn-2 palmitate on fecal lipid metabolome and microbiota profiles in healthy 
term infants: a parallel-controlled study.

Yu J(1)(2), Liu Z(3), Yang M(4), Bi Q(1)(2), Zhou X(3), Qiu Y(4), Ye X(3), Wei 
W(1)(2), Wang X(1)(2).

Author information:
(1)State Key Lab of Food Science and Resources, Jiangnan University, Wuxi 
214122, China..
(2)Collaborative Innovation Center of Food Safety and Quality Control in Jiangsu 
Province, School of Food Science and Technology, Jiangnan University, Wuxi 
214122, China.
(3)Inner Mongolia Mengniu Dairy (Group) Limited by Share Ltd, Hohhot, Inner 
Mongolia, 011500, China..
(4)Nutrition and Health, School of Public Health, Sun Yat-sen University, 
Guangzhou 510080, China.

Infant formula fortified with medium- and long-chain triacylglycerols and sn-2 
palmitate showed a comparable fecal lipidomics pattern to breast milk. 
Significant correlations were observed between fecal lipid and beneficial 
microbiota such as Bifidobacterium. The results indicated that structured lipids 
influenced lipid metabolism and infant gut health.

DOI: 10.1039/d5fo04757a
PMID: 41874534 [Indexed for MEDLINE]


25. Food Chem. 2026 May 30;512:148890. doi: 10.1016/j.foodchem.2026.148890. Epub 
2026 Mar 16.

Enhancing carotenoid bioaccessibility in food matrices using modified mango 
cotyledon starch microcapsules: Contribution to the recommended daily intake.

Torres-Gallo R(1), Aranda-Bustos M(2), Oyarzún P(2), F Tirado D(3), 
Chávez-Salazar A(4), J Castellanos-Galeano F(5).

Author information:
(1)Department of Agroindustrial Engineering, Faculty of Engineering, Universidad 
del Atlántico, 30th Avenue No. 8-49, Puerto Colombia, 081001, Atlántico, 
Colombia; Doctorate in Engineering, Universidad de Caldas, Calle 65 No. 26-10, 
Manizales 170001, Caldas, Colombia. Electronic address: 
.
(2)Food and Drug Research Laboratory, Faculty of Chemistry and Farmacy, 
Pontificia Universidad Católica de Chile, Chile.
(3)Biological Engineering Program, Dirección Académica, Universidad Nacional de 
Colombia, Sede de La Paz, La Paz 202017, Colombia.
(4)Research Center Ci2DT2, Department of Food Engineering, Faculty of Artificial 
Intelligence and Engineering, Universidad de Caldas, Calle 65 No. 26-10, 
Manizales 170001, Caldas, Colombia.
(5)Technological Development Center for Bioprocesses and Agroindustry Plant, 
Department of Food Engineering, Faculty of Artificial Intelligence and 
Engineering, Universidad de Caldas, Calle 65 No. 26-10, Manizales 170001, 
Caldas, Colombia.

This study evaluated starch extracted from mango cotyledons as a wall material 
for microencapsulating a carotenoid-rich pumpkin suspension. The resulting 
microcapsules were incorporated into water, yogurt, and gelatin to develop 
carotenoid-fortified foods. The effects of wall material and matrix on 
carotenoid bioaccessibility during in vitro digestion and their contribution to 
the recommended dietary allowance (RDA) across age groups were assessed. 
Carotenoid content and bioaccessibility varied significantly with encapsulant 
type and matrix. The combination of ultrasound-modified and dual-modified starch 
improved carotenoid bioaccessibility up to 27% (20 mg/100 g) in water, 16% 
(27 mg/100 g) in gelatin and 8% (49 mg/100 g) in yogurt; providing enhanced 
encapsulation and stability. Yogurt yielded the highest bioaccessibility, likely 
due to lipid-protein interactions enhancing carotenoid stability under 
gastrointestinal conditions. A 200 mL serving of fortified yogurt could provide 
26%-36% of adult and 39%-62% of child (ages 4-13) vitamin A RDA. Utilizing mango 
processing residues as wall materials offers a sustainable, value-added strategy 
for developing functional foods.

Copyright © 2024. Published by Elsevier Ltd.

DOI: 10.1016/j.foodchem.2026.148890
PMID: 41855840 [Indexed for MEDLINE]

Conflict of interest statement: Declaration of competing interest The authors 
declare the following financial interests/personal relationships which may be 
considered as potential competing interests: Ramiro Torres Gallo reports 
financial support was provided by Science, Technology, and Innovation Fund 
(FCTI), of the Sistema General de Ragalías (SGR). Ramiro Torres Gallo reports 
financial support was provided by University of Caldas - Colombia.BPIN framework 
project 2,019,000,100,035. Ramiro Torres Gallo reports equipment, drugs, or 
supplies was provided by National Fund for Scientific & Technological 
Development (FONDECYT). Ramiro Torres Gallo reports was provided by Fondequip 
Fondo de Equipamiento Científico y Tecnológico. Ramiro Torres Gallo reports 
equipment, drugs, or supplies was provided by International Cooperation. If 
there are other authors, they declare that they have no known competing 
financial interests or personal relationships that could have appeared to 
influence the work reported in this paper.


26. J Food Sci. 2026 Mar;91(3):e70978. doi: 10.1111/1750-3841.70978.

Developing Indigenous Plant-Enriched Yogurt to Enhance Iron Solubility-A 
Collaboration With Senegalese Women Farmers.

Ifeh C(1), Allan E(1), Ndiaye A(2), Miles MP(1), Kuo WY(1).

Author information:
(1)Department of Food Systems, Nutrition, and Kinesiology, Montana State 
University, Bozeman, Montana, USA.
(2)National Union of Chambers of Commerce, Dakar, Senegal.

This study utilized collaborations with Senegalese women farmers to investigate 
the impact of lactic acid bacteria (LAB) fermentation of sorghum, baobab, and 
milk on nutritional and sensory qualities for plant-enriched yogurt. Using a 
simplex lattice mixture design, five samples of varying weight ratios of 
sorghum/baobab (17.5/32.5-47.5/2.5, w.b.) were prepared by mixing sorghum/baobab 
blend (50 g, w.b.), milk (500 mL), Lactobacillus bulgaricus species and 
Streptococcus thermophilus species, and fermenting at 40°C for 9 h. After only 
fermenting milk, one Control was prepared by adding sorghum/baobab (32.5/17.5). 
The samples were evaluated for pH, texture, LAB count, and iron solubility after 
simulated digestion. A total of 125 panelists, including 32 Africans, evaluated 
the samples flavored with bananas after fermentation, using a nine-point hedonic 
scale and check-all-that-apply (CATA) method. Data were analyzed using variance 
analysis, least significant difference test, agglomerative hierarchical 
clustering, and Fisher's exact test. All samples had a pH below 4.6 and a LAB 
count of 108, except for sample 17.5/32.5 (104), likely due to baobab's acidity 
hindering LAB growth. Fermenting sorghum, baobab, and milk together increased 
iron solubility by 64.1% compared to the Control. From the CATA results, sweet 
flavor and creamy mouthfeel increased overall liking, while tart flavor and 
gritty mouthfeel decreased liking. Cluster analysis showed that Africans favored 
samples with both less and more baobab equally (40/10, 25/25, overall liking of 
6.8), while Caucasians preferred less baobab (40/10, overall liking of 4.8). 
Both clusters disfavored the Control (overall liking 6.0 and 4.1), which had the 
lowest instrumental texture consistency.

© 2026 The Author(s). Journal of Food Science published by Wiley Periodicals LLC 
on behalf of Institute of Food Technologists.

DOI: 10.1111/1750-3841.70978
PMCID: PMC12988478
PMID: 41830445 [Indexed for MEDLINE]

Conflict of interest statement: The authors declare no conflicts of interest.


27. Pediatr Res. 2026 Mar 13. doi: 10.1038/s41390-026-04893-w. Online ahead of 
print.

Beyond numbers: rethinking human milk fortification in preterm nutrition.

Morniroli D(1), Agostoni C(2)(3), Giannì ML(1)(4).

Author information:
(1)Department of Clinical Sciences and Community Health, University of Milan, 
Milan, Italy.
(2)Department of Clinical Sciences and Community Health, University of Milan, 
Milan, Italy..
(3)Pediatric Unit, Fondazione IRCCS Ca' Granda Ospedale Maggiore Policlinico, 
Milan, Italy..
(4)Neonatal Intensive Care Unit, Fondazione IRCCS Ca' Granda Ospedale Maggiore 
Policlinico, Milan, Italy.

As neonatal care continues to evolve, there is growing recognition of the need 
for individualized, functionally targeted nutrition in very preterm infants 
(VPIs). While human milk remains the gold standard, it frequently fails to meet 
the metabolic requirements of this high-risk group, requiring the adoption of 
fortification strategies. In this context, the study by Rasmussen et al., 
published in Pediatric Research, offers important insights into the metabolic 
implications of fortifying human milk with bovine colostrum (BC). The observed 
elevations in plasma concentrations of amino acids such as arginine, glutamine, 
and tyrosine suggest that BC may serve as a fortifier with potential clinical 
value. However, the disconnection between enhanced biochemical profiles and 
clear clinical benefit persists. Nutrient utilization depends not only on intake 
but also on inflammation, illness, organ immaturity, and individual metabolic 
variations. Infants' anthropometrics, mainly head circumference and length, 
remain essential reference values, mirroring brain and lean mass development. A 
shift toward precision strategies incorporating biomarkers and long-term 
developmental follow-up is needed. This approach might hopefully be translated 
into sustained clinical benefit for VPIs. IMPACT: This commentary emphasizes the 
importance of evaluating neonatal nutrition through both surrogate biomarkers 
and functional clinical outcomes to better capture physiological relevance. It 
highlights the limitations of relying exclusively on compositional or 
biochemical analyses when assessing nutritional adequacy in preterm infants. The 
article contextualizes the findings of Rasmussen et al. within the metabolic 
complexity characteristic of preterm physiology. It underscores the role of 
clinical heterogeneity and variable nutrient utilization in shaping metabolic 
responses. The commentary advocates for integrated, technology-supported 
strategies to enable individualized, real-time precision nutrition in neonatal 
care.

© 2026. The Author(s), under exclusive licence to the International Pediatric 
Research Foundation, Inc.

DOI: 10.1038/s41390-026-04893-w
PMID: 41826705

Conflict of interest statement: Competing interests: The authors declare no 
competing interests.


28. iScience. 2026 Feb 9;29(3):114968. doi: 10.1016/j.isci.2026.114968. eCollection 
2026 Mar 20.

Early-life microbiota disruption-induced deficits in the social brain are 
sensitive to diet.

Ratsika A(1)(2), Bergmann CA(1), Valderrama B(1)(2), Bastiaanssen TFS(1), 
Sharvin BL(1)(2), Renes IB(3), Knol J(4)(3), Gunnigle E(1), Clarke G(1)(5), 
Cryan JF(1)(2).

Author information:
(1)APC Microbiome Ireland, Biosciences Institute, University College Cork, Cork 
T12 YT20, Ireland.
(2)Department of Anatomy and Neuroscience, University College Cork, Cork T12 
YT20, Ireland.
(3)Danone Global Research & Innovation Centre, Utrecht, CT 3584, the 
Netherlands.
(4)APC Microbiome Ireland, School of Microbiology, University College Cork, Cork 
T12 YT20, Ireland.
(5)Department of Psychiatry and Neurobehavioral Science, University College 
Cork, Cork T12 YT20, Ireland.

Diet is one of the major modulators of the microbiota-gut-brain axis across the 
lifespan. Milk bioactive components, including human milk oligosaccharides such 
as fucosyllactose and sialyllactose, and prebiotics, including GOS and FOS, 
promote the viability of commensal bacteria, fortify the intestinal barrier, and 
improve cognitive development. Here, we investigate the ability of these dietary 
components alone or in combination to counter the behavioral and physiological 
effects of early-life microbiota depletion via broad-spectrum antibiotics in 
mice. Microbiota depletion impaired social recognition in juvenile mice, which 
was reversed by supplementation with human milk oligosaccharides, GOS/FOS, and 
their combination. Transcriptomic analysis in brain areas linked to social 
memory (amygdala and prefrontal cortex), revealed that pathways for central 
nervous system development, learning, learning and memory are sensitive only to 
the combined supplementation. Together, our data show that prebiotics and milk 
bioactive components exert beneficial effects on the host by reversing 
microbiota depletion-related deficits on the brain and behavior.

© 2026 The Author(s).

DOI: 10.1016/j.isci.2026.114968
PMCID: PMC12969037
PMID: 41809042

Conflict of interest statement: The work in the article “Early-life Microbiota 
Disruption-Induced Deficits in the Social Brain are Sensitive to Diet” was 
funded by a research grant from Danone to J.F.C. J.F.C. has received research 
funding from IFF, Reckitt, and Danone Nutricia. J.K. and I.B.R. are employed by 
Danone Global Research & Innovation Center. G.C. has received honoraria from 
Janssen, Probi, Apsen, and Ingelheim Boehringer as an invited speaker; is in 
receipt of research funding from Pharmavite, Fonterra, Reckitt, Nestle, and Tate 
and Lyle; and has been paid for consultancy work by Yakult, Zentiva, Bayer 
Healthcare, and Heel Pharmaceuticals. This support neither influenced nor 
constrained the contents of this article. A.R., C.A.B., B.V., T.F.B., B.L.S., 
and E.G. declare no conflict of interest.


29. Semin Fetal Neonatal Med. 2026 Feb 27:101710. doi: 10.1016/j.siny.2026.101710. 
Online ahead of print.

Fortification of human milk for feeding preterm infants: What is the ideal 
strategy?

Moya F(1).

Author information:
(1)Pediatrix Medical Group of Austin, Clinical Professor of Pediatrics, Texas A 
& M College of Medicine, 900 E. 30th Street, Suite 211, Austin, TX, 78705, USA. 
Electronic address:.

Promoting optimal growth in preterm infants is of paramount importance. Multiple 
reports have associated better growth during their Neonatal Intensive Care Unit 
(NICU) stay with improved neurodevelopmental outcome and a lesser risk of 
certain morbidities. Nowadays, most preterm infants receive human milk (HM) 
feedings, either mother's own or donor milk. Widespread use of HM to feed 
preterm infants has contributed to a decrease in complications like necrotizing 
enterocolitis. This notwithstanding, HM often does not provide sufficient 
protein and other nutrients to promote adequate growth in these infants and, 
therefore, must be fortified with specially designed human milk fortifiers of 
human or bovine origin. These provide added amounts of protein, minerals like 
sodium, calcium, and phosphorus as well as vitamins and other micronutrients. 
Which fortification strategy, i.e. type of fortifier, timing of fortification, 
must be used to promote optimal growth and neurodevelopment is still a matter of 
debate. The following articles by the well-recognized experts in neonatal 
nutrition, Drs. Amy Hair and Brenda Poindexter, will describe alternate 
strategies to fortify HM and their potential benefits, and are a reflection of 
the scholarly discussions held during the 2025 Neonatal Insights: Best Evidence 
and Practice in Clinical Neonatology Conference.

Copyright © 2026. Published by Elsevier Ltd.

DOI: 10.1016/j.siny.2026.101710
PMID: 41796447

Conflict of interest statement: Declaration of competing interest Fernando Moya 
is a member of the speaker's bureau and has received grant funding for studies 
related to human milk fortification from Mead Johnson Nutrition.


30. Food Res Int. 2026 Apr 30;230:118596. doi: 10.1016/j.foodres.2026.118596. Epub 
2026 Feb 4.

Beneficial effects of Ribes nigrum L. polyphenols on yogurt: Sensory quality, 
probiotic proliferation, pathogen inhibition and bioactivity.

Yang Y(1), Liu H(1), Zhao M(1), Guo P(2), Bai J(3).

Author information:
(1)College of Art and Science, Northeast Agricultural University, Harbin 150030, 
PR China.
(2)College of Art and Science, Northeast Agricultural University, Harbin 150030, 
PR China. Electronic address:.
(3)College of Art and Science, Northeast Agricultural University, Harbin 150030, 
PR China. Electronic address:.

Yogurt is a globally popular fermented dairy product with excellent nutritional 
value. Its well-documented health benefits are predominantly associated with the 
probiotic strains involved in the fermentation process, which play key roles in 
gut microbiota modulation and metabolic homeostasis regulation. Concurrently, 
polyphenols derived from Ribes nigrum L. (RMAP) are likewise recognized for 
their bioactive potential, particularly their antioxidant and antimicrobial 
activities. To meet growing consumer interest and demand for value-added yogurt, 
the polyphenols extracted from Ribes nigrum L. (RMAP) were processed into 
freeze-dried powder and incorporated into yogurt to develop a novel functional 
product. After 21 days of storage at 4 °C, the addition of RMAP (0.25% ∼ 4%, 
w/w) in yogurt significantly promoted the viability of probiotic lactobacilli 
(L. plantarum, L. casei, L. rhamnosus, and L. acidophilus), improved the sensory 
property, antioxidant and α-glucosidase inhibitory activities, and inhibited the 
biofilm formation of pathogenic bacteria (S. aureus, E. coli and Salmonella). 
Moreover, the incorporation of RMAP suppressed protein degradation and 
stabilized the structure of protein in yogurt. Especially, the yogurt with 2% 
RMAP displayed the superior sensory quality, physicochemical characteristics and 
stability. UPLC-ESI-MS analysis revealed that yogurt supplemented with RMAP 
contained elevated levels of antioxidant compounds, including protocatechuic 
acid, rutin, and hyperoside. Principal component analysis demonstrated that the 
microbial stability, flavor quality, antioxidant activity, and overall sensory 
of RMAP-fortified yogurt were closely associated with the storage time. 
Therefore, RMAP may be a promising supplement for the development of innovative 
functional dairy products with enhanced nutritional and bioactive profiles.

Copyright © 2026 Elsevier Ltd. All rights reserved.

DOI: 10.1016/j.foodres.2026.118596
PMID: 41794477 [Indexed for MEDLINE]

Conflict of interest statement: Declaration of competing interest The authors 
declare that they have no known competing financial interests or personal 
relationships that could have appeared to influence the work reported in this 
paper.


31. Food Res Int. 2026 Apr 1;229:118513. doi: 10.1016/j.foodres.2026.118513. Epub 
2026 Jan 27.

Study on rheological behavior of stirred yoghurt for dysphagia patients.

Dong Q(1), Xu J(2), Liu D(3).

Author information:
(1)College of Biosystems Engineering and Food Science, Zhejiang University, 
Hangzhou 310058, China; Innovation Center of Yangtze River Delta, Zhejiang 
University, Jiaxing 314102, China.
(2)College of Biosystems Engineering and Food Science, Zhejiang University, 
Hangzhou 310058, China; Innovation Center of Yangtze River Delta, Zhejiang 
University, Jiaxing 314102, China. Electronic address:.
(3)College of Biosystems Engineering and Food Science, Zhejiang University, 
Hangzhou 310058, China; Innovation Center of Yangtze River Delta, Zhejiang 
University, Jiaxing 314102, China. Electronic address:.

For dysphagia patients, diet management is a highly effective and well-accepted 
therapeutic method. Stirred yoghurt is particularly suitable due to its 
mouthfeel, flavor, and nutritional benefits, especially its increased viscosity 
compared to milk. However, despite extensive research on yoghurt rheology, a 
comprehensive understanding of its thixotropy and yield behaviors remains 
limited, hindering the rational design of texture-modified foods for dysphagia 
management. This study first investigated the viscosity and viscoelasticity of 
four yoghurts with different formulas (rich-textured "Indulgent" yoghurt, Y1; 
starch-containing yoghurt, Y2; light-texture fermented yoghurt, Y3; vitamin 
D-fortified yoghurt, Y4) through steady-state shear, frequency sweep and 
creep-recovery tests. Then, the hysteresis loop and thixotropic recovery tests 
were used to determine the thixotropy of yoghurts. Yield behavior was further 
evaluated and compared across six methods, involving creep, stress ramps, step 
shear rate, steady-state shear, amplitude sweep and stress bifurcation. 
According to the International Dysphagia Diet Standardisation Initiative 
framework, Y1, Y2 and Y4 were all classified as level 4. However, rheological 
analysis revealed that Y2 exhibited significantly higher thixotropy and yield 
stress, corresponding with its higher link density as determined by confocal 
laser scanning microscopy. And the yield of yoghurt was a process, with Y2 and 
Y4 exhibiting brittle yield, while Y1 and Y3 being ductile yield. This study 
provides novel insights into yoghurt microstructure-rheology relationships and 
offers scientifically robust guidance for tailoring diets to dysphagia patients 
with diverse conditions.

Copyright © 2026 Elsevier Ltd. All rights reserved.

DOI: 10.1016/j.foodres.2026.118513
PMID: 41763829 [Indexed for MEDLINE]

Conflict of interest statement: Declaration of competing interest The authors 
declare that they have no known competing financial interests or personal 
relationships that could have appeared to influence the work reported in this 
paper.


32. Food Res Int. 2026 Apr 1;229:118470. doi: 10.1016/j.foodres.2026.118470. Epub 
2026 Jan 21.

Colostrum-specific amino acids and lipids elevating lutein bioaccessibility at 
the micellization stage relative to mature milk and infant formula.

Wang Y(1), He X(2), Chen Y(1), Kan J(3), Du M(3), Takács K(4), Zalán Z(4), Cai 
T(5), Chen K(6).

Author information:
(1)College of Food Science, Southwest University, No. 2, Tiansheng Road, Beibei, 
Chongqing 400715, PR China; Chongqing Key Laboratory of Specialty Food Co-built 
by Sichuan and Chongqing, Chongqing 400715, PR China.
(2)Department of Obstetrics and Gynecology, The First Affiliated Hospital of 
Chongqing Medical University, Chongqing 400016, PR China.
(3)College of Food Science, Southwest University, No. 2, Tiansheng Road, Beibei, 
Chongqing 400715, PR China; Chinese-Hungarian Cooperative Research Centre for 
Food Science, Chongqing 400715, PR China; Chongqing Key Laboratory of Specialty 
Food Co-built by Sichuan and Chongqing, Chongqing 400715, PR China.
(4)Food Science and Technology Institute, Hungarian University of Agriculture 
and Life Sciences, Buda Campus, Villányi road 29-43, H-1118 Budapest, Hungary.
(5)School of Chemistry and Chemical Engineering, Southwest University, No. 2, 
Tiansheng Road, Beibei, Chongqing 400715, PR China; Chinese-Hungarian 
Cooperative Research Centre for Food Science, Chongqing 400715, PR China. 
Electronic address:.
(6)College of Food Science, Southwest University, No. 2, Tiansheng Road, Beibei, 
Chongqing 400715, PR China; Chinese-Hungarian Cooperative Research Centre for 
Food Science, Chongqing 400715, PR China; Chongqing Key Laboratory of Specialty 
Food Co-built by Sichuan and Chongqing, Chongqing 400715, PR China. Electronic 
address:.

Lutein's bioaccessibility varies between infant formula (IF) and milk and among 
lactation stages, and controlling its micellization in matrices and the benefits 
colostrum (CM) at equal lutein pre-digestion levels remain unaddressed. Hence, 
this study used an infant-relevant dynamic in vitro gastrointestinal model to 
equalize lutein levels in digestive fluids, compare digesta's and mixed 
micelles' physicochemical stability, and investigate release dynamics in CM, 
mature milk, and IF. Untargeted and targeted omics profiling of micellar 
fractions was then conducted. IF exhibited >20% lutein loss, which was twice 
that of human milk at matched lutein concentration. CM showed the highest 
micellization efficiency and lutein bioaccessibility, and five amino acids and 
six lipids were enriched in CM-derived micelles. These results suggest that 
functional molecules enhance lutein delivery and human-milk matrices facilitate 
lipophilic nutrient delivery, providing a foundation for next-generation IF 
fortification strategies.

Copyright © 2026 Elsevier Ltd. All rights reserved.

DOI: 10.1016/j.foodres.2026.118470
PMID: 41763792 [Indexed for MEDLINE]

Conflict of interest statement: Declaration of competing interest The authors 
declare that they have no known competing financial interests or personal 
relationships that could have appeared to influence the work reported in this 
paper.


33. Food Chem. 2026 Apr 30;509:148337. doi: 10.1016/j.foodchem.2026.148337. Epub 
2026 Feb 19.

Estimating daily intakes of toxic metals and essential elements from infant 
formula, gruel, porridge and ready-to-eat baby food.

Gustin K(1), Trask M(1), Bjermo H(2), Levi M(1), Lignell S(2), Vahter M(1), 
Kippler M(3).

Author information:
(1)Institute of Environmental Medicine, Karolinska Institutet, Stockholm, 
Sweden.
(2)Department of Risk and Benefit Assessment, Swedish Food Agency, Uppsala, 
Sweden.
(3)Institute of Environmental Medicine, Karolinska Institutet, Stockholm, 
Sweden. Electronic address:.

The commercial production of infant food is undergoing rapid development, but 
control is limited. This study measured several toxic and essential elements in 
77 food products intended for infants (<1 year). Concentrations of toxic metals 
in infant formulas were generally low, but cadmium and cobalt were higher than 
in breast milk, as were iron, manganese and selenium concentrations. Elevated 
arsenic in a rice-based dairy- and gluten-free porridge (3.2 μg/portion) and 
cadmium in some porridges (∼1 μg/portion) raise concern. Further, manganese in 
gruels and porridges with ∼1 mg/portion (not fortified), and even formulas 
(∼0.1 mg/portion; fortified) reached concerning exposure levels, especially if 
mixed with water containing manganese. Estimated intake of inorganic arsenic 
(median 1.7 μg/day, excluding fish) and cadmium (median 3.4 μg/day) from mixed 
foods at 9 months of age exceeded health-based guidance values. Continued 
control of contaminants and additives in infant food and evaluation of related 
health risks are warranted.

Copyright © 2026 The Authors. Published by Elsevier Ltd.. All rights reserved.

DOI: 10.1016/j.foodchem.2026.148337
PMID: 41747551 [Indexed for MEDLINE]

Conflict of interest statement: Declaration of competing interest The authors 
declare that they have no known competing financial interests or personal 
relationships that could have appeared to influence the work reported in this 
paper.


34. Food Microbiol. 2026 Aug;137:105045. doi: 10.1016/j.fm.2026.105045. Epub 2026 
Jan 27.

Chemical profiling, molecular docking, and harnessing Manuka honey's 
antimicrobial and prebiotic potential in probiotic milk and yoghurt.

Ibrahim N(1), Moussa AY(1), Taher EM(2), Luo J(3), Xu B(4), Ibrahim Ahmed L(2).

Author information:
(1)Department of Pharmacognosy, Faculty of Pharmacy, Ain Shams University, 
Abbassia, 11566, Cairo, Egypt.
(2)Department of Food Hygiene and Control, Faculty of Veterinary Medicine, Cairo 
University, Giza, 12211, Egypt.
(3)Food Science and Technology Program, Department of Life Sciences, Beijing 
Normal-Hong Kong Baptist University, Zhuhai, Guangdong 519087, China.
(4)Food Science and Technology Program, Department of Life Sciences, Beijing 
Normal-Hong Kong Baptist University, Zhuhai, Guangdong 519087, China. Electronic 
address:.

The nutritional and therapeutic properties of Manuka honey (MH) constituted a 
major interest of the scientific community. The current study characterized the 
chemical composition of New Zealand MH via LC-HRMS/MS unveiling its richness in 
59 compounds. MH antimicrobial activity was assessed against 7 microbial strains 
where E. coli was the most sensitive followed by P. aeruginosa. Two longitudinal 
studies were conducted, one examined MH potential use as a prebiotic with 
Bifidobacterium bifidum in functional probiotic milk, while the other examined 
its feasibility as a natural antimicrobial additive against E. coli ATCC 25922 
in lab-manufactured yoghurt while maintaining the starter culture activity. MH 
(10 %) had no adverse effect on yoghurt starter culture activity despite 
significantly enhancing B. bifidum growth in probiotic milk (p > 0.05). 
MH-fortified yoghurt showed higher sensory acceptability with no significant 
difference compared to control group (p < 0.05) despite the lower overall 
acceptability score of MH-fortified milk. Furthermore, E. coli viability was 
significantly reduced by two logs in 10 % MH-fortified pasteurized milk by the 
end of the 14th day and completely disappeared by the 5th day of storage period 
of 10 % MH-fortified yoghurt (p < 0.05). Additionally, molecular docking studies 
in E. coli DNA gyrase and Pseudomonas quorum sensing QscR enzymes manifested the 
role of eleven of MH components as powerful binders of these enzymatic pockets; 
thus, further confirming its antimicrobial and quorum sensing inhibitor 
activities in the biological system. Finally, this study emphasized that MH is 
an exquisite natural functional alternative, which opens new avenues for MH's 
application in dairy sector, not only as a natural sweetener but also as a 
prebiotic and natural antimicrobial food preservative.

Copyright © 2026 Elsevier Ltd. All rights reserved.

DOI: 10.1016/j.fm.2026.105045
PMID: 41722988 [Indexed for MEDLINE]

Conflict of interest statement: Declaration of competing interest The authors 
declare that they have no known competing financial interests or personal 
relationships that could have appeared to influence the work reported in this 
paper.


35. Braz J Med Biol Res. 2026 Feb 16;59:e14750. doi: 10.1590/1414-431X2025e14750. 
eCollection 2026.

Perinatal and laboratory determinants of fortified human milk use: a 
case-control study.

Tavares CBG(1), Zacarias JMV(1)(2), Souza VH(3), Visentainer JV(4), Visentainer 
JEL(1).

Author information:
(1)Programa de Pós-Graduação em Biociências e Fisiopatologia, Universidade 
Estadual de Maringá, Maringá, PR, Brasil.
(2)Departamento de Cadeiras Básicas, Faculdade de Medicina de Marília, Marília, 
SP, Brasil.
(3)Departamento de Medicina, Universidade Cesumar (UNICESUMAR), Maringá, PR, 
Brasil.
(4)Programa de Pós-Graduação em Ciências de Alimentos, Universidade Estadual de 
Maringá, Maringá, PR, Brasil.

This case-control study compared perinatal and laboratory factors between 
preterm infants who received unfortified human milk (HM) and those who received 
HM fortified with FM85®. The sample included 38 low birth weight (LBW) preterm 
infants admitted to a neonatal intensive care unit between June 2022 and May 
2023, excluding those with comorbidities or formula use. Perinatal variables 
including gestational age, birth weight, head circumference (HC), and mode of 
delivery were analyzed for associations with fortifier use using chi-squared and 
Student's t-tests. Laboratory values were compared across nutritional stages 
using the Kruskal-Wallis and Mann-Whitney tests. Very preterm infants 
(confidence interval [CI]: 4.02-21.10; P<0.001) and those with LBW (CI: 
1.30-5.75; P=0.009) were more likely to receive the fortifier, particularly 
those with poor weight progression during trophic feeding (CI: 1.12-16.30; 
P=0.039). Altered HC (CI: 1.49-8.24; P=0.004) and cesarean delivery (CI: 
2.24-11.30; P<0.001) were also associated with FM85® use. Both groups showed 
laboratory abnormalities with neutrophils, initially elevated, decreasing 
(P<0.0001), while lymphocytes increased (P<0.0001). Red blood cell (P=0.00026) 
and hemoglobin levels (P=0.00007) worsened. Sodium and calcium levels rose 
significantly (P<0.0001 for both). HM fortification did not significantly alter 
most laboratory values, except for red blood cell count. LBW, altered HC, and 
cesarean delivery were associated with a greater likelihood of fortifier use. 
Immunological changes occurred even with fortification. Homologous fortifiers 
may represent a more physiological alternative.

DOI: 10.1590/1414-431X2025e14750
PMCID: PMC12919753
PMID: 41711763 [Indexed for MEDLINE]


36. Syst Rev. 2026 Feb 19;15(1):98. doi: 10.1186/s13643-025-03027-3.

Effect of nutrient supplementation on somatic growth in very low birth weight 
infants: a protocol for a systematic review and network meta-analysis.

Satardien M(1), Mccaul M(2), Van Niekerk E(3), van Weissenbruch M(4), Van Wyk 
L(1).

Author information:
(1)Department of Paediatrics and Child Health, Faculty of Medicine and Health 
Sciences, Stellenbosch University, Cape Town, South Africa.
(2)Division of Epidemiology and Biostatistics, Stellenbosch University, Cape 
Town, South Africa..
(3)Department of Dietetics, Faculty of Medicine and Health Sciences, 
Stellenbosch University, Cape Town, South Africa..
(4)Emma Children's Hospital, Division IC Neonatology, VU University, Amsterdam, 
The Netherlands..

AIM: No consensus exists on the ideal nutrition for preterm infants. This leads 
to significant practice variation. Fortification of expressed breast milk (EBM) 
and nutrient supplementation of the preterm infant's diet have become common 
practices to enhance growth. This systematic review aims to review the 
comparative effectiveness of oral macro- and micronutrient supplements in 
enhancing post-natal growth for preterm infants. Additionally, to identify 
critical gaps in the current recommendations for nutrient supplements.
METHODS: We will conduct a systematic review and network meta-analysis (NMA). 
Websites and databases will be searched for randomised controlled trials and 
prospective observational studies investigating oral macro and micro-nutrient 
supplementation to improve in-hospital somatic growth for premature very low 
birth weight infants (VLBWIs, < 1500 g). Two review authors will assess 
full-text English publications between 2010-2024 for potentially relevant 
studies for inclusion, independently and in duplicate, utilising an eligibility 
form based on the inclusion criteria. The selection process will be demonstrated 
graphically utilising a PRISMA flow diagram. Details and characteristics of 
excluded studies will be provided. A network meta-analysis (NMA) will be done 
using a frequentist approach and multivariate meta-analysis. Random-effect 
models will be employed to estimate all relative treatment effects. Stata will 
be used for data analysis. All possible comparisons, containing the effect size 
and 95% CIs, will be reported in tabular form. If the assumptions that preserve 
the validity of the NMA are not met, a pairwise meta-analysis will be done. If 
the criteria for pair-wise meta-analysis are not met, only direct comparisons 
will be made, and a narrative description of the findings will be presented 
employing the synthesis without meta-analysis guidelines. The GRADE approach 
will be used to review the certainty of the evidence. For each comparison, the 
overall certainty of evidence for the primary outcomes will be evaluated. 
Estimates of the direct and/or indirect evidence of the NMA will be provided.
RESULTS: The NMA will generate comparative rankings of nutrient supplementation 
interventions and evaluate their effectiveness in improving in-hospital growth. 
Results will be presented as summary tables and SUCRA rankings.
CONCLUSION: This NMA will provide evidence-based guidance on optimal nutrient 
supplementation strategies to enhance postnatal growth in VLBWIs, addressing a 
critical knowledge gap in neonatal nutrition practices.
SYSTEMATIC REVIEW REGISTRATION: This protocol has been registered with PROSPERO.
REGISTRATION NUMBER: CRD420250650341.

© 2026. The Author(s).

DOI: 10.1186/s13643-025-03027-3
PMCID: PMC13020165
PMID: 41709340 [Indexed for MEDLINE]

Conflict of interest statement: Declarations. Ethics approval and consent to 
participate: This study protocol was reviewed and approved by the Health 
Research Ethics Committee of Stellenbosch University, Cape Town, South Africa 
(Approval Number: S24/03/052). As this is a systematic review and network 
meta-analysis based on previously published studies, no new human participants 
were recruited, and therefore, written informed consent was not required. 
Competing interests: The authors have no conflicts of interest to declare.


37. Curr Opin Clin Nutr Metab Care. 2026 May 1;29(3):299-305. doi: 
10.1097/MCO.0000000000001214. Epub 2026 Feb 18.

Measuring nutrient content in human milk: are we there yet?

Geddes DT(1)(2)(3), Stinson L(1)(2)(3).

Author information:
(1)School of Molecular Sciences, The University of Western Australia, Crawley.
(2)ABREAST Network, Perth.
(3)UWA Centre for Human Lactation Research and Translation, Crawley, Western 
Australia, Australia.

PURPOSE OF REVIEW: Precise nutrient quantification is essential for developing 
reference values, guiding fortification strategies and preventing growth 
deficits, particularly in preterm infants. This review examines current 
methodologies for measuring nutrient content in human milk, evaluating accuracy, 
practicality, and limitations of existing analytical approaches.
RECENT FINDINGS: Current analytical methods for human milk nutrients demonstrate 
a trade-off between accuracy and clinical practicality. While some methods 
provide the high accuracy for macronutrient measurement, they require large 
sample volumes and extensive laboratory time. Alternative approaches such as 
spectroscopic and colorimetric methods offer improved efficiency and smaller 
sample requirements but with varying degrees of accuracy. Significant 
methodological challenges persist across all approaches, including the lack of 
standardized sampling protocols that account for temporal variation in milk 
composition, and difficulties in adapting analytical technologies originally 
designed for other matrices. These limitations are particularly problematic for 
micronutrient analysis, where sample degradation and storage conditions could 
impact results.
SUMMARY: While multiple analytical approaches for human milk exist, significant 
methodological limitations compromise accuracy and clinical utility. The field 
requires standardized workflows encompassing rigorous sampling protocols, 
validated storage conditions, and comprehensive method validation. Future 
developments should bridge the gap between research-grade accuracy and clinical 
practicality.

Copyright © 2026 Wolters Kluwer Health, Inc. All rights reserved.

DOI: 10.1097/MCO.0000000000001214
PMID: 41704036 [Indexed for MEDLINE]


38. JPGN Rep. 2025 Nov 3;7(1):150-158. doi: 10.1002/jpr3.70103. eCollection 2026 
Feb.

Current enteral feeding practices for premature infants in neonatal intensive 
care units in the Middle East and North Africa.

Metwally MH(1), Awad H(1), Samy G(1), Ayed M(2), Swanson J(3), Al-Nemri A(4), 
Salah M(5), Elliott M(6), Sullivan S(7), Al-Mudeer A(8), Bahbah W(9).

Author information:
(1)Department of Pediatrics and Neonatology Ain Shams University Cairo Egypt.
(2)Neonatal Department Maternity Hospital Andalous Kuwait.
(3)Division of Neonatology University of Virginia Children's Hospital 
Charlottesville Virginia USA.
(4)Pediatric Department King Saud University College of Medicine, King Saud 
University Medical City Riyadh Saudi Arabia.
(5)Head of Egyptian Academy of Pediatrics Cairo Egypt.
(6)Pediatrix Medical Group, Prolacta Bioscience Rockville Maryland USA.
(7)Envision Health Nashville Tennessee USA.
(8)Department of Pediatrics King Saud University Medical City Riyadh Saudi 
Arabia.
(9)Department of Pediatrics, Faculty of Medicine Menoufia University Shebin 
El-Kom Egypt.

OBJECTIVES: We aimed to gather insights from physicians regarding feeding 
practices for premature infants in the Middle East and North Africa.
METHODS: An online survey was distributed among physicians who managed premature 
infants. Descriptive analyses were used to evaluate the responses.
RESULTS: In total, 1000 out of 1300 participants from Egypt, Iraq, Kuwait, 
Qatar, Saudi Arabia, Syria, and the United Arab Emirates completed the survey. 
As reported, the participants included neonatologists (n = 678), general 
pediatricians (n = 258), and pediatric gastroenterologists (n = 14). Nearly half 
the participants (n = 466; 49.2%) had over 10 years of experience. Overall, 
53.4% of participants followed available feeding protocols, 16.8% inconsistently 
adhered to available protocols, and 14.8% reported unavailability of feeding 
protocols in their practice. Some participants (36%) had concerns about 
feeding-related complications with early initiation of enteral feeding. We 
observed variations in feeding practices as well as the management and 
monitoring of feeding-related complications in preterm infants among the 
participants. Additionally, awareness of human milk fortifiers was variable and 
correlated with clinical experience.
CONCLUSION: There is a need for training healthcare professionals, standardizing 
feeding protocols, and optimizing care for preterm infants across the region to 
reduce feeding-related complications and improve long-term health outcomes.

© 2025 The Author(s). JPGN Reports published by Wiley Periodicals LLC on behalf 
of The European Society for Pediatric Gastroenterology, Hepatology, and 
Nutrition and the North American Society for Pediatric Gastroenterology, 
Hepatology, and Nutrition.

DOI: 10.1002/jpr3.70103
PMCID: PMC12894073
PMID: 41695050

Conflict of interest statement: Melinda Elliot is affiliated with Prolacta 
Bioscience. The remaining authors declare no conflicts of interest.


39. Iodine.

Drugs and Lactation Database (LactMed®) [Internet]. Bethesda (MD): National 
Institute of Child Health and Human Development; 2006–.
2026 Feb 15.

Iodine is an essential trace nutrient for all infants that is a normal component 
of breastmilk. Infant requirements are estimated to be 15 mcg/kg daily in 
full-term infants and 30 mcg/kg daily in preterm infants.[1] Breastmilk iodine 
concentration correlates well with maternal urinary iodine concentration and may 
be a useful index of iodine sufficiency in infants under 2 years of age, but 
there is no clear agreement on a value that indicates iodine sufficiency, and 
may not correlate with infant thyroid function tests.[2-4] A milk iodine value 
of 100 to 200 mcg/L has been suggested to indicate adequate iodine status in 
lactating women,[5] although a range as wide as 60 to 465 mcg/L has been 
proposed.[4] Systematic reviews and studies on iodine nutrition found that 
iodine in breastmilk is adequate in iodine-sufficient countries, but in 
countries with iodine fortification of foods, many mothers did not obtain 
adequate iodine and that additional supplementation was desirable.[6-10] In 
iodine-deficient areas, supplementation of breastfeeding mothers with iodine 
appears to be more effective than direct supplementation of the infant in 
reducing infant iodine deficiency.[11] In the United States, a total iodine 
intake of 290 mcg daily is recommended during lactation; the WHO recommends a 
total intake of 250 mcg daily.[9] The American Thyroid Association recommends 
that breastfeeding women should supplement their diet with a daily oral 
supplement that contains 150 mcg of iodine, but sustained iodine intake while 
breastfeeding that exceeds 500 to 1100 mcg daily should be avoided.[12] Studies 
of lactating women in the United States found that the median urinary iodine 
concentration between 2001 and 2020 was 103.0 mcg/L (95% CI 86.1 to 143.5), 
indicating that many women fell below the minimum level of 100 mcg/L.[13] Vegan 
and vegetarian mothers appear to have reduced iodine concentrations in their 
milk and should use iodine supplements.[14] A survey in the United States 
between 2011 and 2014 found that only 19% of lactating women used a dietary 
supplement that contained iodine.[15] One study found improved neurodevelopment 
in the infants of mothers who received 150 mcg of iodine daily during 
breastfeeding, but a dose of 300 mcg daily did not further improve the 
outcome.[16] The use of excessive amounts of iodine in the mother near term and 
during breastfeeding (e.g., seaweed soup) can increase breastmilk iodine levels 
and cause transient hypothyroidism in breastfed infants.[17-20] Follow-up at 3 
years of age found no long-term thyroid dysfunction in infants exposed to high 
iodine levels in breastmilk.[21] Supplementing mothers with Grave disease in 
appropriate doses potassium iodide usually does not adversely affect their 
breastfed infants, although a few may develop mild hypothyroidism.[22,23] The 
absorption of iodine can be marked after application to open wounds or mucous 
membranes. Exposure of mothers to unnecessary iodine who are or will be 
breastfeeding should be avoided or minimized to the extent possible by avoiding 
its use on maternal mucous membranes (e.g., vaginal use, wound therapy), 
avoiding prolonged contact time, avoiding repeated applications, and applying it 
to the smallest possible surface areas of the body. It is possible that maternal 
exposure to iodine near term could interfere with thyroid studies done as a part 
of newborn screening tests. When treating lactating people after radiation 
exposure, one dose of potassium iodine 130 mg is recommended at the lowest 
threshold exposure of >5 Gy to minimize the risk of blocking thyroid function in 
the fetus or nursing infant. In cases of severe contamination in lactating 
people, repeat dosing may be given and nursing neonates should be monitored for 
signs of illness.[24]

PMID: 30000537


40. Food Chem. 2026 Apr 15;508(Pt A):148378. doi: 10.1016/j.foodchem.2026.148378. 
Epub 2026 Feb 10.

Citrus flavonoid-enriched milk protects against intestinal oxidative stress and 
barrier dysfunction in mice.

He Y(1), Qin Y(1), Xu W(1), Yan J(1), Shen Y(1), Ye T(1), Niu H(1), Tan J(1), 
Jiang L(2), Zhao Y(3).

Author information:
(1)Beijing Key Laboratory of Dairy Cow Nutrition, College of Animal Science and 
Technology, Beijing University of Agriculture, Beijing 102206, China.
(2)Beijing Key Laboratory of Dairy Cow Nutrition, College of Animal Science and 
Technology, Beijing University of Agriculture, Beijing 102206, China. Electronic 
address:.
(3)Beijing Key Laboratory of Dairy Cow Nutrition, College of Animal Science and 
Technology, Beijing University of Agriculture, Beijing 102206, China. Electronic 
address:.

This study investigated the impact of dietary citrus flavonoid extract (CFE) 
supplementation in dairy cows on milk composition and its protective effects 
against intestinal oxidative stress. Supplementation with 150 g/d CFE 
significantly enriched bovine milk with bioactive flavonoids, including 
hesperidin, narirutin, naringin, a their conjugated metabolites, as well as 
polymethoxyflavones such as nobiletin and tangeretin. These changes were 
accompanied by elevated total phenolic and flavonoid concentrations and enhanced 
milk antioxidant capacity. In a lipopolysaccharide (LPS)-challenged mouse model, 
CFE-enriched milk improved systemic and intestinal antioxidant enzyme 
activities, reduced inflammatory cytokine production, and upregulated tight 
junctions and Nrf2/HO-1/NQO1 pathway gene expression. Histological analyses 
confirmed restoration of intestinal morphology and goblet cell abundance. 
Moreover, supplementation modulated gut microbiota, increasing beneficial taxa 
(e.g., Lactobacillus, Bacteroides) and reducing harmful genera (e.g., 
Desulfovibrio). Collectively, these findings demonstrate that citrus 
flavonoid-fortified milk confers antioxidant, anti-inflammatory, and 
gut-protective effects, supporting its potential as a functional dairy product.

Copyright © 2026 Elsevier Ltd. All rights reserved.

DOI: 10.1016/j.foodchem.2026.148378
PMID: 41687380 [Indexed for MEDLINE]

Conflict of interest statement: Declaration of competing interest The authors 
declare that they have no known competing financial interests or personal 
relationships that could have appeared to influence the work reported in this 
paper.


41. Arch Dis Child Fetal Neonatal Ed. 2026 Feb 12:fetalneonatal-2025-329670. doi: 
10.1136/archdischild-2025-329670. Online ahead of print.

International survey on enteral nutrition, supplementation and probiotic 
practices for preterm infants.

Klingenberg C(1)(2), Aunsholt L(3)(4), Van Den Akker CHP(5), Häertel C(6), Sáenz 
de Pipaón M(7), Giannoni E(8), O'Donnell CPF(9)(10), Fenton T(11), Lehtonen 
L(12)(13), Bohlin K(14)(15), Heiring C(3), Pereira-Da-Silva L(16)(17), Kuschel 
CA(18), Embleton ND(19).

Author information:
(1)Research Group for Child and Adolescent Health, Department of Clinical 
Medicine, UiT The Arctic University of Norway, Tromsø, Norway 
.
(2)Department of Paediatric and Adolescent Medicine, University Hospital of 
North Norway, Tromsø, Norway.
(3)Department of Neonatology and Pediatric Intensive Care, Copenhagen University 
Hospital, Rigshospitalet, Copenhagen, Denmark.
(4)Faculty of Health and Medical Sciences, University of Copenhagen, Copenhagen, 
Denmark.
(5)Department of Pediatrics-Neonatology, Emma Children's Hospital, Amsterdam 
UMC, University of Amsterdam, Amsterdam, The Netherlands.
(6)Department of Pediatrics, University Hospital of Würzburg, Wurzburg, Germany.
(7)Neonatology, Hospital Universitario La Paz, Madrid, Spain.
(8)Clinic of Neonatology, Department Mother-Woman-Child, Lausanne University 
Hospital and University of Lausanne, Lausanne, Switzerland.
(9)Neonatal Unit, National Maternity Hospital Neonatal Unit, Dublin, Ireland.
(10)School of Medicine, University College Dublin, Dublin, Ireland.
(11)Community Health Sciences, O'Brien Institute of Public Health, Cumming 
School of Medicine, Alberta Children's Hospital Research Institute, University 
of Calgary, Calgary, Alberta, Canada.
(12)Department of Paediatrics and Adolescent Medicine, Turku University 
Hospital, Turku, Finland.
(13)Department of Clinical Medicine, University of Turku, Turku, Finland.
(14)Department of Clinical Science, Intervention and Technology, Karolinska 
Institutet, Stockholm, Sweden.
(15)Department of Neonatology, Karolinska University Hospital, Stockholm, 
Sweden.
(16)Medicine of Woman, Childhood and Adolescence Academic Area, NOVA Medical 
School, Faculdade de Ciências Médicas, Universidade Nova de Lisboa, Lisbon, 
Portugal.
(17)Neonatal Intensive Care Unit, Department of Pediatrics, Hospital Dona 
Estefânia, Unidade Local de Saúde São José, Lisbon, Portugal.
(18)Neonatal Services, Royal Women's Hospital, Parkville, Victoria, Australia.
(19)Neonatology, Newcastle Upon Tyne Hospitals NHS Foundation Trust, Newcastle 
Upon Tyne, UK.

OBJECTIVE: To evaluate enteral feeding practices in preterm infants in neonatal 
intensive care units (NICUs) in high-income countries across three continents, 
and compare results with a similar survey 13 years earlier.
METHODS: Web-based survey distributed to neonatologists at 258 NICUs across 15 
countries in Europe, Australia, New Zealand and Canada, October 2023 and 
February 2024. Survey domains focused on availability of human milk, onset of 
enteral feeding, breast milk fortification (BMF), cytomegalovirus (CMV) 
screening and enteral supplements including probiotics. Results were compared 
with a similar survey performed in 2010.
RESULTS: Replies were received from 185 (72%) NICUs. Access to donor human milk 
(DHM) was high (91%). Across all NICUs, feeds were started on day 1 in 64%, 73% 
and 85% among infants born <25, 25-27 and 28-31 weeks' gestation, respectively. 
Bovine milk-based BMF was routinely used in 88% of NICUs, with large variation 
in when it was commenced and discontinued. Routine use of human milk-based BMF 
was uncommon (4%). Maternal CMV status was routinely determined in 33% of all 
NICUs who then pasteurised or froze milk if the mother was CMV-seropositive. 
Probiotics were provided in 66% of the NICUs, with large variations in products 
and birth weight/gestational age criteria.
CONCLUSIONS: Compared with our survey from 2010, more NICUs now start feeding 
preterm infants on day 1, and DHM availability has increased in some countries. 
Substantial variation remains in the use of BMF, probiotics and CMV screening. A 
stronger evidence base is needed to update guidelines, aiming ultimately to 
improve growth and long-term neurodevelopment.

© Author(s) (or their employer(s)) 2026. No commercial re-use. See rights and 
permissions. Published by BMJ Group.

DOI: 10.1136/archdischild-2025-329670
PMID: 41679961

Conflict of interest statement: Competing interests: NDE discloses research 
grants paid to his institution for investigator-initiated studies from Prolacta 
Biosciences (2017–2021), Danone Early Life Nutrition (2017–2021), NeoKare 
(2021–2022); research fees paid to his institution for acting as investigator 
for ELGAN pharma (2024 ongoing); in the last 3 years, speaker or consultancy 
fees from Chiesi, Nestlé Nutrition Institute, Abbott all donated to charity; 
medicolegal consultancy for US legal firms involved in class action for NEC, 
fees donated to charity; non-remunerated advice to WHO for a probiotic trial in 
which he is not involved. CHPvdA has participated as a clinical investigator, 
and/or consultant, and/or speaker for ELGAN Pharma, Nestlé Nutrition Institute 
and Nutricia Early Life Nutrition. Reimbursements used as research funds within 
the hospital. MSdP discloses research grants paid to his institution for 
investigator-initiated studies from Mead Johnson (2021–2022); research fees paid 
for acting as investigator for ELGAN pharma (2024 ongoing); in the last 3 years, 
speaker or consultancy fees from Chiesi, Nestlé and Nutricia. None of the other 
authors report any relevant conflicts of interest to declare.


42. Lancet Planet Health. 2026 Feb;10(2):101423. doi: 10.1016/j.lanplh.2025.101423. 
Epub 2026 Feb 9.

Nutritional and health impacts of alternative animal source foods: a scoping 
review.

Beal T(1), McLaren R(2), Saxena T(3), Sokourenko K(3), Nordhagen S(4), 
Lambertini E(2), DiGiovine E(3), Xipsiti M(5), Neufeld LM(5).

Author information:
(1)Global Alliance for Improved Nutrition, Washington, DC, USA. Electronic 
address:.
(2)Global Alliance for Improved Nutrition, Washington, DC, USA.
(3)Cornell University, Ithaca, NY, USA.
(4)Global Alliance for Improved Nutrition, Geneva, Switzerland.
(5)Food and Agriculture Organization of the United Nations, Viale delle Terme di 
Caracalla, Rome, Italy.

Despite rapid growth in the availability and consumption of alternative animal 
source foods (Alt-ASFs), evidence remains fragmented and uneven across food 
types, outcomes, and settings, with few studies directly comparing their 
nutritional quality and health impacts with those of conventional animal 
products in equivalent substitutions-creating uncertainty for dietary guidance 
and policy. This scoping review addresses these gaps by systematically 
evaluating 247 studies comparing the nutritional quality and health impacts of 
Alt-ASFs with those of conventional animal products. Alt-ASFs showed mixed 
nutritional profiles. Plant-based and fungi-based alternatives generally 
provided high fibre and low saturated fat, with improvements in lipid profiles 
and cardiovascular markers. However, plant-based and fungi-based alternatives 
typically contained low bioavailable protein (except soy milk), vitamin B12, 
zinc, and iron. Plant-based milks, except fortified soy, were nutritionally 
inferior to cow's milk, particularly in terms of protein and key micronutrients. 
Insect-based foods showed diverse nutritional profiles, and in some cases, they 
provided protein and mineral contents similar or superior to those of 
conventional meats. Research was overwhelmingly concentrated in high-income 
countries (76%), with no studies conducted in low-income settings. Considerable 
gaps remain in the evidence base for algae-based and cell-based foods, long-term 
health outcomes, and underserved populations. Although Alt-ASFs offer promise 
for supporting healthy and sustainable diets, their integration into dietary 
patterns requires careful consideration, and potentially incorporation of 
complementary food choices or targeted fortification, to prevent nutrient 
inadequacies, particularly among nutritionally at-risk groups.

Copyright © 2025 The Author(s). Published by Elsevier Ltd.. All rights reserved.

DOI: 10.1016/j.lanplh.2025.101423
PMID: 41679326 [Indexed for MEDLINE]

Conflict of interest statement: Declaration of interests We declare no competing 
interests.


43. Food Chem. 2026 Apr 15;508(Pt A):148316. doi: 10.1016/j.foodchem.2026.148316. 
Epub 2026 Feb 4.

Evaluating amino acid fortification in low-dairy ready-to-use therapeutic food 
formulations: Insights from a meta-analysis re-evaluation.

Berhane M(1), Toni AT(2), Gonzales GB(3).

Author information:
(1)Jimma University Clinical and Nutrition Research Center (JUCAN), Jimma 
University, Jimma P.O Box: 378, Ethiopia; Department of Pediatrics and Child 
Health, Faculty of Medical Sciences, Institute of Health, Jimma University, 
Jimma, Ethiopia; Department of Public Health and Primary Care, Faculty of 
Medicine and Health Sciences, Ghent University, C. Heymanslaan 10, 9000 Ghent, 
Belgium.
(2)Department of Pediatrics and Child Health, College of Medicine and Health 
Sciences, University of Gondar, Gondar, Ethiopia; Division of Human Nutrition 
and Health, Wageningen University and Research, Wageningen, the Netherlands.
(3)Department of Public Health and Primary Care, Faculty of Medicine and Health 
Sciences, Ghent University, C. Heymanslaan 10, 9000 Ghent, Belgium. Electronic 
address:.

Severe wasting is treated with ready-to-use therapeutic food (RUTF), wherein the 
protein content primarily comes from milk and peanuts. As these ingredients are 
the main drivers of RUTF's cost, global initiatives seek alternatives using 
locally available, sustainable ingredients to reduce cost and increase coverage. 
To compensate for reduced/removed dairy protein, current practice is to add 
amino acids to meet Codex protein quality requirements. In this meta-analysis 
re-evaluation, we compared RUTF formulations with <50% dairy protein, with or 
without amino acid fortification, against standard milk-peanut RUTF for the 
outcome of weight gain. Growth outcomes were similar between fortified and 
unfortified RUTFs when compared to standard RUTF. These findings suggest that 
elevating protein quality scores by adding amino acids may be insufficient to 
ensure clinical efficacy in novel RUTF formulations. Additionally, the findings 
suggest that Protein Digestibility Corrected Amino Acid Score (PDCAAS) 
compliance alone may not guarantee clinical efficacy. Future formulation efforts 
should address digestibility, antinutritional factors, and increasing total 
protein content to achieve effective, sustainable novel RUTF formulations.

Copyright © 2026 Elsevier Ltd. All rights reserved.

DOI: 10.1016/j.foodchem.2026.148316
PMID: 41672015 [Indexed for MEDLINE]

Conflict of interest statement: Declaration of competing interest The authors 
declare that they have no known competing financial interests or personal 
relationships that could have appeared to influence the work reported in this 
paper.


44. Eur J Clin Nutr. 2026 Mar;80(3):263-269. doi: 10.1038/s41430-026-01701-w. Epub 
2026 Feb 7.

Long-term growth effects of post-discharge formula in moderate-to-late preterm 
infants: follow-up at 24 months corrected age of a randomized controlled trial.

Muts J(1)(2), Ruys CA(2)(3), Finken MJJ(2)(4), Rotteveel J(4), van Goudoever 
JB(1)(2), van den Akker CHP(2)(3), van de Lagemaat M(5)(6); LEGO study group.

Collaborators: van den Akker CHP, van de Lagemaat M, Ruys CA, Schrijver-Levie 
NS, Boonstra V, von Lindern JS, de Winter P, van Brakel MJ, Drewes AJ, Westra M.

Author information:
(1)Department of Pediatrics, Emma Children's Hospital, Amsterdam UMC, University 
of Amsterdam, Amsterdam, The Netherlands.
(2)Amsterdam Reproduction & Development Research Institute, Amsterdam UMC, 
Amsterdam, The Netherlands.
(3)Department of Pediatrics-Neonatology, Emma Children's Hospital, Amsterdam 
UMC, University of Amsterdam, Amsterdam, The Netherlands.
(4)Department of Pediatric Endocrinology, Emma Children's Hospital, Amsterdam 
UMC, VU University, Amsterdam, The Netherlands.
(5)Amsterdam Reproduction & Development Research Institute, Amsterdam UMC, 
Amsterdam, The Netherlands..
(6)Department of Pediatrics-Neonatology, Emma Children's Hospital, Amsterdam 
UMC, University of Amsterdam, Amsterdam, The Netherlands. 
.

BACKGROUND: Moderate-to-late preterm (MLP) infants (i.e., <37 weeks' gestation) 
are at risk for suboptimal neonatal nutrition and growth. We compared 
anthropometry and body composition at 24 months corrected age (CA; i.e., after 
term equivalent age (TEA)) between MLP infants fed either an isocaloric, 
protein- and mineral-enriched post-discharge formula (PDF) or standard term 
formula (STF) between TEA and 6 months CA.
METHODS: After enrollment (≤7 d postpartum), MLP infants received PDF if 
(fortified) mother's own milk was insufficient. At TEA, those receiving >25% of 
intake as formula were randomized to either continue the same PDF or switch to 
STF until 6 months CA. At 24 months CA, anthropometry was assessed and 
air-displacement plethysmography was used to estimate body composition.
RESULTS: From the original study (n = 157), only 64 infants had anthropometry 
assessed and 34 infants had body composition estimated at 24 months CA. At the 
follow-up visit, PDF-fed infants had slightly larger head circumference compared 
to STF-fed infants (49.0 ± 1.5 and 48.3 ± 1.5 cm, p < 0.05). Despite similar 
total body weight at 24 months CA, PDF-fed infants had lower fat mass and higher 
fat-free mass compared to STF-fed infants (fat mass: 2392 ± 923 and 
3615 ± 1359 g; fat-free mass: 10461 ± 1030 and 10108 ± 1700 g; respectively, 
p < 0.05).
CONCLUSIONS: Providing MLP infants with PDF for 6 months after TEA was 
associated with lower fat mass and higher fat-free mass, with similar 
bodyweights at 24 months CA compared to those fed STF. Our findings should be 
interpreted cautiously, due to high loss-to-follow-up.

© 2026. The Author(s), under exclusive licence to Springer Nature Limited.

DOI: 10.1038/s41430-026-01701-w
PMID: 41654679 [Indexed for MEDLINE]

Conflict of interest statement: Competing interests: M. van de Lagemaat, CA 
Ruys, J Muts, MJJ Finken, and J Rotteveel: all report no conflicts of interest. 
JB van Goudoever: Member of the Dutch National Health Institute and founder and 
director of the Dutch National Human Milk Bank. CHP van den Akker reports 
receipt of speakers' and consultancy honoraria from Nestlé Nutrition Institute, 
Nutricia Early Life Nutrition, and Baxter; used as research funds. He is 
vice-director of the Dutch National Human Milk Bank and a member of the National 
Breastfeeding Council since 2024. Ethics approval: The LEGO study was reviewed 
and approved by the Medical Research Ethics Committee of Amsterdam UMC, location 
Vrije Universiteit Amsterdam, Amsterdam, The Netherlands (NTR NL4979).


45. Food Res Int. 2026 Mar 1;227:118124. doi: 10.1016/j.foodres.2025.118124. Epub 
2025 Dec 20.

Comparative analysis of protein composition, digestion dynamics, and amino acid 
release between preterm infant formula and human milk: Implications for preterm 
infant formula optimization.

Liu Y(1), Mu G(1), Yang X(2), Cheng X(2), Kang J(2), Zhu X(3).

Author information:
(1)School of Food Science and Technology, Dalian Polytechnic University, Dalian, 
PR China.
(2)Dalian Women and Children's Medical Group, Dalian, PR China.
(3)School of Food Science and Technology, Dalian Polytechnic University, Dalian, 
PR China. Electronic address:.

To advance precision nutrition and optimize feeding strategies for preterm 
infants, it is essential to further clarify the differences and interrelations 
in protein composition and digestive behavior between infant formulas (IFs) and 
human milk (HM). In this study, an integrated approach combining protein 
compositional analysis, protein digestion profiling, and targeted amino acid 
metabolomics was employed to systematically investigate these differences from 
the perspective of "protein composition-in vitro digestion behavior-amino acid 
release patterns." Our findings revealed that β-lactoglobulin (4-11 % of total 
protein in IFs, absent in HM) exhibited pronounced aggregation during digestion, 
markedly enhancing its resistance to peptic hydrolysis. Caseins, present at 
higher levels in IFs (37-48 % vs. 9.5-11 % in HM), further impeded enzymatic 
cleavage. In contrast, HM samples-particularly colostrum without human milk 
fortifier (HMF)-underwent a more gradual proteolytic process and displayed a 
more balanced amino acid release profile. Excessive release of certain amino 
acids (e.g., lysine, histidine, phenylalanine) in specific IFs may induce acute 
nitrogen metabolic imbalance in preterm infants. Glutamine and glutamic acid 
emerged as major contributors to the divergent amino acid release patterns 
between IF and HM, with their reduced liberation potentially linked to calcium 
soap formation and peptide masking. Additionally, the incorporation of 
full-strength HMF into HM appeared to obscure arginine-rich cleavage sites, 
which may partially account for the increased lactoferrin excretion observed in 
mixed feeding low-weight infants. Enhancing casein phosphopeptide content or 
aligning protein composition more closely with that of HM in IFs may represent 
promising strategies to reduce the nutritional outcome disparities between 
formula-fed and breastfed preterm infants.

Copyright © 2025 Elsevier Ltd. All rights reserved.

DOI: 10.1016/j.foodres.2025.118124
PMID: 41652728 [Indexed for MEDLINE]

Conflict of interest statement: Declaration of competing interest The authors 
declare that they have no known competing financial interests or personal 
relationships that could have appeared to influence the work reported in this 
paper.


46. J Food Sci. 2026 Feb;91(2):e70886. doi: 10.1111/1750-3841.70886.

Dictyophora Polysaccharides in Low-Fat Yogurt: Effects on Rheology, Texture, and 
Sensory Properties.

Li H(1), Liu J(2), Fan J(3), Tang L(1).

Author information:
(1)School of Public Health, Zunyi Medical University, Zunyi, Guizhou, People's 
Republic of China.
(2)Institute of Food Processing Technology, Guizhou Academy of Agricultural 
Sciences, Guiyang, People's Republic of China.
(3)School of Life and Health Science, Kaili University, Kaili, Guizhou, People's 
Republic of China.

The potential of Dictyostelium polysaccharide (DPS) as a fat substitute in 
low-fat yogurt was investigated in this study by analyzing its effects on 
rheology, physicochemical characteristics, textural properties, sensory quality, 
and microstructure. Compared with the control, DPS addition significantly 
enhanced the water-holding capacity and reduced syneresis of low-fat yogurt. 
Moreover, DPS did not alter the yogurt's color but improved its textural 
attributes and sensory acceptance. Rheological analysis showed that all low-fat 
yogurt samples exhibited weak gel-like behavior (G' > G″), and both storage 
modulus (G') and loss modulus (G″) were markedly higher in DPS-fortified samples 
than in the control. Scanning electron microscopy revealed that DPS 
incorporation resulted in a more compact and homogenous cross-linking between 
polysaccharides and casein. In conclusion, DPS demonstrates promising potential 
as a natural fat replacer in low-fat dairy products. PRACTICAL APPLICATIONS: DPS 
significantly enhanced the physicochemical properties and organoleptic quality 
of low-fat yogurt; DPS with medicine food homology can be exploited as a fat 
substitute for low-fat yogurt.

© 2026 Institute of Food Technologists.

DOI: 10.1111/1750-3841.70886
PMID: 41645878 [Indexed for MEDLINE]


47. J Food Sci. 2026 Feb;91(2):e70870. doi: 10.1111/1750-3841.70870.

Yogurt Fortification With Lyophilized Liposomes Coencapsulating Vitamins D(3) 
and B(12): Physicochemical Characterization, Sensory Evaluation and Static In 
Vitro Digestion.

Ferreira LS(1), Luvizzotti EH(1), Ghiraldi M(1), Chaves MA(1)(2), Pinho SC(1).

Author information:
(1)Laboratory of Encapsulation and Functional Foods (LEnAlis), Department of 
Food Engineering, School of Animal Science and Food Engineering (FZEA), 
University of São Paulo (USP), Pirassununga, Brazil.
(2)Laboratory of Molecular Morphophysiology and Development (LMMD), Department 
of Veterinary Medicine, School of Animal Science and Food Engineering (FZEA), 
University of São Paulo (USP), Pirassununga, Brazil.

Liposomes are lipid-based delivery systems of interest in pharmaceutical, food, 
and nutraceutical industries due to their ability to encapsulate bioactive 
compounds, such as vitamins, improving stability, solubility, and 
bioavailability. This study aimed to develop functional yogurt enriched with 
vitamins D3 and B12 by encapsulating these micronutrients in liposomal 
dispersions produced with food-grade phospholipids, coated with pectin, and 
subjected to lyophilization. An optimal sucrose concentration was selected as a 
cryoprotectant to minimize detrimental effects of freeze-drying on vesicle 
integrity. Lyophilized vesicles were characterized by bioactive retention and 
physicochemical properties. High retention of both vitamins was observed across 
most formulations, except those containing only hydrogenated saturated 
phospholipids, which showed reduced encapsulation efficiency. These vesicles 
were incorporated into yogurt, and the enriched product was evaluated for 
bioactive content, physicochemical and rheological properties, and sensory 
acceptability. The addition of vesicles did not negatively affect yogurt 
stability or consumer acceptance. In vitro digestion using the INFOGEST 2.0 
protocol revealed increased vitamin D3 bioaccessibility after the gastric phase 
only in samples containing Phospholipon 90G-based liposomes; however, after the 
intestinal phase, all formulations showed enhanced availability. Vitamin B12 
bioaccessibility remained consistent throughout digestion. Overall, results 
demonstrate a viable approach for coencapsulation and protection of hydrophobic 
and hydrophilic vitamins within a commonly consumed dairy product, offering a 
promising strategy for developing functional foods supporting daily nutritional 
supplementation. PRACTICAL APPLICATIONS: Pectin-coated and uncoated liposomes 
were successfully freeze-dried with sucrose as a cryoprotectant. Purified 
phospholipids improved the bioaccessibility of vitamin D3 after gastric 
digestion. Liposome enrichment did not alter yogurt stability or sensory 
acceptance. The strategy allows the co-administration of hydrophilic and 
hydrophobic vitamins in dairy products.

© 2026 The Author(s). Journal of Food Science published by Wiley Periodicals LLC 
on behalf of Institute of Food Technologists.

DOI: 10.1111/1750-3841.70870
PMCID: PMC12877996
PMID: 41645859 [Indexed for MEDLINE]

Conflict of interest statement: The authors declare no conflicts of interest.


48. Nutr Clin Pract. 2026 Jan 29. doi: 10.1002/ncp.70097. Online ahead of print.

Osmolality of fortified human milk with contemporary human milk fortifiers.

Gardiner A(1)(2), Flores González I(3)(4), Smith C(3), Wessel J(5), Kim 
JH(6)(7), Fu TT(6)(7).

Author information:
(1)Department of Pediatrics, Cincinnati Children's Hospital Medical Center, 
Cincinnati, Ohio, USA.
(2)Division of Neonatology, University of Tennessee Health Science Center, 
Memphis, Tennessee, USA.
(3)Division of Nutrition Therapy, Cincinnati Children's Hospital Medical Center, 
Cincinnati, Ohio, USA.
(4)Department of Neurology, Rockefeller Neuroscience Institute, West Virginia 
University, Morgantown, West Virginia, USA.
(5)Division of Pediatric Surgery, Cincinnati Children's Hospital Medical Center, 
Cincinnati, Ohio, USA.
(6)Perinatal Institute, Division of Neonatology, Cincinnati Children's Hospital 
Medical Center, Cincinnati, Ohio, USA.
(7)Department of Pediatrics, University of Cincinnati College of Medicine, 
Cincinnati, Ohio, USA.

BACKGROUND: Human milk fortification is critical to support preterm infant 
growth. Human milk fortifed to caloric densities above manufacturers' 
recommendations are often necessary to achieve adequate growth. Fortified milk 
may be prepared 12-24 h before feeding, although practice variations exist. We 
investigated how human milk osmolality is affected by contemporary bovine and 
human milk-derived human milk fortifiers (HMFs) immediately and temporally for 
standard and higher-calorie recipes.
METHODS: This study measured the osmolality of human milk fortified with four 
current HMFs across a range of caloric concentrations (22-30 kcal/oz) in fresh, 
previously frozen, and pasteurized human milk. Osmolality was tested at 0, 12, 
and 24 h after preparation, conducted in triplicate. Differences in initial 
osmolality were compared between fortifiers. Percentage change in osmolality at 
12 vs 24 h was tested with mixed-effects linear regression to study the 
effect/interaction of time, fortifier, and caloric density.
RESULTS: Initial osmolality was significantly different between manufacturers 
(range, 315-620 mOsm/kg across caloric densities). There was a median 0.3% (IQR, 
-0.8 to 1.5) difference in change in osmolality at 24 vs 12 h (P = 0.02). In 
mixed-effects modeling, there appeared to be an interaction between time and 
fortifier, but after post hoc adjustments, no pairwise comparisons remained 
significant. At 24 h, there was an interaction between fortifier and milk type 
(P = 0.003), with up to 4% greater change in osmolality with fresh vs frozen 
milk.
CONCLUSION: Contemporary HMFs increase the osmolality of human milk to 
variable degrees, with some temporal effects observed, although clinical 
significance of these findings is unclear.

© 2026 The Author(s). Nutrition in Clinical Practice published by Wiley 
Periodicals LLC on behalf of American Society for Parenteral and Enteral 
Nutrition.

DOI: 10.1002/ncp.70097
PMID: 41608871


49. RSC Adv. 2026 Jan 26;16(6):5641-5651. doi: 10.1039/d5ra08644b. eCollection 2026 
Jan 21.

Regulation of infant gut microbiota and metabolic pathways by human milk 
oligosaccharides: a multi-omics investigation.

Luo L(1)(2)(3), Yang B(2)(3)(4)(5), Zhao J(2)(3), Li X(2)(3), Qiao W(2)(3), 
Zhang M(2)(3), Tian L(2)(3), Chen L(1)(2)(3).

Author information:
(1)Key Laboratory of Dairy Science, Ministry of Education, Food College, 
Northeast Agricultural University Harbin 150030 China.
(2)National Engineering Center of Dairy for Maternal and Child Health, Beijing 
Sanyuan Foods Co. Ltd Beijing 100163 China.
(3)Beijing Engineering Research Center of Dairy, Beijing Sanyuan Foods Co. Ltd 
Beijing 100163 China.
(4)School of Food and Health, Beijing Technology and Business University Beijing 
100048 China.
(5)National Center of Technology Innovation for Dairy Hohhot 010100 China.

The third-most solid component in breast milk is human milk oligosaccharides 
(HMOs)-crucial for the growth and development of infants. HMOs are divided into 
three main categories: fucosylated, nonfucosylated, and sialylated HMOs. 
However, comparative studies evaluating the variations of the impact of in vitro 
fermentation of these typical or dominant HMOs on the metabolic functions of an 
infant's gut microbiome remain limited. Therefore, we systematically analyzed 
the role of three typical HMOs-2'-fucosyl lactose, lactose-N-tetrose, and 
3'-sialyllactose-in the structural reshaping of the infant intestinal flora; we 
also explored their impact on short-chain fatty acid (SCFA) production and 
metabolism, as well as their overall metabolic spectrum. We found that HMOs 
aided the growth of beneficial microorganisms, such as Bifidobacterium, 
Lactobacillus, and Enterococcus, while also reducing the number of harmful 
bacteria, including Escherichia-Shigella. Further, HMOs had a substantial impact 
on amino acid, purine, and lipid metabolic pathways and significantly increased 
SCFA levels. Correlation analyses revealed significant associations between 
Bifidobacterium and multiple lipid metabolites. Positive correlations between 
Lactobacillus and amino acid derivatives, as well as close links between 
Bacteroides and acetate production, were detected. Overall, these findings 
indicate that the gut flora-metabolite interplay is central to HMO function, and 
different HMOs variably regulate the gut microbiota composition and metabolic 
pathways. This study provides a theoretical basis for optimizing HMOs' 
fortification strategies in infant formulas.

This journal is © The Royal Society of Chemistry.

DOI: 10.1039/d5ra08644b
PMCID: PMC12834247
PMID: 41602199

Conflict of interest statement: The authors declare that they have no known 
competing financial interests or personal relationships that could have appeared 
to influence the work reported in this paper.


50. Nutrients. 2026 Jan 10;18(2):224. doi: 10.3390/nu18020224.

Vitamin D Deficiency Mediates the Link Between Dietary Patterns, Inflammatory 
Biomarkers, and Iron Status Indicators (Ferritin and Hemoglobin) in Metabolic 
Syndrome.

Cortes-Álvarez SI(1), Delgado-Enciso I(1)(2)(3), Hernández-Fuentes GA(1)(2)(4), 
Guzmán-Esquivel J(5), Diaz-Martinez J(3), Rodríguez-Hernández A(1), 
Martinez-Fierro ML(6), Rodríguez-Sánchez IP(7), Melnikov V(1), Flores-Ruelas 
Y(8), Garza-Veloz I(6), Cruz-Ruiz M(9), Ramos-Organillo ÁA(4), Sánchez-Ramírez 
CA(1).

Author information:
(1)Department of Molecular Medicine, School of Medicine, University of Colima, 
Colima 28040, Mexico.
(2)Colima State Institute of Cancerology, IMSS-Bienestar, Colima 28085, Mexico.
(3)Research Center in Minority Institutions, Robert Stempel College of Public 
Health, Florida International University, Miami, FL 33199, USA.
(4)Faculty of Chemical Sciences, University of Colima, Coquimatlan 28400, 
Mexico.
(5)Clinical Epidemiology Research Unit, Mexican Institute of Social Security, 
Villa de Alvarez, Colima 28984, Mexico.
(6)Molecular Medicine Laboratory, Academic Unit of Human Medicine and Health 
Sciences, Autonomous University of Zacatecas, Zacatecas 98160, Mexico.
(7)Molecular and Structural Physiology Laboratory, School of Biological 
Sciences, Universidad Autónoma de Nuevo León, San Nicolás de los Garza 66455, 
Mexico.
(8)Institute of Human Nutrition, University Center for Health Sciences, 
University of Guadalajara, Salvador Quevedo y Zubieta No. 750, Hospital Civil de 
Guadalajara, Annex Building, 3rd Floor, Independencia Neighborhood, Guadalajara 
44340, Mexico.
(9)Oficina de Investigación en Salud, Servicios de Salud del Instituto Mexicano 
del Seguro Social Para el Bienestar (IMSS-BIENESTAR), Colima 28085, Mexico.

Background/Objectives: Chronic low-grade inflammation and nutritional 
deficiencies, particularly vitamin D deficiency, have emerged as important 
contributors to Metabolic syndrome (MetS) pathogenesis but remain underexplored. 
This study aimed to comprehensively evaluate the associations between dietary 
intake, vitamin D status, and inflammatory biomarkers (high-sensitivity 
C-reactive protein -CRP- and ferritin) in patients with MetS. Methods: A 
cross-sectional observational study was conducted on 141 adult MetS patients at 
a Mexican hospital. Clinical, anthropometric, dietary (using a validated food 
frequency questionnaire), and biochemical data including serum 25-hydroxyvitamin 
D, CRP, ferritin, and neutrophil-to-lymphocyte ratio (NLR) were collected. 
Vitamin D deficiency was defined as serum 25(OH)D < 20 ng/mL, and high 
inflammation as CRP ≥ 3 mg/L. Logistic regression models adjusted for 
confounders were used to analyze associations. Mediation analysis assessed 
whether vitamin D deficiency mediated the link between dietary intake and high 
CRP or ferritin. Results: Patients with elevated CRP had significantly lower 
serum vitamin D levels (14.0 ± 5.1 vs. 22.1 ± 7.0 ng/mL; p < 0.001). 
Multivariable analysis showed vitamin D deficiency (adjusted OR 7.1; 95% CI 
2.5-19.4; p < 0.001) and hyperferritinemia (ferritin ≥ 200 μg/L; aOR 8.0, 95% CI 
3.5-18.2, p < 0.001) as predictors of high CRP. Conversely, hyperferritinemia 
was predicted by vitamin D deficiency (aOR 24.69; 95% CI 3.76-162.16; p = 
0.001), elevated CRP (aOR 5.06; p = 0.014), Hb (aOR 63.23; p < 0.001), and 
inversely by grade 2 obesity (aOR 0.11; 95% CI 0.02-0.60; p = 0.03), confirming 
bidirectional CRP-ferritin associations and hyperferritinemia as an inflammation 
marker rather than iron overload indicator. Although Hb > 14.3 g/dL associated 
with hyperferritinemia, it did not independently predict CRP in multivariate 
analyses. Frequent consumption of vitamin D-rich foods (milk, fish, Manchego and 
Oaxaca cheese) was associated with lower inflammation. Mediation analysis 
confirmed that vitamin D deficiency mediated dietary intake-CRP and dietary 
intake-ferritin links (Sobel test p < 0.05). Conclusions: Vitamin D deficiency 
is a key mediator linking inadequate dietary vitamin D intake to systemic 
inflammation in MetS. Nutritional strategies emphasizing vitamin D repletion and 
consumption of vitamin D fortified foods may effectively reduce chronic 
inflammation and improve metabolic outcomes.

DOI: 10.3390/nu18020224
PMCID: PMC12845497
PMID: 41599837 [Indexed for MEDLINE]

Conflict of interest statement: The authors declare no conflicts of interest.


51. Foods. 2026 Jan 15;15(2):317. doi: 10.3390/foods15020317.

Characterization of Milk Fat Globule Membrane Phospholipids in Colostrum of 
Holstein cows, Yaks and Buffaloes as Well as in Yak Colostrum and Mature Milk.

Luo J(1), Cao Y(1), Zhou H(1), Yan F(1), Wu S(2), Zhang H(3), Fan X(1).

Author information:
(1)College of Food Science and Technology, Hunan Agricultural University, 
Changsha 410128, China.
(2)Inner Mongolia Enterprise Key Laboratory of Dairy Nutrition, Health & Safety, 
Inner Mongolia Mengniu Dairy (Group) Co., Ltd., Hohhot 011500, China.
(3)College of Food Science and Nutritional Engineering, China Agricultural 
University, Beijing 100083, China.

Milk fat globule membrane (MFGM) phospholipids could promote the development of 
infants' brain, nervous system and digestive system. This research conducted a 
comparative analysis of phospholipid composition in MFGM of colostrum from 
different bovine species (Holstein cattle, yak, and Buffalo), with a particular 
focus on analyzing phospholipid variations in yak MFGM across different 
lactation stages. Chromatographic quantification revealed phosphatidylcholine 
(PC) as the predominant phospholipid class (34.7-47.44%) in all examined 
species. Notably, Holstein cow milk contains significantly higher levels of 
phosphatidylethanolamine (PE). Distinct phospholipid profiles emerged between 
species: yak milk demonstrated significantly higher concentrations of 
sphingomyelin (SM), lysophosphatidylethanolamine (LPE), 
dimethylphosphatidylethanolamine (dMePE), and bis-methylphosphatidic acid 
(BisMePA), whereas buffalo milk showed preferential accumulation of 
phosphatidylinositol (PI), phosphatidylserine (PS), phosphatidylglycerol (PG), 
and lysophosphatidylcholine (LPC). Longitudinal analysis revealed dynamic 
changes in yak milk phospholipids during lactation: as the lactation period 
in-creases, PC, PS, LPC, LPE, methylphosphatidylcholine (MePC), BisMePA, and 
dMePE exhibited progressive decline, while PE, SM, PI and PG showed incremental 
increases. Analysis of phospholipid metabolism pathways indicates that yak 
colostrum supports early calf development by enriching phospholipids associated 
with immune and neuroprotection, while mature milk shifts toward maintaining 
membrane stability. These compositional characteristics position yak milk as a 
promising phospholipid-fortified alternative to human breast milk.

DOI: 10.3390/foods15020317
PMCID: PMC12840161
PMID: 41596916

Conflict of interest statement: The authors declare no conflicts of interest.


52. Front Nutr. 2026 Jan 7;12:1669809. doi: 10.3389/fnut.2025.1669809. eCollection 
2025.

Long-term follow-up of a randomized controlled trial on individualized 
fortification of human milk.

Quan M(1), Zhang Y(1), Zhang L(1), Wang C(1), Li Z(1).

Author information:
(1)Department of Pediatrics, State Key Laboratory of Complex Severe and Rare 
Disease, Peking Union Medical College Hospital, Chinese Academy of Medical 
Science and Peking Union Medical College, Beijing, China.

BACKGROUND AND AIMS: To evaluate short- and long-term outcomes of preterm 
infants from a previous RCT on individualized fortification (IF).
METHODS: Preterm infants from the Peking Union Medical College Hospital (PUMCH) 
IF study were enrolled. Data before age 2 were collected from medical records. 
Long-term follow-up was conducted at ages 6-10 years. Growth and development 
outcomes as well as biochemical indexes were compared between IF and standard 
fortification (SF) groups.
RESULTS: Fifty-one infants completed 2-year follow-up. The IF group showed 
significantly better outcomes in head circumference z-scores at CA 6 months 
(p = 0.035), hemoglobin at CA 12 months (p = 0.044), and weight z-scores at CA 
24 months (p = 0.027). Twenty-five children completed long-term follow-up. No 
significant differences were found in lung function, cognitive scores, ferritin, 
hemoglobin, IGF, 25-OH-vitamin D, bone age, or metabolic parameters including 
BMI and lipid profile.
CONCLUSION: PUMCH IF improved early growth outcomes, but no significant 
long-term health differences were observed.

Copyright © 2026 Quan, Zhang, Zhang, Wang and Li.

DOI: 10.3389/fnut.2025.1669809
PMCID: PMC12819227
PMID: 41572973

Conflict of interest statement: The author(s) declared that this work was 
conducted in the absence of any commercial or financial relationships that could 
be construed as a potential conflict of interest.


53. Pediatr Res. 2026 Jan 21. doi: 10.1038/s41390-025-04680-z. Online ahead of 
print.

Effects of extensive protein hydrolysate in supporting intestinal barrier 
function in vitro.

Bozzetti V(1)(2), Brosnan R(3), Verma S(3), Tran T(3), Sadreyev R(4), Cetinbas 
M(4), Murguia-Peniche T(5)(6), Simmons R(7), Gross G(8), Fasano A(3)(5).

Author information:
(1)Mucosal Immunology and Biology Research Center, Massachusetts General 
Hospital for Children, Boston, MA, USA..
(2)Experimental Gastroenterology Lab, Division of Immunology, Transplantation 
and Infectious Disease, Department of Gastroenterology and Digestive Endoscopy, 
IRCSS San Raffaele Hospital, Milan, Italy..
(3)Mucosal Immunology and Biology Research Center, Massachusetts General 
Hospital for Children, Boston, MA, USA.
(4)Department of Molecular Biology, Massachusetts General Hospital and 
Department of Pathology, Massachusetts General Hospital and Harvard Medical 
School, Boston, MA, USA.
(5)R&D Medical Sciences, Mead Johnson Nutrition Institute/Reckitt, Evansville, 
IN, USA.
(6)School of Medicine, Indiana University, Evansville, IN, USA.
(7)R&D Nutrition Science Platforms, Mead Johnson Nutrition Institute/Reckitt, 
Slough, UK.
(8)R&D Nutrition Science Platforms, Mead Johnson Nutrition Institute/Reckitt, 
Nijmegen, The Netherlands.

Erratum in
    Pediatr Res. 2026 Feb 6. doi: 10.1038/s41390-026-04807-w.

BACKGROUND: Maternal breast milk is the optimal nutrition for preterm infants, 
supporting immune and gut maturation. When unavailable, alternatives include 
infant formulas or pasteurized donor milk, the latter conferring greater 
protection against feeding intolerance and necrotizing enterocolitis. This study 
examined the impact of cow's milk-derived extensively hydrolyzed proteins (eHP), 
widely used in formulas and human milk fortifiers, on intestinal barrier 
function and gene expression in fetal human intestinal organoid-derived 
monolayers, modeling premature epithelium.
METHODS: Monolayers were exposed to free amino acids (AA), intact cow milk 
proteins (WP), or eHP at different concentrations, followed by inflammatory 
cytokines or commensal bacteria, to mimic physiologic gut conditions. Barrier 
integrity, permeability, and viability were measured using FITC-Dextran 
diffusion and LDH release. RNA-seq assessed transcriptional changes.
RESULTS: eHP at low and high concentrations decreased epithelial permeability at 
baseline, AA showed no effect, and WP only at high concentrations. Under 
inflammatory conditions, eHP significantly reduced epithelial barrier 
permeability; both eHP and AA improved cell viability at low concentrations. 
Transcriptomic analysis revealed modulation of proliferation- and 
regulation-related pathways, including NOTCH and WNT signaling.
CONCLUSION: In this gut model, eHP enhanced barrier function under baseline and 
inflammatory conditions, supporting their role in nutrition, though further 
validation is needed.
IMPACT: Extensively hydrolyzed proteins (eHP) derived from cow's milk improve 
intestinal barrier function and cell viability in a fetal organoid-derived model 
of premature human intestine. This is the first study to assess the functional 
and transcriptomic impact of eHP on primary intestinal epithelial cells derived 
from fetal organoids, a highly relevant model for preterm gut physiology. It 
demonstrates that eHP-unlike intact proteins or free amino acids-can mitigate 
inflammation-induced permeability and modulate pathways involved in epithelial 
proliferation and repair (e.g., NOTCH, WNT, E2F, G2M checkpoint). These findings 
highlight the potential of eHP as a nutritional strategy to enhance gut 
integrity and limit inflammatory damage in preterm infants, who are at high risk 
of intestinal complications such as NEC. This study paves the way for further 
mechanistic and clinical research on how peptide-based formulas might support 
immature gut function and reduce extraintestinal risks.

© 2026. The Author(s).

DOI: 10.1038/s41390-025-04680-z
PMID: 41565842

Conflict of interest statement: Competing interests: G.G., T.M., and R.S. have 
all been employees of Reckitt/Mead Johnson Nutrition at the time of study 
execution. A.F. is actually employed by Mead Johnson Nutrition.


54. Adv Nutr. 2026 Apr;17(4):100585. doi: 10.1016/j.advnut.2025.100585. Epub 2026 
Jan 12.

Food and Drug Administration Expert Panel on Infant Formula "Operation Stork 
Speed" June 2025: Part 3, Marketing of Infant Formulas, Breastfeeding Support, 
Hypoallergenic Formulas, and Nutrition for Preterm Infants.

Abrams SA(1), Brenna JT(2), Clemens R(3), Cohran VC(4), Du N(5), Gilbaugh A(6), 
Goran MI(7), Guild A(8), Kerner JA Jr(9), Knudsen TB(10), Krishna S(11), 
Sentongo T(12).

Author information:
(1)Department of Pediatrics, Dell Medical School at the University of Texas, 
Austin, TX, United States. Electronic address:.
(2)Department of Pediatrics, Dell Medical School at the University of Texas, 
Austin, TX, United States.
(3)Department of Regulatory and Quality Sciences, USC Mann School of Pharmacy 
and Pharmaceutical Sciences, University of Southern California, Los Angeles, CA, 
United States.
(4)Department of Pediatrics, The Ann & Robert H. Lurie Children's Hospital of 
Chicago, Feinberg School of Medicine, Chicago, IL, United States.
(5)Center for Nutrition, Division of Gastroenterology, Hepatology, and 
Nutrition, Boston Children's Hospital, Boston, MA, United States.
(6)Clinical Nutrition, Stanford Medicine Children's Health, Palo Alto, CA, 
United States.
(7)Department of Pediatrics, Children's Hospital Los Angeles and Keck School of 
Medicine, University of Southern California, Los Angeles, CA, United States.
(8)Global Gastroschisis Foundation, United States.
(9)Division of Gastroenterology, Hepatology, and Nutrition and Division of 
Neonatology and Developmental Medicine, Stanford University Medical Center, Palo 
Alto, CA, United States.
(10)Department of Intelligent Systems Engineering, Indiana University, 
Bloomington, IN, United States.
(11)Division of Neonatology, New York-Presbyterian-Weill Cornell Medicine, New 
York, NY, United States.
(12)Department of Pediatrics, University of Chicago, Pritzker School of 
Medicine, Chicago, IL, United States.

The United States formula industry uses aggressive marketing strategies that 
exploit parental anxieties, undermine breastfeeding, and violate ethical 
international standards, all of which contribute to early formula introduction 
and disparities in breastfeeding. In addition, misleading or complex labeling 
contributes to caregiver and provider confusion regarding the content of 
formulas. A centralized, Food and Drug Administration-maintained database of 
infant formulas could support caregivers and healthcare professionals in 
countering misinformation and making evidence-based decisions. Human milk 
remains the reference standard for infant nutrition, with strong evidence 
supporting its role in reducing morbidity and mortality, especially in the 
neonatal intensive care unit (NICU). Despite this, disparities persist in 
breastfeeding rates, particularly among Black and Hispanic mothers. These are 
driven by structural barriers, including lack of paid parental leave and 
inadequate lactation support. Donor human milk (DHM) is the recommended 
alternative for high-risk infants in the NICU when the mother's own milk is 
unavailable. However, due to lack of federal funding and oversight, high cost 
and inconsistent insurance coverage, the use of DHM is limited, especially in 
safety-net hospitals. Specialized hypoallergenic and metabolic formulas are 
essential for managing medical conditions including cow's milk-protein allergy 
and inborn errors of metabolism. These are regulated as exempt formulas but must 
meet rigorous clinical standards. In the NICU, adequate early nutrition for 
preterm infants is crucial to reduce morbidity and mortality and improve 
long-term neurodevelopmental outcomes. Human milk fortifiers are composed of 
similar ingredients to infant formula and are used to improve the nutritional 
content of feedings given in the NICU and postdischarge. The use of live 
biotherapeutic products or probiotics as additives to preterm infants has been 
demonstrated in multiple studies to reduce the incidence of necrotizing 
enterocolitis, late-onset sepsis, and all-cause mortality.

Copyright © 2025 The Author(s). Published by Elsevier Inc. All rights reserved.

DOI: 10.1016/j.advnut.2025.100585
PMCID: PMC13100751
PMID: 41565224 [Indexed for MEDLINE]

Conflict of interest statement: Conflict of interest JTB has received honoraria 
or travel funds from the Global Organization for EPA and DHA, the global dairy 
platform, Danone, and research support from the National Cattlemen’s Beef 
Association. MIG has previously served as a scientific advisor for Begin Health 
and Bobbi. VCC has served on the Speaker’s Bureau for Nutricia and Abbott 
Nutrition. SAA is Editor-in-Chief of Advances in Nutrition and played no role in 
the journal’s evaluation of the manuscript. Other authors declare that they have 
no known competing financial interests or personal relationships that could have 
appeared to influence the work reported in this paper.


55. Acta Paediatr. 2026 May;115(5):1048-1062. doi: 10.1111/apa.70448. Epub 2026 Jan 
20.

Human Milk Practices in Swedish Neonatal Units: Results From a Nationwide 
Survey.

Blomqvist YT(1)(2), Lund AM(3), Ahlsson F(1)(2), Ericson J(4), Karlsson C(5), 
Lundström J(6), Sandström I(6), Gustafsson A(7).

Author information:
(1)Neonatal Intensive Care Unit, University Children's Hospital, Uppsala, 
Sweden.
(2)Department of Women's and Children's Health, Uppsala University, Uppsala, 
Sweden.
(3)Department of Clinical Sciences, Paediatrics, Skåne University Hospital, Lund 
University, Lund, Sweden.
(4)Department of Health and Welfare, Dalarna University, Falun, Sweden.
(5)Department of Clinical Microbiology and Infection Prevention and Control, 
Kronoberg Region, Central Hospital, Växjö, Sweden.
(6)Department of Neonatology, Sachsska Children and Youth Hospital, Stockholm, 
Sweden.
(7)Department of Women's and Children's Health, Karolinska Institutet, 
Stockholm, Sweden.

AIM: The aim of this study was to describe the routines for handling mother's 
own milk and donor human milk in Swedish neonatal care units related to national 
guidelines.
METHODS: A web-based survey of 24 of 38 neonatal units and 19 of 28 human milk 
banks in Sweden.
RESULTS: Despite national guidelines, breast milk handling varied widely. All 
units offered lactation support and free breast pumps during hospital stays; 
nine continued post-discharge. Use of mother's milk differed; some prioritised 
fresh, others mixed or used frozen milk by age. Donor milk was mainly given to 
infants < 34-35 weeks or post-surgery, though criteria varied. Some also 
provided it for hypoglycaemia, growth restriction or hypothermia treatment. 
Fortification routines varied in both target groups and preparation, ranging 
from bedside at each feed to once daily. Most milk banks tested donor milk 
before pasteurisation; one tested both before and after. Five units never 
declined donations, while others did due to storage limits, surplus supply or 
time constraints.
CONCLUSION: Substantial variation in practices indicates a need to clarify and 
update national guidelines and strengthen milk banking, with relevance both 
nationally and internationally.

© 2026 The Author(s). Acta Paediatrica published by John Wiley & Sons Ltd on 
behalf of Foundation Acta Paediatrica.

DOI: 10.1111/apa.70448
PMCID: PMC13063361
PMID: 41557602 [Indexed for MEDLINE]

Conflict of interest statement: The authors declare no conflicts of interest.


56. Pediatr Res. 2026 Jan 19. doi: 10.1038/s41390-026-04768-0. Online ahead of 
print.

Bovine colostrum as a human milk fortifier for very preterm infants with slow 
feeding advancement: a randomized controlled trial.

Jiang PP(#)(1)(2), Huang HY(#)(3), Zhao L(4), He Z(3), Li X(3), Yan X(5), Li 
Y(1)(6), Möller S(7)(8), Zachariassen G(2)(7)(8), Zhou P(9), Sangild 
PT(10)(11)(12).

Author information:
(1)Department of Veterinary and Animal Sciences, University of Copenhagen, 
Frederiksberg, Denmark.
(2)Hans Christian Andersen Children's Hospital, Odense University Hospital, 
Odense, Denmark.
(3)Department of Neonatology, Nanshan People's Hospital/Union Hospital, Huazhong 
University of Science and Technology, Shenzhen, China.
(4)Department of Neonatology, Shenzhen Baoan Women's and Children's Hospital, 
Shenzhen, China.
(5)Department of Neonatology, Shenzhen People's Hospital, Jinan University & 
Southern University of Science and Technology, Shenzhen, China.
(6)NBCD A/S, Søborg, Denmark.
(7)Department of Clinical Research, University of Southern Denmark, Odense, 
Denmark.
(8)Open Patient data Explorative Network, Odense University Hospital, Odense, 
Denmark.
(9)Department of Neonatology, Shenzhen Baoan Women's and Children's Hospital, 
Shenzhen, China..
(10)Department of Veterinary and Animal Sciences, University of Copenhagen, 
Frederiksberg, Denmark..
(11)Hans Christian Andersen Children's Hospital, Odense University Hospital, 
Odense, Denmark..
(12)Department of Neonatology, Rigshospitalet, Copenhagen University Hospital, 
Copenhagen, Denmark..
(#)Contributed equally

BACKGROUND: Fortification of human milk for very preterm infants (VPIs) with 
alternatives to conventional bovine milk-based fortifiers remains minimally 
studied. This trial tested whether fortification with protein-rich bovine 
colostrum (BC) improves feeding intolerance and clinical variables in VPIs 
receiving enteral nutrition with a relatively slow advancement.
METHODS: In this unblinded, two-centre, randomised, controlled trial (FortiColos 
CN), VPIs (gestational age, 26 + 0 to 31 + 6 weeks) were fed human milk 
fortified with BC (n = 74) or a conventional fortifier (CF, FM85, Nestlé, 
n = 72) for at least two weeks, starting when enteral feeding volume reached 
80-100 mL/kg body weight/d. Incidence of feeding intolerance, nutrition intake, 
body growth, morbidities and clinical biochemical parameters were compared 
between the two groups.
RESULTS: No statistically significant difference was found in the incidence of 
feeding intolerance or in most of the nutritional or body growth parameters 
(p > 0.05). All recorded morbidity incidences and haematological and blood 
biochemical parameters were also similar between groups. Amino acids (Phe, Pro, 
Ser, Tyr, Val) showed higher levels in the infants receiving BC.
CONCLUSIONS: BC appeared safe when used as a fortifier to human milk for VPIs 
with slow feeding advancement, but did not improve feeding tolerance or clinical 
variables.
IMPACT: Fortifying human milk with bovine colostrum (BC) in very preterm infants 
(VPIs) is safe but did not improve feeding tolerance, growth or clinical 
outcomes, compared with a conventional fortifier (CF), when used during slow 
enteral feeding advancement. This study adds to the limited clinical evidence on 
the use of BC as a human milk fortifier in VPIs receiving enteral feeding with 
different feeding protocols. The findings support the safety of BC as a human 
milk fortifier in VPIs but suggest limited short-term clinical benefits over 
currently used fortifiers.

© 2026. The Author(s).

DOI: 10.1038/s41390-026-04768-0
PMID: 41554861

Conflict of interest statement: Competing interests: This study is partially 
financially supported by Biofiber Damino A/S, a Danish company that donated the 
bovine colostrum used for the study. University of Copenhagen holds a patent 
(Eur. Patent no. EP2858653A1) on the use of bovine colostrum for preterm 
infants. Per Torp Sangild, listed as the sole inventor, has declined any share 
of potential revenue arising from the commercialisation of such a patent. Other 
authors have no conflicts of interest or financial relationships relevant to 
this article to disclose. Consent statement: The study was conducted according 
to the guidelines of the Declaration of Helsinki and approved by the local 
ethical committee. Informed consent was obtained from the parents of all 
participants involved in the study.


57. Arch Dis Child Fetal Neonatal Ed. 2026 Jan 19:fetalneonatal-2024-327677. doi: 
10.1136/archdischild-2024-327677. Online ahead of print.

Effect of different feeding approaches on growth, neonatal morbidities, 
mortality and neurodevelopmental outcome in preterm infants: a systematic review 
and network meta-analysis.

Hamouda N(1)(2), AboEL-Azm Y(3)(4), Elsamman K(3)(5), Nabil S(3)(6), Tarek 
M(3)(7), Wasia FE(3)(8), Amin AM(3)(7), Elshahat A(3)(7), Hassan N(3)(9), 
Mektebi A(3)(10), Khaled A(9), Shahin HN(11), Aldemerdash MA(3)(12), Refaey 
N(3)(13), Khalil M(3)(9), Hendi NI(3)(14), Hafez S(3)(15), Alabdallat YJ(3)(16), 
Abdellatif M(3)(17).

Author information:
(1)Negida Research Academy, Arlington, Virginia, USA 
.
(2)NICU, El Sahel Teaching Hospital, Cairo, Egypt.
(3)Negida Research Academy, Arlington, Virginia, USA.
(4)Zagazig University Faculty Medicine, Zagazig, Ash Sharqia Governorate, Egypt.
(5)Biochemistry and Nutrition, Faculty of Women for Science, Ain Shams 
University, Cairo, Egypt.
(6)Internal Medicine, Beni Suef University Hospital, Beni Suef, Beni Suef 
Governorate, Egypt.
(7)Al-Azhar University Faculty of Medicine, Nasr City, Cairo, Egypt.
(8)Faculty of Medicine Kafrelsheikh University, Kafr El-sheikh, Egypt.
(9)Zagazig University Faculty of Human Medicine, Zagazig, Egypt.
(10)Faculty of Medicine Kutahya Health Science University, Kutahya, Turkey.
(11)South Valley University Faculty of Medicine, Qena, Egypt.
(12)Sohag University Faculty of Medicine, Sohag, Egypt.
(13)Physical Therapy for Women's Health, Cairo University Faculty of Physical 
Therapy, Dokki, Giza, Egypt.
(14)Ain Shams University Faculty of Medicine, Cairo, Egypt.
(15)Minia University Faculty of Medicine, Minia, Egypt.
(16)The Hashemite University Faculty of Medicine, Zarqa, Zarqa, Jordan.
(17)Pediatrics and Neonatology, Muscat Private Hospital, Muscat, Muscat, Oman.

OBJECTIVE: To compare feeding strategies on preterm infants' growth during 
hospitalisation, neonatal morbidities, mortality and neurodevelopmental outcome 
(NDO) at 18-26 months corrected age.
METHODS: We followed the Preferred Reporting Items for Systematic Reviews and 
Meta-Analyses extension statement for network meta-analysis . We searched five 
medical databases for randomised controlled trials comparing different feeding 
approaches in preterm infants and their effects on growth, neonatal morbidities, 
mortality and NDO. The Cochrane Collaboration's tool was used to assess the risk 
of bias. We used a random-effects model. Pooled mean differences (MD) or risk 
ratios with 95% CIs were calculated.
RESULTS: Ninety-five studies (9663 infants) were included.Human milk (HM) with 
bovine milk fortifier (BMF) (adjusted according to blood urea nitrogen) achieved 
the best length increment (MD=0.56 cm/week; 95% CI 0.19 to 0.93). Notably, 
HM+BMF (3.5 gm/kg/d protein) showed the best head circumference growth (MD 
0.46 cm/week; 95% CI 0.10 to 0.81) but no significant difference in weight gain. 
There were no significant differences in neonatal morbidities/mortality. While 
MOM|+PTF (supp) displayed significantly lower NDO delay in the domain of mild 
cognitive delay.
CONCLUSION: Overall, there is a lack of strong evidence to support a specific 
enteral feeding strategy and further high-quality research is required. Targeted 
HM fortification appears to improve head growth, while adjusted fortification 
enhances length. Given the significant inconsistency detected, which may 
compromise the reliability of the network estimates, these results must be 
interpreted carefully.

© Author(s) (or their employer(s)) 2026. No commercial re-use. See rights and 
permissions. Published by BMJ Group.

DOI: 10.1136/archdischild-2024-327677
PMID: 41554628

Conflict of interest statement: Competing interests: None declared.


58. Pediatr Res. 2026 Jan 15. doi: 10.1038/s41390-025-04754-y. Online ahead of 
print.

Enhanced fortification of human milk to meet preterm infant nutritional targets.

Fu TT(1)(2), King CM(3), Kim JH(3)(4), Meinzen-Derr J(4)(5).

Author information:
(1)Division of Neonatology, Perinatal Institute, Cincinnati Children's Hospital 
Medical Center, Cincinnati, OH, USA..
(2)Department of Pediatrics, University of Cincinnati College of Medicine, 
Cincinnati, OH, USA..
(3)Division of Neonatology, Perinatal Institute, Cincinnati Children's Hospital 
Medical Center, Cincinnati, OH, USA.
(4)Department of Pediatrics, University of Cincinnati College of Medicine, 
Cincinnati, OH, USA.
(5)Division of Biostatistics and Epidemiology, Cincinnati Children's Hospital 
Medical Center, Cincinnati, OH, USA.

BACKGROUND: Many preterm infants need nutrition beyond 24 kcal/oz (standard 
fortification) to support growth. Individualized targeted fortification improves 
growth but is labor-intensive. Universal enhanced fortification to presumed 
26 kcal/oz for very low birth weight infants was clinically implemented. We 
determined how often donor breast milk (DBM) and preterm maternal breast milk 
(MBM) met nutritional targets with enhanced fortification.
METHODS: MBM/DBM samples were collected for a prospective cohort study of 
infants born <1500 g and <33 weeks. Macronutrients were measured using a 
mid-infrared analyzer. The frequency at which MBM/DBM samples met intake goals 
(4.8-8.1 g/kg/day fat, 11.6-13.2 g/kg/day carbohydrate, 3.5-4.5 g/kg/day 
protein) with enhanced and standard fortification was compared.
RESULTS: Among 198 MBM samples and 168 DBM samples, MBM had higher protein, fat, 
and energy (p < 0.0001). Regardless of fortification method, MBM samples met 
lower and mid-range fat goals more often (p < 0.01 and p = 0.03, respectively). 
Collectively, more samples achieved protein targets with enhanced fortification: 
all samples reached 3.5 and 4 g/kg/day, 56% (147 MBM, 58 DBM) attained 
4.5 g/kg/day. With standard fortification, 11% achieved 4 g/kg/day protein; none 
attained 4.5 g/kg/day.
CONCLUSIONS: Enhanced fortification is an efficient method that meets enteral 
nutrition goals for preterm infants and delivers desired protein intake more 
consistently.
IMPACT: Enhanced fortification is a feasible and efficient alternative to 
targeted fortification that can also achieve enteral nutrition goals for preterm 
infants. Enhanced fortification delivers desired protein intake more 
consistently than standard fortification, especially with donor breast milk. 
Regardless of fortification strategy, maternal breast milk is more likely to 
reach fat intake goals, although additional enrichment may be needed, depending 
on the fortifier product.

© 2026. The Author(s).

DOI: 10.1038/s41390-025-04754-y
PMID: 41540198

Conflict of interest statement: Competing interests: The authors declare no 
direct conflicts but do share the following financial disclosures. T.T.F. has a 
speaking agreement with Medela. J.H.K. is a consultant for Medela, Mother’s Milk 
is Best, Infant Biotherapeutics, Biomilq, has a sponsored research grant with 
Fresenius Kabi, and is a shareholder for Astarte Medical and Nicolette. He is an 
unpaid Scientific Advisor to the NEC Society. The other authors have no 
financial disclosures. Research was conducted independently of these 
relationships. Consent to participate: Parental consent was obtained for the 
participation of both the lactating parent and infant.


59. Food Res Int. 2026 Feb 28;226:118195. doi: 10.1016/j.foodres.2025.118195. Epub 
2025 Dec 22.

Ohmic heating-assisted extraction of ginger bioactives: Synergistic impacts on 
extract composition and on the quality of high-protein milk beverages.

Vieira GP(1), Martins CPC(1), Pereira TRC(1), Cipriano LC(1), Goulart MB(1), 
Guimarães JT(1), Pagani MM(2), Duarte MCKH(1), Cruz AG(3), Mano SB(1), Mársico 
ET(1), Esmerino EA(1).

Author information:
(1)Federal Fluminense University (UFF), Faculty of Veterinary, 2430-340, 
Niterói, Rio de Janeiro, Brazil.
(2)Federal Rural University of Rio de Janeiro (UFRRJ), Institute of Technology, 
23890-000, Seropédica, Rio de Janeiro, Brazil.
(3)Federal Institute of Education, Science and Technology of Rio de Janeiro 
(IFRJ), Food Department, 20270-021, Rio de Janeiro, Rio de Janeiro, Brazil.

Leveraging the potential of ohmic heating (OH) for phytochemical extraction, 
this study evaluated how different electric field strengths influence the 
extraction of ginger bioactives and the subsequent quality of a lemon-flavored 
high-protein milk beverage. OH markedly improved thermal efficiency. at 140 V 
(25.45 V/cm) the extract reached 80 °C with an 87.7% reduction in heating time 
compared with the conventional method. Extracts obtained at 100 V (18.18 V/cm) 
and 140 V exhibited ∼44-45% DPPH radical-scavenging activity, outperforming the 
control and showing activity comparable to the conventionally heated extract. 
Although total phenolic content did not increase under OH, beverages fortified 
with OH extracts displayed superior antioxidant stability during storage, 
maintaining higher activity than other treatments from day 7 to day 21. OH also 
produced a distinct volatile profile compared with conventional heating, 
suggesting enhanced release of aroma-active compounds. When incorporated into 
the dairy matrix, OH extracts significantly affected beverage rheology, 
increasing the consistency index (up to 0.10 Pa·sⁿ) and apparent viscosity (up 
to 45 mPa·s) compared with beverages containing conventional or control extracts 
(29-31 mPa·s). The dynamic sensory profile revealed that OH beverages were more 
frequently associated with "lemon flavor," "sweet taste," and "creamy/viscous 
texture," consistent with instrumental measurements. Consumer acceptance 
remained high for all OH formulations and was accompanied by more positive 
emotional responses - particularly "happy/satisfied" and "relaxed/calm" -whereas 
the conventionally heated extract elicited less favorable associations. Overall, 
the findings demonstrate that OH, particularly at higher electric field 
strengths, is an efficient and advantageous technique for extracting ginger 
bioactives and enhancing the physicochemical, functional, and sensory quality of 
high-protein milk beverages. The technology shows strong potential for the 
development of novel functional products with improved stability and consumer 
appeal.

Copyright © 2025 Elsevier Ltd. All rights reserved.

DOI: 10.1016/j.foodres.2025.118195
PMID: 41539814 [Indexed for MEDLINE]

Conflict of interest statement: Declaration of competing interest The authors 
declare that they have no known competing financial interests or personal 
relationships that could have appeared to influence the work reported in this 
paper.


60. Indian Pediatr. 2026 Jan 13. doi: 10.1007/s13312-026-00266-y. Online ahead of 
print.

Amino Acid Levels of Preterm Infants on Parenteral and Enteral Nutrition: A 
Cross-Sectional Study.

Varikkottil MM(1), Bhat ZA(1), Pournami F(2), Prabhakar J(1), Prithvi AK(1), 
Kolisambeevi AA(1), Jain N(1).

Author information:
(1)Department of Neonatology, KIMS Health, Trivandrum, Kerala, India.
(2)Department of Neonatology, KIMS Health, Trivandrum, Kerala, India. 
.

OBJECTIVE: To assess the serum amino acid (AA) levels among preterm infants 
receiving parenteral and enteral nutrition.
METHODS: This cross-sectional study included very preterm neonates (≤ 32 weeks) 
and healthy term breastfed neonates as reference group between December 2022 and 
June 2024. Parenteral nutrition (PN) was commenced on day 1 of life for preterm 
neonates delivered at ≤ 30 weeks gestation. Neonates delivered at 31-32 weeks 
gestation did not receive PN. Standard feed regimens were followed for enteral 
nutrition with "early" commencement of human milk fortifier (HMF). AA levels 
were estimated by liquid chromatography tandem mass spectrometry after at least 
one week of full enteral nutrition for 31-32 weeks group, or parenteral followed 
by full enteral nutrition for ≤ 30 weeks group. AA levels in preterm infants 
were considered inadequate if found to be below the lowest level of 
corresponding AA levels in healthy term breastfed babies.
RESULTS: A total of 84 preterm neonates delivered at ≤ 30 weeks gestation, 56 
preemies at 31-32 weeks gestation, and 100 term neonates were studied. 
Compared to term neonates, preterm infants ≤ 30 weeks on PN and EN had 
inadequate levels of phenylalanine, methionine, histidine, valine, glycine, 
glutamine, arginine, alanine, glutamic acid, and aspartic acid. Deficiencies 
were noted in > 10% of this group for phenylalanine, methionine, histidine and 
glycine; in 5-10% for valine, leucine/isoleucine, glutamine, alanine, glutamic 
acid; and in < 5% for tyrosine, arginine, serine, and aspartic acid.
CONCLUSIONS: Despite use of parenteral and enteral nutrition protocols based on 
current science, preterm (≤ 30 weeks) infants had amino acid levels 
significantly lesser than in term healthy breastfed neonates.

© 2026. The Author(s), under exclusive licence to Indian Academy of Pediatrics.

DOI: 10.1007/s13312-026-00266-y
PMID: 41528685

Conflict of interest statement: Conflict of interest: None. Ethics Approval: 
Institute Human Ethics Committee, KIMS Health, Trivandrum, Ref. No. 
KIMS/ACAD/TP-035/2022 dated December 05, 2022. Trial Registry: Clinical Trials 
Registry of India (CTRI/2023/06/054310).


61. Pediatrics. 2026 Feb 1;157(2):e2025073625. doi: 10.1542/peds.2025-073625.

Promoting Human Milk and Breastfeeding for the Very Low Birth Weight Infant: 
Clinical Report.

Parker MG(1), Stellwagen L(2), Miller ER(3), Noble L(4)(5), Corkins MR(6), Hudak 
ML(7); Committee on Fetus and Newborn; Section on Breastfeeding; Committee on 
Nutrition.

Collaborators: Eichenwald E, Ambalavanan N, Guillory C, Hudak ML, Kaufman D, 
Martin C, Lucke A, Parker MG, Pramanik A, Wade K, Bunik M, Awelewa T, Hannon E, 
Long S, Vaidyanathan L, Williams T, Corkins MR, Blanco CL, Larson-Nath C, 
Peebles R, Peterson ALH, Suchdev P, Baidal JW.

Author information:
(1)Department of Pediatrics, UMass Chan School of Medicine, Worcester, 
Massachusetts.
(2)Department of Pediatrics, University of California San Diego School of 
Medicine, La Jolla, California.
(3)Division of Neonatology, Department of Pediatrics, Cincinnati Children's 
Hospital Medical Center, University of Cincinnati, Cincinnati, Ohio.
(4)Department of Pediatrics, Icahn School of Medicine at Mount Sinai, New York 
City, New York.
(5)New York City Health+Hospitals/Elmhurst, Elmhurst, New York.
(6)Division of Pediatric Gastroenterology, Hepatology, and Nutrition, Department 
of Pediatrics, The University of Tennessee Health Science Center, Memphis, 
Tennessee.
(7)Department of Pediatrics, University of Florida College of Medicine - 
Jacksonville, Florida.

Mother's own milk (MOM), appropriately fortified, provides optimal nutrition for 
hospitalized very low birth weight (VLBW [≤1500 g]) infants in the neonatal 
intensive care unit (NICU) and confers a myriad of other short- and long-term 
health benefits. Every family of a VLBW infant should receive information about 
the critical importance of MOM to the health of their infants and receive 
intensive support throughout the full course of their infant's hospitalization 
to achieve their personal lactation goals. Provision of pasteurized donor human 
milk (PDHM) is recommended when MOM is not available, insufficient in volume, or 
is contraindicated. PDHM should optimally be continued until the infant's risk 
to develop necrotizing enterocolitis is low (approximately 34-36 weeks' 
postmenstrual age). Preterm infant formula is recommended when MOM is not 
available and PDHM is either not available or the family declines use. Promotion 
of human milk and breastfeeding for VLBW infants requires access to efficient, 
effective, and comfortable double electric breast pumps in the hospital and at 
home and multidisciplinary and system-wide adoption of lactation support 
practices, including early and frequent milk expression, skin-to-skin care 
(SSC), and direct breastfeeding. Addressing social inequities in provision of 
human milk at the local level requires access to PDHM, peer lactation support, 
maximizing use of interpreter services as appropriate, and implementation of 
standardized approaches to identify and address unmet basic needs.

Copyright © 2026 by the American Academy of Pediatrics.

DOI: 10.1542/peds.2025-073625
PMID: 41520943 [Indexed for MEDLINE]


62. J Food Sci. 2026 Jan;91(1):e70805. doi: 10.1111/1750-3841.70805.

Micronized Dispersible Ferric Pyrophosphate (MDFP) in Extruded Rice Desserts: 
High Iron Bioavailability and Good Sensory Acceptance by Children.

Ferreira DCM(1), de Oliveira TV(2), Moreira MEC(2), Pirozi MR(1), Martino 
HSD(3), de Oliveira EB(1).

Author information:
(1)Department of Food Technology, Federal University of Viçosa, Viçosa, MG, 
Brazil.
(2)Faculty of Chemical Engineering, Federal University of Uberlândia, Patos de 
Minas, MG, Brazil.
(3)Department of Nutrition and Health, Federal University of Viçosa, Viçosa, MG, 
Brazil.

Iron deficiency in children is a persistent global public health problem. This 
study evaluated the efficacy of an extruded rice matrix as a carrier for 
micronized dispersible ferric pyrophosphate (MDFP), aiming to increase iron 
bioavailability in traditional rice-based desserts formulated with cow's milk 
(CM) or soy soluble protein extract (SSPE). Four experimental diets were 
administered to the rats: (i) fortified with CM and MDFP incorporated into 
extruded rice; (ii) fortified with SSPE and MDFP incorporated into extruded 
rice; (iii) control with ferrous sulfate (AIN-93G); and (iv) a control with pure 
MDFP (AIN-93G). Both fortified groups showed significantly higher recovery of 
iron nutritional status (p < 0.05), demonstrating that the different liquid 
ingredients did not compromise iron bioavailability. Gene expression analyses of 
key iron metabolism proteins (DMT-1, DcytB, ferroportin, hephaestin, 
transferrin, and ferritin) confirmed that the extruded rice matrix increased the 
bioavailability of MDFP compared to the compound in its pure form (p < 0.05), 
possibly reducing aggregation and favoring direct interaction with intestinal 
absorption sites. Furthermore, the fortified formulations demonstrated 
satisfactory sensory acceptance among school-ageRd children. These results 
indicate that incorporating MDFP into an extruded rice is an effective and 
technologically viable strategy for developing fortified rice desserts, with 
potential application in programs to combat iron deficiency anemia, specially in 
schoo-laged children at poor regions worldwide. PRACTICAL APPLICATIONS: Rice 
desserts fortified with micronized ferric pyrophosphate (MDFP) in an extruded 
rice matrix effectively improved iron availability without diminish sensory 
acceptance by children. Hence, the technology here reported offers a practical, 
low-cost strategy for school feeding programs as well as for food industry to 
develop fortified products to fight against iron deficiency anemia.

© 2026 The Author(s). Journal of Food Science published by Wiley Periodicals LLC 
on behalf of Institute of Food Technologists.

DOI: 10.1111/1750-3841.70805
PMCID: PMC12790682
PMID: 41520239 [Indexed for MEDLINE]

Conflict of interest statement: The authors declare no conflicts of interest.


63. Nutrients. 2025 Dec 27;18(1):95. doi: 10.3390/nu18010095.

Identifying Predictors for the Acquisition of Tolerance to Cow's Milk Protein in 
Infants with Food Protein-Induced Allergic Proctocolitis (FPIAP): Multifactorial 
Analysis of Two Italian Cohorts.

Scavella A(1), Ferrigno C(2), Baù M(1), Colombo A(2), Tavernelli CI(1), Zobele 
M(2), Borgetto R(1), Maggi A(2), Baronti A(1), Francone A(1), Zuccotti GV(2)(3), 
Agosti M(4), D'Auria E(2)(3), Salvatore S(5).

Author information:
(1)Post Graduate School of Pediatrics, University of Insubria, 21100 Varese, 
Italy.
(2)Department of Pediatrics, Vittore Buzzi Children's Hospital, 20154 Milan, 
Italy.
(3)Department of Biomedical and Clinical Sciences, University of Milan, 20157 
Milan, Italy.
(4)Department of Medicine and Surgery, University of Insubria, 21100 Varese, 
Italy.
(5)Pediatric Unit, Department of Medicine and Technological Innovation, Hospital 
"F. Del Ponte", University of Insubria, 21100 Varese, Italy.

Background/Objectives: Food protein-induced allergic proctocolitis (FPIAP) is a 
non-IgE-mediated gastrointestinal food allergy. Although tolerance to the 
culprit food is usually achieved within the first year of life, late acquisition 
occurs and remains poorly predictable. This study aimed to analyze clinical 
characteristics and explore factors that may potentially function as predictors 
of late tolerance acquisition to cow's milk (CM). Methods: We conducted a 
cross-sectional study at two Italian pediatric clinics (2020-2024), including 
infants diagnosed with FPIAP. Clinical, dietary, and immunological variables; 
onset and duration of rectal bleeding (visible blood in the stools); and time to 
CM tolerance were analyzed. Late tolerance was defined as acquisition after 19 
months according to the distribution of tolerance achievement in our population. 
Statistical analyses included χ2, Mann-Whitney U, Spearman's correlation, and 
logistic regression. Results: Ninety-four infants were included (median age at 
onset 2.9 months [IQR 1.9-4.7]); 58 (62%) were exclusively breastfed and 18 
(19%) were born preterm (<37 completed weeks of gestation). CM was the culprit 
food in all cases; tolerance was achieved in all infants at a median age of 12 
months. Family history of atopy and atopic dermatitis were reported in 44% and 
19% of infants, respectively. Late CM tolerance was associated with preterm 
birth, fortification of human milk, early antibiotic exposure, growth faltering, 
and recurrent infections. Logistic regression identified family history of atopy 
(OR 5.4 [95% CI 1.2-25.4]; p = 0.031), atopic dermatitis (OR 8.2 [1.7-40.7]; p = 
0.010), rectal bleeding >18 days before elimination diet (OR 5.9 [1.3-27.7]; p = 
0.023), and IgE sensitization (OR 6.4 [1.2-35.0]; p = 0.034) as factors that may 
potentially function as predictors of late tolerance acquisition to CM. 
Conclusions: Identification of factors that may potentially function as 
predictors of late tolerance acquisition to CM in infants with FPIAP may help 
providing a personalized clinical management for these patients.

DOI: 10.3390/nu18010095
PMCID: PMC12787599
PMID: 41515212 [Indexed for MEDLINE]

Conflict of interest statement: The authors declare no conflicts of interest.


64. Indian J Pediatr. 2026 Feb;93(2):196. doi: 10.1007/s12098-025-05938-4. Epub 2026 
Jan 10.

Effect of Human Milk-Based Fortifier on Weight Gain and Morbidity Among Preterm 
Low Birth Weight Infants.

Chhina AS(1), Hiremath N(2).

Author information:
(1)Department of Neonatology, Cloudnine Hospital, Old Airport Road, Bangalore, 
560017, Karnataka, India..
(2)Department of Neonatology, Cloudnine Hospital, Old Airport Road, Bangalore, 
560017, Karnataka, India.

DOI: 10.1007/s12098-025-05938-4
PMID: 41513927

Conflict of interest statement: Declarations. Conflict of Interest: None.


65. J Dairy Res. 2026 Feb;93(1):1-20. doi: 10.1017/S002202992510174X.

A review on the holistic exploration of health benefits and technological 
advancements in yoghurt incorporating fruit peels.

Kariyawasam KP(1)(2), Somaratne G(2)(3), Senaratne S(2), Jayasekara M(3), Roy 
D(1), Walallawita U(4).

Author information:
(1)Riddet Institute, Massey University, Palmerston North, New Zealand.
(2)Postgraduate Institute of Agriculture, University of Peradeniya, Peradeniya, 
Sri Lanka.
(3)Department of Food Science and Technology, Faculty of Agriculture, University 
of Peradeniya, Peradeniya, Sri Lanka.
(4)Research Operations, Massey University, Palmerston North, New Zealand.

The trend towards health-conscious consumption has led the food industry to 
enhance the nutritional content of staple products like yoghurt. This review 
explores the integration of nutrient-rich fruit peels into yoghurt, emphasizing 
both health benefits and technological advancements. Fruit peels, which are 
frequently discarded as agro-industrial waste, are abundant in nutrients, 
antioxidants and dietary fibres, which can boost yoghurt's nutritional profile. 
Incorporating fruit peels not only supports waste reduction and sustainability 
but also contributes to the added value of yoghurt products. Technological 
innovations have made it possible to efficiently process and incorporate these 
peels while preserving their benefits. The review evaluates various methods such 
as fermentation, drying and grinding and their effects on yoghurt's taste, 
texture and shelf-life. Additionally, it considers consumer acceptance and 
market potential for these enhanced yoghurts. Overall, this approach highlights 
the promise of fruit peels as functional ingredients, promoting health benefits 
and advancing yoghurt production technologies.

DOI: 10.1017/S002202992510174X
PMID: 41502303 [Indexed for MEDLINE]


66. Crit Rev Food Sci Nutr. 2026 Jan 5:1-20. doi: 10.1080/10408398.2025.2609753. 
Online ahead of print.

Nature-inspired oligosaccharides: plant and algal routes to human milk 
functionality.

Cheong KL(1), Chen W(1), Li J(1), Zhong HJ(2), Wang D(3), Zhong S(1).

Author information:
(1)College of Food Science and Technology, Guangdong Ocean University, Guangdong 
Provincial Key Laboratory of Aquatic Product Processing and Safety, Guangdong 
Province Engineering Laboratory for Marine Biological Products, Guangdong 
Provincial Engineering Technology Research Center of Seafood, Guangdong 
Provincial Engineering Technology Research Center of Prefabricated Seafood 
Processing and Quality Control, Zhanjiang, China.
(2)State Key Laboratory of Bioactive Molecules and Druggability Assessment, 
International Cooperative Laboratory of Traditional Chinese Medicine 
Modernization and Innovative Drug Development of Chinese Ministry of Education 
(MOE) of China, School of Pharmacy, Jinan University, Guangzhou, China.
(3)Department of Obstetrics and Gynecology, The First Affiliated Hospital of 
Jinan University, Guangzhou, China.

Human milk oligosaccharides (HMO) shape early-life health by guiding microbiome 
assembly, fortifying the mucosal barrier, calibrating immune responses by 
enhancing secretory IgA at mucosal surfaces, blocking pathogen adhesion, and 
influencing the microbiota-gut-brain axis. Direct deployment of structurally 
diverse HMO at population scale remains constrained by manufacturing complexity 
and cost. This review examines "nature-inspired" oligosaccharides derived from 
plant and marine algal as affordable, scalable HMO mimetics, mapping chemistry 
to biology across five functional domains: microbiota architecture & metabolism, 
inhibition of pathogen adhesion via glycan decoy mechanisms, immune education, 
mucosal barrier enhancement and gut-brain regulation. We synthesize and 
critically appraise evidence on plant-derived fructo-, xylo-, and 
mannan-oligosaccharides, as well as on red-algal agaro-oligosaccharides and 
brown-algal fuco-oligosaccharides. Although these glycans do not fully replicate 
individual HMO structures, they can exhibit convergent mechanisms: selective 
fueling of saccharolytic consortia to produce functional metabolites (SCFAs), 
multivalent fucose-rich decoying of viral/bacterial adhesins, dendritic-cell/T- 
and B-cell tuning, epithelial junction and mucus programs, and 
neuroactive/metabolite signaling along the gut-brain axis. Framed as functional 
mimicry rather than strict structural emulation, plant and marine 
oligosaccharides emerge as promising "glycobiotics" to deliver HMO-like benefits 
across the lifespan, with opportunities for precision, personalized, and 
sustainable formulations guided by multi-omics and glycoinformatics.

DOI: 10.1080/10408398.2025.2609753
PMID: 41489355


67. Dietary Total Fat Intake and Dietary Polyunsaturated Fatty Acid Intake and Child 
Growth and Development Outcomes: Systematic Review [Internet].

Jungbauer RM, Stadler DD, Hsu F, Gillingham M, Yu Y, Fu R, Alex Foster B, Wood 
EK, Mackiewicz-Seghete K, O'Reilly CD, Totten AM.

Rockville (MD): Agency for Healthcare Research and Quality (US); 2026 Jan. 
Report No.: 26-EHC004.

BACKGROUND: Dietary fats are an important source of energy and essential fatty 
acids (linoleic acid and alpha-linolenic acid), and serve as a medium for 
fat-soluble vitamins (vitamins A, D, E, and K) absorption, transport, function, 
and storage. As part of a nutritionally complete and balanced diet, dietary fats 
support optimal fetal growth and development during pregnancy, and physical 
growth and cognitive development throughout infancy and childhood. Based on 
studies demonstrating reduced rates of preterm birth (<37 weeks of gestational 
age, GA) and early preterm birth (<34 weeks GA) among individuals consuming 
higher amounts of docosahexaenoic acid (DHA), an omega-3 polyunsaturated fatty 
acid (PUFA), during pregnancy, DHA was added to some prenatal vitamin 
supplements. Similarly, in 2002, voluntary fortification of infant formulas with 
DHA in amounts similar to those in mature human milk (0.2 to 0.5% by weight of 
total fatty acids) and arachidonic acid (ARA), also an omega-3 PUFA, at or above 
the amount of DHA, was approved by the U.S. Food and Drug Administration to 
support infant brain development and visual acuity. To inform updates to and 
expansion of the 2005 U.S.-Canadian Dietary Reference Intakes (DRIs), this 
systematic review was designed to identify and summarize the evidence published 
since 1990 on the relationships between consumption of omega-6 and/or omega-3 
(n-6 and n-3, respectively) PUFAs during pregnancy and risk of preterm birth 
(Key Question 1); between intake during pregnancy and/or lactation and infant 
and child growth and development (Key Question 2); and between intake of total 
fat (Key Question 3) or n-6 and/or n-3 PUFA (Key Question 4) by children between 
birth through 18 years of age and growth and developmental outcomes.
METHODS: A medical research librarian designed and conducted a search of Ovid 
MEDLINE®, CINAHL®, and Cochrane CENTRAL from 1990 through August 2025. 
Researchers manually reviewed reference lists and responses to a Federal 
Register Notice request for information. Dual, independent review of abstracts 
and full-text articles was conducted using a priori specified inclusion and 
exclusion criteria. Established methods were used in dual, independent 
assessment of risk of bias. Data were synthesized quantitatively using 
meta-analyses when possible; randomized controlled trials (RCTs) and 
nonrandomized studies (NRS) not included in the meta-analyses were summarized 
qualitatively in tables and text. For each set of analyses with sufficient data, 
we assessed the strength of evidence (SOE) using set criteria and applicability 
of findings to a generally healthy population from the United States and Canada.
RESULTS: Searches identified 8,929 abstracts. Eighty-six studies in 126 
publications were included across the four Key Questions. Of all the 
publications, 108 described RCTs and 18 described NRS; over half were rated high 
risk of bias (k=66, 52%), followed by moderate risk of bias (k=39, 31%) and low 
risk of bias ratings (k=21, 17%). The populations studied were generally 
healthy, with no medical conditions impacting fat consumption, digestion, 
absorption, or metabolism, or unusual dietary patterns. In the 86 parent or 
standalone studies, the majority (k=61, 71%) assessed the effects of dietary 
supplementation on outcomes, followed by diet only (k=22, 26%), and then a 
combination of diet and dietary supplementation (k=3, 4%). The bulk of the 
evidence centered on supplement studies during pregnancy or lactation and growth 
outcomes measured from birth to 6 months of age. The duration of interventional 
exposure was generally less than 6 months, and only 4 studies followed offspring 
into early adolescence (up to 13 years). Overall, supplemental PUFA or total fat 
intake in healthy populations resulted in similar risk of preterm birth and 
growth and developmental outcomes when compared with differing amounts or no 
additional amount of PUFA and total fat according to feeding guidelines. There 
was low SOE of no difference for PUFA supplementation above general dietary 
consumption and risk of preterm birth <34 weeks or <37 weeks, growth outcomes 
(weight, length [height], head circumference), or developmental outcomes 
(cognition, movement, language, social emotional). The exception was studies 
evaluating n-3 PUFA and n-3+n-6 PUFA supplementation, with consumption of only 
n-3 PUFA during pregnancy associated with lower risk of preterm birth <37 weeks, 
and higher weight and length at birth compared with consumption of n-3+n-6 PUFA 
supplements. There was insufficient or no evidence linking most growth and 
developmental outcomes to total fat intake according to infant and child feeding 
guidelines among infants consuming adequate energy.
CONCLUSIONS: Supplementation of PUFA and higher total fat intake in healthy 
populations, at the amounts studied, may not confer demonstrable advantages over 
baseline intake or exposure, and the consistent finding of no difference 
suggests no harm. There were few findings of differences across PUFA 
supplementation or dietary intake of fatty acids and growth and developmental 
outcomes in a healthy population during pregnancy and childhood. The largest 
body of evidence consisted of RCTs with moderate or high risk of bias evaluating 
PUFA supplementation during pregnancy or infancy and preterm birth and growth 
outcomes in the first year of life. Of outcomes with sufficient evidence, 
heterogeneity in study design, exposures and comparators, and outcomes resulted 
in the SOE being rated mostly as low. Additional research in older children and 
adolescents, studies enrolling people during pregnancy with low DHA 
concentrations at baseline, studies with earlier initiation during pregnancy and 
longer exposure durations, and populations who inherently consume diets low in 
PUFA or who have nutrition-related conditions that may benefit from supplemental 
PUFA or higher total fat consumption is needed to inform future DRIs.

DOI: 10.23970/AHRQEPCSR_DIETARYFATCHILD
PMID: 42013221


68. Food Chem. 2026 Feb 28;503:147786. doi: 10.1016/j.foodchem.2025.147786. Epub 
2025 Dec 27.

Improving fermented milk antioxidant properties by functional enrichment with 
Vitis Vinifera leaf extract cv Monica loaded in nanovesicles.

Dessì D(1), Casula L(1), Fais G(2), Pittiu A(1), Schlich M(1), Sinico C(1), Lai 
F(3), Sarais G(4).

Author information:
(1)Department of Life and Environmental Sciences, University of Cagliari, 
Cagliari, Italy.
(2)Interdepartmental Center of Environmental Science and Engineering (CINSA), 
University of Cagliari, Via San Giorgio 12, 09124 Cagliari, Italy; Department of 
Mechanical, Chemical and Materials Engineering, University of Cagliari, Piazza 
d'Armi, 09123 Cagliari, Italy.
(3)Department of Life and Environmental Sciences, University of Cagliari, 
Cagliari, Italy. Electronic address:.
(4)Department of Life and Environmental Sciences, University of Cagliari, 
Cagliari, Italy. Electronic address:.

Kefir is increasingly appreciated for its probiotic-rich composition and health 
benefits, aligning with the growing demand for functional foods. This study 
introduces an innovative approach to fortify kefir with grapevine leaves 
extract, an underutilized viticultural by-product, thereby promoting circular 
economy practices. The extract, characterized by a high total polyphenol content 
and remarkable antioxidant capacity, was analysed by HPLC-DAD, which identified 
caftaric acid, quercetin-3-O-glucuronide, and peonidin-3-O-glucoside as major 
phenolic constituents. The extract was encapsulated in nanovesicles prepared via 
a gentle high-shear homogenization method, a milder alternative to sonication 
that preserves thermosensitive compounds. The resulting nanovesicles exhibited 
encapsulation efficiencies ranging from 28.8 % to 73.9 % and demonstrated good 
physical stability. Moreover, they effectively mitigated the degradation of the 
extract components in simulated gastrointestinal conditions and protected 
enterocytes from oxidative stress in vitro. Incorporation of the loaded 
nanovesicles into the fermented drink enhanced its antioxidant activity in a 
dose-dependent manner without altering rheological properties.

Copyright © 2026 The Authors. Published by Elsevier Ltd.. All rights reserved.

DOI: 10.1016/j.foodchem.2025.147786
PMID: 41475261 [Indexed for MEDLINE]

Conflict of interest statement: Declaration of competing interest The authors 
declare that they have no known competing financial interests or personal 
relationships that could have appeared to influence the work reported in this 
paper.


69. Nutrients. 2025 Dec 12;17(24):3896. doi: 10.3390/nu17243896.

Associations of Lactoferrin-Fortified Formula with Infant Growth and Gut 
Microbiota: A Real-World Observational Study.

Shi X(1), Liu B(2), Ye W(2), Qi X(2), Xi M(3), Liu S(1), Zhu Q(1), Zheng L(1), 
Zhao A(1)(4).

Author information:
(1)Vanke School of Public Health, Tsinghua University, Beijing 100084, China.
(2)Inner Mongolia Dairy Technology Research Institute Co., Ltd., Hohhot 010110, 
China.
(3)College of Food Science and Engineering, Changchun University, Changchun 
130022, China.
(4)Institute for Healthy China, Tsinghua University, Beijing 100084, China.

BACKGROUND/OBJECTIVES: Lactoferrin, a key bioactive component in human milk, may 
bridge functional gaps in infant formula; however, its long-term effects on 
growth and the gut microbiota in term infants remain underexplored, particularly 
in real-world settings.
METHODS: This real-world evidence (RWE) study assessed the impact of 
lactoferrin-fortified formula (LF) on infant growth, the gut microbiota, and 
feeding tolerance compared with control formula (CF) and exclusive breastfeeding 
(BF). After propensity score matching (PSM) for maternal education level and 
infant age, 111 matched Chinese infants (37 per group: LF, CF, and BF; age: 6-12 
months) were analyzed. Growth was evaluated using WHO Z-scores (WAZ, LAZ, WLZ, 
and zBMI). The gut microbiota was profiled via 16S rRNA sequencing (n = 81). 
Feeding challenges were quantified using the Montreal Children's Hospital 
Feeding Scale (MCH-FS).
RESULTS: The LF group exhibited significantly higher length-for-age Z-scores 
(LAZ) compared with both the BF and CF groups (p < 0.001), indicating superior 
linear growth. LF infants also showed reduced MCH-FS scores (18.0 vs. 36.2 in 
CF; p < 0.001), signifying fewer feeding difficulties. Gut microbiota analysis 
revealed enrichment of Bifidobacterium breve and butyrate-producing taxa (e.g., 
Faecalibacterium and Ruminococcaceae), higher alpha diversity, and metabolic 
divergence, involving enhanced lysine fermentation to acetate/butyrate in LF 
infants, suggesting a higher level of short-chain fatty acid (SCFA) production. 
Beta diversity analysis demonstrated that the LF microbiota clustered close to 
BF.
CONCLUSIONS: Lactoferrin-fortified formula was associated with improved linear 
growth and feeding tolerance while shaping a healthy gut microbiota, showing 
similarities to breastfed infants' microbiota. These findings support LF 
fortification as a strategy to improve functional outcomes in formula-fed 
infants.

DOI: 10.3390/nu17243896
PMCID: PMC12736245
PMID: 41470843 [Indexed for MEDLINE]

Conflict of interest statement: The funding sources (the National Key Research 
and Development Program and the National Natural Science Foundation of China) 
played no role in the study design, data collection, analysis, interpretation, 
or in the writing of the manuscript. The authors affiliated with the Inner 
Mongolia Dairy Technology Research Institute Co., Ltd. (B.L., W.Y., X.Q.) were 
not involved in the study design, data collection, data analysis and 
interpretation or manuscript drafting. However, all authors had full access to 
the data and are responsible for the final decision to submit for publication.


70. J Nutr. 2026 Feb;156(2):101290. doi: 10.1016/j.tjnut.2025.101290. Epub 2025 Dec 
27.

Protein Supplements and Protein-Containing Product Use in Ice Cream: A Review.

Mykhalevych A(1), Polishchuk G(2), Kluz M(3), Buniowska-Olejnik M(4).

Author information:
(1)Problem Scientific and Research Laboratory, National University of Food 
Technologies, Kyiv, Ukraine. Electronic address:.
(2)Department of Milk and Dairy Products Technology, Educational and Scientific 
Institute of Food Technologies, National University of Food Technologies, Kyiv, 
Ukraine.
(3)School of Medical and Health Sciences, VIZJA University, Warsaw, Poland; 
Andrzej Frycz Modrzewski Krakow University, Krakow, Poland.
(4)School of Medical and Health Sciences, VIZJA University, Warsaw, Poland.

The increasing demand for high-protein diets, driven by health-conscious 
consumers and global nutritional challenges, has sparked interest in fortifying 
popular food products such as ice cream with protein ingredients. Ice cream, a 
widely consumed dessert, typically offers limited nutritional value, 
particularly in protein content. This review provides a comprehensive overview 
of protein supplementation in ice cream production, focusing on functional and 
technological effects of conventional and emerging protein sources. The article 
critically evaluates milk-derived proteins, as well as underexplored 
alternatives including plant-based, microbial, insect-derived, and aquatic 
proteins. It also addresses legislative constraints, sensory and textural 
challenges, and formulation strategies for optimizing product quality and 
consumer acceptance. The integration of proteins into ice cream not only 
enhances its nutritional profile but also offers potential for market innovation 
aligned with sustainability and health trends. However, the successful 
incorporation of these ingredients into ice cream systems requires navigating 
complex challenges related to formulation performance, sensory quality, 
regulatory compliance, and consumer acceptance. Functional variability, sensory 
trade-offs, and limited industrial validation continue to hinder mainstream 
adoption, especially for novel protein sources. Future advances will depend on a 
deeper mechanistic understanding of protein behavior in multiphase frozen 
matrices, as well as the development of formulation strategies that reconcile 
nutritional value with sensory appeal.

Copyright © 2026 The Authors. Published by Elsevier Inc. All rights reserved.

DOI: 10.1016/j.tjnut.2025.101290
PMID: 41461270 [Indexed for MEDLINE]

Conflict of interest statement: Conflicts of interest The authors report no 
conflicts of interest.


71. Int Breastfeed J. 2025 Dec 25;21(1):9. doi: 10.1186/s13006-025-00801-w.

Impact of three mammalian milk-derived fortifiers on morbidity and mortality in 
preterm infants: a systematic review and network meta-analysis.

Feng Y(#)(1), Gao Y(#)(1), Gu Y(1), Liu Y(1), Xu Y(2), Li S(3).

Author information:
(1)School of Nursing, Guilin Medical University, No.109 Huancheng North 2nd 
Road, Qixing District, Guilin, Guangxi Zhuang Autonomous Region, China.
(2)Affiliated Hospital of Guilin Medical University, No.15 Lequn Road, Diecai 
District, Guilin, Guangxi Zhuang Autonomous Region, China.
(3)School of Nursing, Guilin Medical University, No.109 Huancheng North 2nd 
Road, Qixing District, Guilin, Guangxi Zhuang Autonomous Region, China. 
.
(#)Contributed equally

BACKGROUND: Fortification of breast milk with milk-derived fortifiers is 
essential for the growth and developmental outcomes of preterm infants, yet the 
ideal fortifier remains uncertain. This network meta-analysis assessed the 
effects of bovine milk-derived fortifier (BMF), human milk-derived fortifier 
(HMF), and donkey milk-derived fortifier (DMF) on morbidity and mortality rates 
of preterm infants within neonatal intensive care units (NICUs).
METHODS: We conducted a comprehensive search of multiple databases, including 
MEDLINE, CINAHL, PubMed, CNKI, and the Cochrane Central Register of Controlled 
Trials. The risk of bias for each study was evaluated using the Cochrane Risk of 
Bias 2 (RoB 2) tool.
RESULTS: Ten studies involving 545 preterm infants were identified for 
qualitative descriptive analysis. network meta-analysis was conducted, 
incorporating data from five studies and three different types of milk-based 
fortifiers. The network meta-analysis suggested that HMF may be associated with 
a lower incidence of necrotizing enterocolitis. Compared with HMF, BMF was 
associated with an increased risk of necrotizing enterocolitis (RR = 2.33, 95% 
CI [1.22–4.44]). Cumulative ranking probabilities indicated that HMF appeared to 
be the most favourable fortifier for reducing both necrotizing enterocolitis 
incidence and mortality. DMF might represent a potential option for reducing the 
risk of intraventricular haemorrhage and neonatal sepsis.
CONCLUSION: HMF may be associated with a reduced incidence of necrotizing 
enterocolitis, whereas DMF may offer potential anti-inflammatory benefits, 
particularly for high-risk populations susceptible to intraventricular 
haemorrhage. This study provides direct and comparative evidence of the 
associations between different animal-derived fortifiers and morbidity and 
mortality in preterm infants. It provides a preliminary evaluation of DMF, for 
which limited evidence previously existed, and offers insight into the potential 
mechanisms that connect milk composition to clinical outcomes. However, these 
findings are primarily based on a limited number of studies with small sample 
sizes. Further randomized controlled trials are necessary to validate these 
observations.
SUPPLEMENTARY INFORMATION: The online version contains supplementary material 
available at 10.1186/s13006-025-00801-w.

DOI: 10.1186/s13006-025-00801-w
PMCID: PMC12849099
PMID: 41449441

Conflict of interest statement: Declarations. Ethics approval and consent to 
participate: Not applicable. Consent for publication: Not applicable. Competing 
interests: The authors declare no competing interests.


72. Anal Bioanal Chem. 2026 Feb;418(4):1301-1316. doi: 10.1007/s00216-025-06284-2. 
Epub 2025 Dec 22.

Performance of a targeted LC-MS/MS method for the quantification of peanut, egg, 
and milk in various allergen-fortified or incurred foods.

Butler KE(1), Xiong W(2), Fiedler KL(3).

Author information:
(1)U.S. Food and Drug Administration, Human Foods Program, Office of Laboratory 
Operations and Applied Science, Office of Chemistry and Toxicology, Division of 
Analytical Chemistry, 5001 Campus Drive, College Park, MD, USA.
(2)U.S. Food and Drug Administration, Human Foods Program, Office of Laboratory 
Operations and Applied Science, Office of Chemistry and Toxicology, Division of 
Bioanalytical Chemistry, 5001 Campus Drive, College Park, MD, USA.
(3)U.S. Food and Drug Administration, Human Foods Program, Office of Laboratory 
Operations and Applied Science, Office of Chemistry and Toxicology, Division of 
Analytical Chemistry, 5001 Campus Drive, College Park, MD, USA. 
.

Despite the availability of numerous analytical methods for the quantification 
of food allergens, the global food allergen research community has recognized 
that many challenges in the field remain unresolved. Recently, guidelines have 
been released by multiple entities to align best practices within the field, 
particularly for method validation. With these recommendations in mind, a 
previously published liquid chromatography-tandem mass spectrometry (LC-MS/MS) 
method for the detection and quantification of peanut, egg, and milk was 
evaluated in four food matrices with varied chemical complexity and sample 
processing: fortified oat flour, fortified frozen dessert, incurred sugar 
cookie, and incurred gluten-free (GF) cupcake. Test materials were prepared at 
concentrations of 10, 15, 25, and 100 mg food allergen/kg matrix; then, 
previously established criteria were used to determine if the food allergen was 
detectable and/or quantifiable within each sample. Percent recovery for each 
quantifiable food allergen was determined per matrix and generally met 
recommended method performance requirements. The suitability of the method was 
also compared to target limits of quantification (LOQ) determined for each 
matrix using the ad hoc Joint FAO/WHO Expert Consultation on Risk Assessment of 
Food Allergens reference doses combined with the Reference Amounts Customarily 
Consumed (RACC). The method was able to quantify at the target LOQs for (1) 
peanut in oat flour, frozen dessert, and GF cupcake, (2) egg in oat flour and 
frozen dessert, and (3) milk in frozen dessert, GF cupcake, and cookie, 
demonstrating method performance at current reference doses.

© 2025. This is a U.S. Government work and not under copyright protection in the 
US; foreign copyright protection may apply.

DOI: 10.1007/s00216-025-06284-2
PMID: 41428214 [Indexed for MEDLINE]

Conflict of interest statement: Declarations. Competing interests: The authors 
declare no competing interests. Disclaimer: This article is not an official US 
Food and Drug Administration (FDA) guidance or policy statement. No official 
support or endorsement by the US FDA is intended or should be inferred.


73. J Glob Health. 2025 Dec 22;15:04340. doi: 10.7189/jogh.15.04340.

Neonatal diet and growth outcomes in hospitalised very preterm infants: an 
observational study in middle income countries in Africa, Asia, and Latin 
America.

Belfort MB(1), Ehret DE(2)(3), Greenberg LT(2)(4), Lee AC(5), Procianoy RS(6), 
Semrau KE(7), Silveira RC(6), Tooke L(8), Edwards EM(2)(3)(4).

Author information:
(1)Harvard Medical School and Brigham and Women's Hospital, Department of 
Pediatrics, Boston, Massachusetts, USA.
(2)Vermont Oxford Network, Burlington, Vermont, USA.
(3)University of Vermont Robert Larner, MD College of Medicine, Department of 
Pediatrics, Burlington, Vermont, USA.
(4)University of Vermont College of Engineering and Mathematical Sciences, 
Department of Mathematics and Statistics, Burlington, Vermont, USA.
(5)Warren Alpert Medical School of Brown University, Brown Global Alliance of 
Infant and Maternal Health Research, Department of Pediatrics, Providence, Rhode 
Island, USA.
(6)Universidade Federal do Rio Grande do Sul and Hospital de Clinicas de Porto 
Alegre, Department of Pediatrics, Neonatal Section, Porto Alegre, Rio Grande do 
Sul, Brazil.
(7)Harvard T.H. Chan School of Public Health and Brigham and Women's Hospital, 
Ariadne Labs, Boston, Massachusetts, USA.
(8)University of Cape Town, Department of Paediatrics and Child Health, Cape 
Town, South Africa.

BACKGROUND: Up-to-date data from middle-income countries are needed to inform 
in-hospital feeding practices among small, vulnerable newborns. We aimed to 
quantify growth indicators and their associations with in-hospital diet for 
infants born very preterm or with a very low birth weight (VLBW) of 401-1500 g 
in 12 middle income countries in Africa, Asia, and Latin America.
METHODS: We performed an observational cohort study of infants born at 22-29 
weeks' gestation or VLBW from 2018 to 2024 among Vermont Oxford Network member 
hospitals in 12 middle income countries. We used linear regression to estimate 
adjusted mean change in weight and head circumference (z-scores based on Fenton 
reference) from birth to hospital discharge by category of enteral diet at 
discharge/transfer (human milk only, mixed (human milk with formula and/or 
fortifier), and formula only), adjusting for confounders, in the entire cohort 
and stratified by birth weight and foetal growth status.
RESULTS: Among 35 843 infants, the median length of stay was 50 days 
(interquartile range = 37, 65). Eighty-four percent were receiving at least some 
human milk at discharge (34% human milk only, 50% mixed, 16% formula only). 
Adjusted mean weight z-score declined by 1.40 in the human milk only group, 1.32 
in the mixed group, and 1.17 in the formula only group. The adjusted estimated 
difference between mixed diet and formula only groups was 0.15 z-scores (95% 
confidence interval (CI) = 0.10, 0.20), and between the mixed and human milk 
only groups was 0.08 (95% CI = 0.00, 0.15). Head growth differed little between 
groups.
CONCLUSIONS: Human milk use was high at discharge in this vulnerable population 
of infants born at 22-29 weeks or VLBW in middle- income countries. Infants fed 
a mixed diet gain weight more slowly than infants fed only formula. Rigorous 
intervention studies are needed determine optimal nutrient delivery strategies 
for infants fed human milk in this context.

Copyright © 2025 by the Journal of Global Health. All rights reserved.

DOI: 10.7189/jogh.15.04340
PMCID: PMC12720493
PMID: 41427603 [Indexed for MEDLINE]

Conflict of interest statement: Disclosure of interest: The authors completed 
the ICMJE Disclosure of Interest Form (available upon request from the 
corresponding author) and disclose no relevant interests’


74. BMC Pediatr. 2025 Dec 16;26(1):59. doi: 10.1186/s12887-025-06411-z.

Prevalence and determinants of vitamin D deficiency among children aged 9 months 
to 12 years at a tertiary care center in western India: a cross-sectional study.

Bhonsle SM(1), Makwana NK(2), Vyas BR(1), Vaishnav MR(1).

Author information:
(1)Department of Paediatrics, Shri M P Shah Government Medical College and 
G.G.G. Hospital, Jamnagar, Gujarat, India.
(2)Department of Paediatrics, Shri M P Shah Government Medical College and 
G.G.G. Hospital, Jamnagar, Gujarat, India..

BACKGROUND: Vitamin D deficiency (VDD) is highly prevalent in children in India 
despite abundant sunlight. Multiple behavioral and dietary factors contribute to 
inadequate vitamin D status. We estimated the prevalence of vitamin D deficiency 
and insufficiency among apparently healthy children and explored associated 
sociodemographic, environmental, and dietary factors.
METHODS: We conducted a cross-sectional study at the Department of Pediatrics, 
G.G. Hospital, Jamnagar (April 2023-June 2025). Every fourth eligible child aged 
9 months-12 years attending the immunization clinic or accompanying as a healthy 
sibling was enrolled after consent. A structured proforma captured demographics, 
socioeconomic status, sun exposure (duration/body surface area), sunscreen use, 
and dietary practices (milk, eggs, vegetarian intake). Serum 25-hydroxyvitamin D 
[25(OH)D] was measured by chemiluminescent immunoassay. Vitamin D status 
followed IAP revised cut-offs: deficient < 20 ng/mL; inadequate 20-29 ng/mL; 
sufficient ≥ 30 ng/mL. Associations were analyzed using Chi-square or Fisher's 
exact tests (p < 0.05).
RESULTS: Of 277 children (mean age 5.9 ± 3.3 years; 55.6% male), 34.7% were 
vitamin D deficient and 43.0% inadequate; only 22.4% were sufficient. Vitamin D 
status showed no significant association with age category (p = 0.757), gender 
(p = 0.652), or religion (p = 0.971). A strong dose-response was observed for 
sun exposure: <1 h/day (deficient 50.0%, sufficient 13.0%), 1-2 h/day (deficient 
32.7%, sufficient 21.8%), and > 2 h/day (deficient 9.1%, sufficient 72.7%) 
(p < 0.01). Sunscreen use was associated with higher deficiency (50.0% vs. 
31.8%, p = 0.018). Regular milk intake correlated with better status 
(p = 0.008); moderate egg consumption (3-5 days/week) showed the most favorable 
profile (p < 0.01). Socioeconomic class trended toward association (p = 0.08), 
with the small upper-class subgroup having highest deficiency.
CONCLUSIONS: Over three-quarters of children exhibited suboptimal vitamin D 
status. Modifiable correlates-including sun exposure duration, sunscreen use, 
milk intake, and moderate egg consumption-were significantly associated with 
25(OH)D levels. Pragmatic strategies combining safe sun exposure guidance, 
dietary counseling, food fortification, and targeted supplementation are 
warranted.

© 2025. The Author(s).

DOI: 10.1186/s12887-025-06411-z
PMCID: PMC12828979
PMID: 41402832 [Indexed for MEDLINE]

Conflict of interest statement: Declarations. Ethics approval and consent to 
participate: The study was conducted in full accordance with the ethical 
principles outlined in the Declaration of Helsinki ( 
https://www.wma.net/policies-post/wma-declaration-of-helsinki/ ). Ethical 
approval was obtained from the Institutional Ethics Committee of M.P. Shah 
Government Medical College and Guru Gobindsingh Hospital, Jamnagar (Approval 
No.: 66/01/2023). Written informed consent was obtained from all participants 
(or from parents/legally authorized guardians in case of minors) prior to 
enrolment in the study. Consent for publication: Not applicable — no individual 
person’s data in any form is presented. Competing interests: The authors declare 
no competing interests.


75. Nat Commun. 2025 Dec 12;17(1):237. doi: 10.1038/s41467-025-66930-2.

A young child formula with Limosilactobacillus reuteri and GOS modulates gut 
microbiome and enhances bone and muscle development: a randomized trial.

Bonnet N(1), Capeding MR(2), Siegwald L(1), Garcia-Garcera M(1), Desgeorges 
T(1), Tytgat HLP(1), Krattinger LF(3), Lebumfacil J(4), Phee LC(2), Moll JM(5), 
Gudjonsson A(5), Rodriguez-Garcia P(5), Baruchet M(1), Feige JN(1), Jankovic 
I(6), Chen Y(6), Egli D(6), Horcajada MN(7).

Author information:
(1)Nestlé Institute of Health Sciences, Nestlé Research, Vers-Chez-Les-Blanc & 
EPFL innovation Park, Lausanne, Switzerland.
(2)Clinical Research Unit, Asian Foundation for Tropical Medicine Inc. (AFTMI), 
Muntinlupa City, Philippines.
(3)Clinical Research Unit, Nestlé Research, Lausanne, Switzerland.
(4)Wyeth Nutrition, Makati City, Philippines.
(5)Cmbio, Copenhagen, Denmark.
(6)Nestlé Product Technology Center-Nutrition, Société des Produits Nestlé S.A., 
Vevey, Switzerland.
(7)Nestlé Institute of Health Sciences, Nestlé Research, Vers-Chez-Les-Blanc & 
EPFL innovation Park, Lausanne, Switzerland. 
.

In this randomized, double-blind controlled trial, 182 Filipino children aged 
2-3 years received either an experimental young child formula (EYCF) containing 
a combination of Limosilactobacillus reuteri DSM 17938 and 
galacto-oligosaccharides (GOS; n = 91) or a minimally fortified milk (CM; 
n = 91) for 6 months. Primary outcome was tibia speed of sound and secondary 
outcomes were muscle strength, blood vitamin D levels, bone turnover markers, 
gut microbiota, fecal calcium fatty acid soaps and gastro-intestinal tolerance. 
Compared to CM, those in the EYCF group showed increased tibia speed of sound 
after 3 and 6 months. The intervention remodeled the stool microbiome 
composition, assessed by shotgun metagenomics, with enrichment of L. reuteri and 
higher bifidobacteria presence in the EYCF group. Increased L. reuteri abundance 
after 6 months of EYCF consumption associates with higher bone quality and 
muscle strength. Stool metabolomics show 45 metabolites modulated by EYCF 
consumption and associated to microbiome compositional changes, leading to 
enrichment of tryptophane and indole metabolism. In summary, consumption of EYCF 
containing a L. reuteri + GOS synbiotic improves musculoskeletal development in 
toddlers via modulation of microbiota composition and function. These results 
provide insights on gut-musculoskeletal crosstalk during early life. 
Clinicaltrial.gov NCT04799028.

© 2025. The Author(s).

DOI: 10.1038/s41467-025-66930-2
PMCID: PMC12783733
PMID: 41387706 [Indexed for MEDLINE]

Conflict of interest statement: Competing interests: Some authors (N.B., L.S., 
M.G.G., T.D., H.L.P.T., L.F.K., J.L., M.B., J.N.F., I.J., Y.C., D.E., and 
M.N.H.) are employees from Société des Produits Nestlé SA. Cmbio employees 
(J.M.M., A.G., and P.R.G.) contributed within a service agreement contract. 
M.R.C. and L.C.P. had no conflict of interest to report.


76. Acta Paediatr. 2026 Apr;115(4):865-872. doi: 10.1111/apa.70414. Epub 2025 Dec 
12.

Breastfeeding Ambulatory Supplementation With Preterm Formula Versus Fortifiers 
Achieves Similar Growth at 40 Weeks: A Pilot Study in Preterm Infants.

Cadavid AM(1)(2), Lombo A(1)(2), Montealegre A(1)(2)(3), Angel DA(4), Quevedo 
A(5), Charpak N(2).

Author information:
(1)Integral Kangaroo Mother Care Program, Medellín, Colombia.
(2)Kangaroo Foundation, Bogotá, Colombia.
(3)San Ignacio Teaching Hospital, Javeriana University, Bogotá, Colombia.
(4)Integral Kangaroo Mother Care Program, Bogotá, Colombia.
(5)Department of Pediatrics, Universidad de Antioquia, Medellín, Colombia.

AIM: This study evaluates whether liquid formula supplementation for preterm 
infants was non-inferior to breast milk fortifiers regarding growth and 
adherence to exclusive breastfeeding in an outpatient Kangaroo Mother Care 
Program from Colombia.
METHODS: An open-label randomised trial was initiated in 2016 and interrupted in 
2018 because of the lack of fortifiers in the country and included 81 preterm 
infants born between 32 + 0 and 36 + 6 weeks/days of gestational age. Infants 
were assigned to receive either breast milk fortifiers or liquid formula, with 
outcomes assessed up to 40 weeks.
RESULTS: No significant differences in anthropometric measurements were observed 
between the groups. More than two-thirds (69%) achieved exclusive breastfeeding, 
with higher rates in the fortifier group (73.0% vs. 53.0%, p = 0.06). This 
tendency persisted up to 6 months of corrected age. Breastfeeding was 
discontinued more frequently in the liquid formula group despite showing better 
gastrointestinal tolerance, ease of administration, and lower cost and 
contamination for home use.
CONCLUSIONS: Liquid formula supplementation showed growth outcomes comparable to 
those of breast milk fortifiers at 40 weeks. Further research with larger 
samples is recommended to validate outcomes.

© 2025 Foundation Acta Paediatrica. Published by John Wiley & Sons Ltd.

DOI: 10.1111/apa.70414
PMID: 41386682 [Indexed for MEDLINE]


77. Food Chem. 2026 Jan 30;500:147466. doi: 10.1016/j.foodchem.2025.147466. Epub 
2025 Dec 8.

Collagen peptides increase curcumin bioaccessibility in milk protein-stabilized 
emulsions by increasing mixed micelle solubilization capacity.

Hu S(1), Dong W(2), Zou Y(1), Tian C(2), McClements DJ(3), Zhao Y(4), Li Y(5).

Author information:
(1)College of Food Science and Technology, Huazhong Agricultural University, 
Wuhan 430070, China.
(2)Shanghai M-Action Health Technology Co., LTD, Shanghai 200120, China.
(3)Department of Food Science, University of Massachusetts, Amherst, MA 01003, 
USA.
(4)Shanghai M-Action Health Technology Co., LTD, Shanghai 200120, China. 
Electronic address:.
(5)College of Food Science and Technology, Huazhong Agricultural University, 
Wuhan 430070, China; Key Laboratory of Environment Correlative Dietology 
(Huazhong Agricultural University), Ministry of Education, China. Electronic 
address:.

Collagen peptides (CPTs) are common ingredients in protein-rich functional 
beverages. However, the impact of CPTs on the digestion behavior of 
nutraceutical-fortified functional beverages remains poorly understood. We 
therefore investigated the effects of CPT on the structural, physicochemical, 
and digestion properties of curcumin-loaded milk protein concentrate 
(MPC)-stabilized emulsions. CPT addition had little impact on the 
physicochemical stability or encapsulation efficiency (>95 %) of MPC emulsions. 
In vitro digestion experiments showed that CPT addition increased curcumin 
bioaccessibility (1.14- to 1.35-fold), but slightly decreased lipid hydrolysis 
of MPC-emulsions. Microstructural analysis of the digesta indicated that the 
presence of CPT induced the formation of hollow vesicle structures in the small 
intestinal phase. The presence of CPT in the soluble fraction of the digesta 
increased the solubilization of oleic acid in a dose-dependent manner. The 
results suggest that CPT enhance the bioaccessibility of curcumin in fortified 
beverages, thereby enhancing the nutritional profile of these products.

Copyright © 2025 Elsevier Ltd. All rights reserved.

DOI: 10.1016/j.foodchem.2025.147466
PMID: 41380566 [Indexed for MEDLINE]

Conflict of interest statement: Declaration of competing interest The authors 
declare that they have no known competing financial interests or personal 
relationships that could have appeared to influence the work reported in this 
paper.


78. Food Chem. 2026 Jan 30;500:147462. doi: 10.1016/j.foodchem.2025.147462. Epub 
2025 Dec 8.

Interactions and mechanisms in iron-fortified skim milk: Physicochemical 
properties, calcium bioavailability, and cellular responses.

Li Y(1), Cheng Y(1), Tang N(2).

Author information:
(1)College of Food Science and Nutritional Engineering, China Agricultural 
University, Beijing 100083, China; Beijing Key Laboratory of Functional Food 
from Plant Resources, Beijing 100083, China.
(2)College of Food Science and Nutritional Engineering, China Agricultural 
University, Beijing 100083, China; Beijing Key Laboratory of Functional Food 
from Plant Resources, Beijing 100083, China. Electronic address: 
.

Iron fortification of skim milk offers a promising strategy to address global 
micronutrient deficiencies, however, its effects on the physicochemical 
properties, mineral distribution, and bioavailability of milk remains poorly 
understood. The present study provides a comprehensive analysis of the 
interactions and mechanisms involved in iron-fortified skim milk systems. Iron 
salts (ferrous chloride, iron(II) lactate, and iron(II) gluconate) induced a 
concentration-dependent pH reduction, with ferrous chloride causing the most 
significant acidification. Iron distribution varied by salt type, with iron(II) 
gluconate showing higher serum iron levels. Calcium redistribution revealed a 
22-28 % aqueous-phase depletion due to competitive iron-phosphate binding. 
Particle size remained stable, while ζ-potential decreased, reflecting 
iron-induced charge neutralization. Viscosity increased with iron concentration, 
indicating enhanced casein cross-linking. In vitro digestion and cellular uptake 
studies demonstrated that skim milk protein hydrolysates (MH) improved calcium 
bioavailability by forming soluble peptide‑calcium complexes, upregulating 
calcium transporters (TRPV6 and Claudin-2) and mitigating iron-induced oxidative 
stress. MH also enhanced antioxidant activity, reducing ROS and lipid 
peroxidation while upregulating ferroportin (FPN1) via Nrf2 activation. These 
findings highlight skim milk as an effective carrier for iron fortification, 
enabling simultaneous calcium and iron supplementation while maintaining 
stability and bioavailability.

Copyright © 2025 Elsevier Ltd. All rights reserved.

DOI: 10.1016/j.foodchem.2025.147462
PMID: 41380559 [Indexed for MEDLINE]

Conflict of interest statement: Declaration of competing interest The authors 
declare that they do not have any known competing financial interests or 
personal relationships that could have appeared to influence the work reported 
in this paper.


79. Nutrients. 2025 Nov 30;17(23):3765. doi: 10.3390/nu17233765.

Dairy Intake and Iodine Status in Pregnant and Lactating Women: A Systematic 
Review and Meta-Analysis.

Razmpoosh E(1), Geronimo K(1), Gauntlett C(1), Vicente I(1), Pham P(1), Shah 
A(1), Musa-Veloso K(1).

Author information:
(1)Intertek Health Sciences Inc., 2233 Argentia Road, Suite 201, Mississauga, ON 
L5N 2X7, Canada.

Background/Objectives: Despite salt iodization, iodine deficiency during 
pregnancy and lactation is re-emerging in many industrialized countries, 
necessitating an evaluation of the role of dairy in supporting iodine status 
during these critical periods. Methods: We conducted a systematic review and 
meta-analysis in accordance with PRISMA guidelines. Ten databases were searched 
through March 2025 using ProQuest Dialog™ (Version 75.0). Study quality was 
assessed using either the Quality Evaluation for Observational Studies tool or 
the Risk of Bias 2 tool, depending on study design. Random-effects models were 
applied, with certainty of evidence rated using the GRADE framework. Publication 
bias, sensitivity analyses, and subgroup analyses were also performed. Results: 
Fifty-one publications met the eligibility criteria, including 50 publications 
of observational studies and 1 publication of a randomized controlled trial 
(RCT), with most studies conducted on pregnant women. Higher dairy intake was 
associated with significantly greater urinary iodine concentration (UIC), (23 
studies; standardized mean difference: 0.326; 95% confidence interval [CI]: 
0.228, 0.424; p < 0.001; I2 = 57.31%; low-certainty), and significantly lower 
odds of iodine deficiency (11 studies; odds ratio: 0.58; 95% CI: 0.48, 0.70; p < 
0.001; I2 = 0%; moderate-certainty). Associations were stronger in studies 
conducted during later pregnancy, in higher-quality studies, and when the dairy 
food was specifically milk. Dairy contributed to ~27% of iodine intake from 
foods. Breast milk iodine concentration (BMIC) findings were inconsistent, 
though in one RCT, iodine-fortified milk improved BMIC and UIC. Conclusions: 
Dairy intake supports adequate iodine status during pregnancy and lactation. 
RCTs would be valuable in further investigating the role of dairy in supporting 
iodine status, particularly in lactating women. (PROSPERO CRD420251054576).

DOI: 10.3390/nu17233765
PMCID: PMC12693841
PMID: 41374057 [Indexed for MEDLINE]

Conflict of interest statement: The authors are employees of Intertek Health 
Sciences Inc., which has provided, and continues to provide, paid services to 
Dairy Management Inc. The work presented in this manuscript was funded by the 
National Dairy Council, which is managed by Dairy Management Inc. Neither the 
National Dairy Council nor Dairy Management Inc. had any role in the design of 
the study; in the collection, analyses, or interpretation of data; in the 
writing of the manuscript; or in the decision to publish the manuscript.


80. Nutrients. 2025 Nov 29;17(23):3757. doi: 10.3390/nu17233757.

Is Vitamin D Fortification of Dairy Products Effective for Improving Vitamin D 
Status? A Systematic Review and Meta-Analysis of Randomised Controlled Trials.

Wong CL(1), Givens DI(2), Turpeinen AM(3), Liu X(1), Guo J(1).

Author information:
(1)School of Food Science and Nutrition, Faculty of Environment, University of 
Leeds, Leeds LS2 9JT, UK.
(2)Institute for Food, Nutrition and Health, University of Reading, Reading RG6 
6EU, UK.
(3)Valio Ltd., R&D, 00370 Helsinki, Finland.

Background/objectives: Given the suboptimal vitamin D intake and status among 
the United Kingdom population, the Scientific Advisory Committee on Nutrition is 
seeking suitable food vehicles for vitamin D fortification. Thus, this study 
aimed to examine the efficacy of vitamin D-fortified dairy products in improving 
serum 25-hydroxyvitamin D [25(OH)D] concentration using data from randomised 
controlled trials (RCTs). Methods: PubMed, Embase, and Web of Science were 
searched from inception until October 2024. Studies were included if they were 
RCTs with intervention groups administered vitamin D-fortified dairy products 
and control groups administered unfortified dairy products, as well as examining 
the effects on serum 25(OH)D concentration. Results: There were 35 RCTs eligible 
for inclusion, involving 4965 participants (intervention: 2526; control: 2439). 
The results showed that serum 25(OH)D concentrations were significantly 
increased by vitamin D3-fortified milk/milk powder (n = 15, mean difference 
(MD): 18.31 nmol/L, 95% confidence interval (CI): 13.30-23.33 nmol/L, I2 = 95%), 
vitamin D3-fortified yoghurt/yoghurt drinks (n = 11, MD: 26.22 nmol/L, 95% CI: 
18.67-33.77 nmol/L, I2 = 97%), vitamin D2-fortified milk/milk powder (n = 3, MD: 
11.61 nmol/L, 95% CI: 9.31-13.91 nmol/L, I2 = 0%), vitamin D-fortified (type not 
specified) milk/milk powder (n = 8, MD: 13.59 nmol/L, 95% CI: 8.54-18.64, I2 = 
98%), and vitamin D-fortified (type not specified) yoghurt/yoghurt drinks (n = 
4, MD: 27.74 nmol/L, 95% CI: 16.83-38.64 nmol/L, I2 = 91%), but insignificantly 
increased by vitamin D3-fortified cheese (n = 5, MD: 16.78 nmol/L, 95% CI: 
-3.61-37.16, I2 = 99%). However, the results of vitamin D3-fortified cheese 
became significant when leave-one-out analysis was performed by omitting one RCT 
(MD: 24.13 nmol/L, 95% CI: 4.69-43.58, I2 = 90%). Conclusions: These findings 
provide evidence that vitamin D-fortified dairy products have the potential to 
improve serum 25(OH)D concentrations in populations.

DOI: 10.3390/nu17233757
PMCID: PMC12694000
PMID: 41374048 [Indexed for MEDLINE]

Conflict of interest statement: The authors declare that this research was 
supported by a grant from the Consumer Lab funded by the Biotechnology and 
Biological Sciences Research Council, which provided financial support for 
workshops and the employment of a research assistant (C.L.W.). The funding body 
had no role in the study design, data collection and analysis, decision to 
publish, or preparation of the manuscript. The authors also declare that A.T., 
from Valio Ltd. (Finland), was our formal partner, providing industry input and 
in-kind analytical support. Author D.I.G. also declares that he has received 
travel expenses and honoraria in connection with lectures and meetings from the 
UK Dairy Council (now Dairy UK), the Dutch Dairy Association, the European Dairy 
Association, the International Dairy Federation, Centre National 
Interprofessionnel de l’Industrie Laitière, and the US Dairy Research Institute.


81. Ultrason Sonochem. 2026 Jan;124:107702. doi: 10.1016/j.ultsonch.2025.107702. 
Epub 2025 Dec 4.

Red cabbage anthocyanin-rich natural colorant: Optimization of 
ultrasound-assisted extraction, microencapsulation, bioaccessibility, and 
functional yogurt development.

Kaur S(1), Rani A(1), Thakur R(1), Sharma A(1), Kuksal K(1), Sharma A(1), Nile 
AS(2), Nile SH(3).

Author information:
(1)Division of Food and Nutritional Biotechnology, BRIC-National Agri-Food and 
Biomanufacturing Institute (Formerly National Agri-Food Biotechnology 
Institute), Sector-81, Mohali 140306 Punjab, India.
(2)Department of Food Science and Technology, School of Health Sciences, Amity 
University, Sector-82A, Sahibzada Ajit Singh Nagar, 140306 Punjab, India.
(3)Division of Food and Nutritional Biotechnology, BRIC-National Agri-Food and 
Biomanufacturing Institute (Formerly National Agri-Food Biotechnology 
Institute), Sector-81, Mohali 140306 Punjab, India. Electronic address: 
.

Anthocyanins from red cabbage (Brassica oleracea var. capitata f. rubra) are 
promising natural pigments with notable health benefits, but their thermal and 
gastrointestinal instability limits functional food applications. This study 
employed ultrasound-assisted extraction (UAE) and spray-drying 
microencapsulation to enhance their stability, bioaccessibility, and 
functionality. Optimization using a Box-Behnken design (pH 3.0-5.0, 30-60 °C, 
20-90  min, 0.025-0.05  g/mL solid-liquid ratio) identified optimal UAE 
conditions (pH 3.5, 45 °C, 55  min, 0.0375  g/mL), yielding 125.72 ± 0.83  mg/L 
anthocyanins. Freeze drying retained the highest anthocyanin content 
(129.02 ± 3.36  mg/L), while spray-drying with 10 % maltodextrin achieved 
excellent encapsulation efficiency (96.36 ± 0.80 %) and yield (99.40 %). FTIR 
confirmed the presence of phenolic groups, and SEM revealed spherical 
microcapsules (∼10.27  µm) with smooth surfaces. Encapsulation significantly 
improved thermal stability at 70 °C, reducing the degradation rate constant 
(0.00043  min-1) and increasing half-life (1919.02  min), compared to 
non-encapsulated samples. During simulated digestion, bioaccessibility of 
encapsulated anthocyanins was lower in the oral (53.28 %) and intestinal 
(25.37 %) phases than free forms, but stability was enhanced. Antioxidant 
activity (DPPH: 66.90 %, ABTS: 71.76 %) and enzyme inhibition (α-amylase IC50: 
46.00  µg/mL, α-glucosidase IC50: 51.51  µg/mL) indicated potent bioactivity. 
Cytotoxicity testing on HEK-293 cells showed > 90 % viability, confirming 
safety. Yogurt fortified with 3 % encapsulated powder received high sensory 
scores (8.30 ± 0.86), especially for color and taste. These findings validate 
the integrated UAE and encapsulation approach as a scalable method to stabilize 
red cabbage anthocyanins for functional food applications.

Copyright © 2025 The Author(s). Published by Elsevier B.V. All rights reserved.

DOI: 10.1016/j.ultsonch.2025.107702
PMCID: PMC12732316
PMID: 41351955 [Indexed for MEDLINE]

Conflict of interest statement: Declaration of competing interest The authors 
declare that they have no known competing financial interests or personal 
relationships that could have appeared to influence the work reported in this 
paper.


82. Clin Nutr ESPEN. 2026 Feb;71:102835. doi: 10.1016/j.clnesp.2025.11.155. Epub 
2025 Nov 28.

Impact of fish oil-enriched high protein peptide-based formula on free fatty 
acid profiles and metabolomic changes in home enterally fed patients: A pilot 
study.

Mohamed Elfadil O(1), Johnson DA(1), Pattinson AK(1), Hurt RT(2), Mundi MS(3).

Author information:
(1)Division of Endocrinology, Diabetes, Metabolism and Nutrition, Mayo Clinic, 
Rochester, MN, USA.
(2)Division of Endocrinology, Diabetes, Metabolism and Nutrition, Mayo Clinic, 
Rochester, MN, USA; Division of General Internal Medicine, Mayo Clinic, 
Rochester, MN, USA; Division of Gastroenterology and Hepatology, Mayo Clinic, 
Rochester, MN, USA.
(3)Division of Endocrinology, Diabetes, Metabolism and Nutrition, Mayo Clinic, 
Rochester, MN, USA. Electronic address:.

BACKGROUND: Peptide-based formulas (PBFs) are safe and effective and are 
commonly used for home enteral nutrition (HEN) therapy, especially in patients 
with EN intolerance. There is a lack of data regarding biomolecular changes with 
EN, particularly concerning PBFs. This pilot study examines tolerance and 
changes in metabolic panels, including fatty acids and metabolomics, during the 
transition to a high-protein (100 % whey), fish oil-fortified, peptide-based 
formula (HP/FO w-PBF).
METHODS: A pilot single-center prospective cohort study was conducted at a 
quaternary care center. Adult HEN-dependent participants switched from a 
standard polymeric formula (SPF) to HP/FO w-PBF for 14 days during their 
outpatient HEN follow-up routine visits. Participants completed a daily 
consumption and tolerance diary and collection of plasma at enrollment and the 
end of the study.
RESULTS: Twenty-five participants (72 % female) with a mean age of 60.3 ± 16.3 
years completed the study, with 18 completing baseline and end-of-study blood 
tests for metabolomics and 17 for metabolic panel and fatty acid profile. 
Compared to baseline, C-peptide and insulin levels increased by a median of 36 % 
and 13 %, respectively, possibly reflecting an insulinotropic effect of whey 
protein. Omega-3 fatty acids, EPA and DHA, plasma levels increased significantly 
by a median of 469 % and 143 %, respectively. Metabolomics analyses noted 
essential amino acid enrichment. A significant rise in arginine metabolites, 
such as ornithine, was observed, indicating possible utilization by cells such 
as enterocytes.
CONCLUSIONS: A high-protein (100 % whey), fish-oil-enriched, peptide-based 
formula was tolerated by participants and demonstrated increased levels of 
essential amino acids, as well as a boost in ω-3 fatty acids. However, further 
research is necessary to determine the long-term clinical impact given the short 
duration of this pilot study.

Copyright © 2025 The Authors. Published by Elsevier Ltd.. All rights reserved.

DOI: 10.1016/j.clnesp.2025.11.155
PMID: 41320157 [Indexed for MEDLINE]

Conflict of interest statement: Conflict of interest Mundi, M.S., has research 
grants from Nestlé Health Science, Fresenius Kabi, VectivBio, Rockfield, and 
Realfood Blends. He is on NorthSea’s advisory board. Hurt, R.T., is a consultant 
for Nestlé Health Science and has research funding from Zealand Pharm. Mohamed 
Elfadil, O., is on Nutricia’s advisory board. Johnson, D.A., and Pattinson, A.K. 
have no relevant conflicts of interest to declare.


83. J Dairy Sci. 2026 May;109(5):4973-4987. doi: 10.3168/jds.2025-27345. Epub 2025 
Nov 27.

Fortification of chamomile and saffron in dry yogurt production: Effects on its 
physicochemical and sensory properties.

Shabbir J(1), Itrat N(2), Nazir A(1), Israr B(3), Ahmad AMR(4).

Author information:
(1)Department of Nutrition and Dietetics, The University of Faisalabad, 
Faisalabad, Pakistan, 38000.
(2)Institute of Home Sciences, Faculty of Food, Nutrition and Home Sciences, 
University of Agriculture, Faisalabad, Pakistan, 38000. Electronic address: 
.
(3)Institute of Home Sciences, Faculty of Food, Nutrition and Home Sciences, 
University of Agriculture, Faisalabad, Pakistan, 38000.
(4)Department of Human Nutrition and Dietetics, School of Health Sciences, 
National University of Sciences and Technology (NUST), Islamabad, Pakistan, 
44000. Electronic address:.

The study aimed to formulate fortified dry yogurt with chamomile and saffron, 
making a functional, probiotic, and therapeutic food. Fortified dry yogurt was 
prepared in 7:2:1 ratios of dry yogurt, chamomile, and saffron powders. 
Flavoring of fortified dry yogurt was done at 7:3 ratios of fortified dry yogurt 
and each flavor (mint, cumin seeds, fennel seeds, and black pepper powder). The 
prepared fortified dry yogurt was acidic (pH 3-4) and the total soluble solids 
were 49.97% of the product concentrate. Plain yogurt (100% natural plain yogurt) 
had the highest sensory scores regarding appearance, texture, color, taste, 
aroma, flavor, consistency, mouthfeel, and overall acceptability, whereas 
fortified dry yogurt (FDY) had the lowest scores. Among 4 flavors of FDY, the 
cumin-flavored fortified dry yogurt (CFFDY) showed the highest organoleptic 
potential and was selected for further analysis. The CFFDY proximate profile 
contains a low fiber concentration but is rich in moisture (7.55%), ash (8.13%), 
protein (9.22%), fat (21.85%), and carbohydrates (52.360%), and 442.73 kcal/100 
g. The mineral profile of CFFDY contains low magnesium, but calcium (397.71 
mg/100 g) and sodium (776.89 mg/100 g) were high. The total phenolic content 
(582.78 mg of gallic acid equivalents/100 g), flavonoid content (204.92 mg of 
quercetin equivalents/100 g), and ferric reducing antioxidant power (2,022.82 mg 
Eq of FeSO4/100 g) show the stronger antioxidant potential of CFFDY. Pathogenic 
bacterial species was not detected in CFFDY, but the total plate count was 2.5 × 
103 cfu/g, indicating the presence of probiotic species. Gram staining detected 
small chains of gram-positive cocci cells and medium chains of gram-positive 
palisades that are Bacillus and Bifidobacterium species, whereas glucose 
fermentation and lactose fermentation tests both confirmed the presence of 
Lactobacillus in CFFDY. A storage study of FDY was also conducted and revealed 
high potential organoleptic properties at d 90. Therefore, a novel formulation 
of flavored FDY may be adopted in the dairy industry, serving as a functional 
alternative and shelf-stable dairy product.

The Authors. Published by Elsevier Inc. on behalf of the American Dairy Science 
Association®. This is an open access article under the CC BY license 
(http://creativecommons.org/licenses/by/4.0/).

DOI: 10.3168/jds.2025-27345
PMID: 41317849 [Indexed for MEDLINE]


84. Plant Foods Hum Nutr. 2025 Nov 28;80(4):202. doi: 10.1007/s11130-025-01437-1.

Impact of Spirulina Extract-Loaded Quinoa Protein Isolate Nanoparticles on the 
Quality and Stability of Functional Set Yoghurt.

Maged EMAE(1), Elhendy HA(1), Ahmed SM(1), Darwish AMG(2)(3), El-Enshasy 
HA(4)(5)(6), Soliman TN(7).

Author information:
(1)Home Economics Department, Faculty of Agriculture, Alexandria University, 
Alexandria, Egypt.
(2)Food Industry Technology Program, Faculty of Industrial and Energy 
Technology, Borg Al Arab Technological University (BATU), Alexandria, Egypt. 
.
(3)Food Technology Department, Arid Lands Cultivation Research Institute 
(ALCRI), City of Scientific Research and Technological Applications (SRTA-City), 
New Borg El Arab, P.O. Box 21934, Alexandria, Egypt..
(4)Innovation Centre in Agritechnology for Advanced Bioprocessing (ICA), 
Universiti Teknologi Malaysia (UTM), Johor, Malaysia.
(5)Faculty of Chemical and Energy Engineering, Universiti Teknologi Malaysia 
(UTM), Johor Bahru, Johor, Malaysia.
(6)City of Scientific Research and Technology Application (SRTA-City), New Burg 
Al Arab, Alexandria, Egypt.
(7)Department of Dairy, Food Industries and Nutrition Research Division, 
National Research Centre, Dokki, Cairo, Egypt..

Protein and active compounds play a key role in maintaining human health as 
functional nutrients and as tools in structuring enhanced food products quality. 
This study explored the potentials of novel quinoa protein isolate nanoparticles 
(QPI-NPS) loaded with spirulina ethanol extract (SE) with different 
concentrations (0.2, 0.4, and 0.6%), and its impact on enhanced properties 
assessing; chemical, physical, functional and sensory attributes of set yoghurt 
on time intervals up to 21 days of cold storage. Characterization of the 
(QPI-NPS) using dynamic light scattering (DLS) revealed particle sizes ranging 
from 219.90 to 720.40 nm and the polydispersity indices (PDI) was between 0.270 
and 0.386, with transmission electron microscopy (TEM) defining nanoparticle 
microstructure formation. Encapsulation efficiency reached a maximum of 89.75%. 
High-Performance Liquid Chromatography (HPLC) analysis identified gallic acid, 
chlorogenic acid, catechin, methyl gallate, naringenin, and quercetin as 
dominant phenolic compounds in SE and quinoa flour (QF). Results indicated that 
incorporating 0.4% spirulina extract quinoa protein isolate nanoparticles 
(SE-QPI-NPS) optimized particle size, stability, homogeneity, and encapsulation 
efficiency. The QPI-NPS fortification resulted enhanced the fortified yoghurt 
chemical, viscosity, antioxidant potentials and sensory attributes. This study 
highlights the promising applications of novel QPI-NPS as a delivery vehicle for 
spirulina extract, improving the functionality and consumer acceptability of 
functional food products.

© 2025. The Author(s).

DOI: 10.1007/s11130-025-01437-1
PMCID: PMC12662863
PMID: 41315134 [Indexed for MEDLINE]

Conflict of interest statement: Declarations. Ethical Approval: Not applicable. 
Conflict of Interests: The authors declare no competing interests.


85. Acta Paediatr. 2026 Mar;115(3):686-696. doi: 10.1111/apa.70385. Epub 2025 Nov 
27.

Nutritional Strategies in Preterm Infants: The Role of Human Milk in 
Bronchopulmonary Dysplasia.

Merino-Hernández A(1), Rodríguez-Corrales E(1), Caballero-Martín S(1), 
Ramos-Navarro C(1), González-Pacheco N(1), de Orgaz MCS(1), Sánchez-Luna M(1).

Author information:
(1)Department of Neonatology, Gregorio Marañon University Hospital, Madrid, 
Spain.

AIM: Bronchopulmonary dysplasia (BPD) is a major cause of morbidity and impaired 
growth in preterm infants. This study compared the incidence of grade 2-3 BPD in 
infants predominantly fed with mother's own milk (MOM) or pasteurised donor milk 
(PDM) and examined associations between nutrition and BPD severity.
METHODS: Prospective single-centre study including infants < 32 weeks' gestation 
admitted between January 2023 and June 2024. Infants were classified by 
predominant feeding type (> 50% MOM or PDM) and BPD severity (no BPD/1 or BPD 
2-3). Nutritional intake, feeding tolerance, fortification timing and growth 
were analysed.
RESULTS: Of 158 infants, 88 (56%) received > 50% MOM and 70 (44%) > 50% PDM. BPD 
2-3 occurred in 27 (17.1%) infants with no significant group difference (19% vs. 
14%, p = 0.41). Feeding intolerance (44% vs. 11%, p < 0.01), delayed 
fortification (12.5 vs. 7.0 days, p < 0.01), and later enteral feeding (12.0 vs. 
6.0 days, p < 0.01) were associated with BPD 2-3.
CONCLUSION: No significant difference in BPD 2-3 incidence was found between MOM 
and PDM groups. Feeding intolerance, delayed fortification, and slower enteral 
transition were independent risk factors for grade 2-3 BPD.

© 2025 Foundation Acta Paediatrica. Published by John Wiley & Sons Ltd.

DOI: 10.1111/apa.70385
PMID: 41307366 [Indexed for MEDLINE]


86. Nutrients. 2025 Nov 18;17(22):3606. doi: 10.3390/nu17223606.

Mapping Iodine Sources for Human Nutrition in Portugal Considering Geography, 
Seasonality, and Processing: Milk and Plant-Based Milk Alternatives.

Machado SI(1), Machado A(2)(3), Bordalo AA(2)(3), Roque S(1), Borges N(4), Palha 
JA(1).

Author information:
(1)Life and Health Sciences Research Institute (ICVS), School of Medicine, 
University of Minho, 4710-057 Braga, Portugal.
(2)Laboratory of Hydrobiology and Ecology, Institute of Biomedical Sciences 
(ICBAS-UP), University of Porto, 4050-313 Porto, Portugal.
(3)Interdisciplinary Centre of Marine and Environmental Research (CIIMAR), 
University of Porto, 4450-208 Matosinhos, Portugal.
(4)Faculty of Nutrition and Food Sciences, University of Porto, 4150-180 Porto, 
Portugal.

BACKGROUND: Iodine is a micronutrient essential for the synthesis of thyroid 
hormones and crucial throughout life. Milk is potentially one of the major 
contributors to iodine intake in many countries, including Portugal, due to its 
consumption patterns.
OBJECTIVES: This study characterizes iodine content seasonality in milk and 
plant-based milk alternatives commercially available in Portugal.
RESULTS: Milk products representing summer and winter seasonal pastures and 
plant-based alternatives were obtained from major Portuguese supermarkets. 
Iodine was quantified by the Sandell-Kolthoff reaction in 146 winter and 142 
summer milk products, as well as in 128 plant-based alternatives. Cow's milk 
contained relevant iodine levels (Md = 19.9, IQR = 9.9 µg/100 mL), with no 
influence of thermal processing, fat or lactose content, season, or being 
organic and/or from pasture. However, regional differences were observed. The 
iodine concentration in plant-based drinks was residual, except for four 
fortified products.
CONCLUSIONS: This study provides evidence that milk is still a relevant source 
of iodine in Portugal, whereas most plant-based alternatives provide negligible 
iodine, unless fortified. Given shifts towards plant-based diets, monitoring 
iodine sources and adjusting health policies to fulfil nutritional requirements 
are pivotal to ensure iodine adequacy.

DOI: 10.3390/nu17223606
PMCID: PMC12655576
PMID: 41305656 [Indexed for MEDLINE]

Conflict of interest statement: The authors declare no conflicts of interest.


87. Res Sq [Preprint]. 2025 Oct 7:rs.3.rs-7652186. doi: 10.21203/rs.3.rs-7652186/v1.

Macronutrient and Bioactive Profiles of Donor Milk Differ in Commercial vs 
Non-Profit Milk Banks.

Young B, Tarkleson T(1), Cole L(1), Srivastava M(1), Scholer L(1), Gale C(1), 
Meyers J(2), D'Angio C(2), Ackley D(3).

Author information:
(1)University of Rochester School of Medicine and Dentistry.
(2)University of Rochester Medical Center.
(3)University of Rochester.

Update in
    J Perinatol. 2026 Apr 21. doi: 10.1038/s41372-026-02679-w.

OBJECTIVE: The objective was to explore variation of macronutrient and bioactive 
content between levels of human milk-derived human milk fortifier (HM-HMF), and 
between donor human milk (DHM) from commercial vs non-profit banks.
STUDY DESIGN: We analyzed the concentration of macronutrients, Immunoglobulin A 
(IgA), and cortisol in multiple lots of each HM-HMF product (20, 24, 26, 28, and 
30 kcal/oz) using multiple methods.
RESULT: At each level of caloric density (p<0.0001), protein, carbohydrate, and 
caloric concentration in HM-HMF increased with minor exceptions while fat and 
cortisol concentrations did not differ. Total IgA concentrations differed by 
product type (p <0.0001). Protein concentration did not differ between 
commercial and non-profit donor milk banks while carbohydrates, fat, calories, 
and cortisol were higher, and IgA was lower. Lot-to-lot variability of all DHM 
components was lower in commercial DHM.
CONCLUSION: This study expands data on the variability in the composition of DHM 
originating from various sources.

DOI: 10.21203/rs.3.rs-7652186/v1
PMCID: PMC12632588
PMID: 41282058

Conflict of interest statement: Declarations Competing Interest Disclosures 
(includes financial disclosures): None


88. Food Res Int. 2025 Dec;222(Pt 2):117741. doi: 10.1016/j.foodres.2025.117741. 
Epub 2025 Oct 28.

Macromineral and trace element concentrations of dairy products and plant-based 
imitations in the UK: implications for population intakes.

Wall RJ(1), Clegg M(2), Zou Y(3), Stergiadis S(4).

Author information:
(1)Department of Animal Sciences, School of Agriculture, Policy and Development, 
University of Reading, PO Box 237, Earley Gate, Reading RG6 6EU, United Kingdom.
(2)School of Food and Nutritional Sciences, University College Cork, Ireland.
(3)Technical Services, Research and Innovation Directorate, University of 
Reading, Whiteknights House, Reading RG6 6UR, United Kingdom.
(4)Department of Animal Sciences, School of Agriculture, Policy and Development, 
University of Reading, PO Box 237, Earley Gate, Reading RG6 6EU, United Kingdom. 
Electronic address:.

UK diets rely on dairy to support mineral adequacy. Sales of plant-based dairy 
imitations (PBDI) are increasing. However, they are not nutritionally equivalent 
to dairy, and there is no UK analytical data on their mineral composition to 
assess potential dietary intake risks. We performed the first large-scale 
analysis of mineral concentrations in the most commonly consumed dairy products 
and PBDI, comparing semi-skimmed milk (n = 40), yoghurt (n = 80) and cheese 
(n = 52) with fortified almond, coconut, oat and soya plant-based beverages 
(PBB; n = 138), coconut and soya plant-based yoghurt imitations (PBYI; n = 49) 
and coconut oil plant-based cheese imitations (PBCI; n = 40). The data were used 
to estimate changes in the UK population's mineral intakes based on current 
consumption. PBDI were highly varied, with mineral concentrations determined by 
fortification and plant base. While fortified PBB contained similar calcium 
concentrations to milk, the fortification of PBYI was inconsistent, resulting in 
differences between soya and coconut-based yoghurt imitations. PBCI contained 
significantly less calcium than cheese despite frequent fortification, likely 
due to nutritional compromises made to preserve meltability. PBDIs were 
inadequately fortified with iodine. In PBCI, despite low levels of declared 
fortification, iodine concentrations suggested that other ingredients in the 
formulation enhanced the overall iodine content. PBDI were lower in potassium 
and zinc. Replacement of dairy with PBDI was more expensive and nutritionally 
detrimental, as adequate intakes of iodine and calcium could become inadequate 
in some age groups, and prevalence of low intakes of magnesium, potassium, and 
zinc could be exacerbated, increasing the risk of clinically significant 
deficiency outcomes.

Copyright © 2025 The Author(s). Published by Elsevier Ltd.. All rights reserved.

DOI: 10.1016/j.foodres.2025.117741
PMID: 41271345 [Indexed for MEDLINE]

Conflict of interest statement: Declaration of competing interest The PhD 
scholarship for RJW was funded by the University of Reading and Arla Foods. Arla 
Foods is a dairy cooperative that produces both dairy and plant-source products. 
Arla Foods did not contribute to conceptualisation, methodology, validation, 
data collection and analysis, or writing or reviewing the manuscript. RJW 
reports a relationship with Joint SACN (Scientific Advisory Committee on 
Nutrition) - COT (Committee on Toxicity of Chemicals in Food, Consumer Products 
and the Environment) Group on Plant-Based Drinks that includes: consulting or 
advisory. SS reports a relationship with Dairy UK, which includes speaking and 
lecture fees; the Dairy Council for Northern Ireland, which includes speaking 
and lecture fees; and the Dutch Dairy Association, which includes speaking and 
lecture fees and travel reimbursement. These organizations did not contribute to 
the funding of this research, or to the conceptualization, methodology, 
validation, data collection and analysis, writing or reviewing the manuscript. 
The other authors declare that they have no known competing financial interests 
or personal relationships that could have appeared to influence the work 
reported in this paper.


89. Food Res Int. 2025 Dec;222(Pt 2):117725. doi: 10.1016/j.foodres.2025.117725. 
Epub 2025 Nov 2.

Fortification of yogurt with Solanum melongena phenolic extract: Physicochemical 
stability, bioaccessibility, bioavailability, and antioxidant effects.

Añibarro-Ortega M(1), Gómez-García R(2), Machado M(2), Pereira A(3), Costa 
EM(2), Pintado M(2), Barros L(3), Pinela J(4).

Author information:
(1)CIMO, LA SusTEC, Instituto Politécnico de Bragança, Campus de Santa Apolónia, 
5300-253, Bragança, Portugal; Nutrition and Bromatology Group, Department of 
Analytical and Food Chemistry, Faculty of Food Science and Technology, 
University of Vigo, Ourense Campus, E-32004 Ourense, Spain. Electronic address: 
.
(2)Centro de Biotecnologia e Química Fina (CBQF)-Laboratório Associado, Escola 
Superior de Biotecnologia, Universidade Católica Portuguesa, Rua Diogo Botelho 
1327, 4169-005 Porto, Portugal.
(3)CIMO, LA SusTEC, Instituto Politécnico de Bragança, Campus de Santa Apolónia, 
5300-253, Bragança, Portugal.
(4)CIMO, LA SusTEC, Instituto Politécnico de Bragança, Campus de Santa Apolónia, 
5300-253, Bragança, Portugal; National Institute for Agricultural and Veterinary 
Research (INIAV, I.P.), Rua dos Lágidos, Lugar da Madalena, 4485-655 Vairão, 
Vila do Conde, Portugal. Electronic address:.

This study investigated the impact of fortifying yogurt with Solanum melongena 
phenolic extract by assessing its physicochemical, biochemical, and antioxidant 
stability, including its behavior during in vitro simulated digestion. 
Fortification influenced the yogurt's pH, titratable acidity, and color, and 
improved its cohesiveness during shelf life. The fortified yogurt exhibited 
higher glucose levels and reduced lactic acid production compared to the 
control. During simulated digestion, the extract modulated alterations in the 
protein and fatty acid profiles of the yogurt. Among phenolic compounds, 
3-O-caffeoylquinic acids exhibited good stability during the oral and gastric 
phases but decreased in the intestinal digest. Some antioxidant effects were 
enhanced during digestion, particularly against lipid peroxidation, but declined 
during storage. Furthermore, phenolic acids were absorbed in an in vitro Caco-2 
cell model, supporting their bioavailability. Overall, yogurt fortification with 
S. melongena extract improved nutritional and functional properties, 
highlighting its potential as a sustainable functional food. Future studies 
should address sensory acceptance, safety aspects, and in vivo health outcomes.

Copyright © 2025 The Author(s). Published by Elsevier Ltd.. All rights reserved.

DOI: 10.1016/j.foodres.2025.117725
PMID: 41271339 [Indexed for MEDLINE]

Conflict of interest statement: Declaration of competing interest The authors 
declare that they have no known competing financial interests or personal 
relationships that could have appeared to influence the work reported in this 
paper.


90. Food Res Int. 2025 Dec;222(Pt 1):117655. doi: 10.1016/j.foodres.2025.117655. 
Epub 2025 Oct 13.

Edible matrices developed by ionic gelation improve the acceptability of iron 
sources for mammals: model weaned pigs.

Villarroel Á(1), Villanueva V(1), Sepúlveda M(1), Pinilla V(1), Guzmán-Pino 
SA(1), Valenzuela C(2).

Author information:
(1)Departamento de Fomento de la Producción Animal, Facultad de Ciencias 
Veterinarias y Pecuarias, Universidad de Chile. Avda. Santa Rosa 11.735, La 
Pintana, Santiago, Chile.
(2)Departamento de Fomento de la Producción Animal, Facultad de Ciencias 
Veterinarias y Pecuarias, Universidad de Chile. Avda. Santa Rosa 11.735, La 
Pintana, Santiago, Chile. Electronic address:.

The objective of this study was to evaluate pigs' preference for feed fortified 
with heme iron (HI) and non-heme iron (NHI), to develop edible matrices (EMs) as 
delivery vehicles for both iron sources, and to assess pigs' feeding behavior 
towards these matrices. Pigs showed equal preference for the control feed and 
HI-fortified feed (both 50 %), but a lower preference for NHI-fortified feed 
(31 %) compared to the control (69 %). The EMs were prepared by ionic gelation, 
using sweet whey and sodium alginate as base materials. Three types of EMs were 
developed: a control without iron (EMC), one with NHI (EM-NHI), and another 
containing a mixture of NHI and HI (EMplus). The EMs consisted mainly of 
non‑nitrogenous extract and protein. Structurally, the EMs were compact, and 
iron was successfully incorporated at concentrations ranging from 0.8 to 2.3 mg 
of iron per g. The type of EMs (EMC, EM-NHI, EMplus) did not significantly 
affect interaction time, consumption, or acceptability in pigs. However, the 
animals showed a clear preference for the whole EMs over their ground form. We 
conclude that encapsulation via ionic gelation enables the development of EMs 
capable of delivering NHI and NHI/HI mixtures, increasing pigs' acceptance of 
NHI. These findings suggest that EMs have potential as a delivery system for 
unpalatable compounds in both animals and potentially humans, pending further 
validation.

Copyright © 2025 Elsevier Ltd. All rights reserved.

DOI: 10.1016/j.foodres.2025.117655
PMID: 41267263 [Indexed for MEDLINE]

Conflict of interest statement: Declaration of competing interest The authors 
declare that they have no known competing financial interests or personal 
relationships that could have appeared to influence the work reported in this 
paper.


91. J Dairy Sci. 2025 Nov 17:S0022-0302(25)00949-X. doi: 10.3168/jds.2025-27386. 
Online ahead of print.

Multidimensional evaluation of the similarity between infant formulas and human 
milk based on macronutrients, fatty acid composition and positional 
distribution, and sn-2 palmitoyl triacylglycerols.

Liu P(1), Zhang W(1), Jiang X(1), Wang S(2), Huang H(2), Cao J(3).

Author information:
(1)Key Laboratory of Food Nutrition and Functional Food of Hainan Province, 
School of Food Science and Engineering, Hainan University, Haikou 570228, China.
(2)The First Affiliated Hospital of Hainan Medical University, Haikou 570102, 
China.
(3)Key Laboratory of Food Nutrition and Functional Food of Hainan Province, 
School of Food Science and Engineering, Hainan University, Haikou 570228, China. 
Electronic address:.

A multidimensional similarity evaluation was conducted between 12 commercial 
infant formulas (6 1,3-dioleoyl-2-palmitoylglycerol [OPO]-fortified samples; 6 
nonfortified samples) and 205 human milk samples from Hainan Island, China, in 
this study. The results showed that 12 brands of infant formulas had high 
similarity scores to human milk at the nutrient composition and energy supply 
(GNE > 88), total fatty acid composition (GFA > 80), and sn-2 palmitoyl 
triacylglycerol (GOPO and GOPL > 82) levels. However, the similarity scores of 
infant formulas at the sn-2 FA level were generally low (Gsn-2 FA < 69). Further 
analysis showed that, in terms of sn-2 FA levels, arachidonic acid and 
docosahexaenoic acid were the main reasons for the low similarity score across 
12 infant formulas. Furthermore, 4-dimensional multivariate statistical analysis 
further visually demonstrated that OPO-fortified infant formulas had higher 
similarities to human milk than nonfortified samples did. In summary, although 
fortifying OPO can improve the overall similarity between infant formulas and 
human milk, there are still significant differences in the sn-2 FA between the 
two. It is suggested that future studies should focus on optimizing the 
positional distribution of FA (especially arachidonic acid and docosahexaenoic 
acid) in infant formulas, rather than merely emphasizing their content, so as to 
achieve a comprehensive and scientific infant formula design.

© 2026, The Authors. Published by Elsevier Inc. on behalf of the American Dairy 
Science Association®. This is an open access article under the CC BY license 
(http://creativecommons.org/licenses/by/4.0/).

DOI: 10.3168/jds.2025-27386
PMID: 41260475


92. Mol Nutr Food Res. 2025 Dec;69(24):e70312. doi: 10.1002/mnfr.70312. Epub 2025 
Nov 17.

Vitamin D(3) Emulsions Stabilized With Pea Proteins and Arabinoxylans as a 
Fortification Strategy for Plant-Based Beverages.

Bravo-Núñez Á(1)(2)(3)(4), Salvia-Trujillo L(2)(3), Ramezani M(2)(3), Reboul 
E(4), Martín-Belloso O(2)(3).

Author information:
(1)University of Valladolid, Valladolid, Spain.
(2)Department of Food Technology, Engineering and Science, University of Lleida, 
Lleida, Spain.
(3)Agrotecnio - CERCA Center, Lleida, Spain.
(4)Aix Marseille Univ, INRAE, INSERM, C2VN, Marseille, France.

This study aimed at evaluating the potential of pea protein isolate (PPI) and 
corn arabinoxylans (CAX) as emulsifiers to develop plant-based drinks enriched 
in vitamin D3. The effect of PPI and CAX as emulsifiers on structural changes of 
emulsions during digestion, lipid digestion kinetics, final lipid digestion, and 
vitamin D3 bioaccessibility, alone or included in an oat milk (OM) drink, was 
assessed. PPI was used alone or in combination with corn arabinoxylans (PPI+CAX) 
and Tween 80 was used as a control emulsifier. All resulting emulsions presented 
small droplet sizes (D(3,2) = 0.57-2.58 µm). The incorporation of the emulsions 
into OM reduced the bridging phenomena observed in emulsions stabilized with 
PPI+CAX, hence presenting a higher colloidal stability. This incorporation also 
prevented droplet coalescence during digestion of emulsions, as shown in the 
microcopy images during oral and gastric digestion in the presence of PPI or 
PPI+CAX as emulsifiers. However, the incorporation of PPI and PPI+CAX emulsions 
into OM resulted in a decreased digestion rate compared to the emulsions alone. 
However, no significant differences were found in the final fatty acid or the 
bioaccessibility of vitamin D3. Taken together, these results underline the 
potential of PPI+CAX in the development of plant-based beverages efficiently 
supplying vitamin D3.

© 2025 The Author(s). Molecular Nutrition & Food Research published by Wiley‐VCH 
GmbH.

DOI: 10.1002/mnfr.70312
PMCID: PMC12700063
PMID: 41251176 [Indexed for MEDLINE]

Conflict of interest statement: The authors declare no conflicts of interest.


93. JPEN J Parenter Enteral Nutr. 2026 Jan;50(1):48-55. doi: 10.1002/jpen.70026. 
Epub 2025 Nov 12.

Association between nonacidified standard and high-protein human milk fortifiers 
and increased weight velocity relative to acidified human milk fortifiers in 
preterm infants: A retrospective cohort study.

Bhatt AN(1), Bingham R(1), Gaillard P(2), Stansfield BK(1).

Author information:
(1)Department of Pediatrics, Division of Neonatology, Children's Hospital of 
Georgia, Augusta, Georgia, USA.
(2)Department of Biostatistics, Data Science, and Epidemiology, Augusta 
University, Augusta, Georgia, USA.

BACKGROUND: Fortification of human milk is necessary for preterm infant growth. 
We evaluated the impact of an acidified human milk fortifier and nonacidified 
human milk fortifiers on growth velocity in preterm infants.
METHODS: This retrospective, single-center study compared preterm infants 
(N = 375) receiving either acidified human milk fortifier (n = 163) or a 
combination of standard-protein and high-protein human milk fortifier (n = 212) 
in a tertiary neonatal intensive care unit. Infants with a birth weight of 
500-1800 g and feeding initiation by day of life 7 were eligible for inclusion. 
The coprimary outcome was weight velocity (g/kg/day) at day of life 28 and 36 
weeks' postmenstrual age. Multivariable linear regression model for the 
coprimary outcomes and growth-related secondary outcomes were adjusted for 
gestational age, birth weight, sex, and race.
RESULTS: Infants receiving standard-protein/high-protein human milk fortifiers 
demonstrated higher weight velocity at day of life 28 (12.09 ± 3.27 vs 
10.92 ± 3.06 g/kg/day, P < 0.001) and 36 weeks' postmenstrual age (13.31 ± 2.23 
vs 12.67 ± 2.28 g/kg/day, P = 0.02) compared with those receiving acidified 
human milk fortifier. Length velocity at day of life 28 was greater in the 
standard-protein/high-protein group (1.09 ± 0.54 vs 0.97 ± 0.43 cm/week, 
P = 0.02) but not at 36 weeks' postmenstrual age (P = 0.06). Human milk exposure 
and incidence of comorbidities were similar between groups.
CONCLUSIONS: Standard- and high-protein human milk fortifiers were associated 
with improved weight and length growth in preterm infants.

© 2025 The Author(s). Journal of Parenteral and Enteral Nutrition published by 
Wiley Periodicals LLC on behalf of American Society for Parenteral and Enteral 
Nutrition.

DOI: 10.1002/jpen.70026
PMCID: PMC12775719
PMID: 41229074 [Indexed for MEDLINE]

Conflict of interest statement: Brian K. Stansfield has received grant funding 
from Mead Johnson Nutrition and is a member of the Mead Johnson Nutrition 
Speaker's Bureau. Mead Johnson Nutrition had no role in the execution of the 
experiments or interpretation of the results.


94. Nutrients. 2025 Nov 4;17(21):3472. doi: 10.3390/nu17213472.

Nutrition Strategies for the Preterm Infant with Bronchopulmonary Dysplasia.

Trindade GS(1), Benincasa BC(1), Procianoy GS(2), Silveira RC(1), Procianoy 
RS(1).

Author information:
(1)Department of Pediatrics, Newborn Section, Hospital de Clínicas de Porto 
Alegre, Universidade Federal do Rio Grande do Sul, Porto Alegre 90010-150, RS, 
Brazil.
(2)Medical School, Universidade Federal de Ciências da Saúde de Porto Alegre, 
Porto Alegre 90050-170, RS, Brazil.

Background/Objectives: Bronchopulmonary dysplasia (BPD) is a common chronic 
complication of prematurity, associated with significant morbidity. Nutrition is 
a key modifiable factor influencing lung growth, repair, and overall 
development. This review summarizes current evidence on nutritional strategies 
for BPD prevention and management. Methods: Narrative review was conducted with 
literature search in major databases using relevant keywords. Results: Early 
nutritional deficits are strongly associated with BPD. Higher early protein 
(3.5-4 g/kg/day) and energy intake (>60 kcal/kg/day in the first week, with 
progressive increases) reduce ventilator dependence. Lipids are essential to 
achieve caloric goals. Fluid restriction may reduce BPD risk but often results 
in undernutrition. Nutrient density, rather than fluid volume, is critical. 
Enteral nutrition, particularly mother's own milk, consistently reduces BPD 
risk, whereas formula feeding is linked to higher BPD incidence. In established 
BPD, nutritional requirements are substantially increased. Feeding is frequently 
complicated by fluid restriction, gastroesophageal reflux, and poor oral 
coordination. Management strategies include higher energy intake (130-150 
kcal/kg/day), adequate protein provision (3.5-4 g/kg/day), and careful use of 
lipid-based energy sources. Fortified human milk or enriched preterm formulas 
are essential, with individualized fortification improving growth. Micronutrient 
support is critical, and long-term follow-up is required, as post-discharge 
growth remains vulnerable and predicts later outcomes. Conclusions: Nutritional 
strategies to mitigate BPD should focus on early optimization of protein and 
energy intake, prioritization of nutrient density and promotion of human milk 
feeding. Targeted micronutrient support, individualized fortification and 
multidisciplinary care are essential to improve pulmonary and neurodevelopmental 
outcomes.

DOI: 10.3390/nu17213472
PMCID: PMC12608204
PMID: 41228544 [Indexed for MEDLINE]

Conflict of interest statement: The authors declare no conflicts of interest.


95. Nutrients. 2025 Oct 28;17(21):3384. doi: 10.3390/nu17213384.

Human Milk Fortification and Necrotizing Enterocolitis in Very Low Birthweight 
Infants: State of Evidence and Systematic Review with Meta-Analysis.

Reyes SM(1), Paul TL(1), Ferry J(2).

Author information:
(1)Rev Bioscience, LLC, Boise, ID 83712, USA.
(2)Pediatrix Medical Group Tampa Regional Practices, St. Joseph's Women's 
Hospital, Tampa, FL 33607, USA.

Background: Necrotizing enterocolitis (NEC) remains a leading cause of morbidity 
and mortality in very low birthweight (VLBW) infants. Human milk feeding and 
standardized feeding protocols are protective, but clinical practice varies, 
particularly in fortifier choice. Whether human milk-derived fortifiers reduce 
NEC risk compared with cow milk-derived fortifiers remains unclear. Methods: We 
conducted a systematic state-of-evidence review and meta-analysis, searching 
PubMed, Web of Science, and Scopus through July 2025. Eligible studies included 
RCTs and observational cohorts of VLBW infants comparing an exclusive human milk 
diet (EHMD) including human milk-derived fortifiers to cow milk-derived diets. 
Two reviewers independently screened and extracted data. Both RCTs and 
observational studies were included to evaluate consistency of effect estimates 
across designs and to account for heterogeneity in control group feeding 
practices. Pooled odds ratios (ORs) with 95% CIs were calculated using a 
Sidik-Jonkman random-effects model. Sensitivity analyses by study design and 
exclusion of infant formula from controls were performed. Results: Twenty 
studies (five RCTs, 15 observational; n = 6794 infants) met inclusion criteria, 
most enrolling infants born ≤1250 g. Compared with cow milk-containing diets, 
EHMD was associated with lower odds of Bell Stage ≥ 2 NEC (OR: 0.59; 95% CI: 
0.42, 0.81; p < 0.001; n = 4625) and surgical NEC (OR: 0.43; 95% CI: 0.32, 0.58; 
p < 0.0001; n = 4754). In direct comparisons of fortifier type with a base diet 
of human milk, estimates suggested lower odds of Bell Stage ≥ 2 NEC by 35% (OR: 
0.65; 95% CI: 0.44, 0.97; p = 0.03, n = 2102) and surgical NEC by 49% (OR: 0.51; 
95% CI: 0.26, 0.98; p = 0.04; n = 1659) with human milk-derived fortifiers. 
Effect estimates were generally consistent across study designs, although 
precision and statistical significance varied. Conclusions: EHMD with human 
milk-derived fortifiers was associated with lower odds of medical and surgical 
NEC in VLBW infants, with most evidence from infants born ≤1250 g, reflecting 
current clinical use in the highest-risk population. Although the number and 
sample sizes of RCTs remain limited, the consistency of effect estimates across 
both RCTs and observational studies, together with significance of pooled 
analyses, strengthens confidence in these findings. Pragmatic and registry-based 
studies using standardized fortification protocols may provide the most 
efficient pathway to strengthen the evidence base.

DOI: 10.3390/nu17213384
PMCID: PMC12609769
PMID: 41228457 [Indexed for MEDLINE]

Conflict of interest statement: Support for this work was as follows: S.M.R. 
serves as a consultant for Prolacta Bioscience and received financial support to 
conduct the research and write the manuscript. T.L.P. serves as a consultant for 
Rev Bioscience and received financial support to help conduct the research. J.F. 
does not have any conflicts of interest to disclose.


96. Food Sci Nutr. 2025 Nov 10;13(11):e70828. doi: 10.1002/fsn3.70828. eCollection 
2025 Nov.

Evaluation of Mineral Content in Preterm Human Milk and Infant Formulas in 
Qatar: Assessing Compliance With Dietary Recommendations and Label Accuracy.

Haji Z(1), Mohammed R(1), Fardan H(1), Vatankhah N(1), Ismail S(1), Bassil M(1), 
Shi Z(1), Ibrahim MIA(2), Attieh G(1), Al-Dewik N(3)(4)(5)(6), Karam L(1).

Author information:
(1)Department of Nutrition Sciences, College of Health Sciences, QU Health Qatar 
University Doha Qatar.
(2)Central Laboratories Unit Qatar University Doha Qatar.
(3)Department of Neonatology, Neonatal Intensive Care Unit, Newborn Screening 
Unit Women's Wellness and Research Center, Hamad Medical Corporation Doha Qatar.
(4)Genomics and Precision Medicine (GPM), College of Health & Life Science 
(CHLS) Hamad Bin Khalifa University (HBKU) Doha Qatar.
(5)Department of Research Women's Wellness and Research Center, Hamad Medical 
Corporation Doha Qatar.
(6)Department of Translational & Precision Medicine Research Facility Women's 
Wellness & Research Center (WWRC), Hamad Medical Corporation (HMC) Doha Qatar.

Preterm infants have higher energy and nutrient needs compared to term infants, 
with human milk recommended as the primary feeding choice and infant formula as 
the secondary option. This study aimed to evaluate the concentration of 
essential trace minerals (manganese, copper, iron, and zinc) in preterm human 
milk and infant formulas in Qatar, and assess their nutrition label accuracy and 
compliance with nutritional requirements. Mineral analysis was performed using 
Inductively Coupled Plasma Mass Spectrometry (ICP-MS). Samples included 50 
liquid human milk samples from lactating mothers of preterm infants, 42 powder 
infant formulas from local markets and pharmacies, and 10 water samples commonly 
used in Qatar. All human milk and infant formulas were below the recommended 
zinc and iron ranges as per the American Society for Parenteral and Enteral 
Nutrition (ASPEN). Additionally, 96% (48/50) of human milk and 95% (40/42) of 
infant formulas were below the recommended copper range. Furthermore, 34% 
(17/50) of human milk samples for manganese were below the recommended range, 
whereas 86% (36/42) of infant formulas exceeded it. Water samples showed mineral 
levels below detection limits, and thus had no contribution to mineral levels in 
reconstituted formulas. Significant differences were found between label and 
laboratory-tested values for copper (p = 0.0039) and zinc (p = 0.0000), with 
label values higher than laboratory results. No significant differences were 
observed for manganese (p = 0.7564) or iron (p = 0.1966). Reconstituted formulas 
had significantly higher manganese, zinc, and iron laboratory levels (p < 0.001) 
than human milk, whereas copper showed no significant difference (p = 0.324). 
These findings highlight mineral imbalances in both human milk and infant 
formulas for preterm infants, demonstrating the need for human milk fortifiers, 
improved nutrient formulation, accurate labeling, and further research to ensure 
optimal health outcomes.

© 2025 The Author(s). Food Science & Nutrition published by Wiley Periodicals 
LLC.

DOI: 10.1002/fsn3.70828
PMCID: PMC12598410
PMID: 41221094

Conflict of interest statement: The authors declare no conflicts of interest.


97. Sci Rep. 2025 Nov 6;15(1):38939. doi: 10.1038/s41598-025-22850-1.

Vitamin D modulates gene expression in four major muscle tissues in Atlantic 
salmon.

Gorman CE(1), Egan F(2), Alarcón-López FJ(3), Jakobsen J(4), McGinnity P(5), 
Hulsey CD(6).

Author information:
(1)School of Biology and Environmental Science, University College Dublin, 
Belfield, Dublin, Ireland..
(2)Marine Institute, Newport, Ireland.
(3)Department of Biology and Geology, University of Almería, 04120, Almería, 
Spain.
(4)National Food Institute, Technical University of Denmark, 2800, Lyngby, 
Denmark.
(5)School of Biological, Earth and Environmental Sciences, University College 
Cork, Cork, Ireland.
(6)School of Biology and Environmental Science, University College Dublin, 
Belfield, Dublin, Ireland.

Vitamin D is essential to muscle health but could affect different muscle 
tissues in distinct ways. In humans, vitamin D is primarily synthesized in the 
skin via sun exposure but can also be obtained through ingesting foods that are 
fortified (e.g., milk) or naturally high in vitamin D (e.g., salmon). Like 
humans, salmon vitamin D content can vary considerably and can be altered 
through dietary consumption of vitamin D. We experimentally determined how 
vitamin D manipulation alters gene expression in four salmon muscle tissues: 
skeletal, craniofacial, smooth, and cardiac. Vitamin D induced changes in 
expression were exceptionally profound in the heart, where there were also 
several gene duplicates expressed unidirectionally. Five genes were 
differentially expressed in multiple muscles and many genes impacted by our 
vitamin D treatments in salmon show extensive evolutionary conservation as they 
are influenced by vitamin D titers in mammals, including humans. Salmon could 
provide a powerful model for understanding how variation in vitamin D impacts 
all vertebrates.
SUPPLEMENTARY INFORMATION: The online version contains supplementary material 
available at 10.1038/s41598-025-22850-1.

DOI: 10.1038/s41598-025-22850-1
PMCID: PMC12592442
PMID: 41198894

Conflict of interest statement: Declarations. Competing interests: The authors 
declare no competing interests.


98. Am J Clin Nutr. 2025 Nov;122(5):1317-1325. doi: 10.1016/j.ajcnut.2025.07.031. 
Epub 2025 Sep 18.

A cross-sectional study of macronutrients, vitamins, minerals, and bioactive 
components in human milk of approved milk bank donors from low-, middle-, and 
high-income countries.

Perrin MT(1), Mansen K(2), Israel-Ballard K(2), Bode L(3), Hampel D(4), 
Shahab-Ferdows S(5), Allen LH(4), Maggio FC(6), Njuguna EM(7), Tran HT(8), 
Wesolowska A(9).

Author information:
(1)Department of Nutrition, University of North Carolina, Greensboro, NC, United 
States. Electronic address:.
(2)Primary Health Care Department, Integrated Maternal and Child Health and 
Development Team, PATH, Seattle, WA, United States.
(3)Department of Pediatrics, Larsson-Rosenquist Foundation Mother-Milk-Infant 
Center of Research Excellence (MOMI CORE) and the Human Milk Institute (HMI), 
University of California, San Diego, La Jolla, CA ,United States.
(4)Institute for Global Nutrition, Department of Nutrition, University of 
California, Davis, CA, United States; United States Department of 
Agriculture/Agriculture Research Service, Western Human Nutrition Research 
Center, Davis, CA, United States.
(5)United States Department of Agriculture/Agriculture Research Service, Western 
Human Nutrition Research Center, Davis, CA, United States.
(6)Department of Neonatology Service, Complejo Asistencial Dr. Sótero del Rio, 
Santiago, Chile.
(7)Primary Health Care Department, Integrated Maternal and Child Health and 
Development Team, PATH, Seattle, WA, United States; Department of Pediatrics, 
Pumwani Maternity and Referral Hospital, Nairobi, Kenya.
(8)Neonatal Unit and Human Milk Bank, Da Nang Hospital for Women and Children, 
Da Nang, Vietnam; Department of Pediatrics, School of Medicine and Pharmacy, The 
University of Da Nang, Da Nang, Vietnam.
(9)Laboratory of Human Milk and Lactation Research at Regional Human Milk Bank 
in Holy Family Hospital, Department of Medical Biology, Medical University of 
Warsaw, Warsaw, Poland.

BACKGROUND: Currently, clinical care of preterm infants is limited by a dearth 
of information on the nutritional and bioactive composition of donor human milk 
(DHM).
OBJECTIVES: The primary objective is to assess and compare the macronutrients, 
vitamins, minerals, human milk oligosaccharides (HMOs), and antimicrobial 
proteins in human milk from approved milk banks in a variety of low-, middle-, 
and high-income settings. The secondary objective is to explore factors that 
influence composition, including gestational stage (preterm compared with term), 
lactation stage, maternal age, lifetime donation volume, and storage duration.
METHODS: We conducted a cross-sectional study of human milk collected 
systematically from unique, approved milk bank donors (n = 600) in Chile, Kenya, 
Poland, the United States, and Vietnam. True protein, total fat, lactose, HMOs, 
vitamins, minerals, and antimicrobial proteins were assessed. Analysis of 
variance and Tukey's tests were used to evaluate normally distributed data; 
Kruskal-Wallis and Dunn's tests were used for nonparametric data. Multiple 
regression was used to explore factors associated with milk composition.
RESULTS: The only site with predominantly preterm donors was Kenya (26/50). 
Kenya also had an early donation model (mean lactation stage 1.6 ± 1.1 wk), 
whereas the remaining sites had a mature donation model (mean lactation stage 
ranged 14.2-21.4 wk). Most nutrients differed significantly (P < 0.05) between 
geographies. For instance, true protein in Kenya was 1.3 ± 0.3 g/dL compared 
with 0.8 ± 0.2 at all United States banks. Many minerals were higher in the 
early donation model whereas lactose and several water-soluble vitamins were 
higher in the mature donation model. Although the lactation stage typically had 
the most predictive value, most nutrient variation (>60%) remained unexplained.
CONCLUSIONS: Clinical protocols for feeding DHM to preterm infants, including 
setting-specific fortifier products, may need to take into consideration the 
local donor milk banking model. Multidonor pooling warrants further 
investigation as a tool for milk banks to ensure minimum nutrient standards for 
DHM.

Copyright © 2025 The Author(s). Published by Elsevier Inc. All rights reserved.

DOI: 10.1016/j.ajcnut.2025.07.031
PMCID: PMC12614297
PMID: 41193033 [Indexed for MEDLINE]

Conflict of interest statement: Conflict of interest MTP reports financial 
support was provided by National Institute of Child Health and Human 
Development. LB reports a relationship with Family Larsson-Rosenquist Foundation 
that includes: funding grants. MTP serves on the Scientific Advisory Board for 
the Human Milk Banking Association of North America, and on the WHO Guidelines 
Development Group for Donor Milk Banking in unpaid capacities. KI-B and KM serve 
as coordinators for the WHO Standards on Human Milk Banking. All other authors 
report no conflicts of interest.


99. Pediatrics. 2025 Dec 1;156(6):e2024069320. doi: 10.1542/peds.2024-069320.

Improving and Sustaining VLBW Infant Growth Through a Quality Collaborative in 
California.

Morris M(1), Cui X(1)(2), Breault C(1)(2), Kim JH(3), Hetherton MC(4), Quinn 
J(5), Profit J(1)(2), Payton KSE(1)(6).

Author information:
(1)California Perinatal Quality Care Collaborative, Stanford, California.
(2)Division of Neonatal and Developmental Medicine, Department of Pediatrics, 
Stanford University School of Medicine, Stanford, California.
(3)Department of Pediatrics, Perinatal Institute, Cincinnati Children's Hospital 
Medical Center, University of Cincinnati College of Medicine, Cincinnati, Ohio.
(4)UCSF Benioff Children's Hospital, San Francisco, California.
(5)NorthBay Medical Center, Fairfield, California.
(6)Division of Neonatology, Department of Pediatrics, Cedars-Sinai Medical 
Center, Los Angeles, California.

Comment in
    Pediatrics. 2025 Dec 1;156(6):e2025073798. doi: 10.1542/peds.2025-073798.

INTRODUCTION: Very low birth weight (VLBW) infants experience inadequate 
postnatal growth, which may be associated with poor neurodevelopmental outcomes. 
This statewide quality improvement collaborative aimed to improve VLBW infant 
growth.
METHODS: The collaborative was conducted from October 2018 to March 2020. The 
aim was to reduce the primary outcome measure of proportion of weight <10th 
percentile at discharge among VLBW infants by 20% in 1 year. The secondary 
outcome measure was a z-score deficit ≥0.8 from birth. Process measures were 
early fortification, nutrition rounds, and feeding guideline compliance. 
Balancing measures were necrotizing enterocolitis and human milk feeding at 
discharge. Measures were analyzed with statistical process control charts. 
Outcome measures were compared among participant and nonparticipant neonatal 
intensive care units (NICUs) for a total of 6 years including 2 years after the 
collaborative.
RESULTS: 22 NICUs participated in the collaborative. The 78-month analysis 
included 7856 VLBW infants. The participant group reduced growth restriction at 
discharge (45% to 37.6%) and z-score deficit ≥0.8 (44.4% to 33.3%). 
Postcollaborative improvement was sustained for 24 months. The 114 
nonparticipant NICUs did not reduce rates of growth outcome measures. Several 
process measures showed sustained improvement. Balancing measures were unchanged 
during the collaborative period. However, post hoc analysis showed a higher 
necrotizing enterocolitis rate when 2 years after the collaborative were 
included.
CONCLUSION: Quality improvement collaborative NICUs improved nutrition processes 
and VLBW growth. Improvement was sustained for 24 months after the 
collaborative. Nonparticipant NICUs did not show similar improvement. 
Implementing NICU nutrition processes may lead to sustained improvements in VLBW 
infant growth.

Copyright © 2025 by the American Academy of Pediatrics.

DOI: 10.1542/peds.2024-069320
PMID: 41192624 [Indexed for MEDLINE]


100. Plant Foods Hum Nutr. 2025 Nov 5;80(4):186. doi: 10.1007/s11130-025-01429-1.

Utilization of Grape Pomace as A Value-added Phenolic Component for Oat Milk 
Production.

Hassan MZ(1)(2), Abidi KH(3), Kovaleva EG(3).

Author information:
(1)Research, Educational and Innovation Center of Chemical and Pharmaceutical 
Technologies, Institute of Chemical Technology, Ural Federal University, 
Ekaterinburg, Russia..
(2)Bangladesh Livestock Research Institute, Savar, Dhaka, 1341, Bangladesh. 
.
(3)Research, Educational and Innovation Center of Chemical and Pharmaceutical 
Technologies, Institute of Chemical Technology, Ural Federal University, 
Ekaterinburg, Russia.

Grape pomace (GP), a wine industry byproduct rich in dietary fiber and 
polyphenols, was aimed to enhance the nutritional, phytochemical, and sensory 
qualities of oat milk. Oat milk, a sustainable dairy alternative rich in 
β-glucan and minerals, but its low phenolic and antioxidant content limits its 
functional potentiality. This work incorporated GP extract into oat milk to 
improve its bioactive profile, stability, and consumer appeal, meeting the 
demand for plant-based functional beverages. The study assessed the effects of 
GP fortification on phenolic composition, antioxidant activity, and 
acceptability. Oat milk exhibited stable emulsion properties at pH 6.77 ± 0.12 
and titratable acidity 0.43 ± 0.21%. GP supplementation significantly increased 
bioactive potential: total phenolic content rose from 35.08 ± 2.30 to 
101.43 ± 25 mg GAE/g DW, total flavonoids from 69.56 ± 1.5 to 180.11 ± 18 mg 
QE/g DW, and antioxidant activity to 48 ± 8 mM Trolox/g DW. Anthocyanin content 
also increased, highlighting GP's role in enhancing phenolic delivery. Sensory 
evaluation showed that 0.2% GP-fortified oat milk achieved the highest 
acceptability in flavor, color, aroma, mouthfeel, and overall preference. Higher 
GP levels (0.3-0.5%) intensified flavor and color but reduced consumer 
acceptance. Overall, GP fortification, particularly at 0.2%, improves oat milk's 
nutritional and functional properties as a value-added ingredient for functional 
beverages within a circular economy.

© 2025. The Author(s), under exclusive licence to Springer Science+Business 
Media, LLC, part of Springer Nature.

DOI: 10.1007/s11130-025-01429-1
PMID: 41191155 [Indexed for MEDLINE]

Conflict of interest statement: Declarations. Ethical Approval: Not applicable 
due to no animal experiments was done. Competing Interests: The authors declare 
no competing interests.


101. Food Res Int. 2025 Dec;221(Pt 4):117594. doi: 10.1016/j.foodres.2025.117594. 
Epub 2025 Sep 25.

Encapsulation of hydrophobic functional components in yogurt: technology, 
product characteristics and health benefits.

Meng G(1), Zhang M(1), Wu Z(1), Zeng X(1), Zhao G(2), Bao W(3), Zhou C(4), Pan 
D(1), Du Q(5).

Author information:
(1)State Key Laboratory for Managing Biotic and Chemical Threats to the Quality 
and Safety of Agro-products, Ningbo University, Ningbo, China; Key Laboratory 
for Food Microbiology and Nutrition of Zhejiang Province, College of Food 
Science and Engineering, Ningbo University, Ningbo 315211, China; 
Zhejiang-Malaysia Joint Research Laboratory for Agricultural Product Processing 
and Nutrition, Ningbo University, Ningbo, China.
(2)Hangzhou New Hope Shuangfeng Dairy Products Co., Ltd., Hangzhou, China.
(3)Jinhua Yinhe Biological Technology Co., Ltd., Jinhua, China.
(4)State Key Laboratory for Managing Biotic and Chemical Threats to the Quality 
and Safety of Agro-products, Ningbo University, Ningbo, China; Key Laboratory 
for Food Microbiology and Nutrition of Zhejiang Province, College of Food 
Science and Engineering, Ningbo University, Ningbo 315211, China; 
Zhejiang-Malaysia Joint Research Laboratory for Agricultural Product Processing 
and Nutrition, Ningbo University, Ningbo, China. Electronic address: 
.
(5)State Key Laboratory for Managing Biotic and Chemical Threats to the Quality 
and Safety of Agro-products, Ningbo University, Ningbo, China; Key Laboratory 
for Food Microbiology and Nutrition of Zhejiang Province, College of Food 
Science and Engineering, Ningbo University, Ningbo 315211, China; 
Zhejiang-Malaysia Joint Research Laboratory for Agricultural Product Processing 
and Nutrition, Ningbo University, Ningbo, China. Electronic address: 
.

Yogurt, a widely consumed functional food, presents a physical incompatibility 
between its hydrophilic matrix and hydrophobic functional components, resulting 
in uneven dispersion, oxidative degradation and reduced bioavailability. To 
address these challenges, encapsulation systems such as emulsions, liposomes and 
micro- or nanoparticles are commonly employed to enhance the stability and 
efficacy of hydrophobic bioactives in fortified yogurt. The stability of 
encapsulation systems is governed by microscopic parameters, including 
interfacial layer, particle coating and particle size. Strategies such as 
promoting electrostatic repulsion to prevent casein micelle aggregation, filling 
voids within the casein network with particles and optimising droplet size 
notably influence the physicochemical properties of yogurt and enhance the 
delivery efficiency of functional components. Yogurt fortified with hydrophobic 
functional components benefits from a physical barrier formed by its 
encapsulation structure, which delays the oxidative degradation of active 
ingredients. This barrier enhances delivery efficiency and bioavailability in 
the gastrointestinal tract through controlled-release mechanisms. Such 
improvements contribute to positive health outcomes, including cardiovascular 
support, antioxidant defence, anti-inflammatory activity and bone health, and 
may help reduce the risk of chronic diseases. Future in vivo studies, 
particularly those involving human clinical trials, are needed to inform the 
development of functional yogurt products and to support the application of 
related delivery systems across other dairy matrices.

Copyright © 2025 Elsevier Ltd. All rights reserved.

DOI: 10.1016/j.foodres.2025.117594
PMID: 41185344 [Indexed for MEDLINE]

Conflict of interest statement: Declaration of competing interest The authors 
declare that they have no known competing financial interests or personal 
relationships that could have appeared to influence the work reported in this 
paper.


102. J Cachexia Sarcopenia Muscle. 2025 Dec;16(6):e70080. doi: 10.1002/jcsm.70080.

Postinactivity Exercise Training Improves Sarcopenia Traits in 40-60-Year-Old 
Women Regardless of Fortified Milk Supplementation.

Trezise J(1), Lima RM(2)(3), Poppitt SD(4), Fanning AC(5), Devine A(6), 
Blazevich AJ(7).

Author information:
(1)Human Performance Centre, Southern Institute of Technology, Te Pūkenga, 
Southland, New Zealand.
(2)Department of Nutrition, University of Brasília, Brasília, Brazil.
(3)Department of Physical Education, University of Brasília, Brasília, Brazil.
(4)Human Nutrition Unit, School of Biological Sciences, The University of 
Auckland, Auckland, New Zealand.
(5)Fonterra Research and Development Centre, Palmerston North, New Zealand.
(6)Nutrition and Health Innovation Research Institute, School of Medical and 
Health Sciences, Joondalup, WA, Australia.
(7)Centre for Precision Health, School of Medical and Health Sciences, Edith 
Cowan University, Joondalup, WA, Australia.

BACKGROUND: The term sarcopenia was introduced to describe the age-related 
decline in muscle mass, but current definitions also include measures of muscle 
strength and function. Menopause increases sarcopenia risk and may exacerbate 
the adverse effects of physical inactivity. Exercise training is a potent 
stimulus to restore muscle health, and nutritional supplementation can further 
improve the outcomes. The purposes of this study were to examine the changes in 
a set of sarcopenia-related phenotypes induced by a 14-day step-reduction period 
followed by a 12-week exercise training programme, with or without fortified 
milk supplementation in healthy 40-60-year-old females, and to determine whether 
menopausal status interacted with these changes.
METHODS: In this double-blind, placebo-controlled randomized trial, females aged 
40-60 years were evaluated before and after 2 weeks of reduced activity 
monitored through pedometer and after a subsequent 12-week exercise + nutrition 
programme with ingestion of a fortified milk product (FMP) or placebo. Muscle 
volume (dual-energy X-ray absorptiometry [DXA] and peripheral quantitative 
computed tomography), handgrip (hydraulic handheld dynamometer), knee extensor 
and plantar flexor strength (isokinetic dynamometer) and a variety of physical 
function measures were assessed at all timepoints.
RESULTS: Eighty-three self-reported healthy females (50.7 ± 5.3 years; 52 
postmenopausal) completed the reduced-activity period, and 67 completed the 
subsequent exercise training + nutrition phase. At baseline, participants 
averaged 8323 ± 3077 daily steps and then decreased to 1876 ± 729 during the 
reduced-activity period. Mean sarcopenia outcomes declined after 2 weeks of 
activity restriction, with significant changes (p < 0.05) in shank muscle 
cross-sectional area (CSA) (67.7 ± 9.9 to 66.5 ± 9.9 cm2), handgrip strength 
(25.3 ± 5.0 to 24.0 ± 5.3 kg), knee extensor peak torque (134.7 ± 36.2 to 
122.7 ± 34.5 Nm) and stair ascending time (3.6 ± 0.6 to 3.7 ± 0.6 s) and power 
(367.6 ± 67.6 to 356.7 ± 67.7 W), with no significant time × menopause 
interactions (p > 0.05 for all variables). All muscle mass, strength and 
function outcomes were not only improved after exercise training + nutrition but 
also significantly increased compared to preintervention baseline (all 
p < 0.05). No training × menopause or training × supplementation interactions 
were observed for any variable (both p > 0.05).
CONCLUSIONS: Two weeks of step reduction negatively affected muscle mass, 
strength and physical function in 40-60-year-old females. A 12-week training 
programme including strength exercises and dietary supplementation not only 
recovered muscular health but also promoted improvements above the baseline 
before the physical activity restriction. Menopause status did not influence 
changes in response to step reduction or exercise, and the addition of a 
fortified milk product during the training period did not influence the induced 
adaptations.

© 2025 The Author(s). Journal of Cachexia, Sarcopenia and Muscle published by 
Wiley Periodicals LLC.

DOI: 10.1002/jcsm.70080
PMCID: PMC12582911
PMID: 41184220 [Indexed for MEDLINE]

Conflict of interest statement: S.D.P. held the Fonterra Chair in Human 
Nutrition at the University of Auckland, New Zealand.


103. Sci Rep. 2025 Oct 31;15(1):38102. doi: 10.1038/s41598-025-22091-2.

Enhancing yogurt health benefits with moringa and black seed oil nanoemulsions 
to improve fatty acids and microbial viability.

Abd-Rabou HS(#)(1), Soliman TN(#)(2), Galal Darwish AM(3)(4), Awad S(5), Allam 
MG(6).

Author information:
(1)Food Technology Department, Arid Lands Cultivation Research Institute, The 
City of Scientific Research and Technological Application (SRTA- City), 
Alexandria, Egypt.
(2)Dairy Department, Food Industries and Nutrition Research Institute, National 
Research Centre, 12622, Dokki, Giza, Egypt..
(3)Food Technology Department, Arid Lands Cultivation Research Institute, The 
City of Scientific Research and Technological Application (SRTA- City), 
Alexandria, Egypt..
(4)Food Industry Technology Program, Faculty of Industrial and Energy 
Technology, Borg Al Arab Technological University (BATU), Alexandria, Egypt. 
.
(5)Department of Dairy Science, Faculty of Agriculture, Alexandria University, 
El-Shatby, Alexandria, Egypt.
(6)Food Science Department, Faculty of Agriculture, Saba Basha, Alexandria 
University, Alexandria, Egypt.
(#)Contributed equally

The increasing consumer demand for natural functional foods has prompted the 
development of fortified dairy products with improved nutritional and health 
benefits. This study designed and evaluated yoghurts fortified with cold-pressed 
moringa seed oil (MSO) and black seed oil (BSO) at 1.5 g /L, as well as their 
water-based nanoemulsions at 3 g Nanoemulsion powder/L, compared to a plain 
control. Phenolic profiles characterized by HPLC-MS revealed high levels of 
ellagic acid (23.5 mg/100 g), rutin (18.2 mg/100 g), and chlorogenic acid 
(21.4 mg/100 g) in MSO, and chlorogenic acid (19.8 mg/100 g), apigenin 
(16.7 mg/100 g), and naringenin (14.3 mg/100 g) in BSO. Nanoemulsions with 5% 
oil showed droplet sizes of 69.1 nm (MSO) and 38.1 nm (BSO) and zeta potentials 
above - 30 mV, confirming good colloidal stability over 7 days. Cytotoxicity 
assays indicated a safe dose up to 80 µg/mL. Yoghurt fortified with 
nanoemulsions exhibited a significant increase in total solids (+ 12%), 
unsaturated fatty acids (notably omega-3 increased by 25%), and antioxidant 
capacity (DPPH radical scavenging improved by 30%) while reducing acidity and 
syneresis relative to the control. Lactic acid bacteria viability remained 
unaffected. Sensory evaluation showed improved color, texture, and overall 
acceptance for yoghurts with 1.5 g nanoemulsion addition per liter. These 
findings demonstrate that incorporation of MSO and BSO nanoemulsions at this 
level effectively enhances yoghurt's nutritional and functional properties 
without compromising microbial or sensory quality.

© 2025. The Author(s).

DOI: 10.1038/s41598-025-22091-2
PMCID: PMC12579260
PMID: 41173981 [Indexed for MEDLINE]

Conflict of interest statement: Competing interests: The authors declare no 
competing interests. Ethical approval: The sensory evaluation has been reviewed 
and approved by the Medical Research Ethics Committee (MREC) of the National 
Research Centre. The MREC’s review ensures that all aspects of this research 
meet the highest ethical standards, as guided by the principles of the 
Declaration of Helsinki and Good Clinical Practice (GCP), which aim to protect 
all research participants’ rights, safety, and well-being. The approval number 
for this study is 13050205-1.


104. J Food Sci. 2025 Nov;90(11):e70618. doi: 10.1111/1750-3841.70618.

Mechanisms of Calcium Precipitation in Cheese-Enriched Meat Products: A Review 
of Factors, Challenges, and Potential Solutions.

Okedoyin D(1), Boyle E(1).

Author information:
(1)Department of Animal Science and Industry, Kansas State University, 
Manhattan, Kansas, USA.

Calcium, an essential nutrient, plays a crucial role in various physiological 
functions in humans and other organisms. Diets of poor quality can lead to 
deficiencies in several essential nutrients, with calcium being one of the most 
commonly deficient. This review explores the phenomenon of calcium precipitation 
in cheese-enriched meat products, a pressing challenge in the development of 
stable and nutritionally fortified food matrices. The review explains the 
physicochemical, structural, and environmental interactions underlying calcium 
redistribution and crystal formation when cheese is incorporated into meat 
systems. The core drivers, such as pH shifts, thermal treatments, 
protein-calcium binding, and phosphate and chelator additives' roles, are 
discussed in detail. This review identifies critical control points and presents 
emerging technological interventions, from encapsulated calcium systems to 
modified atmosphere packaging (MAP). Synthesizing current knowledge and 
outlining future directions calls for precision formulation approaches to 
transform calcium precipitation for innovation in functional food design.

© 2025 Institute of Food Technologists.

DOI: 10.1111/1750-3841.70618
PMID: 41164913 [Indexed for MEDLINE]


105. Food Funct. 2025 Nov 10;16(22):8720-8763. doi: 10.1039/d5fo01106j.

The impact of (poly)phenol-rich foods and extracts on flow-mediated dilation 
(FMD): a narrative review.

Murphy BÓ(1), Neill HR(1), Magee N(1), Rosbotham EJ(1), Pourshahidi LK(1), 
Richardson P(2), Pouchieu C(3), Gaudout D(3), Ray S(1)(4), Bresciani L(5), Del 
Rio D(5), Rodriguez-Mateos A(6), Crozier A(7)(8), Gill CIR(1).

Author information:
(1)Nutrition Innovation Centre for Food and Health (NICHE), School of Biomedical 
Sciences, Ulster University, Coleraine, UK..
(2)Centre for Optometry and Vision Science Research, School of Biomedical 
Sciences, Ulster University, Coleraine, UK.
(3)Activ'Inside, Beychac-et-Caillau, France.
(4)NNEdPro Global Institute for Food Nutrition and Health, Cambridge, UK.
(5)Human Nutrition Unit, Department of Food and Drug, University of Parma, 
Parma, Italy.
(6)Department of Nutritional Sciences, School of Life Course and Population 
Health Sciences, Faculty of Life Sciences and Medicine, King's College London, 
UK.
(7)Department of Chemistry, King Saud University Riyadh, Saudi Arabia.
(8)School of Medicine, Dentistry and Nursing, University of Glasgow, Glasgow, 
UK.

Cardiovascular disease (CVD) remains the leading cause of morbidity and 
mortality worldwide, with endothelial dysfunction as a key precursor. 
Flow-mediated dilation (FMD), the gold-standard measure of endothelial function, 
is improved by (poly)phenol-rich foods and extracts, with increases of 1% FMD 
representing 13% reduced cardiovascular risk. This narrative review aims to 
evaluate the efficacy of various (poly)phenol-rich foods and extracts on 
endothelial function as measured by flow-mediated dilation (FMD) and assesses 
the feasibility of a food-first approach. Literature was systematically searched 
from databases including PubMed and Web of Science, focusing on human clinical 
trials. While all (poly)phenol-rich food groups demonstrate variable effects, 
berries (0.9-2.6%), cocoa (0.7-5.9%), and tea (1.2-4.8%) have the most robust 
evidence, consistently improving FMD, with chronic intake sustaining benefits. A 
large variance (0.8-8.7%) was observed with grape-derived (poly)phenols, making 
their effects difficult to substantiate without detailed compositional or 
metabolomic data; however, a few key studies highlight their potential. Citrus 
polyphenols also exhibit variable FMD responses (0.2-7.2%). However, strong 
mechanistic evidence supports their role in vascular health and nitric oxide 
(NO) bioavailability. Coffee exhibits a variable response, initially impairing 
FMD, likely due to caffeine, before later improving endothelial function as 
phenolic metabolites increase. Although estimated (poly)phenol intake in Western 
populations is high (∼1000-1200 mg day-1), it is primarily derived from tea, 
coffee, and cocoa, limiting exposure to diverse bioactive compounds. Moreover, 
the food matrix significantly influences bioavailability, with co-consumed 
components such as milk or sugar attenuating FMD responses. Interestingly, 
fortification and enrichment maintain bioactivity and may optimize intake, 
ensuring consistent and diverse delivery. Future research should refine dietary 
guidelines, establish intake thresholds, and explore fortification strategies to 
maximize cardiovascular benefits while considering dose-response relationships 
and long-term efficacy.

DOI: 10.1039/d5fo01106j
PMID: 41140262 [Indexed for MEDLINE]


106. J Med Econ. 2025 Dec;28(1):1899-1909. doi: 10.1080/13696998.2025.2578101. Epub 
2025 Nov 1.

Cost-utility analysis of an exclusive human milk diet for very low birthweight 
infants in Japan.

Igarashi A(1)(2), Reyes SM(3), Mizuno K(4).

Author information:
(1)Department of Health Data Science, Graduate School of Data Science, Yokohama 
City University, Yokohama, Japan.
(2)Health Outcome Research Institute, Ichikawa, Japan.
(3)Rev Bioscience, LLC, Boise, ID, USA.
(4)Department of Pediatrics, Showa University School of Medicine, Tokyo, Japan.

OBJECTIVE: To assess the cost-utility of an exclusive human milk diet (EHMD) 
compared to conventional nutrition for very low birthweight (VLBW, <1500 g) 
infants from Japan's healthcare payer perspective.
METHODS: We conducted a cost-utility analysis using used a Markov 
state-transition model built with TreeAge. Effectiveness was assessed using 
Quality-Adjusted Life Years (QALYs). The primary outcome was the incremental 
cost-effectiveness ratio (ICER) expressed as cost per QALY gained. Using 2024 
prices, EHMD cost was JPY 2,000,000 (USD 13,220), varying up to JPY 4,000,000 
(USD 26,440) in sensitivity analyses. Deterministic and probabilistic 
sensitivity analyses were performed.
RESULTS: Base case analysis showed an ICER of JPY 2,949,278 (USD 19,495) per 
QALY gained, well below the hypothetical willingness-to-pay threshold value of 
JPY5,000,000 to JPY7,500,000 (USD 33,050 to USD 49,575). Sensitivity analysis 
indicated EHMD remained cost-effective up to JPY 4,750,000 (USD 31,398) per 
course against a revised JPY 7,500,000 (USD 49,575) per QALY threshold.
CONCLUSION: This cost-utility analysis demonstrated that an EHMD may be a 
cost-effective option for VLBW infants in Japan at 2024 costs, and provides 
evidence to inform decision makers considering its adoption.

DOI: 10.1080/13696998.2025.2578101
PMID: 41117636 [Indexed for MEDLINE]


107. Pediatr Neonatol. 2025 Oct 3:S1875-9572(25)00163-9. doi: 
10.1016/j.pedneo.2025.08.006. Online ahead of print.

Recent advances in human milk bioactives: osteopontin.

Chang SW(1), Lien CH(2), Yeung CY(3).

Author information:
(1)Department of Gastroenterology, Hepatology and Nutrition, MacKay Children's 
Hospital, Taipei, Taiwan.
(2)Department of Gastroenterology, Hepatology and Nutrition, Hsinchu Municipal 
MacKay Children's Hospital, Hsinchu, Taiwan. Electronic address: 
.
(3)Department of Gastroenterology, Hepatology and Nutrition, MacKay Children's 
Hospital, Taipei, Taiwan; Department of Gastroenterology, Hepatology and 
Nutrition, Hsinchu Municipal MacKay Children's Hospital, Hsinchu, Taiwan; 
Department of Medicine, MacKay Medical University, New Taipei City, Taiwan. 
Electronic address:.

Osteopontin, also referred to as bone sialoprotein I or secreted phosphoprotein 
1, is a multifunctional glycoprotein implicated in numerous physiological and 
pathological processes. Initially identified in the early 1980s in osseous 
tissue, OPN plays a critical role in biomineralization and bone remodeling. It 
is expressed in a wide range of tissues and exerts its biological effects 
through binding to integrins and CD44 receptors on target cell surfaces, thereby 
modulating intracellular signaling pathways involved in immune regulation and 
inflammatory responses. Functionally, OPN acts as a cytokine that mediates 
cell-matrix interactions, facilitating cell adhesion, migration, and immune cell 
activation. Recent studies have proposed that the inclusion of OPN in infant 
formula may confer immunological and gastrointestinal benefits comparable to 
those observed in breastfed infants. Consequently, OPN is being investigated as 
a functional ingredient to enhance the nutritional and developmental efficacy of 
infant formula. While exclusive breastfeeding remains the gold standard for 
infant nutrition and developmental support, OPN-fortified formula may serve as a 
viable alternative when breast milk is unavailable or insufficient.

Copyright © 2025 Taiwan Pediatric Association. Published by Elsevier B.V. All 
rights reserved.

DOI: 10.1016/j.pedneo.2025.08.006
PMID: 41107081


108. Int J Food Sci Nutr. 2025 Nov;76(7):709-724. doi: 10.1080/09637486.2025.2570365. 
Epub 2025 Oct 16.

Nutrient quality of beverages: comparing the Nordic Keyhole, Nutri-Score and 
Nutrient Rich Food indices.

Bianchi M(1), Moshtaghian H(1), Lindroos AK(2)(3), Hallström E(1), Winkvist 
A(3)(4).

Author information:
(1)Department of Food Research and Innovation, RISE Research Institutes of 
Sweden, Gothenburg, Sweden.
(2)Department of Food Studies, Nutrition and Dietetics, Uppsala University, 
Uppsala, Sweden.
(3)Department of Internal Medicine and Clinical Nutrition, The Sahlgrenska 
Academy, University of Gothenburg, Gothenburg, Sweden.
(4)Department of Public Health and Clinical Medicine, Sustainable Health, Umeå 
University, Umeå, Sweden.

The nutrient quality of 96 beverages in Sweden was assessed based on the Keyhole 
(KH), Nutri-Score (NS) and Nutrient Rich Food (NRF) index. Of milk- and 
plant-based beverages, 20% were similarly rated by all three as either high or 
low quality. Keyhole evaluated plain milk more coherently to the dietary 
guidelines, whilst NS assessed plant-based beverages more consistently than 
other indicators. Of water-based beverages, 61% were similarly rated by NS and 
NRF, with highest alignment in sugar-sweetened and non-nutritive sweeteners 
containing beverages. According to NS, water, unsweetened beverages, freshly 
squeezed fruit juices and vegetable juices qualified as nutritious choices. NRF 
index evaluated beverages less coherently to the guidelines and the rating of 
products largely mirrored fortification. Thus, agreement among indicators varies 
across beverage groups. NS is suitable to rate the nutrient quality of most 
beverages consistently with the dietary guidelines, except for milk where KH 
outperforms NS.

DOI: 10.1080/09637486.2025.2570365
PMID: 41102873 [Indexed for MEDLINE]


109. Nutrients. 2025 Sep 25;17(19):3057. doi: 10.3390/nu17193057.

Freeze-Dried Donor Milk for Fortification of Mother's Own Milk in Preterm 
Infants: A Preliminary Observational Study.

Rochow N(1)(2), Weiss GA(1), Knab K(1), Prothmann I(1), Schäfer S(1), Zimmermann 
JL(1), Meis A(1), Lohmüller-Weiß S(1), Simon K(1), Schäfer S(1), Welsch J(1), 
Fusch C(1)(3).

Author information:
(1)Department of Pediatrics, Paracelsus Medical University, Breslauer Str. 201, 
90471 Nürnberg, Germany.
(2)Department of Pediatrics, Rostock University Medical Center, 18057 Rostock, 
Germany.
(3)Department of Pediatrics, McMaster University, Hamilton, ON L8S 4L8, Canada.

Background/Objectives: Freeze-dried high-temperature short-time pasteurized 
human milk fortifiers offer potential for exclusive human milk feeding in 
preterm infants while providing necessary nutritional supplementation. However, 
clinical data on safety, tolerability, and growth outcomes remain limited. This 
study evaluated donor milk fortification compared to conventional bovine 
protein-based fortification. Methods: We conducted a prospective 
non-interventional observational cohort study with a retrospectively matched 
comparison cohort at University Children's Hospital of Nuremberg. Preterm 
infants ≥ 30 weeks gestational age requiring mother's own milk fortification 
were included. The exposed cohort (n = 32) received freeze-dried 
high-temperature short-time pasteurized donor milk fortifier at 1.6-4.8 g/100 mL 
of mother's own milk; the matched comparison cohort (n = 32) received bovine 
protein-based fortifier. Primary outcomes included feeding tolerance, safety 
parameters, and anthropometric measurements. Cohorts were matched for birth 
weight (±10%), gestational age (±5 days), and fortified feeding. Results: 
Baseline characteristics were not significantly different: gestational ages 32.8 
± 1.0 versus 33.0 ± 1.2 weeks; birth weights 1900 ± 380 g versus 1840 ± 370 g. 
Excellent feeding tolerance was demonstrated across >3100 feedings. No 
necrotizing enterocolitis, abdominal complications, or serious adverse events 
occurred. Blood glucose, triglycerides, and urea remained normal. Birth weights, 
lengths, and head circumferences showed no significant differences. Discharge 
parameters including weight, length, head circumference, and length of stay were 
also not significantly different. Conclusions: Freeze-dried human milk 
fortification demonstrates excellent safety and tolerability in preterm infants 
≥ 30 weeks gestational age, achieving anthropometric outcomes not significantly 
different to bovine protein-based fortification. However, the suboptimal 
protein-to-energy ratio may limit applicability for very low birth weight 
infants. Therefore, freeze-dried high-temperature short-time pasteurized human 
milk fortification is suggested to provide appropriate nutritional 
supplementation for preterm infants with a birth weight over 1500 g.

DOI: 10.3390/nu17193057
PMCID: PMC12526076
PMID: 41097134 [Indexed for MEDLINE]

Conflict of interest statement: The authors declare no conflicts of interest.


110. Food Res Int. 2025 Nov;220:117080. doi: 10.1016/j.foodres.2025.117080. Epub 2025 
Jul 28.

Oxidative stability of polyunsaturated fatty acid-enriched infant flours based 
on teff and cowpea: Impact of natural antioxidants from amaranth leaves and 
black rice bran.

Cancalon M(1), Hemery Y(2), Dormoy N(3), Barouh N(4), Baréa B(4), Durand E(4), 
Barbar R(5), Antoine-Assor C(5), Micard V(5), Lhomond L(5), Reau A(5), 
Villeneuve P(6), Bourlieu-Lacanal C(7).

Author information:
(1)CIRAD, UMR Qualisud, F34398 Montpellier, France; Qualisud, Univ Montpellier, 
Avignon Université, CIRAD, Institut Agro, IRD, Université de La Réunion, 
Montpellier, France; UMR IATE, UM Montpellier, INRAE, Institut Agro, F34060 
Montpellier, France.
(2)Qualisud, Univ Montpellier, Avignon Université, CIRAD, Institut Agro, IRD, 
Université de La Réunion, Montpellier, France; IRD, CIRAD Qualisud, F34398 
Montpellier, France.
(3)CIRAD, UMR Qualisud, F34398 Montpellier, France.
(4)CIRAD, UMR Qualisud, F34398 Montpellier, France; Qualisud, Univ Montpellier, 
Avignon Université, CIRAD, Institut Agro, IRD, Université de La Réunion, 
Montpellier, France.
(5)UMR IATE, UM Montpellier, INRAE, Institut Agro, F34060 Montpellier, France.
(6)CIRAD, UMR Qualisud, F34398 Montpellier, France; Qualisud, Univ Montpellier, 
Avignon Université, CIRAD, Institut Agro, IRD, Université de La Réunion, 
Montpellier, France. Electronic address:.
(7)UMR IATE, UM Montpellier, INRAE, Institut Agro, F34060 Montpellier, France. 
Electronic address:.

From 4/6 months of age, complementary foods such as infant flours (IF) are 
introduced in the infant diet to supplement the nutritional intake of breast 
milk and/or infant formula that are no longer sufficient. However, the lipid 
profile and micronutrient content of IF are often unbalanced. In addition, the 
current lack of knowledge about oxidation mechanisms in low-moisture systems 
makes it difficult to control them in this type of food. The aim of this study 
was to better understand oxidation mechanisms in IF with optimized lipid profile 
for infant nutritional needs using climate smart crops, and with or without, the 
addition of vegetal sources of natural antioxidants. Five IF were formulated 
using climate smart crops, i.e. cowpea and teff flours, as sources of essential 
omega-3 fatty acids to complement soy flours. Four IF contained heat treated 
cowpea flour to evaluate the impact of lipoxygenases inactivation in this flour 
on global IF oxidative stability. Three IF were also fortified in long chain 
polyunsaturated fatty acid that are essential to support infant development to 
check whether this staple food could be good nutritional vector. Among these IF, 
two were enriched in natural antioxidant fractions with the addition of either 
powdered amaranth leaves or black rice bran, bringing carotenoids and phenolic 
compounds. The oxidative stability of IF was monitored over a 6-month storage 
period at 25 °C. The results showed that including teff and cowpea flours in the 
IF formulation enabled a balanced lipid profile (omega-6/omega-3 ratio close to 
5) and ensured relatively good overall oxidative stability. However, 50 % of 
vitamin A was lost after 2 weeks of storage. Black rice bran and amaranth leaves 
acted as vitamin A stabilizers, delaying losses. This study unveiled that 
oxidation is triggered mainly during initial flour milling and propagated at a 
relatively slow rate over storage in these low moisture foods. Oxidation rate 
was related to the antioxidants composition and was faster in the presence of 
photosensitive compounds.

Copyright © 2025 The Authors. Published by Elsevier Ltd.. All rights reserved.

DOI: 10.1016/j.foodres.2025.117080
PMID: 41074306 [Indexed for MEDLINE]

Conflict of interest statement: Declaration of competing interest The authors 
declare that they have no known competing financial interests or personal 
relationships that could have appeared to influence the work reported in this 
paper.


111. Front Nutr. 2025 Sep 24;12:1650108. doi: 10.3389/fnut.2025.1650108. eCollection 
2025.

Enhancing human milk iodine concentration: a data-driven action plan for Latvia.

Aumeistere L(1), Kovada K(2), Driksna A(1), Kozačenko J(3), Ciproviča I(1).

Author information:
(1)Faculty of Agriculture and Food Technology, Latvia University of Life 
Sciences and Technologies, Jelgava, Latvia.
(2)Faculty of Medicine and Life Sciences, University of Latvia, Riga, Latvia.
(3)Institute of Public Health, Rīga Stradiņš University, Riga, Latvia.

BACKGROUND: Iodine deficiency remains a significant public health challenge in 
Europe. However, there is currently a lack of data on iodine status among 
lactating women and infants in Latvia.
METHODS: This cross-sectional study assessed the impact of maternal diet on 
iodine concentration in human milk. The data obtained were used as reference 
points for planning targeted interventions to improve iodine supply. Between 
October 2024 and April 2025, pooled 24-h human milk samples, 72-h food diaries 
and questionnaires on anthropometric, sociodemographic characteristics were 
collected from 55 exclusively breastfeeding women residing in Latvia. Human milk 
iodine concentration was determined using inductively coupled plasma mass 
spectrometry, while dietary data were analyzed using the Estonian open-access 
dietary analysis program NutriData.
RESULTS: Human milk iodine concentration among participants varied considerably 
(15.00-291.00 μg L-1) with a median value (86.00 μg L-1) below the level 
indicating optimal iodine supply for an exclusively breastfed infant (i.e., 
150 μg L-1). A significant positive association was observed between maternal 
iodine intake and human milk iodine concentration (ρ = 0.407, p = 0.002). The 
participants' diets lacked iodine-rich sources (i.e., fish, dairy, iodized salt) 
and, without supplementation, did not reach the adequate intake for iodine 
(i.e., 200 μg per day).
CONCLUSION: Preliminary data indicate that current maternal dietary practices in 
Latvia are insufficient to guarantee optimal iodine supply to exclusively 
breastfed infants. Study results highlight the need for policy reforms and 
improved nutritional guidance to address the iodine deficiency problem in 
Latvia.

Copyright © 2025 Aumeistere, Kovada, Driksna, Kozačenko and Ciproviča.

DOI: 10.3389/fnut.2025.1650108
PMCID: PMC12504860
PMID: 41070018

Conflict of interest statement: The authors declare that the research was 
conducted in the absence of any commercial or financial relationships that could 
be construed as a potential conflict of interest.


112. Curr Opin Clin Nutr Metab Care. 2026 Jan 1;29(1):89-97. doi: 
10.1097/MCO.0000000000001178. Epub 2025 Nov 7.

Amino acids and lean mass accretion in preterm neonates.

Ramos Dos Santos AC(1), Fiorotto ML(2), Davis TA(1)(2).

Author information:
(1)Institute for Advancing Health Through Agriculture, Texas A&M AgriLife, Texas 
A&M University, College Station.
(2)Department of Pediatrics, Children's Nutrition Research Center, Baylor 
College of Medicine, Houston, Texas, USA.

PURPOSE OF REVIEW: This review evaluates recent evidence on how total protein 
and energy intake, amino acid composition, and nutrient delivery modality 
influence protein synthesis in preterm neonates, with the goal of informing 
nutritional strategies to support optimal lean mass accretion and growth.
RECENT FINDINGS: Preterm neonates exhibit anabolic resistance to feeding due to 
impaired insulin and amino acid signaling in skeletal muscle. Enteral nutrition, 
especially fortified human milk, supports better lean mass accretion and 
neurodevelopment than parenteral nutrition. Intermittent bolus feeding and 
pulsatile leucine supplementation during continuous feeding further enhance 
mTORC1-dependent translation initiation signaling and protein synthesis 
efficiency in skeletal muscle, offering promising strategies to optimize lean 
tissue growth.
SUMMARY: Strategies that prioritize early enteral feeding, appropriate milk 
fortification, and nutrient delivery patterns that mimic feeding pulsatility may 
overcome anabolic resistance and enhance lean mass accretion, supporting healthy 
growth trajectories in preterm infants.

Copyright © 2025 The Author(s). Published by Wolters Kluwer Health, Inc.

DOI: 10.1097/MCO.0000000000001178
PMCID: PMC12700696
PMID: 41057241 [Indexed for MEDLINE]

Conflict of interest statement: All authors report no conflicts of interest.


113. BMJ Open. 2025 Oct 5;15(10):e105609. doi: 10.1136/bmjopen-2025-105609.

MaxiMoM InForM: individualised fortification of human milk for very low 
birthweight infants- protocol of a three-arm randomised clinical trial.

Beggs MR(1)(2), Pichardo D(1), Chrzaniecki A(1)(2), Kotsopoulos K(3), Bishara 
R(4), Ng E(4)(5), Tomlinson C(5)(6), Campbell D(5)(7), Vaz S(8), Kiss A(9), 
Unger S(10)(11)(12), O'Connor DL(13)(2)(14).

Author information:
(1)Translational Medicine, The Hospital for Sick Children, Toronto, Ontario, 
Canada.
(2)Department of Nutritional Sciences, University of Toronto, Toronto, Ontario, 
Canada.
(3)Allied Health Disciplines, Sinai Health, Toronto, Ontario, Canada.
(4)DAN Women & Babies Program, Sunnybrook Health Sciences Centre, Toronto, 
Ontario, Canada.
(5)Department of Pediatrics, University of Toronto, Toronto, Ontario, Canada.
(6)Neonatal Intensive Care Unit, The Hospital for Sick Children, Toronto, 
Ontario, Canada.
(7)St. Michael's Hospital, Unity Health Toronto, Toronto, Ontario, Canada.
(8)Department of Pediatrics, William Osler Health System, Brampton, Ontario, 
Canada.
(9)Institute of Health Policy, Management and Evaluation, University of Toronto, 
Toronto, Ontario, Canada.
(10)Neonatology, IWK Health Centre, Halifax, Nova Scotia, Canada.
(11)Department of Pediatrics, Dalhousie University, Halifax, Nova Scotia, 
Canada.
(12)Rogers Hixon Ontario Human Milk Bank, Sinai Health, Toronto, Ontario, 
Canada.
(13)Translational Medicine, The Hospital for Sick Children, Toronto, Ontario, 
Canada.
(14)Department of Pediatrics, Sinai Health, Toronto, Ontario, Canada.

INTRODUCTION: To meet the elevated nutritional requirements of very low 
birthweight (<1500 g, VLBW) infants during hospitalisation, many neonatal units 
routinely add a fixed dose of multinutrient fortifier to human milk. 
Individualised approaches that either tailor macronutrient additions to routine 
analysis of milk composition (target fortification) or an infant's metabolic 
response to nutrition (blood urea nitrogen (BUN) adjustable fortification) show 
short-term improvement in growth, but important outcomes such as longer-term 
neurodevelopment and major in-hospital morbidity have not been systematically 
assessed. The aim of our study is to determine whether target or BUN adjustable 
fortification of human milk, compared with standard fortification, during 
hospitalisation improves cognitive (primary), language and motor development of 
VLBW infants at 18-24 months corrected age (CA). Secondary outcomes include 
growth, body composition, mortality and serious morbidity.
METHODS AND ANALYSIS: This is a three-arm, pragmatic, multicentre, double-blind, 
randomised clinical trial of 615 human milk-fed infants born either (1) ≤1250 g 
or (2) <30 weeks and <1500 g. Once enteral feeds reach 100 mL/kg/day, and until 
36 weeks corrected gestational age (CGA), infants are fed according to one of 
three fortification protocols: (1) standard fortification, in which a fixed dose 
of nutrients is added to human milk; (2) target fortification, in which values 
from twice weekly direct analysis of mother's milk and each batch of donor milk 
are used to provide 4 and 4.8 g/kg/day true protein and fat, respectively; and 
(3) BUN adjustable fortification, in which weekly BUN values are used to adjust 
protein fortification of milk. Neurodevelopment is assessed using the Bayley-IV 
scale at 18-24 months CA. Growth, morbidities and nutrient intakes are recorded 
daily during hospitalisation, and skinfolds are measured at 36 weeks CGA and at 
4 months CA. At 4 months CA, body composition is determined by air displacement 
plethysmography on a subset of infants.
ETHICS AND DISSEMINATION: Ethical approval was obtained from Clinical Trials 
Ontario (CTO) and local research ethics boards that are not CTO members. Study 
findings will be disseminated to clinicians at seminars and conferences and in 
peer-reviewed publications.
TRIAL REGISTRATION NUMBER: NCT05308134.

© Author(s) (or their employer(s)) 2025. Re-use permitted under CC BY-NC. No 
commercial re-use. See rights and permissions. Published by BMJ Group.

DOI: 10.1136/bmjopen-2025-105609
PMCID: PMC12506200
PMID: 41047263 [Indexed for MEDLINE]

Conflict of interest statement: Competing interests: None declared.


114. Clin Nutr. 2025 Nov;54:71-82. doi: 10.1016/j.clnu.2025.09.006. Epub 2025 Sep 23.

Docosahexaenoic acid supplementation in pregnancy and early lactation: Impacts 
on breast milk docosahexaenoic acid and Maternal-Infant gut microbiota - A 
randomized controlled trial.

He T(1), Zhang J(1), Zhao A(2), Luo S(3), Jiang H(4), Tang M(5), Zhang Y(6).

Author information:
(1)Department of Nutrition and Food Hygiene, School of Public Health, Peking 
University, Beijing 100191, China.
(2)Department of Nutrition and Food Hygiene, School of Public Health, Peking 
University, Beijing 100191, China; Vanke School of Public Health, Tsinghua 
University, Beijing 100084, China.
(3)Maternal and Child Health Hospital of Changsha County, Changsha City 410100, 
Hunan province, China.
(4)School of Nursing, Peking University, Beijing 100091, China.
(5)Department of Food Science and Human Nutrition, Colorado State University, 
Fort Collins, CO 80521, USA.
(6)Department of Nutrition and Food Hygiene, School of Public Health, Peking 
University, Beijing 100191, China. Electronic address:.

AIM: This study aimed to evaluate whether docosahexaenoic acid (DHA) 
supplementation during pregnancy increases DHA concentrations in colostrum among 
Chinese women, and to determine whether continued postpartum supplementation is 
necessary to sustain DHA levels throughout lactation. Additionally, the study 
examined the impact of maternal DHA supplementation on the gut microbiota of 
both mothers and their infants.
METHODS: 79 pregnant women were enrolled in this multicenter, double-blinded, 
randomized, placebo-controlled, parallel trial and were randomized to three 
groups (CC, control group; FC, DHA supplementation in the third trimester; FF, 
DHA supplementation from the third trimester to 42 days postpartum). 
Participants in the FF and FC groups were administered 100 mg/day DHA via milk 
powder and those in the CC group were administered milk powder without DHA 
fortification. Breast milk was collected - on days 3 and 42 postpartum. Fatty 
acid composition in breast milk was measured with gas chromatography and 
expressed as percentage of total fatty acids by weight. Fecal samples from 
mothers (at gestational week 28, 38 and 42 days postpartum) and infants (at 3 
and 42 days of age) were collected, then analyzed using 16S rRNA sequencing.
RESULTS: The FF group had significantly higher levels of DHA in colostrum than 
the other two groups (FF: 1.49%, FC: 1.27%, CC: 1.11, P-ANCOVA = 0.018) but not 
in early mature milk (FF: 0.94%, FC: 0.82%, CC: 0.85%, P-ANCOVA = 0.646). At 42 
days postpartum, both mothers and infants in the FF group exhibited a higher 
abundance of the genus Lactobacillus in the fecal microbiome.
CONCLUSION: In this cohort of Chinese women, maternal supplementation with 100 
mg/day of DHA starting from the third trimester of pregnancy significantly 
increased DHA concentrations in colostrum. However, 100 mg/day may be 
insufficient to sustain DHA levels beyond colostrum. Additionally, maternal DHA 
supplementation was found to influence Lactobacillus abundance at 42 days 
postpartum of both mothers and their infants. Future studies should explore 
higher DHA doses for long-term lactation stability, and investigate dynamic 
changes in other gut microbiota taxa and microbiota-mediated health effects.
CLINICAL TRIAL REGISTRATION: ChinaClinicalTrial.gov (ChiCTR1800019461).

Copyright © 2025 Elsevier Ltd and European Society for Clinical Nutrition and 
Metabolism. All rights reserved.

DOI: 10.1016/j.clnu.2025.09.006
PMID: 41045839 [Indexed for MEDLINE]


115. Front Pediatr. 2025 Sep 17;13:1679676. doi: 10.3389/fped.2025.1679676. 
eCollection 2025.

Analysis of NEC cases registered in the human milk bank database.

Mizuno K(1), Wada YS(2), Sakurai M(3), Tani Y(4), Miyata M(5), Shindo J(6), 
Nishimaki S(7), Den H(8).

Author information:
(1)Department of Pediatrics, Showa Medical University, Tokyo, Japan.
(2)Division of Neonatology, Center for Maternal-Fetal, Neonatal, and 
Reproductive Medicine, National Center for Child Health and Development, Tokyo, 
Japan.
(3)Department of Pediatrics and Neonatology, Kameda Medical Center, Chiba, 
Japan.
(4)Department of Pediatrics, Nara Medical University, Nara, Japan.
(5)Department of Pediatrics, Fujita Health University School of Medicine, Aichi, 
Japan.
(6)Department of Neonatology, Tokyo Metropolitan Children's Medical Center, 
Tokyo, Japan.
(7)Department of Pediatrics, Yokohama City University, Yokohama, Japan.
(8)Department of Hygiene, Public Health, and Preventative Medicine, Showa 
Medical University, Tokyo, Japan.

BACKGROUND: Necrotizing enterocolitis (NEC) remains a major cause of morbidity 
and mortality in extremely low birth weight infants (ELBWIs), despite widespread 
donor human milk (DHM) use. This study examined NEC cases among DHM recipients 
to explore potential contributing factors.
METHODS: We retrospectively analyzed 1,425 infants registered in Japan's human 
milk bank database (2018-2023). NEC cases at Bell stage ≥ II were confirmed by 
attending physicians. Infants who received DHM only after NEC onset were 
excluded. Cases were categorized by onset timing and associated clinical 
factors.
RESULTS: Among 1,324 very low birth weight infants, 21 (1.58%) developed NEC, 
with 20 requiring surgical intervention. Median gestational age and birth weight 
were 25.1 weeks and 637 g, respectively. NEC onset was classified as follows: 
within 7 days of birth (n = 5), after 2 months (n = 5), after formula or 
fortifier use (n = 6), associated with hemodynamic changes (n = 7), or of 
unknown etiology (n = 4). Common factors included symptomatic PDA, congenital 
heart disease, infection, formula exposure, and ophthalmologic procedures.
CONCLUSION: NEC can develop despite DHM use, often in association with early 
infections, PDA, or fortification. Strategies to further reduce NEC incidence 
should include management of hemodynamic instability, delayed formula 
introduction, and use of exclusive human milk-based diets. Further research 
should explore potential roles of ophthalmologic interventions and human milk 
fortifiers in NEC development.

© 2025 Mizuno, Wada, Sakurai, Tani, Miyata, Shindo, Nishimaki and Den.

DOI: 10.3389/fped.2025.1679676
PMCID: PMC12484215
PMID: 41040642

Conflict of interest statement: The authors declare that the research was 
conducted in the absence of any commercial or financial relationships that could 
be construed as a potential conflict of interest.


116. Breastfeed Med. 2025 Dec;20(12):888-894. doi: 10.1177/15568253251383521. Epub 
2025 Sep 30.

Impact of Short-Term Freezing at -20°C on Macronutrient Content in Human Milk 
from Preterm Infants.

Livaoglu Say B(1), Hatipoglu HU(2), Uras HB(3), Uras N(4).

Author information:
(1)Pediatrics Department, Alanya Alaaddin Keykubat Unıversity, Antalya, Turkey.
(2)Department of Child Health and Diseases, Hamidiye Medical Faculty of Health 
Sciences University, Istanbul Haseki Health Application and Research Center, 
Istanbul, Turkey.
(3)Psychology Department, Texas Christian University, Fort Worth, Texas, USA.
(4)Pediatrics Department, Istinye University Hospital, Istanbul, Turkey.

Objectives: Breast milk is an ideal food for newborns born term and preterm. The 
short-term storage of human milk (HM) involves freezing at low temperatures; 
however, its effects on macronutrients remain unclear. The macronutrients in HM 
are generally thought to be unaffected by short-term freezing. This study aimed 
to analyze the impact of freezing HM from preterm infants at -20°C for 5 days on 
macronutrient content. Methods: HM samples were collected from 54 mothers of 
preterm infants. Each sample was divided into three aliquots and stored at -20°C 
for 1 and 5 days. After thawing and homogenization, energy and macronutrient 
contents were measured using a HM infrared spectroscopy analyzer. Results: We 
analyzed 162 samples (from 54 mothers of preterm infants). Mean baseline 
concentrations in fresh milk were protein 1.55 ± 0.61 g/100 mL, carbohydrates 
6.87 ± 0.81 g/100 mL, fat 4.29 ± 2.16 g/100 mL, and energy 75.9 ± 27.3 kcal/100 
mL. Freezing at -20°C reduced protein to 1.33 ± 0.35 g/100 mL after 24 hours and 
1.21 ± 0.38 g/100 mL after 5 days (14.2% and 21.9% decrease, p < 0.001), fat to 
3.23 ± 1.15 g/100 mL and 3.04 ± 1.40 g/100 mL (24.7% and 29.1% decrease, p < 
0.001), and energy to 64.3 ± 12.8 kcal/100 mL and 61.7 ± 16.1 kcal/100 mL, 
respectively (p < 0.001); carbohydrate content remained unchanged (p = 0.41). 
Conclusions: Freezing HM at -20°C for 5 days can significantly reduce critical 
nutrients, including fat and protein, suggesting the need for individualized 
fortification strategies to ensure optimal growth in preterm infants.

DOI: 10.1177/15568253251383521
PMID: 41027670 [Indexed for MEDLINE]


117. Mymensingh Med J. 2025 Oct;34(4):1110-1117.

Knowledge on Vitamin A and Household Consumption Frequency of Vitamin A-Rich 
Foods among Rural Population in Some Selected Villages, Chandina, Cumilla, 
Bangladesh.

Shakil M(1), Sultana T, Yeasmin S.

Author information:
(1)Dr Mohammad Shakil, Associate Professor, Department of Community Medicine, 
Army Medical College, Cumilla, Bangladesh;.

Vitamin A (retinol) is an essential, fat soluble nutrient stored in body organs, 
mainly the liver. Vitamin A is an essential nutrient for maintaining eye health 
and vision, growth, immune function and survival. Vitamin A deficiency is one of 
the most serious health problems in developing countries. This study was carried 
out to assess the knowledge on vitamin A and household consumption frequency of 
vitamin A-rich foods in some selected villages Chandina, Cumilla, Bangladesh. 
This cross-sectional study was conducted among 480 adult populations residing in 
some selected villages in Chandina, Cumilla, Bangladesh using a 
self-administered questionnaire from October 2021 to December 2021. Data were 
analyzed using computer software like Microsoft Word, Microsoft Excel. The study 
revealed that most of the respondents were in more than 45 years age groups 
(42.0%) and majority of them were female 291(60.62%) with Muslim predominant 
440(91.67%). In concern of education more than 40.0% of them were completed 
secondary level 161(33.54%) and maximum of them were housewife 261(54.38%). More 
than 40.0% respondent's monthly family income was BDT 10,000-20,000(41.88%). 
About 66.88% participants had knowledge on vitamin A and maximum of them got 
this inform from health assistant 178(55.45%). According to vitamin A capsule 
given to your children, most the respondents told that they have given to their 
children (90.03%). According to their specific knowledge about vitamin A, most 
the respondents told about vitamin A maintains only eye health 171(53.27%), then 
followed by vitamin A deficiency only causes night blindness 95(29.60%), vitamin 
A helps in immunity 24(7.48%), vitamin A helps in digestion 13(4.05%), vitamin A 
maintains only skin health 11(3.43%) and vitamin A makes us healthy 7(2.18%). 
Regarding household consumption of vitamin A-rich foods, most of the respondents 
told that they eat green leafy vegetables less than 2 kg/week, carrot less than 
2 kg/week, sweet potato less than 2 kg/week, pumpkin less than 2 kg/week, tomato 
2-3 kg/week, papaya/coloured fruits less than 2 kg/week, liver less than 1 
kg/week, small fish 1-2 kg/week, whole milk 1-2 L/week, egg 1-2 dozen/week. For 
cooking most of the respondents used vitamin A fortified soybean oil regularly 
244(76.01%). In general the knowledge of study population regarding the 
importance of vitamin A was quiet satisfactory but frequency of consumption 
vitamin A-rich foods was still poorer in rural areas.

PMID: 41024549 [Indexed for MEDLINE]


118. Int J Mol Sci. 2025 Sep 10;26(18):8813. doi: 10.3390/ijms26188813.

Antioxidant Activity, Total Polyphenol Content, and Mineral Composition of Milk 
Beverages Fortified with Spice Mixtures (Clove, Cinnamon, and Turmeric) and 
Natural Sweeteners (Erythritol and Stevia): Evidence of Synergistic or 
Antagonistic Effects of Compounds.

Rak K(1), Kolniak-Ostek J(2), Gajda R(1), Marcinkiewicz K(1), Nemś A(3), 
Raczkowska E(1).

Author information:
(1)Department of Human Nutrition, Faculty of Biotechnology and Food Science, 
Wrocław University of Environmental and Life Sciences, 51-630 Wrocław, Poland.
(2)Department of Fruit, Vegetable and Plant Nutraceutical Technology, Faculty of 
Biotechnology and Food Sciences, Wrocław University of Environmental and Life 
Sciences, 51-630 Wrocław, Poland.
(3)Department of Food Storage and Technology, Faculty of Biotechnology and Food 
Science, Wrocław University of Environmental and Life Sciences, 51-630 Wrocław, 
Poland.

Extensive research is underway on a variety of functional foods that support 
consumer health. A promising combination is milk (a drink with high nutritional 
value), fortified with spices (naturally rich in antioxidant compounds) and 
supplemented with low-calorie, health-promoting sweeteners. The aim was to 
analyze the antioxidant activity (AA), total polyphenol content (TPC), and 
mineral composition of milk beverages fortified with a mixture of spices and 
sweeteners and to verify the interactions between these ingredients. Twenty-four 
variants of milk drinks were prepared with the addition of three types of spice 
mixtures (1:1) of clove (Cl), cinnamon (Ci), and turmeric (Tu) with the shares 
of 2.5%, 5%, 7.5%, and 10%, and two types of sweeteners-erythritol (E) and 
stevia (S)-as well as six control samples, including three 10% aqueous solutions 
of spice mixtures. AA was measured using the ABTS, FRAP, and DPPH methods. TPC 
was determined using the Folin-Ciocalteu method. Mineral content (Ca, Mg, Fe, 
Cu, and Zn) was assessed using the FAAS/FAES method. The highest AA was 
demonstrated for beverages with Cl-Ci and Cl-Tu, whereas the highest TPC was 
found in those with Cl-Tu. AA and TPC values increased with the increase in the 
share of spices in the beverages, and both measurements were significantly 
higher in beverages with erythritol compared to those with stevia. Despite the 
4-5 times lower TPC, 3-8 times higher AA was demonstrated in beverages with the 
addition of 10% spice mixtures than their corresponding control samples (aqueous 
solutions with the addition of 10% spice mixtures), which suggests the great 
role of interactions between nutrients in food. For beverages with 10% 
Cl-Ci/Cl-Tu mixtures, significantly higher ABTS, FRAP, and DPPH were observed 
than would result from the simple sum of AA of the components (synergistic 
effect). The opposite (antagonistic) effect was observed for beverages with 
Ci-Tu. AA was positively correlated with the content of Fe, Mg, and Cu; 
negatively with Ca; and not correlated with Zn. The results suggest that the 
most health-promoting properties are exhibited by the milk drink with the 
addition of 10% Cl-Tu and erythritol, demonstrating the highest AA and TPC, the 
strongest synergistic effect of the components, and the highest content of Mg 
and Zn. This study highlights the importance of carefully selecting and 
combining ingredients to maximize the antioxidant properties of functional 
beverages. However, further research is needed to expand knowledge on this 
issue.

DOI: 10.3390/ijms26188813
PMCID: PMC12469906
PMID: 41009382 [Indexed for MEDLINE]

Conflict of interest statement: The authors declare no conflicts of interest.


119. Children (Basel). 2025 Sep 13;12(9):1227. doi: 10.3390/children12091227.

Does Breastfeeding Small for Gestational Age Neonates Promote a Healthier Growth 
Pattern? A Narrative Review.

Atzemoglou N(1), Tzavellas NP(1), Dermitzaki N(1), Baltogianni M(1), Balomenou 
F(1), Serbis A(2), Giapros V(1).

Author information:
(1)Neonatal Intensive Care Unit, University Hospital of Ioannina, 45500 
Ioannina, Greece.
(2)Department of Pediatrics, University Hospital of Ioannina, 45500 Ioannina, 
Greece.

Background: Small for gestational age neonates represent a population at risk of 
growth failure or deviant growth patterns and long-term metabolic complications. 
Breastfeeding has been identified as a critical factor in promoting healthier 
growth and long-term metabolic health in both full-term and preterm appropriate 
for gestational age infants, but similar studies in small for gestational age 
infants are limited. The aim of this narrative review is to assess the impact of 
breastfeeding on growth and body composition in small for gestational age 
neonates. Methods: The PubMed and Google Scholar databases were screened for the 
relevant literature. The following terms, were used: "low birth weight", "in 
utero growth restriction", "small for gestational age", "human milk", and 
"growth". The initial screening identified 57 relevant studies. Thirteen of them 
fulfilled the eligibility criteria and were included in this narrative review. 
Results: In preterm small for gestational age neonates, human milk nutrition was 
associated with healthier catch-up growth without excessive fat accumulation. 
Fortification strategies were associated with enhanced growth outcomes without 
increased incidence of neonatal morbidities. In the context of full-term, small 
for gestational age neonates, exclusive breastfeeding has been demonstrated to 
be associated with healthy catch-up growth. Furthermore, human milk nutrition 
has been shown to mitigate the predisposition of these children to obesity and 
cardiometabolic complications. Conclusions: According to the limited extant 
literature, human milk feeding has been identified as a potentially protective 
factor for small for gestational age neonates, promoting healthier growth 
patterns and long-term cardiometabolic health. However, larger prospective 
studies are needed to evaluate human milk feeding and human milk fortification 
in association with growth and long-term outcomes in small for gestational age 
infants.

DOI: 10.3390/children12091227
PMCID: PMC12468219
PMID: 41007092

Conflict of interest statement: The authors declare no conflict of interest.


120. Nutr Clin Pract. 2025 Sep 23. doi: 10.1002/ncp.70037. Online ahead of print.

Enteral nutrition safety practices in pediatric transplant: Perspectives of 
dietitians in pediatric transplant.

Sun T(1), Leonard J(1), Potvin R(1), Bastardi H(2), Hron BM(1).

Author information:
(1)Division of Gastroenterology, Hepatology, and Nutrition and Center for 
Nutrition, Boston Children's Hospital, Boston, Massaachusetts, USA.
(2)Department of Cardiology, Advanced Cardiac Therapies, Boston Children's 
Hospital, Boston, Massachusetts, USA.

BACKGROUND: Our objective was to characterize enteral nutrition safety practices 
and education for pediatric solid organ transplant recipients and compare 
practices with the 2017 American Society for Parenteral and Enteral Nutrition 
(ASPEN) Safe Practices for Enteral Nutrition Therapy.
METHODS: A 43-question electronic survey was distributed through the national 
registered dietitian pediatric transplant listserv. Questions reviewed formula 
hang-time, preparation, storage during initial transplant admission, and 
discharge education.
RESULTS: Sixty-six of 216 (31%) individuals completed at least one survey 
section. Forty-one of 47 (87%) reported a standard inpatient policy, and 40/40 
(100%) reported ASPEN Safe Practices compliance for nonsterile powder formula 
with or without additives or unfortified and fortified human milk, whereas 33/39 
(85%) complied for sterile liquid formula in an open system. Hospital size, 
type, and location did not predict compliance practices. Discharge education was 
primarily provided by dietitians (98%) and nurses (37%). Four-hour hang-time 
education was provided by 18/42 (43%) respondents for sterile formula in an open 
system, 31/42 (74%) for nonsterile powder formula in an open system, and 35/42 
(83%) for nonsterile formula with additives. Educator type (dietitian vs 
non-dietitian or nurse vs non-nurse) did not predict compliance for sterile 
liquid in open system or nonsterile powder formula in an open system.
CONCLUSION: Inpatient policies for formula hang-time are highly compliant with 
2017 ASPEN recommendations. However, formula hang-time discharge education 
varied, particularly for sterile liquid formula in an open system. Standardizing 
enteral nutrition safety education for transplant patients is critical for 
minimizing infection risk within this immunocompromised population.

© 2025 American Society for Parenteral and Enteral Nutrition.

DOI: 10.1002/ncp.70037
PMID: 40985927


121. J Sci Food Agric. 2026 Jan 15;106(1):563-578. doi: 10.1002/jsfa.70191. Epub 2025 
Sep 23.

In vitro dynamic infant digestion model: effects of 1,3-dioleoyl-2-palmitoyl 
glycerol, lactoferrin, and casein phosphopeptide fortification on infant formula 
protein and fat digestion.

Anwar S(1), Zhang L(1)(2), Zhong Y(1), Peng X(3), Guo D(3), Wang J(3), Li W(3), 
Zhou P(1)(2).

Author information:
(1)School of Food Science & Technology, Jiangnan University, Wuxi, China.
(2)State Key Laboratory of Food Science & Resources, Jiangnan University, Wuxi, 
China.
(3)Hunan Engineering and Technology Research Center for Nutrition and Health 
Products, Innovation Platform and Talent Plan, Ausnutria Dairy (China) Co., 
Ltd., Ausnutria Institute of Food and Nutrition, Changsha, Hunan Province, 
China.

BACKGROUND: This study investigated the effects of fortifying infant formulas 
(IFs) with casein phosphopeptide (CPP), lactoferrin (LTF), and 
1,3-dioleoyl-2-palmitoyl glycerol (OPO) on protein and fat digestion, using an 
in vitro infant dynamic digestion model with human milk (HM) as the standard. 
Four formulas were examined: an unfortified control (CC) and three fortified 
variants (F1, F2, and F3), each containing the same fortifying components at 
varying concentrations.
RESULTS: Sodium dodecyl sulfate-polyacrylamide gel electrophoresis (SDS-PAGE) 
showed that most proteins were completely hydrolyzed during gastrointestinal 
digestion, except β-lactoglobulin (β-Lg). After 180 min, protein digestibility 
ranged from 97.6% to 99.08%, with F2 reaching the highest value (99.08%), 
comparable to HM (98.92%). Amino acid fit analysis indicated strong similarity 
to HM in F2 (96.67%) and F3 (94.87%). Protein hydrolysis and free amino acid 
(FAA) release were significantly greater (P < 0.05) in F2 and F3 than in CC and 
F1. The fortified IFs showed peptide profiles and lipolysis degrees (LDs) closer 
to HM than the control, although HM had the greatest peptide release. In total, 
145 bioactive peptides were identified, 77 of which were common to all IFs.
CONCLUSION: These findings suggest that fortification with CPP, LTF, and OPO can 
modulate the protein and fat digestion patterns in IF, increasing its similarity 
to HM. © 2025 Society of Chemical Industry.

© 2025 Society of Chemical Industry.

DOI: 10.1002/jsfa.70191
PMID: 40984756 [Indexed for MEDLINE]


122. Arch Osteoporos. 2025 Sep 22;20(1):128. doi: 10.1007/s11657-025-01611-7.

Effect of vitamin D-fortified foods on bone health and osteoporosis: a scoping 
review and future policies focusing on the United Arab Emirates.

Iqbal S(1), Abuajlan M(2), Alktheri A(2), Malik ZI(3), Ahmad AMR(3), Qudah T(2), 
Abumweis S(4).

Author information:
(1)College of Pharmacy, Al Ain University, Abu Dhabi Campus, Abu Dhabi, United 
Arab Emirates..
(2)College of Pharmacy, Al Ain University, Abu Dhabi Campus, Abu Dhabi, United 
Arab Emirates.
(3)Department of Human Nutrition and Dietetics, NUST School of Health Sciences, 
National University of Sciences & Technology (NUST) Sector H-12, Islamabad, 
Pakistan.
(4)Department of Clinical Nutrition and Dietetics, Faculty of Applied Medical 
Sciences, The Hashemite University, Zarqa, Jordan.

BACKGROUND: Osteoporosis is a major public health issue characterized by reduced 
bone mass and increased fracture risk particularly affecting the older 
population.
OBJECTIVE: This scoping review aims to summarize the existing literature 
focusing on the impact of vitamin D-fortified foods in reducing the risk of 
osteoporosis-related outcomes and improving the overall bone health.
METHODS: Considering the study objectives, two databases, i.e., PubMed and 
Scopus, were selected to search for literature. A search term "vitamin D" OR 
"vitamin D2" OR "vitamin D3" OR "cholecalciferol" OR "ergocalciferol" AND 
"fortified food" OR "fortification" OR "oil fortification" OR "ghee 
fortification" OR "cereal fortification" OR "margarine fortification" OR "milk 
fortification" OR "egg fortification" AND "osteoporosis" OR "bone mineral 
density" OR "bone health" was applied to retrieve the available literature. 
Preferred Reporting Items for Systematic Reviews and Meta-Analyses extension for 
Scoping Reviews (PRISMA-ScR) guidelines were followed for the methodology and 
results compilation.
RESULTS: A total of 14 papers were included after applying the 
inclusion/exclusion criteria. Evidence from most of the studies (n = 9) 
indicates that regular consumption of vitamin D-fortified foods improves serum 
25(OH)D levels, enhances bone mineral density, and lowers the risk of fractures. 
However, different factors such as age, fortification dosage, and food vehicle 
(type of food) appear to affect the result outcomes.
CONCLUSION: The integration of vitamin D-fortified foods into daily diets 
appears to be a promising nutritional intervention strategy to support bone 
health and prevent osteoporosis. Further research is needed for standardization 
of recommendations for different age groups. Moreover, for countries like the 
UAE, where vitamin D deficiency remains prevalent and fortification is not yet 
standardized, adopting an evidence-based policy could be a decisive step toward 
mitigating future public health and economic burdens related to osteoporosis.

© 2025. The Author(s), under exclusive licence to the International Osteoporosis 
Foundation and the Bone Health and Osteoporosis Foundation.

DOI: 10.1007/s11657-025-01611-7
PMID: 40982112 [Indexed for MEDLINE]

Conflict of interest statement: Declarations. Conflicts of interest: None.


123. Front Pediatr. 2025 Sep 5;13:1658550. doi: 10.3389/fped.2025.1658550. 
eCollection 2025.

Transition phase of nutrition-optimizing nutrient administration.

Alur P(1), Ramarao S(2).

Author information:
(1)Department of Pediatrics, Penn State College of Medicine, Penn State Health, 
Hampden Medical Center, Enola, PA, United States.
(2)Department of Pediatrics, University of Mississippi Medical Center, Jackson, 
MS, United States.

This discussion explores the complex aspects of managing nutrition for preterm 
infants during the critical transition from exclusive parenteral nutrition (PN) 
to full enteral feeding (EN). The primary objectives of nutritional care in this 
very low birth weight infants (VLBW) population are to promote growth rates 
comparable to those in utero and enhance key neurodevelopmental milestones. The 
transition phase is characterized by the gradual increase of enteral feeds 
concurrently with the reduction and eventual cessation of parenteral nutrition. 
This period presents several key challenges in clinical practice, marked by 
notable variability: (A) determining the appropriate timing and criteria for 
initiating enteral feeds; (B) optimizing the rate at which enteral feed volumes 
are safely increased; (C) deciding the specific enteral volume threshold when to 
initiate human milk fortification to meet increasing caloric and protein 
demands; (D) establishing the optimal timing for discontinuing intravenous lipid 
emulsions (ILE); and (E) identifying the precise enteral volume threshold that 
dictates when to cease parenteral amino acid administration. To navigate these 
complexities and ensure seamless nutrient administration, practical 
recommendations for effective management are crucial. These include advocating 
for early fortification of human milk, judicious use of concentrated parenteral 
nutrition to provide adequate nutrients in lower fluid volumes, and 
strategically minimizing the overall duration of the transition phase. 
Implementing these evidence-informed steps aims to ensure smooth nutrition 
management, optimize nutrient delivery, and significantly reduce the pervasive 
risk of postnatal growth failure in extremely low birth weight infants.

© 2025 Alur and Ramarao.

DOI: 10.3389/fped.2025.1658550
PMCID: PMC12446270
PMID: 40980127

Conflict of interest statement: The authors declare that the research was 
conducted in the absence of any commercial or financial relationships that could 
be construed as a potential conflict of interest.


124. Adv Food Nutr Res. 2025;116:103-138. doi: 10.1016/bs.afnr.2025.04.010. Epub 2025 
May 19.

Current research on plant-based milk alternatives and impact on health.

Gilani SJ(1), Asif M(2), Fatima K(3).

Author information:
(1)Department of Pharmaceutical Sciences, College of Pharmacy, Princess Nourah 
bint Abdulrahman University, Riyadh, Saudi Arabia. Electronic address: 
.
(2)Faculty of Pharmacy, Lachoo Memorial College of Science and Technology, 
Jodhpur, Rajasthan, India. Electronic address:.
(3)Department of Pharmacology, Faculty of Pharmacy, Maulana Azad University, 
Jodhpur, Rajasthan, India.

The consumption of Plant-based Milk Alternatives (PBMAs) is experiencing rapid 
growth due to evolving food preferences, the rise of veganism, environmental 
concerns, and issues associated with animal-based milk such as lactose 
intolerance, milk protein allergy, and galactosemia. PBMAs are beverages derived 
from the extraction of cereals, nuts, seeds, and legumes, with examples 
including soy milk, almond milk, oat milk, and various others. Despite the 
numerous benefits of PBMAs, their nutritional profile differs slightly from 
animal-based milk, particularly in terms of calcium and protein content. 
Consequently, fortification of these alternatives using appropriate methods is 
necessary. The promotion of PBMAs is warranted due to their diverse advantages, 
including addressing problems related to animal-based milk, reducing economic 
burden, offering health benefits for conditions like diabetes and cardiovascular 
diseases, minimizing environmental impact, and reaching populations where 
livestock management is challenging. This review examines various aspects of 
PBMAs, including their nutritional profile, associated health benefits, 
bioavailability concerns, safety profile, and environmental considerations.

Copyright © 2025. Published by Elsevier Inc.

DOI: 10.1016/bs.afnr.2025.04.010
PMID: 40940086 [Indexed for MEDLINE]


125. Am J Clin Nutr. 2025 Nov;122(5):1161-1173. doi: 10.1016/j.ajcnut.2025.09.005. 
Epub 2025 Sep 10.

The effect of milk fat globule membrane phospholipid supplementation on 
cognitive function in older adults with subjective memory complaints-a 
randomized, double-blind, placebo-controlled trial.

Zajac IT(1), Kakoschke N(2), Evas S(3), Kuhn-Sherlock B(4), Wood K(2), King 
R(5), Arnoldy L(5), Dekker J(6), Gunn C(6), Fuad M(6), Scholey A(7), MacGibbon 
A(6), White DJ(5).

Author information:
(1)Health & Biosecurity, Commonwealth Scientific and Industrial Research 
Organisation, Adelaide, Australia; Flinders University, College of Education 
Psychology and Social Work, Adelaide, South Australia. Electronic address: 
.
(2)Health & Biosecurity, Commonwealth Scientific and Industrial Research 
Organisation, Adelaide, Australia.
(3)Health & Biosecurity, Commonwealth Scientific and Industrial Research 
Organisation, Adelaide, Australia; School of Psychology, Faculty of Health and 
Medical Sciences, University of Adelaide, Adelaide, Australia.
(4)BKS Consulting, Hamilton, New Zealand, Australia.
(5)Swinburne Neuroimaging, Centre for Mental Health and Brain Sciences, School 
of Health Sciences, Swinburne University of Technology, Melbourne, Australia.
(6)Fonterra Research and Development Centre, New Zealand, Australia.
(7)Swinburne Neuroimaging, Centre for Mental Health and Brain Sciences, School 
of Health Sciences, Swinburne University of Technology, Melbourne, Australia; 
Monash University, Nutrition Dietetics and Food, School of Clinical Sciences, 
Victoria, Australia; Division of Psychology, Northumbria University, 
Newcastle-upon-Tyne NE1 8ST UK.

BACKGROUND: Milk fat globule membrane (MFGM) phospholipids have been linked to 
improved cognitive function, especially in infants, but evidence in healthy 
older adults is limited.
OBJECTIVES: This project investigated MFGM phospholipid supplementation in older 
adults and focused on clinically meaningful cognitive function and other 
health-related outcomes.
METHODS: This study was a multicenter, repeated-measures, parallel groups, 
randomized, double-blind, placebo-controlled trial. A total of N = 263 
participants aged 55-75 y were randomly assigned to receive: 1) powdered milk 
fortified with low-dose MFGM (1.7-g phospholipid/d); 2) powdered milk fortified 
with high-dose MFGM (4.0-g phospholipid/d); or 3) rice starch powder (Control). 
Randomization used minimization to balance groups across study sites, sex, age, 
and Montreal Cognitive Assessment scores (MoCA). Participants consumed the 
product daily for 16 wk and attended their local study site at baseline, week 8, 
and endpoint. At each visit, they completed various cognitive, behavioral, 
biochemical, and physical measures.
RESULTS: For the primary cognitive outcome, Repeatable Battery for 
Neuropsychological Status (RBANS), there were no significant treatment effects, 
nor for the secondary cognitive outcome, Computerized Mental Performance 
Assessment System (COMPASS); however, alertness (Bond-Lader Visual Analogue 
Scales) was better maintained during cognitive testing at endpoint in the 
high-dose group. TNF-α was reduced in the high-dose group, whereas 
γ-aminobutyric acid (GABA) and serotonin increased in both MFGM groups. 
Cholesterol and low-density lipoprotein (LDL) were also reduced in both MFGM 
groups. Active B12 increased, and homocysteine tended to decrease in the 
high-dose group.
CONCLUSIONS: Although no significant effects were evident for cognitive 
function, subjective alertness during cognitive testing was improved in the 
high-dose group. Reduced inflammation, cholesterol, and LDL impacts for 
long-term cardiovascular health, linked with cognitive decline in aging. MFGM 
supplementation positively impacted active B12 (high and low dose) and 
homocysteine (high dose only), which have been associated with brain health. 
Longer study durations are needed to demonstrate cognitive benefits. This trial 
was registered at Australian New Zealand Clinical Trials Registry as 
ACTRN12620000270910; www.anzctr.org.au.

Crown Copyright © 2025. Published by Elsevier Inc. All rights reserved.

DOI: 10.1016/j.ajcnut.2025.09.005
PMCID: PMC12799387
PMID: 40939691 [Indexed for MEDLINE]

Conflict of interest statement: Conflict of interest The authors report no 
conflicts of interest.


126. Food Res Int. 2025 Nov;219:116905. doi: 10.1016/j.foodres.2025.116905. Epub 2025 
Jun 20.

Ferrous sulfate microparticles as a food fortification strategy: Application in 
plant-based yogurt and bioaccessibility assessment.

Rebellato AP(1), da Paixão Teixeira JL(2), Milani RF(2), Alvim ID(2), da Silva 
Santana TM(2), Galland F(2), Morgano MA(2).

Author information:
(1)Institute of Food Technology, Av. Brasil 2880, Jd. Chapadão, P.O. Box 139, 
Campinas, SP 13070-178, Brazil. Electronic address:.
(2)Institute of Food Technology, Av. Brasil 2880, Jd. Chapadão, P.O. Box 139, 
Campinas, SP 13070-178, Brazil.

This study evaluated the feasibility of using ferrous sulfate microparticles 
(FSM), produced through the combination of spray drying and spray chilling 
techniques, to fortify plant-based yogurt and increase dietary iron intake. The 
stability of FSM was assessed, and iron bioavailability was estimated using the 
standardized INFOGEST in vitro digestion method, followed by Caco-2 cell culture 
assays. FSM showed moisture content and water activity (Aw <0.3) comparable to 
control particles without iron (CP). Despite similar average particle sizes 
(D50), FSM exhibited lower luminescence and a more greenish-yellow hue than CP. 
The iron content in FSM and CP was 519 ± 23 mg/kg and 13.5 ± 0.8 mg/kg, 
respectively. FSM contained 309 mg of soluble iron/kg of particles, 
corresponding to 59.5 % bioaccessibility. After 180 days of storage, FSM showed 
slightly higher moisture, Aw, and color values compared to CP, but overall 
stability remained similar. Total and soluble iron contents did not 
significantly differ, confirming FSM's suitability for yogurt fortification. 
When both plant-based and dairy yogurts were fortified with FSM and isolated 
ferrous sulfate salt, total iron content ranged from 9.83 to 19.77 mg/kg, with 
bioaccessible iron ranging from 1.40 to 13.2 mg/kg. Bioaccessibility percentages 
varied depending on the type of iron (isolated vs. encapsulated), matrix origin 
(plant-based vs. dairy), and plant-based formulation. Notably, 
strawberry-flavored plant-based yogurt fortified with FSM showed higher 
bioavailability than yogurt fortified with isolated salt, emphasizing FSM's 
effectiveness in improving iron availability in plant-based yogurts.

Copyright © 2025 Elsevier Ltd. All rights reserved.

DOI: 10.1016/j.foodres.2025.116905
PMID: 40922140 [Indexed for MEDLINE]

Conflict of interest statement: Declaration of competing interest The authors 
declare that they have no known competing financial interests or personal 
relationships that could have appeared to influence the work reported in this 
paper.


127. PLoS One. 2025 Sep 8;20(9):e0331790. doi: 10.1371/journal.pone.0331790. 
eCollection 2025.

Cost-effectiveness of oil and milk fortification by scale for reducing Vitamin A 
and Vitamin D deficiency in India.

Kumar A(1), Ahluwalia K(2), Edathil AP(2), Sankar R(3), Arlappa N(4), Nair S(5), 
Duggal M(6), Joe W(2).

Author information:
(1)Department of Economics, School of Liberal Education, FLAME University, 
Lavale, Maharashtra, India.
(2)Institute of Economic Growth, Delhi University Enclave (North Campus), Delhi, 
India.
(3)Senior Advisor, Partnerships For Impact, Delhi, India.
(4)Division of Public Health Nutrition, ICMR-National Institute of Nutrition 
(ICMR-NIN), Tarnaka, Hyderabad, Telangana, India.
(5)Department of Foods and Nutrition, Faculty of Family and Community Sciences, 
M S University of Baroda, Vadodara, Gujarat, India.
(6)Indian Council of Medical Research, National Institute for Research in 
Reproductive and Child Health, Mumbai, Maharashtra, India.

BACKGROUND: Although broad-scale data might suggest low prevalence, millions of 
children in India still suffer from Vitamin A and Vitamin D deficiencies despite 
India's existing guidelines for Vitamin A deficiency. To address the issue, the 
Government of India has recommended fortification of oil and milk to improve 
Vitamin A and Vitamin D consumption. However, there is limited information on 
the health benefits and cost-effectiveness of fortifying oil and milk at scale.
OBJECTIVE: To estimate the health benefits and cost-effectiveness of 
supplementation programme and fortification of milk and oil among children under 
5 years, pregnant women, women in the reproductive age group, and the elderly.
DATA AND METHODS: To measure the health benefits associated with supplementation 
and fortification of oil and milk, the number of DALYs that are currently lost 
due to Vitamin A and Vitamin D deficiencies were estimated. For Vitamin A 
related mortality, a reduction of 4%, 12% and 23% were assumed while the 
assumptions for estimating morbidity benefits were derived from Global Burden of 
Disease. For the costing exercise, we considered the following two scenarios: 
(1) high-dose vitamin A supplementation for children and pregnant women; (2) 
industrial fortification of oil for children, pregnant women, women in the 
reproductive age group, and the elderly.
RESULTS: Overall, intervention related to Vitamin A could avert 1,119,044 Years 
of Life Lost (YLLs) at a 23% reduction, 194,616 YLLs at 4%, and 583,849 YLLs at 
12% and 28,534 YLDs. Intervention related to Vitamin D could avert 99,219 YLDs. 
The total cost for supplying supplements to approximately 109,965 thousand 
children and 26,920 thousand pregnant women is around 26 million USD. The cost 
to fortify is 7.6 million USD for oil and 9.8 million USD for milk fortification 
for children and women. The overall cost effectiveness ratio of the 
fortification programme is 150.
CONCLUSION: Fortification could emerge as a potentially superior long-term 
solution, considering the widespread consumption of oil and milk, offering a 
broader reach to the population.

Copyright: © 2025 Kumar et al. This is an open access article distributed under 
the terms of the Creative Commons Attribution License, which permits 
unrestricted use, distribution, and reproduction in any medium, provided the 
original author and source are credited.

DOI: 10.1371/journal.pone.0331790
PMCID: PMC12416695
PMID: 40920683 [Indexed for MEDLINE]

Conflict of interest statement: The authors have declared that no competing 
interests exist.


128. Biotechnol Appl Biochem. 2026 Apr;73(2):568-575. doi: 10.1002/bab.70047. Epub
