## Supplementary material for "Cross-LLM AI platform meta-research: Non-inferiority of bovine milk-based fortifiers to human milk-based fortifiers": SI2_130.Meta&SystematicR.abstract.txt

1. Nutr Clin Pract. 2026 Apr 29. doi: 10.1002/ncp.70127. Online ahead of print.

Growth and human milk intake outcomes from postdischarge feedings among 
very-low-birth-weight preterm infants: A systematic review.

Rebele T(1), Gomes A(1), Newkirk M(1), Ghajar M(1), Byham-Gray L(1), Ziegler 
J(1).

Author information:
(1)Department of Clinical and Preventive Nutrition Sciences, Rutgers, The State 
University of New Jersey, New Brunswick, New Jersey, USA.

BACKGROUND: Preterm infants of <34 weeks gestation or <1500 g are at risk for 
postnatal growth failure and often require nutrition support after discharge. 
Despite expert recommendations for postdischarge human milk (HM) fortification 
or nutrient-enriched formula feeding, the effects of current feeding strategies 
on growth and HM intake remain unclear. This systematic review evaluated the 
impact of postdischarge fortified expressed HM or direct breastfeeding (BF) with 
postdischarge formula (PDF) bottle feeds compared to unfortified HM or BF with 
standard term formula bottle feeds on growth and HM intake outcomes through 6 
and 12 months corrected age (CA).
METHODS: A systematic search of eight databases and grey literature sources was 
conducted for studies published in English since 2012 identifying 3146 studies. 
Risk of bias was assessed using standardized Cochrane RoB 2 and ROBINS-I tools. 
Meta-analysis was not conducted due to heterogeneity in interventions, outcome 
timing, and outcome metrics; results were narratively synthesized.
RESULTS: Four studies (n = 2 randomized controlled trials, n = 2 cohorts), 
including 548 preterm infants, met eligibility criteria. Two studies showed 
significantly greater weight gain, and one showed significantly greater head 
circumference in the fortified groups at 6 months CA compared to the unfortified 
groups. Length differences were not significant. Three studies reporting HM 
intake found comparable or greater HM intake in the fortified groups.
CONCLUSION: Current evidence suggests that postdischarge fortified HM or direct 
BF with supplemental PDF bottle feeds may support improved weight and head 
circumference outcomes without compromising HM intake. However, methodological 
variability highlights the need for standardized, prospective studies.

© 2026 The Author(s). Nutrition in Clinical Practice published by Wiley 
Periodicals LLC on behalf of American Society for Parenteral and Enteral 
Nutrition.

DOI: 10.1002/ncp.70127
PMID: 42051106


2. Food Res Int. 2026 Jun 1;233(Pt 2):118943. doi: 10.1016/j.foodres.2026.118943. 
Epub 2026 Mar 15.

Global evidence on vitamins in human milk: A 2002-2022 systematic review with a 
focus on China and implications for maternal diet and infant formula.

Yang Y(1), Ni X(1), Liu N(1), Li Q(1), Fang Q(1), Szeto IM(2), Duan S(2), Wei 
T(1), Rao T(1), Deng Z(3), Li T(4), Li J(5).

Author information:
(1)State Key Laboratory of Food Science and Resources, Nanchang University, 
Nanchang, Jiangxi 330047, China.
(2)National Center of Technology Innovation for Dairy, Hohhot 010110, China; 
Inner Mongolia Yili Industrial Group Co., Ltd, Hohhot, PR China.
(3)State Key Laboratory of Food Science and Resources, Nanchang University, 
Nanchang, Jiangxi 330047, China; International Institute of Food Innovation, 
Nanchang University, Nanchang, Jiangxi 330051, China. Electronic address: 
.
(4)Inner Mongolia Dairy Technology Research Institute Co. Ltd., Hohhot 010110, 
China; National Center of Technology Innovation for Dairy, Hohhot 010110, China. 
Electronic address:.
(5)State Key Laboratory of Food Science and Resources, Nanchang University, 
Nanchang, Jiangxi 330047, China; International Institute of Food Innovation, 
Nanchang University, Nanchang, Jiangxi 330051, China. Electronic address: 
.

Current knowledge on vitamin composition of human milk in China remains 
fragmented, with most evidence derived from small-scale studies focusing on 
individual vitamins. To provide a comprehensive perspective relevant to 
nutrition science and infant formula design, this study systematically reviewed 
global trends in human milk vitamins over the past two decades (2002-2022), with 
attention to regional disparities, maternal dietary patterns, and local 
standards. Literature was retrieved from PubMed, Web of Science, ScienceDirect, 
Chongqing VIP Information, China National Knowledge Infrastructure (CNKI), and 
Wanfang databases following predefined inclusion criteria. Fifty-seven studies 
encompassing 9833 human milk samples from 33 countries were included. Across 
lactation, concentrations of water-soluble vitamins generally increased, whereas 
fat-soluble vitamins A (VA) and E (VE) exhibited a declining tendency. Compared 
with Chinese samples, non-Chinese human milk contained higher levels of vitamin 
B1 (VB1), vitamin B6 (VB6), VA, and VE; while vitamin B5 (VB5) was consistently 
higher in Chinese human milk. Temporal analysis within China revealed elevated 
levels of VA (2016-2017), B vitamins (2014), and VE (2022), likely reflecting 
shifts in maternal dietary intake and nutritional awareness. Concentrations of 
VA, VE, VB5, and vitamin VB7 (VB7) in Chinese human milk met national adequate 
intake (AI) recommendations for infants aged 0-6 months. However, these 
concentrations remained substantially lower than the levels specified in China's 
national standard for infant formula. Notably, vitamin D (VD) emerged as the 
nutrient most vulnerable to inadequacy among exclusively breastfed infants, 
indicating a persistent gap between physiological supply from human milk and 
recommended intake. These findings emphasize the need for nationally 
representative, lactation-stage-specific reference ranges for human milk 
vitamins in China. Such evidence is essential for optimizing maternal dietary 
strategies, for guiding rational vitamin fortification, and formulation of 
next-generation infant formulas that better reflect physiological nutrition.

Copyright © 2026 Elsevier Ltd. All rights reserved.

DOI: 10.1016/j.foodres.2026.118943
PMID: 41956646 [Indexed for MEDLINE]

Conflict of interest statement: Declaration of competing interest The authors 
declare that they have no known competing financial interests or personal 
relationships that could have appeared to influence the work reported in this 
paper.


3. Food Chem. 2026 Apr 15;508(Pt A):148316. doi: 10.1016/j.foodchem.2026.148316. 
Epub 2026 Feb 4.

Evaluating amino acid fortification in low-dairy ready-to-use therapeutic food 
formulations: Insights from a meta-analysis re-evaluation.

Berhane M(1), Toni AT(2), Gonzales GB(3).

Author information:
(1)Jimma University Clinical and Nutrition Research Center (JUCAN), Jimma 
University, Jimma P.O Box: 378, Ethiopia; Department of Pediatrics and Child 
Health, Faculty of Medical Sciences, Institute of Health, Jimma University, 
Jimma, Ethiopia; Department of Public Health and Primary Care, Faculty of 
Medicine and Health Sciences, Ghent University, C. Heymanslaan 10, 9000 Ghent, 
Belgium.
(2)Department of Pediatrics and Child Health, College of Medicine and Health 
Sciences, University of Gondar, Gondar, Ethiopia; Division of Human Nutrition 
and Health, Wageningen University and Research, Wageningen, the Netherlands.
(3)Department of Public Health and Primary Care, Faculty of Medicine and Health 
Sciences, Ghent University, C. Heymanslaan 10, 9000 Ghent, Belgium. Electronic 
address:.

Severe wasting is treated with ready-to-use therapeutic food (RUTF), wherein the 
protein content primarily comes from milk and peanuts. As these ingredients are 
the main drivers of RUTF's cost, global initiatives seek alternatives using 
locally available, sustainable ingredients to reduce cost and increase coverage. 
To compensate for reduced/removed dairy protein, current practice is to add 
amino acids to meet Codex protein quality requirements. In this meta-analysis 
re-evaluation, we compared RUTF formulations with <50% dairy protein, with or 
without amino acid fortification, against standard milk-peanut RUTF for the 
outcome of weight gain. Growth outcomes were similar between fortified and 
unfortified RUTFs when compared to standard RUTF. These findings suggest that 
elevating protein quality scores by adding amino acids may be insufficient to 
ensure clinical efficacy in novel RUTF formulations. Additionally, the findings 
suggest that Protein Digestibility Corrected Amino Acid Score (PDCAAS) 
compliance alone may not guarantee clinical efficacy. Future formulation efforts 
should address digestibility, antinutritional factors, and increasing total 
protein content to achieve effective, sustainable novel RUTF formulations.

Copyright © 2026 Elsevier Ltd. All rights reserved.

DOI: 10.1016/j.foodchem.2026.148316
PMID: 41672015 [Indexed for MEDLINE]

Conflict of interest statement: Declaration of competing interest The authors 
declare that they have no known competing financial interests or personal 
relationships that could have appeared to influence the work reported in this 
paper.


4. Arch Dis Child Fetal Neonatal Ed. 2026 Jan 19:fetalneonatal-2024-327677. doi: 
10.1136/archdischild-2024-327677. Online ahead of print.

Effect of different feeding approaches on growth, neonatal morbidities, 
mortality and neurodevelopmental outcome in preterm infants: a systematic review 
and network meta-analysis.

Hamouda N(1)(2), AboEL-Azm Y(3)(4), Elsamman K(3)(5), Nabil S(3)(6), Tarek 
M(3)(7), Wasia FE(3)(8), Amin AM(3)(7), Elshahat A(3)(7), Hassan N(3)(9), 
Mektebi A(3)(10), Khaled A(9), Shahin HN(11), Aldemerdash MA(3)(12), Refaey 
N(3)(13), Khalil M(3)(9), Hendi NI(3)(14), Hafez S(3)(15), Alabdallat YJ(3)(16), 
Abdellatif M(3)(17).

Author information:
(1)Negida Research Academy, Arlington, Virginia, USA 
.
(2)NICU, El Sahel Teaching Hospital, Cairo, Egypt.
(3)Negida Research Academy, Arlington, Virginia, USA.
(4)Zagazig University Faculty Medicine, Zagazig, Ash Sharqia Governorate, Egypt.
(5)Biochemistry and Nutrition, Faculty of Women for Science, Ain Shams 
University, Cairo, Egypt.
(6)Internal Medicine, Beni Suef University Hospital, Beni Suef, Beni Suef 
Governorate, Egypt.
(7)Al-Azhar University Faculty of Medicine, Nasr City, Cairo, Egypt.
(8)Faculty of Medicine Kafrelsheikh University, Kafr El-sheikh, Egypt.
(9)Zagazig University Faculty of Human Medicine, Zagazig, Egypt.
(10)Faculty of Medicine Kutahya Health Science University, Kutahya, Turkey.
(11)South Valley University Faculty of Medicine, Qena, Egypt.
(12)Sohag University Faculty of Medicine, Sohag, Egypt.
(13)Physical Therapy for Women's Health, Cairo University Faculty of Physical 
Therapy, Dokki, Giza, Egypt.
(14)Ain Shams University Faculty of Medicine, Cairo, Egypt.
(15)Minia University Faculty of Medicine, Minia, Egypt.
(16)The Hashemite University Faculty of Medicine, Zarqa, Zarqa, Jordan.
(17)Pediatrics and Neonatology, Muscat Private Hospital, Muscat, Muscat, Oman.

OBJECTIVE: To compare feeding strategies on preterm infants' growth during 
hospitalisation, neonatal morbidities, mortality and neurodevelopmental outcome 
(NDO) at 18-26 months corrected age.
METHODS: We followed the Preferred Reporting Items for Systematic Reviews and 
Meta-Analyses extension statement for network meta-analysis . We searched five 
medical databases for randomised controlled trials comparing different feeding 
approaches in preterm infants and their effects on growth, neonatal morbidities, 
mortality and NDO. The Cochrane Collaboration's tool was used to assess the risk 
of bias. We used a random-effects model. Pooled mean differences (MD) or risk 
ratios with 95% CIs were calculated.
RESULTS: Ninety-five studies (9663 infants) were included.Human milk (HM) with 
bovine milk fortifier (BMF) (adjusted according to blood urea nitrogen) achieved 
the best length increment (MD=0.56 cm/week; 95% CI 0.19 to 0.93). Notably, 
HM+BMF (3.5 gm/kg/d protein) showed the best head circumference growth (MD 
0.46 cm/week; 95% CI 0.10 to 0.81) but no significant difference in weight gain. 
There were no significant differences in neonatal morbidities/mortality. While 
MOM|+PTF (supp) displayed significantly lower NDO delay in the domain of mild 
cognitive delay.
CONCLUSION: Overall, there is a lack of strong evidence to support a specific 
enteral feeding strategy and further high-quality research is required. Targeted 
HM fortification appears to improve head growth, while adjusted fortification 
enhances length. Given the significant inconsistency detected, which may 
compromise the reliability of the network estimates, these results must be 
interpreted carefully.

© Author(s) (or their employer(s)) 2026. No commercial re-use. See rights and 
permissions. Published by BMJ Group.

DOI: 10.1136/archdischild-2024-327677
PMID: 41554628

Conflict of interest statement: Competing interests: None declared.


5. Dietary Total Fat Intake and Dietary Polyunsaturated Fatty Acid Intake and Child 
Growth and Development Outcomes: Systematic Review [Internet].

Jungbauer RM, Stadler DD, Hsu F, Gillingham M, Yu Y, Fu R, Alex Foster B, Wood 
EK, Mackiewicz-Seghete K, O'Reilly CD, Totten AM.

Rockville (MD): Agency for Healthcare Research and Quality (US); 2026 Jan. 
Report No.: 26-EHC004.

BACKGROUND: Dietary fats are an important source of energy and essential fatty 
acids (linoleic acid and alpha-linolenic acid), and serve as a medium for 
fat-soluble vitamins (vitamins A, D, E, and K) absorption, transport, function, 
and storage. As part of a nutritionally complete and balanced diet, dietary fats 
support optimal fetal growth and development during pregnancy, and physical 
growth and cognitive development throughout infancy and childhood. Based on 
studies demonstrating reduced rates of preterm birth (<37 weeks of gestational 
age, GA) and early preterm birth (<34 weeks GA) among individuals consuming 
higher amounts of docosahexaenoic acid (DHA), an omega-3 polyunsaturated fatty 
acid (PUFA), during pregnancy, DHA was added to some prenatal vitamin 
supplements. Similarly, in 2002, voluntary fortification of infant formulas with 
DHA in amounts similar to those in mature human milk (0.2 to 0.5% by weight of 
total fatty acids) and arachidonic acid (ARA), also an omega-3 PUFA, at or above 
the amount of DHA, was approved by the U.S. Food and Drug Administration to 
support infant brain development and visual acuity. To inform updates to and 
expansion of the 2005 U.S.-Canadian Dietary Reference Intakes (DRIs), this 
systematic review was designed to identify and summarize the evidence published 
since 1990 on the relationships between consumption of omega-6 and/or omega-3 
(n-6 and n-3, respectively) PUFAs during pregnancy and risk of preterm birth 
(Key Question 1); between intake during pregnancy and/or lactation and infant 
and child growth and development (Key Question 2); and between intake of total 
fat (Key Question 3) or n-6 and/or n-3 PUFA (Key Question 4) by children between 
birth through 18 years of age and growth and developmental outcomes.
METHODS: A medical research librarian designed and conducted a search of Ovid 
MEDLINE®, CINAHL®, and Cochrane CENTRAL from 1990 through August 2025. 
Researchers manually reviewed reference lists and responses to a Federal 
Register Notice request for information. Dual, independent review of abstracts 
and full-text articles was conducted using a priori specified inclusion and 
exclusion criteria. Established methods were used in dual, independent 
assessment of risk of bias. Data were synthesized quantitatively using 
meta-analyses when possible; randomized controlled trials (RCTs) and 
nonrandomized studies (NRS) not included in the meta-analyses were summarized 
qualitatively in tables and text. For each set of analyses with sufficient data, 
we assessed the strength of evidence (SOE) using set criteria and applicability 
of findings to a generally healthy population from the United States and Canada.
RESULTS: Searches identified 8,929 abstracts. Eighty-six studies in 126 
publications were included across the four Key Questions. Of all the 
publications, 108 described RCTs and 18 described NRS; over half were rated high 
risk of bias (k=66, 52%), followed by moderate risk of bias (k=39, 31%) and low 
risk of bias ratings (k=21, 17%). The populations studied were generally 
healthy, with no medical conditions impacting fat consumption, digestion, 
absorption, or metabolism, or unusual dietary patterns. In the 86 parent or 
standalone studies, the majority (k=61, 71%) assessed the effects of dietary 
supplementation on outcomes, followed by diet only (k=22, 26%), and then a 
combination of diet and dietary supplementation (k=3, 4%). The bulk of the 
evidence centered on supplement studies during pregnancy or lactation and growth 
outcomes measured from birth to 6 months of age. The duration of interventional 
exposure was generally less than 6 months, and only 4 studies followed offspring 
into early adolescence (up to 13 years). Overall, supplemental PUFA or total fat 
intake in healthy populations resulted in similar risk of preterm birth and 
growth and developmental outcomes when compared with differing amounts or no 
additional amount of PUFA and total fat according to feeding guidelines. There 
was low SOE of no difference for PUFA supplementation above general dietary 
consumption and risk of preterm birth <34 weeks or <37 weeks, growth outcomes 
(weight, length [height], head circumference), or developmental outcomes 
(cognition, movement, language, social emotional). The exception was studies 
evaluating n-3 PUFA and n-3+n-6 PUFA supplementation, with consumption of only 
n-3 PUFA during pregnancy associated with lower risk of preterm birth <37 weeks, 
and higher weight and length at birth compared with consumption of n-3+n-6 PUFA 
supplements. There was insufficient or no evidence linking most growth and 
developmental outcomes to total fat intake according to infant and child feeding 
guidelines among infants consuming adequate energy.
CONCLUSIONS: Supplementation of PUFA and higher total fat intake in healthy 
populations, at the amounts studied, may not confer demonstrable advantages over 
baseline intake or exposure, and the consistent finding of no difference 
suggests no harm. There were few findings of differences across PUFA 
supplementation or dietary intake of fatty acids and growth and developmental 
outcomes in a healthy population during pregnancy and childhood. The largest 
body of evidence consisted of RCTs with moderate or high risk of bias evaluating 
PUFA supplementation during pregnancy or infancy and preterm birth and growth 
outcomes in the first year of life. Of outcomes with sufficient evidence, 
heterogeneity in study design, exposures and comparators, and outcomes resulted 
in the SOE being rated mostly as low. Additional research in older children and 
adolescents, studies enrolling people during pregnancy with low DHA 
concentrations at baseline, studies with earlier initiation during pregnancy and 
longer exposure durations, and populations who inherently consume diets low in 
PUFA or who have nutrition-related conditions that may benefit from supplemental 
PUFA or higher total fat consumption is needed to inform future DRIs.

DOI: 10.23970/AHRQEPCSR_DIETARYFATCHILD
PMID: 42013221


6. Int Breastfeed J. 2025 Dec 25;21(1):9. doi: 10.1186/s13006-025-00801-w.

Impact of three mammalian milk-derived fortifiers on morbidity and mortality in 
preterm infants: a systematic review and network meta-analysis.

Feng Y(#)(1), Gao Y(#)(1), Gu Y(1), Liu Y(1), Xu Y(2), Li S(3).

Author information:
(1)School of Nursing, Guilin Medical University, No.109 Huancheng North 2nd 
Road, Qixing District, Guilin, Guangxi Zhuang Autonomous Region, China.
(2)Affiliated Hospital of Guilin Medical University, No.15 Lequn Road, Diecai 
District, Guilin, Guangxi Zhuang Autonomous Region, China.
(3)School of Nursing, Guilin Medical University, No.109 Huancheng North 2nd 
Road, Qixing District, Guilin, Guangxi Zhuang Autonomous Region, China. 
.
(#)Contributed equally

BACKGROUND: Fortification of breast milk with milk-derived fortifiers is 
essential for the growth and developmental outcomes of preterm infants, yet the 
ideal fortifier remains uncertain. This network meta-analysis assessed the 
effects of bovine milk-derived fortifier (BMF), human milk-derived fortifier 
(HMF), and donkey milk-derived fortifier (DMF) on morbidity and mortality rates 
of preterm infants within neonatal intensive care units (NICUs).
METHODS: We conducted a comprehensive search of multiple databases, including 
MEDLINE, CINAHL, PubMed, CNKI, and the Cochrane Central Register of Controlled 
Trials. The risk of bias for each study was evaluated using the Cochrane Risk of 
Bias 2 (RoB 2) tool.
RESULTS: Ten studies involving 545 preterm infants were identified for 
qualitative descriptive analysis. network meta-analysis was conducted, 
incorporating data from five studies and three different types of milk-based 
fortifiers. The network meta-analysis suggested that HMF may be associated with 
a lower incidence of necrotizing enterocolitis. Compared with HMF, BMF was 
associated with an increased risk of necrotizing enterocolitis (RR = 2.33, 95% 
CI [1.22–4.44]). Cumulative ranking probabilities indicated that HMF appeared to 
be the most favourable fortifier for reducing both necrotizing enterocolitis 
incidence and mortality. DMF might represent a potential option for reducing the 
risk of intraventricular haemorrhage and neonatal sepsis.
CONCLUSION: HMF may be associated with a reduced incidence of necrotizing 
enterocolitis, whereas DMF may offer potential anti-inflammatory benefits, 
particularly for high-risk populations susceptible to intraventricular 
haemorrhage. This study provides direct and comparative evidence of the 
associations between different animal-derived fortifiers and morbidity and 
mortality in preterm infants. It provides a preliminary evaluation of DMF, for 
which limited evidence previously existed, and offers insight into the potential 
mechanisms that connect milk composition to clinical outcomes. However, these 
findings are primarily based on a limited number of studies with small sample 
sizes. Further randomized controlled trials are necessary to validate these 
observations.
SUPPLEMENTARY INFORMATION: The online version contains supplementary material 
available at 10.1186/s13006-025-00801-w.

DOI: 10.1186/s13006-025-00801-w
PMCID: PMC12849099
PMID: 41449441

Conflict of interest statement: Declarations. Ethics approval and consent to 
participate: Not applicable. Consent for publication: Not applicable. Competing 
interests: The authors declare no competing interests.


7. Nutrients. 2025 Nov 30;17(23):3765. doi: 10.3390/nu17233765.

Dairy Intake and Iodine Status in Pregnant and Lactating Women: A Systematic 
Review and Meta-Analysis.

Razmpoosh E(1), Geronimo K(1), Gauntlett C(1), Vicente I(1), Pham P(1), Shah 
A(1), Musa-Veloso K(1).

Author information:
(1)Intertek Health Sciences Inc., 2233 Argentia Road, Suite 201, Mississauga, ON 
L5N 2X7, Canada.

Background/Objectives: Despite salt iodization, iodine deficiency during 
pregnancy and lactation is re-emerging in many industrialized countries, 
necessitating an evaluation of the role of dairy in supporting iodine status 
during these critical periods. Methods: We conducted a systematic review and 
meta-analysis in accordance with PRISMA guidelines. Ten databases were searched 
through March 2025 using ProQuest Dialog™ (Version 75.0). Study quality was 
assessed using either the Quality Evaluation for Observational Studies tool or 
the Risk of Bias 2 tool, depending on study design. Random-effects models were 
applied, with certainty of evidence rated using the GRADE framework. Publication 
bias, sensitivity analyses, and subgroup analyses were also performed. Results: 
Fifty-one publications met the eligibility criteria, including 50 publications 
of observational studies and 1 publication of a randomized controlled trial 
(RCT), with most studies conducted on pregnant women. Higher dairy intake was 
associated with significantly greater urinary iodine concentration (UIC), (23 
studies; standardized mean difference: 0.326; 95% confidence interval [CI]: 
0.228, 0.424; p < 0.001; I2 = 57.31%; low-certainty), and significantly lower 
odds of iodine deficiency (11 studies; odds ratio: 0.58; 95% CI: 0.48, 0.70; p < 
0.001; I2 = 0%; moderate-certainty). Associations were stronger in studies 
conducted during later pregnancy, in higher-quality studies, and when the dairy 
food was specifically milk. Dairy contributed to ~27% of iodine intake from 
foods. Breast milk iodine concentration (BMIC) findings were inconsistent, 
though in one RCT, iodine-fortified milk improved BMIC and UIC. Conclusions: 
Dairy intake supports adequate iodine status during pregnancy and lactation. 
RCTs would be valuable in further investigating the role of dairy in supporting 
iodine status, particularly in lactating women. (PROSPERO CRD420251054576).

DOI: 10.3390/nu17233765
PMCID: PMC12693841
PMID: 41374057 [Indexed for MEDLINE]

Conflict of interest statement: The authors are employees of Intertek Health 
Sciences Inc., which has provided, and continues to provide, paid services to 
Dairy Management Inc. The work presented in this manuscript was funded by the 
National Dairy Council, which is managed by Dairy Management Inc. Neither the 
National Dairy Council nor Dairy Management Inc. had any role in the design of 
the study; in the collection, analyses, or interpretation of data; in the 
writing of the manuscript; or in the decision to publish the manuscript.


8. Nutrients. 2025 Nov 29;17(23):3757. doi: 10.3390/nu17233757.

Is Vitamin D Fortification of Dairy Products Effective for Improving Vitamin D 
Status? A Systematic Review and Meta-Analysis of Randomised Controlled Trials.

Wong CL(1), Givens DI(2), Turpeinen AM(3), Liu X(1), Guo J(1).

Author information:
(1)School of Food Science and Nutrition, Faculty of Environment, University of 
Leeds, Leeds LS2 9JT, UK.
(2)Institute for Food, Nutrition and Health, University of Reading, Reading RG6 
6EU, UK.
(3)Valio Ltd., R&D, 00370 Helsinki, Finland.

Background/objectives: Given the suboptimal vitamin D intake and status among 
the United Kingdom population, the Scientific Advisory Committee on Nutrition is 
seeking suitable food vehicles for vitamin D fortification. Thus, this study 
aimed to examine the efficacy of vitamin D-fortified dairy products in improving 
serum 25-hydroxyvitamin D [25(OH)D] concentration using data from randomised 
controlled trials (RCTs). Methods: PubMed, Embase, and Web of Science were 
searched from inception until October 2024. Studies were included if they were 
RCTs with intervention groups administered vitamin D-fortified dairy products 
and control groups administered unfortified dairy products, as well as examining 
the effects on serum 25(OH)D concentration. Results: There were 35 RCTs eligible 
for inclusion, involving 4965 participants (intervention: 2526; control: 2439). 
The results showed that serum 25(OH)D concentrations were significantly 
increased by vitamin D3-fortified milk/milk powder (n = 15, mean difference 
(MD): 18.31 nmol/L, 95% confidence interval (CI): 13.30-23.33 nmol/L, I2 = 95%), 
vitamin D3-fortified yoghurt/yoghurt drinks (n = 11, MD: 26.22 nmol/L, 95% CI: 
18.67-33.77 nmol/L, I2 = 97%), vitamin D2-fortified milk/milk powder (n = 3, MD: 
11.61 nmol/L, 95% CI: 9.31-13.91 nmol/L, I2 = 0%), vitamin D-fortified (type not 
specified) milk/milk powder (n = 8, MD: 13.59 nmol/L, 95% CI: 8.54-18.64, I2 = 
98%), and vitamin D-fortified (type not specified) yoghurt/yoghurt drinks (n = 
4, MD: 27.74 nmol/L, 95% CI: 16.83-38.64 nmol/L, I2 = 91%), but insignificantly 
increased by vitamin D3-fortified cheese (n = 5, MD: 16.78 nmol/L, 95% CI: 
-3.61-37.16, I2 = 99%). However, the results of vitamin D3-fortified cheese 
became significant when leave-one-out analysis was performed by omitting one RCT 
(MD: 24.13 nmol/L, 95% CI: 4.69-43.58, I2 = 90%). Conclusions: These findings 
provide evidence that vitamin D-fortified dairy products have the potential to 
improve serum 25(OH)D concentrations in populations.

DOI: 10.3390/nu17233757
PMCID: PMC12694000
PMID: 41374048 [Indexed for MEDLINE]

Conflict of interest statement: The authors declare that this research was 
supported by a grant from the Consumer Lab funded by the Biotechnology and 
Biological Sciences Research Council, which provided financial support for 
workshops and the employment of a research assistant (C.L.W.). The funding body 
had no role in the study design, data collection and analysis, decision to 
publish, or preparation of the manuscript. The authors also declare that A.T., 
from Valio Ltd. (Finland), was our formal partner, providing industry input and 
in-kind analytical support. Author D.I.G. also declares that he has received 
travel expenses and honoraria in connection with lectures and meetings from the 
UK Dairy Council (now Dairy UK), the Dutch Dairy Association, the European Dairy 
Association, the International Dairy Federation, Centre National 
Interprofessionnel de l’Industrie Laitière, and the US Dairy Research Institute.


9. Nutrients. 2025 Oct 28;17(21):3384. doi: 10.3390/nu17213384.

Human Milk Fortification and Necrotizing Enterocolitis in Very Low Birthweight 
Infants: State of Evidence and Systematic Review with Meta-Analysis.

Reyes SM(1), Paul TL(1), Ferry J(2).

Author information:
(1)Rev Bioscience, LLC, Boise, ID 83712, USA.
(2)Pediatrix Medical Group Tampa Regional Practices, St. Joseph's Women's 
Hospital, Tampa, FL 33607, USA.

Background: Necrotizing enterocolitis (NEC) remains a leading cause of morbidity 
and mortality in very low birthweight (VLBW) infants. Human milk feeding and 
standardized feeding protocols are protective, but clinical practice varies, 
particularly in fortifier choice. Whether human milk-derived fortifiers reduce 
NEC risk compared with cow milk-derived fortifiers remains unclear. Methods: We 
conducted a systematic state-of-evidence review and meta-analysis, searching 
PubMed, Web of Science, and Scopus through July 2025. Eligible studies included 
RCTs and observational cohorts of VLBW infants comparing an exclusive human milk 
diet (EHMD) including human milk-derived fortifiers to cow milk-derived diets. 
Two reviewers independently screened and extracted data. Both RCTs and 
observational studies were included to evaluate consistency of effect estimates 
across designs and to account for heterogeneity in control group feeding 
practices. Pooled odds ratios (ORs) with 95% CIs were calculated using a 
Sidik-Jonkman random-effects model. Sensitivity analyses by study design and 
exclusion of infant formula from controls were performed. Results: Twenty 
studies (five RCTs, 15 observational; n = 6794 infants) met inclusion criteria, 
most enrolling infants born ≤1250 g. Compared with cow milk-containing diets, 
EHMD was associated with lower odds of Bell Stage ≥ 2 NEC (OR: 0.59; 95% CI: 
0.42, 0.81; p < 0.001; n = 4625) and surgical NEC (OR: 0.43; 95% CI: 0.32, 0.58; 
p < 0.0001; n = 4754). In direct comparisons of fortifier type with a base diet 
of human milk, estimates suggested lower odds of Bell Stage ≥ 2 NEC by 35% (OR: 
0.65; 95% CI: 0.44, 0.97; p = 0.03, n = 2102) and surgical NEC by 49% (OR: 0.51; 
95% CI: 0.26, 0.98; p = 0.04; n = 1659) with human milk-derived fortifiers. 
Effect estimates were generally consistent across study designs, although 
precision and statistical significance varied. Conclusions: EHMD with human 
milk-derived fortifiers was associated with lower odds of medical and surgical 
NEC in VLBW infants, with most evidence from infants born ≤1250 g, reflecting 
current clinical use in the highest-risk population. Although the number and 
sample sizes of RCTs remain limited, the consistency of effect estimates across 
both RCTs and observational studies, together with significance of pooled 
analyses, strengthens confidence in these findings. Pragmatic and registry-based 
studies using standardized fortification protocols may provide the most 
efficient pathway to strengthen the evidence base.

DOI: 10.3390/nu17213384
PMCID: PMC12609769
PMID: 41228457 [Indexed for MEDLINE]

Conflict of interest statement: Support for this work was as follows: S.M.R. 
serves as a consultant for Prolacta Bioscience and received financial support to 
conduct the research and write the manuscript. T.L.P. serves as a consultant for 
Rev Bioscience and received financial support to help conduct the research. J.F. 
does not have any conflicts of interest to disclose.


10. Acta Paediatr. 2025 Dec;114(12):3309-3316. doi: 10.1111/apa.70260. Epub 2025 Jul 
29.

Absence of Evidence of Beneficial Effects of Human Milk-Based Fortifier: A 
Bayesian Model-Averaged Meta-Analysis.

Huizing MJ(1), Vizzari G(2), Bartoš F(3), Cavallaro G(2), Gianni ML(2)(4), 
Villamor E(1).

Author information:
(1)Division of Neonatology, Department of Pediatrics, MosaKids Children's 
Hospital, Maastricht University Medical Center (MUMC+), School for Oncology and 
Reproduction (GROW), Maastricht University, Maastricht, the Netherlands.
(2)Fondazione IRCCS ca' Granda Ospedale Maggiore Policlinico, NICU, Milan, 
Italy.
(3)Department of Psychology, University of Amsterdam, Amsterdam, the 
Netherlands.
(4)Department of Clinical Sciences and Community Health, University of Milan, 
Milan, Italy.

AIM: Human milk-based fortifiers (HMBFs) have been adopted in neonatal care 
despite limited efficacy data. Our objective was to conduct a Bayesian 
re-analysis of the current evidence on the protective effect of HMBF against 
necrotising enterocolitis (NEC, stage II-III).
METHODS: We searched for randomised controlled trials (RCTs) comparing HMBF 
versus bovine milk-based fortifier (BMBF) in infants with gestational age 
< 32 weeks or birthweight < 1500 g. Bayesian model-averaged (BMA) meta-analysis 
was used to calculate Bayes factors (BFs). The BF10 is the ratio of the 
probability of the data under the alternative hypothesis (H1, presence of 
effect) over the probability of the data under the null hypothesis (H0, absence 
of effect).
RESULTS: We included 3 RCTs (405 infants). The BMA analysis did not show 
conclusive evidence in favour of H1 (BF10 > 3) or in favour of H0 (BF10 < 1/3) 
for NEC (BF10 = 0.86), mortality (BF10 = 1.07), late onset sepsis (BF10 = 0.69), 
bronchopulmonary dysplasia (BPD, BF10 = 0.92), or severe retinopathy of 
prematurity (ROP, BF10 = 0.75).
CONCLUSIONS: The extant corpus of RCTs does not provide sufficient evidence to 
either confirm or exclude a potential effect of HMBF compared with BMBF on the 
risk of NEC, mortality, sepsis, BPD, or severe ROP.

© 2025 The Author(s). Acta Paediatrica published by John Wiley & Sons Ltd on 
behalf of Foundation Acta Paediatrica.

DOI: 10.1111/apa.70260
PMCID: PMC12599611
PMID: 40729426 [Indexed for MEDLINE]

Conflict of interest statement: The authors declare no conflicts of interest.


11. J Neonatal Perinatal Med. 2026 Jan;19(1):10-21. doi: 10.1177/19345798251350987. 
Epub 2025 Jun 19.

Human breast milk fortification with human milk fortifier vs preterm infant 
formula: A systematic review and meta-analysis.

Aryadevi NNB(1), Putri RSI(1), Marsubrin PMT(1)(2)(3).

Author information:
(1)Faculty of Medicine, Universitas Indonesia, Jakarta, Indonesia.
(2)Department of Child Health, Faculty of Medicine, Dr. Cipto Mangunkusumo 
National General Hospital, Universitas Indonesia, Jakarta, Indonesia.
(3)Universitas Indonesia Hospital, Depok, Indonesia.

BackgroundThe limited availability and high cost of human milk fortifier (HMF) 
in developing countries represent significant challenges. Preterm formula (PTF) 
fortification of breast milk is a potential alternative, but its safety and 
impact on growth are still uncertain. This study compares the safety and effect 
of breast milk fortification with PTF versus HMF on growth velocity in very low 
birth weight preterm infants.MethodsA thorough literature search was conducted 
in the PubMed, Scopus, Google Scholar, ClinicalTrials.gov, and Europe PMC 
databases up to March 24, 2024. Relevant randomized controlled trials (RCTs) 
studying fortifying breast milk with PTF or HMF were identified and their 
quality assessed using the Cochrane Risk of Bias 2 tool. Statistical analysis 
was performed in Review Manager Web, and certainty of evidence was assessed with 
GRADE.ResultsSix RCTs were included, of which one exhibited low bias risk, three 
had moderate concerns, and two were classed as high risk. The six RCTs, 
involving 434 preterm infants, revealed no significant differences in the gain 
of weight (SMD 0, 95% CI -0.19 to 0.19), length (MD -0.01 cm/wk, 95% CI -0.06 to 
0.05), or head circumference (MD -0.01 cm/wk, 95% CI -0.06 to 0.04) between PTF 
and HMF. There were comparable risks of morbidities.ConclusionVery low certainty 
evidence suggests that breast milk fortification with PTF may be a safe 
alternative to HMF, with similar safety profiles and effects on growth.

DOI: 10.1177/19345798251350987
PMID: 40534564 [Indexed for MEDLINE]

Conflict of interest statement: Declaration of conflicting interestsThe 
author(s) declared no potential conflicts of interest with respect to the 
research, authorship, and/or publication of this article.


12. Pediatr Res. 2025 Nov;98(5):1696-1710. doi: 10.1038/s41390-025-04115-9. Epub 
2025 Jun 5.

Systematic review and meta-analysis of enteral protein intake effects on growth 
in preterm infants.

Sanchez-Holgado M(1), Johnson MJ(2)(3)(4), Witte Castro A(1), Criado Camargo 
S(1), van den Akker CHP(5)(6), Alvarez-García P(1), Cabrera-Lafuente M(1), 
Jiménez Varas MÁ(7), Saenz de Pipaon M(1).

Author information:
(1)Department of Neonatology. La Paz University Hospital. Hospital La Paz 
Institute for Health Research-IdIPAZ, Madrid, Spain.
(2)Faculty of Medicine, University of Southampton, Southampton, UK. 
.
(3)Department of Neonatal Medicine, University Hospital Southampton NHS 
Foundation Trust, Southampton, UK..
(4)NIHR Biomedical Research Centre Southampton, University Hospital Southampton 
NHS Foundation Trust and University of Southampton, Southampton, UK. 
.
(5)Department of Pediatrics-Neonatology, Emma Children's Hospital, Amsterdam 
UMC, University of Amsterdam, Amsterdam, The Netherlands.
(6)Amsterdam Reproduction & Development Research Institute and Amsterdam 
Gastroenterology, Endocrinology & Metabolism Research Institute, Amsterdam, The 
Netherlands.
(7)Information specialist. La Paz University Hospital, Madrid, Spain.

Comment in
    Pediatr Res. 2025 Nov;98(5):1615-1616. doi: 10.1038/s41390-025-04270-z.

BACKGROUND: The optimal enteral protein intake for adequate growth in preterm 
infants remains unclear. This systematic review evaluates the impact of protein 
intake from fortified human milk on growth in very preterm infants.
METHODS: Randomized clinical trials from January 2005 to August 2024 were 
included. Eligible studies measured true enteral protein intake in preterm 
infants. Searches were conducted in PubMed, Embase, and Cochrane CENTRAL. Risk 
of bias was evaluated using the revised Cochrane Risk of Bias Tool.
RESULTS: Ten randomized clinical trials (n = 646) were included. Meta-regression 
revealed a significant linear relationship between protein intake and weight 
gain (5.73 g/kg/day weight gain for each gram of protein/kg/day, p = 0.001), but 
not with head circumference or length gain. After adjustment for energy intake, 
significant relationships were found between protein intake and both weight gain 
and length growth. In contrast, the forest plot meta-analysis comparing high 
versus low protein intake showed no significant differences in weight or head 
circumference gain. However, infants receiving higher protein intake had greater 
weight at discharge (SMD 0.35, 95% CI 0.12 to 0.57, n = 312, 4 studies, high 
certainty) and more length growth (SMD 0.5, 95% CI 0.08 to 0.92, n = 174, 3 
studies, moderate certainty).
DISCUSSION AND CONCLUSION: Moderate to high-certainty evidence suggests that 
increased enteral protein intake improves growth outcomes in very preterm 
infants.
REGISTRATION: PROSPERO CRD42022287991.
IMPACT: This systematic review is the first to evaluate the impact of enteral 
protein intake on growth in preterm infants <32 weeks, using studies that 
measured actual intake. A positive correlation was found between protein intake 
and weight gain. Meta-regression suggests most premature infants may require 
4.0-4.5 g/kg/day to achieve in utero growth rates, rather than 3.5-4.0 g/kg/day. 
The meta-analysis indicates a positive relationship between protein intake, 
growth in length, and discharge weight. These findings underscore the critical 
role of adequate protein intake in growth outcomes and highlight the need to 
maintain appropriate energy: protein ratios.

© 2025. The Author(s).

DOI: 10.1038/s41390-025-04115-9
PMCID: PMC12602309
PMID: 40473805 [Indexed for MEDLINE]

Conflict of interest statement: Consent statement: This manuscript does not 
contain individual personal data from patients. Patient consent was not 
required. Competing interests: C.H.P. vdA declares no personal financial gain. 
All speaker and consultancy honoraria received from Nestlé Nutrition Institute, 
Nutricia Early Life Nutrition, and Baxter were paid directly to his hospital and 
allocated as research funds. His hospital is also participating in an 
industry-sponsored trial conducted by ELGAN, with reimbursements similarly 
directed to his hospital and designated for research purposes. C.H.P. vdA serves 
as the unpaid Vice-Director of the Dutch National Human Milk Bank and has been 
an unpaid member of the National Breastfeeding Council since 2024. All other 
authors declare no competing interests.


13. Nutrients. 2025 May 12;17(10):1651. doi: 10.3390/nu17101651.

Effect of Multi-Nutrient Milk Fortification on Preterm Neonate Outcomes: A 
Network Meta-Analysis.

Campbell-Yeo M(1)(2), Gullickson C(3), McCulloch H(2), Disher T(4), Hughes B(5).

Author information:
(1)School of Nursing, Dalhousie University, Halifax, NS B3H 4R2, Canada.
(2)Mom-Linc Lab, IWK Health, Halifax, NS B3K 6R8, Canada.
(3)Faculty of Medicine, Dalhousie University, Halifax, NS B3H 4R2, Canada.
(4)Faculty of Computer Science, Dalhousie University, Halifax, NS B3H 4R2, 
Canada.
(5)School of Nursing, University of Prince Edward Island, Charlottetown, PEI C1A 
4P3, Canada.

Background/Objectives: Optimal feeding regimens for preterm neonates, including 
the role of multi-nutrient fortification, are unknown, leading to large practice 
variation in comparing different feeding regimens that include fortification and 
their impact on outcomes for preterm infants. Methods: Using a network 
meta-analyses design, two reviewers independently extracted data. A Cochrane 
CENTRAL, Medline, Embase, and CINAHL search was conducted for all studies 
published up to 27 June 2023. Randomized clinical trials of feeding regimens for 
preterm infants that included multi-nutrient fortification were included. 
Outcomes were mortality, necrotizing enterocolitis (NEC), retinopathy of 
prematurity (ROP), sepsis, periventricular leukomalacia (PVL), bronchopulmonary 
dysplasia (BPD), time to full enteral feeds, and the Bayley II MDI developmental 
score. Results: Fifty-nine studies were included. For mortality, NEC, and time 
to reach full enteral feeds, the top-ranked treatment class was the mother's own 
milk with donor milk and a human-milk-based fortifier. For ROP and BPD, the 
top-ranked treatment class was mother's own milk with a phosphorus fortifier. 
For sepsis, the top-ranked treatment class was mother's own milk with formula. 
For PVL, the top-ranked treatment classes were mother's own milk and mother's 
own milk with donor milk and a bovine fortifier in the two disconnected 
networks. For the Bayley II MDI score, the top-ranked treatment class was 
mother's own milk with formula and bovine fortification. Conclusions: Treatment 
rankings are consistent with the underlying hypothesis that increased mother's 
own milk intake appears to be associated with better clinical outcomes. This 
review provides the first global view of interventions and highlights 
insufficient high-quality evidence to support or refute one fortification 
feeding regimen over another.

DOI: 10.3390/nu17101651
PMCID: PMC12113778
PMID: 40431391 [Indexed for MEDLINE]

Conflict of interest statement: The authors declare no conflicts of interest.


14. Public Health Nurs. 2025 May-Jun;42(3):1302-1314. doi: 10.1111/phn.13521. Epub 
2025 Jan 7.

Unraveling the Relationship Between Vitamin D and Noncommunicable Diseases: A 
Systemic Review and Meta-Analysis.

Dumbre D(1), Upendra S(1), Zacharias BS(1).

Author information:
(1)Symbiosis College of Nursing, Symbiosis International (Deemed University), 
Pune, India.

BACKGROUND: Subclinical vitamin D insufficiency is frequent in both developing 
and developed countries. Even after rickets was eliminated in the 1930s by 
fortifying milk, up to 1 billion people worldwide suffer from subclinical 
vitamin D deficiency (VDD). Numerous noncommunicable diseases (NCDs), including 
depression, autoimmune illnesses, diabetes, osteoporosis, and cardiovascular 
disease, are linked to this deficiency.
METHODOLOGY: The Preferred Reporting Items for Systematic Reviews and 
Meta-Analyses (PRISMA) protocol was applied when conducting a systematic review. 
Relevant studies published between 2010 and 2023 were found by searching 
databases such as Web of Science, Scopus, and PubMed. To find correlations 
between deficiency of vitamin D and NCDs, data was extracted from the studies 
and analyzed using the Mixed Method Appraisal Tools (MMAT). In meta-analysis 
data combined from multiple observational Unraveling the Relationship Between 
Vitamin D and Noncommunicable Diseases-A Systemic Review and Meta-Analysis 
studies to evaluate the relationship between vitamin D insufficiency and health 
outcomes. Statistical techniques, such as calculating effect sizes and 
constructing confidence ranges, are employed to determine combined outcomes and 
investigate possible causal connections.
RESULTS: Fourteen studies-including cross-sectional studies, prospective 
cohorts, randomized controlled trials, and longitudinal studies-met the 
inclusion criteria. Significant correlations among deficiency of vitamin D and a 
number of NCDs, including anemia, diabetes, hypertension, rheumatoid arthritis 
(RA), mental health issues, and autoimmune hypothyroidism, were found in the 
review. The results highlight the prevalence of deficiency of vitamin D 
worldwide and its detrimental impact on health. In order to lower the risk of 
numerous NCDs, this review emphasizes the need for greater awareness and 
possible interventions to maintain appropriate vitamin D levels. A meta-analysis 
was conducted to investigate the potential causal connections between vitamin D 
insufficiency and different health disorders. It elucidates the connections 
between deficient levels of vitamin D and conditions such as mental health 
disorders, anemia, diabetes, hypertension, and autoimmune diseases. The study 
prioritizes the examination of temporal correlations, biological plausibility, 
and the removal of confounding factors to provide support for these 
associations. While the text acknowledges that experimental data, such as 
randomized controlled trials, would enhance the argument for causation, it 
mostly depends on observational studies.

© 2025 Wiley Periodicals LLC.

DOI: 10.1111/phn.13521
PMID: 39777920 [Indexed for MEDLINE]


15. Cureus. 2024 Nov 10;16(11):e73380. doi: 10.7759/cureus.73380. eCollection 2024 
Nov.

Strategies for the Fortification of Human Milk in Preterm Infants: A Systematic 
Review.

Contreras Chova F(1), Villanueva-García A(1), González-Boyero JL(1), 
Campos-Martínez AM(1), Blanca-Jover E(1), Jerez-Calero AE(1), Uberos-Fernández 
J(1).

Author information:
(1)Pediatrics and Neonatology, Hospital Universitario San Cecilio, Granada, ESP.

Breast milk is the best source of nourishment for both full-term and preterm 
newborns. However, in preterm newborns, exclusive breastfeeding can lead to 
nutritional deficiencies, with short- and long-term consequences on growth and 
neurocognitive development. Breast milk fortification is a widely used strategy 
to provide an adequate nutritional profile to these patients. There is currently 
a great heterogeneity in terms of fortification patterns and the type of 
fortifier. The present systematic review summarizes the current scientific 
evidence to evaluate the different strategies for fortification of human milk in 
preterm infants, their possible differences on benefits, and the hypothetical 
impact on prematurity outcomes.

Copyright © 2024, Contreras Chova et al.

DOI: 10.7759/cureus.73380
PMCID: PMC11630059
PMID: 39659332

Conflict of interest statement: Conflicts of interest: In compliance with the 
ICMJE uniform disclosure form, all authors declare the following: 
Payment/services info: All authors have declared that no financial support was 
received from any organization for the submitted work. Financial relationships: 
All authors have declared that they have no financial relationships at present 
or within the previous three years with any organizations that might have an 
interest in the submitted work. Other relationships: All authors have declared 
that there are no other relationships or activities that could appear to have 
influenced the submitted work.


16. Neonatology. 2025;122(Suppl 1):209-223. doi: 10.1159/000542154. Epub 2024 Nov 
26.

Nutritional Management of Low Birth Weight and Preterm Infants in Low- and Low 
Middle-Income Countries.

Azhar M(1), Yasin R(1), Hanif S(1), Bughio SA(1), Das JK(1)(2), Bhutta ZA(1)(3).

Author information:
(1)Institute for Global Health and Development, The Aga Khan University, 
Karachi, Pakistan.
(2)Division of Women and Child Health, The Aga Khan University, Karachi, 
Pakistan.
(3)Center for Global Child Health, Hospital for Sick Children, Toronto, Ontario, 
Canada.

INTRODUCTION: Preterm and low birth weight (LBW) infants are at an increased 
risk of morbidity and mortality compared with their term counterparts, with more 
than 20 million LBW infants born each year, the majority in lower middle-income 
countries (LMICs). Given the increased vulnerability and higher nutritional 
needs of these infants, optimizing feeding strategies may play a crucial role in 
improving their health outcomes.
METHODS: We updated evidence of Every Newborn Series published in The Lancet 
2014 by identifying relevant systematic reviews, extracting low-income country 
(LIC) and LMIC data, and conducting revised meta-analysis for these contexts.
RESULTS: We found 15 reviews; the evidence showed that early initiation of 
enteral feeding reduced neonatal mortality overall, but not in LIC/LMIC 
settings. Breastfeeding promotion interventions increased the prevalence of 
early initiation of breastfeeding and exclusive breastfeeding at 3 and 6 months 
of age in LMIC settings. There was an increased risk of neonatal mortality with 
formula milk in LIC/LMIC settings. Despite contributing to greater weight gain, 
there was a higher risk of necrotizing enterocolitis with formula milk overall. 
Breast milk fortification and nutrient-enriched formula improved growth 
outcomes. Iron and vitamin A supplementation reduced anemia and mortality rates 
(LMIC), respectively. The evidence also suggested that benefits of various 
different micronutrient supplementation interventions such as zinc, 
calcium/phosphorous, and vitamin D, outweigh the risks since our review 
demonstrates little to no adverse effects deriving from their supplementation, 
particularly for a breastfed preterm and/or LBW infant.
CONCLUSION: Early adequate nutritional support of preterm or LBW infant is 
paramount to averse adverse health outcomes, contribute to normal growth, 
resistance to infection, and optimal development. Breast milk feeding and 
micronutrient supplementation are crucial to reduce diarrhea incidence and 
mortality respectively while feed fortification or nutrient-enriched formula, 
when breast milk is not available, to enhance better growth especially in LMICs 
where there is higher population of growth restriction and stunting. This review 
also highlights need for randomized trials in LMICs at large scale to further 
strengthen the evidence.
INTRODUCTION: Preterm and low birth weight (LBW) infants are at an increased 
risk of morbidity and mortality compared with their term counterparts, with more 
than 20 million LBW infants born each year, the majority in lower middle-income 
countries (LMICs). Given the increased vulnerability and higher nutritional 
needs of these infants, optimizing feeding strategies may play a crucial role in 
improving their health outcomes.
METHODS: We updated evidence of Every Newborn Series published in The Lancet 
2014 by identifying relevant systematic reviews, extracting low-income country 
(LIC) and LMIC data, and conducting revised meta-analysis for these contexts.
RESULTS: We found 15 reviews; the evidence showed that early initiation of 
enteral feeding reduced neonatal mortality overall, but not in LIC/LMIC 
settings. Breastfeeding promotion interventions increased the prevalence of 
early initiation of breastfeeding and exclusive breastfeeding at 3 and 6 months 
of age in LMIC settings. There was an increased risk of neonatal mortality with 
formula milk in LIC/LMIC settings. Despite contributing to greater weight gain, 
there was a higher risk of necrotizing enterocolitis with formula milk overall. 
Breast milk fortification and nutrient-enriched formula improved growth 
outcomes. Iron and vitamin A supplementation reduced anemia and mortality rates 
(LMIC), respectively. The evidence also suggested that benefits of various 
different micronutrient supplementation interventions such as zinc, 
calcium/phosphorous, and vitamin D, outweigh the risks since our review 
demonstrates little to no adverse effects deriving from their supplementation, 
particularly for a breastfed preterm and/or LBW infant.
CONCLUSION: Early adequate nutritional support of preterm or LBW infant is 
paramount to averse adverse health outcomes, contribute to normal growth, 
resistance to infection, and optimal development. Breast milk feeding and 
micronutrient supplementation are crucial to reduce diarrhea incidence and 
mortality respectively while feed fortification or nutrient-enriched formula, 
when breast milk is not available, to enhance better growth especially in LMICs 
where there is higher population of growth restriction and stunting. This review 
also highlights need for randomized trials in LMICs at large scale to further 
strengthen the evidence.

© 2024 The Author(s). Published by S. Karger AG, Basel.

DOI: 10.1159/000542154
PMCID: PMC11875417
PMID: 39591949 [Indexed for MEDLINE]

Conflict of interest statement: The authors declare no conflict of interest.


17. J Nutr. 2024 Nov;154(11):3446-3456. doi: 10.1016/j.tjnut.2024.09.010. Epub 2024 
Sep 26.

A Systematic Review on the Impact of Plant-Based Milk Consumption on Growth and 
Nutrition in Children and Adolescents.

Soczynska I(1), da Costa BR(2), O'Connor DL(3), Jenkins DJ(4), Birken CS(5), 
Keown-Stoneman CD(6), D'Hollander C(1), Calleja S(7), Maguire JL(8).

Author information:
(1)Department of Nutritional Sciences, Faculty of Medicine, University of 
Toronto, Toronto, Canada; Department of Pediatrics, St. Michael's Hospital, 
Toronto, Canada; Joannah and Brian Lawson Centre for Child Nutrition, University 
of Toronto, Toronto, Canada.
(2)Institute of Health Policy, Management and Evaluation, University of Toronto, 
Toronto, Canada; Clinical Trial Service Unit and Epidemiological Studies Unit, 
Nuffield Department of Population Health, University of Oxford, Oxford, United 
Kingdom.
(3)Department of Nutritional Sciences, Faculty of Medicine, University of 
Toronto, Toronto, Canada; Joannah and Brian Lawson Centre for Child Nutrition, 
University of Toronto, Toronto, Canada; Translational Medicine, SickKids 
Research Institute, Toronto, Canada.
(4)Department of Nutritional Sciences, Faculty of Medicine, University of 
Toronto, Toronto, Canada; Li Ka Shing Knowledge Institute, St. Michael's 
Hospital, Toronto, Canada; Department of Medicine, Temerty Faculty of Medicine, 
University of Toronto, Toronto, Canada; Clinical Nutrition and Risk Factor 
Modification Centre, St. Michael's Hospital, Toronto, Canada; Division of 
Endocrinology and Metabolism, St. Michael's Hospital, Toronto, Canada.
(5)Joannah and Brian Lawson Centre for Child Nutrition, University of Toronto, 
Toronto, Canada; Department of Paediatrics, Temerty Faculty of Medicine, 
University of Toronto, Toronto, Ontario; Child Health and Evaluative Sciences, 
SickKids Research Institute, Toronto, Canada; Division of Paediatric Medicine, 
Hospital for Sick Children, Toronto, Canada.
(6)Li Ka Shing Knowledge Institute, St. Michael's Hospital, Toronto, Canada; 
Applied Health Research Centre, St. Michael's Hospital, Toronto, Canada; 
Department of Biostatistics, Dalla Lana School of Public Health, University of 
Toronto, Toronto, Canada.
(7)Schulich Library of Physical Sciences, Life Sciences and Engineering, McGill 
University, Montreal, Canada.
(8)Department of Nutritional Sciences, Faculty of Medicine, University of 
Toronto, Toronto, Canada; Department of Pediatrics, St. Michael's Hospital, 
Toronto, Canada; Joannah and Brian Lawson Centre for Child Nutrition, University 
of Toronto, Toronto, Canada; Institute of Health Policy, Management and 
Evaluation, University of Toronto, Toronto, Canada; Li Ka Shing Knowledge 
Institute, St. Michael's Hospital, Toronto, Canada; Department of Paediatrics, 
Temerty Faculty of Medicine, University of Toronto, Toronto, Ontario. Electronic 
address:.

BACKGROUND: Children are increasingly consuming plant-based milks, yet the 
impact on their growth and nutrition is unclear.
OBJECTIVE: This systematic review aimed to summarize the available evidence on 
the impact of plant-based milk consumption on growth and nutrition in children 
and adolescents.
METHODS: MEDLINE, Embase (Excerpta Medica Database), EBM Reviews - Cochrane 
Central Register of Controlled Trials and Cochrane Database of Systematic 
Reviews, Cumulative Index to Nursing and Allied Health Literature, Child 
Development and Adolescent Studies, and Scopus were comprehensively searched 
from 2000 to 2024 to identify studies evaluating the growth and nutritional 
effects of plant-based milk consumption in children aged 1-18 y. Two reviewers 
independently screened full-text articles, assessed their quality, and extracted 
data.
RESULTS: A total of 6 studies were identified: 3 cross-sectional studies, 1 
prospective cohort study, and 2 clinical trials (total n = 15,815). 
Observational studies found that consumption of plant-based milk was associated 
with lower childhood body mass index (BMI), height, and serum vitamin D 
concentrations compared with cow milk. No association was found between soy milk 
consumption and BMI in adolescent girls. Low-quality clinical trials showed 
minimal effects on growth, and 1 study found that adolescent girls with low 
calcium intake who consumed fortified soy milk had higher bone density compared 
with those who did not consume soy milk.
CONCLUSIONS: Available evidence suggests that children who consume plant-based 
milk may have lower BMI, height, and micronutrient intake compared with those 
who consume cow milk, whereas fortified soy milk may support bone health in 
adolescents who do not drink cow milk. Longitudinal studies and randomized 
controlled trials are needed to determine whether these associations persist 
over time, differ between children and adolescents or among those who consume 
soy milk, and to understand the potential underlying mechanisms. This trial was 
registered at PROSPERO as CRD42022367269.

Copyright © 2024 American Society for Nutrition. Published by Elsevier Inc. All 
rights reserved.

DOI: 10.1016/j.tjnut.2024.09.010
PMID: 39332772 [Indexed for MEDLINE]

Conflict of interest statement: Conflict of interest JLM received financial 
support from Canadian Institutes of Health Research; received an unrestricted 
research grant for a completed investigator-initiated study from the Dairy 
Farmers of Canada (2011-2012) and D-drops provided non-financial support 
(vitamin D supplements) for initiated study on vitamin D and respiratory tract 
infections (2011–2015); received research grants from Saskatchewan & Alberta 
Pulse Growers Associations, the Agricultural Bioproducts Innovation Program 
through the Pulse Research Network, the Advanced Foods and Material Network, 
Loblaw Companies Ltd., Unilever Canada and the Netherlands, Barilla, the Almond 
Board of California, Agriculture and Agri-food Canada, Pulse Canada, Kellogg’s 
Company, Canada, Quaker Oats, Canada, Procter & Gamble Technical Centre Ltd., 
Bayer Consumer Care, Springfield, NJ, Pepsi/Quaker, International Nut & Dried 
Fruit Council (INC), Soy Foods Association of North America, the Coca-Cola 
Company (investigator initiated, unrestricted grant), Solae, Haine Celestial, 
the Sanitarium Company, Orafti, the International Tree Nut Council Nutrition 
Research and Education Foundation, the Peanut Institute, Soy Nutrition Institute 
(SNI), the Canola and Flax Councils of Canada, the Calorie Control Council, the 
Canadian Institutes of Health Research (CIHR), the Canada Foundation for 
Innovation (CFI), and the Ontario Research Fund (ORF); received in-kind supplies 
for trials as a research support from the Almond board of California, Walnut 
Council of California, the Peanut Institute, Barilla, Unilever, Unico, Primo, 
Loblaw Companies, Quaker (Pepsico), Pristine Gourmet, Bunge Limited, Kellogg 
Canada, and WhiteWave Foods; has been on the speaker’s panel, served on the 
scientific advisory board and/or received travel support and/or honoraria from 
Nutritional Fundamentals for Health (NFH)-Nutramedica, Saint Barnabas Medical 
Center, The University of Chicago, 2020 China Glycemic Index (GI) International 
Conference, Atlantic Pain Conference, Academy of Life Long Learning, the Almond 
Board of California, Canadian Agriculture Policy Institute, Loblaw Companies 
Ltd, the Griffin Hospital (for the development of the NuVal scoring system), the 
Coca-Cola Company, Epicure, Danone, Diet Quality Photo Navigation (DQPN), Better 
Therapeutics (FareWell), Verywell, True Health Initiative (THI), Heali AI Corp, 
Institute of Food Technologists (IFT), Soy Nutrition Institute (SNI), Herbalife 
Nutrition Institute (HNI), Saskatchewan & Alberta Pulse Growers Associations, 
Sanitarium Company, Orafti, the International Tree Nut Council Nutrition 
Research and Education Foundation, the Peanut Institute, Herbalife 
International, Pacific Health Laboratories, Barilla, Metagenics, Bayer Consumer 
Care, Unilever Canada and the Netherlands, Solae, Kellogg, Quaker Oats, Procter 
& Gamble, Abbott Laboratories, Dean Foods, the California Strawberry Commission, 
Haine Celestial, PepsiCo, the Alpro Foundation, Pioneer Hi-Bred International, 
DuPont Nutrition and Health, Spherix Consulting and WhiteWave Foods, the 
Advanced Foods and Material Network, the Canola and Flax Councils of Canada, 
Agri-Culture and Agri-Food Canada, the Canadian Agri-Food Policy Institute, 
Pulse Canada, the Soy Foods Association of North America, the Nutrition 
Foundation of Italy (NFI), Nutra-Source Diagnostics, the McDougall Program, the 
Toronto Knowledge Translation Group (St. Michael’s Hospital), the Canadian 
College of Naturopathic Medicine, The Hospital for Sick Children, the Canadian 
Nutrition Society (CNS), the American Society of Nutrition (ASN), Arizona State 
University, Paolo Sorbini Foundation and the Institute of Nutrition, Metabolism 
and Diabetes; received an honorarium from the United States Department of 
Agriculture to present the 2013 W.O. Atwater Memorial Lecture. He received the 
2013 Award for Excellence in Research from the International Nut and Dried Fruit 
Council; received funding and travel support from the Canadian Society of 
Endocrinology and Metabolism to produce mini cases for the Canadian Diabetes 
Association (CDA); member of the International Carbohydrate Quality Consortium 
(ICQC); and His wife, Alexandra L Jenkins, is a director and partner of INQUIS 
Clinical Research for the Food Industry, his 2 daughters, Wendy Jenkins and Amy 
Jenkins, have published a vegetarian book that promotes the use of the foods 
described here, The Portfolio Diet for Cardiovascular Risk Reduction (Academic 
Press/Elsevier 2020 ISBN:978-0-12-810510-8) and his sister, Caroline Brydson, 
received funding through a grant from the St. Michael’s Hospital Foundation to 
develop a cookbook for one of his studies. All authors report no conflicts of 
interest.


18. Cochrane Database Syst Rev. 2024 Sep 6;9(9):CD002971. doi: 
10.1002/14651858.CD002971.pub6.

Donor human milk for preventing necrotising enterocolitis in very preterm or 
very low-birthweight infants.

Quigley M(1), Embleton ND(2), Meader N(3), McGuire W(4).

Author information:
(1)National Perinatal Epidemiology Unit, University of Oxford, Oxford, UK.
(2)Newcastle Neonatal Service , Newcastle Hospitals NHS Foundation Trust and 
Newcastle University, Newcastle upon Tyne, UK.
(3)University of Newcastle, Newcastle upon Tyne, UK.
(4)Centre for Reviews and Dissemination , University of York, York, UK.

Update of
    Cochrane Database Syst Rev. 2019 Jul 19;7:CD002971. doi: 
10.1002/14651858.CD002971.pub5.

BACKGROUND: When sufficient maternal milk is not available, donor human milk or 
formula are the alternative forms of enteral nutrition for very preterm or very 
low-birthweight (VLBW) infants. Donor human milk may retain the non-nutritive 
benefits of maternal milk and has been proposed as a strategy to reduce the risk 
of necrotising enterocolitis (NEC) and associated mortality and morbidity in 
very preterm or VLBW infants.
OBJECTIVES: To assess the effectiveness of donor human milk compared with 
formula for preventing NEC and associated morbidity and mortality in very 
preterm or VLBW infants when sufficient maternal milk is not available.
SEARCH METHODS: We searched the Cochrane Central Register of Controlled Trials 
(CENTRAL), MEDLINE, Embase, the Maternity and Infant Care (MIC) database, and 
the Cumulative Index to Nursing and Allied Health Literature (CINAHL), from the 
earliest records to February 2024. We searched clinical trials registries and 
examined the reference lists of included studies.
SELECTION CRITERIA: Randomised or quasi-randomised controlled trials comparing 
feeding with donor human milk versus formula in very preterm (< 32 weeks' 
gestation) or VLBW (< 1500 g) infants.
DATA COLLECTION AND ANALYSIS: Two review authors evaluated the risk of bias in 
the trials, extracted data, and synthesised effect estimates using risk ratio, 
risk difference, and mean difference, with associated 95% confidence intervals. 
The primary outcomes were NEC, late-onset invasive infection, and all-cause 
mortality before hospital discharge. The secondary outcomes were growth 
parameters and neurodevelopment. We used the GRADE approach to assess the 
certainty of the evidence for our primary outcomes.
MAIN RESULTS: Twelve trials with a total of 2296 infants fulfilled the inclusion 
criteria. Most trials were small (average sample size was 191 infants). All 
trials were performed in neonatal units in Europe or North America. Five trials 
were conducted more than 40 years ago; the remaining seven trials were conducted 
in the year 2000 or later. Some trials had methodological weaknesses, including 
concerns regarding masking of investigators and selective reporting. 
Meta-analysis showed that donor human milk reduces the risk of NEC (risk ratio 
(RR) 0.53, 95% confidence interval (CI) 0.37 to 0.76; I² = 4%; risk difference 
(RD) -0.03, 95% CI -0.05 to -0.01; 11 trials, 2261 infants; high certainty 
evidence). Donor human milk probably has little or no effect on late-onset 
invasive infection (RR 1.12, 0.95 to 1.31; I² = 27%; RD 0.03, 95% CI -0.01 to 
-0.07; 7 trials, 1611 infants; moderate certainty evidence) or all-cause 
mortality (RR 1.00, 95% CI 0.76 to 1.31; I² = 0%; RD -0.00, 95% CI -0.02 to 
0.02; 9 trials, 2116 infants; moderate certainty evidence).
AUTHORS' CONCLUSIONS: The evidence shows that donor human milk reduces the risk 
of NEC by about half in very preterm or VLBW infants. There is probably little 
or no effect on late-onset invasive infection or all-cause mortality before 
hospital discharge.

Copyright © 2024 The Cochrane Collaboration. Published by John Wiley & Sons, 
Ltd.

DOI: 10.1002/14651858.CD002971.pub6
PMCID: PMC11378496
PMID: 39239939 [Indexed for MEDLINE]

Conflict of interest statement: MQ was a deputy Co‐ordinating Editor for 
Cochrane Common Mental Disorders; she did not participate in the acceptance or 
editorial processes for this review. NDE declares research grants paid to his 
employer for a trial using commercially prepared, human milk derived products 
from Prolacta Bioscience, 2017; a grant from Danone Early Life Nutrition for a 
study on feeding in late and moderately preterm infants, 2018; and a grant for a 
trial of human milk‐derived fortifier from NeoKare, 2022. NDE declares lecture 
honoraria from Nestle Nutrition Institute donated to charity in 2021 and 2023. 
NDE declares no financial relationships or benefits. NDE is a member of the UK 
Association for Milk Banking. NM has nothing to declare. WM is a Co‐ordinating 
Editor for Cochrane Neonatal; he did not participate in the acceptance or 
editorial processes for this review.


19. BMC Med. 2024 Aug 22;22(1):336. doi: 10.1186/s12916-024-03524-7.

A systematic review and meta-analysis of randomized trials of substituting 
soymilk for cow's milk and intermediate cardiometabolic outcomes: understanding 
the impact of dairy alternatives in the transition to plant-based diets on 
cardiometabolic health.

Erlich MN(1)(2), Ghidanac D(1)(2), Blanco Mejia S(1)(2), Khan TA(1)(2), 
Chiavaroli L(1)(2)(3), Zurbau A(1)(2), Ayoub-Charette S(1)(2), Almneni A(4), 
Messina M(5), Leiter LA(1)(2)(3)(6)(7), Bazinet RP(1), Jenkins 
DJA(1)(2)(3)(6)(7), Kendall CWC(1)(2)(8), Sievenpiper JL(9)(10)(11)(12)(13).

Author information:
(1)Department of Nutritional Sciences, Temerty Faculty of Medicine, University 
of Toronto, Toronto, ON, Canada.
(2)Toronto 3D Knowledge Synthesis and Clinical Trials Unit, Clinical Nutrition 
and Risk Factor Modification Centre, St. Michael's Hospital, Toronto, ON, 
Canada.
(3)Li Ka Shing Knowledge Institute, St. Michael's Hospital, Toronto, ON, Canada.
(4)Royal College of Surgeons in Ireland, Dublin, Ireland.
(5)Soy Nutrition Institute Global, Washington, DC, USA.
(6)Department of Medicine, Temerty Faculty of Medicine, University of Toronto, 
Toronto, ON, Canada.
(7)Division of Endocrinology and Metabolism, Department of Medicine, St. 
Michael's Hospital, Toronto, ON, Canada.
(8)College of Pharmacy and Nutrition, University of Saskatchewan, Saskatoon, SK, 
Canada.
(9)Department of Nutritional Sciences, Temerty Faculty of Medicine, University 
of Toronto, Toronto, ON, Canada..
(10)Toronto 3D Knowledge Synthesis and Clinical Trials Unit, Clinical Nutrition 
and Risk Factor Modification Centre, St. Michael's Hospital, Toronto, ON, 
Canada..
(11)Li Ka Shing Knowledge Institute, St. Michael's Hospital, Toronto, ON, 
Canada..
(12)Department of Medicine, Temerty Faculty of Medicine, University of Toronto, 
Toronto, ON, Canada..
(13)Division of Endocrinology and Metabolism, Department of Medicine, St. 
Michael's Hospital, Toronto, ON, Canada..

BACKGROUND: Dietary guidelines recommend a shift to plant-based diets. Fortified 
soymilk, a prototypical plant protein food used in the transition to plant-based 
diets, usually contains added sugars to match the sweetness of cow's milk and is 
classified as an ultra-processed food. Whether soymilk can replace minimally 
processed cow's milk without the adverse cardiometabolic effects attributed to 
added sugars and ultra-processed foods remains unclear. We conducted a 
systematic review and meta-analysis of randomized controlled trials, to assess 
the effect of substituting soymilk for cow's milk and its modification by added 
sugars (sweetened versus unsweetened) on intermediate cardiometabolic outcomes.
METHODS: MEDLINE, Embase, and the Cochrane Central Register of Controlled Trials 
were searched (through June 2024) for randomized controlled trials of ≥ 3 weeks 
in adults. Outcomes included established markers of blood lipids, glycemic 
control, blood pressure, inflammation, adiposity, renal disease, uric acid, and 
non-alcoholic fatty liver disease. Two independent reviewers extracted data and 
assessed risk of bias. The certainty of evidence was assessed using GRADE 
(Grading of Recommendations, Assessment, Development, and Evaluation). A 
sub-study of lactose versus sucrose outside of a dairy-like matrix was conducted 
to explore the role of sweetened soymilk which followed the same methodology.
RESULTS: Eligibility criteria were met by 17 trials (n = 504 adults with a range 
of health statuses), assessing the effect of a median daily dose of 500 mL of 
soymilk (22 g soy protein and 17.2 g or 6.9 g/250 mL added sugars) in 
substitution for 500 mL of cow's milk (24 g milk protein and 24 g or 12 g/250 mL 
total sugars as lactose) on 19 intermediate outcomes. The substitution of 
soymilk for cow's milk resulted in moderate reductions in non-HDL-C (mean 
difference, - 0.26 mmol/L [95% confidence interval, - 0.43 to - 0.10]), systolic 
blood pressure (- 8.00 mmHg [- 14.89 to - 1.11]), and diastolic blood pressure 
(- 4.74 mmHg [- 9.17 to - 0.31]); small important reductions in LDL-C 
(- 0.19 mmol/L [- 0.29 to - 0.09]) and c-reactive protein (CRP) (- 0.82 mg/L 
[- 1.26 to - 0.37]); and trivial increases in HDL-C (0.05 mmol/L [0.00 to 
0.09]). No other outcomes showed differences. There was no meaningful effect 
modification by added sugars across outcomes. The certainty of evidence was high 
for LDL-C and non-HDL-C; moderate for systolic blood pressure, diastolic blood 
pressure, CRP, and HDL-C; and generally moderate-to-low for all other outcomes. 
We could not conduct the sub-study of the effect of lactose versus added sugars, 
as no eligible trials could be identified.
CONCLUSIONS: Current evidence provides a good indication that replacing cow's 
milk with soymilk (including sweetened soymilk) does not adversely affect 
established cardiometabolic risk factors and may result in advantages for blood 
lipids, blood pressure, and inflammation in adults with a mix of health 
statuses. The classification of plant-based dairy alternatives such as soymilk 
as ultra-processed may be misleading as it relates to their cardiometabolic 
effects and may need to be reconsidered in the transition to plant-based diets.
TRIAL REGISTRATION: ClinicalTrials.gov identifier, NCT05637866.

© 2024. The Author(s).

DOI: 10.1186/s12916-024-03524-7
PMCID: PMC11340166
PMID: 39169353 [Indexed for MEDLINE]

Conflict of interest statement: TAK reports receiving grants from Institute for 
the Advancement of Food and Nutrition Sciences (IAFNS, formerly ILSI North 
America) and National Honey Board (USDA Checkoff program). He has received 
honorariums from Advancement of Food and Nutrition Sciences (IAFNS), the 
International Food Information Council (IFIC), the Calorie Control Council 
(CCC), the International Sweeteners Association (ISA), and AmCham Dubai. He has 
received funding from the Toronto 3D Knowledge Synthesis and Clinical Trials 
foundation. LC has received research support from the Canadian Institutes of 
health Research (CIHR), Protein Industries Canada (a Government of Canada Global 
Innovation Clusters), The United Soybean Board (USDA soy “Checkoff” program), 
and the Alberta Pulse Growers Association. AZ is a part-time research associate 
at INQUIS Clinical Research, Ltd., a contract research organization. She has 
received consulting fees from Glycemic Index Foundation Inc. SA-C has received 
an honorarium from the International Food Information Council (IFIC) for a talk 
on artificial sweeteners, the gut microbiome, and the risk for diabetes. MM was 
employed by the Soy Nutrition Institute Global, an organization that receives 
funding from the United Soybean Board (USB) and from members involved in the soy 
industry. RPB has received industrial grants, including those matched by the 
Canadian government, and/or travel support or consulting fees largely related to 
work on brain fatty acid metabolism or nutrition from Arctic Nutrition, Bunge 
Ltd., Dairy Farmers of Canada, DSM, Fonterra Inc, Mead Johnson, Natures Crops 
International, Nestec Inc. Pharmavite, Sancero Inc., and Spore Wellness Inc. 
Moreover, Dr. Bazinet is on the executive of the International Society for the 
Study of Fatty Acids and Lipids and held a meeting on behalf of Fatty Acids and 
Cell Signaling, both of which rely on corporate sponsorship. Dr. Bazinet has 
given expert testimony in relation to supplements and the brain. DJAJ has 
received research grants from Saskatchewan & Alberta Pulse Growers Associations, 
the Agricultural Bioproducts Innovation Program through the Pulse Research 
Network, the Advanced Foods and Material Network, Loblaw Companies Ltd., 
Unilever Canada and Netherlands, Barilla, the Almond Board of California, 
Agriculture and Agri-food Canada, Pulse Canada, Kellogg’s Company, Canada, 
Quaker Oats, Canada, Procter & Gamble Technical Centre Ltd., Bayer Consumer 
Care, Springfield, NJ, Pepsi/Quaker, International Nut & Dried Fruit Council 
(INC), Soy Foods Association of North America, the Coca-Cola Company 
(investigator initiated, unrestricted grant), Solae, Haine Celestial, the 
Sanitarium Company, Orafti, the International Tree Nut Council Nutrition 
Research and Education Foundation, the Peanut Institute, Soy Nutrition Institute 
(SNI), the Canola and Flax Councils of Canada, the Calorie Control Council, the 
Canadian Institutes of Health Research (CIHR), the Canada Foundation for 
Innovation (CFI), and the Ontario Research Fund (ORF). He has received in-kind 
supplies for trials as a research support from the Almond board of California, 
Walnut Council of California, the Peanut Institute, Barilla, Unilever, Unico, 
Primo, Loblaw Companies, Quaker (Pepsico), Pristine Gourmet, Bunge Limited, 
Kellogg Canada, and WhiteWave Foods. He has been on the speaker’s panel, served 
on the scientific advisory board and/or received travel support and/or honoraria 
from Lawson Centre Nutrition Digital Series, Nutritional Fundamentals for Health 
(NFH)-Nutramedica, Saint Barnabas Medical Center, The University of Chicago, 
2020 China Glycemic Index (GI) International Conference, Atlantic Pain 
Conference, Academy of Life Long Learning, the Almond Board of California, 
Canadian Agriculture Policy Institute, Loblaw Companies Ltd, the Griffin 
Hospital (for the development of the NuVal scoring system), the Coca-Cola 
Company, Epicure, Danone, Diet Quality Photo Navigation (DQPN), Better 
Therapeutics (FareWell), Verywell, True Health Initiative (THI), Heali AI Corp, 
Institute of Food Technologists (IFT), Soy Nutrition Institute (SNI), Herbalife 
Nutrition Institute (HNI), Saskatchewan & Alberta Pulse Growers Associations, 
Sanitarium Company, Orafti, the International Tree Nut Council Nutrition 
Research and Education Foundation, the Peanut Institute, Herbalife 
International, Pacific Health Laboratories, Barilla, Metagenics, Bayer Consumer 
Care, Unilever Canada and Netherlands, Solae, Kellogg, Quaker Oats, Procter & 
Gamble, Abbott Laboratories, Dean Foods, the California Strawberry Commission, 
Haine Celestial, PepsiCo, the Alpro Foundation, Pioneer Hi-Bred International, 
DuPont Nutrition and Health, Spherix Consulting and WhiteWave Foods, the 
Advanced Foods and Material Network, the Canola and Flax Councils of Canada, 
Agri-Culture and Agri-Food Canada, the Canadian Agri-Food Policy Institute, 
Pulse Canada, the Soy Foods Association of North America, the Nutrition 
Foundation of Italy (NFI), Nutra-Source Diagnostics, the McDougall Program, the 
Toronto Knowledge Translation Group (St. Michael’s Hospital), the Canadian 
College of Naturopathic Medicine, The Hospital for Sick Children, the Canadian 
Nutrition Society (CNS), the American Society of Nutrition (ASN), Arizona State 
University, Paolo Sorbini Foundation, and the Institute of Nutrition, Metabolism 
and Diabetes. He received an honorarium from the United States Department of 
Agriculture to present the 2013 W.O. Atwater Memorial Lecture. He received the 
2013 Award for Excellence in Research from the International Nut and Dried Fruit 
Council. He received funding and travel support from the Canadian Society of 
Endocrinology and Metabolism to produce mini cases for the Canadian Diabetes 
Association (CDA). He is a member of the International Carbohydrate Quality 
Consortium (ICQC). His wife, Alexandra L Jenkins, is a director and partner of 
INQUIS Clinical Research for the Food Industry, his 2 daughters, Wendy Jenkins 
and Amy Jenkins, have published a vegetarian book that promotes the use of the 
foods described here, The Portfolio Diet for Cardiovascular Risk Reduction 
(Academic Press/Elsevier 2020 ISBN:978–0-12–810510-8), and his sister, Caroline 
Brydson, received funding through a grant from the St. Michael’s Hospital 
Foundation to develop a cookbook for one of his studies. He is also a vegan. 
CWCK has received grants or research support from the Advanced Food Materials 
Network, Agriculture and Agri-Foods Canada (AAFC), Almond Board of California, 
Barilla, Canadian Institutes of Health Research (CIHR), Canola Council of 
Canada, International Nut and Dried Fruit Council, International Tree Nut 
Council Research and Education Foundation, Loblaw Brands Ltd, the Peanut 
Institute, Pulse Canada, and Unilever. He has received in-kind research support 
from the Almond Board of California, Barilla, California Walnut Commission, 
Kellogg Canada, Loblaw Companies, Nutrartis, Quaker (PepsiCo), the Peanut 
Institute, Primo, Unico, Unilever, and WhiteWave Foods/Danone. He has received 
travel support and/or honoraria from the Barilla, California Walnut Commission, 
Canola Council of Canada, General Mills, International Nut and Dried Fruit 
Council, International Pasta Organization, Lantmannen, Loblaw Brands Ltd, 
Nutrition Foundation of Italy, Oldways Preservation Trust, Paramount Farms, the 
Peanut Institute, Pulse Canada, Sun-Maid, Tate & Lyle, Unilever, and White Wave 
Foods/Danone. He has served on the scientific advisory board for the 
International Tree Nut Council, International Pasta Organization, McCormick 
Science Institute, and Oldways Preservation Trust. He is a founding member of 
the International Carbohydrate Quality Consortium (ICQC), Executive Board Member 
of the Diabetes and Nutrition Study Group (DNSG) of the European Association for 
the Study of Diabetes (EASD), is on the Clinical Practice Guidelines Expert 
Committee for Nutrition Therapy of the EASD, and is a Director of the Toronto 3D 
Knowledge Synthesis and Clinical Trials foundation. JLS has received research 
support from the Canadian Foundation for Innovation, Ontario Research Fund, 
Province of Ontario Ministry of Research and Innovation and Science, Canadian 
Institutes of health Research (CIHR), Diabetes Canada, American Society for 
Nutrition (ASN), National Honey Board (U.S. Department of Agriculture [USDA] 
honey “Checkoff” program), Institute for the Advancement of Food and Nutrition 
Sciences (IAFNS), Pulse Canada, Quaker Oats Center of Excellence, INC 
International Nut and Dried Fruit Council Foundation, The United Soybean Board 
(USDA soy “Checkoff” program), Protein Industries Canada (a Government of Canada 
Global Innovation Cluster), Almond Board of California, European Fruit Juice 
Association, The Tate and Lyle Nutritional Research Fund at the University of 
Toronto, The Glycemic Control and Cardiovascular Disease in Type 2 Diabetes Fund 
at the University of Toronto (a fund established by the Alberta Pulse Growers), 
The Plant Protein Fund at the University of Toronto (a fund which has received 
contributions from IFF among other donors), The Plant Milk Fund at the 
University of Toronto (a fund established by the Karuna Foundation through Vegan 
Grants), and The Nutrition Trialists Network Fund at the University of Toronto 
(a fund established by donations from the Calorie Control Council and Physicians 
Committee for Responsible Medicine). He has received food donations to support 
randomized controlled trials from the Almond Board of California, California 
Walnut Commission, Danone, Nutrartis, Soylent, and Dairy Farmers of Canada. He 
has received travel support, speaker fees and/or honoraria from Danone, 
FoodMinds LLC, Nestlé, Abbott, General Mills, Nutrition Communications, 
International Food Information Council (IFIC), Arab Beverages, International 
Sweeteners Association, Association Calorie Control Council, and Phynova. He has 
or has had ad hoc consulting arrangements with Perkins Coie LLP, Tate & Lyle, 
Ingredion, and Brightseed. He is on the Clinical Practice Guidelines Expert 
Committees of Diabetes Canada, European Association for the study of Diabetes 
(EASD), Canadian Cardiovascular Society (CCS), and Obesity Canada/Canadian 
Association of Bariatric Physicians and Surgeons. He serves as an unpaid member 
of the Board of Trustees of IAFNS. He is a Director at Large of the Canadian 
Nutrition Society (CNS), founding member of the International Carbohydrate 
Quality Consortium (ICQC), Executive Board Member of the Diabetes and Nutrition 
Study Group (DNSG) of the EASD, and Director of the Toronto 3D Knowledge 
Synthesis and Clinical Trials foundation. His spouse is an employee of AB 
InBev. All other authors declare that they have no competing interests.


20. Clin Pharmacokinet. 2024 Jun;63(6):773-818. doi: 10.1007/s40262-024-01377-0. 
Epub 2024 May 28.

Together or Apart? Revealing the Impact of Dietary Interventions on 
Bioavailability of Quinolones: A Systematic Review with Meta-analyses.

Wiesner A(1)(2), Zagrodzki P(2), Gawalska A(3), Paśko P(4).

Author information:
(1)Doctoral School of Medical and Health Sciences, Jagiellonian University 
Medical College, św. Łazarza 16, 31-530, Krakow, Poland.
(2)Department of Food Chemistry and Nutrition, Faculty of Pharmacy, Jagiellonian 
University Medical College, Medyczna 9, 30-688, Krakow, Poland.
(3)Department of Medicinal Chemistry, Faculty of Pharmacy, Jagiellonian 
University Medical College, Medyczna 9, 30-688, Krakow, Poland.
(4)Department of Food Chemistry and Nutrition, Faculty of Pharmacy, Jagiellonian 
University Medical College, Medyczna 9, 30-688, Krakow, Poland. 
.

BACKGROUND AND OBJECTIVE: Managing drug-food interactions is essential for 
optimizing the effectiveness and safety profile of quinolones. Following PRISMA 
guidelines, we systematically reviewed the influence of dietary interventions on 
the bioavailability of 22 quinolones.
METHODS: All studies describing or investigating the impact of food, beverages, 
antacids, and mineral supplements on pharmacokinetic parameters or 
pharmacokinetic/pharmacodynamic indices of orally taken quinolones were 
considered for inclusion. We excluded reviews, in vitro and in silico studies, 
studies performed on animals, and those involving alcohol. We performed the 
search in Medline (via PubMed), Embase, and Cochrane Library, covering reports 
from database inception to December 2022. We used the following tools to assess 
the risk of bias: version 2 of the Cochrane risk-of-bias tool for parallel 
trials, the Cochrane risk-of-bias tool for cross-over studies, and the NIH 
quality assessment tool for before-after studies. We performed quantitative 
analyses for each quinolone if two or more food-effect studies with specified 
and comparable study designs were available. If meta-analyses were not 
applicable, we qualitatively summarized the results.
RESULTS: We included 109 studies from 101 reports. Meta-analyses were conducted 
for 12 antibiotics and qualitative synthesis was employed for the remaining 
drugs. Of the studies, 60.5% were open-label, cross-over, as recommended by FDA. 
We judged 46% of studies as having a high risk of bias and only 4% of having a 
low risk of bias. Among 19 quinolones with available food impact data, 14 (74%) 
had potentially clinically important interactions. For nalidixic acid, oxolinic 
acid, and tosufloxacin, food exerted a high positive impact on bioavailability 
(AUC or Cmax increased by > 45%), whereas, for all the remaining drugs, 
postprandial absorption was lower. The most significant negative influence of 
food (AUC or Cmax decreased by > 40%) occurred for delafloxacin capsules and 
norfloxacin, whereas the moderate influence (AUC or Cmax decreased by 30-40%) 
occurred for nemonoxacin and rufloxacin. All 14 analysed quinolones showed a 
substantial reduction in bioavailability when co-administered with antacids and 
mineral supplements, except for calcium preparations. The impact of beverages 
was evaluated for 10 quinolones, with 50% experiencing significantly reduced 
absorption in the presence of milk (the highest negative impact for 
ciprofloxacin). Moreover, both ciprofloxacin and levofloxacin demonstrated 
compromised bioavailability when consumed with orange juice, particularly 
calcium-fortified.
DISCUSSION: Several factors may influence interactions, including the 
physicochemical characteristics of quinolones, the type of intervention, drug 
formulation, and the patient's health status. We assessed the quality of 
evidence as low due to the poor actuality of included studies, their 
methodological diversity, and uneven data availability for individual drugs.

© 2024. The Author(s).

DOI: 10.1007/s40262-024-01377-0
PMCID: PMC11222276
PMID: 38807006 [Indexed for MEDLINE]

Conflict of interest statement: The authors have no competing interests to 
declare that are relevant to the content of this article.


21. Food Res Int. 2024 Jul;187:114420. doi: 10.1016/j.foodres.2024.114420. Epub 2024 
Apr 26.

Microencapsulation of vitamins: A review and meta-analysis of coating materials, 
release and food fortification.

Espinoza-Espinoza LA(1), Muñoz-More HD(2), Nole-Jaramillo JM(3), Ruiz-Flores 
LA(3), Arana-Torres NM(3), Moreno-Quispe LA(4), Valdiviezo-Marcelo J(3).

Author information:
(1)Departamento de Ingeniería, Universidad Nacional de Barranca, Barranca 15169, 
Peru.
(2)Laboratorio de Alimentos Funcionales y Bioprocesos - Facultad de Ingeniería 
de Industrias alimentarias, Universidad Nacional de Frontera, Sullana 20100, 
Peru. Electronic address:.
(3)Laboratorio de Alimentos Funcionales y Bioprocesos - Facultad de Ingeniería 
de Industrias alimentarias, Universidad Nacional de Frontera, Sullana 20100, 
Peru.
(4)Facultad de Ciencias empresariales y Turismo, Universidad Nacional de 
Frontera, Sullana 20100, Peru.

Vitamins are responsible for providing biological properties to the human body; 
however, their instability under certain environmental conditions limits their 
utilization in the food industry. The objective was to conduct a systematic 
review on the use of biopolymers and lipid bases in microencapsulation 
processes, assessing their impact on the stability, controlled release, and 
viability of fortified foods with microencapsulated vitamins. The literature 
search was conducted between the years 2013-2023, gathering information from 
databases such as Scopus, PubMed, Web of Science and publishers including Taylor 
& Francis, Elsevier, Springer and MDPI; a total of 49 articles were compiled The 
results were classified according to the microencapsulation method, considering 
the following information: core, coating material, solvent, formulation, process 
conditions, particle size, efficiency, yield, bioavailability, bioaccessibility, 
in vitro release, correlation coefficient and references. It has been evidenced 
that gums are the most frequently employed coatings in the protection of 
vitamins (14.04%), followed by alginate (10.53%), modified chitosan (9.65%), 
whey protein (8.77%), lipid bases (8.77%), chitosan (7.89%), modified starch 
(7.89%), starch (7.02%), gelatin (6.14%), maltodextrin (5.26%), zein (3.51%), 
pectin (2.63%) and other materials (7.89%). The factors influencing the release 
of vitamins include pH, modification of the coating material and crosslinking 
agents; additionally, it was determined that the most fitting mathematical model 
for release values is Weibull, followed by Zero Order, Higuchi and 
Korsmeyer-Peppas; finally, foods commonly fortified with microencapsulated 
vitamins were described, with yogurt, bakery products and gummy candies being 
notable examples.

Copyright © 2024 Elsevier Ltd. All rights reserved.

DOI: 10.1016/j.foodres.2024.114420
PMID: 38763670 [Indexed for MEDLINE]

Conflict of interest statement: Declaration of competing interest The authors 
declare that they have no known competing financial interests or personal 
relationships that could have appeared to influence the work reported in this 
paper.


22. J Nutr. 2024 Jun;154(6):1827-1841. doi: 10.1016/j.tjnut.2024.04.031. Epub 2024 
Apr 28.

The Relationship Between Vitamin D Intake and Serum 25-hydroxyvitamin D in Young 
Children: A Meta-Regression to Inform WHO/FAO Vitamin D Intake Recommendations.

Rios-Leyvraz M(1), Martino L(2), Cashman KD(3).

Author information:
(1)Consultant, Department of Nutrition and Food Safety, WHO, Geneva, 
Switzerland. Electronic address:.
(2)Department of Risk Assessment Services, Methodology and Scientific Support 
Unit, European Food Safety Authority, Parma, Italy.
(3)Department of Medicine, Cork Centre for Vitamin D and Nutrition Research, 
School of Food and Nutritional Sciences, University College Cork, Cork, Ireland.

BACKGROUND: This work was commissioned by the World Health Organization and Food 
and Agriculture Organization to inform their update on the vitamin D 
requirements for children aged <4 y.
OBJECTIVES: The objective of this work was to undertake multilevel and 
multivariable dose-response modeling of serum 25-hydroxyvitamin D (25OHD) to 
total vitamin D intake in children aged <4 y with the goal of deriving updated 
vitamin D requirements for young children.
METHODS: Systematically identified randomized controlled trials among healthy 
children from 2 wk up to 3.9 y of age provided with daily vitamin D supplements 
or vitamin D-fortified foods were included. Linear and nonlinear random effects 
multilevel meta-regression models with and without covariates were fitted and 
compared. Interindividual variability was included by simulating the individual 
serum 25OHD responses. The percentage of individuals reaching set minimal and 
maximal serum 25OHD thresholds was calculated and used to derive vitamin D 
requirements.
RESULTS: A total of 31 trials with 186 data points from North America, Europe, 
Asia, and Australasia/Oceania, with latitudes ranging from 61°N to 38°S, and 
with participants of likely mostly light or medium skin pigmentation, were 
included. In 29 studies the children received vitamin D supplements and in 2 
studies the children received vitamin D-fortified milk with or without 
supplements. The dose-response relationship between vitamin D intake and serum 
25OHD was best fitted with the unadjusted quadratic model. Adding additional 
covariates, such as age, did not significantly improve the model. At a vitamin D 
intake of 10 μg/d, 97.3% of the individuals were predicted to achieve a minimal 
serum 25OHD threshold of 28 nmol/L. At a vitamin D intake of 35 μg/d, 1.4% of 
the individuals predicted to reach a maximal serum 25OHD threshold of 200 
nmol/L.
CONCLUSIONS: In conclusion, this paper details the methodological steps taken to 
derive vitamin D requirements in children aged <4 y, including the addition of 
an interindividual variability component.

Copyright © 2024. Published by Elsevier Inc.

DOI: 10.1016/j.tjnut.2024.04.031
PMCID: PMC11217029
PMID: 38685317 [Indexed for MEDLINE]


23. Nutrients. 2024 Mar 21;16(6):910. doi: 10.3390/nu16060910.

Association of Fortification with Human Milk versus Bovine Milk-Based Fortifiers 
on Short-Term Outcomes in Preterm Infants-A Meta-Analysis.

Galis R(1)(2), Trif P(1)(3), Mudura D(1), Mazela J(2), Daly MC(4), Kramer BW(2), 
Diggikar S(5).

Author information:
(1)Department of Neonatology, Emergency County Hospital Bihor, 50 Clujului 
Street, 410053 Oradea, Romania.
(2)Department of Neonatology, Poznan University of Medical Sciences, 10 Fredry 
Street, 61-701 Poznan, Poland.
(3)Doctoral School of Biomedical Sciences, University of Oradea, 1 University 
Street, 410087 Oradea, Romania.
(4)Irish Neonatal Health Alliance (INHA), 26 Oak Glen View, Southern Cross, A98 
Y234 Bray, Ireland.
(5)Oyster Woman and Child Hospital, Bengaluru 560043, India.

This meta-analysis assessed short-term outcomes after using human milk-derived 
fortifiers (HMFs) compared with bovine milk fortifiers (BMFs) in preterm infants 
fed an exclusive human milk (HM) diet, either mother's own milk (MOM) or donor 
human milk (DHM). We searched PubMed, Embase, Google Scholar, CENTRAL and CINHAL 
between January 2015 and August 2023 for studies reporting outcomes in infants 
with ≤28 weeks gestation and/or birthweight ≤ 1500 g on an exclusive human milk 
diet fortified with HMF versus BMF. The primary outcomes were death and NEC 
(stage ≥ 2). Four studies with a total of 681 infants were included. Mortality 
was significantly lower in infants fed with an HM-HMFs diet (four studies, 681 
infants; RR = 0.50, 95% CI = 0.26-0.94; p = 0.03; I2 = 0%), NEC was similar 
between the two groups (four studies, 681 infants; RR = 0.48, 95% CI = 
0.20-1.17; p = 0.11; I2= 39%). BPD was higher in the HM-BMFs group (four 
studies, 663 infants; RR = 0.83, 95% CI = 0.69-1.000; p = 0.05, I2 = 0%), 
although not statistically significant. No differences were found for sepsis (RR 
= 0.97, 95% CI = 0.66-1.42; p = 0.96; I2 = 26%) or combined ROP (four studies, 
671 infants; RR = 0.64, 95% CI = 0.53-1.07; p = 0.28; I2 = 69%). An HM-HMFs diet 
could possibly be associated with decreased mortality with no association with 
NEC, BPD, sepsis, or ROP. This meta-analysis was limited by the small number of 
studies included. However, the results should not be refuted for this reason as 
they provide an impetus for subsequent clinical trials to assess the observed 
associations.

DOI: 10.3390/nu16060910
PMCID: PMC10975992
PMID: 38542821 [Indexed for MEDLINE]

Conflict of interest statement: The authors declare no conflict of interest.


24. J Steroid Biochem Mol Biol. 2024 Apr;238:106448. doi: 
10.1016/j.jsbmb.2023.106448. Epub 2023 Dec 21.

Strategic food vehicles for vitamin D fortification and effects on vitamin D 
status: A systematic review and meta-analysis of randomised controlled trials.

Cashman KD(1), O'Neill CM(2).

Author information:
(1)Cork Centre for Vitamin D and Nutrition Research, School of Food and 
Nutritional Sciences, University College Cork, Cork, Ireland. Electronic 
address:.
(2)Cork Centre for Vitamin D and Nutrition Research, School of Food and 
Nutritional Sciences, University College Cork, Cork, Ireland.

There has been growing interest in the potential of vitamin D food fortification 
in Europe as a means of addressing low vitamin D status. The WHO-FAO suggest 
that choosing a suitable food vehicle and ensuring the combination of the food 
vehicle and the fortificant will be efficacious and effective are of key 
importance to a successful food fortification programme. Our key objective was 
to conduct a systematic review and meta-analysis to investigate the effect of 
various animal- and plant-based food vehicles fortified with vitamin D (as D3 or 
D2) on circulating 25-hydroxyvitamin D [25(OH)D] concentrations. A list of 
prioritised food vehicles was established and we searched PubMed, Embase, Scopus 
and Web of Science for randomised controlled trials (RCTs) which used these 
vehicles individually, and which met prespecified criteria. The searches 
identified 49 papers which described suitable RCTs using vitamin D-fortified 
bread/savoury biscuits (n = 5), orange juice (n = 5), UV-mushrooms (n = 8), 
cheese (n = 3), yogurt (n = 5), fluid milk (n = 13), powdered milk (n = 5), eggs 
(n = 2), edible oils (n = 4), or breakfast cereal (n = 1). No suitable RCTs were 
identified for rice, maize flour, butter, margarine or dairy spreads, 
plant-based milk or yogurt alternatives. Random-effects meta-analyses of each 
food vehicle individually indicated weighted mean differences (WMD) in 25(OH)D 
in the range ∼9-35 nmol/L (3-15 RCT arms, depending on vehicle), and all 
statistically significant (P < 0.01-0.0001), with the exception of UV-mushrooms 
(P = 0.06). Heterogeneity was variable (I2 =33-99%, depending on vehicle), but 
subgroup analysis based on vitamer and dose reduced it in some instances. 
Sub-group analysis on the basis of whether the food vehicles were from 
plant-based or animal-based origin showed no significant difference in WMD (15.2 
versus 15.9 nmol/L, respectively; P = 0.48). These results support the use of 
various animal- and plant-based food vehicles for vitamin D fortification to 
improve circulating 25(OH)D concentrations in populations. This work was 
registered with PROSPERO as CRD42023439883.

Copyright © 2024 Elsevier Ltd. All rights reserved.

DOI: 10.1016/j.jsbmb.2023.106448
PMID: 38141736 [Indexed for MEDLINE]


25. Adv Nutr. 2023 Sep;14(5):1187-1196. doi: 10.1016/j.advnut.2023.06.010. Epub 2023 
Jul 4.

The Effects of Dairy Product Supplementation on Bone Health Indices in Children 
Aged 3 to 18 Years: A Meta-Analysis of Randomized Controlled Trials.

Hidayat K(1), Zhang LL(2), Rizzoli R(3), Guo YX(2), Zhou Y(2), Shi YJ(4), Su 
HW(4), Liu B(5), Qin LQ(6).

Author information:
(1)Department of Nutrition and Food Hygiene, School of Public Health, Soochow 
University, Suzhou, China. Electronic address:.
(2)Department of Nutrition and Food Hygiene, School of Public Health, Soochow 
University, Suzhou, China.
(3)Division of Bone Diseases, Department of Internal Medicine, Geneva University 
Hospitals and Faculty of Medicine, Geneva, Switzerland.
(4)Branch Company, Inner Mongolia Yili Industrial Group Co, Ltd, Hohhot, China.
(5)Branch Company, Inner Mongolia Yili Industrial Group Co, Ltd, Hohhot, China. 
Electronic address:.
(6)Department of Nutrition and Food Hygiene, School of Public Health, Soochow 
University, Suzhou, China. Electronic address:.

Childhood and adolescence are critical periods for optimizing skeletal growth. 
Dairy products are valuable sources of bone-beneficial nutrients, particularly 
calcium and protein. A random-effects meta-analysis of published randomized 
controlled trials was performed to quantitatively assess the effects of dairy 
supplementation on bone health indices in children and adolescents. The PubMed 
and Web of Science databases were searched. Dairy supplementation increased 
whole-body bone mineral content (BMC) (+25.37 g) and areal bone mineral density 
(aBMD) (+0.016 g/cm2), total hip BMC (+0.49 g) and aBMD (+0.013 g/cm2), femoral 
neck BMC (+0.06 g) and aBMD (+0.030 g/cm2), lumbar spine BMC (+0.85 g) and aBMD 
(+0.019 g/cm2), and height (0.21 cm). When expressed as a percentage difference, 
whole-body BMC was increased by 3.0%, total hip BMC by 3.3%, femoral neck BMC by 
4.0%, lumbar spine BMC by 4.1%, whole-body aBMD by 1.8%, total hip aBMD by 1.2%, 
femoral neck aBMD by 1.5%, and lumbar spine aBMD by 2.6%. Dairy supplementation 
increased serum insulin-like growth factor I concentrations (19.89 nmol/L) and 
reduced concentrations of urinary deoxypyridinoline (-1.78 nmol/mmol creatinine) 
and serum parathyroid hormone (-10.46 pg/mL) but did not significantly affect 
the serum concentrations of osteocalcin, bone alkaline phosphatase, and 
C-terminal telopeptide of type 1 collagen. Serum 25-hydroxyvitamin D 
concentrations (+4.98 ng/mL) increased with vitamin D-fortified dairy 
supplementation. The positive effects on bone mineral mass parameters and height 
were generally consistent across subgroups defined by sex, geographical region, 
baseline calcium intake, calcium from the supplementation, trial duration, and 
Tanner stages. In summary, dairy supplementation during growth leads to a small 
but significant increase in bone mineral mass parameters, and these findings are 
generally supported by the changes in several biochemical parameters related to 
bone health.

Copyright © 2023 The Authors. Published by Elsevier Inc. All rights reserved.

DOI: 10.1016/j.advnut.2023.06.010
PMCID: PMC10509403
PMID: 37414219 [Indexed for MEDLINE]


26. Clin Nutr ESPEN. 2023 Apr;54:1-11. doi: 10.1016/j.clnesp.2023.01.001. Epub 2023 
Jan 4.

Effect of intake of iron-fortified milk on levels of ferritin and hemoglobin in 
preschoolers: A systematic review and meta-analysis.

Diniz Largueza CB(1), Mocellin MC(2), Nunes JC(3), Ribas SA(4).

Author information:
(1)Postgraduate Program in Food and Nutritional Security, Federal University of 
the State of Rio de Janeiro, Rio de Janeiro, Brazil. Electronic address: 
.
(2)Fundamental Nutrition Department, Federal University of the State of Rio de 
Janeiro, Rio de Janeiro, Brazil.
(3)Post-graduate Program in Food and Nutritional Security, Food Science 
Department, Federal University of the State of Rio de Janeiro, Rio de Janeiro, 
Brazil.
(4)Post-graduate Program in Food and Nutritional Security, Program in Food and 
Nutritional Security, Federal University of the State of Rio de Janeiro, Rio de 
Janeiro, Brazil.

BACKGROUND: Food fortification is often the simplest way to increase iron intake 
on a broad and sustainable basis. It is one of the most cost-effective global 
development efforts.
OBJECTIVE: To systematize data from randomized and nonrandomized controlled 
trials investigating the effect of consumption of iron-fortified milk (IFM) on 
serum iron status in preschoolers.
METHODS: The trials were systematically searched in five electronic databases 
that evaluated the effect of iron-fortified milk (IFM) consumption on hemoglobin 
and ferritin in children aged 1-6 years. The randomized the meta-analysis model 
was used to calculate total and stratified effects.
RESULTS: Seven trials representing 1210 preschoolers showed a significant effect 
of IFM on hemoglobin [difference in means (MD) 0.33 g/dL (95% CI 0.23, 0.44; 
I2 = 3.85%, p (Q statistic) < 0.01)] and ferritin concentrations [effect size 
(SMD) 0.57 (95% CI 0.19, 0.95; I2 = 84.63%, p (Q statistic) < 0.01)]. In 
stratified analyses, hemoglobin showed an increase when subjects received iron 
with other co-interventions [MD 0.35 g/dL (95% CI 0.22, 0.48; I2 = 11.01%; p (Q 
statistic) = 0.36)]; and when the iron dose was above 5 mg/day [MD 0.34 g/dL 
(95% CI 0.23, 0.45; I2 = 2.33%; p (Q statistic) = 0.37)] and the intervention 
time was over 6 months [MD 0.39 g/dL (95% CI 0.18, 0.60; I2 = 37.90%; p (Q 
statistic) = 0.20)]. For ferritin, stratified analyses showed a larger effect 
size when they used dose higher than 5 mg/day [SMD 0.60 (95% CI 0.02, 1.18; 
I2 = 91.06, p (Q statistic) < 0.01)] and intervention was conducted longer than 
6 months [SMD 0.96 (95% CI - 0.16, 1.76; I2 = 93.38%, p (Q statistic) < 0.01)].
CONCLUSIONS: Our findings indicate that although milk iron fortification 
modestly increases serum hemoglobin levels or the effect size of serum ferritin 
of participants, it cannot be considered in the coadjuvant treatment for anemia. 
Register: CRD42020213604.

Copyright © 2023 European Society for Clinical Nutrition and Metabolism. 
Published by Elsevier Ltd. All rights reserved.

DOI: 10.1016/j.clnesp.2023.01.001
PMID: 36963849 [Indexed for MEDLINE]

Conflict of interest statement: Declaration of competing interest The authors 
have declared no conflicts of interest.


27. Nutrients. 2023 Feb 21;15(5):1076. doi: 10.3390/nu15051076.

Effectiveness of Dietary Management for Moderate Wasting among Children > 6 
Months of Age-A Systematic Review and Meta-Analysis Exploring Different Types, 
Quantities, and Durations.

Cichon B(1), Das JK(2)(3), Salam RA(4), Padhani ZA(2), Stobaugh HC(5)(6), Mughal 
M(1), Pajak P(1), Rutishauser-Perera A(1), Bhutta ZA(2)(3)(7), Black RE(8).

Author information:
(1)Action Against Hunger UK, Operations Department, London SE10 0ER, UK.
(2)Institute for Global Health and Development, Aga Khan University, Karachi 
74800, Pakistan.
(3)Division of Women and Child Health, Aga Khan University, Karachi 74800, 
Pakistan.
(4)The Daffodil Centre, The University of Sydney, Sydney 2006, Australia.
(5)Action Against Hunger USA, Technical Services and Innovation Department, 
Washington, DC 20463, USA.
(6)Friedman School of Nutrition Science and Policy, Tufts University, Boston, MA 
02111, USA.
(7)Centre for Global Child Health, Hospital for Sick Children, Toronto, ON M5G 
0A4, Canada.
(8)Bloomberg School of Public Health, Johns Hopkins University, Baltimore, MD 
21205, USA.

Currently, no World Health Organization guidelines exist for the management of 
approximately 31.8 million moderately wasted children globally. The objective of 
this review was to synthesise evidence on the optimal type, quantity, and 
duration of dietary treatment for moderate wasting. Ten electronic databases 
were searched until the 23rd of August 2021. Experimental studies comparing 
interventions for the dietary management of moderate wasting were included. 
Meta-analyses were conducted and results were presented as risk ratios or mean 
differences with 95% confidence intervals. Seventeen studies comparing specially 
formulated foods were included involving 23,005 participants. Findings suggest 
little or no difference in recovery between Fortified Blended Foods (FBFs) with 
improved micronutrient and/or milk content (enhanced FBFs) and lipid-based 
nutrient supplements (LNS), whereas children treated with non-enhanced FBFs 
(locally produced FBFs or standard corn-soy blend) may have lower recovery rates 
than those treated with LNS. There was no difference in recovery when 
ready-to-use therapeutic and ready-to-use supplementary food were compared. 
Other outcomes mostly aligned with results for recovery. In conclusion, LNSs 
improve recovery compared to non-enhanced FBFs, but are comparable to enhanced 
FBFs. Programmatic choice of supplement should consider factors such as cost, 
cost-effectiveness, and acceptability. Further research is required to determine 
optimal dosing and duration of supplementation.

DOI: 10.3390/nu15051076
PMCID: PMC10005276
PMID: 36904076 [Indexed for MEDLINE]

Conflict of interest statement: B.C. and H.S. were co-authors of some of the 
studies included in this review. As stated in the acknowledgements, the PICO 
question was developed by the WHO Organization and the Guideline Development 
Group for the revision of the guidelines on the prevention and treatment of 
wasting.


28. Matern Child Nutr. 2023 Jul;19(3):e13488. doi: 10.1111/mcn.13488. Epub 2023 Feb 
26.

The effect of interventions distributing home fortification products on infant 
and young child feeding (IYCF) practices: A systematic narrative review.

Locks LM(1)(2), Newell KB(1), Imohe A(3), Moloney GM(3), Shaker-Berbari L(3), 
Paudyal N(4), Jefferds MED(5).

Author information:
(1)Department of Health Sciences, College of Health and Rehabilitation Sciences: 
Sargent College, Boston University, Boston, Massachusetts, USA.
(2)Department of Global Health, Boston University, Boston, Massachusetts, USA.
(3)United Nation Children's Fund (UNICEF) Headquarters, New York City, New York, 
USA.
(4)United Nation Children's Fund (UNICEF) Nepal Country Office, Kathmandu, 
Nepal.
(5)Centers for Disease Control and Prevention, Atlanta, Georgia, USA.

Interventions distributing micronutrient powders (MNPs) and small-quantity 
lipid-based nutrient supplements (SQ-LNS), or home fortification products 
(HFPs), have the potential to improve infant and young child feeding (IYCF) 
practices and children's nutrition. We systematically searched for studies on 
the effect of interventions distributing HFP on IYCF practices. We identified 12 
(8 MNP, 4 SQ-LNS) studies: seven programmes with IYCF behaviour change 
communications (BCC) and MNP (IYCF-MNP) and one provided MNP without IYCF BCC 
(MNP only). Three SQ-LNS studies came from randomised trials without an IYCF 
component (SQ-LNS only) and one from a programme with both IYCF BCC and SfQ-LNS 
(IYCF-SQ-LNS). Five IYCF-MNP programmes reported positive associations with some 
IYCF practices-four with minimum dietary diversity, two with minimum meal 
frequency, four with minimum acceptable diet, and three with the initiation of 
complementary foods at 6 months. Two reported no association between MNP and 
IYCF indicators, and one reported a decline in IYCF practices during the 
intervention, although it also reported significant changes to the IYCF 
programme during the evaluation period. Two studies from interventions that 
distributed SQ-LNS (one from a related set of randomised controlled trials and 
the sole IYCF-SQ-LNS programme) reported a positive association with IYCF 
practices; one trial reported no change in breast milk intake with the provision 
of SQ-LNS and one found no association with IYCF practices. SQ-LNS and MNP can 
address nutrient gaps for young children in low-resource settings; our findings 
indicate that programmes that combine HFP with IYCF interventions may also 
contribute to improved IYCF practices in some settings.

© 2023 The Authors. Maternal & Child Nutrition published by John Wiley & Sons 
Ltd. This article has been contributed to by U.S. Government employees and their 
work is in the public domain in the USA.

DOI: 10.1111/mcn.13488
PMCID: PMC10262895
PMID: 36842164 [Indexed for MEDLINE]

Conflict of interest statement: The findings and conclusions in this report are 
those of the authors and do not necessarily represent the official position of 
the Centres for Disease Control and Prevention or UNICEF. The authors have no 
conflict of interest to declare.


29. Nutrients. 2022 Nov 28;14(23):5060. doi: 10.3390/nu14235060.

Effect of Fortified Formula on Growth and Nutritional Status in Young Children: 
A Systematic Review and Meta-Analysis.

Brooker PG(1), Rebuli MA(1), Williams G(1), Muhlhausler BS(1).

Author information:
(1)Health and Biosecurity, Commonwealth Scientific and Industrial Research 
Organisation (CSIRO), Adelaide, BC 5000, Australia.

Previous reviews of the effect of young child formulas on health outcomes in 
infants and toddlers have been inconclusive. In this study, we undertook a 
contemporary synthesis of studies investigating the effects of consuming 
fortified milk beverages (compared to cow’s milk or unfortified comparator 
formula) on growth and/or nutritional status in children 1−3 years of age. Five 
electronic databases were searched (PubMed, Web of Science, Scopus, ProQuest, 
and Cochrane Library) for randomised controlled trials comparing fortified milk 
against control milk in young children (9−48 months), published between January 
1990 and June 2022. Outcomes were growth, body composition, biochemical markers, 
and/or nutritional status. Mean differences (MD) were pooled using 
random-effects meta-analysis where there were ≥3 studies. The risk of bias was 
assessed using the Cochrane Risk of Bias 2.0 tool. Nineteen articles (12 
studies; n = 4795) met the inclusion criteria. Heterogeneity was substantial, 
likely attributable to considerable variation in study characteristics. 
Fortified milk was associated with increased weight gain (MD = 0.14 kg [95% CI 
0.06, 021], p = 0.0003) compared with control milk. Subgroup analyses 
demonstrated increases in weight in lower-income countries, and in studies with 
intervention periods > 6 months. There were no effects of fortified milks on 
other anthropometric measures. Haemoglobin (MD = 3.76 g/L [95% CI 0.17, 7.34], p 
= 0.04) and ferritin (MD = 0.01 nmol/L [95% CI 0.00, 0.02], p = 0.02) 
concentrations were increased in infants consuming fortified milks. Fortified 
milk beverages appear to offer a safe and acceptable source of complementary 
nutrition as a short-term strategy for addressing nutritional deficits and may 
modestly promote weight gain in vulnerable populations when provided for periods 
> 6 months. This study was prospectively registered with PROSPERO 
(CRD42022339920) and funded by the Infant Nutrition Council.

DOI: 10.3390/nu14235060
PMCID: PMC9737957
PMID: 36501090 [Indexed for MEDLINE]

Conflict of interest statement: The funders had no role in the design of the 
study; in the collection, analyses, or interpretation of data; in the writing of 
the manuscript; or in the decision to publish the results. P.G.B., M.A.R. and 
G.W. declare no conflicts of interest. B.S.M. serves on the advisory board of 
the Nestle Nutrition Institute (NNI) of Australia and New Zealand as an 
independent expert research advisor. NNI had no involvement in the funding, 
design or execution of this project.


30. Nutr Rev. 2023 Mar 10;81(4):428-440. doi: 10.1093/nutrit/nuac067.

Linkage between a plant-based diet and age-related eye diseases: a systematic 
review and meta-analysis.

Cirone C(1), Cirone KD(2), Malvankar-Mehta MS(3).

Author information:
(1)is with the School of Nutrition, Toronto Metropolitan University, Toronto, 
Ontario, Canada.
(2)is with the Schulich School of Medicine and Dentistry, The University of 
Western Ontario, London, Ontario, Canada.
(3)is with the Department of Ophthalmology, and the Department of Epidemiology 
and Biostatistics, Schulich School of Medicine and Dentistry, The University of 
Western Ontario, London, Ontario, Canada.

CONTEXT: An increased risk of age-related eye disease has been observed in 
individuals lacking a balanced diet. Following a plant-based diet may result in 
nutritional insufficiencies and negatively affect health if an effort is not 
made to ensure the consumption of fortified foods or specific supplements.
OBJECTIVE: The purpose of this article is to characterize the relationship 
between plant-based diets and age-related ocular outcomes among adults.
DATA SOURCES: A comprehensive literature review was performed using the MEDLINE, 
Embase, Cochrane, and PubMed databases up until December 19, 2021.
STUDY SELECTION: Studies that focused on observed visual changes due to a 
reduced intake of animal products and that included a minimum of 50 eyes were 
eligible for inclusion.
DATA EXTRACTION: Two levels of screening, quality assessment, and data 
extraction were conducted by 2 reviewers independently. The 21 studies 
identified from 814 unique studies progressed to data extraction and 15 were 
included in the quantitative analysis using STATA 15.0 fixed-effect and 
random-effect models computed on the basis of heterogeneity.
RESULTS: The 15 (n = 51 695 participants) assessed the impact of fish 
consumption, 8 studies (n = 28 753 participants) analyzed the effect of red meat 
intake, and 3 studies (n = 7723 participants) assessed the impact of omission of 
skim milk, poultry, and non-meat animal products and the presence of disease 
incidence as indicated by age-related macular degeneration or cataract 
development. Meta-analysis indicated regular consumption of fish (odds ratio 
[OR], 0.70; 95%CI, 0.62-0.79) and skim milk, poultry, and non-meat animal 
products (OR, 0.70; 95%CI, 0.61-0.79) reduced the risk of age-related eye 
disease development among adults. Consumption of red meat (OR, 1.41; 95%CI, 
1.07-1.86) may increase the risk of age-related eye disease development.
CONCLUSION: A pescatarian diet is associated with the most favorable visual 
outcomes among adults, whereas consumption of red meat negatively affects ocular 
health. Results suggest a need for more initiatives promoting a healthy and 
balanced diet.
SYSTEMATIC REVIEW REGISTRATION: PROSPERO registration no. CRD42021269925.

© The Author(s) 2022. Published by Oxford University Press on behalf of the 
International Life Sciences Institute. All rights reserved. For permissions, 
please.

DOI: 10.1093/nutrit/nuac067
PMID: 36102832 [Indexed for MEDLINE]


31. Front Pediatr. 2022 May 17;10:874976. doi: 10.3389/fped.2022.874976. eCollection 
2022.

Evaluation of Risk and Preventive Factors for Necrotizing Enterocolitis in 
Premature Newborns. A Systematic Review of the Literature.

Campos-Martinez AM(1), Expósito-Herrera J(1), Gonzalez-Bolívar M(1), 
Fernández-Marin E(1), Uberos J(1)(2).

Author information:
(1)Neonatal Intensive Care Unit, San Cecilio Clinical Hospital, Granada, Spain.
(2)Pediatrics Department, Medicine Faculty, Granada, Spain.

Necrotizing enterocolitis (NEC) is a serious condition related to prematurity 
and the initiation of enteral feeding. In this article, we review the evidence 
published in recent years on necrotizing enterocolitis risk factors 
(prematurity, feeding with low-weight formula, existence of intestinal 
dysbiosis) and protective factors (human milk or donated milk supply, 
supplementation of human milk with oligosaccharides, probiotics administration, 
and the determination of disease predictive biomarkers). A systematic review was 
conducted of preventive, risk and predictive factors for necrotizing 
enterocolitis in neonates prior to 37 weeks' gestational age, based on a 
literature search for clinical trials, meta-analyses, randomized controlled 
trials and systematic reviews published between January 2018 and October 2021. 
For this purpose, the PubMed, MEDLINE, and Cochrane Library databases were 
consulted. The literature search obtained 113 articles, of which 19 were 
selected for further analysis after applying the inclusion and exclusion 
criteria. The conclusions drawn from this analysis were that adequate knowledge 
of risk factors that can be prevented or modified (such as alteration of the 
intestinal microbiota, oxidative stress, metabolic dysfunction at birth, or 
alteration of the immunity modulation) can reduce the incidence of NEC in 
premature infants. These factors include the supplementation of enteral 
nutrition with human milk oligosaccharides (with prebiotic and immunomodulatory 
effects), the combined administration of probiotics (especially the 
Lactobacillus spp and Bifidobacterium spp combination, which inhibits bacterial 
adhesion effects, improves the intestinal mucosa barrier function, strengthens 
the innate and adaptive immune system and increases the secretion of bioactive 
metabolites), the supplementation of human milk with lactoferrin and the use of 
donated milk fortified in accordance with the characteristics of the premature 
newborn. The determination of factors that can predict the existence of NEC, 
such as fecal calprotectin, increased TLR4 activity, and IL6 receptor, can lead 
to an early diagnosis of NEC. Although further studies should be conducted to 
determine the values of predictive biomarkers of NEC, and/or the recommended 
doses and strains of probiotics, lactoferrin or oligosaccharides, the knowledge 
acquired in recent years is encouraging.

Copyright © 2022 Campos-Martinez, Expósito-Herrera, Gonzalez-Bolívar, 
Fernández-Marin and Uberos.

DOI: 10.3389/fped.2022.874976
PMCID: PMC9152085
PMID: 35656377

Conflict of interest statement: The authors declare that the research was 
conducted in the absence of any commercial or financial relationships that could 
be construed as a potential conflict of interest.


32. Nutrients. 2022 Apr 21;14(9):1724. doi: 10.3390/nu14091724.

Efficacy of Donated Milk in Early Nutrition of Preterm Infants: A Meta-Analysis.

Li Y(1)(2), Chi C(2), Li C(2), Song J(2), Song Z(3), Wang W(2), Sun J(3)(4).

Author information:
(1)School of Nursing, Weifang Medical University, Weifang 261042, China.
(2)School of Nursing, Jining Medical University, Jining 272067, China.
(3)School of Medicine and Dentistry, Griffith University, Gold Coast, QLD 4215, 
Australia.
(4)Menzies Health Institute Queensland, Griffith University, Gold Coast, QLD 
4215, Australia.

Background: Preterm birth is associated with an increased risk of many 
complications, which is a main public health problem worldwide with social and 
economic consequences. Human milk from breast feeding has been proved to be the 
optimal nutrition strategy for preterm infants when available. However, the lack 
of human milk from mothers makes formula widely used in clinical practice. In 
recent years, donated breast milk has gained popularity as an alternative choice 
which can provide human milk oligosaccharides and other bioactive substances. 
Objective: We aimed to conduct a systematic review and meta-analysis to evaluate 
the nutritional effects of donated breast milk on preterm infants compared with 
formula. Method: In the present study, we searched Medline, Web of Science, 
Embase, clinicaltrials.gov, the China national knowledge infrastructure, and the 
Cochrane central register of controlled trials for candidate randomized 
controlled trials (RCTs). Results: A total of 1390 patients were enrolled in 11 
RCTs and meta-analysis results showed that donated breast milk is also more 
advantageous in reducing the incidence of necrotizing enterocolitis (NEC, RR = 
0.67, 95% CI = 0.48 to 0.93, p = 0.02), reducing the duration of parenteral 
nutrition (MD = −2.39, 95% CI = −3.66 to −1.13, p = 0.0002) and the time of full 
enteral feeding (MD = −0.33, 95% CI = −3.23 to 2.57, p = 0.0002). In comparison, 
formula significantly promotes the growth of premature infants, including their 
weight gain (MD = −3.45, 95% CI = −3.68 to −3.21, p < 0.00001), head growth (MD 
= −0.07, 95% CI = −0.08 to −0.06, p < 0.00001) and body length (MD = −0.13, 95% 
CI = −0.15 to −0.11, p < 0.00001), and reduces the time it takes for premature 
infants to regain birth weight (MD = 6.60, 95% CI = 6.11 to 7.08, p < 0.00001. 
Conclusion: Compared with formula, donated breast milk could significantly 
reduce the incidence of NEC, the duration of parenteral nutrition, and the time 
of full enteral feeding. Adding fortifiers in donated milk could make it as 
effective as formula in promoting the physical growth of premature infants.

DOI: 10.3390/nu14091724
PMCID: PMC9105142
PMID: 35565692 [Indexed for MEDLINE]

Conflict of interest statement: The authors declare no conflict of interest.


33. Front Pediatr. 2022 Feb 25;9:793311. doi: 10.3389/fped.2021.793311. eCollection 
2021.

Exclusive Maternal Milk Compared With Exclusive Formula on Growth and Health 
Outcomes in Very-Low-Birthweight Preterm Infants: Phase II of the Pre-B Project 
and an Evidence Analysis Center Systematic Review.

Taylor SN(1), Fenton TR(2)(3)(3), Groh-Wargo S(4), Gura K(5), Martin CR(6), 
Griffin IJ(7)(8), Rozga M(9), Moloney L(9).

Author information:
(1)Division of Neonatology, Department of Pediatrics, Yale School of Medicine, 
New Haven, CT, United States.
(2)Community Health Sciences, Institute of Public Health, Alberta Children's 
Hospital Research Institute, Cumming School of Medicine, University of Calgary, 
Calgary, AB, Canada.
(3)Nutrition Services, Alberta Health Services, Calgary, AB, Canada.
(4)Departments of Nutrition and Pediatrics, Case Western Reserve University at 
MetroHealth Medical Center, Cleveland, OH, United States.
(5)Clinical Research Program, Department of Pharmacy, Boston Children's 
Hospital, Boston, MA, United States.
(6)Division of Translational Research, Department of Neonatology, Harvard 
Medical School, Neonatal Intensive Care Unit (NICU), Beth Israel Deaconess 
Medical Center, Boston, MA, United States.
(7)Biomedical Research Institute of New Jersey, Cedar Knolls, NJ, United States.
(8)Department of Pediatrics, Morristown Medical Center, Morristown, NJ, United 
States.
(9)Academy of Nutrition and Dietetics, Evidence Analysis Center, Chicago, IL, 
United States.

As part of the Pre-B Project, a systematic review was conducted to evaluate 
associations between exclusive maternal milk (≥75%) intake and exclusive formula 
intake and growth and health outcomes in very-low-birthweight (VLBW) preterm 
infants. The protocols from the Academy of Nutrition and Dietetics' Evidence 
Analysis Center and the Preferred Reporting Items for Systematic Reviews and 
Meta-Analyses (PRISMA) checklist were followed. Thirteen observational studies 
were included; 11 studies reported data that could be synthesized in a pooled 
analysis. The evidence is very uncertain (very low quality) about the effect of 
exclusive maternal milk on all outcomes due to observational study designs and 
risk of selection, performance, detection, and reporting bias in most of the 
included studies. Very-low-quality evidence suggested that providing VLBW 
preterm infants with exclusive maternal milk was not associated with mortality, 
risk of necrotizing enterocolitis, sepsis, or developing bronchopulmonary 
dysplasia, as compared with exclusive preterm formula, but exclusive maternal 
milk was associated with a lower risk of retinopathy of prematurity (very low 
certainty). Results may change when additional studies are conducted. There was 
no difference in weight, length, and head circumference gain between infants fed 
fortified exclusive maternal milk and infants receiving exclusive preterm 
formula; however, weight and length gain were lower in infants fed non-fortified 
exclusive maternal milk. Given the observational nature of human milk research, 
cause-and-effect evidence was lacking for VLBW preterm infants.
SYSTEMATIC REVIEW REGISTRATION: 
https://www.crd.york.ac.uk/prospero/display_record.php?RecordID=86829, PROSPERO 
ID: CRD42018086829.

Copyright © 2022 Taylor, Fenton, Groh-Wargo, Gura, Martin, Griffin, Rozga and 
Moloney.

DOI: 10.3389/fped.2021.793311
PMCID: PMC8913886
PMID: 35280446

Conflict of interest statement: The authors declare that the research was 
conducted in the absence of any commercial or financial relationships that could 
be construed as a potential conflict of interest.


34. Afr Health Sci. 2021 Sep;21(3):1346-1354. doi: 10.4314/ahs.v21i3.46.

Ingestion of supplements and fortified food with iodine on the breast milk 
iodine concentration in deficiency areas: a systematic review.

Machamba AAL(1), Priore SE(2), Macedo MS(3), Franceschini SDCC(4).

Author information:
(1)Almeida A. L. Machamba: PhD Student in the Postgraduate Program in Nutrition 
Science, Department of Nutrition and Health, Federal University of Viçosa (UFV), 
Viçosa, Brazil.
(2)Full PhD Professor in the the Postgraduate Program in Nutrition Science, 
Department of Nutrition and Health, Federal University of Viçosa (UFV), Viçosa, 
Brazil.
(3)Post-Doctoral in the Postgraduate Program in Nutrition Science, Department of 
Nutrition and Health, Federal University of Viçosa (UFV), Viçosa, Brazil.
(4)Full PhD Professor in the the Postgraduate Program in Nutrition Science, 
Department of Nutrition and Health and Pro-Rector of community affairs, Federal 
University of Viçosa (UFV), Viçosa, Brazil.

INTRODUCTION: The level of iodine in breast milk may be inadequate and 
compromise the health of this, both due to excess and lack, some population 
groups remain deficient because of the low consumption of iodate salt, because 
there is an increase in consumption of other sources of iodine, such as 
supplements and fortified foods.
OBJECTIVE: To evaluate the effect of the consumption of fortified foods and 
nutritional supplements with iodine on maternal milk levels.
METHODOLOGY: Systematic review based on the Prism method, using the descriptors 
provided by DeCS. The reading, selection and analysis of the methodological 
quality of the articles was done by two researchers independently.
RESULTS: From 346 abstracts, 6 were eligible. The median iodination range 
between the studies ranged from 75 to 600 µg in supplements and 150 and 225 µg 
in fortified foods with effect on increased iodine concentration of breastmilk 
(BMIC), achieving the adequacy of the median BMIC in 4 of the 6 studies.
CONCLUSION: Iodine ingestion through supplements or fortified foods results in 
improved iodine levels in breast milk.

© 2021 Machamba AAL et al.

DOI: 10.4314/ahs.v21i3.46
PMCID: PMC8843278
PMID: 35222600 [Indexed for MEDLINE]


35. Children (Basel). 2022 Jan 24;9(2):150. doi: 10.3390/children9020150.

Effects of Formula Milk Feeding in Premature Infants: A Systematic Review.

Moreira-Monteagudo M(1), Leirós-Rodríguez R(2), Marqués-Sánchez P(2).

Author information:
(1)Faculty of Physical Therapy, Universidade de Vigo, Campus a Xunqueira, s/n, 
36005 Pontevedra, Spain.
(2)SALBIS Research Group, Faculty of Health Sciences, University of León, 
Astorga Ave. s/n, 24401 Ponferrada, Spain.

The preterm baby is born at a critical period for the growth and development of 
the gastrointestinal and neuromotor systems. Breast milk is the food of choice 
for infants during the first months of life, as it provides multiple short- and 
long-term benefits to preterm and sick newborns. Despite this, breastfeeding is 
often nutritionally insufficient, requiring the addition of fortifiers. In other 
cases, it is important to ensure the necessary nutrients and calories, which can 
be provided by formula milk or pasteurized and fortified donated human milk. 
However, the specific guidelines for the use of formula milk have not yet been 
determined. Therefore, a systematic search was considered necessary in order to 
identify the effects of feeding with formula milk in preterm infants. A 
systematic search in Scopus, Medline, Pubmed, Cinahl, ClinicalTrials and Web of 
Science with the terms Infant Formula and Infant Premature was conducted. A 
total of 18 articles were selected, of which, eight were experimental and ten 
were observational studies. Among the objectives of the analyzed investigations, 
we distinguished nine that compared the effects of feeding with formula milk, 
breast milk and donated human milk, five that evaluated the effects of different 
compositions of formula milk and/or fortifiers and four investigations that 
compared the effects of formula milk and donated human milk. In conclusion, when 
breast milk is insufficient or unavailable, formula milk is a good nutritional 
option, due to its higher caloric density and protein content. Nevertheless, the 
preterm infant's diet should incorporate breast milk to reduce the incidence of 
morbidities such as necrotizing enterocolitis and sepsis (related to hospital 
handling of fortifiers and formula milk).

DOI: 10.3390/children9020150
PMCID: PMC8870637
PMID: 35204871

Conflict of interest statement: The authors declare no conflict of interest.


36. Adv Nutr. 2022 Aug 1;13(4):1186-1199. doi: 10.1093/advances/nmab136.

The Effects of Milk Supplementation on Bone Health Indices in Adults: A 
Meta-Analysis of Randomized Controlled Trials.

Hidayat K(1), Chen JS(1), Wang TC(1), Liu YJ(1), Shi YJ(2), Su HW(2), Liu B(2), 
Qin LQ(1).

Author information:
(1)Department of Nutrition and Food Hygiene, School of Public Health, Soochow 
University, Suzhou, China.
(2)Branch Company, Inner Mongolia Yili Industrial Group Co. Ltd., Hohhot, China.

Milk contains a number of bone-beneficial nutrients. However, milk, due to the 
D-galactose content, might have unfavorable effects on bone health. A 
meta-analysis of randomized controlled trials (RCTs) was performed to clarify 
the effects of milk supplementation on bone mineral density (BMD), bone turnover 
markers [N-terminal telopeptide of type I collagen (NTx), C-terminal telopeptide 
of type 1 collagen (CTx), osteocalcin, bone alkaline phosphatase (BALP), and 
procollagen type 1 N-propeptide (P1NP)], and hormonal indices related to bone 
metabolism [parathyroid hormone (PTH), 25-hydroxyvitamin D [25(OH)D], and 
insulin-like growth factor 1 (IGF-1)] in adults. The PubMed and Web of Science 
databases were searched. A random-effects model was used to estimate the pooled 
effect sizes. A total of 20 RCTs were included. The trial duration ranged from 1 
mo to 36 mo. Milk supplementation resulted in a small but significant increase 
in BMD at the hip (+0.004 g/cm2; n = 9 RCTs) and lumbar spine (+0.025 g/cm2; 
n = 7), but did not significantly affect whole-body BMD (n = 3) and femoral neck 
BMD (n = 7). Milk supplementation reduced the concentrations of P1NP 
(-5.20 ng/mL; n = 9), CTx (-0.16 ng/mL; n = 9), and NTx (-8.66 nmol bone 
collagen equivalents/mmol creatinine; n = 3). The concentrations of osteocalcin 
(n = 9) and BALP (n = 3) were not affected by milk supplementation. Reduced 
parathyroid hormone PTH (-1.01 pg/mL; n = 13) concentrations and increased IGF-1 
(+1.79 nmol/l; n = 4) concentrations were observed with milk supplementation. 
25(OH)D (+3.73 ng/mL; n = 11) concentrations were increased with vitamin-D 
fortified milk supplementation. The addition of milk to the diet may potentially 
increase the likelihood of preventing bone loss by restoring bone homeostasis 
through the modulation of the calcium-vitamin D-PTH axis, bone remodeling rate, 
and growth hormone/IGF-1 axis.

© The Author(s) 2021. Published by Oxford University Press on behalf of the 
American Society for Nutrition.

DOI: 10.1093/advances/nmab136
PMCID: PMC9340984
PMID: 34792092 [Indexed for MEDLINE]


37. J Paediatr Child Health. 2022 Jan;58(1):30-38. doi: 10.1111/jpc.15810. Epub 2021 
Oct 20.

Early versus delayed introduction of human milk fortification in enterally fed 
preterm infants: A systematic review and meta-analysis.

Hilditch C(1)(2)(3)(4), Keir A(1)(2)(3)(4), Collins CT(1)(2), Middleton 
P(1)(2)(3), Gomersall J(1)(5).

Author information:
(1)SAHMRI Women and Kids, South Australian Health and Medical Research 
Institute, Adelaide, South Australia, Australia.
(2)Adelaide Medical School, The University of Adelaide, Adelaide, South 
Australia, Australia.
(3)Robinson Research Institute, The University of Adelaide, Adelaide, South 
Australia, Australia.
(4)Women's and Children's Health Network, Adelaide, South Australia, Australia.
(5)School of Public Health, The University of Adelaide, Adelaide, South 
Australia, Australia.

AIM: To assess effects of early versus delayed introduction of human milk 
fortification in preterm infants.
METHODS: We searched Cochrane Central Register of Controlled Trials, MEDLINE, 
Embase, PubMed and CINAHL for randomised controlled trials evaluating start time 
for human milk fortification in preterm infants (March 2020). Two authors 
assessed trial eligibility and risk of bias, extracted data and assessed 
evidence certainty.
RESULTS: We identified 1307 publications and included three trials (378 
infants). Meta-analysis comparing fortification commencing at an enteral feed 
volume of ≤40 mL/kg/day versus ≥75 mL/kg/day, showed little to no difference in 
rates of necrotising enterocolitis (3 trials), sepsis (3 trials), feeding 
intolerance (2 trials) (low-quality evidence) and infant growth (1 trial, very 
low-quality evidence).
CONCLUSIONS: Whether early introduction of fortification, at an enteral feed 
volume of ≤40 mL versus delayed at ≥75 mL/kg/day improves growth or influences 
adverse feeding outcomes is very uncertain.

© 2021 Paediatrics and Child Health Division (The Royal Australasian College of 
Physicians).

DOI: 10.1111/jpc.15810
PMID: 34669996 [Indexed for MEDLINE]


38. BMJ. 2021 Oct 13;375:n2202. doi: 10.1136/bmj.n2202.

Conduct and reporting of formula milk trials: systematic review.

Helfer B(1)(2), Leonardi-Bee J(3), Mundell A(4), Parr C(1), Ierodiakonou D(5), 
Garcia-Larsen V(1)(6), Kroeger CM(7), Dai Z(7)(8), Man A(1), Jobson J(1), Dewji 
F(1), Kunc M(1), Bero L(9), Boyle RJ(10)(11).

Author information:
(1)National Heart and Lung Institute, Imperial College London, London, UK.
(2)Institute of Psychology, University of Wroclaw, Wroclaw, Poland.
(3)Centre for Evidence Based Healthcare, Faculty of Medicine and Health 
Sciences, University of Nottingham, Nottingham, UK.
(4)Imperial College Healthcare NHS Trust, London, UK.
(5)Department of Primary Care and Population Health, University of Nicosia 
Medical School, Nicosia, Cyprus.
(6)Program in Human Nutrition, Department of International Health, Johns Hopkins 
University, Baltimore, MD, USA.
(7)Faculty of Medicine and Health, University of Sydney, Sydney, NSW, Australia.
(8)Australian Institute of Health Innovation, Faculty of Medicine, Health and 
Human Sciences, Macquarie University, Sydney, NSW, Australia.
(9)Center for Bioethics and Humanities, Schools of Medicine and Public Health, 
University of Colorado Anschutz Medical Center, Aurora, CO, USA.
(10)National Heart and Lung Institute, Imperial College London, London, UK 
.
(11)Centre of Evidence Based Dermatology, University of Nottingham, Nottingham, 
UK.

Erratum in
    BMJ. 2021 Nov 24;375:n2890. doi: 10.1136/bmj.n2890.

OBJECTIVE: To systematically review the conduct and reporting of formula trials.
DESIGN: Systematic review.
DATA SOURCES: Medline, Embase, and Cochrane Central Register of Controlled 
Trials (CENTRAL) were searched from 1 January 2006 to 31 December 2020.
REVIEW METHODS: Intervention trials comparing at least two formula products in 
children less than three years of age were included, but not trials of human 
breast milk or fortifiers of breast milk. Data were extracted in duplicate and 
primary outcome data were synthesised for meta-analysis with a random effects 
model weighted by the inverse variance method. Risk of bias was evaluated with 
Cochrane risk of bias version 2.0, and risk of undermining breastfeeding was 
evaluated according to published consensus guidance. Primary outcomes of the 
trials included in the systematic review were identified from clinical trial 
registries, protocols, or trial publications.
RESULTS: 22 201 titles were screened and 307 trials were identified that were 
published between 2006 and 2020, of which 73 (24%) trials in 13 197 children 
were prospectively registered. Another 111 unpublished but registered trials in 
17 411 children were identified. Detailed analysis was undertaken for 125 trials 
(23 757 children) published since 2015. Seventeen (14%) of these recently 
published trials were conducted independently of formula companies, 26 (21%) 
were prospectively registered with a clear aim and primary outcome, and authors 
or sponsors shared prospective protocols for 11 (9%) trials. Risk of bias was 
low in five (4%) and high in 100 (80%) recently published trials, mainly because 
of inappropriate exclusions from analysis and selective reporting. For 68 
recently published superiority trials, a pooled standardised mean difference of 
0.51 (range -0.43 to 3.29) was calculated with an asymmetrical funnel plot 
(Egger's test P<0.001), which reduced to 0.19 after correction for asymmetry. 
Primary outcomes were reported by authors as favourable in 86 (69%) trials, and 
115 (92%) abstract conclusions were favourable. One of 38 (3%) trials in 
partially breastfed infants reported adequate support for breastfeeding and 14 
of 87 (16%) trials in non-breastfed infants confirmed the decision not to 
breastfeed was firmly established before enrolment in the trial.
CONCLUSIONS: The results show that formula trials lack independence or 
transparency, and published outcomes are biased by selective reporting.
SYSTEMATIC REVIEW REGISTRATION: PROSPERO 2018 CRD42018091928.

© Author(s) (or their employer(s)) 2019. Re-use permitted under CC BY-NC. No 
commercial re-use. See rights and permissions. Published by BMJ.

DOI: 10.1136/bmj.n2202
PMCID: PMC8513520
PMID: 34645600 [Indexed for MEDLINE]

Conflict of interest statement: Competing interests: All authors have completed 
the ICMJE uniform disclosure form at www.icmje.org/disclosure-of-interest/ and 
declare support from Imperial Health Charity: RJB received personal fees from 
Cochrane, DBV Technologies, and Prota Therapeutics, and from expert witness work 
in cases of food anaphylaxis and class actions related to infant formula health 
claims, outside the submitted work, and received personal fees from Public 
Health England as a member of the UK Nutrition and Health Claims Committee and 
the Maternal and Child Nutrition Subgroup of the Scientific Advisory Committee 
on Nutrition. JL-B received fees from Danone Nutricia Research and the Food 
Standards Agency, outside of the submitted work. The University of Colorado 
receives remuneration for LB’s work as senior editor, Cochrane, which is outside 
the submitted work. The authors report no other relationships or activities that 
could appear to have influenced the submitted work.


39. Clin Nutr ESPEN. 2021 Oct;45:26-32. doi: 10.1016/j.clnesp.2021.08.019. Epub 2021 
Sep 3.

Lactoferrin reduces the risk of respiratory tract infections: A meta-analysis of 
randomized controlled trials.

Ali AS(1), Hasan SS(2), Kow CS(3), Merchant HA(4).

Author information:
(1)Department of Medicine, Aga Khan University Hospital, Karachi, Pakistan; 
School of Public Health, Dow University of Health Sciences, Karachi, Pakistan.
(2)Department of Pharmacy, School of Applied Sciences, University of 
Huddersfield, Huddersfield, United Kingdom; School of Biomedical Sciences and 
Pharmacy, University of Newcastle, Newcastle, Australia.
(3)School of Pharmacy, Monash University Malaysia, Selangor, Malaysia.
(4)Department of Pharmacy, School of Applied Sciences, University of 
Huddersfield, Huddersfield, United Kingdom. Electronic address: 
.

BACKGROUND: Lactoferrin (Lf) is one of the key immunomodulatory substances found 
naturally in various body fluids, such as saliva, tears, and breast milk, and 
forms a vital part of the innate defense against invading pathogens. Various 
studies have demonstrated antibacterial, antifungal, and antiviral properties of 
Lf and its protective role against respiratory tract infections (RTIs). The 
present meta-analysis aims to elucidate the association of Lf administration in 
reducing the risk of RTIs by systematically reviewing the data from randomized 
controlled trials (RCTs).
METHODS: We systematically searched PubMed, Cochrane Library, Medline & CINAHL, 
Turning Research into Practice (TRIP), ProQuest Theses & Dissertations 
Databases, and China National Knowledge Infrastructure (CNKI) from inception 
till March 15, 2021. The primary outcome measure was a reduction in respiratory 
illness; decrease in frequency, symptoms, and duration. Random-effects model was 
used to estimate the odds ratio (OR) and 95% confidence interval (CI). We used 
Cochrane's RoB-2 to appraise the risk of bias of included RCTs.
RESULTS: A total of nine RCTs were eligible for this review, of which six were 
included in the meta-analysis. Overall, two studies demonstrated a high risk of 
bias. The meta-analysis revealed a significantly reduced odds of developing 
respiratory infections with the use of Lf relative to the control (pooled odds 
ratio = 0.57; 95% confidence interval 0.44 to 0.74, n = 1,194), with sufficient 
evidence against the hypothesis of 'no significant difference' at the current 
sample size.
CONCLUSIONS: The administration of Lf shows promising efficacy in reducing the 
risk of RTIs. Current evidence also favours Lf fortification of infant formula. 
Lf may also have a beneficial role in managing symptoms and recovery of patients 
suffering from RTIs and may have potential for use as an adjunct in COVID-19, 
however this warrants further evidence from a large well-designed RCT.

Copyright © 2021 European Society for Clinical Nutrition and Metabolism. 
Published by Elsevier Ltd. All rights reserved.

DOI: 10.1016/j.clnesp.2021.08.019
PMID: 34620326 [Indexed for MEDLINE]

Conflict of interest statement: Declaration of competing interest HAM is on a 
scientific advisory board for a lactoferrin supplement product but does not have 
any financial or other affiliation with the company and has neither received any 
funding or financial reimbursements. HAM is committed to promote the science 
behind Lf and its immunomodulatory role in the prevention and management of 
infections.


40. Cochrane Database Syst Rev. 2021 Sep 26;9(9):CD013092. doi: 
10.1002/14651858.CD013092.pub2.

Nutrition-specific interventions for preventing and controlling anaemia 
throughout the life cycle: an overview of systematic reviews.

da Silva Lopes K(1), Yamaji N(2), Rahman MO(2), Suto M(3), Takemoto Y(4), 
Garcia-Casal MN(5), Ota E(6).

Author information:
(1)Graduate School of Public Health, St. Luke's International University, Tokyo, 
Japan.
(2)Global Health Nursing, Graduate School of Nursing Science, St. Luke's 
International University, Tokyo, Japan.
(3)Department of Health Policy, National Center for Child Health and 
Development, Tokyo, Japan.
(4)Department of Obstetrics and Gynaecology, School of Medicine, Juntendo 
University, Tokyo, Japan.
(5)Department of Nutrition and Food Safety, World Health Organization, Geneva , 
Switzerland.
(6)Global Health Nursing, Graduate School of Nursing Science, St. Luke's 
International University , Tokyo, Japan.

Update of
    doi: 10.1002/14651858.CD013092.

BACKGROUND: Anaemia is a prevalent health problem worldwide. Some types are 
preventable or controllable with iron supplementation (pills or drops), 
fortification (sprinkles or powders containing iron added to food) or 
improvements to dietary diversity and quality (e.g. education or counselling).
OBJECTIVES: To summarise the evidence from systematic reviews regarding the 
benefits or harms of nutrition-specific interventions for preventing and 
controlling anaemia in anaemic or non-anaemic, apparently healthy populations 
throughout the life cycle.
METHODS: In August 2020, we searched MEDLINE, Embase and 10 other databases for 
systematic reviews of randomised controlled trials (RCTs) in anaemic or 
non-anaemic, apparently healthy populations. We followed standard Cochrane 
methodology, extracting GRADE ratings where provided. The primary outcomes were 
haemoglobin (Hb) concentration, anaemia, and iron deficiency anaemia (IDA); 
secondary outcomes were iron deficiency (ID), severe anaemia and adverse effects 
(e.g. diarrhoea, vomiting).
MAIN RESULTS: We included 75 systematic reviews, 33 of which provided GRADE 
assessments; these varied between high and very low. Infants (6 to 23 months; 13 
reviews) Iron supplementation increased Hb levels and reduced the risk of 
anaemia and IDA in two reviews. Iron fortification of milk or cereals, 
multiple-micronutrient powder (MMNP), home fortification of complementary foods, 
and supplementary feeding increased Hb levels and reduced the risk of anaemia in 
six reviews. In one review, lipid-based nutrient supplementation (LNS) reduced 
the risk of anaemia. In another, caterpillar cereal increased Hb levels and IDA 
prevalence. Food-based strategies (red meat and fortified cow's milk, beef) 
showed no evidence of a difference (1 review). Preschool and school-aged 
children (2 to 10 years; 8 reviews) Daily or intermittent iron supplementation 
increased Hb levels and reduced the risk of anaemia and ID in two reviews. One 
review found no evidence of difference in Hb levels, but an increased risk of 
anaemia and ID for the intermittent regime. All suggested that zinc plus iron 
supplementation versus zinc alone, multiple-micronutrient (MMN)-fortified 
beverage versus control, and point-of-use fortification of food with 
iron-containing micronutrient powder (MNP) versus placebo or no intervention may 
increase Hb levels and reduce the risk of anaemia and ID. Fortified dairy 
products and cereal food showed no evidence of a difference on the incidence of 
anaemia (1 review). Adolescent children (11 to 18 years; 4 reviews) Compared 
with no supplementation or placebo, five types of iron supplementation may 
increase Hb levels and reduce the risk of anaemia (3 reviews). One review on 
prevention found no evidence of a difference in anaemia incidence on iron 
supplementation with or without folic acid, but Hb levels increased. Another 
suggested that nutritional supplementation and counselling reduced IDA. One 
review comparing MMN fortification with no fortification observed no evidence of 
a difference in Hb levels. Non-pregnant women of reproductive age (19 to 49 
years; 5 reviews) Two reviews suggested that iron therapy (oral, intravenous 
(IV), intramuscular (IM)) increased Hb levels; one showed that iron folic acid 
supplementation reduced anaemia incidence; and another that daily iron 
supplementation with or without folic acid or vitamin C increased Hb levels and 
reduced the risk of anaemia and ID. No review reported interventions related to 
fortification or dietary diversity and quality. Pregnant women of reproductive 
age (15 to 49 years; 23 reviews) One review apiece suggested that: daily iron 
supplementation with or without folic acid increased Hb levels in the third 
trimester or at delivery and in the postpartum period, and reduced the risk of 
anaemia, IDA and ID in the third trimester or at delivery; intermittent iron 
supplementation had no effect on Hb levels and IDA, but increased the risk of 
anaemia at or near term and ID, and reduced the risk of side effects; vitamin A 
supplementation alone versus placebo, no intervention or other micronutrient 
might increase maternal Hb levels and reduce the risk of maternal anaemia; MMN 
with iron and folic acid versus placebo reduced the risk of anaemia; 
supplementation with oral bovine lactoferrin versus oral ferrous iron 
preparations increased Hb levels and reduced gastrointestinal side effects; MNP 
for point-of-use fortification of food versus iron and folic acid 
supplementation might decrease Hb levels at 32 weeks' gestation and increase the 
risk of anaemia; and LNS versus iron or folic acid and MMN increased the risk of 
anaemia. Mixed population (all ages; 22 reviews) Iron supplementation versus 
placebo or control increased Hb levels in healthy children, adults, and elderly 
people (4 reviews). Hb levels appeared to increase and risk of anaemia and ID 
decrease in two reviews investigating MMN fortification versus placebo or no 
treatment, iron fortified flour versus control, double fortified salt versus 
iodine only fortified salt, and rice fortification with iron alone or in 
combination with other micronutrients versus unfortified rice or no 
intervention. Each review suggested that fortified versus non-fortified 
condiments or noodles, fortified (sodium iron ethylenediaminetetraacetate; 
NaFeEDTA) versus non-fortified soy sauce, and double-fortified salt versus 
control salt may increase Hb concentration and reduce the risk of anaemia. One 
review indicated that Hb levels increased for children who were anaemic or had 
IDA and received iron supplementation, and decreased for those who received 
dietary interventions. Another assessed the effects of foods prepared in iron 
pots, and found higher Hb levels in children with low-risk malaria status in two 
trials, but no difference when comparing food prepared in non-cast iron pots in 
a high-risk malaria endemicity mixed population. There was no evidence of a 
difference for adverse effects. Anaemia and malaria prevalence were rarely 
reported. No review focused on women aged 50 to 65 years plus or men (19 to 65 
years plus).
AUTHORS' CONCLUSIONS: Compared to no treatment, daily iron supplementation may 
increase Hb levels and reduce the risk of anaemia and IDA in infants, preschool 
and school-aged children and pregnant and non-pregnant women. Iron fortification 
of foods in infants and use of iron pots with children may have prophylactic 
benefits for malaria endemicity low-risk populations. In any age group, only a 
limited number of reviews assessed interventions to improve dietary diversity 
and quality. Future trials should assess the effects of these types of 
interventions, and consider the requirements of different populations.

Copyright © 2021 The Cochrane Collaboration. Published by John Wiley & Sons, 
Ltd.

DOI: 10.1002/14651858.CD013092.pub2
PMCID: PMC8464655
PMID: 34564844 [Indexed for MEDLINE]

Conflict of interest statement: Katharina da Silva Lopes: none known Noyuri 
Yamaji: none known Md Obaidur Rahman: none known Maiko Suto: none known Yo 
Takemoto: none known Maria Nieves García‐Casal (MNGC) is a full‐time member of 
staff at the Department of Nutrition and Food Safety, World Health Organization 
(WHO). MNGC declares no other known conflicts of interest. Erika Ota's 
institution received funding (2017/725990) for this work from the Evidence and 
Programme Guidance, Department of Nutrition for Health and Development, WHO, 
Switzerland. EO, KL and MNGC are authors of included reviews (EO: Abe 2016, 
Rumbold 2015, Suchdev 2020; KL: Suchdev 2020; MNGC: Garcia‐Casal 2018, 
Peña‐Rosas 2015b, Peña‐Rosas 2019). EO and MNGC were not involved in review 
selection, data extraction and assessment of methodological quality using AMSTAR 
(Shea 2007a; Shea 2007b; Shea 2009). KL was involved in these tasks but not in 
relation to her own reviews; in those cases, these tasks were performed by two 
other independent review authors (OR, NY). Disclaimer: The review authors alone 
are responsible for the views expressed in this publication and they do not 
necessarily represent the decisions, policy or views of the WHO.


41. Nutrients. 2021 Jun 18;13(6):2089. doi: 10.3390/nu13062089.

A Systematic Review and Meta-Analysis of Human Milk Feeding and Short-Term 
Growth in Preterm and Very Low Birth Weight Infants.

Suganuma M(1), Rumbold AR(1)(2), Miller J(1)(3), Chong YF(3), Collins CT(1)(2).

Author information:
(1)SAHMRI Women and Kids, South Australian Health and Medical Research 
Institute, Adelaide, SA 5006, Australia.
(2)Adelaide Medical School, The University of Adelaide, Adelaide, SA 5006, 
Australia.
(3)College of Nursing and Health Sciences, Flinders University, Adelaide, SA 
5001, Australia.

Human milk (HM) is the gold standard for feeding infants but has been associated 
with slower growth in preterm infants compared with preterm formula. This 
systematic review and meta-analysis summarises the post-1990 literature to 
examine the effect of HM feeding on growth during the neonatal admission of 
preterm infants with birth weight ≤1500 g and/or born ≤28 weeks' gestation. 
Medline, PubMed, CINAHL, and Scopus were searched, and comparisons were grouped 
as exclusive human milk (EHM) vs. exclusive preterm formula (EPTF), any HM vs. 
EPTF, and higher vs. lower doses of HM. We selected studies that used fortified 
HM and compared that with a PTF; studies comparing unfortified HM and term 
formula were excluded. Experimental and observational studies were pooled 
separately. The GRADE system was used to evaluate risk of bias and certainty of 
evidence. Forty-four studies were included with 37 (n = 9963 infants) included 
in the meta-analyses. In general, due to poor quality studies, evidence of the 
effect of any HM feeds or higher versus lower doses of HM was inconclusive. 
There was a possible effect that lower doses of HM compared with higher doses of 
HM improved weight gain during the hospital admission, and separately, a 
possible effect of increased head circumference growth in infants fed EPTF vs. 
any HM. The clinical significance of this is unclear. There was insufficient 
evidence to determine the effects of an exclusive HM diet on any outcomes.

DOI: 10.3390/nu13062089
PMCID: PMC8234723
PMID: 34207261 [Indexed for MEDLINE]

Conflict of interest statement: C.C. salary was supported by a Research 
Fellowship (ID 1132596) from the National Health and Medical Research Council 
(NHMRC), Australia. The views expressed in this article are solely the 
responsibility of the authors and do not reflect the views of the NHMRC. C.C. is 
an author on an included paper (Jacobi-Polishook 2016). All other authors 
declare no conflict of interest.


42. Nutr Clin Pract. 2021 Dec;36(6):1144-1162. doi: 10.1002/ncp.10663. Epub 2021 Jun 
8.

Comparative effectiveness of liquid human milk fortifiers: A systematic review 
and meta-analysis.

Bridges KM(1)(2), Newkirk M(3), Byham-Gray L(1), Chung M(1)(4).

Author information:
(1)Department of Clinical and Preventive Nutrition Sciences, Rutgers, The State 
University of New Jersey, Newark, New Jersey, USA.
(2)Neonatal Intensive Care Unit, Beaumont Children's Hospital, Royal Oak, 
Michigan, USA.
(3)Neonatal Intensive Care Unit, Johns Hopkins All Children's Hospital, Saint 
Petersburg, Florida, USA.
(4)Department of Public Health and Community Medicine, Tufts University School 
of Medicine, Boston, Massachusetts, USA.

OBJECTIVE: To determine the effectiveness of liquid human milk fortifiers (HMFs) 
derived from exclusive HM or hydrolyzed protein on growth, necrotizing 
enterocolitis (NEC), or late-onset sepsis in North American very 
low-birth-weight (VLBW) infants compared with powder HMFs (control).
METHODS: Prospective trials published between 2009 and 2020 were systematically 
reviewed, and meta-analysis was conducted by using a random-effects model.
RESULTS: Five studies were identified for up to 591 participants across 39 
centers. Study treatments included whey or casein hydrolysate HMF and exclusive 
HM HMF. Infants fed whey or casein hydrolysate HMF had growth differences 
compared with the control. No differences were found across treatments in regard 
to NEC or sepsis.
CONCLUSION: Very low-quality evidence suggests greater linear growth in VLBW 
infants fed whey hydrolysate liquid HMF, as well as greater weight gain in those 
fed casein hydrolysate HMF, compared with the control. Additional prospective, 
multicenter randomized controlled trials are needed to confirm these estimates 
because of sparsity of evidence. There is insufficient evidence to support HMF 
decisions regarding NEC or late-onset sepsis prophylaxis.

© 2021 American Society for Parenteral and Enteral Nutrition.

DOI: 10.1002/ncp.10663
PMID: 34101248 [Indexed for MEDLINE]


43. Gut Microbes. 2021 Jan-Dec;13(1):1-33. doi: 10.1080/19490976.2021.1884514.

A systematic review of the factors influencing microbial colonization of the 
preterm infant gut.

Aguilar-Lopez M(1), Dinsmoor AM(1), Ho TTB(2), Donovan SM(1)(3).

Author information:
(1)Division of Nutritional Sciences, University of Illinois at Urbana-Champaign, 
Urbana, USA.
(2)Department of Pediatrics, Morsani College of Medicine, University of South 
Florida, Tampa, USA.
(3)Department of Food Science and Human Nutrition, University of Illinois at 
Urbana-Champaign, Urbana, USA.

Prematurity coupled with the necessary clinical management of preterm (PT) 
infants introduces multiple factors that can interfere with microbial 
colonization. This study aimed to review the perinatal, physiological, 
pharmacological, dietary, and environmental factors associated with gut 
microbiota of PT infants. A total of 587 articles were retrieved from a search 
of multiple databases. Sixty studies were included in the review after removing 
duplicates and articles that did not meet the inclusion criteria. Review of this 
literature revealed that evidence converged on the effect of postnatal age, mode 
of delivery, use of antibiotics, and consumption of human milk in the 
composition of gut microbiota of PT infants. Less evidence was found for 
associations with race, sex, use of different fortifiers, macronutrients, and 
other medications. Future studies with rich metadata are needed to further 
explore the impact of the PT exposome on the development of the microbiota in 
this high-risk population.

DOI: 10.1080/19490976.2021.1884514
PMCID: PMC8023245
PMID: 33818293 [Indexed for MEDLINE]


44. Matern Child Nutr. 2021 Jul;17(3):e13176. doi: 10.1111/mcn.13176. Epub 2021 Mar 
17.

The effect of milk type and fortification on the growth of low-birthweight 
infants: An umbrella review of systematic reviews and meta-analyses.

North K(1), Marx Delaney M(2), Bose C(1), Lee ACC(3)(4), Vesel L(2), Adair L(5), 
Semrau K(2)(4).

Author information:
(1)Department of Pediatrics, University of North Carolina at Chapel Hill, Chapel 
Hill, North Carolina, USA.
(2)Ariadne Labs, Brigham Women's Hospital and Harvard TH Chan School of Public 
Health, Boston, Massachusetts, USA.
(3)Department of Pediatric Newborn Medicine, Brigham and Women's Hospital, 
Boston, Massachusetts, USA.
(4)Harvard Medical School, Boston, Massachusetts, USA.
(5)Department of Nutrition, Carolina Population Center, Gillings School of 
Public Health, University of North Carolina at Chapel Hill, Chapel Hill, North 
Carolina, USA.

Approximately 15% of infants worldwide are born with low birthweight (<2500 g). 
These children are at risk for growth failure. The aim of this umbrella review 
is to assess the relationship between infant milk type, fortification and growth 
in low-birthweight infants, with particular focus on low- and lower 
middle-income countries. We conducted a systematic review in PubMed, CINAHL, 
Embase and Web of Science comparing infant milk options and growth, grading the 
strength of evidence based on standard umbrella review criteria. Twenty-six 
systematic reviews qualified for inclusion. They predominantly focused on 
infants with very low birthweight (<1500 g) in high-income countries. We found 
the strongest evidence for (1) the addition of energy and protein fortification 
to human milk (donor or mother's milk) leading to increased weight gain (mean 
difference [MD] 1.81 g/kg/day; 95% confidence interval [CI] 1.23, 2.40), linear 
growth (MD 0.18 cm/week; 95% CI 0.10, 0.26) and head growth (MD 0.08 cm/week; 
95% CI 0.04, 0.12) and (2) formula compared with donor human milk leading to 
increased weight gain (MD 2.51 g/kg/day; 95% CI 1.93, 3.08), linear growth (MD 
1.21 mm/week; 95% CI 0.77, 1.65) and head growth (MD 0.85 mm/week; 95% CI 0.47, 
1.23). We also found evidence of improved growth when protein is added to both 
human milk and formula. Fat supplementation did not seem to affect growth. More 
research is needed for infants with birthweight 1500-2500 g in low- and lower 
middle-income countries.

© 2021 The Authors. Maternal & Child Nutrition published by John Wiley & Sons 
Ltd.

DOI: 10.1111/mcn.13176
PMCID: PMC8189224
PMID: 33733580 [Indexed for MEDLINE]

Conflict of interest statement: The authors declare that they have no conflicts 
of interest.


45. Cochrane Database Syst Rev. 2021 Mar 9;3(3):CD012413. doi: 
10.1002/14651858.CD012413.pub3.

High versus standard volume enteral feeds to promote growth in preterm or low 
birth weight infants.

Abiramalatha T(1), Thomas N(2), Thanigainathan S(3).

Author information:
(1)Neonatology, Sri Ramachandra Institute of Higher Education and Research, 
Chennai, India.
(2)Neonatology, Joan Kirner Women's and Children's at Sunshine Hospital, Western 
Health, St Albans, Australia.
(3)Neonatology, All India Institute of Medical Sciences, Jodhpur, India.

Update of
    Cochrane Database Syst Rev. 2017 Sep 12;9:CD012413. doi: 
10.1002/14651858.CD012413.pub2.

BACKGROUND: Human milk is the best enteral nutrition for preterm infants. 
However, human milk, given at standard recommended volumes, is not adequate to 
meet the protein, energy, and other nutrient requirements of preterm or low 
birth weight infants. One strategy that may be used to address the potential 
nutrient deficits is to give a higher volume of enteral feeds. High volume feeds 
may improve nutrient accretion and growth, and in turn may improve 
neurodevelopmental outcomes. However, there are concerns that high volume feeds 
may cause feed intolerance, necrotising enterocolitis, or complications related 
to fluid overload such as patent ductus arteriosus and chronic lung disease. 
This is an update of a review published in 2017.
OBJECTIVES: To assess the effect on growth and safety of high versus standard 
volume enteral feeds in preterm or low birth weight infants. In infants who were 
fed fortified human milk or preterm formula, high and standard volume feeds were 
defined as > 180 mL/kg/day and ≤ 180 mL/kg/day, respectively. In infants who 
were fed unfortified human milk or term formula, high and standard volume feeds 
were defined as > 200 mL/kg/day and ≤ 200 mL/kg/day, respectively.
SEARCH METHODS: We used the standard search strategy of Cochrane Neonatal to 
search Cochrane Central Register of Controlled Trials (CENTRAL; 2020 Issue 6) in 
the Cochrane Library; Ovid MEDLINE (1946 to June 2020); Embase (1974 to June 
2020); and CINAHL (inception to June 2020); Maternity & Infant Care Database 
(MIDIRS) (1971 to April 2020); as well as previous reviews, and trial 
registries.
SELECTION CRITERIA: We included randomised controlled trials (RCTs) that 
compared high versus standard volume enteral feeds for preterm or low birth 
weight infants.
DATA COLLECTION AND ANALYSIS: Two review authors assessed trial eligibility and 
risk of bias and independently extracted data. We analysed treatment effects in 
individual trials and reported risk ratio (RR) and risk difference for 
dichotomous data, and mean difference (MD) for continuous data, with respective 
95% confidence intervals (CIs). We used the GRADE approach to assess the 
certainty of evidence. The primary outcomes were weight gain, linear and head 
growth during hospital stay, and extrauterine growth restriction at discharge.
MAIN RESULTS: We included two new RCTs (283 infants) in this update. In total, 
we included three trials (347 infants) in this updated review. High versus 
standard volume feeds with fortified human milk or preterm formula Two trials 
(283 infants) met the inclusion criteria for this comparison. Both were of good 
methodological quality, except for lack of masking. Both trials were performed 
in infants born at < 32 weeks' gestation. Meta-analysis of data from both trials 
showed high volume feeds probably improves weight gain during hospital stay (MD 
2.58 g/kg/day, 95% CI 1.41 to 3.76; participants = 271; moderate-certainty 
evidence). High volume feeds may have little or no effect on linear growth (MD 
0.05 cm/week, 95% CI -0.02 to 0.13; participants = 271; low-certainty evidence), 
head growth (MD 0.02 cm/week, 95% CI -0.04 to 0.09; participants = 271; 
low-certainty evidence), and extrauterine growth restriction at discharge (RR 
0.71, 95% CI 0.50 to 1.02; participants = 271; low-certainty evidence). We are 
uncertain of the effect of high volume feeds with fortified human milk or 
preterm formula on the risk of necrotising enterocolitis (RR 0.74, 95% CI 0.12 
to 4.51; participants = 283; very-low certainty evidence). High versus standard 
volume feeds with unfortified human milk or term formula One trial with 64 very 
low birth weight infants met the inclusion criteria for this comparison. This 
trial was unmasked but otherwise of good methodological quality. High volume 
feeds probably improves weight gain during hospital stay (MD 6.2 g/kg/day, 95% 
CI 2.71 to 9.69; participants = 61; moderate-certainty evidence). The trial did 
not provide data on linear and head growth, and extrauterine growth restriction 
at discharge. We are uncertain as to the effect of high volume feeds with 
unfortified human milk or term formula on the risk of necrotising enterocolitis 
(RR 1.03, 95% CI 0.07 to 15.78; participants = 61; very low-certainty evidence).
AUTHORS' CONCLUSIONS: High volume feeds (≥ 180 mL/kg/day of fortified human milk 
or preterm formula, or ≥ 200 mL/kg/day of unfortified human milk or term 
formula) probably improves weight gain during hospital stay. The available data 
is inadequate to draw conclusions on the effect of high volume feeds on other 
growth and clinical outcomes. A large RCT is needed to provide data of 
sufficient quality and precision to inform policy and practice.

Copyright © 2021 The Cochrane Collaboration. Published by John Wiley & Sons, 
Ltd.

DOI: 10.1002/14651858.CD012413.pub3
PMCID: PMC8092452
PMID: 33733486 [Indexed for MEDLINE]

Conflict of interest statement: TA has no interest to declare. NT was the 
principal investigator in one study included in this review (Thomas 2012). 
However, TA performed the 'Risk of bias' assessment and data extraction for the 
trial. NT received no funding for Thomas 2012. ST has no interest to declare. 
Core editorial and administrative support for this review has been provided by a 
grant from The Gerber Foundation. The Gerber Foundation is a separately endowed, 
private foundation, independent from the Gerber Products Company. The grantor 
has no input on the content of the review or the editorial process (see Sources 
of support).


46. J Spec Oper Med. 2021 Spring;21(1):115-119. doi: 10.55460/ETA1-NLQP.

Vitamin A and Bone Fractures.

Knapik JJ, Hoedebecke SS.

Vitamin A is a generic term describing compounds that have the same biological 
activity as retinol. Dietary vitamin A can be obtained from "provitamin A" 
carotenoids (e.g., ß-carotene) found in plant foods such as carrots, 
cantaloupes, and sweet peppers, or as "preformed vitamin A" found in many 
dietary supplements, animal livers, and vitamin A-fortified foods, such as 
breakfast cereals, milk, cheese, and yogurt. Low consumption of vitamin A can 
cause night blindness, reduce immune function, and have detrimental 
developmental effects. Several lines of evidence suggest that excessive dietary 
intake of vitamin A might be associated with an increased risk of bone 
fractures. Meta-analysis of observational human studies that have examined 
vitamin A and fractures suggests that dietary consumption of large amounts of 
vitamin A in the form of ß-carotene likely has a protective effect, reducing the 
risk of fractures. On the other hand, meta-analyses that have specifically 
examined hip fractures have shown that total vitamin A (all types) or retinol 
consumption may increase the risk of hip fractures. Until more information is 
available, it is advisable to consume vitamin A primarily from plant sources, 
avoid excessive consumption from dietary supplements and animal sources, and 
lower consumption from fortified foods.

2021.

DOI: 10.55460/ETA1-NLQP
PMID: 33721319 [Indexed for MEDLINE]


47. Nutrients. 2021 Jan 22;13(2):316. doi: 10.3390/nu13020316.

Effect of Calcium Fortified Foods on Health Outcomes: A Systematic Review and 
Meta-Analysis.

Cormick G(1)(2)(3), Betran AP(4), Romero IB(3), Cormick MS(5), Belizán JM(1)(2), 
Bardach A(2)(6), Ciapponi A(2)(6).

Author information:
(1)Department of Mother and Child Health Research, Institute for Clinical 
Effectiveness and Health Policy (IECS-CONICET), Ciudad de Buenos Aires 1414, 
Argentina.
(2)Centro de Investigaciones Epidemiolóicas y Salud Púlica (CIESP-IECS). 
CONICET, Ciudad de Buenos Aires 1414, Argentina.
(3)Departament de Salud, Universidad Nacional de La Matanza (UNLAM), San Justo 
1903, Argentina.
(4)UNDP/UNFPA/UNICEF/WHO/World Bank Special Programme of Research, Development 
and Research Training in Human Reproduction (HRP), Department of Sexual and 
Reproductive Health and Research, World Health Organization, 1211 Geneva 27, 
Switzerland.
(5)Departament de Diagnóstico por Imágenes, Fleni, Montañeses 2325, Ciudad de 
Buenos Aires C1428, Argentina.
(6)Centro Cochrane Argentino-Instituto de Efectividad Clínica y Sanitaria 
(IECS-CONICET), Ciudad de Buenos Aires 1414, Argentina.

Calcium supplementation and fortification are strategies widely used to prevent 
adverse outcome in population with low-calcium intake which is highly frequent 
in low-income settings. We aimed to determine the effectiveness and 
cost-effectiveness of calcium fortified foods on calcium intake and related 
health, or economic outcomes. We performed a systematic review and meta-analysis 
involving participants of any age or gender, drawn from the general population. 
We searched PubMed, Agricola, EMBASE, CINAHL, Global Health, EconLit, the FAO 
website and Google until June 2019, without language restrictions. Pair of 
reviewers independently selected, extracted data and assessed the risk of bias 
of included studies using Covidence software. Disagreements were resolved by 
consensus. We performed meta-analyses using RevMan 5.4 and subgroup analyses by 
study design, age group, and fortification levels. We included 20 studies of 
which 15 were randomized controlled trials (RCTs), three were non-randomised 
studies and two were economic evaluations. Most RCTs had high risk of bias on 
randomization or blinding. Most represented groups were women and children from 
1 to 72 months, most common intervention vehicles were milk and bakery products 
with a fortification levels between 96 and 1200 mg per 100 g of food. Calcium 
intake increased in the intervention groups between 460 mg (children) and 1200 
mg (postmenopausal women). Most marked effects were seen in children. Compared 
to controls, height increased 0.83 cm (95% CI 0.00; 1.65), plasma parathyroid 
hormone decreased -1.51 pmol/L, (-2.37; -0.65), urine:calcium creatinine ratio 
decreased -0.05, (-0.07; -0.03), femoral neck and hip bone mineral density 
increased 0.02 g/cm2 (0.01; 0.04) and 0.03 g/cm2 (0.00; 0.06), respectively. The 
largest cost savings (43%) reported from calcium fortification programs came 
from prevented hip fractures in older women from Germany. Our study highlights 
that calcium fortification leads to a higher calcium intake, small benefits in 
children's height and bone health and also important evidence gaps for other 
outcomes and populations that could be solved with high quality experimental or 
quasi-experimental studies in relevant groups, especially as some evidence of 
calcium supplementation show controversial results on the bone health benefit on 
older adults.

DOI: 10.3390/nu13020316
PMCID: PMC7911363
PMID: 33499250 [Indexed for MEDLINE]

Conflict of interest statement: The authors declare no conflict of interest.


48. Indian Pediatr. 2021 Mar 15;58(3):253-258. Epub 2021 Jan 2.

Fortification of Human Milk With Infant Formula for Very Low Birth Weight 
Preterm Infants: A Systematic Review.

Kumar M(1), Upadhyay J(2), Basu S(3).

Author information:
(1)Department of Pediatrics, All India Institute of Medical Sciences, Rishikesh, 
Uttarakhand, India.
(2)Department of Neonatology, All India Institute of Medical Sciences, 
Rishikesh, Uttarakhand, India.
(3)Department of Neonatology, All India Institute of Medical Sciences, 
Rishikesh, Uttarakhand, India. Correspondence to: Sriparna Basu, Department of 
Neonatology, All India Institute of Medical Sciences, Rishikesh, Uttarakhand 
-249203, India..

BACKGROUND: Off-label fortification of expressed human milk (HM) with infant 
milk formula (IMF) is common in developing countries, though its benefits and 
safety are unclear.
OBJECTIVE: To study the effects of IMF fortification of HM on growth of very low 
birth weight (VLBW) preterm infants.
DESIGN: Systematic review and meta-analysis of randomized and quasi-randomized 
controlled trials (RCTs).
DATA SOURCES AND SELECTION CRITERIA: MEDLINE, EMBASE, CINAHL, CENTRAL and other 
databases were searched for articles published in English language from 
inception to December 2019, evaluating the effects of HM fortified with IMF as 
intervention, compared to unfortified HM or HM fortified with human milk 
fortifier (HMF).
PARTICIPANTS: Five RCTs including 423 VLBW preterm infants.
INTERVENTION: Feeding with HM fortified with IMF compared to unfortified or 
HMF-fortified HM.
OUTCOME MEASURES: Primary outcome measure was assessment of growth as weight, 
length and head circumference (HC) gain velocity. Secondary outcome measures 
were incidences of feed intolerance (FI), necrotizing enterocolitis (NEC), time 
to reach full feeds, concentration of nutritional biomarkers, duration of 
hospital-stay and cost of intervention.
RESULTS: Of the five studies included in the review, pooled effects regarding 
weight gain velocity (SMD 0.27 g/day; 95% CI 0.08 to 0.62), length gain (MD 
0.07cm/week; 95% CI 0.02 to 0.16) and HC gain (MD 0.05 cm/wk; 95% CI 0.01 to 
0.11), were not statistically significant. Sensitivity analysis by pooling 
studies using unfortified milk as comparator yielded a statistically significant 
result for all growth parameters. Risk of FI or NEC was comparable. Length of 
hospitalstay was reduced in the intervention group.
CONCLUSIONS: A very-low quality evidence suggested that IMF fortification of HM 
is superior to unfortified milk and may be a safe alternative for HMF for short 
term growth of VLBW preterm infants.

PMID: 33408285 [Indexed for MEDLINE]


49. Cochrane Database Syst Rev. 2020 Nov 23;11(11):CD013465. doi: 
10.1002/14651858.CD013465.pub2.

Individualized versus standard diet fortification for growth and development in 
preterm infants receiving human milk.

Fabrizio V(1)(2), Trzaski JM(1)(2), Brownell EA(3)(4), Esposito P(1), Lainwala 
S(1)(2), Lussier MM(1), Hagadorn JI(1)(2).

Author information:
(1)Division of Neonatology, Connecticut Children's, Hartford, CT, USA.
(2)Department of Pediatrics, University of Connecticut School of Medicine, 
Farmington, CT, USA.
(3)School of Nursing, University of Texas Health Science Center, San Antonio, 
TX, USA.
(4)Department of Pediatrics, University of Texas Health Science Center, San 
Antonio, Texas, USA.

Comment in
    Dev Med Child Neurol. 2021 Dec;63(12):1371-1373. doi: 10.1111/dmcn.15057.

BACKGROUND: Human milk as compared to formula reduces morbidity in preterm 
infants but requires fortification to meet their nutritional needs and to reduce 
the risk of extrauterine growth failure. Standard fortification methods are not 
individualized to the infant and assume that breast milk is uniform in 
nutritional content. Strategies for individualizing fortification are available; 
however it is not known whether these are safe, or if they improve outcomes in 
preterm infants.
OBJECTIVES: To determine whether individualizing fortification of breast milk 
feeds in response to infant blood urea nitrogen (adjustable fortification) or to 
breast milk macronutrient content as measured with a milk analyzer (targeted 
fortification) reduces mortality and morbidity and promotes growth and 
development compared to standard, non-individualized fortification for preterm 
infants receiving human milk at < 37 weeks' gestation or at birth weight < 2500 
grams.
SEARCH METHODS: We used the standard search strategy of Cochrane Neonatal to 
search the Cochrane Central Register of Controlled Trials (CENTRAL; 2019, Issue 
9), in the Cochrane Library; Ovid MEDLINE(R) and Epub Ahead of Print, In-Process 
& Other Non-Indexed Citations, Daily and Versions(R); and the Cumulative Index 
to Nursing and Allied Health Literature (CINAHL), on September 20, 2019. We also 
searched clinical trials databases and the reference lists of retrieved articles 
for pertinent randomized controlled trials (RCTs) and quasi-randomized trials.
SELECTION CRITERIA: We considered randomized, quasi-randomized, and 
cluster-randomized controlled trials of preterm infants fed exclusively breast 
milk that compared a standard non-individualized fortification strategy to 
individualized fortification using a targeted or adjustable strategy. We 
considered studies that examined any use of fortification in eligible infants 
for a minimum duration of two weeks, initiated at any time during enteral 
feeding, and providing any regimen of human milk feeding.
DATA COLLECTION AND ANALYSIS: Data were collected using the standard methods of 
Cochrane Neonatal. Two review authors evaluated the quality of the studies and 
extracted data. We reported analyses of continuous data using mean differences 
(MDs), and dichotomous data using risk ratios (RRs). We used the GRADE approach 
to assess the certainty of evidence.
MAIN RESULTS: Data were extracted from seven RCTs, resulting in eight 
publications (521 total participants were enrolled among these studies), with 
duration of study interventions ranging from two to seven weeks. As compared to 
standard non-individualized fortification, individualized (targeted or 
adjustable) fortification of enteral feeds probably increased weight gain during 
the intervention (typical mean difference [MD] 1.88 g/kg/d, 95% confidence 
interval [CI] 1.26 to 2.50; 6 studies, 345 participants), may have increased 
length gain during the intervention (typical MD 0.43 mm/d, 95% CI 0.32 to 0.53; 
5 studies, 242 participants), and may have increased head circumference gain 
during the intervention (typical MD 0.14 mm/d, 95% CI 0.06 to 0.23; 5 studies, 
242 participants). Compared to standard non-individualized fortification, 
targeted fortification probably increased weight gain during the intervention 
(typical MD 1.87 g/kg/d, 95% CI 1.15 to 2.58; 4 studies, 269 participants) and 
may have increased length gain during the intervention (typical MD 0.45 mm/d, 
95% CI 0.32 to 0.57; 3 studies, 166 participants). Adjustable fortification 
probably increased weight gain during the intervention (typical MD 2.86 g/kg/d, 
95% CI 1.69 to 4.03; 3 studies, 96 participants), probably increased gain in 
length during the intervention (typical MD 0.54 mm/d, 95% CI 0.38 to 0.7; 3 
studies, 96 participants), and increased gain in head circumference during the 
intervention (typical MD 0.36 mm/d, 95% CI 0.21 to 0.5; 3 studies, 96 
participants). We are uncertain whether there are differences between 
individualized versus standard fortification strategies in the incidence of 
in-hospital mortality, bronchopulmonary dysplasia, necrotizing enterocolitis, 
culture-proven late-onset bacterial sepsis, retinopathy of prematurity, 
osteopenia, length of hospital stay, or post-hospital discharge growth. No study 
reported severe neurodevelopmental disability as an outcome. One study that was 
published after our literature search was completed is awaiting classification.
AUTHORS' CONCLUSIONS: We found moderate- to low-certainty evidence suggesting 
that individualized (either targeted or adjustable) fortification of enteral 
feeds in very low birth weight infants increases growth velocity of weight, 
length, and head circumference during the intervention compared with standard 
non-individualized fortification. Evidence showing important in-hospital and 
post-discharge clinical outcomes was sparse and of very low certainty, 
precluding inferences regarding safety or clinical benefits beyond short-term 
growth.

Copyright © 2020 The Cochrane Collaboration. Published by John Wiley & Sons, 
Ltd.

DOI: 10.1002/14651858.CD013465.pub2
PMCID: PMC8094236
PMID: 33226632 [Indexed for MEDLINE]

Conflict of interest statement: VF on the Mothers' Milk Bank Northeast Advisory 
Board in a voluntary capacity. This is a non‐profit community milk bank that 
provides donated, pasteurized human milk. JMT has no interests to declare. EAB 
has no interests to declare. PE has no interests to declare. SL has no interests 
to declare. MML has no interests to declare. JIH has no interests to declare. 
Core editorial and administrative support for this review has been provided by a 
grant from The Gerber Foundation. The Gerber Foundation is a separately endowed, 
private foundation, independent from the Gerber Products Company. The grantor 
has no input on the content of the review or the editorial process (see Sources 
of support).


50. Cochrane Database Syst Rev. 2020 Nov 20;11(11):CD007090. doi: 
10.1002/14651858.CD007090.pub2.

Comparison of different protein concentrations of human milk fortifier for 
promoting growth and neurological development in preterm infants.

Gao C(1), Miller J(1), Collins CT(1)(2), Rumbold AR(1).

Author information:
(1)SAHMRI Women and Kids, South Australian Health and Medical Research 
Institute, Adelaide, Australia.
(2)Discipline of Paediatrics, Adelaide Medical School, The University of 
Adelaide, Adelaide, Australia.

Update of
    doi: 10.1002/14651858.CD007090.

BACKGROUND: Human milk alone may provide inadequate amounts of protein to meet 
the growth requirements of preterm infants because of restrictions in the amount 
of fluid they can tolerate. It has become common practice to feed preterm 
infants with breast milk fortified with protein and other nutrients but there is 
debate about the optimal concentration of protein in commercially available 
fortifiers.
OBJECTIVES: To compare the effects of different protein concentrations in human 
milk fortifier, fed to preterm infants, on growth and neurodevelopment.
SEARCH METHODS: We used the standard search strategy of Cochrane Neonatal to 
search CENTRAL (2019, Issue 8), Ovid MEDLINE and CINAHL on 15 August 2019. We 
also searched clinical trials databases and the reference lists of retrieved 
articles for randomised controlled trials and quasi-randomised trials.
SELECTION CRITERIA: We included all published and unpublished randomised, 
quasi-randomised and cluster-randomised trials comparing two different 
concentrations of protein in human milk fortifier. We included preterm infants 
(less than 37 weeks' gestational age). Participants may have been exclusively 
fed human milk or have been supplemented with formula. The concentration of 
protein was classified as low (< 1g protein/100 mL expressed breast milk (EBM)), 
moderate (≥ 1g to < 1.4g protein/100 mL EBM) or high (≥ 1.4g protein/100 mL 
EBM). We excluded trials that compared two protein concentrations that fell 
within the same category.
DATA COLLECTION AND ANALYSIS: We undertook data collection and analyses using 
the standard methods of Cochrane Neonatal. Two review authors independently 
evaluated trials. Primary outcomes included growth, neurodevelopmental outcome 
and mortality. Data were synthesised using risk ratios (RR), risk differences 
and mean differences (MD), with 95% confidence intervals (CI). We used the GRADE 
approach to assess the certainty of the evidence.
MAIN RESULTS: We identified nine trials involving 861 infants. There is one 
trial awaiting classification, and nine ongoing trials. The trials were mostly 
conducted in infants born < 32 weeks' gestational age or < 1500 g birthweight, 
or both. All used a fortifier derived from bovine milk. Two trials fed infants 
exclusively with mother's own milk, three trials gave supplementary feeds with 
donor human milk and four trials supplemented with preterm infant formula. 
Overall, trials were small but generally at low or unclear risk of bias. High 
versus moderate protein concentration of human milk fortifier There was moderate 
certainty evidence that a high protein concentration likely increased 
in-hospital weight gain compared to moderate concentration of human milk 
fortifier (MD 0.66 g/kg/day, 95% CI 0.51 to 0.82; trials = 6, participants = 
606). The evidence was very uncertain about the effect of high versus moderate 
protein concentration on length gain (MD 0.01 cm/week, 95% CI -0.01 to 0.03; 
trials = 5, participants = 547; very low certainty evidence) and head 
circumference gain (MD 0.00 cm/week, 95% CI -0.01 to 0.02; trials = 5, 
participants = 549; very low certainty evidence). Only one trial reported 
neonatal mortality, with no deaths in either group (participants = 45). Moderate 
versus low protein concentration of human milk fortifier A moderate versus low 
protein concentration fortifier may increase weight gain (MD 2.08 g/kg/day, 95% 
CI 0.38 to 3.77; trials = 2, participants = 176; very low certainty evidence) 
with little to no effect on head circumference gain (MD 0.13 cm/week, 95% CI 
0.00 to 0.26; I² = 85%; trials = 3, participants = 217; very low certainty 
evidence), but the evidence is very uncertain. There was low certainty evidence 
that a moderate protein concentration may increase length gain (MD 0.09 cm/week, 
95% CI 0.05 to 0.14; trials = 3, participants = 217). Only one trial reported 
mortality and found no difference between groups (RR 0.48, 95% CI 0.05 to 5.17; 
participants = 112). No trials reported long term growth or neurodevelopmental 
outcomes including cerebral palsy and developmental delay.
AUTHORS' CONCLUSIONS: Feeding preterm infants with a human milk fortifier 
containing high amounts of protein (≥ 1.4g/100 mL EBM) compared with a fortifier 
containing moderate protein concentration (≥ 1 g to < 1.4 g/100 mL EBM) results 
in small increases in weight gain during the neonatal admission. There may also 
be small increases in weight and length gain when infants are fed a fortifier 
containing moderate versus low protein concentration (< 1 g protein/100 mL EBM). 
The certainty of this evidence is very low to moderate; therefore, results may 
change when the findings of ongoing studies are available. There is insufficient 
evidence to assess the impact of protein concentration on adverse effects or 
long term outcomes such as neurodevelopment. Further trials are needed to 
determine whether modest increases in weight gain observed with higher protein 
concentration fortifiers are associated with benefits or harms to long term 
growth and neurodevelopment.

Copyright © 2020 The Cochrane Collaboration. Published by John Wiley & Sons, 
Ltd.

DOI: 10.1002/14651858.CD007090.pub2
PMCID: PMC8092673
PMID: 33215474 [Indexed for MEDLINE]

Conflict of interest statement: CG received an Adelaide Graduate Research 
Scholarship from the University of Adelaide to support her PhD. JM is an author 
of the following included studies: Miller 2012; Reid 2018. CTC is an author of 
the following included studies: Miller 2012; Reid 2018. CTC is supported by a 
National Health and Medical Research Council (NHMRC) Translating Research into 
Practice Fellowship (APP1132596). The views expressed in this article are solely 
the responsibility of the authors and do not reflect the views of the NHMRC. JM 
and CTC declared the following conflicts of interest: Nestle Nutrition donated 
half of the human milk fortifier, and Nutricia donated Polyjoule and Protifar, 
used in a randomised controlled trial (RCT) of increased protein concentration 
human milk fortifier (Reid 2018). Study product for a separate RCT of human milk 
fortifier was manufactured and donated by Nestle Product Technology Centre, 
Konolfingen, Switzerland (Miller 2012). The sponsors had no role (in either 
Miller 2012 or Reid 2018), in the design of the study, in the collection, 
analyses or interpretation of data, in writing of the manuscript, or the 
decision to publish the results. Data extraction of these two studies (Miller 
2012; Reid 2018) was completed by CG. AR has no interests to declare. Core 
editorial and administrative support for this review was provided by a grant 
from The Gerber Foundation. The Gerber Foundation is a separately endowed, 
private foundation, independent from the Gerber Products Company. The grantor 
had no input on the content of the review or the editorial process. The review 
authors did not receive any direct funding from The Gerber Foundation (Sources 
of support).


51. Cochrane Database Syst Rev. 2020 Sep 23;9(9):CD000433. doi: 
10.1002/14651858.CD000433.pub3.

Protein supplementation of human milk for promoting growth in preterm infants.

Amissah EA(1), Brown J(2), Harding JE(1).

Author information:
(1)Liggins Institute, University of Auckland, Auckland, New Zealand.
(2)Department of Obstetrics and Gynaecology, The University of Auckland, 
Auckland, New Zealand.

Update of
    Cochrane Database Syst Rev. 2018 Jun 22;6:CD000433. doi: 
10.1002/14651858.CD000433.pub2.

BACKGROUND: Preterm infants require high protein intake to achieve adequate 
growth and development. Although breast milk feeding has many benefits for this 
population, the protein content is highly variable, and inadequate to support 
rapid infant growth. This is a 2020 update of a Cochrane Review first published 
in 1999.
OBJECTIVES: To determine whether protein-supplemented human milk compared with 
unsupplemented human milk, fed to preterm infants, improves growth, body 
composition, cardio-metabolic, and neurodevelopmental outcomes, without 
significant adverse effects.
SEARCH METHODS: We used the standard search strategy of Cochrane Neonatal to 
search Cochrane Central Register of Controlled Trials (CENTRAL 2019, Issue 8) in 
the Cochrane Library and MEDLINE via PubMed on 23 August 2019. We also searched 
clinical trials databases and the reference lists of retrieved articles for 
randomised controlled trials and quasi-randomised trials.
SELECTION CRITERIA: Published and unpublished RCTs were eligible if they used 
random or quasi-random methods to allocate hospitalised preterm infants who were 
being fed human milk, to additional protein supplementation or no 
supplementation.
DATA COLLECTION AND ANALYSIS: Two review authors independently abstracted data, 
assessed risk of bias and the quality of evidence at the outcome level, using 
GRADE methodology. We performed meta-analyses, using risk ratio (RR) for 
dichotomous data, and mean difference (MD) for continuous data, with their 
respective 95% confidence intervals (CIs). We used a fixed-effect model and had 
planned to explore potential causes of heterogeneity via subgroup or sensitivity 
analyses.
MAIN RESULTS: We included six RCTs, involving 204 preterm infants. The risk of 
bias for most methodological domains was unclear as there was insufficient 
detail reported. Low-quality evidence showed that protein supplementation of 
human milk may increase in-hospital rates of growth in weight (MD 3.82 g/kg/day, 
95% CI 2.94 to 4.7; five RCTs, 101 infants; I² = 73%), length (MD 0.12 cm/wk, 
95% CI 0.07 to 0.17; four RCTs, 68 infants; I² = 89%), and head circumference 
(MD 0.06 cm/wk, 95% CI 0.01 to 0.12; four RCTs, 68 infants; I² = 84%). Protein 
supplementation may lead to longer hospital stays (MD 18.5 days, 95% CI 4.39 to 
32.61; one RCT, 20 infants; very low-quality evidence). Very low quality 
evidence means that the effect of protein supplementation on the risk of feeding 
intolerance (RR 2.70, 95% CI 0.13 to 58.24; one RCT, 17 infants), or necrotizing 
enterocolitis (RR 1.11, 95% CI 0.07 to 17.12; one RCT, 76 infants) remains 
uncertain. No data were available about the effects of protein supplementation 
on neurodevelopmental outcomes.
AUTHORS' CONCLUSIONS: Low-quality evidence showed that protein supplementation 
of human milk, fed to preterm infants, increased short-term growth. However, the 
small sample sizes, low precision, and very low-quality evidence regarding 
duration of hospital stay, feeding intolerance, and necrotising enterocolitis 
precluded any conclusions about these outcomes. There were no data on outcomes 
after hospital discharge. Our findings may not be generalisable to low-resource 
settings, as none of the included studies were conducted in these settings. 
Since protein supplementation of human milk is now usually done as a component 
of multi-nutrient fortifiers, future studies should compare different amounts of 
protein in multi-component fortifiers, and be designed to determine the effects 
on duration of hospital stay and safety, as well as on long-term growth, body 
composition, cardio-metabolic, and neurodevelopmental outcomes.

Copyright © 2020 The Cochrane Collaboration. Published by John Wiley & Sons, 
Ltd.

DOI: 10.1002/14651858.CD000433.pub3
PMCID: PMC8094919
PMID: 32964431 [Indexed for MEDLINE]

Conflict of interest statement: EA receives a scholarship in the form of a 
stipend from the University of Auckland as a PhD student. JB is currently 
employed by a medical writing company. The preparation of this review took place 
prior to this employment and her current work is not related to the topic of 
this review. JH has partial salary support from research grants from the Health 
Research Council of New Zealand. The Council has no role in the production of 
this review. An undirected research grant is pending from Biomed Ltd, Auckland, 
New Zealand. This company makes dextrose gel. The methods section of this review 
is based on a standard template used by Cochrane Neonatal. Core editorial and 
administrative support for the 2020 update of this review has been provided by a 
grant from The Gerber Foundation. The Gerber Foundation is a separately endowed, 
private foundation, independent from the Gerber Products Company. The grantor 
has no input on the content of the review or the editorial process.


52. Arch Dis Child Fetal Neonatal Ed. 2021 Mar;106(2):137-142. doi: 
10.1136/archdischild-2020-319406. Epub 2020 Sep 17.

Safety and efficacy of human milk-based fortifier in enterally fed preterm 
and/or low birthweight infants: a systematic review and meta-analysis.

Grace E(1)(2)(3), Hilditch C(3)(4), Gomersall J(4)(5), Collins CT(3)(4), Rumbold 
A(4)(6), Keir AK(7)(3)(4)(6).

Author information:
(1)Department of Neonatal Medicine, Women's and Children's Hospital, North 
Adelaide, South Australia, Australia.
(2)Women and Kids Theme, South Australian Health and Medical Research Institute, 
Adelaide, South Australia, Australia.
(3)Adelaide Medical School, University of Adelaide, Adelaide, South Australia, 
Australia.
(4)SAHMRI Women and Kids, South Australian Health and Medical Research 
Institute, North Adelaide, South Australia, Australia.
(5)School of Public Health, University of Adelaide, Adelaide, South Australia, 
Australia.
(6)Robinson Research Institute, University of Adelaide, Adelaide, South 
Australia, Australia.
(7)Department of Neonatal Medicine, Women's and Children's Hospital, North 
Adelaide, South Australia, Australia.

OBJECTIVE: To conduct a systematic review and meta-analysis of the efficacy and 
safety of fortification of human milk with human milk-based fortifier versus 
cow's milk-based fortifier for use in preterm and/or very low birthweight 
infants.
DESIGN: Randomised or quasi-randomised controlled trials comparing the effect of 
human milk fortification with human milk-based milk fortifier versus cow's 
milk-based fortifier in infants born <34 weeks' gestation and/or with birth 
weight <1500 g were identified by searching databases, clinical trial registries 
and reference lists until 5 November 2019. Two authors independently extracted 
data and assessed evidence quality. Meta-analyses were conducted using fixed or 
random effects models, as appropriate.
MAIN OUTCOME MEASURES: Necrotising enterocolitis (Bell's stage II or higher) and 
late-onset sepsis.
RESULTS: Of 863 unique records identified, 16 full-text trials were screened and 
2 trials involving 334 infants were included. Primary outcome data were 
available for 332 infants. Use of human milk-based fortifier compared with cow's 
milk-based fortifier reduced the risk of necrotising enterocolitis (risk ratio 
0.47, 95% CI 0.22 to 0.98). There was no clear evidence of an effect on 
late-onset sepsis or any other outcomes. The quality of evidence was low to very 
low due to imprecision and lack of blinding in one study.
CONCLUSIONS: Findings suggest that there is a reduction in the incidence of 
necrotising enterocolitis with human milk-based fortifiers compared with cow's 
milk-based fortifiers. The overall quality of evidence is low. Further 
appropriately powered trials are required before this intervention can be 
routinely recommended for preterm infants.

© Author(s) (or their employer(s)) 2021. No commercial re-use. See rights and 
permissions. Published by BMJ.

DOI: 10.1136/archdischild-2020-319406
PMID: 32943531 [Indexed for MEDLINE]

Conflict of interest statement: Competing interests: None declared.


53. Cochrane Database Syst Rev. 2020 Sep 8;9(9):CD000280. doi: 
10.1002/14651858.CD000280.pub3.

Carbohydrate supplementation of human milk to promote growth in preterm infants.

Amissah EA(1), Brown J(2), Harding JE(1).

Author information:
(1)Liggins Institute, University of Auckland, Auckland, New Zealand.
(2)Department of Obstetrics and Gynaecology, The University of Auckland, 
Auckland, New Zealand.

Update of
    Cochrane Database Syst Rev. 2018 Aug 23;8:CD000280. doi: 
10.1002/14651858.CD000280.pub2.

BACKGROUND: Preterm infants are born with low glycogen stores and require higher 
glucose intake to match fetal accretion rates. In spite of the myriad benefits 
of breast milk for preterm infants, it may not adequately meet the needs of 
these rapidly growing infants. Supplementing human milk with carbohydrates may 
help. However, there is a paucity of data on assessment of benefits or harms of 
carbohydrate supplementation of human milk to promote growth in preterm infants. 
This is a 2020 update of a Cochrane Review first published in 1999.
OBJECTIVES: To determine whether human milk supplemented with carbohydrate 
compared with unsupplemented human milk fed to preterm infants improves growth, 
body composition, and cardio-metabolic and neurodevelopmental outcomes without 
significant adverse effects.
SEARCH METHODS: We used the standard search strategy of Cochrane Neonatal to 
search Cochrane Central Register of Controlled Trials (CENTRAL 2019, Issue 8) in 
the Cochrane Library and MEDLINE via PubMed on 22 August 2019. We also searched 
clinical trials databases and the reference lists of retrieved articles for 
randomised controlled trials and quasi-randomised trials.
SELECTION CRITERIA: Published and unpublished controlled trials were eligible if 
they used random or quasi-random methods to allocate preterm infants in hospital 
fed human milk to supplementation or no supplementation with additional 
carbohydrate.
DATA COLLECTION AND ANALYSIS: Two review authors independently abstracted data 
and assessed trial quality and the quality of evidence at the outcome level 
using the Grading of Recommendations Assessment, Development and Evaluation 
(GRADE) method. We planned to perform meta-analyses using risk ratios (RRs) for 
dichotomous data and mean differences (MDs) for continuous data, with their 
respective 95% confidence intervals (CIs). We planned to use a fixed-effect 
model and to explore potential causes of heterogeneity via sensitivity analyses. 
We contacted study authors for additional information.
MAIN RESULTS: One unblinded, quasi-randomised controlled trial (RCT) assessing 
effects of carbohydrate supplementation of human milk in the form of a prebiotic 
in 75 preterm infants was eligible for inclusion in this review. We identified 
two publications of the same trial, which reported different methods regarding 
blinding and randomisation. Study authors confirmed that these publications 
pertain to the same trial, but they have not yet clarified which method is 
correct. We were unable to reproduce analyses from the data presented. At 30 
days of age, the mean weight of preterm infants in the trial was greater in the 
prebiotic carbohydrate-supplemented group than in the unsupplemented group (MD 
160.4 grams, 95% CI 12.4 to 308.4 grams; one RCT, N = 75; very low-quality 
evidence). We found no evidence of a clear difference in risk of feeding 
intolerance (RR 0.64, 95% CI 0.36 to 1.15; one RCT, N = 75 infants; very 
low-quality evidence) or necrotising enterocolitis (NEC) (RR 0.2, 95% CI 0.02 to 
1.3; one RCT, N = 75 infants; very low-quality evidence) between the 
prebiotic-supplemented group and the unsupplemented group. Duration of hospital 
stay was shorter in the prebiotic group than in the control group at a median 
(range) of 16 (9 to 45) days (95% CI 15.34 to 24.09) and 25 (11 to 80) days (95% 
CI 25.52 to 34.39), respectively. No other data were available for assessing 
effects of carbohydrate supplementation on short- and long-term growth, body 
mass index, body composition, and neurodevelopmental or cardio-metabolic 
outcomes.
AUTHORS' CONCLUSIONS: We found insufficient evidence on the short- and long-term 
effects of carbohydrate supplementation of human milk in preterm infants. The 
only trial included in this review presented very low-quality evidence, and 
study authors provided uncertain information about study methods and analysis. 
The evidence may be limited in its applicability because researchers included a 
small sample of preterm infants from a single centre. However, the outcomes 
assessed are common to all preterm infants, and this trial demonstrates the 
feasibility of prebiotic carbohydrate supplementation in upper-middle-income 
countries. Future trials should assess the safety and efficacy of different 
types and concentrations of carbohydrate supplementation for preterm infants fed 
human milk. Although prebiotic carbohydrate supplementation in preterm infants 
is currently a topic of active research, we do not envisage that further trials 
of digestible carbohydrates will be conducted, as this is currently done as a 
component of multi-nutrient human milk fortification. Hence we do not plan to 
publish any further updates of this review.

Copyright © 2020 The Cochrane Collaboration. Published by John Wiley & Sons, 
Ltd.

DOI: 10.1002/14651858.CD000280.pub3
PMCID: PMC8094174
PMID: 32898300 [Indexed for MEDLINE]

Conflict of interest statement: EA receives a scholarship in the form of a 
stipend from the University of Auckland as a PhD student. JB is currently 
employed by a medical writing company. The preparation of this review took place 
prior to this employment and her current work is not related to the topic of 
this review. JH has partial salary support from research grants from the Health 
Research Council of New Zealand. The Council has no role in the production of 
this review. An undirected research grant is pending from Biomed Ltd, Auckland, 
New Zealand. This company makes dextrose gel. Core editorial and administrative 
support for the 2020 update of this review has been provided by a grant from The 
Gerber Foundation. The Gerber Foundation is a separately endowed, private 
foundation, independent from the Gerber Products Company. The grantor has no 
input on the content of the review or the editorial process (see Sources of 
support).


54. Cochrane Database Syst Rev. 2020 Aug 25;8(8):CD000341. doi: 
10.1002/14651858.CD000341.pub3.

Fat supplementation of human milk for promoting growth in preterm infants.

Amissah EA(1), Brown J(2), Harding JE(1).

Author information:
(1)Liggins Institute, University of Auckland, Auckland, New Zealand.
(2)Department of Obstetrics and Gynaecology, The University of Auckland, 
Auckland, New Zealand.

Update of
    Cochrane Database Syst Rev. 2018 Jun 19;6:CD000341. doi: 
10.1002/14651858.CD000341.pub2.

BACKGROUND: As preterm infants do not experience the nutrient accretion and 
rapid growth phase of the third trimester of pregnancy, they are vulnerable to 
postnatal nutritional deficits, including of fat. Consequently, they require 
higher fat intakes compared to their full term counterparts to achieve adequate 
growth and development. Human milk fat provides the major energy needs of the 
preterm infant and also contributes to several metabolic and physiological 
functions. Although human milk has many benefits for this population, its fat 
content is highly variable and may be inadequate for their optimum growth and 
development. This is a 2020 update of a Cochrane Review last published in 2000.
OBJECTIVES: To determine whether supplementation of human milk with fat compared 
with unsupplemented human milk fed to preterm infants improves growth, body 
composition, cardio-metabolic, and neurodevelopmental outcomes without 
significant adverse effects.
SEARCH METHODS: We used the standard search strategy of Cochrane Neonatal to 
search Cochrane Central Register of Controlled Trials (CENTRAL 2019, Issue 8) in 
the Cochrane Library and MEDLINE via PubMed on 23 August 2019. We also searched 
clinical trials databases and the reference lists of retrieved articles for 
randomised controlled trials and quasi-randomised trials.
SELECTION CRITERIA: Published and unpublished randomised controlled trials were 
eligible if they used random or quasi-random methods to allocate preterm infants 
fed human milk in hospital to supplementation or no supplementation with 
additional fat.
DATA COLLECTION AND ANALYSIS: No new randomised controlled trials matching the 
selection criteria were found but we extracted data from the previously included 
trial due to changes in review outcomes from when the protocol was first 
published. Two reviewers independently abstracted data, assessed trial quality, 
and the quality of evidence at the outcome level using the Grading of 
Recommendations Assessment, Development and Evaluation (GRADE) criteria. We 
planned to perform meta-analyses using risk ratio (RR) for dichotomous data and 
mean difference (MD) for continuous data, with their respective 95% confidence 
intervals (CIs). We planned to use a fixed-effect model and to explore potential 
causes of heterogeneity via sensitivity analyses.
MAIN RESULTS: One randomised trial involving 14 preterm infants was included. 
There was no evidence of a clear difference between the fat-supplemented and 
unsupplemented groups in in-hospital rates of growth in weight (MD 0.6 g/kg/day, 
95% CI -2.4 to 3.6; 1 RCT, n = 14 infants, very low-quality evidence), length 
(MD 0.1 cm/week, 95% CI -0.08 to 0.3; 1 RCT, n = 14 infants, very low-quality 
evidence) and head circumference (MD 0.2 cm/week, 95% CI -0.07 to 0.4; 1 RCT n = 
14 infants, very low-quality evidence). There was no clear evidence that fat 
supplementation increased the risk of feeding intolerance (RR 3.0, 95% CI 0.1 to 
64.3; 1 RCT, n = 16 infants, very low-quality evidence). No data were available 
regarding the effects of fat supplementation on long-term growth, body mass 
index, body composition, neurodevelopmental, or cardio-metabolic outcomes.
AUTHORS' CONCLUSIONS: The one included trial suggests no evidence of an effect 
of fat supplementation of human milk on short-term growth and feeding 
intolerance in preterm infants. However, the very low-quality evidence, small 
sample size, few events, and low precision diminishes our confidence that these 
results reflect the true effect of fat supplementation of human milk in preterm 
infants, and no long-term outcomes were reported. Further high-quality research 
should evaluate the effect on short and long-term growth, neurodevelopmental and 
cardio-metabolic outcomes in the context of the development of multicomponent 
fortifiers. Optimal dosage, adverse effects, and delivery practices should also 
be evaluated.

Copyright © 2020 The Cochrane Collaboration. Published by John Wiley & Sons, 
Ltd.

DOI: 10.1002/14651858.CD000341.pub3
PMCID: PMC8236752
PMID: 32842164 [Indexed for MEDLINE]

Conflict of interest statement: EA receives a scholarship in the form of a 
stipend from the University of Auckland as a PhD student. JB is currently 
employed by a medical writing company. The preparation of this review took place 
prior to this employment and her current work is not related to the topic of 
this review. JH has partial salary support from research grants from the Health 
Research Council of New Zealand. The Council has no role in the production of 
this review. An undirected research grant is pending from Biomed Ltd, Auckland, 
New Zealand. This company makes dextrose gel. Core editorial and administrative 
support for the 2020 update of this review has been provided by a grant from The 
Gerber Foundation. The Gerber Foundation is a separately endowed, private 
foundation, independent from the Gerber Products Company. The grantor has no 
input on the content of the review or the editorial process (see Sources of 
support).


55. Cochrane Database Syst Rev. 2020 Jul 29;7(7):CD013392. doi: 
10.1002/14651858.CD013392.pub2.

Early fortification of human milk versus late fortification to promote growth in 
preterm infants.

Thanigainathan S(1), Abiramalatha T(2).

Author information:
(1)Neonatology, Jawaharlal Institute of Postgraduate Medical Education and 
Research, Puducherry, India.
(2)Neonatology, Sri Ramachandra Institute of Higher Education and Research, 
Chennai, India.

Update of
    doi: 10.1002/14651858.CD013392.

BACKGROUND: Uncertainty exists about the optimal point at which multi-component 
fortifier should be added to human milk for promoting growth in preterm infants. 
The most common practice is to start fortification when the infant's daily 
enteral feed volume reaches 100 mL/kg body weight. Another approach is to 
commence fortification earlier, in some cases as early as the first enteral 
feed. Early fortification of human milk could increase nutrient intake and 
growth rates but may increase the risk of feed intolerance and necrotising 
enterocolitis (NEC).
OBJECTIVES: To assess effects on growth and safety of early fortification of 
human milk versus late fortification in preterm infants To assess whether 
effects vary based upon gestational age (≤ 27 weeks; 28 to 31 weeks; ≥ 32 
weeks), birth weight (< 1000 g; 1000 to 1499 g; ≥ 1500 g), small or appropriate 
for gestational age, or type of fortifier (bovine milk-based human milk 
fortifier (HMF); human milk-based HMF; formula powder) SEARCH METHODS: We used 
the standard strategy of Cochrane Neonatal to search the Cochrane Central 
Register of Controlled Trials (CENTRAL; 2019, Issue 8); OVID MEDLINE (R) and 
Epub Ahead of Print, In-Process & Other Non-Indexed Citations, Daily and 
Versions (R) (1946 to 15 August 2019); MEDLINE via PubMed (1 August 2018 to 15 
August 2019) for the previous year; and the Cumulative Index to Nursing and 
Allied Health Literatue (CINAHL) (1981 to 15 August 2019). We searched clinical 
trials databases and reference lists of included studies.
SELECTION CRITERIA: We included randomised controlled trials that compared early 
versus late fortification of human milk in preterm infants. We defined early 
fortification as fortification started at < 100 mL/kg/d enteral feed volume or < 
7 days postnatal age, and late fortification as fortification started at ≥ 100 
mL/kg/d feeds or ≥ 7 days postnatal age.
DATA COLLECTION AND ANALYSIS: Both review authors assessed trial eligibility and 
risk of bias and independently extracted data. We analysed treatment effects in 
individual trials, and we reported risk ratio (RR) for dichotomous data and mean 
difference (MD) for continuous data, with respective 95% confidence intervals 
(CIs). We used the GRADE approach to assess the certainty of evidence.
MAIN RESULTS: We included two trials with a total of 237 infants. All 
participants were very low birth weight infants (birth weight < 1500 g). Early 
fortification was started at 20 mL/kg/d enteral feeds in one study and 40 
mL/kg/d in the other study. Late fortification was started at 100 mL/kg/d feeds 
in both studies. One study used bovine milk-based fortifier, and the other used 
human milk-based fortifier. Meta-analysis showed that early fortification may 
have little or no effect on growth outcomes including time to regain birth 
weight (MD -0.06 days, 95% CI -1.32 to 1.20 days), linear growth (MD 0.10 
cm/week, 95% CI -0.03 to 0.22 cm/week), or head growth (MD -0.01 cm/week, 95% CI 
-0.07 to 0.06 cm/week) during the initial hospitalisation period. Early 
fortification may have little or no effect on the risk of NEC (MD -0.01, 95% CI 
-0.07 to 0.06). The certainty of evidence was low for these outcomes due to risk 
of bias (lack of blinding) and imprecision (small sample size). Early 
fortification may have little or no effect on incidence of surgical NEC, time to 
reach full enteral feeds, extrauterine growth restriction at discharge, 
proportion of infants with feed interruption episodes, duration of total 
parenteral nutrition (TPN), duration of central venous line usage, or incidence 
of invasive infection, all-cause mortality, and duration of hospital stay. The 
certainty of evidence was low for these outcomes due to risk of bias (lack of 
blinding) and imprecision (small sample size). We did not have data for other 
outcomes such as subsequent weight gain after birth weight is regained, 
parenteral nutrition-associated liver disease, postdischarge growth, and 
neurodevelopmental outcomes.
AUTHORS' CONCLUSIONS: Available evidence is insufficient to support or refute 
early fortification of human milk in preterm infants. Further large trials would 
be needed to provide data of sufficient quality and precision to inform policy 
and practice.

Copyright © 2020 The Cochrane Collaboration. Published by John Wiley & Sons, 
Ltd.

DOI: 10.1002/14651858.CD013392.pub2
PMCID: PMC7390609
PMID: 32726863 [Indexed for MEDLINE]

Conflict of interest statement: ST has no conflict of interest to declare. TA 
has no conflict of interest to declare. Core editorial and administrative 
support for this review has been provided by a grant from The Gerber Foundation. 
The Gerber Foundation is a separately endowed, private foundation that is 
independent from the Gerber Products Company. The grantor has no input on the 
content of the review nor on the editorial process (Sources of support).


56. Nutrients. 2020 Jul 9;12(7):2044. doi: 10.3390/nu12072044.

Feeding Interventions for Infants with Growth Failure in the First Six Months of 
Life: A Systematic Review.

Rana R(1)(2)(3), McGrath M(4), Gupta P(1), Thakur E(1), Kerac M(2)(5).

Author information:
(1)Indian Institute of Public Health Gandhinagar, Gujarat 382042, India.
(2)Department of Population Health, London School of Hygiene & Tropical 
Medicine, London WC1E 7HT, UK.
(3)Nutrition Research Advisor (MAMI), GOAL Global, A96 C7W7 Dublin, Ireland.
(4)Emergency Nutrition Network, Oxford OX5 2DN, UK.
(5)Centre for Maternal, Adolescent, Reproductive & Child Health (MARCH), London 
School of Hygiene & Tropical Medicine, London WC1E 7HT, UK.

(1) Introduction: Current evidence on managing infants under six months with 
growth failure or other nutrition-related risk is sparse and low quality. This 
review aims to inform research priorities to fill this evidence gap, focusing on 
breastfeeding practices. (2) Methods: We searched PubMed, CINAHL Plus, and 
Cochrane Library for studies on feeding interventions that aim to restore or 
improve the volume or quality of breastmilk and breastfeeding when breastfeeding 
practices are sub-optimal or prematurely stopped. We included studies from both 
low- and middle-income countries and high-income countries. (3) Results: 
Forty-seven studies met the inclusion criteria. Most were from high-income 
countries (n = 35, 74.5%) and included infants who were at risk of growth 
failure at birth (preterm infants/small for gestational age) and newborns with 
early growth faltering. Interventions included formula fortification or 
supplementation (n = 31, 66%), enteral feeds (n = 8, 17%), cup feeding (n = 2, 
4.2%), and other (n = 6, 12.8%). Outcomes included anthropometric change (n = 
40, 85.1%), reported feeding practices (n = 16, 34%), morbidity (n = 11, 23.4%), 
and mortality (n = 5, 10.6%). Of 31 studies that assessed formula fortification 
or supplementation, 30 reported anthropometric changes (n = 17 no effect, n = 9 
positive, n = 4 mixed), seven morbidity (n = 3 no effect, n = 2 positive, n = 2 
negative), five feeding (n = 2 positive, n = 2 no effect, n = 1 negative), and 
four mortality (n = 3 no effect, n = 1 negative). Of eight studies that assessed 
enteral feed interventions, seven reported anthropometric changes (n = 4 
positive, n = 3 no effect), five feeding practices (n = 2 positive, n = 2 no 
effect, n = 1 negative), four morbidity (n = 4 no effect), and one reported 
mortality (n = 1 no effect). Overall, interventions with positive effects on 
feeding practices were cup feeding compared to bottle-feeding among preterm; 
nasogastric tube feed compared to bottle-feeding among low birth weight preterm; 
and early progressive feeding compared to delayed feeding among extremely low 
birth weight preterm. Bovine/cow milk feeding and high volume feeding 
interventions had an unfavourable effect, while electric breast pump and 
Galactagogue had a mixed effect. Regarding anthropometric outcomes, overall, 
macronutrient fortified formula, cream supplementation, and fortified human milk 
formula had a positive effect (weight gain) on preterm infants. Interventions 
comparing human breastmilk/donor milk with formula had mixed effects. Overall, 
only human milk compared to formula intervention had a positive effect on 
morbidity among preterm infants, while none of the interventions had any 
positive effect on mortality. Bovine/cow milk supplementation had unfavourable 
effects on both morbidity and mortality. (4) Conclusion: Future research should 
prioritise low- and middle-income countries, include infants presenting with 
growth failure in the post-neonatal period and record effects on morbidity and 
mortality outcomes.

DOI: 10.3390/nu12072044
PMCID: PMC7400880
PMID: 32660020 [Indexed for MEDLINE]

Conflict of interest statement: The authors declare no conflict of interest.


57. Iron from Supplements Consumed During Infancy and Toddlerhood and Growth, Size, 
and Body Composition: A Systematic Review [Internet].

Dewey K(1), Bazzano L(2), Davis T(3), Donovan S(4), Taveras E(5), Kleinman R(6), 
Güngör D(7), Venkatramanan S(7), Madan E(7), English LK(7), Terry N(8), Butera 
G(9), Obbagy J(10).

Alexandria (VA): USDA Nutrition Evidence Systematic Review; 2020 Jul.
USDA Nutrition Evidence Systematic Reviews.

Author information:
(1)Chair, Birth to 24 Months Subcommittee, 2020 Dietary Guidelines Advisory 
Committee; University of California, Davis
(2)Member, Birth to 24 Months Subcommittee, 2020 Dietary Guidelines Advisory 
Committee; Tulane University and Ochsner Health System
(3)Member, Birth to 24 Months Subcommittee, 2020 Dietary Guidelines Advisory 
Committee; Baylor College of Medicine
(4)Member, Birth to 24 Months Subcommittee, 2020 Dietary Guidelines Advisory 
Committee; University of Illinois, Urbana-Champaign
(5)Member, Birth to 24 Months Subcommittee, 2020 Dietary Guidelines Advisory 
Committee; Massachusetts General Hospital, Harvard Medical School, and Harvard 
T.H. Chan School of Public Health
(6)Vice-Chair, 2020 Dietary Guidelines Advisory Committee; Massachusetts General 
Hospital, Harvard Medical School
(7)Systematic review analyst, Nutrition Evidence Systematic Review (NESR) team; 
Panum Group under contract with the Food and Nutrition Service (FNS), U.S. 
Department of Agriculture (USDA)
(8)Biomedical librarian, NESR team; National Institutes of Health Library, U.S. 
Department of Health and Human Services
(9)Systematic review librarian, NESR team; Panum Group under contract with the 
FNS, USDA
(10)Project Lead, NESR team; Nutrition Guidance and Analysis Division, Center 
for Nutrition Policy and Promotion, FNS, USDA

BACKGROUND: This important public health question was identified by the U.S. 
Departments of Agriculture (USDA) and Health and Human Services (HHS) to be 
examined by the 2020 Dietary Guidelines Advisory Committee. The 2020 Dietary 
Guidelines Advisory Committee, Birth to 24 Months Subcommittee, conducted a 
systematic review to answer this question with support from the Nutrition 
Evidence Systematic Review (NESR) team. The goal of this systematic review was 
to examine the following question: What is the relationship between iron from 
supplements consumed during infancy and toddlerhood and growth, size, and body 
composition?
CONCLUSION STATEMENTS AND GRADES: Moderate evidence indicates that human 
milk-fed infants who are supplemented with iron do not have greater growth, and 
may have slower growth, than human milk-fed infants not supplemented with iron. 
(Grade: Moderate). Insufficient evidence is available to determine the 
relationship between iron from supplements consumed during infancy and body 
composition during infancy. (Grade: Grade not assignable). Insufficient evidence 
is available to determine the relationship between iron from supplements 
consumed during infancy and growth, size, and body composition beyond age 12 
months. (Grade: Grade not assignable). Insufficient evidence is available to 
determine the relationship between iron from supplements consumed after age 12 
months and growth, size, and body composition. (Grade: Grade not assignable).
METHODS: A literature search was conducted using 4 databases (PubMed, Embase, 
CINAHL, and Cochrane) to identify articles that evaluated the intervention or 
exposure of iron from supplements consumed during infancy and toddlerhood and 
growth, size, and body composition outcomes. A manual search was conducted to 
identify articles that may not have been included in the electronic databases 
searched. Articles were screened by two NESR analysts independently for 
inclusion based on pre-determined criteria. Data extraction and risk of bias 
assessment were conducted for each included study, and both were checked for 
accuracy. The Committee qualitatively synthesized the body of evidence to inform 
development of conclusion statements, and graded the strength of evidence using 
pre-established criteria for risk of bias, consistency, directness, precision, 
and generalizability.
SUMMARY OF THE EVIDENCE: Ten articles met the inclusion criteria for this 
systematic review, which presented evidence from 8 randomized controlled trials, 
1 non-randomized controlled trial, and 1 study that did not clearly describe its 
prospective study design. The intervention or exposure of interest was iron from 
supplements consumed during infancy and toddlerhood. Dietary supplements are 
products that contain one or more dietary ingredients (in this case, iron) 
intended to be taken by mouth to supplement the diet. Nine studies examined iron 
supplementation during infancy, and only 1 study examined iron supplementation 
during toddlerhood. The comparators of interest were different dosages of iron 
from supplements and iron from fortified foods. The outcomes of interest were 
measures of growth, size, and body composition at any age. However, no articles 
were identified that examined outcomes beyond 24 months. The articles presented 
evidence about growth (i.e., change in size between birth or baseline and 
follow-up) and size (i.e., attained size at follow-up). However, no articles 
presented evidence about body composition (e.g., percent fat mass, skinfold 
thickness). Moderate evidence, from 5 studies that compared iron from 
supplements with no iron from supplements, indicated that human milk-fed infants 
who are supplemented with iron do not have greater growth, and may have slower 
growth, than human milk-fed infants not supplemented with iron. Inconsistencies 
in the evidence may be explained by differences in the risk of iron deficiency 
between the populations studied, differences in participants’ consumption of 
iron-fortified formula or iron-rich foods, and differences in the timing of iron 
supplementation. This heterogeneity, the small number of studies, and the small 
sample sizes, were the primary factors limiting the ability to draw stronger 
conclusions. Evidence available from 3 studies was insufficient to determine 
whether a relationship exists between iron from supplements, compared with a 
different dosage or duration of iron from supplements, and growth or size, 
because the studies used heterogeneous interventions that could not be compared. 
Evidence available from 2 studies was insufficient to determine whether a 
relationship exists between iron from supplements, compared with iron from 
fortified foods, and growth or size, because the studies used heterogeneous 
interventions that could not be compared.

DOI: 10.52570/NESR.DGAC2020.SR0303
PMID: 35324134


58. Vitamin D from Supplements Consumed during Infancy and Toddlerhood and Bone 
Health: A Systematic Review [Internet].

Dewey K(1), Bazzano L(2), Davis T(3), Donovan S(4), Taveras E(5), Kleinman R(6), 
Güngör D(7), Venkatramanan S(7), Madan E(7), English LK(7), Terry N(8), Butera 
G(9), Obbagy J(10).

Alexandria (VA): USDA Nutrition Evidence Systematic Review; 2020 Jul.
USDA Nutrition Evidence Systematic Reviews.

Author information:
(1)Chair, Birth to 24 Months Subcommittee, 2020 Dietary Guidelines Advisory 
Committee; University of California, Davis
(2)Member, Birth to 24 Months Subcommittee, 2020 Dietary Guidelines Advisory 
Committee; Tulane University and Ochsner Health System
(3)Member, Birth to 24 Months Subcommittee, 2020 Dietary Guidelines Advisory 
Committee; Baylor College of Medicine
(4)Member, Birth to 24 Months Subcommittee, 2020 Dietary Guidelines Advisory 
Committee; University of Illinois, Urbana-Champaign
(5)Member, Birth to 24 Months Subcommittee, 2020 Dietary Guidelines Advisory 
Committee; Massachusetts General Hospital, Harvard Medical School, and Harvard 
T.H. Chan School of Public Health
(6)Vice-Chair, 2020 Dietary Guidelines Advisory Committee; Massachusetts General 
Hospital, Harvard Medical School
(7)Systematic review analyst, Nutrition Evidence Systematic Review (NESR) team; 
Panum Group under contract with the Food and Nutrition Service (FNS), U.S. 
Department of Agriculture (USDA)
(8)Biomedical librarian, NESR team; National Institutes of Health Library, U.S. 
Department of Health and Human Services
(9)Systematic review librarian, NESR team; Panum Group under contract with the 
FNS, USDA
(10)Project Lead, NESR team; Nutrition Guidance and Analysis Division, Center 
for Nutrition Policy and Promotion, FNS, USDA

BACKGROUND: This important public health question was identified by the U.S. 
Departments of Agriculture (USDA) and Health and Human Services (HHS) to be 
examined by the 2020 Dietary Guidelines Advisory Committee. The 2020 Dietary 
Guidelines Advisory Committee, Birth to 24 Months Subcommittee conducted a 
systematic review to answer this question with support from the Nutrition 
Evidence Systematic Review (NESR) team. The goal of this systematic review was 
to examine the following question: What is the relationship between vitamin D 
from supplements consumed during infancy and toddlerhood and bone health?
CONCLUSION STATEMENTS AND GRADES: Limited evidence suggests no relationship 
between consumption of 400 IU per day of vitamin D from supplements before age 
12 months, compared with higher dosages of up to 1600 IU per day, and biomarkers 
of bone metabolism in children up to age 36 months. (Grade: Limited). 
Insufficient evidence is available to determine the relationship between 400 IU 
per day of vitamin D from supplements, compared with higher dosages, and bone 
mass, rickets, or fracture. (Grade: Grade not assignable). Insufficient evidence 
is available to determine the relationship between 400 IU per day of vitamin D 
from supplements, compared with lower dosages, and bone mass, biomarkers of bone 
metabolism, rickets, or fracture. (Grade: Grade not assignable). Insufficient 
evidence is available to determine the relationship between vitamin D from 
supplements, compared with no vitamin D from supplements, and bone mass, 
biomarkers of bone metabolism, rickets, or fracture. (Grade: Grade not 
assignable). Insufficient evidence is available to determine the relationship 
between vitamin D from supplements, compared with vitamin D from fortified 
foods, and bone mass, biomarkers of bone metabolism, rickets, or fracture. 
(Grade: Grade not assignable).
METHODS: A literature search was conducted using 4 databases (PubMed, Cochrane, 
Embase, and CINAHL) to identify articles that evaluated the intervention or 
exposure of vitamin D from supplements consumed during infancy and toddlerhood 
and bone health outcomes. A manual search was conducted to identify articles 
that may not have been included in the electronic databases searched. Articles 
were screened by two NESR analysts independently for inclusion based on 
pre-determined criteria. Data extraction and risk of bias assessment were 
conducted for each included study, and both were checked for accuracy. The 
Committee qualitatively synthesized the body of evidence to inform development 
of conclusion statements, and graded the strength of evidence using 
pre-established criteria for risk of bias, consistency, directness, precision, 
and generalizability.
SUMMARY OF THE EVIDENCE: Six articles met the inclusion criteria for this 
systematic review, which presented evidence from 5 independent randomized 
controlled trials (1 research group published 2 articles about the same trial). 
The intervention of interest was vitamin D from supplements consumed during 
infancy or toddlerhood. Dietary supplements are products that contain one or 
more dietary ingredients (in this case, vitamin D) intended to be taken by mouth 
to supplement the diet. In the United States, 400 IU of vitamin D per day is the 
AI for infants younger than age 12 months, whereas the RDA for ages 12 to 24 
months of age is 600 IU per day. To meet this need, the American Academy of 
Pediatrics currently recommends a supplement of 400 IU per day for infants fed 
human milk (with the possible exception of infants whose mothers are taking 
supplements of about 6000 IU per day; maternal vitamin D supplementation during 
lactation was outside of the scope of this systematic review). The comparators 
of interest were different dosages of vitamin D from supplements and vitamin D 
from fortified foods. However, no articles were identified that included 
fortified food comparators. The outcomes of interest were bone mass, biomarkers 
of bone metabolism, rickets, and fracture through adolescence (i.e., birth 
through age 18 years). However, no articles were identified that examined 
fracture or outcomes beyond age 36 months. Limited evidence from 3 studies 
suggests no relationship between 400 IU per day of vitamin D from supplements, 
compared with higher dosages, and biomarkers of bone metabolism in children up 
to age 36 months. The ability to draw a stronger conclusion was primarily 
limited by a small number of studies, small sample sizes, heterogeneous methods, 
and limited generalizability. Evidence available from 4 studies was insufficient 
to determine whether a relationship exists between 400 IU per day of vitamin D 
from supplements, compared with higher dosages, and bone mass. The ability to 
draw a conclusion was hindered by inconsistent findings from a small number of 
studies. No studies were available that examined the relationship between 400 IU 
per day of vitamin D from supplements, compared with higher dosages, and rickets 
or bone fracture. Evidence available from 1 study was insufficient to determine 
whether a relationship exists between 400 IU per day of vitamin D from 
supplements, compared with lower dosages, and bone mass or biomarkers of bone 
metabolism. No studies were available that examined the relationship between 400 
IU per day of vitamin D from supplements, compared with lower dosages, and 
rickets or fracture. Evidence available from 1 study was insufficient to 
determine whether a relationship exists between 200 IU per day of vitamin D from 
supplements for different durations, compared with no vitamin D from 
supplements, and biomarkers of bone metabolism or rickets. No studies were 
available that examined the relationship between 200 IU per day of vitamin D 
from supplements for different durations, compared with no vitamin D from 
supplements, and bone mass or fracture. No studies were available that compared 
other dosages of vitamin D from supplements with no supplementation. It is 
likely that the evidence that led to the current supplementation recommendation 
pre-dates our literature search date range of January 2000 to January 2020.

DOI: 10.52570/NESR.DGAC2020.SR0304
PMID: 35315998


59. Folic Acid from Fortified Foods and/or Supplements during Pregnancy and 
Lactation and Health Outcomes: A Systematic Review [Internet].

Donovan S(1), Dewey K(2), Novotny R(3), Stang J(4), Taveras E(5), Kleinman R(6), 
Spill M(6), Kim JH(7), Nevins J(7), Raghavan R(7), Scinto-Madonich S(7), Butera 
G(8), Terry N(9), Obbagy J(10).

Alexandria (VA): USDA Nutrition Evidence Systematic Review; 2020 Jul.
USDA Nutrition Evidence Systematic Reviews.

Author information:
(1)Chair, Pregnancy and Lactation Subcommittee, 2020 Dietary Guidelines Advisory 
Committee; University of Illinois, Urbana-Champaign
(2)Member, Pregnancy and Lactation Subcommittee, 2020 Dietary Guidelines 
Advisory Committee; University of California, Davis
(3)Member, Pregnancy and Lactation Subcommittee, 2020 Dietary Guidelines 
Advisory Committee; University of Hawaii
(4)Member, Pregnancy and Lactation Subcommittee, 2020 Dietary Guidelines 
Advisory Committee; University of Minnesota
(5)Member, Pregnancy and Lactation Subcommittee, 2020 Dietary Guidelines 
Advisory Committee; Massachusetts General Hospital, Harvard Medical School, and 
Harvard T.H. Chan School of Public Health
(6)Vice-Chair, 2020 Dietary Guidelines Advisory Committee; Massachusetts General 
Hospital, Harvard Medical School
(7)Systematic review analyst, Nutrition Evidence Systematic Review (NESR) team; 
Panum Group under contract with the Food and Nutrition Service (FNS), U.S. 
Department of Agriculture (USDA)
(8)Biomedical librarian, NESR team; National Institutes of Health Library, U.S. 
Department of Health and Human Services
(9)Systematic review librarian, NESR team; Panum Group under contract with the 
FNS, USDA
(10)Project Lead, NESR team; Nutrition Guidance and Analysis Division, Center 
for Nutrition Policy and Promotion, FNS, USDA

The goal of this systematic review was to examine the following questions: ○. 
What is the relationship between folic acid from supplements and/or fortified 
foods consumed before and during pregnancy and lactation and micronutrient 
status? ○. What is the relationship between folic acid from supplements and/or 
fortified foods consumed before and during pregnancy and risk of gestational 
diabetes? ○. What is the relationship between folic acid from supplements and/or 
fortified foods consumed before and during pregnancy and risk of hypertensive 
disorders during pregnancy? ○. What is the relationship between folic acid from 
supplements and/or fortified foods consumed before and during pregnancy and 
lactation and human milk composition? ○. What is the relationship between folic 
acid from supplements and/or fortified foods consumed before and during 
pregnancy and lactation and developmental milestones, including neurocognitive 
development, in the child?

DOI: 10.52570/NESR.DGAC2020.SR0205
PMID: 35289987


60. Syst Rev. 2020 Jun 16;9(1):144. doi: 10.1186/s13643-020-01360-3.

The impact of vitamin D food fortification and health outcomes in children: a 
systematic review and meta-regression.

Al Khalifah R(1)(2), Alsheikh R(3), Alnasser Y(4)(3)(5), Alsheikh R(3), Alhelali 
N(4), Naji A(4), Al Backer N(4)(6).

Author information:
(1)Division of Pediatric Endocrinology, Pediatric Department, College of 
Medicine, King Saud University, Riyadh, Saudi Arabia..
(2)College of Medicine, King Saud University, Riyadh, Saudi Arabia. 
.
(3)Pediatric Department, King Saud University, Riyadh, Saudi Arabia.
(4)College of Medicine, King Saud University, Riyadh, Saudi Arabia.
(5)Pediatric Department, BC Children's Hospital, Vancouver, BC, Canada.
(6)Division of Developmental-Behavioral Pediatric, Pediatric Department, College 
of Medicine, King Saud University, Riyadh, Saudi Arabia.

OBJECTIVE: Vitamin D (vitD) deficiency is a global childhood health problem. 
Food fortification is a promising strategy to curb vitD deficiency. We aimed to 
assess the effectiveness of utilizing vitD fortification in staple foods to 
improve 25hydroxyvitamin D (25(OH)D) concentration and to reduce the prevalence 
of vitD deficiency among healthy children.
METHODS: We conducted a systematic review and meta-analysis of randomized 
controlled trials (RCTs) evaluating the use of vitD fortified food products 
compared to no fortification among healthy children aged 1-18 years old. We 
searched Medline, Embase, Global Health, and Cochrane (CENTRAL) databases from 
database inception until May 2019. Independently, six reviewers in pairs 
screened titles and abstracts, assessed the full text for eligibility, and 
performed data extraction and quality assessment. The primary outcome is the 
impact of fortification on 25(OH)D concentration. The secondary outcomes 
included the impact of fortification on the prevalence of vitD deficiency, 
school performance, cognitive function, school absences, infection rate, 
hospital admission length, and compliance with fortified food product 
consumption.
RESULTS: We identified 2229 articles. After assessing eligibility, 20 RCTs met 
the inclusion criteria. The eligible RCTs assessed the fortification of milk, 
cereal, juice, bread, yogurt, and cheese compared with no fortification. All 
RCTs, except for three, had a low risk of bias. Food fortification improved 
25(OH)D concentration by a mean difference (MD) of 15.51 nmol/L (95% confidence 
interval (CI) 6.28, 24.74; I2 = 99%), which resulted in a mean increase of 
3 nmol/l for every 100 IU of vitD, when adjusted for baseline 25(OH)D 
concentration and country latitude. Additionally, the prevalence of vitD 
deficiency decreased by a risk ratio of 0.53 (95% CI 0.41, 0.69; I2 = 95%), and 
cognitive function improved by a MD of 1.22 intelligence quotient (IQ) points 
(95% CI 0.65, 1.79; I2 = 0%). The overall evidence quality was high.
CONCLUSION: VitD food fortification is an effective way to improve 25(OH)D 
concentration, prevent vitD deficiency, and improve IQ levels.
SYSTEMATIC REVIEW REGISTRATION: PROSPERO CRD42017057631.

DOI: 10.1186/s13643-020-01360-3
PMCID: PMC7298752
PMID: 32546259 [Indexed for MEDLINE]

Conflict of interest statement: The authors declare that they have no competing 
interests.


61. Cochrane Database Syst Rev. 2020 Jun 3;6(6):CD000343. doi: 
10.1002/14651858.CD000343.pub4.

Multi-nutrient fortification of human milk for preterm infants.

Brown JV(1), Lin L(2), Embleton ND(3), Harding JE(2), McGuire W(4).

Author information:
(1)Mental Health and Addiction Research Group, Department of Health Sciences, 
University of York, York, UK.
(2)Liggins Institute, University of Auckland, Auckland, New Zealand.
(3)Newcastle Neonatal Service, Newcastle Hospitals NHS Foundation Trust and 
University of Newcastle, Newcastle upon Tyne, UK.
(4)Centre for Reviews and Dissemination, University of York, York, UK.

Update of
    doi: 10.1002/14651858.CD000343.pub3.

BACKGROUND: Human breast milk-fed preterm infants can accumulate nutrient 
deficits leading to extrauterine growth restriction. Feeding preterm infants 
with multi-nutrient fortified human milk could increase nutrient accretion and 
growth rates and improve neurodevelopmental outcomes. Concern exists, however, 
that multi-nutrient fortifiers are associated with adverse events such as feed 
intolerance and necrotising enterocolitis.
OBJECTIVES: To determine whether multi-nutrient fortified human milk, compared 
with unfortified human milk, affects important outcomes (including growth rate 
and neurodevelopment) of preterm infants without increasing the risk of adverse 
effects (such as feed intolerance and necrotising enterocolitis).
SEARCH METHODS: We searched the Cochrane Central Register of Controlled Trials 
(CENTRAL; 2019, Issue 9), MEDLINE via PubMed (1966 to 26 September 2019), Embase 
(1980 to 26 September 2019), and the Cumulative Index to Nursing and Allied 
Health Literature (CINAHL; 1982 to 26 September 2019). We searched clinical 
trials databases, conference proceedings, and reference lists of retrieved 
articles for randomised controlled trials and quasi-randomised trials.
SELECTION CRITERIA: Randomised and quasi-randomised controlled trials that 
compared feeding preterm infants with multi-nutrient (protein and energy plus 
minerals, vitamins, or other nutrients) fortified human breast milk versus 
unfortified (no added protein or energy) breast milk.
DATA COLLECTION AND ANALYSIS: We used the standard methods of Cochrane Neonatal. 
Two review authors separately evaluated trial quality, extracted data, and 
synthesised effect estimates using risk ratios (RRs), risk differences, and mean 
differences (MDs). We assessed the certainty of the body of evidence at the 
outcome level using "Grading of Recommendations Assessment, Development and 
Evaluation" (GRADE) methods.
MAIN RESULTS: We identified 18 trials in which a total of 1456 preterm infants 
participated. These trials were generally small and methodologically weak. 
Meta-analyses provided low- to moderate-certainty evidence showing that 
multi-nutrient fortification of human milk increases in-hospital rate of weight 
gain (MD 1.76 g/kg/d, 95% confidence interval (CI) 1.30 to 2.22), body length 
(MD 0.11 cm/week, 95% CI 0.08 to 0.15), or head circumference (MD 0.06 cm/week, 
95% CI 0.03 to 0.08) among preterm infants. Few data on growth and developmental 
outcomes assessed beyond infancy are available, and these do not show effects of 
multi-nutrient fortification. The data do not suggest other benefits or harms 
and provide low-certainty evidence suggesting effects of multi-nutrient 
fortification on the risk of necrotising enterocolitis in preterm infants 
(typical RR 1.37, 95% CI 0.72 to 2.63; 13 studies, 1110 infants).
AUTHORS' CONCLUSIONS: Feeding preterm infants with multi-nutrient fortified 
human breast milk compared with unfortified human breast milk is associated with 
modest increases in in-hospital growth rates. Evidence is insufficient to show 
whether multi-nutrient fortification has any effect on long-term growth or 
neurodevelopment.

Copyright © 2020 The Cochrane Collaboration. Published by John Wiley & Sons, 
Ltd.

DOI: 10.1002/14651858.CD000343.pub4
PMCID: PMC7268980
PMID: 35658821

Conflict of interest statement: JVEB: nothing to declare. LL: nothing to 
declare. NDE declares the following: receiving a research grant award for a 
randomised controlled trial of breast milk products from Prolacta Bioscience, 
2017; receiving a grant from Danone Early Life Nutrition to support a study on 
feeding in late and moderately preterm infants, 2018; receiving a grant from 
Nestle Nutrition for transcriptomic analyses of gut tissue, 2016; and presenting 
lectures with Wyeth Nutrition in 2017, Nestle Nutrition Institute in 2017 and 
2018, Philipps in 2017, and Fresenius in 2017. JH: nothing to declare. WM: 
nothing to declare. Core editorial and administrative support for this review 
has been provided by a grant from The Gerber Foundation. The Gerber Foundation 
is a separately endowed, private foundation, independent from the Gerber 
Products Company. The grantor has no input on the content of the review or the 
editorial process (see Sources of support).


62. Eur J Pediatr. 2020 Jul;179(7):1057-1068. doi: 10.1007/s00431-020-03677-6. Epub 
2020 May 27.

Early versus late fortification of breast milk in preterm infants: a systematic 
review and meta-analysis.

Basu S(1), Upadhyay J(2), Singh P(2), Kumar M(3).

Author information:
(1)Department of Neonatology, All India Institute of Medical Sciences, 
Rishikesh, Uttarakhand, 249203, India..
(2)Department of Neonatology, All India Institute of Medical Sciences, 
Rishikesh, Uttarakhand, 249203, India.
(3)Department of Pediatrics, All India Institute of Medical Sciences, Gorakhpur, 
India.

This systematic review and meta-analysis assessed the effects of early 
fortification (EF) versus late fortification (LF) of breast milk (BM) on growth 
of preterm infants. Randomized and quasi-randomized controlled trials (RCTs) 
dealing with the effects of EF versus LF on growth parameters, incidence of 
adverse events, and duration of hospital stay in preterm infants were included. 
Data were pooled using the RevMan 5.3 software. Quality of evidence for 
predefined outcomes was analyzed by GRADE. Available evidence (3 RCTs, 309 
preterm infants) showed no statistically significant difference between EF and 
LF of BM for any of the growth parameters-weight (standardized mean difference 
(SMD) 0.13; 95% confidence interval (CI) - 0.09, 0.36); length (SMD 0.02; 95% CI 
- 0.20, 0.25); and head circumference (SMD - 0.10; 95% CI - 0.33, 0.12). Total 
parenteral nutrition days were similar. Duration of hospital stay was 
significantly higher with EF (MD 4.29; 95% CI 0.84, 7.75) with a trend of 
non-significant increase in feed intolerance and necrotizing enterocolitis 
(NEC).Conclusion: Very low quality evidence did not find any significant 
difference in growth parameters of preterm infants in association with EF or LF 
of BM. A significant increase in hospital stay and non-significant increase in 
feed intolerance and NEC were associated with EF.PROSPERO registration number: 
CRD42019139235What is Known:• Fortification of breast milk with essential macro- 
and micronutrients is necessary for optimization of nutrition in preterm 
infants.• There is no consensus regarding the breast milk feeding volume at 
which fortification should be initiated.What is New:• Very low quality evidence 
showed no significant difference between early and late fortification of breast 
milk on growth parameters of preterm infants.• Early fortification was 
associated with non-significant increase in feed intolerance and necrotizing 
enterocolitis and a significant increase in hospital stay.

DOI: 10.1007/s00431-020-03677-6
PMID: 32458060 [Indexed for MEDLINE]


63. Nutrients. 2020 May 20;12(5):1487. doi: 10.3390/nu12051487.

Effects of Prebiotic and Probiotic Supplementation on Lactase Deficiency and 
Lactose Intolerance: A Systematic Review of Controlled Trials.

Leis R(1)(2)(3)(4), de Castro MJ(1)(2)(5), de Lamas C(4), Picáns R(1), Couce 
ML(1)(2)(4)(5).

Author information:
(1)Department of Pediatrics, University Clinical Hospital of Santiago de 
Compostela, 15704 Santiago de Compostela, Spain.
(2)IDIS-Health Research Institute of Santiago de Compostela, 15704 Santiago de 
Compostela, Spain.
(3)CIBEROBN, Instituto Salud Carlos III, 28029 Madrid, Spain.
(4)Facultad de Medicina, Departamento de Pediatría, Universidad de Santiago de 
Compostela, 15704 Santiago de Compostela, Spain.
(5)CIBERER, Instituto Salud Carlos III, 28029 Madrid, Spain.

Lactose intolerance (LI) is characterized by the presence of primarily 
gastrointestinal clinical signs resulting from colonic fermentation of lactose, 
the absorption of which is impaired due to a deficiency in the lactase enzyme. 
These clinical signs can be modified by several factors, including lactose dose, 
residual lactase expression, concurrent ingestion of other dietary components, 
gut-transit time, and enteric microbiome composition. In many of individuals 
with lactose malabsorption, clinical signs may be absent after consumption of 
normal amounts of milk or, in particular, dairy products (yogurt and cheese), 
which contain lactose partially digested by live bacteria. The intestinal 
microbiota can be modulated by biotic supplementation, which may alleviate the 
signs and symptoms of LI. This systematic review summarizes the available 
evidence on the influence of prebiotics and probiotics on lactase deficiency and 
LI. The literature search was conducted using the MEDLINE (via PUBMED) and 
SCOPUS databases following Preferred Reporting Items for Systematic Reviews and 
Meta-Analyses (PRISMA) guidelines, and included randomized controlled trials. 
For each study selected, the risk of bias was assessed following the Cochrane 
Collaboration methodology. Our findings showed varying degrees of efficacy but 
an overall positive relationship between probiotics and LI in relation to 
specific strains and concentrations. Limitations regarding the wide 
heterogeneity between the studies included in this review should be taken into 
account. Only one study examined the benefits of prebiotic supplementation and 
LI. So further clinical trials are needed in order to gather more evidence.

DOI: 10.3390/nu12051487
PMCID: PMC7284493
PMID: 32443748 [Indexed for MEDLINE]

Conflict of interest statement: The authors declare no conflict of interest.


64. Osteoporos Int. 2020 Aug;31(8):1411-1425. doi: 10.1007/s00198-020-05383-3. Epub 
2020 May 7.

Systematic review and meta-analysis of the association between dairy consumption 
and the risk of hip fracture: critical interpretation of the currently available 
evidence.

Hidayat K(1)(2), Du X(3), Shi BM(4), Qin LQ(5).

Author information:
(1)Department of Endocrinology and Metabolism, The First Affiliated Hospital of 
Soochow University, No. 188 Shizi Street, Suzhou, 215006, China. 
.
(2)Department of Nutrition and Food Hygiene, School of Public Health, Soochow 
University, No. 199 Ren'ai Road, Suzhou, 215123, China..
(3)Department of Endocrinology and Metabolism, The First Affiliated Hospital of 
Soochow University, No. 188 Shizi Street, Suzhou, 215006, China.
(4)Department of Endocrinology and Metabolism, The First Affiliated Hospital of 
Soochow University, No. 188 Shizi Street, Suzhou, 215006, China. 
.
(5)Department of Nutrition and Food Hygiene, School of Public Health, Soochow 
University, No. 199 Ren'ai Road, Suzhou, 215123, China..

In the present meta-analysis, reductions in the risk of hip fracture with milk 
consumption were only observed among American adults, but not among Scandinavian 
adults, possibly because milk products are more commonly fortified with vitamin 
D in the former population than in Scandinavian countries. The reduction in the 
risk of hip fracture was also observed with yogurt consumption, which is often 
associated with healthy lifestyles and dietary patterns that contribute to 
improved bone health.
INTRODUCTION: Although dairy products contain bone-beneficial nutrients, the 
association between dairy consumption and the risk of hip fracture remains 
equivocal. Fueling this uncertainty, the elevated risk of hip fracture in 
association with milk consumption was observed in a cohort of Swedish women. A 
systematic review and meta-analysis of prospective cohort studies was performed 
to critically evaluate the association, or lack thereof, between dairy 
consumption (milk, yogurt, and cheese) and the risk of hip fracture.
METHODS: A random effects model was used to generate the summary relative risks 
(RRs) with their 95% confidence intervals (CIs) for the associations of 
interest.
RESULTS: In the meta-analysis of the highest versus lowest category of 
consumption, higher consumption of yogurt (RR 0.78, 95% CI 0.68, 0.90), but not 
milk (RR 0.86, 95% CI 0.73, 1.02) or cheese (RR 0.85, 95% CI 0.66, 1.08), was 
associated with a lower risk of hip fracture. For milk, the reduced risk of 
fracture with higher milk consumption was observed in the USA (RR 0.75, 95% CI 
0.65, 0.87), but not in Scandinavian countries (RR 1.00, 95% CI 0.85, 1.17). 
These findings were further supported by the fact that American studies (RR 
0.93, 95% CI 0.88, 0.98; per 1 glass/day), but not Scandinavian studies (RR 
1.01, 95% CI 0.95, 1.07; per 1 glass/day), demonstrated a linear association 
between milk consumption and the risk of hip fracture.
CONCLUSIONS: The cumulative evidence from prospective cohort studies 
reassuringly suggests that the risk of hip fracture may not be elevated among 
people who consume milk, yogurt, and cheese, and that a greater consumption of 
milk or yogurt may even be associated with a lower risk of hip fracture 
depending on the factors that may differ across the population of interest.

DOI: 10.1007/s00198-020-05383-3
PMID: 32383066 [Indexed for MEDLINE]


65. Breastfeed Med. 2020 Jun;15(6):377-386. doi: 10.1089/bfm.2019.0273. Epub 2020 
Apr 17.

Economic Analysis of Exclusive Human Milk Diets for High-Risk Neonates, a 
Canadian Hospital Perspective.

van Katwyk S(1)(2), Ferretti E(1)(3), Kumar S(1), Hutton B(1), Harrold J(1)(3), 
Walker M(2)(3), Forster AJ(1)(4)(5), Thavorn K(1)(2).

Author information:
(1)Ottawa Hospital Research Institute, Ontario, Canada.
(2)School of Epidemiology and Public Health, University of Ottawa, Ontario, 
Canada.
(3)Division of Neonatology, The Ottawa Hospital General Campus, Ontario, Canada.
(4)The Ottawa Hospital, Ontario, Canada.
(5)Faculty of Medicine, University of Ottawa, Ontario, Canada.

Background: There is increasing evidence that premature newborns and infants 
with low birth weight can benefit substantially from an exclusive human 
milk-based diet (EHMD), consisting of human milk supplemented with a pasteurized 
donor human milk-derived fortifier. However, compared with the standard infant 
diet, EHMD also represents a significant added cost to the hospital and/or 
health system, thereby raising important questions about the economic 
feasibility of incorporating EHMD into newborn care. Design: We conducted a cost 
analysis and estimated the potential cost savings to a Canadian tertiary 
hospital based on the attributable complications averted from EHMD among 
low-weight neonates. A meta-analysis was performed to derive input parameters. A 
probabilistic analysis was conducted to determine the probability that EHMD is 
cost saving and 95% confidence interval (CI) around our estimates. Results: Our 
findings show that providing EHMD to preterm infants under 750 g at birth and at 
the highest risk of developing major complications is likely to be cost saving 
in the amount of $107,567 (95% CI: -145,229 to 360,362) per year. Extending EHMD 
to higher weight classes may be economically feasible depending on the pricing 
of the human milk-derived fortifier and the baseline risk of complications in 
the hospital setting. Conclusions: This comprehensive study provides critical 
insight for hospital-based decision makers to evaluate the potential gains and 
uncertainties associated with improved nutritional care for neonatal patients.

DOI: 10.1089/bfm.2019.0273
PMID: 32302511 [Indexed for MEDLINE]


66. Adv Nutr. 2020 Sep 1;11(5):1325-1333. doi: 10.1093/advances/nmaa039.

Human Milk-Derived Fortifiers Compared with Bovine Milk-Derived Fortifiers in 
Preterm Infants: A Systematic Review and Meta-Analysis.

Ananthan A(1), Balasubramanian H(2), Rao S(3)(4), Patole S(4)(5).

Author information:
(1)Department of Neonatology, Seth GS Medical College and King Edward Memorial 
Hospital, Mumbai, India.
(2)Department of Neonatology, Surya Mother and Child Care Hospital, Mumbai, 
India.
(3)Department of Neonatal Paediatrics, Perth Children's Hospital, Perth, Western 
Australia, Australia.
(4)School of Medicine, University of Western Australia, Perth, Western 
Australia, Australia.
(5)Neonatal Directorate, King Edward Memorial Hospital for Women, Perth, Western 
Australia, Australia.

Comment in
    Adv Nutr. 2020 Nov 16;11(6):1713-1715. doi: 10.1093/advances/nmaa099.
    Adv Nutr. 2020 Nov 16;11(6):1712-1713. doi: 10.1093/advances/nmaa098.

This systematic review assessed outcomes after using human milk-derived 
fortifier (HMF) compared with bovine milk-derived fortifier (BMF) in preterm 
infants. Six randomized controlled trials (RCTs) were included. Meta-analysis 
using a random-effects model showed the following results: 1) lower risk of 
necrotizing enterocolitis (NEC; ≥Stage II) (RR: 0.38; 95% CI: 0.15, 0.95; 
P = 0.04, I2 = 9%; n = 334, 4 RCTs) and surgical NEC (RR: 0.13; 95% CI: 0.02, 
0.67; P = 0.02, I2 = 0%; n = 209, 3 RCTs) in the HMF group; 2) no significant 
difference in mortality (RR: 0.40; 95% CI: 0.14, 1.15; P = 0.09, I2 = 0%; 
n = 334, 4 RCTs); 3) lower weight gain in the HMF group [mean difference (MD) = 
-1.08 g · kg-1 · d-1; 95% CI: -1.96, -0.21 g · kg-1 · d-1; P = 0.02, I2 = 0%; 
n = 241, 4 RCTs]; 4) no differences for length (MD = -0.11 cm/wk; 95% CI: -0.26, 
0.04 cm/wk; P = 0.14, I2 = 68%) and head circumference (MD = -0.02 cm/wk; 95% 
CI: -0.08, 0.05 cm/wk; P = 0.59, I2 = 23%); and 5) no significant difference in 
late-onset sepsis (RR: 0.96; 95% CI: 0.56, 1.67; P = 0.90, I2 = 63%; n = 334, 4 
RCTs). The beneficial effects of HMF for NEC were no longer significant in 
sensitivity analyses after excluding studies with high risk of bias. Quality of 
evidence as per Grading of Recommendations, Assessment, Development and 
Evaluation (GRADE) analysis was low to very low, and hence the confidence in 
these results is low. In summary, fortification of milk in preterm infants with 
HMF compared with BMF decreased the risk of NEC but was associated with lower 
weight gain. Given the low quality of evidence, adequately powered and 
well-designed RCTs without the influence of industry are required in this field.

Copyright © The Author(s) 2020.

DOI: 10.1093/advances/nmaa039
PMCID: PMC7490161
PMID: 32277813 [Indexed for MEDLINE]


67. Nutrients. 2020 Jan 1;12(1):116. doi: 10.3390/nu12010116.

Effectiveness of Interventions for Managing Acute Malnutrition in Children under 
Five Years of Age in Low-Income and Middle-Income Countries: A Systematic Review 
and Meta-Analysis.

Das JK(1), Salam RA(1), Saeed M(2), Kazmi FA(1), Bhutta ZA(3)(4).

Author information:
(1)Division of Women and Child Health, Aga Khan University Hospital, Karachi 
74800, Pakistan.
(2)Medical Student, Aga Khan University, Karachi 74800, Pakistan.
(3)Centre of Excellence in Women and Child Health, Aga Khan University, Karachi 
74800, Pakistan.
(4)Centre for Global Child Health, the Hospital for Sick Children, Toronto, ON 
M5G 0A4, Canada.

Childhood malnutrition is a major public health concern, as it is associated 
with significant short- and long-term morbidity and mortality. The objective of 
this review was to comprehensively review the evidence for the management of 
severe acute malnutrition (SAM) and moderate acute malnutrition (MAM) according 
to the current World Health Organization (WHO) protocol using facility- and 
community-based approaches, as well as the effectiveness of ready-to-use 
therapeutic food (RUTF), ready-to-use supplementary food (RUSF), prophylactic 
antibiotic use, and vitamin A supplementation. We searched relevant electronic 
databases until 11 February 2019, and performed a meta-analysis. This review 
summarizes findings from a total of 42 studies (48 papers), including 35,017 
children. Limited data show some benefit of integrated community-based 
screening, identification, and management of SAM and MAM on improving recovery 
rate. Facility-based screening and management of uncomplicated SAM has no effect 
on recovery and mortality, while the effect of therapeutic milk F100 for SAM is 
comparable to RUTF for weight gain and mortality. Local food and whey RUSF are 
comparable to standard RUSF for recovery rate and weight gain in MAM, while 
standard RUSF has additional benefits to CSB. Prophylactic antibiotic 
administration in uncomplicated SAM improves recovery rate and probably improves 
weight gain and reduces mortality. Limited data suggest that high-dose vitamin A 
supplementation is comparable with low-dose vitamin A supplementation for weight 
gain and mortality among children with SAM.

DOI: 10.3390/nu12010116
PMCID: PMC7019612
PMID: 31906272 [Indexed for MEDLINE]

Conflict of interest statement: The authors declare no conflict of interest.


68. Cochrane Database Syst Rev. 2019 Dec 18;12(12):CD011400. doi: 
10.1002/14651858.CD011400.pub2.

Food fortification with multiple micronutrients: impact on health outcomes in 
general population.

Das JK(1), Salam RA(1)(2), Mahmood SB(3), Moin A(1), Kumar R(1), Mukhtar K(1), 
Lassi ZS(1)(4), Bhutta ZA(5).

Author information:
(1)Aga Khan University Hospital, Division of Women and Child Health, Stadium 
Road, PO Box 3500, Karachi, Sind, Pakistan.
(2)Healthy Mothers, Babies and Children, South Australian Health and Medical 
Research Institute, Adelaide, Australia.
(3)Aga Khan University Hospital, Department of Paediatrics, Karachi, Sindh, 
Pakistan.
(4)University of Adelaide, Robinson Research Institute, Adelaide, Australia, 
Australia.
(5)The Hospital for Sick Children, Centre for Global Child Health, Toronto, 
Canada.

Update of
    doi: 10.1002/14651858.CD011400.

BACKGROUND: Vitamins and minerals are essential for growth and maintenance of a 
healthy body, and have a role in the functioning of almost every organ. Multiple 
interventions have been designed to improve micronutrient deficiency, and food 
fortification is one of them.
OBJECTIVES: To assess the impact of food fortification with multiple 
micronutrients on health outcomes in the general population, including men, 
women and children.
SEARCH METHODS: We searched electronic databases up to 29 August 2018, including 
the Cochrane Central Register of Controlled Trial (CENTRAL), the Cochrane 
Effective Practice and Organisation of Care (EPOC) Group Specialised Register 
and Cochrane Public Health Specialised Register; MEDLINE; Embase, and 20 other 
databases, including clinical trial registries. There were no date or language 
restrictions. We checked reference lists of included studies and relevant 
systematic reviews for additional papers to be considered for inclusion.
SELECTION CRITERIA: We included randomised controlled trials (RCTs), 
cluster-RCTs, quasi-randomised trials, controlled before-after (CBA) studies and 
interrupted time series (ITS) studies that assessed the impact of food 
fortification with multiple micronutrients (MMNs). Primary outcomes included 
anaemia, micronutrient deficiencies, anthropometric measures, morbidity, 
all-cause mortality and cause-specific mortality. Secondary outcomes included 
potential adverse outcomes, serum concentration of specific micronutrients, 
serum haemoglobin levels and neurodevelopmental and cognitive outcomes. We 
included food fortification studies from both high-income and low- and 
middle-income countries (LMICs).
DATA COLLECTION AND ANALYSIS: Two review authors independently screened, 
extracted and quality-appraised the data from eligible studies. We carried out 
statistical analysis using Review Manager 5 software. We used random-effects 
meta-analysis for combining data, as the characteristics of study participants 
and interventions differed significantly. We set out the main findings of the 
review in 'Summary of findings' tables, using the GRADE approach.
MAIN RESULTS: We identified 127 studies as relevant through title/abstract 
screening, and included 43 studies (48 papers) with 19,585 participants (17,878 
children) in the review. All the included studies except three compared MMN 
fortification with placebo/no intervention. Two studies compared MMN 
fortification versus iodised salt and one study compared MMN fortification 
versus calcium fortification alone. Thirty-six studies targeted children; 20 
studies were conducted in LMICs. Food vehicles used included staple foods, such 
as rice and flour; dairy products, including milk and yogurt; non-dairy 
beverages; biscuits; spreads; and salt. Fourteen of the studies were fully 
commercially funded, 13 had partial-commercial funding, 14 had non-commercial 
funding and two studies did not specify the source of funding. We rated all the 
evidence as of low to very low quality due to study limitations, imprecision, 
high heterogeneity and small sample size. When compared with placebo/no 
intervention, MMN fortification may reduce anaemia by 32% (risk ratio (RR) 0.68, 
95% confidence interval (CI) 0.56 to 0.84; 11 studies, 3746 participants; 
low-quality evidence), iron deficiency anaemia by 72% (RR 0.28, 95% CI 0.19 to 
0.39; 6 studies, 2189 participants; low-quality evidence), iron deficiency by 
56% (RR 0.44, 95% CI 0.32 to 0.60; 11 studies, 3289 participants; low-quality 
evidence); vitamin A deficiency by 58% (RR 0.42, 95% CI 0.28 to 0.62; 6 studies, 
1482 participants; low-quality evidence), vitamin B2 deficiency by 64% (RR 0.36, 
95% CI 0.19 to 0.68; 1 study, 296 participants; low-quality evidence), vitamin 
B6 deficiency by 91% (RR 0.09, 95% CI 0.02 to 0.38; 2 studies, 301 participants; 
low-quality evidence), vitamin B12 deficiency by 58% (RR 0.42, 95% CI 0.25 to 
0.71; 3 studies, 728 participants; low-quality evidence), weight-for-age 
z-scores (WAZ) (mean difference (MD) 0.1, 95% CI 0.02 to 0.17; 8 studies, 2889 
participants; low-quality evidence) and weight-for-height/length z-score 
(WHZ/WLZ) (MD 0.1, 95% CI 0.02 to 0.18; 6 studies, 1758 participants; 
low-quality evidence). We are uncertain about the effect of MMN fortification on 
zinc deficiency (RR 0.84, 95% CI 0.65 to 1.08; 5 studies, 1490 participants; 
low-quality evidence) and height/length-for-age z-score (HAZ/LAZ) (MD 0.09, 95% 
CI 0.01 to 0.18; 8 studies, 2889 participants; low-quality evidence). Most of 
the studies in this comparison were conducted in children. Subgroup analyses of 
funding sources (commercial versus non-commercial) and duration of intervention 
did not demonstrate any difference in effects, although this was a relatively 
small number of studies and the possible association between commercial funding 
and increased effect estimates has been demonstrated in the wider health 
literature. We could not conduct subgroup analysis by food vehicle and funding; 
since there were too few studies in each subgroup to draw any meaningful 
conclusions. When we compared MMNs versus iodised salt, we are uncertain about 
the effect of MMN fortification on anaemia (R 0.86, 95% CI 0.37 to 2.01; 1 
study, 88 participants; very low-quality evidence), iron deficiency anaemia (RR 
0.40, 95% CI 0.09 to 1.83; 2 studies, 245 participants; very low-quality 
evidence), iron deficiency (RR 0.98, 95% CI 0.82 to 1.17; 1 study, 88 
participants; very low-quality evidence) and vitamin A deficiency (RR 0.19, 95% 
CI 0.07 to 0.55; 2 studies, 363 participants; very low-quality evidence). Both 
of the studies were conducted in children. Only one study conducted in children 
compared MMN fortification versus calcium fortification. None of the primary 
outcomes were reported in the study. None of the included studies reported on 
morbidity, adverse events, all-cause or cause-specific mortality.
AUTHORS' CONCLUSIONS: The evidence from this review suggests that MMN 
fortification when compared to placebo/no intervention may reduce anaemia, iron 
deficiency anaemia and micronutrient deficiencies (iron, vitamin A, vitamin B2 
and vitamin B6). We are uncertain of the effect of MMN fortification on 
anthropometric measures (HAZ/LAZ, WAZ and WHZ/WLZ). There are no data to suggest 
possible adverse effects of MMN fortification, and we could not draw reliable 
conclusions from various subgroup analyses due to a limited number of studies in 
each subgroup. We remain cautious about the level of commercial funding in this 
field, and the possibility that this may be associated with higher effect 
estimates, although subgroup analysis in this review did not demonstrate any 
impact of commercial funding. These findings are subject to study limitations, 
imprecision, high heterogeneity and small sample sizes, and we rated most of the 
evidence low to very low quality. and hence no concrete conclusions could be 
drawn from the findings of this review.

Copyright © 2019 The Cochrane Collaboration. Published by John Wiley & Sons, 
Ltd.

DOI: 10.1002/14651858.CD011400.pub2
PMCID: PMC6917586
PMID: 31849042 [Indexed for MEDLINE]

Conflict of interest statement: JKD: no competing interests. RAS: no competing 
interests. SBM: no competing interests. AM: no competing interests. RK: no 
competing interests. KM: no competing interests. ZL: Participated in a Nestlé 
Nutrition Institute workshop on Health and Nutrition in Adolescents and Young 
Women: preparing for the next generation for the related publication Nestlé 
Nutrition Institute Series Volume 80 (2015) ZAB: Participated in a Nestlé 
Nutrition Institute workshop on Health and Nutrition in Adolescents and Young 
Women: preparing for the next generation and co‐edited the related publication 
Nestlé Nutrition Institute Series Volume 80 (2015). ZAB declares previous travel 
support from the Nestlé Nutrition Institute for attendance at a meeting on 
fortification strategies at the University of Winterthur, Winterthur, 
Switzerland, in October 2011. ZAB received an institutional grant from GAIN on 
fortification program evidence review in 2014. The paper is currently in press.


69. Cochrane Database Syst Rev. 2019 Nov 7;2019(11):CD013145. doi: 
10.1002/14651858.CD013145.pub2.

Human milk-derived fortifier versus bovine milk-derived fortifier for prevention 
of mortality and morbidity in preterm neonates.

Premkumar MH(1), Pammi M(1), Suresh G(1).

Author information:
(1)Baylor College of Medicine, Section of Neonatology, Department of Pediatrics, 
Houston, Texas, USA.

Update of
    doi: 10.1002/14651858.CD013145.

BACKGROUND: Preterm infants who are fed breast milk in comparison to infant 
formula have decreased morbidity such as necrotizing enterocolitis. 
Multi-nutrient fortifiers used to increase the nutritional content of the breast 
milk are commonly derived from bovine milk. Human milk-derived multi-nutrient 
fortifier is now available, but it is not clear if it improves outcomes in 
preterm infants fed with breast milk.
OBJECTIVES: To determine whether the fortification of breast milk feeds with 
human milk-derived fortifier in preterm infants reduces mortality, morbidity, 
and promotes growth and development compared to bovine milk-derived fortifier.
SEARCH METHODS: We searched the following databases for relevant trials in 
September 2018. Cochrane Central Register of Controlled Trials (CENTRAL) (the 
Cochrane Library, Issue 9), electronic journal reference databases including 
MEDLINE (1980 to 20 September 2018), PREMEDLINE, Embase (1974 to 20 September 
2018), CINAHL (1982 to 20 September 2018), biological abstracts in the database 
BIOSIS and conference abstracts from 'Proceedings First' (from 1992 to 2011). We 
also included the following clinical trials registries for ongoing or recently 
completed trials: ClinicalTrials.gov (ClinicalTrials.gov), the World Health 
Organization International Clinical Trials Registry Platform (WHO ICTRP; 
www.whoint/ictrp/search/en/) and the ISRCTN Registry (www.isrctn.com/), and 
abstracts of conferences: proceedings of Pediatric Academic Societies (American 
Pediatric Society, Society for Pediatric Research and European Society for 
Paediatric Research) from 1990 in the 'Pediatric Research' journal and 
'Abstracts online' (2000 to 2017).
SELECTION CRITERIA: We included randomized and quasi-randomized controlled 
trials that compared preterm infants fed breast milk fortified with human 
milk-derived fortifier versus those fed with breast milk fortified with bovine 
milk-derived fortifier.
DATA COLLECTION AND ANALYSIS: The data were collected using the standard methods 
of Cochrane Neonatal. Two authors evaluated trial quality of the studies and 
extracted data. We reported dichotomous data using risk ratios (RRs), risk 
differences (RDs), number needed to treat (NNT) where applicable, and continuous 
data using mean differences (MDs). We assessed the quality of evidence using the 
Grading of Recommendations Assessment, Development and Evaluation (GRADE) 
approach.
MAIN RESULTS: One randomized trial with 127 infants met the eligibility criteria 
and had low risk of bias. Human milk-based fortifier did not decrease the risk 
of necrotizing enterocolitis in exclusively breast milk-fed preterm infants (RR 
0.95, 95% CI 0.2 to 4.54; 1 study, 125 infants, low certainty of evidence). 
Human milk-derived fortifiers did not improve growth, decrease feeding 
intolerance, late-onset sepsis, or death.
AUTHORS' CONCLUSIONS: There is insufficient evidence evaluating human 
milk-derived fortifier with bovine milk-derived fortifier in exclusively breast 
milk-fed preterm infants. Low-certainty evidence from one study suggests that in 
exclusively breast milk-fed preterm infants human milk-derived fortifiers in 
comparison with bovine milk-derived fortifier may not change the risk of 
necrotizing enterocolitis, mortality, feeding intolerance, infection, or improve 
growth. Well-designed randomized controlled trials are needed to evaluate 
short-term and long-term outcomes.

Copyright © 2019 The Cochrane Collaboration. Published by John Wiley & Sons, 
Ltd.

DOI: 10.1002/14651858.CD013145.pub2
PMCID: PMC6837687
PMID: 31697857 [Indexed for MEDLINE]

Conflict of interest statement: Muralidhar H Premkumar has no known conflicts of 
interest relevant to this review.  Mohan Pammi has no known conflicts of 
interest relevant to this review.  Gautham Suresh has no known conflicts of 
interest relevant to this review. The methods section of this review is based on 
a standard template used by Cochrane Neonatal. Core editorial and administrative 
support for this review has been provided by a grant from The Gerber Foundation. 
The Gerber Foundation is a separately endowed, private foundation, distinct from 
the Gerber Products Company. The grantor has no input on the content of the 
review or the editorial process. In order to maintain the utmost editorial 
independence for this Cochrane Review, an editor outside of the Cochrane 
Neonatal core editorial team who is not receiving any financial remuneration 
from the grant, William McGuire, was the Sign‐off Editor for this review. 
Additionally, a Senior Editor from the Cochrane Children and Families Network, 
Robert Boyle, assessed and signed off on this Cochrane Review.


70. Pediatr Res. 2020 Jan;87(2):371-377. doi: 10.1038/s41390-019-0630-2. Epub 2019 
Oct 23.

Nutritional interventions to reduce rates of infection, necrotizing 
enterocolitis and mortality in very preterm infants.

Bührer C(1), Fischer HS(2), Wellmann S(3).

Author information:
(1)Department of Neonatology, Charité - Universitätsmedizin Berlin, Berlin, 
Germany..
(2)Department of Neonatology, Charité - Universitätsmedizin Berlin, Berlin, 
Germany.
(3)Division of Neonatology, University Children's Hospital Regensburg (KUNO), 
University of Regensburg, Regensburg, Germany.

Observational studies demonstrating reduced rates of infections, necrotizing 
enterocolitis (NEC), and mortality in preterm infants fed their own mother's 
milk, as opposed to formula, have prompted endeavors to achieve similar effects 
with the right choice of food and food additives. In a systematic review of 
meta-analyses and randomized controlled trials (RCTs), we considered nutritional 
interventions aimed at reducing the rates of infections, NEC, or mortality in 
very preterm infants. The overall effects of particular interventions were 
presented as risk ratios with 95% confidence intervals. In RCTs, pasteurized 
human donor milk, as opposed to formula, reduced NEC but not infections or 
mortality. No differences emerged between infants receiving human or bovine 
milk-based fortifiers. Pooled data of small trials and a recent large RCT 
suggested that bovine lactoferrin reduced rates of fungal sepsis without impact 
on other infections, NEC, or mortality. Pooled data of RCTs assessing the use of 
prebiotic oligosaccharides found reduced infection but not mortality. Enteral 
L-glutamine (six RCTs) lowered infection rates, and enteral L-arginine (three 
RCTs) reduced NEC. A meta-analysis sensitivity approach found multiple-strain 
(but not single-strain) probiotics to be highly effective in reducing NEC and 
mortality. Thus, selected food components may help to improve outcomes in 
preterm infants.

DOI: 10.1038/s41390-019-0630-2
PMID: 31645057 [Indexed for MEDLINE]


71. Breastfeed Med. 2019 Nov;14(9):648-653. doi: 10.1089/bfm.2019.0100. Epub 2019 
Aug 12.

The Practicality of Feeding Defatted Human Milk in the Treatment of Congenital 
Chylothorax.

Concheiro-Guisan A(1), Alonso-Clemente S(2), Suarez-Albo M(1), Duran-Fernandez 
Feijoo C(1), Fiel-Ozores A(1), Fernandez-Lorenzo JR(3).

Author information:
(1)Neonatology Department and Human Milk Bank, Alvaro Cunqueiro University 
Hospital, Vigo, Spain.
(2)Nutrition Department and Human Milk Bank, Alvaro Cunqueiro University 
Hospital, Vigo, Spain.
(3)Pediatrics Department and Human Milk Bank, Alvaro Cunqueiro University 
Hospital, Vigo, Spain.

Introduction: Congenital chylothorax (CC) is a rare and life-threating 
condition. Since its treatment is founded on the elimination of long-chain fatty 
acids from the diet, breastfeeding has been traditionally contraindicated. 
However, breast milk could be very beneficial due to its immunological and 
nutritional benefits. Only limited research has been published about the usage 
of modified-fat breast milk (MBM) in chylothorax treatment. Methods and Results: 
Systematic review methods were used by two independent reviewers. Only a few 
case report studies (quality assessment on the domains of the GRADE approach), 
two small controlled studies, a retrospective study, and some test-tube-based 
laboratory research met the inclusion criteria. Despite this, we have observed a 
widespread clinical adoption of this novel treatment in health institutions. 
Data suggest that modified-fat breast milk does facilitate the resolution of 
chylothoraces. Refrigerated centrifuge (2°C, 3,000 rpm for 15 minutes) and 
syringe fat removal methods were the most efficient options in terms of fat 
reduction. Conclusions: Feeding of human milk is advisable in CC and feasible by 
means of a simple milk defatting procedure. Open questions remain, related to 
length and degree of fat restriction and need for individualized fortification 
of defatted breast milk.

DOI: 10.1089/bfm.2019.0100
PMID: 31403320 [Indexed for MEDLINE]


72. J Dairy Sci. 2019 Oct;102(10):8587-8603. doi: 10.3168/jds.2018-16046. Epub 2019 
Aug 7.

Is vitamin D-fortified yogurt a value-added strategy for improving human health? 
A systematic review and meta-analysis of randomized trials.

Gasparri C(1), Perna S(2), Spadaccini D(3), Alalwan T(2), Girometta C(4), 
Infantino V(5), Rondanelli M(6).

Author information:
(1)Department of Public Health, Experimental and Forensic Medicine, Section of 
Human Nutrition, Endocrinology and Nutrition Unit, Azienda di Servizi alla 
Persona, University of Pavia, 27100 Pavia, Italy. Electronic address: 
.
(2)Department of Biology, College of Science, University of Bahrain, Sakhir 
Campus, PO Box 32038, Kingdom of Bahrain.
(3)Department of Public Health, Experimental and Forensic Medicine, Section of 
Human Nutrition, Endocrinology and Nutrition Unit, Azienda di Servizi alla 
Persona, University of Pavia, 27100 Pavia, Italy.
(4)Department of Earth and Environmental Sciences, Mycology and Plant Pathology 
Laboratory, University of Pavia, 27100 Pavia, Italy.
(5)Department of Biomedical Sciences and Human Oncology, University of Bari Aldo 
Moro, 70121 Bari, Italy.
(6)IRCCS Mondino Foundation, 27100 Pavia, Italy; Department of Public Health, 
Experimental and Forensic Medicine, Unit of Human and Clinical Nutrition, 
University of Pavia, 27100 Pavia, Italy.

Yogurt is a good source of probiotics, calcium, and proteins, but its content of 
vitamin D is low. Therefore, yogurt could be a good choice for vitamin D 
fortification to improve the positive health outcomes associated with its 
consumption. The primary aim of this systematic review and meta-analysis was to 
investigate the effect of vitamin D-fortified yogurt compared with plain yogurt 
on levels of serum 25-hydroxy vitamin D (25OHD). The secondary aim was to 
evaluate the effect of fortified yogurt on parathyroid hormone, anthropometric 
parameters, blood pressure, glucose metabolism, and lipid profile. We searched 
PubMed, Scopus, and Google Scholar for eligible studies; that is, randomized 
controlled trials (RCT) that compared vitamin D-fortified yogurt with control 
treatment without any additional supplement. Random-effects models were used to 
estimate pooled effect sizes and 95% confidence intervals. Findings from 9 RCT 
(n = 665 participants) that lasted from 8 to 16 wk are summarized in this 
review. The meta-analyzed mean differences for random effects showed that 
vitamin D-fortified yogurt (from 400 to 2,000 IU) increased serum 25OHD by 31.00 
nmol/L. In addition, vitamin D-fortified yogurt decreased parathyroid hormone by 
15.47 ng/L, body weight by 0.92 kg, waist circumference by 2.01 cm, HOMA-IR by 
2.18 mass units, fasting serum glucose by 22.54 mg/dL, total cholesterol by 
13.38 mg/dL, and triglycerides by 30.12 mg/dL compared with the controlled 
treatments. No publication bias was identified. Considerable between-study 
heterogeneity was observed for most outcomes. Vitamin D-fortified yogurt may be 
beneficial in improving serum 25OHD, lipid profile, glucose metabolism, and 
anthropometric parameters and decreasing parathyroid hormone level in pregnant 
women and adult and elderly subjects with or without diabetes, prediabetes, or 
metabolic syndrome.

Copyright © 2019 American Dairy Science Association. Published by Elsevier Inc. 
All rights reserved.

DOI: 10.3168/jds.2018-16046
PMID: 31400903 [Indexed for MEDLINE]


73. Neonatology. 2020;117(1):24-32. doi: 10.1159/000501279. Epub 2019 Jul 19.

Early versus Delayed Fortification of Human Milk in Preterm Infants: A 
Systematic Review.

Alyahya W(1), Simpson J(2)(3), Garcia AL(4), Mactier H(5), Edwards CA(4).

Author information:
(1)School of Medicine, Dentistry and Nursing, Human Nutrition, University of 
Glasgow, Glasgow, United Kingdom,.
(2)Neonatal Unit, Royal Hospital for Children, Glasgow, United Kingdom.
(3)NHS Greater Glasgow and Clyde Human Milk Bank, Queen Elizabeth University 
Hospital, Glasgow, United Kingdom.
(4)School of Medicine, Dentistry and Nursing, Human Nutrition, University of 
Glasgow, Glasgow, United Kingdom.
(5)Neonatal Unit, Princess Royal Maternity Hospital, Glasgow, United Kingdom.

Expressed breast milk (EBM) is commonly supplemented with commercially prepared 
human milk fortifier to meet the additional nutritional needs of preterm 
infants. The optimal milk intake at which to introduce fortification is unknown. 
The objective of this systematic review was to compare the effect of early 
fortification (EF) versus that of delayed introduction of human milk fortifier 
(DF) on short-term outcomes including growth, feeding intolerance, length of 
hospital stay, and maturity at discharge in very-low-birth-weight infants. The 
search was carried out until March 2019 using 5 electronic databases (PubMed, 
Ovid Medline, Web of Science, Ovid Embase, and the Cochrane Library). The search 
was supplemented with a search of the clinical trial registry and reference 
lists. Eligible studies involved randomized controlled trials that had been 
designed to compare EF against DF using multi-nutrient fortifier for infants of 
a birth weight of <1,500 g who were fed exclusively or predominantly EBM. Four 
authors independently screened the studies for eligibility. A total of 1,972 
articles were screened; 2 studies met the inclusion criteria and were included 
with a total number of participants of 171. The definition of EF and DF was not 
consistent between the 2 studies. There was no significant impact of EF versus 
DF on all outcomes. In conclusion, current data are limited and do not provide 
evidence on the optimal time to start fortification. The definition of EF and DF 
needs to be agreed upon and further larger randomized controlled trials are 
required.

© 2019 S. Karger AG, Basel.

DOI: 10.1159/000501279
PMID: 31326969 [Indexed for MEDLINE]


74. Cochrane Database Syst Rev. 2019 Jul 19;7(7):CD002971. doi: 
10.1002/14651858.CD002971.pub5.

Formula versus donor breast milk for feeding preterm or low birth weight 
infants.

Quigley M(1), Embleton ND, McGuire W.

Author information:
(1)National Perinatal Epidemiology Unit, University of Oxford, Old Road Campus, 
Oxford, UK, 0X3 7LF.

Update in
    Cochrane Database Syst Rev. 2024 Sep 6;9:CD002971. doi: 
10.1002/14651858.CD002971.pub6.

Comment in
    Arch Dis Child Educ Pract Ed. 2020 Dec;105(6):381-382. doi: 
10.1136/archdischild-2019-318459.

Update of
    Cochrane Database Syst Rev. 2018 Jun 20;6:CD002971. doi: 
10.1002/14651858.CD002971.pub4.

BACKGROUND: When sufficient maternal breast milk is not available, alternative 
forms of enteral nutrition for preterm or low birth weight (LBW) infants are 
donor breast milk or artificial formula. Donor breast milk may retain some of 
the non-nutritive benefits of maternal breast milk for preterm or LBW infants. 
However, feeding with artificial formula may ensure more consistent delivery of 
greater amounts of nutrients. Uncertainty exists about the balance of risks and 
benefits of feeding formula versus donor breast milk for preterm or LBW infants.
OBJECTIVES: To determine the effect of feeding with formula compared with donor 
breast milk on growth and development in preterm or low birth weight (LBW) 
infants.
SEARCH METHODS: We used the Cochrane Neonatal search strategy, including 
electronic searches of the Cochrane Central Register of Controlled Trials 
(CENTRAL; 2019, Issue 5), Ovid MEDLINE, Embase, and the Cumulative Index to 
Nursing and Allied Health Literature (3 May 2019), as well as conference 
proceedings, previous reviews, and clinical trials.
SELECTION CRITERIA: Randomised or quasi-randomised controlled trials (RCTs) 
comparing feeding with formula versus donor breast milk in preterm or LBW 
infants.
DATA COLLECTION AND ANALYSIS: Two review authors assessed trial eligibility and 
risk of bias and extracted data independently. We analysed treatment effects as 
described in the individual trials and reported risk ratios (RRs) and risk 
differences (RDs) for dichotomous data, and mean differences (MDs) for 
continuous data, with respective 95% confidence intervals (CIs). We used a 
fixed-effect model in meta-analyses and explored potential causes of 
heterogeneity in subgroup analyses. We assessed the certainty of evidence for 
the main comparison at the outcome level using GRADE methods.
MAIN RESULTS: Twelve trials with a total of 1879 infants fulfilled the inclusion 
criteria. Four trials compared standard term formula versus donor breast milk 
and eight compared nutrient-enriched preterm formula versus donor breast milk. 
Only the five most recent trials used nutrient-fortified donor breast milk. The 
trials contain various weaknesses in methodological quality, specifically 
concerns about allocation concealment in four trials and lack of blinding in 
most of the trials. Most of the included trials were funded by companies that 
made the study formula.Formula-fed infants had higher in-hospital rates of 
weight gain (mean difference (MD) 2.51, 95% confidence interval (CI) 1.93 to 
3.08 g/kg/day), linear growth (MD 1.21, 95% CI 0.77 to 1.65 mm/week) and head 
growth (MD 0.85, 95% CI 0.47 to 1.23 mm/week). These meta-analyses contained 
high levels of heterogeneity. We did not find evidence of an effect on long-term 
growth or neurodevelopment. Formula feeding increased the risk of necrotising 
enterocolitis (typical risk ratio (RR) 1.87, 95% CI 1.23 to 2.85; risk 
difference (RD) 0.03, 95% CI 0.01 to 0.05; number needed to treat for an 
additional harmful outcome (NNTH) 33, 95% CI 20 to 100; 9 studies, 1675 
infants).The GRADE certainty of evidence was moderate for rates of weight gain, 
linear growth, and head growth (downgraded for high levels of heterogeneity) and 
was moderate for neurodevelopmental disability, all-cause mortality, and 
necrotising enterocolitis (downgraded for imprecision).
AUTHORS' CONCLUSIONS: In preterm and LBW infants, moderate-certainty evidence 
indicates that feeding with formula compared with donor breast milk, either as a 
supplement to maternal expressed breast milk or as a sole diet, results in 
higher rates of weight gain, linear growth, and head growth and a higher risk of 
developing necrotising enterocolitis. The trial data do not show an effect on 
all-cause mortality, or on long-term growth or neurodevelopment.

DOI: 10.1002/14651858.CD002971.pub5
PMCID: PMC6640412
PMID: 31322731 [Indexed for MEDLINE]

Conflict of interest statement: MQ: nothing to declare. NDE declares the 
following: receiving a research grant award for an RCT of breast milk products 
by Prolacta Bioscience, 2017; receiving a grant from Danone Early Life Nutrition 
to support a study on feeding in late and moderately preterm infants, 2018; 
receiving a grant from Nestle Nutrition for transcriptomic analyses of gut 
tissue, 2016; lectures with Wyeth Nutrition in 2017, Nestle Nutrition Institute 
in 2017 and 2018, Philipps in 2017, and Fresenius, 2017. WM: nothing to declare. 
Core editorial and administrative support for this review has been provided by a 
grant from The Gerber Foundation. The Gerber Foundation is a separately endowed, 
private foundation, distinct from the Gerber Products Company. The grantor has 
no input on the content of the review or the editorial process. In order to 
maintain the utmost editorial independence for this Cochrane Review, an editor 
outside of the Cochrane Neonatal core editorial team who is not receiving any 
financial remuneration from the grant, Mohan Pammi, was the Sign‐off Editor for 
this review. Additionally, a Senior Editor from the Cochrane Children and 
Families Network, Robert Boyle, assessed and signed off on this Cochrane Review.


75. Adv Nutr. 2019 May 1;10(suppl_2):S251-S271. doi: 10.1093/advances/nmz001.

Role of Functional Fortified Dairy Products in Cardiometabolic Health: A 
Systematic Review and Meta-analyses of Randomized Clinical Trials.

Soto-Méndez MJ(1), Rangel-Huerta OD(2), Ruiz-López MD(1)(3)(4), Martínez de 
Victoria E(1)(5)(4), Anguita-Ruiz A(4)(6)(7), Gil A(1)(8)(4)(6)(7).

Author information:
(1)Iberoamerican Nutrition Foundation -FINUT-, Granada, Spain.
(2)Department of Nutrition, Institute of Basic Medical Sciences, University of 
Oslo, Oslo, Norway.
(3)Department of Nutrition and Food Sciences, School of Pharmacy.
(4)Institute of Nutrition and Food Technology "José Mataix," Biomedical Research 
Center, University of Granada, Granada, Spain.
(5)Department of Physiology, School of Sciences.
(6)Biomedical Research Institute ibs GRANADA, University Hospital Complex in 
Granada, Granada, Spain.
(7)CIBEROBN (CIBER Physiopathology of Obesity and Nutrition), Instituto de Salud 
Carlos III, Madrid, Spain.
(8)Department of Biochemistry and Molecular Biology II, School of Pharmacy, 
University of Granada, Granada, Spain.

There is insufficient evidence on the role of functional fortified dairy 
products in improving health and in preventing risk factors associated with 
noncommunicable chronic diseases. This systematic review was conducted to 
summarize effects of the consumption of fortified dairy products on biomarkers 
of cardiometabolic risk. MEDLINE and SCOPUS databases were used to perform 
searches to include studies published up to 30 April 2018. Randomized clinical 
trials with human subjects consuming dairy products fortified with phytosterols, 
FAs, vitamins or minerals and relating this consumption with cardiometabolic 
health were included in this review. Risk of bias assessment according to 
Cochrane guidelines was performed to determine the quality of the trials. 
Forty-one studies were finally selected for this synthesis; the selected studies 
tested dairy products fortified with the following nutrients and bioactive 
components: phytosterols (n = 31), FAs (n = 8), and vitamin D (n = 2). We found 
that the consumption of phytosterol-fortified dairy, led to an overall LDL 
cholesterol reduction of -0.36 (-0.41, -0.31) mmol/L, P < 0.001; this decrease 
was mainly related to the dosage. Likewise, consumption of ω-3 FA-fortified 
dairy products resulted in a plasma LDL cholesterol reduction of -0.18 (-0.27, 
-0.09) mmol/L as well as a decrease of -0.18 (-0.32, -0.05) mmol/L in 
triacylglycerols (TG). Performing meta-analyses of the consumption of dairy 
products fortified with vitamin D or FAs other than ω-3 FAs and biomarkers of 
cardiometabolic risk was not possible because of the few available publications. 
Our results indicate that consumption of dairy products fortified with 
phytosterols and ω-3 FAs can lead to a reduction of LDL cholesterol and 
consumption of fortified dairy products fortified with ω-3 FAs can reduce TG 
concentration. However, more studies with homogeneous designs are needed to 
determine the advantages of using dairy products as fortification vehicles to 
prevent cardiometabolic risk.

Copyright © American Society for Nutrition 2019.

DOI: 10.1093/advances/nmz001
PMCID: PMC6518140
PMID: 31089744 [Indexed for MEDLINE]


76. Cochrane Database Syst Rev. 2019 May 10;5(5):CD010068. doi: 
10.1002/14651858.CD010068.pub2.

Fortification of staple foods with vitamin A for vitamin A deficiency.

Hombali AS(1), Solon JA, Venkatesh BT, Nair NS, Peña-Rosas JP.

Author information:
(1)Department of Research, Institute of Mental Health, Block 7, Buangkok View, 
Buangkok Green Medical Park, Singapore, Singapore, 539747.

Update of
    doi: 10.1002/14651858.CD010068.

BACKGROUND: Vitamin A deficiency is a significant public health problem in many 
low- and middle-income countries, especially affecting young children, women of 
reproductive age, and pregnant women. Fortification of staple foods with vitamin 
A has been used to increase vitamin A consumption among these groups.
OBJECTIVES: To assess the effects of fortifying staple foods with vitamin A for 
reducing vitamin A deficiency and improving health-related outcomes in the 
general population older than two years of age.
SEARCH METHODS: We searched the following international databases with no 
language or date restrictions: Cochrane Central Register of Controlled Trials 
(CENTRAL; 2018, Issue 6) in the Cochrane Library; MEDLINE and MEDLINE In Process 
OVID; Embase OVID; CINAHL Ebsco; Web of Science (ISI) SCI, SSCI, CPCI-exp and 
CPCI-SSH; BIOSIS (ISI); POPLINE; Bibliomap; TRoPHI; ASSIA (Proquest); IBECS; 
SCIELO; Global Index Medicus - AFRO and EMRO; LILACS; PAHO; WHOLIS; WPRO; 
IMSEAR; IndMED; and Native Health Research Database. We also searched 
clinicaltrials.gov and the International Clinical Trials Registry Platform to 
identify ongoing and unpublished studies. The date of the last search was 19 
July 2018.
SELECTION CRITERIA: We included individually or cluster-randomised controlled 
trials (RCTs) in this review. The intervention included fortification of staple 
foods (sugar, edible oils, edible fats, maize flour or corn meal, wheat flour, 
milk and dairy products, and condiments and seasonings) with vitamin A alone or 
in combination with other vitamins and minerals. We included the general 
population older than two years of age (including pregnant and lactating women) 
from any country.
DATA COLLECTION AND ANALYSIS: Two authors independently screened and assessed 
eligibility of studies for inclusion, extracted data from included studies and 
assessed their risk of bias. We used standard Cochrane methodology to carry out 
the review.
MAIN RESULTS: We included 10 randomised controlled trials involving 4455 
participants. All the studies were conducted in low- and upper-middle income 
countries where vitamin A deficiency was a public health issue. One of the 
included trials did not contribute data to the outcomes of interest.Three trials 
compared provision of staple foods fortified with vitamin A versus unfortified 
staple food, five trials compared provision of staple foods fortified with 
vitamin A plus other micronutrients versus unfortified staple foods, and two 
trials compared provision of staple foods fortified with vitamin A plus other 
micronutrients versus no intervention. No studies compared staple foods 
fortified with vitamin A alone versus no intervention.The duration of 
interventions ranged from three to nine months. We assessed six studies at high 
risk of bias overall. Government organisations, non-governmental organisations, 
the private sector, and academic institutions funded the included studies; 
funding source does not appear to have distorted the results.Staple food 
fortified with vitamin A versus unfortified staple food We are uncertain whether 
fortifying staple foods with vitamin A alone makes little or no difference for 
serum retinol concentration (mean difference (MD) 0.03 μmol/L, 95% CI -0.06 to 
0.12; 3 studies, 1829 participants; I² = 90%, very low-certainty evidence). It 
is uncertain whether vitamin A alone reduces the risk of subclinical vitamin A 
deficiency (risk ratio (RR) 0.45, 95% CI 0.19 to 1.05; 2 studies; 993 
participants; I² = 33%, very low-certainty evidence). The certainty of the 
evidence was mainly affected by risk of bias, imprecision and inconsistency.It 
is uncertain whether vitamin A fortification reduces clinical vitamin A 
deficiency, defined as night blindness (RR 0.11, 95% CI 0.01 to 1.98; 1 study, 
581 participants, very low-certainty evidence). The certainty of the evidence 
was mainly affected by imprecision, inconsistency, and risk of bias.Staple foods 
fortified with vitamin A versus no intervention No studies provided data for 
this comparison.Staple foods fortified with vitamin A plus other micronutrients 
versus same unfortified staple foods Fortifying staple foods with vitamin A plus 
other micronutrients may not increase the serum retinol concentration (MD 0.08 
μmol/L, 95% CI -0.06 to 0.22; 4 studies; 1009 participants; I² = 95%, 
low-certainty evidence). The certainty of the evidence was mainly affected by 
serious inconsistency and risk of bias.In comparison to unfortified staple 
foods, fortification with vitamin A plus other micronutrients probably reduces 
the risk of subclinical vitamin A deficiency (RR 0.27, 95% CI 0.16 to 0.49; 3 
studies; 923 participants; I² = 0%; moderate-certainty evidence). The certainty 
of the evidence was mainly affected by serious risk of bias.Staple foods 
fortified with vitamin A plus other micronutrients versus no 
interventionFortification of staple foods with vitamin A plus other 
micronutrients may increase serum retinol concentration (MD 0.22 μmol/L, 95% CI 
0.15 to 0.30; 2 studies; 318 participants; I² = 0%; low-certainty evidence). 
When compared to no intervention, it is uncertain whether the intervention 
reduces the risk of subclinical vitamin A deficiency (RR 0.71, 95% CI 0.52 to 
0.98; 2 studies; 318 participants; I² = 0%; very low-certainty evidence) . The 
certainty of the evidence was affected mainly by serious imprecision and risk of 
bias.No trials reported on the outcomes of all-cause morbidity, all-cause 
mortality, adverse effects, food intake, congenital anomalies (for pregnant 
women), or breast milk concentration (for lactating women).
AUTHORS' CONCLUSIONS: Fortifying staple foods with vitamin A alone may make 
little or no difference to serum retinol concentrations or the risk of 
subclinical vitamin A deficiency. In comparison with provision of unfortified 
foods, provision of staple foods fortified with vitamin A plus other 
micronutrients may not increase serum retinol concentration but probably reduces 
the risk of subclinical vitamin A deficiency.Compared to no intervention, staple 
foods fortified with vitamin A plus other micronutrients may increase serum 
retinol concentration, although it is uncertain whether the intervention reduces 
the risk of subclinical vitamin A deficiency as the certainty of the evidence 
has been assessed as very low.It was not possible to estimate the effect of 
staple food fortification on outcomes such as mortality, morbidity, adverse 
effects, congenital anomalies, or breast milk vitamin A, as no trials included 
these outcomes.The type of funding source for the studies did not appear to 
distort the results from the analysis.

DOI: 10.1002/14651858.CD010068.pub2
PMCID: PMC6509778
PMID: 31074495 [Indexed for MEDLINE]

Conflict of interest statement: Aditi S Hombali ‐ none. Bhumika T Venkatesh ‐ 
none. Sreekumar Nair ‐ none. Juan Antonio Solon works for the Nutrition Center 
of the Philippines, a non‐governmental organisation working in the field of 
public health nutrition. The Nutrition Center of the Philippines has several 
studies in the field of food fortification. Two of these studies are included in 
the review; one was excluded. Dr Solon is not an author in any of the included 
studies, and he did not participate in the screening and eligibility of the 
included studies. Juan Pablo Peña‐Rosas is a full‐time WHO staff member. The 
Department of Nutrition for Health and Development, World Health Organization 
receives financial resources from several external sources including the Bill & 
Melinda Gates Foundation (2013‐2019), US Centers for Disease Control and 
Prevention (CDC) (2014‐2019), Nutrition International (2014‐2019), and USAID 
(2014‐2019). Donors do not fund specific guidelines and do not participate in 
any decision related to the guideline development process including the 
composition of policy questions, membership of the guideline groups, the conduct 
and interpretation of systematic reviews, or the formulation of recommendations.


77. Types and Amounts of Complementary Foods and Beverages and Growth, Size, and 
Body Composition: A Systematic Review [Internet].

English LK(1), Obbagy JE(2), Wong YP(3), Psota TL(2), Nadaud P(1), Johns K(1), 
Terry N(4), Butte NF(5), Dewey KG(6), Fleischer DM(7), Fox MK(8), Greer FR(9), 
Krebs NF(10), Scanlon KS(11), Casavale KO(12), Spahn JM(13), Stoody E(13).

Alexandria (VA): USDA Nutrition Evidence Systematic Review; 2019 Apr.
USDA Nutrition Evidence Systematic Reviews.

Author information:
(1)Systematic review analyst, Nutrition Evidence Systematic Review (NESR) team; 
Panum Group under contract with the Food and Nutrition Service (FNS), U.S. 
Department of Agriculture (USDA)
(2)Systematic review analyst, NESR team; Office of Nutrition Guidance and 
Analysis (ONGA), Center for Nutrition Policy and Promotion (CNPP), FNS, USDA
(3)Systematic review librarian, NESR team; ONGA, CNPP, FNS, USDA
(4)Biomedical librarian, NESR team; National Institutes of Health Library, U.S. 
Department of Health and Human Services (HHS)
(5)Member, Complementary Feeding Technical Expert Collaborative, Pregnancy and 
Birth to 24 Months Project; Baylor College of Medicine, Emeritus
(6)Member, Complementary Feeding Technical Expert Collaborative, Pregnancy and 
Birth to 24 Months Project; University of California, Davis
(7)Member, Complementary Feeding Technical Expert Collaborative, Pregnancy and 
Birth to 24 Months Project; University of Colorado Denver School of Medicine, 
Children’s Hospital Colorado
(8)Member, Complementary Feeding Technical Expert Collaborative, Pregnancy and 
Birth to 24 Months Project; Mathematica Policy Research
(9)Member, Complementary Feeding Technical Expert Collaborative, Pregnancy and 
Birth to 24 Months Project; University of Wisconsin, Madison, Emeritus
(10)Member, Complementary Feeding Technical Expert Collaborative, Pregnancy and 
Birth to 24 Months Project; University of Colorado School of Medicine, 
Department of Pediatrics
(11)Member and Federal Expert Group Liaison, Complementary Feeding Technical 
Expert Collaborative, Pregnancy and Birth to 24 Months Project; Office of Policy 
Support, FNS, USDA
(12)Project Lead, Pregnancy and Birth to 24 Months Project; Office of Disease 
Prevention and Health Promotion, HHS
(13)Project Lead, Pregnancy and Birth to 24 Months Project; NESR team, ONGA, 
CNPP, FNS, USDA

BACKGROUND: The goal of this systematic review was to examine the following 
question: What is the relationship between types and amounts of complementary 
foods and beverages (CFB) consumed and growth, size, and body composition? 
Systematic reviews were conducted as part of the U.S. Department of Agriculture 
and Department of Health and Human Services Pregnancy and Birth to 24 Months 
Project. Complementary feeding is the process that starts when human milk or 
infant formula is complemented by other foods and beverages, beginning during 
infancy and typically continuing to 24 months of age. CFB were defined as foods 
and/or beverages other than human milk or infant formula (liquids, semisolids, 
and solids) provided to an infant or young child to provide nutrients and 
energy.
CONCLUSION STATEMENT: Moderate evidence indicates that higher versus lower meat 
intake or meat versus iron-fortified cereal intake over a short duration (~3mo) 
during the complementary feeding period does not favorably or unfavorably 
influence growth, size, and/or body composition. There is insufficient evidence 
to determine a relationship between meat intake and prevalence/incidence of 
overweight or obesity. Limited evidence suggests that type or amount of cereal 
given does not favorably or unfavorably influence growth, size, body 
composition, and/or prevalence/incidence of overweight or obesity. Moderate 
evidence suggests that consumption of complementary foods with different fats 
and/or fatty acid composition does not favorably or unfavorably influence 
growth, size, or body composition. There is not enough evidence to determine a 
relationship between consumption of complementary foods with different fats 
and/or fatty acid composition and/or prevalence/incidence of overweight or 
obesity. Limited evidence suggests that sugar sweetened beverage consumption 
during the complementary feeding period is associated with increased risk of 
obesity in childhood, but is not associated with other measures of growth, size, 
and body composition. Limited evidence showed a positive association between 
juice intake and infant weight-for-length and child BMI z-scores. No conclusion 
could be made about the relationship between other complementary foods 
(vegetables, fruit, dairy products and/or cow’s milk, cereal-based products, 
milk-cereal drink, and/or categories such as “ready-made foods”) and growth, 
size, body composition, and/or prevalence/incidence of overweight or obesity. No 
conclusion could be made about the relationship between distinct dietary 
patterns during the complementary feeding period and growth, size, body 
composition, and/or prevalence/incidence of malnutrition, overweight or obesity. 
Grades: Moderate - Meat; Different Fats and/or Fatty Acid Composition; Limited - 
Juice; Sugar-Sweetened Beverages; Grade Not Assignable - Cereal; Other CFB; 
Dietary Patterns.
METHODS: This systematic review was conducted by a team of staff from the 
Nutrition Evidence Systematic Review in collaboration with a Technical Expert 
Collaborative. Literature search was conducted using 4 databases (PubMed, 
Cochrane, Embase, and CINAHL) to identify articles that evaluated the 
intervention or exposure of types and amounts of CFB consumed and the outcomes 
of growth, size, and/or body composition. A manual search was conducted to 
identify articles that may not have been included in the electronic databases 
searched. Articles were screened by two analysts independently for inclusion 
based on pre-determined criteria. Data extraction and risk of bias assessment 
were conducted for each included study, and both were checked for accuracy. The 
body of evidence was qualitatively synthesized to inform development of a 
conclusion statement(s), and the strength of evidence was graded using 
pre-established criteria evaluating the body of evidence on risk of bias, 
adequacy, consistency, impact, and generalizability.
SUMMARY OF THE EVIDENCE: This review includes 49 articles from 18 randomized 
controlled trials, 1 non-randomized controlled trial, and 30 prospective cohort 
studies. The studies varied in terms of the types and/or amounts of CFB 
examined, which included dietary patterns consumed during the complementary 
feeding period, meat, cereal, foods with different fatty acid composition, 
sugar-sweetened beverages, juice or 100% juice, and other CFB. Gaps and 
limitations in the evidence include the need for randomized controlled trials 
and studies that examine a wider range of specific types and amounts 
complementary foods and beverages, account for the rationale for type/amount of 
complementary foods and beverages given (e.g., reverse causality), and adjust 
for potential confounders (e.g., human milk and/or formula-feeding and baseline 
growth status).

DOI: 10.52570/NESR.PB242018.SR0306
PMID: 35901225


78. Timing of Introduction of Complementary Foods and Beverages and Bone Health: A 
Systematic Review [Internet].

Obbagy JE(1), English LK(2), Psota TL(1), Nadaud P(2), Johns K(2), Wong YP(3), 
Terry N(4), Butte NF(5), Dewey KG(6), Fleischer DM(7), Fox MK(8), Greer FR(9), 
Krebs NF(10), Scanlon KS(11), Casavale KO(12), Spahn JM(13), Stoody E(13).

Alexandria (VA): USDA Nutrition Evidence Systematic Review; 2019 Apr.
USDA Nutrition Evidence Systematic Reviews.

Author information:
(1)Systematic review analyst, Nutrition Evidence Systematic Review (NESR) team; 
Office of Nutrition Guidance and Analysis (ONGA), Center for Nutrition Policy 
and Promotion (CNPP), Food and Nutrition Service (FNS), U.S. Department of 
Agriculture (USDA)
(2)Systematic review analyst, NESR team; Panum Group under contract with FNS, 
USDA
(3)Systematic review librarian, NESR team; ONGA, CNPP, FNS, USDA
(4)Biomedical librarian, NESR team; National Institutes of Health Library, U.S. 
Department of Health and Human Services (HHS)
(5)Member, Complementary Feeding Technical Expert Collaborative, Pregnancy and 
Birth to 24 Months Project; Baylor College of Medicine, Emeritus
(6)Member, Complementary Feeding Technical Expert Collaborative, Pregnancy and 
Birth to 24 Months Project; University of California, Davis
(7)Member, Complementary Feeding Technical Expert Collaborative, Pregnancy and 
Birth to 24 Months Project; University of Colorado Denver School of Medicine, 
Children’s Hospital Colorado
(8)Member, Complementary Feeding Technical Expert Collaborative, Pregnancy and 
Birth to 24 Months Project; Mathematica Policy Research
(9)Member, Complementary Feeding Technical Expert Collaborative, Pregnancy and 
Birth to 24 Months Project; University of Wisconsin, Madison, Emeritus
(10)Member, Complementary Feeding Technical Expert Collaborative, Pregnancy and 
Birth to 24 Months Project; University of Colorado School of Medicine, 
Department of Pediatrics
(11)Member and Federal Expert Group Liaison, Complementary Feeding Technical 
Expert Collaborative, Pregnancy and Birth to 24 Months Project; Office of Policy 
Support, FNS, USDA
(12)Project Lead, Pregnancy and Birth to 24 Months Project; Office of Disease 
Prevention and Health Promotion, HHS
(13)Project Lead, Pregnancy and Birth to 24 Months Project; NESR team, ONGA, 
CNPP, FNS, USDA

BACKGROUND: Complementary feeding is the process that starts when human milk or 
infant formula is complemented by other foods and beverages, beginning during 
infancy and typically continuing to 24 months of age. This systematic review was 
conducted by the Nutrition Evidence Systematic Review (NESR) team as part of the 
U.S. Department of Agriculture and Department of Health and Human Services 
Pregnancy and Birth to 24 Months Project. The goal of this systematic review was 
to answer the following research question: What is the relationship between 
timing of introduction of complementary foods and beverages and bone health?
CONCLUSION STATEMENT AND GRADES: Insufficient evidence is available to draw 
conclusions about the relationship between the timing of introduction of 
complementary foods and beverages and bone health. Grade: [Grade Not 
Assignable].
METHODS: This systematic review was conducted by a team of staff from NESR in 
collaboration with a Technical Expert Collaborative. A literature search was 
conducted using 4 databases (CINAHL, Cochrane, Embase, and PubMed) to identify 
articles published from January 1980 to July 2016 that examined the age when 
complementary foods and beverages (CFB) were first introduced and bone health. 
CFB were defined as foods and beverages other than human milk or infant formula 
provided to an infant or young child. Bone health outcomes included bone, 
biomarkers of bone metabolism, and incidence and prevalence of osteoporosis, 
osteopenia, rickets, and fracture. A manual search was done to identify articles 
that may not have been included in the electronic databases searched. Articles 
were screened in a dual manner, independently by 2 NESR analysts, to determine 
which articles met predetermined criteria for inclusion. Data from each included 
article were extracted, risk of bias was assessed. The body of evidence was 
qualitatively synthesized, a conclusion statement was developed and the strength 
of the evidence (grade) was assessed using pre-established criteria including 
evaluation of the internal validity/risk of bias, adequacy, consistency, impact, 
and generalizability of available evidence. Research recommendations were 
identified.
SUMMARY OF EVIDENCE: Three studies met the inclusion criteria for this 
systematic review, one randomized controlled trial, one prospective cohort 
study, and one case-control study. The randomized controlled trial included a 
small sample of infants fed only vitamin D fortified infant formula, and did not 
follow infants long enough to assess the impact of timing of introduction to CFB 
and bone health, as outcomes were assessed when infants were 26 weeks (wk) of 
age. The observational studies did not measure and/or account for a number 
confounding factors that could have impacted the relationship between timing of 
CFB introduction and bone health later in childhood. The ability to draw 
conclusions about the relationship between the timing of CFB introduction and 
bone health is limited by an overall lack of research, as well as heterogeneity 
in the three studies included in this systematic review with regards to 
methodology, subject populations, and results.

DOI: 10.52570/NESR.PB242018.SR0309
PMID: 35853107


79. Types and Amounts of Complementary Foods and Beverages and Developmental 
Milestones: A Systematic Review [Internet].

English LK(1), Obbagy JE(2), Wong YP(3), Psota TL(2), Nadaud P(1), Johns K(1), 
Terry N(4), Butte NF(5), Dewey KG(6), Fleischer DM(7), Fox MK(8), Greer FR(9), 
Krebs NF(10), Scanlon KS(11), Casavale KO(12), Spahn JM(13), Stoody E(13).

Alexandria (VA): USDA Nutrition Evidence Systematic Review; 2019 Apr.
USDA Nutrition Evidence Systematic Reviews.

Author information:
(1)Systematic review analyst, Nutrition Evidence Systematic Review (NESR) team; 
Panum Group under contract with the Food and Nutrition Service (FNS), U.S. 
Department of Agriculture (USDA)
(2)Systematic review analyst, NESR team; Office of Nutrition Guidance and 
Analysis (ONGA), Center for Nutrition Policy and Promotion (CNPP), FNS, USDA
(3)Systematic review librarian, NESR team; ONGA, CNPP, FNS, USDA
(4)Biomedical librarian, NESR team; National Institutes of Health Library, U.S. 
Department of Health and Human Services (HHS)
(5)Member, Complementary Feeding Technical Expert Collaborative, Pregnancy and 
Birth to 24 Months Project; Baylor College of Medicine, Emeritus
(6)Member, Complementary Feeding Technical Expert Collaborative, Pregnancy and 
Birth to 24 Months Project; University of California, Davis
(7)Member, Complementary Feeding Technical Expert Collaborative, Pregnancy and 
Birth to 24 Months Project; University of Colorado Denver School of Medicine, 
Children’s Hospital Colorado
(8)Member, Complementary Feeding Technical Expert Collaborative, Pregnancy and 
Birth to 24 Months Project; Mathematica Policy Research
(9)Member, Complementary Feeding Technical Expert Collaborative, Pregnancy and 
Birth to 24 Months Project; University of Wisconsin, Madison, Emeritus
(10)Member, Complementary Feeding Technical Expert Collaborative, Pregnancy and 
Birth to 24 Months Project; University of Colorado School of Medicine, 
Department of Pediatrics
(11)Member and Federal Expert Group Liaison, Complementary Feeding Technical 
Expert Collaborative, Pregnancy and Birth to 24 Months Project; Office of Policy 
Support, FNS, USDA
(12)Project Lead, Pregnancy and Birth to 24 Months Project; Office of Disease 
Prevention and Health Promotion, HHS
(13)Project Lead, Pregnancy and Birth to 24 Months Project; NESR team, ONGA, 
CNPP, FNS, USDA

BACKGROUND: Systematic reviews were conducted as part of the U.S. Department of 
Agriculture and Department of Health and Human Services Pregnancy and Birth to 
24 Months Project. The goal of this systematic review was to examine the 
following question: What is the relationship between types and amounts of 
complementary foods and beverages (CFB) consumed and developmental milestones? 
Complementary feeding is the process that starts when human milk or infant 
formula is complemented by other foods and beverages, beginning during infancy 
and typically continuing to 24 months of age. CFB were defined as foods and/or 
beverages other than human milk or infant formula (liquids, semisolids, and 
solids) provided to an infant or young child to provide nutrients and energy.
CONCLUSION STATEMENT: There was insufficient evidence to draw a conclusion about 
the relationships between types and amounts of CFB consumed and developmental 
milestones. Grade: Grade Not Assignable.
METHODS: This systematic review was conducted by a team of staff from the 
Nutrition Evidence Systematic Review in collaboration with a Technical Expert 
Collaborative. Literature search was conducted using 4 databases (PubMed, 
Cochrane, Embase, and CINAHL) to identify articles that evaluated the 
intervention or exposure of timing of CFB introduction and the outcomes of 
developmental milestones. A manual search was conducted to identify articles 
that may not have been included in the electronic databases searched. Articles 
were screened by two analysts independently for inclusion based on 
pre-determined criteria. Data extraction and risk of bias assessment were 
conducted for each included study, and both were checked for accuracy. The body 
of evidence was qualitatively synthesized to inform development of a conclusion 
statement(s), and the strength of evidence was graded using pre-established 
criteria evaluating the body of evidence on risk of bias, adequacy, consistency, 
impact, and generalizability.
SUMMARY OF THE EVIDENCE: This review includes eight studies published since 1980 
that examined the relationship between consuming different types and/or amounts 
of CFB and developmental milestones during childhood through 18 years of age, 
including three randomized controlled trials and five prospective cohort 
studies. The studies varied in terms of the types and/or amounts of CFB 
examined, which included dietary patterns consumed during the complementary 
feeding period, meat and/or fortified-cereal intake, and foods with differing 
levels of DHA or phytate. Different types of developmental milestone outcomes 
were measured between 4 months and 8.5 years of age, including: ○. Communication 
(e.g., sentence repetition). ○. Cognitive (e.g., mental development index). ○. 
Motor (e.g., psychomotor development index). ○. Neurological (e.g., cortical 
processing). Three articles from two observational studies identified positive 
associations between dietary patterns emphasizing vegetables and meats during 
the complementary feeding period, and intelligence quotient (IQ) between ages 
4-8.5y. However, a conclusion could not be drawn due to low generalizability and 
heterogeneity in exposures, observed effects, and potential confounding. Because 
there was substantial variation in how studies were designed, it is difficult to 
compare and contrast the reported results. No conclusion regarding the 
relationship between types and/or amounts of CFB and developmental milestones 
could be drawn due to an inadequate number of studies that were comparable in 
terms of design, the types of CFB examined, how and when developmental 
milestones outcomes were assessed, and reported results.

DOI: 10.52570/NESR.PB242018.SR0308
PMID: 35853106


80. Types and Amounts of Complementary Foods and Beverages and Micronutrient Status: 
A Systematic Review [Internet].

Obbagy JE(1), English LK(2), Psota TL(1), Nadaud P(2), Johns K(2), Wong YP(3), 
Terry N(4), Butte NF(5), Dewey KG(6), Fleischer DM(7), Fox MK(8), Greer FR(9), 
Krebs NF(10), Scanlon KS(11), Casavale KO(12), Spahn JM(13), Stoody E(13).

Alexandria (VA): USDA Nutrition Evidence Systematic Review; 2019 Apr.
USDA Nutrition Evidence Systematic Reviews.

Author information:
(1)Systematic review analyst, Nutrition Evidence Systematic Review (NESR) team; 
Office of Nutrition Guidance and Analysis (ONGA), Center for Nutrition Policy 
and Promotion (CNPP), Food and Nutrition Service (FNS), U.S. Department of 
Agriculture (USDA)
(2)Systematic review analyst, NESR team; Panum Group under contract with FNS, 
USDA
(3)Systematic review librarian, NESR team; ONGA, CNPP, FNS, USDA
(4)Biomedical librarian, NESR team; National Institutes of Health Library, U.S. 
Department of Health and Human Services (HHS)
(5)Member, Complementary Feeding Technical Expert Collaborative, Pregnancy and 
Birth to 24 Months Project; Baylor College of Medicine, Emeritus
(6)Member, Complementary Feeding Technical Expert Collaborative, Pregnancy and 
Birth to 24 Months Project; University of California, Davis
(7)Member, Complementary Feeding Technical Expert Collaborative, Pregnancy and 
Birth to 24 Months Project; University of Colorado Denver School of Medicine, 
Children’s Hospital Colorado
(8)Member, Complementary Feeding Technical Expert Collaborative, Pregnancy and 
Birth to 24 Months Project; Mathematica Policy Research
(9)Member, Complementary Feeding Technical Expert Collaborative, Pregnancy and 
Birth to 24 Months Project; University of Wisconsin, Madison, Emeritus
(10)Member, Complementary Feeding Technical Expert Collaborative, Pregnancy and 
Birth to 24 Months Project; University of Colorado School of Medicine, 
Department of Pediatrics
(11)Member and Federal Expert Group Liaison, Complementary Feeding Technical 
Expert Collaborative, Pregnancy and Birth to 24 Months Project; Office of Policy 
Support, FNS, USDA
(12)Project Lead, Pregnancy and Birth to 24 Months Project; Office of Disease 
Prevention and Health Promotion, HHS
(13)Project Lead, Pregnancy and Birth to 24 Months Project; NESR team, ONGA, 
CNPP, FNS, USDA

BACKGROUND: Complementary feeding is the process that starts when human milk or 
infant formula is complemented by other foods and beverages, beginning during 
infancy and typically continuing to 24 months of age. This systematic review was 
conducted by the Nutrition Evidence Systematic Review team as part of the U.S. 
Department of Agriculture and Department of Health and Human Services Pregnancy 
and Birth to 24 Months Project. The goal of this systematic review was to answer 
the following research question: What is the relationship between types and 
amounts of complementary foods and beverages and micronutrient status?
CONCLUSION STATEMENT AND GRADES: Strong evidence suggests that consuming 
complementary foods and beverages that contain substantial amounts of iron, such 
as meats or iron-fortified cereal, helps maintain adequate iron status or 
prevent iron deficiency during the first year of life among infants with 
insufficient iron stores or breastfed infants who are not receiving adequate 
iron from another source. However, the benefit of these types of complementary 
foods and beverages for infants with sufficient iron stores, such as those 
consuming iron-fortified infant formula, is less evident. There is not enough 
evidence to determine the relationship between other types/amounts of 
complementary foods and beverages containing lesser amounts of iron, such as 
fruits and vegetables, and iron status. Limited evidence suggests that consuming 
complementary foods and beverages that contain substantial amounts of zinc, such 
as meats or cereals fortified with zinc, supports zinc status during the first 
year of life, particularly among breastfed infants who are not receiving 
adequate zinc from another source. However, the benefit of these types of 
complementary foods for infants consuming fortified infant formula is less 
evident. Moderate evidence suggests that consuming complementary foods and 
beverages with differing fatty acid profiles, particularly long-chain 
polyunsaturated fatty acids, can influence fatty acid status. During the second 
year of life, good sources of micronutrients are still needed, but there is 
limited evidence to indicate which types and amounts of complementary foods and 
beverages are associated with adequate micronutrient status. There is not enough 
evidence to determine the relationship between types and amounts of 
complementary foods and beverages and vitamin B12, vitamin D, or folate status. 
Grades: Strong – Iron status; Moderate – Fatty Acid status; Limited – Zinc 
status; Grade Not Assignable – Vitamin B12 status, Vitamin D status, Folate 
status.
METHODS: The systematic review was conducted by a team of staff from the 
Nutrition Evidence Systematic Review in collaboration with a Technical Expert 
Collaborative. A literature search was conducted using 4 databases (CINAHL, 
Cochrane, Embase, and PubMed) to identify articles published from January 1980 
to March 2016 that examined the types and amounts of complementary foods and 
beverages (CFB) consumed and micronutrient status. CFB were defined as foods and 
beverages other than human milk or infant formula provided to an infant or young 
child. Micronutrient status outcomes included iron, zinc, vitamin B12, folate, 
vitamin D, and/or folate status. A manual search was done to identify articles 
that may not have been included in the electronic databases searched. Articles 
were screened in a dual manner, independently by 2 NESR analysts, to determine 
which articles met predetermined criteria for inclusion. Data from each included 
article were extracted, risks of bias were assessed, and both were checked for 
accuracy. The body of evidence was qualitatively synthesized, a conclusion 
statement was developed, and the strength of the evidence (grade) was assessed 
using pre-established criteria including evaluation of the internal 
validity/risk of bias, adequacy, consistency, impact, and generalizability of 
available evidence.
SUMMARY OF EVIDENCE: Thirty-one studies published between 01/1980 and 07/2016 
met the inclusion criteria for this systematic review. ○. Most studies examined 
the relationship between types and/or amounts of CFB and iron status, and the 
CFB examined were largely limited to Fe-fortified cereals and meats. ○. Several 
studies examined zinc and fatty acid status and few studies examined vitamin D, 
vitamin B12, and folate. One randomized controlled trial, conducted in both 
breastfed (BF) and formula fed (FF) infants, showed that consuming meats or Fe- 
and/or Zn-fortified cereals as CFB generally protected against Fe deficiency 
anemia and supported Zn status in the first year of life, though evidence is 
more limited in the second year of life. ○. Among BF infants, meat and Fe- and 
Zn-fortified cereals supported iron and zinc status in later infancy. Meat 
provided a valuable source of trace minerals for BF infants who may not have 
been fed Fe- and Zn-fortified products; in fact, the frequency of meat 
consumption was associated with iron status in the first and second years of 
life. ○. In infants/toddlers whose diets already contained other bioavailable Fe 
and Zn sources (i.e. infant formulas and cereal fortified with iron and/or 
zinc), meat offered little additional benefit for Fe or Zn status, though it is 
an important source of bioavailable Fe and Zn. Dietary sources of fatty acids, 
particularly LC-PUFA, in CFB (i.e., oils, fish, meats, and eggs) influenced the 
plasma fatty acid profile of infants and toddlers. A limitation of some of the 
studies included in this systematic review was lack of accounting for whether 
infants were fed breast milk and/or infant formula, and other aspects of the 
overall diet, including consumption of fortified products and bioavailability of 
nutrients consumed. Another limitation is a lack of studies that examined 
vitamin D, vitamin B12, and folate.

DOI: 10.52570/NESR.PB242018.SR0302
PMID: 35816596


81. Timing of Introduction of Complementary Foods and Beverages and Micronutrient 
Status: A Systematic Review [Internet].

Obbagy JE(1), English LK(2), Psota TL(1), Nadaud P(2), Johns K(2), Wong YP(3), 
Terry N(4), Butte NF(5), Dewey KG(6), Fleischer DM(7), Fox MK(8), Greer FR(9), 
Krebs NF(10), Scanlon KS(11), Casavale KO(12), Spahn JM(13), Stoody E(13).

Alexandria (VA): USDA Nutrition Evidence Systematic Review; 2019 Apr.
USDA Nutrition Evidence Systematic Reviews.

Author information:
(1)Systematic review analyst, Nutrition Evidence Systematic Review (NESR) team; 
Office of Nutrition Guidance and Analysis (ONGA), Center for Nutrition Policy 
and Promotion (CNPP), Food and Nutrition Service (FNS), U.S. Department of 
Agriculture (USDA)
(2)Systematic review analyst, NESR team; Panum Group under contract with FNS, 
USDA
(3)Systematic review librarian, NESR team; ONGA, CNPP, FNS, USDA
(4)Biomedical librarian, NESR team; National Institutes of Health Library, U.S. 
Department of Health and Human Services (HHS)
(5)Member, Complementary Feeding Technical Expert Collaborative, Pregnancy and 
Birth to 24 Months Project; Baylor College of Medicine, Emeritus
(6)Member, Complementary Feeding Technical Expert Collaborative, Pregnancy and 
Birth to 24 Months Project; University of California, Davis
(7)Member, Complementary Feeding Technical Expert Collaborative, Pregnancy and 
Birth to 24 Months Project; University of Colorado Denver School of Medicine, 
Children’s Hospital Colorado
(8)Member, Complementary Feeding Technical Expert Collaborative, Pregnancy and 
Birth to 24 Months Project; Mathematica Policy Research
(9)Member, Complementary Feeding Technical Expert Collaborative, Pregnancy and 
Birth to 24 Months Project; University of Wisconsin, Madison, Emeritus
(10)Member, Complementary Feeding Technical Expert Collaborative, Pregnancy and 
Birth to 24 Months Project; University of Colorado School of Medicine, 
Department of Pediatrics
(11)Member and Federal Expert Group Liaison, Complementary Feeding Technical 
Expert Collaborative, Pregnancy and Birth to 24 Months Project; Office of Policy 
Support, FNS, USDA
(12)Project Lead, Pregnancy and Birth to 24 Months Project; Office of Disease 
Prevention and Health Promotion, HHS
(13)Project Lead, Pregnancy and Birth to 24 Months Project; NESR team, ONGA, 
CNPP, FNS, USDA

BACKGROUND: Complementary feeding is the process that starts when human milk or 
infant formula is complemented by other foods and beverages, beginning during 
infancy and typically continuing to 24 months of age. This systematic review was 
conducted by the Nutrition Evidence Systematic Review team as part of the U.S. 
Department of Agriculture and Department of Health and Human Services Pregnancy 
and Birth to 24 Months Project. The goal of this systematic review was to answer 
the following research question: What is the relationship between timing of 
introduction of complementary foods and beverages and micronutrient status?
CONCLUSION STATEMENT AND GRADES: Moderate evidence suggests that introducing 
complementary foods and beverages at 4 months of age compared to 6 months of age 
offers no long term advantages or disadvantages in terms of iron status among 
healthy, full-term infants who are breastfed, fed iron fortified formula, or 
both. (Grade: Moderate – Iron Status). There is not enough evidence to determine 
the relationship between timing of introduction of complementary foods and 
beverages and zinc, vitamin D, vitamin B12, folate, or fatty acid status. 
(Grade: Grade Not Assignable - Zinc, Vitamin D, Vitamin B12, Folate, and Fatty 
Acid Status).
METHODS: The systematic review was conducted by a team of staff from the 
Nutrition Evidence Systematic Review in collaboration with a Technical Expert 
Collaborative. A literature search was conducted using 4 databases (CINAHL, 
Cochrane, Embase, and PubMed) to identify articles published from January 1980 
to March 2016 that examined the age when complementary foods and beverages (CFB) 
were first introduced and micronutrient status. CFB were defined as foods and 
beverages other than human milk or infant formula provided to an infant or young 
child. Micronutrient status outcomes included iron, zinc, vitamin B12, folate, 
vitamin D, and/or folate status. A manual search was done to identify articles 
that may not have been included in the electronic databases searched. Articles 
were screened in a dual manner, independently by 2 NESR analysts, to determine 
which articles met predetermined criteria for inclusion. Data from each included 
article were extracted, risks of bias were assessed, and both were checked for 
accuracy. The body of evidence was qualitatively synthesized, a conclusion 
statement was developed, and the strength of the evidence (grade) was assessed 
using pre-established criteria including evaluation of the internal 
validity/risk of bias, adequacy, consistency, impact, and generalizability of 
available evidence.
SUMMARY OF EVIDENCE: Nine studies published from 1/1980 to 3/2016 met the 
inclusion criteria for this systematic review, with most studies examining the 
relationship between timing of introduction of CFB and iron status. Few studies 
examined zinc, vitamin D, vitamin B12, folate, and/or fatty acid status. The 
majority of studies reported no significant associations between timing of CFB 
introduction and micronutrient status. Additional factors that need to be 
considered in examining the relationship between the age at which CFB are 
introduced and micronutrient status include: birth weight, post-natal growth, 
type of feeding (breast, formula, or mixed feedings), iron stores at birth, and 
intake and absorption of iron from sources other than human milk, including 
types and amounts of CFB being consumed.

DOI: 10.52570/NESR.PB242018.SR0301
PMID: 35782004


82. Cochrane Database Syst Rev. 2019 Feb 19;2(2):CD012818. doi: 
10.1002/14651858.CD012818.pub2.

Effectiveness of provision of animal-source foods for supporting optimal growth 
and development in children 6 to 59 months of age.

Eaton JC(1), Rothpletz-Puglia P, Dreker MR, Iannotti L, Lutter C, Kaganda J, 
Rayco-Solon P.

Author information:
(1)Brown School/Institute for Public Health, Washington University in St. Louis, 
1 Brookings Drive, St. Louis, Missouri, USA, 63130.

Update of
    doi: 10.1002/14651858.CD012818.

BACKGROUND: Adequate nutrients early in life promote cognitive development and 
are critical for proper growth and functioning. The effect of individual 
nutrients consumed through food is often not the same as consuming the same 
nutrients in supplementary form due to 'food synergy', the biological and 
chemical interrelations that occur between nutrients. Animal-source foods, such 
as eggs, meat, fish, and dairy, are energy dense and contain multiple 
micronutrients and essential fatty acids with high bioavailability. The benefits 
of animal-source foods may include higher food synergy relative to fortified 
foods as well as decreasing dependence on external suppliers of fortified foods.
OBJECTIVES: To assess the effectiveness of animal-source foods compared to any 
other feeding interventions or no intervention in improving growth and 
developmental outcomes in children aged 6 to 59 months.
SEARCH METHODS: We searched CENTRAL, MEDLINE, Embase, CINAHL, 18 other 
databases, and three trials registers up to August 2018. We also contacted 
authors and known experts in the field for assistance in identifying ongoing or 
unpublished data, and searched the reference lists of included studies and 
reviews, and websites of relevant organizations, for other studies that may not 
have been captured by our electronic searches.
SELECTION CRITERIA: We included randomized controlled trials and 
quasi-randomized controlled trials of any duration, where children between 5 
months and 59 months (6 years) of age were provided with an animal-source food 
(e.g. consumption of milk, meat, or eggs), prepared with any cooking method, 
compared with any intervention or no intervention.
DATA COLLECTION AND ANALYSIS: Two review authors independently assessed trial 
eligibility using prespecified criteria, extracted data, assessed risk of bias, 
and graded the quality of the evidence using the GRADE approach.
MAIN RESULTS: Study characteristicsWe included 6 studies that analyzed data from 
3036 children aged 5 to 50 months. The studies were conducted in China, the 
Democratic Republic of Congo, Ecuador, Guatemala, Pakistan, the USA, and Zambia, 
and lasted between 5 and 12 months. Three studies were funded, in part, by 
government entities; one study was supported by a nonprofit organization. Two 
studies did not report a funding source.Three studies compared the effects of 
feeding an animal-source food with a fortified (iron or iron and zinc), or 
unfortified cereal; two used a control group with no intervention; one compared 
a meat-based diet to a dairy-based diet. The types of animal-source foods tested 
included yogurt, eggs, cheese, lyophilized (freeze-dried) beef product, ground 
and frozen pork, puréed and jarred beef with gravy or pork, and powdered whey 
protein.We judged four studies to be at unclear risk of bias overall; three 
studies because they were funded by an industry with a plausible interest in the 
outcome of the intervention; and one study because there was insufficient 
information to assess five of the seven bias 'Risk of bias' domains. We judged 
two of the six studies to be at high risk of bias overall; one study because 
there was significant baseline imbalance in length-for-age z scores (LAZ) 
between groups and evidence of selective reporting; the other study because 
there there was both a significant baseline imbalance in LAZ and weight-for-age 
z scores (WAZ) between groups, and a large-scale social media campaign that may 
have influenced care received at home in the control group.Key 
resultsAnimal-source foods versus cereal-based foods or no interventionFive 
studies (2972 children) measured change in linear growth with either 
height-for-age z scores (HAZ) or LAZ. Three studies (592 children) reported a 
significant increase in HAZ and LAZ in the intervention group compared to the 
control group. Two studies (2380 children) reported a decline in LAZ in both 
groups. In one study (1062 children) there was no difference between the groups 
in the rate of decline; in the other (1318 children) the decrease in LAZ was 
significantly smaller in the intervention group.Five studies (2972 children) 
measured weight gain using WAZ. Three studies (592 children) reported a 
significant increase in WAZ in the intervention group compared to the control 
group. In two studies (2380 children), WAZ decreased in both groups. In one of 
these studies (1318 children), the decrease in the intervention group was 
significantly smaller than in the control group. In the other study (1062 
children), there was no difference between the groups.Three studies (1612 
children) reported impacts on all-cause morbidity, but metrics were inconsistent 
between studies. One study with yogurt (402 children) reported a significant 
reduction in duration and incidence of diarrhea and upper respiratory infections 
in the intervention group. One study with eggs (148 children) reported a 
significant increase in the incidence of diarrhea in the intervention group, but 
this may have been due to cultural associations with eggs and gastrointestional 
problems. There were no other significant differences in fever, respiratory 
infections, or skin conditions between groups. The third study (1062 children) 
found no differences between intervention and control groups across morbidity 
measures.No studies reported data on anemia.Meat-based diet versus dairy-based 
dietOne study (64 children) measured change in LAZ and WAZ in infants fed either 
a meat-based diet or dairy-based diet. There was a significant increase in LAZ 
among infants consuming the meat-based diet and a significant decrease in LAZ 
among infants consuming a dairy-based diet. WAZ increased in both groups, with 
no significant difference between groups.The study did not assess all-cause 
morbidity or anemia.Quality of the evidenceWe rated the quality of the evidence 
as very low overall due to baseline imbalances between intervention and control 
groups, high heterogeneity in meta-analysis, and imprecision due to wide 
confidence intervals and inconsistent direction of effects. We have little 
confidence in the results; further research is likely to change the estimate of 
magnitude and direction of treatment effect.
AUTHORS' CONCLUSIONS: Given the limited quality of the evidence, we are 
uncertain of the effects of the provision of animal-source food versus cereal 
products or no intervention on the growth or development of children. More 
adequately powered trials with deliberately selected animal-source foods are 
needed.

DOI: 10.1002/14651858.CD012818.pub2
PMCID: PMC6380771
PMID: 30779870 [Indexed for MEDLINE]

Conflict of interest statement: Jacob Eaton received financial support from the 
World Health Organization (WHO) for his work on this review and for travel to 
attend the Cochrane/Cornell/WHO Summer Collaborative for Systematic Reviews in 
Nutrition.  Pamela Rothpletz‐Puglia: none known.  Margaret R Dreker: none 
known.  Lora Iannotti and Chessa Lutter are authors of one of the studies 
included in this review (Iannotti 2017). Neither review author was involved in 
study selection, assessment of risk of bias, data extraction, or assessment of 
the quality of the evidence.  Joyceline Kaganda: none known.  Pura Rayco‐Solon 
is a full‐time member of staff at the WHO. Disclaimer: the review authors alone 
are responsible for the views expressed in this publication, and they do not 
necessarily represent the official position, decisions, policy, or views of the 
WHO. The WHO gratefully acknowledges the financial contribution of the Bill & 
Melinda Gates Foundation, Nutrition International (NI; formerly Micronutrient 
Initiative (MI)), the Centers for Disease Control and Prevention (CDC), the US 
Agency for International development (USAID), and the Global Alliance for 
Improved Nutrition (GAIN) towards work in the area of nutrition. Donors do not 
fund specific guidelines and do not participate in any decision related to the 
guideline development process, including the composition of research questions, 
membership of the guideline groups, conduct and interpretation of systematic 
reviews, or formulation of recommendations.


83. Cochrane Database Syst Rev. 2019 Feb 12;2(2):CD010734. doi: 
10.1002/14651858.CD010734.pub2.

Iodine fortification of foods and condiments, other than salt, for preventing 
iodine deficiency disorders.

Santos JAR(1), Christoforou A, Trieu K, McKenzie BL, Downs S, Billot L, Webster 
J, Li M.

Author information:
(1)Office of the Chief Scientist, The George Institute for Global Health, 
University of New South Wales, Sydney, Australia.

Update of
    doi: 10.1002/14651858.CD010734.

BACKGROUND: Iodine deficiency disorders (IDD) affect close to 1.9 billion people 
worldwide, and are a major public health concern in many countries. Among 
children, iodine deficiency is the main cause of potentially preventable 
deficits of central nervous system development and impairment of cognitive 
function, as well as goitre and hypothyroidism in people of all ages. Salt 
iodisation is the preferred strategy for IDD prevention and control, however, in 
some instances where salt is not the major condiment, alternate vehicles for 
iodine fortification have been considered.
OBJECTIVES: To assess the effects of fortifying foods, beverages, condiments, or 
seasonings other than salt with iodine alone or in conjunction with other 
micronutrients, on iodine status and health-related outcomes in all populations.
SEARCH METHODS: Studies were identified through systematic searches of the 
following databases from their start date to January 2018: Cochrane Public 
Health Group Specialised Register; CENTRAL; MEDLINE; MEDLINE in Process; Embase; 
Web of Science; CINAHL; POPLINE; AGRICOLA; BIOSIS; Food Science and Technology 
Abstracts; OpenGrey; Bibliomap and TRoPHI; AGRIS; IBECS; Scielo; Global Index 
Medicus-AFRO and EMRO; LILACS; PAHO; WHOLIS; WPRO; IMSEAR; IndMED; and Native 
Health Research Database. We also searched reference list of relevant articles, 
conference proceedings, and databases of ongoing trials, and contacted experts 
and relevant organisations to identify any unpublished work. We applied no 
language or date restrictions.
SELECTION CRITERIA: Studies were eligible if they were randomised or 
quasi-randomised controlled trials (RCT) with randomisation at either the 
individual or cluster level (including cross-over trials), non-randomised RCTs, 
or prospective observational studies with a control group, such as cohort 
studies, controlled before-and-after studies, and interrupted time series. We 
included studies that examined the effects of fortification of food, beverage, 
condiment, or seasoning with iodine alone, or in combination with other 
micronutrients versus the same unfortified food, or no intervention. We 
considered the following measures: death (all-cause), goitre, physical 
development, mental development, cognitive function and motor skill development, 
cretinism, hypothyroidism, adverse effects (any reported by trialists), urinary 
iodine concentration, thyroid-stimulating hormone (TSH) concentration, and serum 
thyroglobulin concentration. We included all populations, including pregnant 
women, from any country.
DATA COLLECTION AND ANALYSIS: Two review authors independently assessed study 
eligibility, extracted data, and assessed risk of bias of included studies.We 
used random-effects meta-analyses to combine data and generate an overall 
estimate of treatment effect, when more than one study examined the same outcome 
measure. The overall effect estimate was calculated as the mean difference (MD) 
or standardised mean difference (SMD) between the intervention group and the 
comparison group for continuous outcomes, and as odds ratio (OR) for dichotomous 
outcomes. We assessed the level of heterogeneity through the I² statistic. We 
conducted post-hoc subgroup analyses to explore possible sources of 
heterogeneity, and sensitivity analyses to check the robustness of the findings 
from the primary analyses. We assessed the quality of the evidence for each 
outcome using the GRADE framework.Where it was not possible to pool the results 
in a meta-analysis, we provided a narrative summary of the outcomes.
MAIN RESULTS: Eleven studies met the criteria, providing 14 comparisons, and 
capturing data on 4317 participants. Seven studies were RCTs, three were cluster 
non-RCTs, and one was a randomised cross-over design. Seven studies were carried 
out among school children (N = 3636), three among women of reproductive age (N = 
648), and one among infants (N = 33). The studies used diverse types of food as 
vehicle for iodine delivery: biscuits, milk, fish sauce, drinking water, 
yoghourt, fruit beverage, seasoning powder, and infant formula milk. Daily 
amounts of iodine provided ranged from 35 µg/day to 220 µg/day; trial duration 
ranged from 11 days to 48 weeks. Five studies examined the effect of iodine 
fortification alone, two against the same unfortified food, and three against no 
intervention. Six studies evaluated the effect of cofortification of iodine with 
other micronutrients versus the same food without iodine but with different 
levels of other micronutrients. We assessed one study to be at low risk of bias 
for all bias domains, three at low risk of bias for all domains apart from 
selective reporting, and seven at an overall rating of high risk of bias.No 
study assessed the primary outcomes of death, mental development, cognitive 
function, cretinism, or hypothyroidism, or secondary outcomes of TSH or serum 
thyroglobulin concentration. Two studies reported the effects on goitre, one on 
physical development measures, and one on adverse effects. All studies assessed 
urinary iodine concentration.The effects of iodine fortification compared to 
control on goitre prevalence (OR 1.60, 95% CI 0.60 to 4.31; 1 non-RCT, 83 
participants; very low-quality evidence), and five physical development measures 
were uncertain (1 non-RCT, 83 participants; very low-quality evidence): weight 
(MD 0.23 kg, 95% CI -6.30 to 6.77); height (MD -0.66 cm, 95% CI -4.64 to 3.33); 
weight-for-age (MD 0.05, 95% CI -0.59 to 0.69); height-for-age (MD -0.30, 95% CI 
-0.75 to 0.15); and weight-for-height (MD -0.21, 95% CI -0.51 to 0.10). One 
study reported that there were no adverse events observed during the cross-over 
trial (low-quality evidence).Pooled results from RCTs showed that urinary iodine 
concentration significantly increased following iodine fortification (SMD 0.59, 
95% CI 0.37 to 0.81; 6 RCTs, 2032 participants; moderate-quality evidence). This 
is equivalent to an increase of 38.32 µg/L (95% CI 24.03 to 52.61 µg/L). This 
effect was not observed in the meta-analysis of non-RCTs (SMD 0.25, 95% CI -0.16 
to 0.66; 3 non-RCTs, 262 participants; very low-quality evidence). Sensitivity 
analyses did not change the effect observed in the primary analyses.
AUTHORS' CONCLUSIONS: The evidence on the effect of iodine fortification of 
foods, beverages, condiments, or seasonings other than salt on reducing goitre, 
improving physical development measures, and any adverse effects is uncertain. 
However, our findings suggest that the intervention likely increases urinary 
iodine concentration. Additional, adequately powered, high-quality studies on 
the effects of iodine fortification of foods on these, and other important 
outcomes, as well as its efficacy and safety, are required.

DOI: 10.1002/14651858.CD010734.pub2
PMCID: PMC6370918
PMID: 30746700 [Indexed for MEDLINE]

Conflict of interest statement: JW is the Director of the World Health 
Organization Collaborating Centre on Population Salt Reduction, and receives 
funding for her work from the National Health and Medical Research Council, the 
Victorian Health Promotion Foundation, and headquarters and various regional 
offices of the World Health Organization. From 2003 to 2006, she managed the UK 
government's Salt Reduction program while working for the Food Standards Agency. 
She is currently supported by a four‐year National Health and Medical Research 
Council Fellowship/National Heart Foundation fellowship for her work on salt 
reduction. KT receives support from a National Health and Medical Research 
Council postgraduate scholarship, the Victorian Health Promotion Foundation 
(VicHealth), and the World Health Organization. She received financial support 
from the World Health Organization during the conduct of the study. ML ‐ none 
known. JS ‐ none known. AC ‐ none known. BM ‐ none known. SD ‐ none known. LB ‐ 
none known.


84. Support Care Cancer. 2019 Feb;27(2):333-349. doi: 10.1007/s00520-018-4458-9. 
Epub 2018 Sep 10.

Sensory preferences of supplemented food products among cancer patients: a 
systematic review.

Enriquez-Fernández BE(1), Nejatinamini S(1), Campbell SM(2), Mazurak VC(1), 
Wismer WV(3)(4).

Author information:
(1)Department of Agricultural, Food and Nutritional Science, University of 
Alberta, Edmonton, AB, T6G 2E1, Canada.
(2)University of Alberta Libraries, University of Alberta, Edmonton, AB, Canada.
(3)Department of Agricultural, Food and Nutritional Science, University of 
Alberta, Edmonton, AB, T6G 2E1, Canada..
(4)Department of Agricultural, Life and Environmental Sciences, University of 
Alberta, 3-18C Agriculture/Forestry Ctr, Edmonton, AB, T6G 2E1, Canada. 
.

PURPOSE: Oral nutritional supplements and fortified foods, here considered 
supplemented food products (SFP), are recommended as part of nutrition therapy 
guidelines to treat malnutrition among cancer patients. However, their 
successful use is limited by patients' failure to meet recommended intakes. This 
systematic review aimed to identify sensory preferences for SFP among cancer 
patients and evaluate the methodologies employed in sensory preference 
assessment.
METHODS: A systematic search was conducted in several relevant databases 
yielding 1056 papers of which 19 met the inclusion criteria. Two authors 
independently selected papers and extracted findings. The included papers were 
categorized according to the focus of the preference assessment.
RESULTS: Studies comparing sensory preferences for SFP of cancer patients with 
those of a control group suggested that the liking for SFP by cancer patients 
differs from healthy participants. Patient heterogeneity in site and stage of 
tumor, variation in study methodologies, and type of treatment complicated a 
conclusion regarding the effects of cancer treatment and taste changes on taste 
preferences. However, some general results were observed among the studies, such 
as the preference for fresh milk-based supplements when compared with other 
supplement types.
CONCLUSION: This review highlighted the need for consistent reporting and 
control of variables that influence the sensory characteristics of SFP when 
sensory preferences are assessed in the clinical setting. Attention to these 
methodological details will enhance the reliability and accuracy of sensory 
preference assessment among cancer patients for realistic evaluation of SFP 
targeted to their nutritional needs.

DOI: 10.1007/s00520-018-4458-9
PMID: 30203359 [Indexed for MEDLINE]


85. Ann Nutr Metab. 2018;73 Suppl 1:7-13. doi: 10.1159/000490087. Epub 2018 Sep 7.

Impact of Food Fortification on Child Growth and Development during 
Complementary Feeding.

Okeyo DO.

There appears to be increasing evidence of the relationship between infant 
feeding practices and growth during infancy. Effective complementary feeding has 
demonstrated an observable positive effect on the linear growth of a child 
within the first 24 months of life. It appears that improved complementary 
feeding is directly proportional to the linear growth of a child. Fortification 
of commonly used food vehicles provides an opportunity for increasing nutrient 
intake during infancy and has the potential to improve growth and development 
dimensions. This review scanned through 186 articles from common search engines, 
mainly PubMed, BioMed Central, and Google Scholar. The result based on a 
systematic review of articles which met the minimum selection criteria 
identified milk, iodine, maize meal porridge, and vegetable oils as recurring 
fortification vehicles in the context of complementary feeding. A significant 
impact of fortification on linear and cognitive growth was demonstrated 
recurrently across the included empirical studies. However, the review reflects 
outcomes that still do not demonstrate direct cause and effect relationships but 
rather implied meaning in the relationship matrix.

© 2018 S. Karger AG, Basel.

DOI: 10.1159/000490087
PMID: 30196295 [Indexed for MEDLINE]


86. Breastfeed Med. 2018 Sep;13(7):465-472. doi: 10.1089/bfm.2018.0135. Epub 2018 
Aug 27.

Effects of Refrigerated and Frozen Storage on Holder-Pasteurized Donor Human 
Milk: A Systematic Review.

Schlotterer HR(1), Perrin MT(1).

Author information:
(1)Department of Nutrition, University of North Carolina Greensboro , 
Greensboro, North Carolina.

BACKGROUND: Pasteurized donor human milk (PDHM) is the recommended feeding 
alternative for preterm infants when mother's own milk is not available. Use of 
PDHM in United States neonatal hospitals is increasing, although guidelines for 
the refrigerated and frozen storage are limited.
OBJECTIVE: We aimed to review the current evidence for the storage of Holder 
PDHM (HPDHM) under refrigerated and frozen storage conditions.
METHODS: A systematic review of the literature was conducted for studies 
published between 1985 and May 2018. Studies were included if they studied the 
storage of Holder-pasteurized human milk under refrigerated or frozen storage 
conditions.
RESULTS: Fourteen studies met the inclusion criteria. Five studies addressed 
refrigerated storage and nine studies addressed frozen storage. There was little 
overlap in the outcomes measured or the analytical methods employed. There was 
concordance in three studies reporting no microbial growth over 4-9 days of 
refrigerated storage, and in five studies reporting a reduction in fat during 
1-8 months of frozen storage. Only one study assessed the storage of HPDHM that 
had been fortified.
CONCLUSIONS: Long-term refrigerated and frozen storage of HPDHM affects some 
components in milk more than others. While there is evidence of microbial purity 
during four or more days of refrigerated storage in clinical conditions, there 
is limited research on the impact of macro and micronutrients, or the impact of 
fortifiers. More research is needed in these areas.

DOI: 10.1089/bfm.2018.0135
PMID: 30148655 [Indexed for MEDLINE]


87. Cochrane Database Syst Rev. 2018 Aug 23;8(8):CD000280. doi: 
10.1002/14651858.CD000280.pub2.

Carbohydrate supplementation of human milk to promote growth in preterm infants.

Amissah EA(1), Brown J, Harding JE.

Author information:
(1)Liggins Institute, University of Auckland, Auckland, New Zealand.

Update in
    Cochrane Database Syst Rev. 2020 Sep 8;9:CD000280. doi: 
10.1002/14651858.CD000280.pub3.

Update of
    Cochrane Database Syst Rev. 2000;(2):CD000280. doi: 
10.1002/14651858.CD000280.

BACKGROUND: Preterm infants are born with low glycogen stores and require higher 
glucose intake to match fetal accretion rates. In spite of the myriad benefits 
of breast milk for preterm infants, it may not adequately meet the needs of 
these rapidly growing infants. Supplementing human milk with carbohydrates may 
help. However, there is a paucity of data on assessment of benefits or harms of 
carbohydrate supplementation of human milk to promote growth in preterm infants. 
This is a 2018 update of a Cochrane Review first published in 1999.
OBJECTIVES: To determine whether human milk supplemented with carbohydrate 
compared with unsupplemented human milk fed to preterm infants improves growth, 
body composition, and cardio-metabolic and neurodevelopmental outcomes without 
significant adverse effects.
SEARCH METHODS: We used the standard search strategy of the Cochrane Neonatal 
Review Group to search the Cochrane Central Register of Controlled Trials 
(CENTRAL; 2017, Issue 8), MEDLINE via PubMed (1966 to 21 February 2018), Embase 
(1980 to 21 February 2018), and the Cumulative Index to Nursing and Allied 
Health Literature (CINAHL; 1982 to 21 February 2018). We also searched clinical 
trials databases, conference proceedings, and reference lists of retrieved 
articles for randomised controlled trials (RCTs) and quasi-randomised trials.
SELECTION CRITERIA: Published and unpublished controlled trials were eligible if 
they used random or quasi-random methods to allocate preterm infants in hospital 
fed human milk to supplementation or no supplementation with additional 
carbohydrate.
DATA COLLECTION AND ANALYSIS: Two review authors independently abstracted data 
and assessed trial quality and the quality of evidence at the outcome level 
using the Grading of Recommendations Assessment, Development and Evaluation 
(GRADE) method. We planned to perform meta-analyses using risk ratios (RRs) for 
dichotomous data and mean differences (MDs) for continuous data, with their 
respective 95% confidence intervals (CIs). We planned to use a fixed-effect 
model and to explore potential causes of heterogeneity via sensitivity analyses. 
We contacted study authors for additional information.
MAIN RESULTS: One unblinded, quasi-randomised controlled trial (RCT) assessing 
effects of carbohydrate supplementation of human milk in the form of a prebiotic 
in 75 preterm infants was eligible for inclusion in this review. We identified 
two publications of the same trial, which reported different methods regarding 
blinding and randomisation. Study authors confirmed that these publications 
pertain to the same trial, but they have not yet clarified which method is 
correct. We were unable to reproduce analyses from the data presented. At 30 
days of age, the mean weight of preterm infants in the trial was greater in the 
prebiotic carbohydrate-supplemented group than in the unsupplemented group (MD 
160.4 grams, 95% CI 12.4 to 308.4 grams; one RCT, N = 75; very low-quality 
evidence). We found no evidence of a clear difference in risk of feeding 
intolerance (RR 0.64, 95% CI 0.36 to 1.15; one RCT, N = 75 infants; very 
low-quality evidence) or necrotising enterocolitis (NEC) (RR 0.2, 95% CI 0.02 to 
1.3; one RCT, N = 75 infants; very low-quality evidence) between the 
prebiotic-supplemented group and the unsupplemented group. Duration of hospital 
stay was shorter in the prebiotic group than in the control group at a median 
(range) of 16 (9 to 45) days (95% CI 15.34 to 24.09) and 25 (11 to 80) days (95% 
CI 25.52 to 34.39), respectively. No other data were available for assessing 
effects of carbohydrate supplementation on short- and long-term growth, body 
mass index, body composition, and neurodevelopmental or cardio-metabolic 
outcomes.
AUTHORS' CONCLUSIONS: We found insufficient evidence on the short- and long-term 
effects of carbohydrate supplementation of human milk in preterm infants. The 
only trial included in this review presented very low-quality evidence, and 
study authors provided uncertain information about study methods and analysis. 
The evidence may be limited in its applicability because researchers included a 
small sample of preterm infants from a single centre. However, the outcomes 
assessed are common to all preterm infants, and this trial demonstrates the 
feasibility of prebiotic carbohydrate supplementation in upper-middle-income 
countries. Future trials should assess the safety and efficacy of different 
types and concentrations of carbohydrate supplementation for preterm infants fed 
human milk. Although prebiotic carbohydrate supplementation in preterm infants 
is currently a topic of active research, we do not envisage that further trials 
of digestible carbohydrates will be conducted, as this is currently done as a 
component of multi-nutrient human milk fortification. Hence we do not plan to 
publish any further updates of this review.

DOI: 10.1002/14651858.CD000280.pub2
PMCID: PMC6513426
PMID: 30138549 [Indexed for MEDLINE]

Conflict of interest statement: Emma Amissah: none known. Julie Brown: none 
known. Jane Harding: none known.


88. Cochrane Database Syst Rev. 2018 Jun 22;6(6):CD000433. doi: 
10.1002/14651858.CD000433.pub2.

Protein supplementation of human milk for promoting growth in preterm infants.

Amissah EA(1), Brown J, Harding JE.

Author information:
(1)Liggins Institute, University of Auckland, Auckland, New Zealand.

Update in
    Cochrane Database Syst Rev. 2020 Sep 23;9:CD000433. doi: 
10.1002/14651858.CD000433.pub3.

Update of
    Cochrane Database Syst Rev. 2000;(2):CD000433. doi: 
10.1002/14651858.CD000433.

BACKGROUND: Preterm infants require high protein intake to achieve adequate 
growth and development. Although breast milk feeding has many benefits for this 
population, the protein content is highly variable, and inadequate to support 
rapid infant growth. This is a 2018 update of a Cochrane Review first published 
in 1999.
OBJECTIVES: To determine whether protein-supplemented human milk compared with 
unsupplemented human milk, fed to preterm infants, improves growth, body 
composition, cardio-metabolic, and neurodevelopmental outcomes, without 
significant adverse effects.
SEARCH METHODS: We used the standard search strategy of Cochrane Neonatal to 
search CENTRAL, MEDLINE via PubMed, Embase, and CINAHL (February 2018). We also 
searched clinical trials databases, conference proceedings and the reference 
lists of retrieved articles for randomised controlled trials (RCT) and 
quasi-randomised trials.
SELECTION CRITERIA: Published and unpublished RCTs were eligible if they used 
random or quasi-random methods to allocate hospitalised preterm infants who were 
being fed human milk, to additional protein supplementation or no 
supplementation.
DATA COLLECTION AND ANALYSIS: Two review authors independently abstracted data, 
assessed risk of bias and the quality of evidence at the outcome level, using 
GRADE methodology. We performed meta-analyses, using risk ratio (RR) for 
dichotomous data, and mean difference (MD) for continuous data, with their 
respective 95% confidence intervals (CIs). We used a fixed-effect model and had 
planned to explore potential causes of heterogeneity via subgroup or sensitivity 
analyses.
MAIN RESULTS: We included six RCTs, involving 204 preterm infants. Low-quality 
evidence showed that protein supplementation of human milk increased in-hospital 
rates of growth in weight (MD 3.82 g/kg/day, 95% CI 2.94 to 4.7; five RCTs, 101 
infants; I² = 73%), length (MD 0.12 cm/wk, 95% CI 0.07 to 0.17; four RCTs, 68 
infants; I² = 89%), and head circumference (MD 0.06 cm/wk, 95% CI 0.01 to 0.12; 
four RCTs, 68 infants; I² = 84%). There was no evidence of a clear difference in 
rate of growth of skin fold thickness between the supplemented and 
unsupplemented groups (triceps MD 0.06 mm/wk, 95% CI -0.09 to 0.21; one RCT, 20 
infants; or subscapular MD 0.00 mm/wk, 95% CI -0.17 to 0.17; one RCT, 20 
infants). Protein supplementation led to longer hospital stays (MD 18.5 days, 
95% CI 4.39 to 32.61; one RCT, 20 infants; very low-quality evidence), and 
higher blood urea nitrogen concentrations compared to the unsupplemented group 
(MD 0.95 mmol/L, 95% CI 0.81 to 1.09; four RCTs, 81 infants; I² = 56%). Very 
low-quality evidence did not show that protein supplementation clearly increased 
the risk of feeding intolerance (RR 2.70, 95% CI 0.13 to 58.24; one RCT, 17 
infants), or necrotizing enterocolitis (RR 1.11, 95% CI 0.07 to 17.12; one RCT, 
76 infants), or clearly altered serum albumin concentrations (MD 2.5 g/L, 95% CI 
-5.66 to 10.66; one RCT, 11 infants), compared with the unsupplemented groups. 
No data were available about the effects of protein supplementation on long-term 
growth, body mass index, body composition, neurodevelopmental, or 
cardio-metabolic outcomes.
AUTHORS' CONCLUSIONS: Low-quality evidence showed that protein supplementation 
of human milk, fed to preterm infants, increased short-term growth. However, the 
small sample sizes, low precision, and very low-quality evidence regarding 
duration of hospital stay, feeding intolerance, and necrotising enterocolitis 
precluded any conclusions about these outcomes. There were no data on outcomes 
after hospital discharge. Our findings may not be generalisable to low-resource 
settings, as none of the included studies were conducted in these settings.Since 
protein supplementation of human milk is now usually done as a component of 
multi-nutrient fortifiers, future studies should compare different amounts of 
protein in multi-component fortifiers, and be designed to determine the effects 
on duration of hospital stay and safety, as well as on long-term growth, body 
composition, cardio-metabolic, and neurodevelopmental outcomes.

DOI: 10.1002/14651858.CD000433.pub2
PMCID: PMC6513468
PMID: 29931679 [Indexed for MEDLINE]

Conflict of interest statement: Emma Amissah: None known. Julie Brown: None 
known. Jane Harding: None known.


89. Cochrane Database Syst Rev. 2018 Jun 20;6(6):CD002971. doi: 
10.1002/14651858.CD002971.pub4.

Formula versus donor breast milk for feeding preterm or low birth weight 
infants.

Quigley M(1), Embleton ND, McGuire W.

Author information:
(1)National Perinatal Epidemiology Unit, University of Oxford, Old Road Campus, 
Oxford, UK, 0X3 7LF.

Update in
    Cochrane Database Syst Rev. 2019 Jul 19;7:CD002971. doi: 
10.1002/14651858.CD002971.pub5.

Comment in
    Evid Based Nurs. 2019 Jan;22(1):18. doi: 10.1136/eb-2018-102988.

Update of
    Cochrane Database Syst Rev. 2014 Apr 22;(4):CD002971. doi: 
10.1002/14651858.CD002971.pub3.

BACKGROUND: When sufficient maternal breast milk is not available, alternative 
forms of enteral nutrition for preterm or low birth weight (LBW) infants are 
donor breast milk or artificial formula. Donor breast milk may retain some of 
the non-nutritive benefits of maternal breast milk for preterm or LBW infants. 
However, feeding with artificial formula may ensure more consistent delivery of 
greater amounts of nutrients. Uncertainty exists about the balance of risks and 
benefits of feeding formula versus donor breast milk for preterm or LBW infants.
OBJECTIVES: To determine the effect of feeding with formula compared with donor 
breast milk on growth and development in preterm or low birth weight (LBW) 
infants.
SEARCH METHODS: We used the Cochrane Neonatal search strategy, including 
electronic searches of the Cochrane Central Register of Controlled Trials 
(CENTRAL; 2017, Issue 6), Ovid MEDLINE, Embase, and the Cumulative Index to 
Nursing and Allied Health Literature (until 8 June 2017), as well as conference 
proceedings and previous reviews.
SELECTION CRITERIA: Randomised or quasi-randomised controlled trials (RCTs) 
comparing feeding with formula versus donor breast milk in preterm or LBW 
infants.
DATA COLLECTION AND ANALYSIS: Two review authors assessed trial eligibility and 
risk of bias and extracted data independently. We analysed treatment effects as 
described in the individual trials and reported risk ratios (RRs) and risk 
differences (RDs) for dichotomous data, and mean differences (MDs) for 
continuous data, with respective 95% confidence intervals (CIs). We used a 
fixed-effect model in meta-analyses and explored potential causes of 
heterogeneity in subgroup analyses. We assessed the quality of evidence for the 
main comparison at the outcome level using "Grading of Recommendations 
Assessment, Development and Evaluation" (GRADE) methods.
MAIN RESULTS: Eleven trials, in which 1809 infants participated in total, 
fulfilled the inclusion criteria. Four trials compared standard term formula 
versus donor breast milk and seven compared nutrient-enriched preterm formula 
versus donor breast milk. Only the four most recent trials used 
nutrient-fortified donor breast milk. The trials contain various weaknesses in 
methodological quality, specifically concerns about allocation concealment in 
four trials and lack of blinding in most of the trials.Formula-fed infants had 
higher in-hospital rates of weight gain (mean difference (MD) 2.51, 95% 
confidence interval (CI) 1.93 to 3.08 g/kg/day), linear growth (MD 1.21, 95% CI 
0.77 to 1.65 mm/week) and head growth (MD 0.85, 95% CI 0.47 to 1.23 mm/week). We 
did not find evidence of an effect on long-term growth or neurodevelopment. 
Formula feeding increased the risk of necrotising enterocolitis (typical risk 
ratio (RR) 1.87, 95% CI 1.23 to 2.85; risk difference (RD) 0.03, 95% CI 0.01 to 
0.06).The GRADE quality of evidence was moderate for rates of weight gain, 
linear growth, and head growth (downgraded for high levels of heterogeneity) and 
was moderate for neurodevelopmental disability, all-cause mortality, and 
necrotising enterocolitis (downgraded for imprecision).
AUTHORS' CONCLUSIONS: In preterm and LBW infants, feeding with formula compared 
with donor breast milk, either as a supplement to maternal expressed breast milk 
or as a sole diet, results in higher rates of weight gain, linear growth, and 
head growth and a higher risk of developing necrotising enterocolitis. The trial 
data do not show an effect on all-cause mortality, or on long-term growth or 
neurodevelopment.

DOI: 10.1002/14651858.CD002971.pub4
PMCID: PMC6513381
PMID: 29926476 [Indexed for MEDLINE]

Conflict of interest statement: MQ: nothing to declare. NDE has conducted 
research with support from manufacturers of infant formula including Nestec SA 
(Switzerland), Wyeth UK and Nutricia UK but did not receive any payment, support 
or benefit in kind for contribution to this review. WM: nothing to declare.


90. Cochrane Database Syst Rev. 2018 Jun 19;6(6):CD000341. doi: 
10.1002/14651858.CD000341.pub2.

Fat supplementation of human milk for promoting growth in preterm infants.

Amissah EA(1), Brown J, Harding JE.

Author information:
(1)Liggins Institute, University of Auckland, Auckland, New Zealand.

Update in
    Cochrane Database Syst Rev. 2020 Aug 25;8:CD000341. doi: 
10.1002/14651858.CD000341.pub3.

Update of
    Cochrane Database Syst Rev. 2000;(2):CD000341. doi: 
10.1002/14651858.CD000341.

BACKGROUND: As preterm infants do not experience the nutrient accretion and 
rapid growth phase of the third trimester of pregnancy, they are vulnerable to 
postnatal nutritional deficits, including of fat. Consequently, they require 
higher fat intakes compared to their full term counterparts to achieve adequate 
growth and development. Human milk fat provides the major energy needs of the 
preterm infant and also contributes to several metabolic and physiological 
functions. Although human milk has many benefits for this population, its fat 
content is highly variable and may be inadequate for their optimum growth and 
development. This is a 2018 update of a Cochrane Review last published in 2000.
OBJECTIVES: To determine whether supplementation of human milk with fat compared 
with unsupplemented human milk fed to preterm infants improves growth, body 
composition, cardio-metabolic, and neurodevelopmental outcomes without 
significant adverse effects.
SEARCH METHODS: We used the standard search strategy of Cochrane Neonatal to 
search the Cochrane Central Register of Controlled Trials (CENTRAL 2018, Issue 
1), MEDLINE via PubMed (1966 to 08 February 2018), Embase (1980 to 08 February 
2018), and CINAHL (1982 to 08 February 2018). We also searched clinical trials 
databases, conference proceedings, and the reference lists of retrieved articles 
for randomised controlled trials and quasi-randomised trials.
SELECTION CRITERIA: Published and unpublished randomised controlled trials were 
eligible if they used random or quasi-random methods to allocate preterm infants 
fed human milk in hospital to supplementation or no supplementation with 
additional fat.
DATA COLLECTION AND ANALYSIS: No new randomised controlled trials matching the 
selection criteria were found but we extracted data from the previously included 
trial due to changes in review outcomes from when the protocol was first 
published. Two reviewers independently abstracted data, assessed trial quality, 
and the quality of evidence at the outcome level using the Grading of 
Recommendations Assessment, Development and Evaluation (GRADE) criteria. We 
planned to perform meta-analyses using risk ratio (RR) for dichotomous data and 
mean difference (MD) for continuous data, with their respective 95% confidence 
intervals (CIs). We planned to use a fixed-effect model and to explore potential 
causes of heterogeneity via sensitivity analyses.
MAIN RESULTS: One randomised trial involving 14 preterm infants was included. 
There was no evidence of a clear difference between the fat-supplemented and 
unsupplemented groups in in-hospital rates of growth in weight (MD 0.6 g/kg/day, 
95% CI -2.4 to 3.6; 1 RCT, n = 14 infants, very low-quality evidence), length 
(MD 0.1 cm/week, 95% CI -0.08 to 0.3; 1 RCT, n = 14 infants, very low-quality 
evidence) and head circumference (MD 0.2 cm/week, 95% CI -0.07 to 0.4; 1 RCT n = 
14 infants, very low-quality evidence). There was no clear evidence that fat 
supplementation increased the risk of feeding intolerance (RR 3.0, 95% CI 0.1 to 
64.3; 1 RCT, n = 16 infants, very low-quality evidence). No data were available 
regarding the effects of fat supplementation on long-term growth, body mass 
index, body composition, neurodevelopmental, or cardio-metabolic outcomes.
AUTHORS' CONCLUSIONS: The one included trial suggests no evidence of an effect 
of fat supplementation of human milk on short-term growth and feeding 
intolerance in preterm infants. However, the very low-quality evidence, small 
sample size, few events, and low precision diminishes our confidence that these 
results reflect the true effect of fat supplementation of human milk in preterm 
infants, and no long-term outcomes were reported. Further high-quality research 
should evaluate the effect on short and long-term growth, neurodevelopmental and 
cardio-metabolic outcomes in the context of the development of multicomponent 
fortifiers. Optimal dosage, adverse effects, and delivery practices should also 
be evaluated.

DOI: 10.1002/14651858.CD000341.pub2
PMCID: PMC6513401
PMID: 29924388 [Indexed for MEDLINE]

Conflict of interest statement: Emma Amissah: None known. Julie Brown: None 
known. Jane Harding: None known.


91. Nutrients. 2018 Feb 20;10(2):238. doi: 10.3390/nu10020238.

Donor Human Milk Protects against Bronchopulmonary Dysplasia: A Systematic 
Review and Meta-Analysis.

Villamor-Martínez E(1), Pierro M(2), Cavallaro G(3), Mosca F(4), Kramer BW(5), 
Villamor E(6).

Author information:
(1)Department of Pediatrics, Maastricht University Medical Center (MUMC+), 
School for Oncology and Developmental Biology (GROW), 6202 AZ Maastricht, The 
Netherlands..
(2)Neonatal Intensive Care Unit, Alessandro Manzoni Hospital, 23900 Lecco, 
Italy..
(3)Neonatal Intensive Care Unit, Department of Clinical Sciences and Community 
Health, Fondazione IRCCS Cà Granda Ospedale Maggiore Policlinico, Università 
degli Studi di Milano, 20122 Milan, Italy..
(4)Neonatal Intensive Care Unit, Department of Clinical Sciences and Community 
Health, Fondazione IRCCS Cà Granda Ospedale Maggiore Policlinico, Università 
degli Studi di Milano, 20122 Milan, Italy..
(5)Department of Pediatrics, Maastricht University Medical Center (MUMC+), 
School for Oncology and Developmental Biology (GROW), 6202 AZ Maastricht, The 
Netherlands..
(6)Department of Pediatrics, Maastricht University Medical Center (MUMC+), 
School for Oncology and Developmental Biology (GROW), 6202 AZ Maastricht, The 
Netherlands..

Bronchopulmonary dysplasia (BPD) is the most common complication after preterm 
birth. Pasteurized donor human milk (DHM) has increasingly become the standard 
of care for very preterm infants over the use of preterm formula (PF) if the 
mother's own milk (MOM) is unavailable. Studies have reported beneficial effects 
of DHM on BPD. We conducted a systematic review and meta-analysis of randomized 
controlled trials (RCTs) and observational studies on the effects of DHM on BPD 
and other respiratory outcomes. Eighteen studies met the inclusion criteria. 
Meta-analysis of RCTs could not demonstrate that supplementation of MOM with DHM 
reduced BPD when compared to PF (three studies, risk ratio (RR) 0.89, 95% 
confidence interval (CI) 0.60-1.32). However, meta-analysis of observational 
studies showed that DHM supplementation reduced BPD (8 studies, RR 0.78, 95% CI 
0.67-0.90). An exclusive human milk diet reduced the risk of BPD, compared to a 
diet with PF and/or bovine milk-based fortifier (three studies, RR 0.80, 95% CI 
0.68-0.95). Feeding raw MOM, compared to feeding pasteurized MOM, protected 
against BPD (two studies, RR 0.77, 95% CI 0.62-0.96). In conclusion, our data 
suggest that DHM protects against BPD in very preterm infants.

DOI: 10.3390/nu10020238
PMCID: PMC5852814
PMID: 29461479 [Indexed for MEDLINE]

Conflict of interest statement: The authors declare no conflict of interest.


92. Cochrane Database Syst Rev. 2017 Feb 26;2(2):CD003310. doi: 
10.1002/14651858.CD003310.pub2.

Calcium and phosphorus supplementation of human milk for preterm infants.

Harding JE(1), Wilson J(1), Brown J(1).

Author information:
(1)Liggins Institute, The University of Auckland, Auckland, New Zealand, 1001.

Update of
    Cochrane Database Syst Rev. 2001;(4):CD003310. doi: 
10.1002/14651858.CD003310.

BACKGROUND: Preterm infants are born with low skeletal stores of calcium and 
phosphorus. Preterm human milk provides insufficient calcium and phosphorus to 
meet the estimated needs of preterm infants for adequate growth. Supplementation 
of human milk with calcium and phosphorus may improve growth and development of 
preterm infants.
OBJECTIVES: To determine whether addition of calcium and phosphorus supplements 
to human milk leads to improved growth and bone metabolism of preterm infants 
without significant adverse effects.
SEARCH METHODS: We used the standard search strategy of the Cochrane Neonatal 
Review Group to search the Cochrane Central Register of Controlled Trials 
(CENTRAL; 2016, Issue 3), MEDLINE via PubMed (1966 to 14 April 2016), Embase 
(1980 to 14 April 2016) and the Cumulative Index to Nursing and Allied Health 
Literature (CINAHL; 1982 to 14 April 2016). We also searched clinical trials 
databases (11 May 2016) and the reference lists of retrieved articles for 
randomised controlled trials and quasi-randomised trials.
SELECTION CRITERIA: Randomised and quasi-randomised trials comparing 
supplementation of human milk with calcium and/or phosphorus versus no 
supplementation in hospitalised preterm infants were eligible for inclusion in 
this review.
DATA COLLECTION AND ANALYSIS: Two review authors (JB, JW) independently 
extracted data and assessed trial quality using standard methods of the Cochrane 
Neonatal Review Group. We reported dichotomous data as risk ratios (RRs) and 
continuous data as mean differences (MDs) with 95% confidence intervals (CIs). 
We used the Grading of Recommendations Assessment, Development and Evaluation 
(GRADE) approach to assess the quality of evidence.
MAIN RESULTS: This is an update of a 2001 review that identified no eligible 
trials. One trial including 40 infants met the inclusion criteria for this 
review. Using GRADE criteria, we judged the quality of the evidence as low owing 
to risk of bias (inadequate reporting of methods of randomisation, allocation 
concealment and/or blinding) and imprecision (wide confidence intervals and data 
from a single small trial). We found no evidence of a difference between calcium 
and phosphorus supplementation versus no supplementation for neonatal growth 
outcomes (weight, length, head circumference) at any time point reported (two, 
four or six weeks postnatal age). At six weeks postnatal age, supplementation 
with calcium/phosphorus was associated with a decrease in serum alkaline 
phosphatase concentration (MD -56.85 IU/L, 95% CI -101.27 to -12.43; one 
randomised controlled trial (RCT); n = 40 infants). Investigators provided no 
data on growth at 12 to 18 months, neonatal fractures, feed intolerance, 
breastfeeding or any of the prespecified childhood outcomes for this review 
(fractures, growth, neurodevelopmental outcomes).
AUTHORS' CONCLUSIONS: We identified one small trial including only 40 infants 
that compared supplementation of human milk with calcium and phosphorus versus 
no supplementation in hospitalised preterm infants. We judged the evidence to be 
of low quality and found no evidence of differences between groups for 
clinically important outcomes including growth and fractures. Although serum 
alkaline phosphatase concentration was reduced in the group receiving 
supplementation at six weeks postnatal age, this difference is unlikely to be of 
clinical significance. We conclude that evidence is insufficient to determine 
whether benefit or harm ensues when human milk is supplemented with calcium 
and/or phosphorus for the hospitalised preterm infant. We see no advantage of 
conducting further trials of this intervention because with the advent of 
multi-component human milk fortifier, supplementation of human milk with calcium 
and/or phosphorus alone is no longer common practice. Future trials should 
consider assessing effects of multi-component fortifiers with different mineral 
compositions on clinically important outcomes during the neonatal period and in 
later childhood.

DOI: 10.1002/14651858.CD003310.pub2
PMCID: PMC6464224
PMID: 28238222 [Indexed for MEDLINE]

Conflict of interest statement: None known.


93. Clin Perinatol. 2017 Mar;44(1):209-267. doi: 10.1016/j.clp.2016.11.001. Epub 
2016 Dec 29.

"Bed Side" Human Milk Analysis in the Neonatal Intensive Care Unit: A Systematic 
Review.

Fusch G(1), Kwan C(1), Kotrri G(1), Fusch C(2).

Author information:
(1)Division of Neonatology, Department of Pediatrics, McMaster University, 1280 
Main Street West, Hamilton, Ontario L8S 4L8, Canada.
(2)Division of Neonatology, Department of Pediatrics, McMaster University, 1280 
Main Street West, Hamilton, Ontario L8S 4L8, Canada; Department of Pediatrics, 
Paracelsus Medical School, General Hospital of Nuremberg, Breslauer Straße 201, 
Nuremberg 90471, Germany. Electronic address:.

Human milk analyzers can measure macronutrient content in native breast milk to 
tailor adequate supplementation with fortifiers. This article reviews all 
studies using milk analyzers, including (i) evaluation of devices, (ii) the 
impact of different conditions on the macronutrient analysis of human milk, and 
(iii) clinical trials to improve growth. Results lack consistency, potentially 
due to systematic errors in the validation of the device, or pre-analytical 
sample preparation errors like homogenization. It is crucial to introduce good 
laboratory and clinical practice when using these devices; otherwise a 
non-validated clinical usage can severely affect growth outcomes of infants.

Copyright © 2016 Elsevier Inc. All rights reserved.

DOI: 10.1016/j.clp.2016.11.001
PMID: 28159207 [Indexed for MEDLINE]


94. Clin Perinatol. 2017 Mar;44(1):173-178. doi: 10.1016/j.clp.2016.11.011. Epub 
2016 Dec 24.

The Use of Multinutrient Human Milk Fortifiers in Preterm Infants: A Systematic 
Review of Unanswered Questions.

Mimouni FB(1), Nathan N(2), Ziegler EE(3), Lubetzky R(2), Mandel D(4).

Author information:
(1)Department of Neonatology, Shaare Zedek Medical Center, 12 Shmuel Bait 
Street, Jerusalem 913102, Israel; Sackler Faculty of Medicine, Tel Aviv Sourasky 
Medical Center, Tel Aviv University, Tel Aviv, Israel.
(2)Sackler Faculty of Medicine, Tel Aviv Sourasky Medical Center, Tel Aviv 
University, Tel Aviv, Israel; Department of Pediatrics, Dana Dwek Children's 
Hospital, Tel Aviv Sourasky Medical Center, 6 Weizman Street, Tel Aviv 6423906, 
Israel.
(3)Department of Pediatrics, University of Iowa, Iowa City, IA, USA.
(4)Sackler Faculty of Medicine, Tel Aviv Sourasky Medical Center, Tel Aviv 
University, Tel Aviv, Israel; Department of Neonatology, Dana Dwek Children's 
Hospital, Tel Aviv Sourasky Medical Center, 6 Weizman Street, Tel Aviv 6423906, 
Israel. Electronic address:.

There is evidence that multinutrient fortification of human milk increases 
in-hospital growth of preterm infants, but fortification has not been shown to 
improve long-term growth and neurodevelopmental outcome. We aimed to ascertain 
whether randomized controlled trials have determined the effect of early versus 
late introduction of fortifiers on growth and/or other outcomes, and have 
compared the efficacy/adverse effects of human milk-based versus cow milk-based 
fortifiers. We conclude that there is little evidence that early introduction of 
human milk fortification affects important outcomes, and limited evidence that a 
bovine fortifier places the infant at a higher risk of NEC.

Copyright © 2016 Elsevier Inc. All rights reserved.

DOI: 10.1016/j.clp.2016.11.011
PMID: 28159204 [Indexed for MEDLINE]


95. Public Health Nutr. 2017 May;20(7):1214-1225. doi: 10.1017/S1368980016003189. 
Epub 2016 Dec 12.

Effect of fortified milk on growth and nutritional status in young children: a 
systematic review and meta-analysis.

Matsuyama M(1), Harb T(1), David M(2), Davies PS(1), Hill RJ(1).

Author information:
(1)1Children's Nutrition Research Centre,Child Health Research Centre,The 
University of Queensland,Level 6,Centre for Children's Health Research,63 Graham 
Street,South Brisbane,QLD 4101,Australia.
(2)2School of Public Health,The University of Queensland,Herston,QLD,Australia.

OBJECTIVE: Adequate nutrition is critical for optimal growth and development. 
However, young children may be at risk of nutrient deficiencies when 
transitioning to weaning foods for a variety of reasons. Supplementation with 
fortified milk may provide potentially lacking essential nutrients, but effects 
on growth and nutritional status are yet to be established.
DESIGN: Five databases were searched for randomised controlled trials using 
fortified milk against control milk in young children. Outcomes were growth, 
body composition and/or biochemical markers. Pooled differences in means were 
calculated for continuous outcomes and odds ratios for binary outcomes.
SETTING: Randomised controlled trials set in any country.
SUBJECTS: Otherwise healthy children aged 6-47 months.
RESULTS: Fifteen articles met the eligibility criteria. Fortification varied 
from Fe, Zn, vitamins, essential fatty acids, to pre- and/or probiotics.
