## Supplementary material for "Cross-LLM AI platform meta-research: Non-inferiority of bovine milk-based fortifiers to human milk-based fortifiers": SI3_RoB_2.0.pdf

### Revised Cochrane risk-of-bias tool for randomized trials (RoB 2)

Edited by Julian PT Higgins, Jelena Savović, Matthew J Page, Jonathan AC Sterne
on behalf of the RoB2 Development Group

**22 August 2019**

**Dedicated to Professor Douglas G Altman, whose contributions were of fundamental importance to development of risk of bias assessment in systematic reviews**

This work is licensed under a Creative Commons Attribution-NonCommercial-NoDerivatives 4.0 International License.

#### Contents

1 Introduction ... 2
1.1 Signalling questions ... 3
1.2 Risk-of-bias judgements ... 3
1.3 Specifying the nature of the effect of interest ... 5
2 Issues in implementation of RoB 2 ... 6
2.1 Multiple assessments ... 6
2.2 The data collection process ... 7
2.3 Presentation of risk-of-bias assessments ... 7
2.4 Rapid assessments ... 7
3 Detailed guidance: preliminary considerations ... 8
4 Detailed guidance: bias arising from the randomization process ... 10
4.1 Background ... 10
4.2 Empirical evidence of bias arising from the randomization process ... 11
4.3 Using this domain of the tool ... 12
4.4 Signalling questions and criteria for judging risk of bias ... 16
5 Detailed guidance: bias due to deviations from intended interventions ... 21
5.1 Background ... 21
5.2 Empirical evidence of bias due to deviations from intended interventions ... 26
5.3 Using this domain of the tool ... 26
5.4 Signalling questions and criteria for judging risk of bias ... 27
6 Detailed guidance: bias due to missing outcome data ... 39
6.1 Background ... 39
6.2 Empirical evidence of bias due to missing outcome data ... 44
6.3 Using this domain of the tool ... 44
6.4 Signalling questions and criteria for judging risk of bias ... 44
7 Detailed guidance: bias in measurement of the outcome ... 49
7.1 Background ... 49
7.2 Empirical evidence of bias in measurement of the outcome ... 52
7.3 Using this domain of the tool ... 52
7.4 Signalling questions and criteria for judging risk of bias ... 53
8 Detailed guidance: bias in selection of the reported result ... 58
8.1 Background ... 58
8.2 Empirical evidence of bias in selection of the reported result ... 60
8.3 Using this domain of the tool ... 60
8.4 Signalling questions and criteria for judging risk of bias ... 62
9 Acknowledgements ... 68
10 Contributors ... 68
11 References ... 68

### 1 Introduction

The RoB 2 tool provides a framework for considering the risk of bias in the findings of any type of randomized trial. The assessment is specific to a single trial result that is an estimate of the relative effect of two interventions or intervention strategies on a particular outcome. We refer to the interventions as the **experimental intervention** and the **comparator intervention**, although we recognize that the result may sometimes refer to a comparison of two active interventions.

The tool is structured into five domains through which bias might be introduced into the result. These were identified based on both empirical evidence (see Box 1) and theoretical considerations. Because the domains cover all types of bias that can affect results of randomized trials, each is mandatory, and no further domains should be added. The five domains for individually randomized trials (including cross-over trials) are:

(1) bias arising from the randomization process;
(2) bias due to deviations from intended interventions;
(3) bias due to missing outcome data;
(4) bias in measurement of the outcome;
(5) bias in selection of the reported result.

The domain names are direct descriptions of the causes of bias addressed in the domain. We have avoided many of the terms used in version 1 of the tool (e.g. selection bias, performance bias, attrition bias, detection bias) because they do not describe the specific issues addressed and so cause confusion (1).

We offer several templates for addressing these domains, tailored to the following study designs:

(1) randomized parallel-group trials;
(2) cluster-randomized parallel-group trials (including stepped-wedge designs);
(3) randomized cross-over trials and other matched designs.

For cluster-randomized trials, an additional domain is included ((1b) Bias arising from identification or recruitment of individual participants within clusters).

This document describes the main features of the RoB 2 tool and provides guidance for its application to individually randomized parallel-group trials. Supplementary documents address additional considerations for cluster-randomized parallel-group trials and individually-randomized cross-over trials. We have not yet developed a version appropriate for cluster cross-over trials.

**Box 1: Empirical evidence of bias in randomized trials: the role of meta-epidemiology**

| |
|---|
| Empirical evidence of bias in randomized trials comes from a field known as **meta-epidemiology** (2). A meta-epidemiological study analyses the results of a large collection of previous studies to understand how methodological characteristics of the studies are associated with their results. The first well-known meta-epidemiological study examined 33 meta-analyses containing 250 clinical trials (3). Each trial was categorized on the basis of four characteristics: whether sequence generation was reported to have a random component; whether allocation was reported to be adequately concealed, whether the trial was described as double-blind, and whether the trial reported exclusion of participants from its analysis. For each of the four characteristics separately, trials were compared within each meta-analysis to estimate a ratio of odds ratios among the 'better' versus the 'worse' trials, and these ratios of odds ratios were combined across the 33 meta-analyses. Numerous similar studies have been undertaken since, examining many study characteristics that are potentially associated with biases in results. More recent analyses address both the average and the variability in bias associated with the characteristic under investigation. Specifically, they examine also the extent to which the characteristic is associated with increased between-trial heterogeneity and to which the average bias varies between meta-analyses (4). In several places in this document we refer to empirical evidence from meta-epidemiological studies, or systematic reviews of them, to support the selection of domains and signalling questions. |

#### 1.1 Signalling questions

Inclusion of signalling questions within each domain of bias is a key feature of RoB 2. Signalling questions aim to elicit information relevant to an assessment of risk of bias. They seek to be reasonably factual in nature. Responses to these questions feed into algorithms we have developed to guide users of the tool to judgements about the risk of bias.

The **response options for the signalling questions** are:

(1) Yes;
(2) Probably yes;
(3) Probably no;
(4) No;
(5) No information;

To maximize the signalling questions' simplicity and clarity, they are phrased such that a response of 'Yes' may be indicative of either a low or high risk of bias, depending on the most natural way to ask the question.

Responses of 'Yes' and 'Probably yes' have the same implications for risk of bias, as do responses of 'No' and 'Probably no'. The definitive versions ('Yes' and 'No') would typically imply that firm evidence is available in relation to the signalling question; the 'Probably' versions would typically imply that a judgement has been made. If review authors calculate measures of agreement (e.g. kappa statistics) for the answers to the signalling questions, we recommend treating 'Yes' and 'Probably yes' as the same response and 'No' and 'Probably no' as the same response.

The 'No information' response should be used only when both (i) insufficient details are reported to permit a response of 'Probably yes' or 'Probably no', and (ii) in the absence of these details it would be unreasonable to respond 'Probably yes' or 'Probably no' in the circumstances of the trial. For example, in the context of a large trial run by an experienced clinical trials unit, absence of specific information about generation of the randomization sequence, in a paper published in a journal with rigorously enforced word count limits, is likely to result in a response of 'Probably yes' rather than 'No information' to the signalling question about sequence generation. The implications for risk of bias judgements of a 'No information' response to a signalling question differ according to the purpose of the question. If the question seeks to identify evidence of a problem, then 'No information' corresponds to no evidence of that problem. If the question relates to an item that is expected to be reported (such as whether any participants were lost to follow up), then the absence of information leads to concerns about there being a problem.

For signalling questions that are answered only if the response to a previous question implies that they are required, a response option 'Not applicable' is available. Signalling questions should be answered independently: the answer to one question should not affect answers to other questions in the same or other domains other than through determining which subsequent questions are answered.

##### *1.1.1 Free-text boxes alongside signalling questions*

The tool provides space for free text alongside the signalling question. In some instances, when the same information is likely to be used to answer more than one question, one text box covers more than one question. These boxes should be used to provide support for the answer to each signalling question. Brief **direct quotations** from the text of the study report should be used whenever possible.

#### 1.2 Risk-of-bias judgements

##### *1.2.1 Domain-level judgements about risk of bias*

RoB 2 is conceived hierarchically: responses to signalling questions elicit what happened and provide the basis for domain-level judgements about the risk of bias. In turn, these domain-level judgements provide the basis for an overall risk-of-bias judgement for the specific trial result being assessed.

The tool includes algorithms that map responses to signalling questions onto a proposed risk-of-bias judgement for each domain. The possible **risk-of-bias judgements** are:

(1) Low risk of bias;
(2) Some concerns; and
(3) High risk of bias.

Use of the word 'judgement' is important for the risk-of-bias assessment. In particular, the algorithms provide proposed judgements, but users should verify these and change them if they feel this is appropriate. In reaching final judgements, the following considerations apply:

- 'Risk of bias' is to be interpreted as '**risk of material bias**'. That is, concerns should be expressed only about issues that are likely to affect the ability to draw reliable conclusions from the study.
- Domain-level judgements about risk of bias should have the same implication for each of the six domains with respect to concern about the impact of bias on the trustworthiness of the result. A judgement of 'High' risk of bias for any individual domain will lead to the result being at 'High' risk of bias overall, and a judgement of 'Some concerns' for any individual domain will lead to the result being at 'Some concerns', or 'High' risk, overall (see 1.2.3).

##### 1.2.2 *Direction of bias*

The tool includes optional judgements of the direction of the bias for each domain and overall. For some domains, the bias is most easily thought of as being towards or away from the null. For example, high levels of switching of participants from their assigned intervention to the other intervention would be likely to lead to the estimated effect of adhering to intervention being biased towards the null. For other domains, the bias is likely to favour one of the interventions being compared, implying an increase or decrease in the effect estimate depending on which intervention is favoured. Examples include manipulation of the randomization process, awareness of interventions received influencing the outcome assessment and selective reporting of results. **If review authors do not have a clear rationale for judging the likely direction of the bias, they should not guess it.**

##### 1.2.3 *Reaching an overall judgement about risk of bias*

The response options for an overall risk-of-bias judgement are the same as for individual domains. Table 1 shows the basic approach to mapping risk-of-bias judgements within domains to an overall judgement across domains for the outcome.

**Table 1. Reaching an overall risk-of-bias judgement for a specific outcome.**

| Overall risk-of-bias judgement | Criteria |
|---|---|
| Low risk of bias | The study is judged to be at **low risk of bias for all domains** for this result. |
| Some concerns | The study is judged to raise **some concerns** in at least one domain for this result, but not to be at high risk of bias for any domain. |
| High risk of bias | The study is judged to be at **high risk of bias** in at least one domain for this result.<br>Or<br>The study is judged to have **some concerns** for **multiple domains** in a way that substantially lowers confidence in the result. |

**Judging a result to be at a particular level of risk of bias for an individual domain implies that the result has an overall risk of bias at least this severe**. Therefore, a judgement of 'High' risk of bias within any domain should have similar implications for the result as a whole, irrespective of which domain is being assessed. 'Some concerns' in multiple domains may lead the review authors to decide on an overall judgement of 'High' risk of bias for that outcome or group of outcomes.

##### 1.2.4 *Free-text boxes alongside risk-of-bias judgements*

There is space for free text alongside each risk-of-bias judgement to explain the reasoning that underpins the judgement. It is particularly important that reasons are provided for any judgements that do not follow the proposed algorithms.

#### 1.3 Specifying the nature of the effect of interest

Assessments for the domain 'Bias due to deviations from intended interventions' vary according to whether review authors are interested in quantifying:

(1) the effect of **assignment** to the interventions at baseline (regardless of whether the interventions are received during follow-up, sometimes known as the 'intention-to-treat effect'); or
(2) the effect of **adhering to** intervention as specified in the trial protocol (sometimes known as the 'per-protocol effect') (5).

These effects will differ some participants do not receive their assigned intervention or deviate from the assigned intervention after baseline.

Each of these two effects may be of interest. For example, the estimated effect of assignment to intervention may be the most appropriate to inform a health policy question about whether to recommend an intervention in a particular health system (e.g. whether to instigate a screening programme, or whether to prescribe a new cholesterol-lowering drug), whereas the estimated effect of adhering to intervention as specified in the trial protocol may more directly inform a care decision by an individual patient (e.g. whether to be screened, or whether to take the new drug).

Review authors should define the intervention effect in which they are interested, and apply the risk-of-bias tool appropriately to this effect. When assessing the effect of adhering to intervention, review authors should specify what types of deviations from the intended intervention will be examined: these will be one or more of (i) occurrence of non-protocol interventions; (ii) failures in implementing the intervention that could affect the outcome; and (iii) non-adherence by trial participants to their assigned intervention (see section 5.3). For example, the START randomized trial compared immediate with deferred initiation of antiretroviral therapy (ART) in HIV-positive individuals, but 30% of those assigned to deferred initiation started ART earlier than the protocol specified (6). Lodi and colleagues estimated a per-protocol effect that adjusted for these protocol deviations, but not for whether participants continued antiretroviral therapy throughout trial follow-up (7). In such an example, review authors might specify that occurrence of non-protocol interventions, but not non-adherence to assigned intervention by trial participants, would be addressed in their risk of bias assessments.

The effect of principal interest should be specified in the review protocol. On occasion, review authors may be interested in both effects of interest.

Note that specification of the 'effect of interest' in RoB 2 does not relate to the choice of treatment effect metric (odds ratio, risk difference etc.).

##### *1.3.1 Estimating the effect of interest*

Authors of randomized trials can use several analytical approaches to estimate interventions effects. They may not explain the reasons for their choice of analysis approach, or whether their aim is to estimate the effect of assignment or adherence to intervention. We discuss different approaches to analysis and their implications for bias. Because multiple analyses can be reported, we also suggest an order of preference in which estimated effects of intervention should be chosen when review authors are interested in the effect of assignment to intervention.

The effect of **assignment** to intervention should be estimated by an **intention-to-treat (ITT) analysis** that includes all randomized participants (8). The principles of ITT analyses are (9, 10):

(1) analyse participants in the intervention groups to which they were randomized, regardless of the intervention they actually received; and
(2) include all randomized participants in the analysis, which requires measuring all participants' outcomes.

An ITT analysis maintains the benefit of randomization that, on average, the intervention groups do not differ at baseline with respect to measured or unmeasured prognostic factors. Note that the term 'intention-to-treat analysis' does not have a consistent definition, and is used inconsistently in study reports (11-13).

In a blinded, placebo-controlled trial in which there is non-adherence to assigned interventions, an ITT analysis is expected to underestimate the intervention effect that would have been seen had all participants adhered (the per-protocol effect), which is problematic for non-inferiority or equivalence studies. However the ITT effect estimate may overestimate the per-protocol effect in trials comparing two or more active interventions, and when interventions are in different directions for different participants (14, 15). Underestimation of the effect if all participants had adhered to the intervention may be particularly problematic when examining harms (adverse effects) of the experimental intervention. Variable rates of non-adherence to assigned intervention may also be a source of heterogeneity in ITT estimates of intervention effects: we expect greater effectiveness in a trial with perfect adherence than in a trial in which a substantial proportion of participants do not adhere (5).

Patients and other stakeholders are often interested in the effect of adhering to intervention as described in the trial protocol (the 'per protocol effect'). It is possible to use data from a randomized trial to derive an unbiased estimate of the effect of adhering to intervention, but appropriate methods require strong assumptions and published applications are relatively rare to date (5). Importantly, commonly used approaches to adherence adjustment may result in biased estimates of per-protocol effects, , because they do not include adjustment for prognostic factors that may influence whether individuals receive their assigned intervention (14). These include:

- naïve 'per protocol' analyses restricted to individuals in each intervention group who adhered to their assigned intervention; and
- 'as-treated' analyses in which participants are analysed according to the intervention they actually received, even if their assigned intervention group was different.

Trial investigators often estimate the effect of intervention using more than one approach. **We recommend that when the effect of interest is that of assignment to intervention, the trial result included in meta-analyses, and assessed for risk of bias, should be chosen according to the following order of preference:**

(1) The result corresponding to a full ITT analysis, as defined above;
(2) The result corresponding to an analysis (sometimes described as a 'modified intention-to-treat' (mITT) analysis) that adheres to ITT principles except that participants with missing outcome data are excluded (see section 5.3.1). Such an analysis does not prevent bias due to missing outcome data, which is addressed in the corresponding domain;
(3) A result corresponding to an 'as treated' or naïve 'per-protocol' analysis, or an analysis from which eligible trial participants were excluded.

Valid estimation of per-protocol effects usually requires data on what deviations from intended intervention occurred, as well as adjustment for prognostic factors that predict deviations from intended intervention. An increased focus on measuring these factors during trial follow up may facilitate more frequent estimation of per-protocol effects in the future. In trials comparing interventions that are sustained over time, valid estimation will generally require appropriate adjustment for pre- and post-randomization values of prognostic factors. Because conventional statistical methods, such as standard regression models, are not valid if post-randomization prognostic factors are affected by prior intervention, 'g-methods' such as inverse probability weighting and the g-formula must generally be used (5, 14, 15).

For trials with an intervention that is administered only at baseline and with all-or-nothing adherence, methods that use randomization status as an instrumental variable may be used to estimate the per-protocol effect. Instrumental variable methods require data on adherence and based on strong assumptions (5) but, if these assumption are met, bypass the need to adjust for prognostic factors that predict receipt of intervention. For example, a randomized trial of the effect of flexible sigmoidoscopy screening on colorectal cancer incidence and mortality reported both a primary ITT analysis and a secondary, instrumental variable, analysis estimating the effect of screening adjusted for nonadherence (16).

For each effect of interest, a signalling question in the domain 'Bias due to deviations from intended interventions' asks whether appropriate statistical methods were used to estimate that effect.

### 2 Issues in implementation of RoB 2

#### 2.1 Multiple assessments

Trials usually contribute multiple results to a systematic review, mainly through contributing to multiple outcomes. Therefore, several risk-of-bias assessments may be needed for each study. We have not yet formulated recommendations on which results should be targeted with an assessment, or how many results should be assessed. However, these decisions are likely to align with the outcomes included in a Summary of Findings table.

#### 2.2 The data collection process

Assessment of risk of bias is specific to a particular result, for a particular outcome measured at a particular time, from the study. However, some causes of bias (such as biases arising from the randomization process) apply generally to the whole study; some (such as bias due to deviations from intended intervention) apply mainly to the outcome being measured; some (such as bias in measurement of outcomes) apply mainly to the outcome measurement method used; and some (such as bias in selection of the reported result) apply to the specific result. This has implications for how review authors can most efficiently extract information relevant to risk of bias from study reports.

#### 2.3 Presentation of risk-of-bias assessments

We suggest that RoB 2 assessments are presented as follows. More work is required in this area.

- For full transparency of the process, review authors may wish to present the answers, free-text supports and judgements for each assessor separately. Since these may be confusing to the reader, we recommend that they are not presented prominently, so might be included in an appendix or supplementary document.
- Present the domain-level judgements in the main review document (e.g. as a table, or a figure, or within a forest plot of the results). Only consensus judgements across multiple assessors should be presented. If space permits, abridged free-text justifications for each judgement would be an attractive supplement to this within the main review document.
- Provide answers for each signalling question and the free text support for each of these answers in an appendix or supplementary document. Only consensus answers across multiple assessors should be presented.

#### 2.4 Rapid assessments

Because the default overall judgement for the result will be 'High' risk of bias if one of the domains is judged at 'High' risk of bias, users of the tool may be tempted to stop their assessment as soon as one domain is judged as 'High'. We discourage this when the tool is used in the context of a full systematic review, for several reasons. First, many readers of systematic reviews prefer to see full and consistent evaluations of the included evidence, in the interests of transparency. Second, full evaluations of the limitations of existing randomized trials are likely to be useful in the design and conduct of future trials of the intervention(s) in question. Third, there is a drive from the research community to make risk-of-bias assessments of trials produced by review authors publicly available alongside trial results; a fully documented domain-level assessment is needed for this. A more minor consideration for review authors is that meta-epidemiological studies, which re-analyse multiple meta-analyses to learn about the impact of trial design features, and are invaluable sources of information about the size and direction of biases introduced by study limitations, require full assessments for each domain of the tool (17).

We recognize that some users of the tool may need to introduce 'stopping rules' into their assessment when the sole purpose is to reach a rapid judgement about whether the trial is at 'High' risk of bias. We recommend that this be done only when it has been pre-specified in the protocol that trials judged to be at 'High' risk of bias will play no role in the synthesis of evidence. If trials are to be included in sensitivity analyses or subgroup analyses, then we recommend that full assessments be made so that the study can be appropriately characterized.

### 3 Detailed guidance: preliminary considerations

Before completing the risk-of-bias assessment, it is helpful to document important characteristics of the assessment, such as the design of the trial, the outcome being assessed (as well as the specific result being assessed), and whether interest focusses on the effect of *assignment to intervention* or the effect of *adhering to intervention*. Review authors should document the sources that are used to complete the assessment (as many sources as possible should be used in practice). The RoB 2 standard template includes questions to capture these details (Box 2), and to ensure clarity on which intervention is being referred to as ‘experimental’ and which as ‘comparator’ within the assessment.

#### Box 2. The RoB 2 tool (part 1): Preliminary considerations

**Study design**

- ☐ Individually-randomized parallel-group trial
- ☐ Cluster-randomized parallel-group trial
- ☐ Individually randomized cross-over (or other matched) trial

**For the purposes of this assessment, the interventions being compared are defined as**

: Experimental: [ ] Comparator: [ ]

**Specify which outcome is being assessed for risk of bias** [ ]

**Specify the numerical result being assessed.** In case of multiple alternative analyses being presented, specify the numeric result (e.g. RR = 1.52 (95% CI 0.83 to 2.77) and/or a reference (e.g. to a table, figure or paragraph) that uniquely defines the result being assessed. [ ]

**Is the review team's aim for this result…?**

- ☐ to assess the effect of *assignment to intervention* (the 'intention-to-treat' effect)
- ☐ to assess the effect of *adhering to intervention* (the 'per-protocol' effect)

**If the aim is to assess the effect of *adhering to intervention***, select the deviations from intended intervention that should be addressed (at least one must be checked):

- ☐ occurrence of non-protocol interventions
- ☐ failures in implementing the intervention that could have affected the outcome
- ☐ non-adherence to their assigned intervention by trial participants

**Which of the following sources were <u>obtained</u> to help inform the risk-of-bias assessment? (tick as many as apply)**

- ☐ Journal article(s)
- ☐ Trial protocol
- ☐ Statistical analysis plan (SAP)
- ☐ Non-commercial trial registry record (e.g. ClinicalTrials.gov record)
- ☐ Company-owned trial registry record (e.g. GSK Clinical Study Register record)
- ☐ 'Grey literature' (e.g. unpublished thesis)
- ☐ Conference abstract(s) about the trial
- ☐ Regulatory document (e.g. Clinical Study Report, Drug Approval Package)
- ☐ Research ethics application
- ☐ Grant database summary (e.g. NIH RePORTER or Research Councils UK Gateway to Research)
- ☐ Personal communication with trialist
- ☐ Personal communication with the sponsor

### 4 Detailed guidance: bias arising from the randomization process

#### 4.1 Background

If successfully accomplished, randomization avoids an influence of either known or unknown prognostic factors (factors that predict the outcome, such as severity of illness or presence of comorbidities) on intervention group assignment. This means that, on average, the intervention groups have the same prognosis before the start of intervention. If prognostic factors influence the intervention group to which participants are assigned then the estimated effect of intervention will be biased by 'confounding', which occurs when there are common causes of intervention group assignment and outcome. Confounding is an important potential cause of bias in intervention effect estimates from observational studies, because treatment decisions in routine care are often influenced by prognostic factors.

To randomize participants into a study, an allocation sequence that specifies how participants will be assigned to interventions is generated, based on a process that includes an element of chance. We call this process **allocation sequence generation**. Subsequently, steps must be taken to prevent participants or trial personnel from knowing the forthcoming allocations until after recruitment has been confirmed. This process is often called **allocation sequence concealment**.

Knowledge of the next assignment (e.g. if the sequence is openly posted on a bulletin board) can enable selective enrolment of participants on the basis of prognostic factors. Participants who would have been assigned to an intervention deemed to be 'inappropriate' may be rejected. In epidemiological terms this is a type of selection bias. Other participants may be directed to the 'appropriate' intervention, which can be accomplished by delaying their entry into the trial until the desired allocation appears. In epidemiological terms, such manipulation of the assigned intervention may introduce confounding. For this reason, successful allocation sequence concealment is an essential part of randomization.

Allocation concealment should not be confused with blinding of assigned interventions during the trial. Allocation concealment seeks to prevent bias in intervention assignment by preventing trial personnel and participants from knowing the allocation sequence before and until assignment. It can always be successfully implemented, regardless of the study design or clinical area (18, 19). In contrast, blinding (of participants, trial personnel or outcome assessors) seeks to prevent bias subsequent to randomization by continuing the concealment of the assigned intervention after randomization (19, 20), and cannot always be implemented. This is often the situation, for example, in trials comparing surgical with non-surgical interventions. Allocation concealment up to the point of assignment of the intervention and blinding after that point address different sources of bias and differ in their feasibility, Nonetheless, failure to conceal allocation from participants and personnel at the point of assignment implies that these individuals are not blinded to the assignments afterwards.

##### *4.1.1 Approaches to sequence generation*

Randomization with no constraints is called **simple randomization** or **unrestricted randomization**. Sometimes **blocked randomization (restricted randomization)** is used to generate a sequence to ensure that the desired ratio of participants in the experimental and comparator intervention groups (e.g. 1:1) is achieved (21, 22). This is done by ensuring that the numbers of participants assigned to each intervention group is balanced within blocks of specified size (for example, for every 10 consecutively entered participants): the specified number of allocations to experimental and comparator intervention groups is assigned in random order within each block. If the block size is known to trial personnel, then the last allocation within each block can always be predicted. To avoid this problem multiple block sizes may be used, and randomly varied (random permuted blocks).

**Stratified randomization**, in which restricted randomization is performed separately within subsets of participants defined by potentially important prognostic factors, such as disease severity and study centres, is also common. If simple (rather than restricted) randomization is used in each stratum, then stratification offers no benefit, but the randomization is still valid.

**Minimization,** which incorporates both stratification and restricted randomization, can be used to make intervention groups closely similar with respect to specified prognostic factors. Minimization generally includes a random element (at least for participants enrolled when the groups are balanced with respect to the prognostic factors included in the algorithm).

Other adequate types of randomization that are sometimes used are biased coin or urn randomization, replacement randomization, mixed randomization, and maximal randomization (21, 23, 24). If these or other approaches are encountered, consultation with a methodologist may be necessary.

###### 4.1.2 *Allocation concealment and failures of randomization*

Even when the allocation sequence is generated appropriately, knowledge of the next assignment can enable selective enrolment of participants on the basis of prognostic factors. Participants who would have been assigned to an intervention deemed to be inappropriate may be rejected, or participants may be directed to the 'appropriate' intervention, for example by delaying their entry into the trial until the desired allocation appears. For this reason, successful allocation sequence concealment is an essential part of randomization.

Ways in which future assignments can be anticipated, leading to a failure of allocation concealment, include:

(1) knowledge of a deterministic assignment rule, such as by alternation, date of birth or day of admission;
(2) knowledge of the sequence of assignments, whether randomized or not (e.g. if a sequence of random assignments is openly posted on a bulletin board);
(3) ability to predict assignments successfully, based on previous assignments.

The last of these can occur when blocked randomization is used, and when assignments are known to the recruiter after each participant is enrolled into the trial. It may then be possible to predict future assignments, particularly when blocks are of a fixed size and are not divided across multiple recruitment centres (25).

The risk that assignments could be predicted when using minimization leads some methodologists to be cautious about the acceptability of this approach, while others consider it to be attractive, particularly for small trials in which substantial imbalances in baseline characteristics can occur by chance if simple randomization is used (26, 27). To mitigate this risk, minimization approaches are often combined with simple randomization, so that (for example) 80% of allocations are by minimization but the remaining 20% by simple randomization. Allocation concealment when using a minimization-based strategy is further protected in multicentre trials where minimization is done across centres.

Attempts to achieve allocation concealment may be undermined in practice. For example, unsealed allocation envelopes may be opened, while translucent envelopes may be held against a bright light to reveal the contents (3, 28, 29). Personal accounts suggest that many allocation schemes have been deciphered by investigators because the methods of concealment were inadequate (28).

Information about methods for sequence generation and allocation concealment can usually be found in trial protocols, but unfortunately is often not fully reported in publications (27). For example, a Cochrane review on the completeness of reporting of randomized trials found allocation concealment reported adequately in only 45% (393/876) of randomized trials in CONSORT-endorsing journals and in 22% (329/1520) of randomized trials in non-endorsing journals (30). This can sometimes be due to limited word counts in journals, highlighting the importance of looking at multiple information sources. Lack of description of methods of randomization and allocation concealment in a journal article does not necessarily mean that the methods used were inappropriate: details may have been omitted because of limited word counts, oversight or editorial recommendations. (31).

The success of randomization in producing comparable groups is often examined by comparing baseline values of important prognostic factors between intervention groups. In contrast to the under-reporting of randomization methods, baseline characteristics are reported in 95% of RCTs published in CONSORT-endorsing journals and in 87% of RCTs in non-endorsing journals (30). Corbett et al have argued that risk-of-bias assessments should consider whether participant characteristics are balanced between intervention groups (32). RoB 2 includes a signalling question requiring a judgement about whether baseline imbalances suggest that there was a problem with the randomization process (see 4.3.3).

#### 4.2 Empirical evidence of bias arising from the randomization process

A recent meta-analysis of seven meta-epidemiological studies found that an inadequate or unclear (versus adequate) method of **sequence generation** was associated with a small (7%) exaggeration of intervention effect estimates (33). Unexpectedly, the bias was greater in trials reporting subjective outcomes: there was little evidence for bias in trials assessing all-cause mortality and other objective outcomes.

Similarly, a modest (10%) exaggeration of intervention effect estimates was observed for trials with inadequate/unclear (versus adequate) **concealment of allocation**. The average bias associated with inadequate allocation concealment was greatest in trials reporting subjective outcomes and in trials of complementary and alternative medicine, with no evidence of bias in trials of mortality or other objective outcomes. Evidence on baseline imbalances is scarcer. Although three empirical studies found no evidence that baseline imbalances inflate intervention effect estimates (34-36), all estimates were imprecise and these studies also found no evidence that randomization methods were associated with inflated intervention effect estimates. There was little evidence that intervention effect estimates were exaggerated in trials without adjustment for confounders (34) or in unblinded trials with block randomization, in which the last allocation in a block might be predictable (35). However, each characteristic was only examined in a single small study.

#### 4.3 Using this domain of the tool

##### *4.3.1 Assessing random sequence generation*

The use of a random component should be sufficient for adequate sequence generation.

In principle, simple randomization can be achieved by allocating interventions using methods such as repeated coin-tossing, throwing dice, dealing previously shuffled cards, or by referring to a published list of random numbers (21, 22). More usually a list of random assignments is generated by a computer. Risk of bias may be judged in the same way whether or not a trial claims to have stratified its randomization.

> *Example of random sequence generation*: "We generated the two comparison groups using simple randomization, with an equal allocation ratio, by referring to a table of random numbers."
>
> *Example of random sequence generation*: "We used blocked randomization to form the allocation list for the two comparison groups. We used a computer random number generator to select random permuted blocks with a block size of eight and an equal allocation ratio."

Systematic methods, such as alternation, assignment based on date of birth, case record number and date of presentation, which are sometimes referred to as "quasi-random", are inadequate methods of sequence generation. Alternation (or rotation, for more than two intervention groups) might in principle result in similar groups, but many other systematic methods of sequence generation may not. For example, the day on which a patient is admitted to hospital is not solely a matter of chance. An important weakness with all systematic methods is that concealing the allocation schedule is usually impossible, which allows foreknowledge of intervention assignment among those recruiting participants to the study, and biased allocations.

> *Example of non-random sequence generation*: "Patients were randomized by the first letter of the last name of their primary resident (37)."
>
> *Example of non-random sequence generation*: "Those born on even dates were resuscitated with room air (room air group), and those born on odd dates were resuscitated with 100% oxygen (oxygen group) (38)."

###### *4.3.1.1 Assessing sequence generation when insufficient information is provided about the methods used*

A simple statement such as "we randomly allocated" or "using a randomized design" is often insufficient to be confident that the allocation sequence was genuinely randomized. Indeed, it is common for authors to use the term "randomized" even when it is not justified: many trials with declared systematic allocation have been described by the authors as "randomized". In some situations, a reasonable judgement may be made about whether a random sequence was used. For example, , in the context of a large trial run by an experienced clinical trials unit, absence of specific information about generation of the randomization sequence, in a paper published in a journal with rigorously enforced word count limits, is likely to result in a response of 'Probably yes' rather than 'No information'. Alternatively, if other (contemporary) trials by the same investigator team have clearly used non-random sequences, it might be reasonable to assume that the current study was done using similar methods, and answer 'Probably no' to the signalling question. If users of the tool are not able (or insufficiently confident) to make such judgements, an answer of 'No information' should be provided.

Trial investigators may describe their approach to sequence generation incompletely, without confirming that there was a random component. For example, authors may state that blocked allocation was used without an explicit statement that the order of allocation within the blocks was random. In such instances, an answer of 'No information' should generally be provided.

##### *4.3.2 Assessing concealment of allocation sequence*

Among the methods used to conceal allocation, central randomization by a third party is the most desirable. Methods using envelopes are more susceptible to manipulation than other approaches (18, 27). If investigators use envelopes, they should develop and monitor the allocation process to preserve concealment. In addition to use of sequentially numbered, opaque, sealed envelopes, they should ensure that the envelopes are opened sequentially, and only after the envelope has been irreversibly assigned to the participant. When blocking is used, it be may be possible to predict the last intervention assignments within each block. This will be a problem when the person recruiting participants knows the start and end of each block and the allocations are revealed after assignment. The problem is likely to be more serious if block sizes are small and of equal sizes. In such situations, an answer of 'No' or 'Probably no' should be provided for the signalling question concerning whether allocations were concealed.

Table 2 provides minimal criteria for a judgement of adequate concealment of allocation sequence and extended criteria, which provide additional assurance that concealment of the allocation sequence was indeed adequate. Some examples of adequate approaches are provided in Box 3.

**Table 2. Minimal and extended criteria for judging of allocation sequence to be concealed**

| Minimal criteria for a judgement of adequate concealment of the allocation sequence | Extended criteria providing additional assurance |
|---|---|
| Central randomization. | The central randomization office was remote from patient recruitment centres. Participant details were provided, for example, by phone (including interactive voice response systems), email or an interactive online system, and the allocation sequence was concealed to individuals staffing the randomization office until a participant was irreversibly registered. |
| Sequentially numbered drug containers. | Drug containers prepared by an independent pharmacy were sequentially numbered and opened sequentially. Containers were of identical appearance, tamper-proof and equal in weight. |
| Sequentially numbered, opaque, sealed envelopes. | Envelopes were sequentially numbered and opened sequentially only after participant details were written on the envelope. Pressure-sensitive or carbon paper inside the envelope transferred the participant's details to the assignment card. Cardboard or aluminium foil inside the envelope rendered the envelope impermeable to intense light. Envelopes were sealed using tamper-proof security tape. |

**Box 3. Examples of adequate allocation sequence concealment (as compiled by Schulz and Grimes (39))**

"... that combined coded numbers with drug allocation. Each block of ten numbers was transmitted from the central office to a person who acted as the randomization authority in each centre. This individual (a pharmacist or a nurse not involved in care of the trial patients and independent of the site investigator) was responsible for allocation, preparation, and accounting of trial infusion. The trial infusion was prepared at a separate site, then taken to the bedside nurse every 24 h. The nurse infused it into the patient at the appropriate rate. The randomization schedule was thus concealed from all care providers, ward physicians, and other research personnel." (40).

"... concealed in sequentially numbered, sealed, opaque envelopes, and kept by the hospital pharmacist of the two centres." (41).

"Treatments were centrally assigned on telephone verification of the correctness of inclusion criteria..." (42).

"Glenfield Hospital Pharmacy Department did the randomization, distributed the study agents, and held the trial codes, which were disclosed after the study." (43).

##### *4.3.3 Using baseline imbalance to identify problems with the randomization process*

The RoB 2 tool includes consideration of situations in which baseline characteristics indicate that something may have gone wrong with the randomization process. However, only differences that are clearly beyond what is expected due to chance should be interpreted as suggesting problems with the randomization process: see section 4.3.3.1.

Severe baseline imbalances may arise as a result of deliberate attempts to subvert the randomization process (44). They may also occur because of unintentional actions or errors that occurred due to insufficient safeguards: for example an error in a minimization programme such as writing a 'plus' instead of a 'minus', leading to maximizing instead of minimizing differences in one or more prognostic factors between groups.

Assessment of baseline imbalance should be based on data for all randomized participants. If baseline data are presented only for participants who completed the trial (or some other subset of randomized participants) then it is more difficult to assess baseline imbalance, and the proportion of and reasons for missing data need to be considered. The practice of reporting baseline characteristics of analysed participants only is not common in healthcare trials but may be common in other areas such as social care.

###### *4.3.3.1 Chance imbalances at baseline*

In trials using large samples , simple randomization generates intervention groups of relatively similar sizes (21-23). In trials using small samples, simple randomization will sometimes lead to groups that differ substantially, by chance, in size or in the distribution of prognostic factors (45). For example, with 250 participants per group and five important prognostic factors each with 20% prevalence, there is a 23% chance that at least one of them will have >7% difference between groups (46).

Chance imbalances are not a source of systematic bias, and the **RoB 2 tool does not aim to identify imbalances in baseline variables that have arisen due to chance**. A small number of differences identified as 'statistically significant' at the conventional 0.05 threshold should usually be considered to be compatible with chance.

The 95% confidence interval for the effect of intervention incorporates the uncertainty arising from the potential for imbalances in prognostic factors between intervention groups (47). Nonetheless, when chance baseline imbalances in prognostic factors occur it is appropriate to adjust for them (48); preferably in a pre-planned way (e.g. based on a rule specified in a trial analysis plan that is published before unblinded data are available to the investigators) (47).

The average effect of chance imbalances across the trials included in a meta-analysis will be zero, and the confidence interval for the meta-analysis result incorporates their effect. The possible impact on a synthesis of studies with important chance imbalances across rather than within the studies needs to be considered outside of the study-specific risk of bias assessment.

###### 4.3.3.2 *Indications from baseline imbalance that there were problems with the randomization process*

(1) Substantial differences between intervention group sizes, compared with the intended allocation ratio

One example is a 1948 trial comparing anticoagulation medication to conventional treatment for myocardial infarction (49). Anticoagulants were administered to patients admitted on odd admission dates (n=589) and conventional therapy to patients admitted on even admission dates (n=442). Such a large difference in numbers is unlikely given the expected 1:1 allocation ratio (P = 0.001), raising suspicion that investigators manipulated the allocation so that more patients were recruited to the trial on odd dates, when they would receive the new anticoagulant (49).

(2) A substantial excess in statistically significant differences in baseline characteristics between intervention groups, beyond that expected by chance

It is widely understood that statistical tests for differences in baseline characteristics should not be used in truly randomized trials, because the null hypothesis (that there are no systematic differences between the intervention groups) is known to be true. However, such tests can in principle be used to examine whether randomization was implemented successfully. *It is important that such evidence is interpreted appropriately*. Under randomization, one in 20 tests for baseline imbalance are expected to be statistically significant at a 5% level. If a substantially greater proportion of tests for baseline imbalance provide evidence of differences between intervention groups, or if P values are extremely small, this may suggest problems with the randomization process. However, it is possible that trial investigators select the tests for baseline imbalance that are reported, either because they are statistically significant or because they are not statistically significant. Further, different prognostic factors may be correlated (for example a chance imbalance in age may lead to imbalance in other prognostic factors that are influenced by age). Therefore, review authors should be cautious in concluding that there is an excess of statistically significant differences between baseline characteristics.

(3) Imbalance in key prognostic factors, or baseline measures of outcome variables, that are unlikely to be due to chance

These are the factors that might influence those recruiting participants into the study, and therefore have most potential to be manipulated by investigators who (consciously or unconsciously) want to influence the trial results. The review team should, where possible, identify in advance the key prognostic factors that may influence the outcome of interest, for example through the knowledge of subject matter experts who are members of the review group, through initial (scoping) literature reviews, or through discussions with health professionals who make intervention decisions for the target patient or population groups. Based on this knowledge, imbalances in one or more key prognostic factors should be considered to place the study at high risk of bias if the P value for the between-intervention group difference is small enough that they are unlikely to be due to chance (for example, <0.001) and the difference is big enough for the resulting confounding to bias the intervention effect estimate.

Plotting difference in baseline characteristics between intervention arms on a forest plot can be helpful way of visualizing baseline differences between intervention groups across studies. A methodological case study demonstrated that an apparent treatment effect was in fact due to baseline imbalances between intervention groups (50).

(4) Excessive similarity in baseline characteristics that is not compatible with chance

Excessive similarity across intervention groups may provide evidence of flawed or absent methods of randomization, if it is not compatible with the chance differences that arise through randomization. In an examination of baseline data from 5087 randomized trials, Carlisle observed more instances of baseline similarity than would be expected by chance, which could be explained by data fabrication among other reasons (51). Carlisle also observed that the proportion of trials with excessive similarity was higher among trials that had subsequently been retracted. Note that restricted randomization methods (see section 4.3.1) tend to give rise to groups that are more similar at baseline than simple randomization methods.

##### 4.3.4 *Analyses that adjust for baseline imbalances*

If trialists observe baseline imbalances between intervention groups, they may undertake analyses that attempt to control for these by adjusting for baseline values of prognostic variables or the outcome variable. However, if the imbalances were caused by problems in the randomization process, rather than being due to chance, then to remove the risk of bias it would be necessary to adjust for all prognostic factors that influenced intervention group assignment. Because this is unlikely to be possible, such analyses will at best reduce the risk of bias. If review authors wish to assess the risk of bias in a trial that controlled for baseline imbalances in order to mitigate failures of randomization, the study should be treated as non-randomized and assessed using the ROBINS-I tool (Risk of Bias in Non-randomized Studies of Interventions) (52).

#### 4.4 Signalling questions and criteria for judging risk of bias

Signalling questions for this domain are provided in Box 4. Note that the answer to one signalling question should not affect answers to other questions. For example, if the trial has large baseline imbalances, but authors report adequate randomization methods, then sequence generation and allocation concealment should still be assessed on the basis of the reported adequate methods. Concerns about the imbalances should be reflected in the answer to the question about the baseline imbalance and reflected in the domain-level judgement.

Criteria for reaching risk-of-bias judgements are given in Table 3, and an algorithm for implementing these is provided in Table 4 and Figure 1. A judgement of low risk of bias requires that the trial has an adequate method of concealing the allocation sequence from those involved in enrolling participants, and there are no concerns about generation of the allocation sequence. Suggested risk of bias judgements can be overridden if review authors believe this is justified: for example the importance of allocation concealment may depend on the extent to which potential participants in the study have different prognoses, whether strong beliefs exist among investigators and participants regarding the benefits or harms of assigned interventions, and whether uncertainty about the interventions is accepted by all people involved (44).

**Box 4. The RoB 2 tool (part 2): Risk of bias arising from the randomization process**

| Signalling questions | Elaboration | Response options |
|---|---|---|
| **1.1 Was the allocation sequence random?** | Answer 'Yes' if a random component was used in the sequence generation process. Examples include computer-generated random numbers; reference to a random number table; coin tossing; shuffling cards or envelopes; throwing dice; or drawing lots. Minimization is generally implemented with a random element (at least when the scores are equal), so an allocation sequence that is generated using minimization should generally be considered to be random.<br><br>Answer 'No' if no random element was used in generating the allocation sequence or the sequence is predictable. Examples include alternation; methods based on dates (of birth or admission); patient record numbers; allocation decisions made by clinicians or participants; allocation based on the availability of the intervention; or any other systematic or haphazard method.<br><br>Answer 'No information' if the only information about randomization methods is a statement that the study is randomized.<br><br>In some situations a judgement may be made to answer 'Probably no' or 'Probably yes'. For example, , in the context of a large trial run by an experienced clinical trials unit, absence of specific information about generation of the randomization sequence, in a paper published in a journal with rigorously enforced word count limits, is likely to result in a response of 'Probably yes' rather than 'No information'. Alternatively, if other (contemporary) trials by the same investigator team have clearly used non-random sequences, it might be reasonable to assume that the current study was done using similar methods. | Y/PY/PN/N/NI |
| **1.2 Was the allocation sequence concealed until participants were enrolled and assigned to interventions?** | Answer 'Yes' if the trial used any form of remote or centrally administered method to allocate interventions to participants, where the process of allocation is controlled by an external unit or organization, independent of the enrolment personnel (e.g. independent central pharmacy, telephone or internet-based randomization service providers).<br><br>Answer 'Yes' if envelopes or drug containers were used appropriately. Envelopes should be opaque, sequentially numbered, sealed with a tamper-proof seal and opened only after the envelope has been irreversibly assigned to the participant. Drug containers should be sequentially numbered and of identical appearance, and dispensed or administered only after they have been irreversibly assigned to the participant. This level of detail is rarely provided in reports, and a judgement may be required to justify an answer of 'Probably yes' or 'Probably no'.<br><br>Answer 'No' if there is reason to suspect that the enrolling investigator or the participant had knowledge of the forthcoming allocation. | Y/PY/PN/N/NI |
| **1.3 Did baseline differences between intervention groups suggest a problem with the randomization process?** | *Note that differences that are compatible with chance do not lead to a risk of bias. A small number of differences identified as 'statistically significant' at the conventional 0.05 threshold should usually be considered to be compatible with chance.*<br><br>Answer 'No' if no imbalances are apparent or if any observed imbalances are compatible with chance.<br><br>Answer 'Yes' if there are imbalances that indicate problems with the randomization process, including:<br>(1) substantial differences between intervention group sizes, compared with the intended allocation ratio; or<br>(2) a substantial excess in statistically significant differences in baseline characteristics between intervention groups, beyond that expected by chance; or | Y/PY/PN/N/NI |

| | | |
|---|---|---|
| | (3) imbalance in one or more key prognostic factors, or baseline measures of outcome variables, that is very unlikely to be due to chance and for which the between-group difference is big enough to result in bias in the intervention effect estimate.<br>Also answer 'Yes' if there are other reasons to suspect that the randomization process was problematic:<br>(4) excessive similarity in baseline characteristics that is not compatible with chance.<br>Answer 'No information' when there is no *useful* baseline information available (e.g. abstracts, or studies that reported only baseline characteristics of participants in the final analysis).<br>The answer to this question should not influence answers to questions 1.1 or 1.2. For example, if the trial has large baseline imbalances, but authors report adequate randomization methods, questions 1.1 and 1.2 should still be answered on the basis of the reported adequate methods, and any concerns about the imbalance should be raised in the answer to the question 1.3 and reflected in the domain-level risk-of-bias judgement.<br>Trialists may undertake analyses that attempt to deal with flawed randomization by controlling for imbalances in prognostic factors at baseline. To remove the risk of bias caused by problems in the randomization process, it would be necessary to know, and measure, all the prognostic factors that were imbalanced at baseline. It is unlikely that all important prognostic factors are known and measured, so such analyses will at best reduce the risk of bias. If review authors wish to assess the risk of bias in a trial that controlled for baseline imbalances in order to mitigate failures of randomization, the study should be assessed using the ROBINS-I tool. | |
| **Risk-of-bias judgement** | See Table 3, Table 4 and Figure 1. | Low / High / Some concerns |
| Optional: What is the predicted direction of bias arising from the randomization process? | If the likely direction of bias can be predicted, it is helpful to state this. The direction might be characterized either as being towards (or away from) the null, or as being in favour of one of the interventions. | NA / Favours experimental / Favours comparator / Towards null /Away from null / Unpredictable |

**Table 3. Reaching risk-of-bias judgements for bias arising from the randomization process**

| | |
|---|---|
| Low risk of bias | (i) The allocation sequence was adequately concealed<br>AND<br>(ii.1) Any baseline differences observed between intervention groups appear to be compatible with chance<br>OR<br>(ii.2) There is no information about baseline imbalances<br>AND<br>(iii.1) The allocation sequence was random<br>OR<br>(iii.2) There is no information about whether the allocation sequence was random |
| Some concerns | (i.1) The allocation sequence was adequately concealed<br>AND<br>(i.2.1) The allocation sequence was not random<br>OR<br>(i.2.2) Baseline differences between intervention groups suggest a problem with the randomization process<br>OR<br>(ii.1) There is no information about concealment of the allocation sequence<br>AND<br>(ii.2) Any baseline differences observed between intervention groups appear to be compatible with chance<br>OR<br>(iii) There is no information to answer any of the signalling questions |
| High risk of bias | (i) The allocation sequence was not adequately concealed<br>OR<br>(ii.1) There is no information about concealment of the allocation sequence<br>AND<br>(ii.2) Baseline differences between intervention groups suggest a problem with the randomization process |

**Table 4 Mapping of signalling questions to suggested risk-of-bias judgements for bias arising from the randomization process.** This is only a suggested decision tree: all default judgements can be overridden by assessors.

| Signalling question | | | Domain-level judgement | |
|---|---|---|---|---|
| **1.1 Sequence random?** | **1.2 Allocation concealed?** | **1.3 Imbalance suggest problem?** | **Default risk of bias** | **Remarks** |
| Y/PY/NI | Y/PY | NI/N/PN | Low | |
| Y/PY | Y/PY | Y/PY | Some concerns | There is considerable room for judgement here. Substantial baseline imbalance despite apparently sound randomization methods should be investigated carefully, and a judgement of 'Low' risk of bias or 'High' risk of bias might be reached. |
| N/PN/NI | Y/PY | Y/PY | Some concerns | Substantial baseline imbalance may lead to a judgement of 'High' risk of bias, especially if the method of sequence generation is also inappropriate. |
| Any response | NI | N/PN/NI | Some concerns | |
| Any response | NI | Y/PY | High | |
| Any response | N/PN | Any response | High | |

Y/PY = 'Yes' or 'Probably yes'; N/PN = 'No' or 'Probably no'; NI = 'No information'

**Figure 1. Algorithm for suggested judgement of risk of bias arising from the randomization process.**

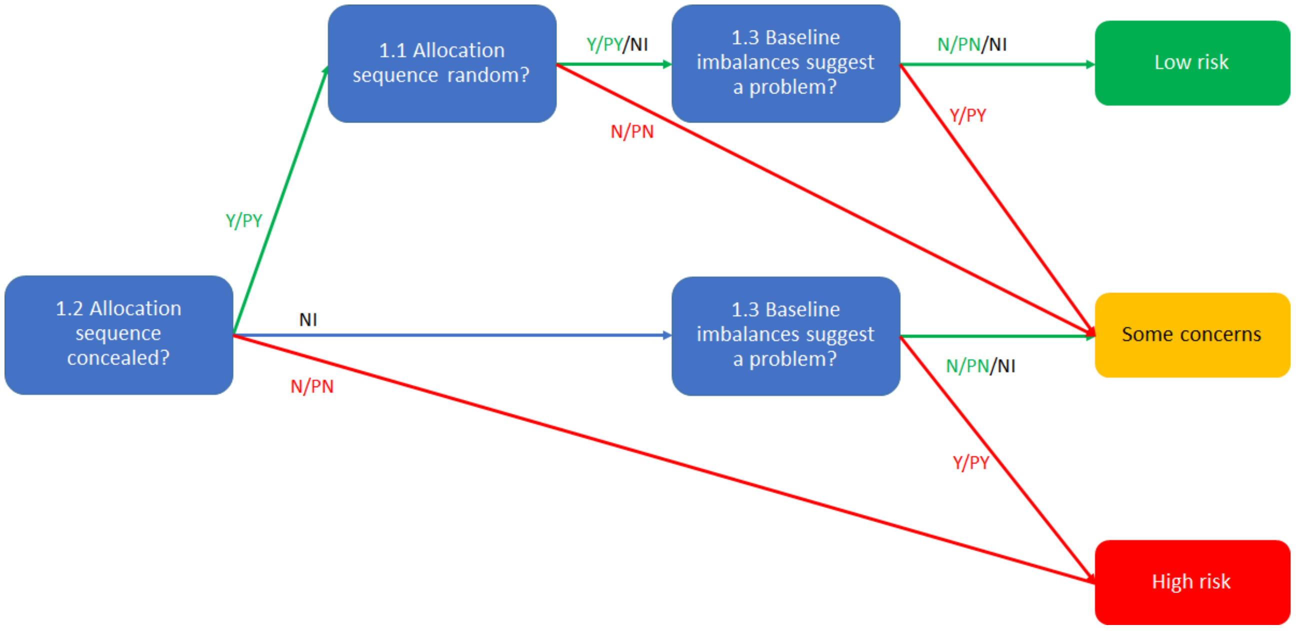

### 5 Detailed guidance: bias due to deviations from intended interventions

#### 5.1 Background

This domain relates to biases that arise when there are deviations from the intended interventions. Such deviations could be the administration of additional interventions that are inconsistent with the trial protocol, failure to implement the protocol interventions as intended, or non-adherence by trial participants to their assigned intervention. Biases that arise due to deviations from intended interventions were referred to as performance biases in the original Cochrane tool for assessing risk of bias in randomized trials.

The interventions that were intended should be fully specified in the trial protocol, although this is often not done, particularly when it is intended that interventions should change or evolve in response to the health of, or events experienced by, trial participants. For example, the investigators may intend that:

- in a trial of a new drug to control symptoms of rheumatoid arthritis, participants experiencing severe toxicities should receive additional care and/or switch to an alternative drug;
- in a trial of a specified cancer drug regimen, participants whose cancer progresses should switch to a second-line intervention; or
- in a trial comparing surgical intervention with conservative management of stable angina, participants who progress to unstable angina receive surgical intervention.

**Such changes to intervention are consistent with the trial protocol, do not cause bias, and should not be considered to be deviations from intended intervention.**

Unfortunately, trial protocols may not fully specify or articulate the circumstances in which deviations from the initial intervention should occur, or distinguish changes to intervention that are consistent with the intentions of the investigators from those that are inconsistent with the protocol and so should be considered as deviations from the intended intervention. For example, a cancer trial protocol may not define progression, or specify the second-line drug that should be used in patients who progress (53). It may therefore be necessary for users of RoB 2 to document changes to intervention that they do and do not consider to be consistent with the trial protocol. Similarly, for trials in which the comparator intervention is "usual care", the protocol may not specify the interventions consistent with usual care or whether they are expected to be used alongside the experimental intervention. Users of the RoB 2 tool may therefore need to describe interventions that are consistent with usual care.

##### *5.1.1 Non-protocol interventions*

Non-protocol interventions that trial participants might receive during trial follow up and that are likely to affect the outcome of interest can lead to bias in estimated intervention effects. If possible, review authors should specify potential non-protocol interventions in advance (at review protocol writing stage). They may be identified through the expert knowledge of members of the review group, via initial (scoping) reviews of the literature, and through discussions with health professionals.

##### *5.1.2 The role of the effect of interest*

As described in section 1.3, assessments for this domain depend on whether the intervention effect of interest to the review authors is

(1) the effect of **assignment** to the interventions at baseline (regardless of whether the interventions are received or adhered to during follow-up, sometimes known as the 'intention-to-treat effect'); or

(2) the effect of **adhering to** intervention as specified in the trial protocol (sometimes known as the 'per-protocol effect').

These effects will differ if some participants do not receive their assigned intervention or deviate from assigned intervention after baseline.

The net effect of assignment to intervention in a particular trial depends on three components: the actual effect of the intervention, the degree and type of adherence to the intervention, and trial-specific recruitment and engagement activities that affect participants' outcomes (54). As an example of the third component, the information provided during the process of securing informed consent may increase participants' awareness of potential ways that behaviour change might improve their prognosis, or increase participants' engagement with health care and propensity to adhere to interventions. By contrast, the net effect of adhering to intervention depends only on the actual effect of the intervention and the trial-specific recruitment and engagement activities that affect participants' outcomes.

The extent of deviations from intended intervention may differ between trials comparing similar interventions, leading to heterogeneity in meta-analyses of intention-to-treat effects (5). For example, rates of non-adherence to assigned intervention may differ according to the care with which the process of randomisation was explained during recruitment of participants.

Participants recruited to trials may not be representative of all individuals who might benefit from the interventions being compared (55). Trial protocols may restrict eligibility, for example by specifying a maximum age or excluding patients with comorbidities. Participants who consent to recruitment may have less strong preferences for particular interventions, or differ in their motivation to adhere to interventions, than participants who do not consent. Further, the quality of health care may differ between trial- and non-trial settings. These issues limit the generalisability of trial results to non-trial settings (its external validity), and have led to initiatives that aim to make trial results more representative, for example by avoiding restrictive eligibility criteria or conducting them in situations typical of routine care (56). However, they do not affect our ability to estimate the intervention effect without bias in trial participants cared for in the healthcare settings of the trial. Therefore, these aspects of generalizability or transportability are not addressed in the RoB 2 tool.

###### *5.1.2.1 Bias in the effect of assignment to intervention*

In some trials, particularly those in which participants or trial personnel were not blinded, non-protocol interventions may arise because of trial recruitment and engagement activities. It is also possible that trial personnel (carers and people delivering the interventions) undermine the trial comparisons by implementing non-protocol interventions or failing to implement the protocol interventions. This could happen because of unconscious processes (e.g. lack of equipoise leading to administration of non-protocol interventions in one group and not the other), or conscious processes (e.g. arising from a conflict of interest).

We use the term **trial context** to refer to effects of recruitment and engagement activities on trial participants and effects of any unconscious or conscious processes on the delivery of interventions by trial personnel. In RoB 2 the only deviations from the intended intervention that are addressed in relation to the effect of *assignment to intervention* are those that:

**(1) arose because of the trial context;**
**(2) were inconsistent with the trial protocol; and**
**(3) influence the outcome.**

For example, in an unblinded study the process of securing informed consent may lead participants subsequently assigned to the comparator group to feel unlucky and therefore seek the experimental intervention, or other interventions that improve their prognosis. Effect estimates from trials with strong recruitment or engagement effects are hard to transport to settings in which those effects do not exist, and for this reason they are assessed in RoB 2.

Similarly, in an open-label study comparing minimally invasive versus open surgery for oesophageal cancer the protocol specified that one-lung ventilation should be used in both groups. However, one-lung mechanical ventilation is thought to increase respiratory complications, including RTIs and surgeons usually used two-lung ventilation in the minimally invasive group. Such non-protocol interventions arose because of surgeons' beliefs about the safety of the interventions (lack of equipoise) then the estimated effect of assignment to intervention would be at risk of bias.

Deviations from intervention that do not arise because of the trial context, such as a patient's choice to stop taking their assigned medication, do not lead to bias in the effect of assignment to intervention.

###### *5.1.2.2 Bias in the effect of adhering to intervention*

All deviations from the intended intervention that are inconsistent with the trial protocol and affect the outcome are addressed in relation to the effect of adhering to intervention, regardless of whether they arose because of the trial context.

It is sometimes not possible to adjust for all deviations from intended intervention. Therefore, when assessing the effect of ***adhering to intervention*** as specified in the trial protocol, review authors should specify, in the preliminary considerations (see section 3), what types of deviations from the intended intervention (departures from the trial protocol) will be examined. These will be one or more of: (1) occurrence of non-protocol interventions that could affect the outcome; (2) failures in implementing the protocol interventions that could affect the outcome; and (3) non-adherence to their assigned intervention by trial participants. For example, the START randomized trial compared immediate with deferred initiation of antiretroviral therapy (ART) in HIV-positive individuals, but 30% of those assigned to deferred initiation started ART earlier than the protocol specified (6). Lodi and colleagues estimated a per-protocol effect that adjusted for these protocol deviations, but not for whether participants continued antiretroviral therapy throughout trial follow-up (7). If such deviations are present, review authors should consider whether appropriate statistical methods were used to adjust for their effects.

Some examples of studies in which there were deviations from the intended interventions are provided in Box 5.

##### *5.1.3 The role of blinding*

The implementation of mechanisms to ensure that participants, carers and trial personnel (i.e. people delivering the interventions) are unaware of the interventions received is referred to as blinding. In some areas (including eye health) the term 'masking' is preferred. If successful, blinding should prevent knowledge of the intervention assignment from influencing contamination (application of one of the interventions in participants intended to receive the other), switches to non-protocol interventions or non-adherence by trial participants. Lack of blinding may cause bias if it leads to deviations from intended intervention that are related to the trial context or, for estimation of per-protocol effects, to departures from the trial protocol that differ between the intervention groups.

Blinding is not appropriate in pragmatic trials whose goal is to compare intervention strategies in individuals who are aware of their care. However, it is essential in trials that aim to eliminate placebo effects and isolate the specific effect of the protocol interventions. For example, trials of acupuncture to treat pain tend to find benefit when it is compared with no treatment, but no important benefit when the comparison is with sham acupuncture and the participants and carers (other than those delivering the intervention) are blinded (57). Similarly, a blinded comparison of a drug with placebo allows estimation of the pharmacological effect of the compound concerned, whereas the effect estimated from a comparison of the drug with no intervention combines the effect of the compound with that of the whole treatment process.

Regardless of whether it is appropriate, blinding trial participants is difficult or impossible in some contexts – for example in a trial comparing a surgical with a non-surgical intervention. Non-blinded ('open') studies may take other measures to avoid deviations from intended intervention, such as treating participants according to strict criteria that prevent administration of non-protocol interventions.

The RoB 2 tool assumes that when participants and trial personnel were blinded during the trial, deviations from intended intervention are not influenced by trial-specific recruitment and engagement activities. Therefore, when interest is in the effect of assignment to intervention the signalling question addressing such deviations is asked only when these groups were not blinded.

Blinding of outcome assessors, to avoid bias in *measuring* the outcome, is considered separately, in the 'Bias in measurement of outcomes' domain (section 7). Bias due to differential rates of drop out (withdrawal from the study) is considered in the 'Bias due to missing outcome data' domain (section 6).

An attempt to blind participants, carers and people delivering the interventions to intervention group does not ensure successful blinding in practice. For many blinded drug trials, the side effects of the drugs allow the possible detection of the intervention being received for some participants, unless the study compares similar interventions, e.g. drugs with similar side effects, or uses an active placebo (58-60). Furthermore, if assignment of intervention was not concealed at the time of randomization (see section 4.1.2), then knowledge of the allocation may also be available during the conduct of the trial, so that carers and people delivering the interventions are not fully blinded.

Several groups have suggested that it would be sensible to ask trial participants at the end of the trial to guess which intervention they had been receiving (61, 62), and some reviews of such reports have been published (61, 63). However, evidence of successful guesses can simply reflect that a good outcome or a marked side effect is more often attributed to an active intervention We would then expect to see some successful "guessing" when there is a difference in either efficacy or adverse effects, but none when the interventions have very similar effects, even when the blinding has been preserved.

Deducing the intervention received does not in itself lead to a risk of bias. As discussed above, cessation of a drug intervention because of toxicity will usually not be considered a deviation from intended intervention. The elaborations that accompany the signalling questions for this domain address this issue in more detail.

Study reports often describe blinding in broad terms, such as "double blind". This term makes it difficult to know who was blinded (20). Such terms are also used very inconsistently (65-67), and the frequency of explicit reporting of the blinding status of study participants and trial personnel remains low even in trials published in top journals (68), despite recommendations in the CONSORT Statement to be explicit (69). A review of methods used for blinding highlights the variety of methods used in practice (58).

**Box 5. Examples of studies with deviations from the intended interventions**

**Example 1: substantial numbers of participants not treated as randomized**
To determine the efficacy of surgery for lumbar intervertebral disc herniation, the SPORT trial (Spine Patient Outcomes Research Trial) randomized patients with lumbar disc herniation to receive surgical treatment (discectomy) or non-operative care (encompassing a variety of interventions including analgesics, education, physiotherapy and acupuncture) (70). An ITT analysis found no evidence that the primary outcome (Short Form-36 bodily pain and physical function scales) differed between intervention groups two years after randomization. However, by that time only 60% of patients assigned to surgical treatment had undergone the procedure and 45% of those in the non-operative group had been treated surgically. The authors performed an 'as treated' analysis which, in contrast to ITT, showed advantages for surgery. This leads to a biased estimate of the effect of adhering to intervention (the 'per-protocol effect') because baseline characteristics of participants who underwent surgery (irrespective of their assigned intervention) differed substantially from characteristics of those who did not.

**Example 2: crossover from comparator group to experimental intervention group**
To determine the efficacy of percutaneous coronary interventions, the FAME 2 trial (Fractional Flow Reserve versus Angiography for Multivessel Evaluation 2 trial) randomized patients with stable, functionally-significant, coronary artery disease to a percutaneous coronary intervention (PCI) with implantation of drug-eluting stents and optimal medical therapy (OMT) or to OMT alone (71). The trial was stopped early because of clear evidence from an ITT analysis that participants assigned to PCI were at reduced risk of the primary composite outcome of death from any cause, nonfatal myocardial infarction, or urgent revascularization (hazard ratio [HR] 0.39; 95% CI 0.26 to 0.57; p<0.001). This was driven by a pronounced 77% reduction in urgent revascularization in the PCI group (HR 0.23; 95% CI 0.14 to 0.38; P<0.001). In contrast, there was only a 21% reduction in the composite of death or myocardial infarction (HR 0.79; 95% CI 0.49 to 1.29). However, 41% of patients allocated to OMT had 'crossed over' to PCI: 40% of these PCI procedures were performed because they were clinically indicated (following a myocardial infarction or unstable angina) but the majority were deviations from the trial protocol. Therefore, it is likely that the effect of assignment to PCI versus OMT (the ITT effect) on death or myocardial infarction was closer to the null than the effect of adhering to intervention (the 'per-protocol effect'). To estimate the per-protocol effect, it would be necessary to censor follow up when non-clinically-indicated PCIs occurred in participants allocated to OMT, and adjust the analysis (for example, through inverse probability weighting) to account for the censoring.

**Example 3: non-protocol interventions not balanced between intervention groups and likely to affect the outcome**
An open-label study compared respiratory tract infection (RTI) rates after minimally invasive or open surgery for oesophageal cancer (72). There were two important differences between intervention groups in the delivery of non-protocol interventions. First, one-lung mechanical ventilation (which is thought to increase respiratory complications, including RTIs) was used in the open surgery group, whereas the minimally invasive group underwent two-lung ventilation (note that both types of mechanical ventilation could have been used for either intervention). Second, epidural analgesia was used more frequently in the open surgery group: patients with epidurals are generally less mobile and thus at increased risk of developing an RTI. If some of these non-protocol interventions were administered because of the trial context then the estimated effect of assignment to intervention was at risk of bias, because the co-interventions were not balanced between intervention groups and likely to impact on the outcome.

**Example 4: interventions not implemented successfully**
Patients with acute appendicitis were randomized to undergo standard laparoscopic appendicectomy or single incision laparoscopic appendicectomy (73). Standard laparoscopic surgery involves three separate incisions (through which two laparoscopic instruments and the laparoscopic camera are placed), whereas single incision surgery involves just one incision through which the camera and one instrument are placed. However, fewer than 50% of the patients randomized to undergo single incision surgery received this procedure as intended: additional instruments and incisions were required in 51% and 10% of procedures, respectively. ITT analyses found no strong evidence of between-intervention differences in the risk of wound infection (primary outcome), time to return to normal diet, length of hospital stay, time to return to normal activity, or duration of post-operative analgesia. Such analyses are appropriate to estimate the effect of assignment to intervention (the ITT effect), but this effect may be closer to the null than the effect of adhering to intervention (the per-protocol effect) because of the failure to implement the intervention successfully.

#### 5.2 Empirical evidence of bias due to deviations from intended interventions

Empirical evidence of bias due to deviations from the intended interventions largely comes from studies exploring whether reporting of "double blinding" is associated with intervention effects. Such studies were restricted to meta-analyses in which at least one trial was and was not blinded, and therefore do not apply to situations in which blinding was not feasible or not appropriate. In the largest meta-epidemiological study conducted to date, lack of or unclear double blinding (versus double blinding) in trials with subjectively assessed outcomes was associated with a 23% exaggeration of odds ratio (74). Lack of or unclear double blinding was also associated with increased between-trial heterogeneity. By comparison, there was little evidence of such bias in trials of mortality or other objectively assessed outcomes, in a meta-analysis of meta-epidemiological studies (33). Two other studies examining subjectively measured continuous outcomes (e.g. patient-rated questionnaires) found that standardized mean differences tended to be exaggerated in trials with lack of or unclear blinding of participants (versus blinding of participants) (75, 76). Because existing empirical evidence does not distinguish blinding of participants and trial personnel from blinding of outcome assessors, it not clear which of these aspects of blinding is most important in preventing bias.

Naïve 'per-protocol' analyses have been found to exaggerate estimates of the effect of assignment to intervention compared with 'intention-to-treat' analyses (77, 78). Reporting the use of a 'modified ITT' analysis (versus ITT) has been associated with exaggerated effects (79). Tierney et al. observed a tendency for analyses conducted after trial investigators excluded participants to favour the experimental intervention, compared with analyses including all participants (80).

Interpretation of empirical studies is difficult because exclusions are often poorly reported, particularly in the pre-CONSORT era before 1996. For example, Schulz observed that the *apparent* lack of exclusions was associated with more "beneficial" effect sizes as well as with less likelihood of adequate allocation concealment (18). Hence, failure to report exclusions in trials in Schulz's study may have been a marker of poor trial conduct rather than true absence of any exclusions.

#### 5.3 Using this domain of the tool

##### *5.3.1 The effect of assignment to intervention (the 'intention to treat effect')*

When assessing the effect of assignment to intervention, the signalling questions address:

(1) whether participants, carers and people delivering the interventions were blinded;
(2) if some of these groups were not blinded, whether deviations from intended intervention arose because of the trial context and were likely to have biased the intervention effect; and
(3) whether an appropriate analysis was used to estimate the effect of assignment to intervention.

An appropriate analysis should follow the principles of ITT that participants are analysed in the intervention groups to which they were randomized (regardless of the intervention they actually received) and that all randomized participants in the analysis (regardless of whether the interventions were implemented as intended and regardless of adherence of the participants). Some authors may report a 'modified intention-to-treat' (mITT) analysis in which participants with missing outcome data are excluded. Such an analysis should be considered appropriate for the purposes of this domain: bias because of the missing outcome data is addressed separately, in the domain "Bias due to missing outcome data" (see section 6). Note that the phrase 'modified intention-to-treat' is used in different ways, and may refer to inclusion of participants who received at least one dose of treatment (81); our use of the term refers to missing data rather than to adherence to intervention.

Inappropriate analyses include naïve 'per-protocol' analyses, 'as-treated' analyses, and other analyses based on excluding eligible trial participants post-randomization (14) (see also section 1.3.1). Other inappropriate reasons for excluding eligible trial participants post-randomization include that they experienced toxicities resulting from intervention or even "lack of efficacy". However, post-randomization exclusions of ineligible participants (when eligibility was not confirmed until after randomization and could not have been influenced by intervention group assignment) can be considered appropriate.

Note that it might be possible to conduct individual participant data meta-analyses that include participants who were excluded by the study authors ("re-inclusions") (82). Review authors are encouraged to do this when possible. In this situation, the risk of bias assessment should apply to the result of the trial as it is included in the synthesis, rather than the result as it is reported by the trialists.

It should be straightforward to answer the signalling questions about analysis, providing that the trial is reported in accordance with the CONSORT statement, and includes a CONSORT flow chart (69). Reports of 'as-treated' analyses are uncommon. Reports of naïve 'per-protocol' analyses are more common, although they are often reported in addition to, rather than instead of, an ITT or mITT analysis. Results of ITT analyses should be preferred for inclusion in risk of bias assessments and meta-analyses, when a choice is available (see section 1.3.1). When the result being assessed is based on a naïve 'per-protocol' or 'as treated' analysis, review authors should assess whether there is potential for a substantial impact on the estimated effect of intervention. A 'per protocol' analysis should be selected in preference to an 'as treated' analysis.

Risk of bias in this domain may be differ between outcomes, even if the same people were aware of intervention assignments during the trial. For example, knowledge of the assigned intervention may impact on behaviour (such as number of clinic visits), while not impacting importantly on physiology (including risk of mortality).

##### *5.3.2 The effect of adhering to intervention (the 'per-protocol effect')*

When assessing the effect of adherence to intervention, the signalling questions address:

(1) whether participants, carers and people delivering the interventions were blinded;
(2) if participants, carers or people delivering the interventions were not blinded, whether important co-interventions were balanced across intervention groups;
(3) whether the intervention was implemented successfully, and whether study participants adhered to the assigned intervention;
(4) if deviations from intended intervention arising from points 2 and 3 above occurred, whether an appropriate analysis was used.

Appropriate analysis approaches are described by Hernán and Robins (5): for example instrumental variable approaches can be used in some circumstances to estimate the effect of intervention among participants who received the assigned intervention (see also section 1.3.1).

#### 5.4 Signalling questions and criteria for judging risk of bias

Signalling questions for the effect of assignment to intervention are provided in Box 6. Criteria for reaching risk-of-bias judgements are given in Table 5, and an algorithm for implementing these is provided in Table 6 and Figure 2. Signalling questions for the effect of adhering to intervention are provided in Box 7. Criteria for reaching risk-of-bias judgements are given in Table 7, and an algorithm for implementing these is provided in Table 8 and Figure 3.

**Box 6. The RoB 2 tool (part 3): Risk of bias due to deviations from the intended interventions (*effect of assignment to intervention*)**

| Signalling questions | Elaboration | Response options |
|---|---|---|
| **2.1. Were participants aware of their assigned intervention during the trial?** | If participants are aware of their assigned intervention it is more likely that health-related behaviours will differ between the intervention groups. Blinding participants, most commonly through use of a placebo or sham intervention, may prevent such differences. If participants experienced side effects or toxicities that they knew to be specific to one of the interventions, answer this question 'Yes' or 'Probably yes'. | Y/PY/PN/N/NI |
| **2.2. Were carers and people delivering the interventions aware of participants' assigned intervention during the trial?** | If carers or people delivering the interventions are aware of the assigned intervention then its implementation, or administration of non-protocol interventions, may differ between the intervention groups. Blinding may prevent such differences. If participants experienced side effects or toxicities that carers or people delivering the interventions knew to be specific to one of the interventions, answer question 'Yes' or 'Probably yes'. If randomized allocation was not concealed, then it is likely that carers and people delivering the interventions were aware of participants' assigned intervention during the trial. | Y/PY/PN/N/NI |
| **2.3. If Y/PY/NI to 2.1 or 2.2: Were there deviations from the intended intervention that arose because of the trial context?** | For the effect of assignment to intervention, this domain assesses problems that arise when changes from assigned intervention that are inconsistent with the trial protocol arose because of the trial context. We use the term trial context to refer to effects of recruitment and engagement activities on trial participants and when trial personnel (carers or people delivering the interventions) undermine the implementation of the trial protocol in ways that would not happen outside the trial. For example, the process of securing informed consent may lead participants subsequently assigned to the comparator group to feel unlucky and therefore seek the experimental intervention, or other interventions that improve their prognosis.<br><br>Answer 'Yes' or 'Probably yes' only if there is evidence, or strong reason to believe, that the trial context led to failure to implement the protocol interventions or to implementation of interventions not allowed by the protocol.<br><br>Answer 'No' or 'Probably no' if there were changes from assigned intervention that are inconsistent with the trial protocol, such as non-adherence to intervention, but these are consistent with what could occur outside the trial context.<br><br>Answer 'No' or 'Probably no' for changes to intervention that are consistent with the trial protocol, for example cessation of a drug intervention because of acute toxicity or use of additional interventions whose aim is to treat consequences of one of the intended interventions.<br><br>If blinding is compromised because participants report side effects or toxicities that are specific to one of the interventions, answer 'Yes' or 'Probably yes' only if there were changes from assigned intervention that are inconsistent with the trial protocol and arose because of the trial context.<br><br>The answer 'No information' may be appropriate, because trialists do not always report whether deviations arose because of the trial context. | NA/Y/PY/PN/N/NI |
| **2.4 If Y/PY to 2.3: Were these deviations likely to have affected the outcome?** | Changes from assigned intervention that are inconsistent with the trial protocol and arose because of the trial context will impact on the intervention effect estimate if they affect the outcome, but not otherwise. | NA/Y/PY/PN/N/NI |

| | | |
|---|---|---|
| **2.5. If Y/PY/NI to 2.4: Were these deviations from intended intervention balanced between groups?** | Changes from assigned intervention that are inconsistent with the trial protocol and arose because of the trial context are more likely to impact on intervention effect estimate if they are not balanced between the intervention groups. | NA/Y/PY/PN/N/NI |
| **2.6 Was an appropriate analysis used to estimate the effect of assignment to intervention?** | Both intention-to-treat (ITT) analyses and modified intention-to-treat (mITT) analyses excluding participants with missing outcome data should be considered appropriate. Both naïve 'per-protocol' analyses (excluding trial participants who did not receive their assigned intervention) and 'as treated' analyses (in which trial participants are grouped according to the intervention that they received, rather than according to their assigned intervention) should be considered inappropriate. Analyses excluding eligible trial participants post-randomization should also be considered inappropriate, but post-randomization exclusions of ineligible participants (when eligibility was not confirmed until after randomization, and could not have been influenced by intervention group assignment) can be considered appropriate. | Y/PY/PN/N/NI |
| **2.7 If N/PN/NI to 2.6: Was there potential for a substantial impact (on the result) of the failure to analyse participants in the group to which they were randomized?** | This question addresses whether the number of participants who were analysed in the wrong intervention group, or excluded from the analysis, was sufficient that there could have been a substantial impact on the result. It is not possible to specify a precise rule: there may be potential for substantial impact even if fewer than 5% of participants were analysed in the wrong group or excluded, if the outcome is rare or if exclusions are strongly related to prognostic factors. | NA/Y/PY/PN/N/NI |
| **Risk-of-bias judgement** | See Table 5, Table 6 and Figure 2. | Low / High / Some concerns |
| Optional: What is the predicted direction of bias due to deviations from intended interventions? | If the likely direction of bias can be predicted, it is helpful to state this. The direction might be characterized either as being towards (or away from) the null, or as being in favour of one of the interventions. | NA / Favours experimental / Favours comparator / Towards null /Away from null / Unpredictable |

**Table 5. Reaching risk-of-bias judgements for bias due to deviations from intended intervention (*effect of assignment to intervention*)**

Because the domain addresses two somewhat distinct issues, we separate the algorithm into two parts and combine them to reach the judgement.

| | Part 1: criteria for questions 2.1 to 2.5 | Part 2: criteria for questions 2.6 and 2.7 | Criteria for the domain |
|---|---|---|---|
| Low risk of bias | (i) Participants, carers and people delivering the interventions were unaware of intervention groups during the trial<br>OR<br>(ii.1) Participants, carers or people delivering the interventions were aware of intervention groups during the trial<br>AND<br>(ii.2) No deviations from intended intervention arose because of the trial context. | An appropriate analysis was used to estimate the effect of assignment to intervention | 'Low' risk of bias for Part 1<br>AND<br>'Low' risk of bias for Part 2 |
| Some concerns | (i) Participants, carers or people delivering the interventions were aware of intervention groups during the trial<br>AND<br>(ii.1) There is no information on whether there were deviations from intended intervention because of the trial context<br>OR<br>(ii.1.1) There were deviations from intended interventions that arose because of the trial context<br>AND<br>(ii.1.1.1) These deviations were not likely to have affected the outcome<br>OR<br>(ii.1.1.2) These deviations were balanced between the intervention groups | (i) An appropriate analysis was not used to estimate the effect of assignment to intervention<br>AND<br>(ii) The potential impact (on the estimated effect of intervention) of the failure to analyse participants in the group to which they were randomized was not substantial | "Some concerns' for Part 1<br>OR<br>"Some concerns' for Part 2<br>AND<br>Part 1 not 'High' risk of bias<br>AND<br>Part 2 not 'High' risk of bias |

| | | | |
|---|---|---|---|
| High risk of bias | (i) Participants, carers or people delivering the interventions were aware of intervention groups during the trial<br>AND<br>(ii) There were deviations from intended interventions that arose because of the trial context<br>AND<br>(iii) These deviations were likely to have affected the outcome<br>AND<br>(iv) These deviations were unbalanced between the intervention groups | (i) An appropriate analysis was not used to estimate the effect of assignment to intervention<br>AND<br>(ii) The potential impact (on the estimated effect of intervention) of the failure to analyse participants in the group to which they were randomized was substantial | 'High' risk of bias for Part 1<br>OR<br>'High' risk of bias for Part 2 |

**Table 6. Mapping of signalling questions to suggested risk-of-bias judgements for bias due to deviations from intended interventions (*effect of assignment to intervention*).** This is only a suggested decision tree: all default judgements can be overridden by assessors.

| Signalling question | | | | | Domain level judgement |
|---|---|---|---|---|---|
| **Part 1: Questions 2.1 to 2.5** | | | | | |
| **2.1 Participants aware?** | **2.2 Personnel aware?** | **2.3 Any deviations?** | **2.4 Affecting outcomes?** | **2.5 Balanced deviations?** | **Default risk of bias for part 1** |
| Both 2.1 & 2.2 N/PN | | NA | NA | NA | Low |
| Either 2.1 or 2.2 Y/PY/NI | | N/PN | NA | NA | Low |
| Either 2.1 or 2.2 Y/PY/NI | | NI | NA | NA | Some concerns |
| Either 2.1 or 2.2 Y/PY/NI | | Y/PY | N/PN | NA | Some concerns |
| Either 2.1 or 2.2 Y/PY/NI | | Y/PY | Y/PY/NI | Y/PY | Some concerns |
| Either 2.1 or 2.2 Y/PY/NI | | Y/PY | Y/PY/NI | N/PN/NI | High |
| **Part 2: Questions 2.6 and 2.7** | | | | | |
| **2.6 Appropriate analysis?** | | **2.7 Potential impact on result due to switching groups in analysis?** | | | **Default risk of bias for part 2** |
| Y/PY | | NA | | | Low |
| N/PN/NI | | N/PN | | | Some concerns |
| N/PN/NI | | Y/PY/NI | | | High |
| **Criteria for the domain** | | | | | |
| 'Low' risk of bias in Part 1 AND 'Low' risk of bias in Part 2 | | | | | Low |
| 'Some concerns' in either Part 1 OR in Part 2, AND NOT 'High' risk in either part | | | | | Some concerns |
| 'High' risk of bias in in either Part 1 OR in Part 2 | | | | | High |

Y/PY = 'Yes' or 'Probably yes'; N/PN = 'No' or 'Probably no'; NI = 'No information'; NA = Not applicable

**Figure 2. Algorithm for suggested judgement of risk of bias due to deviations from the intended interventions (*effect of assignment to intervention*).**

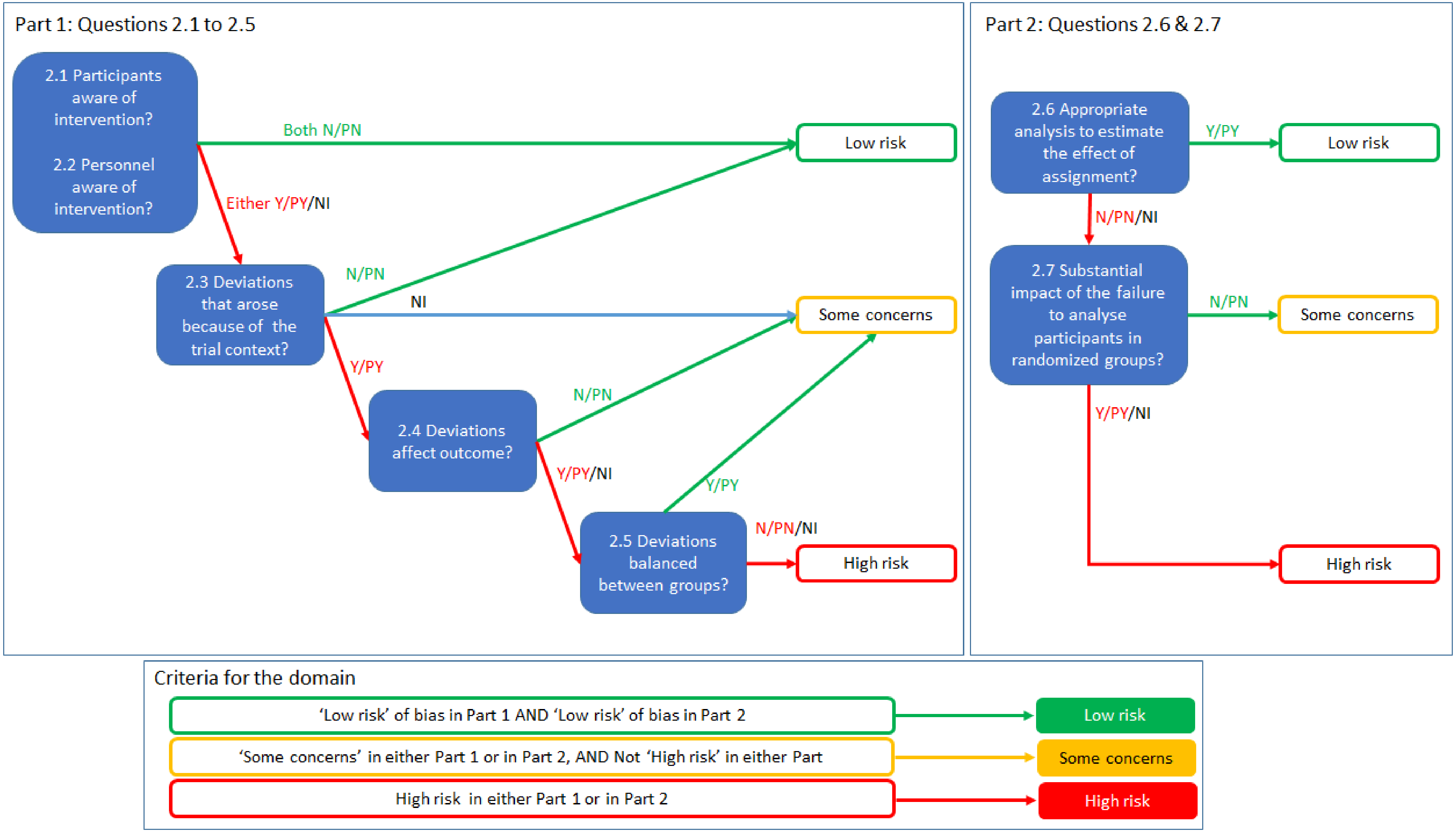

**Box 7. The RoB 2 tool (part 4): Risk of bias due to deviations from the intended interventions (*effect of adhering to intervention*)**

| Signalling questions | Elaboration | Response options |
|---|---|---|
| **2.1. Were participants aware of their assigned intervention during the trial?** | If participants are aware of their assigned intervention it is more likely that health-related behaviours will differ between the intervention groups. Blinding participants, most commonly through use of a placebo or sham intervention, may prevent such differences. If participants experienced side effects or toxicities that they knew to be specific to one of the interventions, answer this question 'Yes' or 'Probably yes'. | Y/PY/PN/N/NI |
| **2.2. Were carers and people delivering the interventions aware of participants' assigned intervention during the trial?** | If carers or people delivering the interventions are aware of the assigned intervention then its implementation, or administration of non-protocol interventions, may differ between the intervention groups. Blinding may prevent such differences. If participants experienced side effects or toxicities that carers or people delivering the interventions knew to be specific to one of the interventions, answer 'Yes' or 'Probably yes'. If randomized allocation was not concealed, then it is likely that carers and people delivering the interventions were aware of participants' assigned intervention during the trial. | Y/PY/PN/N/NI |
| **2.3. [If applicable:] If Y/PY/NI to 2.1 or 2.2: Were important non-protocol interventions balanced across intervention groups?** | This question is asked only if the preliminary considerations specify that the assessment will address imbalance of important non-protocol interventions between intervention groups. Important non-protocol interventions are the additional interventions or exposures that: (1) are inconsistent with the trial protocol; (2) trial participants might receive with or after starting their assigned intervention; and (3) are prognostic for the outcome. Risk of bias will be higher if there is imbalance in such interventions between the intervention groups. | NA/Y/PY/PN/N/NI |
| **2.4. [If applicable:] Were there failures in implementing the intervention that could have affected the outcome?** | This question is asked only if the preliminary considerations specify that the assessment will address failures in implementing the intervention that could have affected the outcome. Risk of bias will be higher if the intervention was not implemented as intended by, for example, the health care professionals delivering care. Answer 'No' or 'Probably no' if implementation of the intervention was successful for most participants. | NA/Y/PY/PN/N/NI |
| **2.5. [If applicable:] Was there non-adherence to the assigned intervention regimen that could have affected participants' outcomes?** | This question is asked only if the preliminary considerations specify that the assessment will address non-adherence that could have affected participants' outcomes. Non-adherence includes imperfect compliance with a sustained intervention, cessation of intervention, crossovers to the comparator intervention and switches to another active intervention. Consider available information on the proportion of study participants who continued with their assigned intervention throughout follow up, and answer 'Yes' or 'Probably yes' if the proportion who did not adhere is high enough to raise concerns. Answer 'No' for studies of interventions that are administered once, so that imperfect adherence is not possible, and all or most participants received the assigned intervention. | NA/Y/PY/PN/N/NI |
| **2.6. If N/PN/NI to 2.3, or Y/PY/NI to 2.4 or 2.5: Was an appropriate analysis used to estimate the effect of** | Both ' naïve 'per-protocol' analyses (excluding trial participants who did not receive their allocated intervention) and 'as treated' analyses (comparing trial participants according to the intervention they actually received) will usually be inappropriate for estimating the effect of adhering to intervention (the 'per-protocol' effect). However, it is possible to use data from a randomized trial to derive an unbiased estimate of the effect of adhering to intervention. Examples of appropriate methods include: (1) instrumental variable analyses to estimate the effect of receiving the assigned intervention | NA/Y/PY/PN/N/NI |

| | | |
|---|---|---|
| **adhering to intervention?** | in trials in which a single intervention, administered only at baseline and with all-or-nothing adherence, is compared with standard care; and (2) inverse probability weighting to adjust for censoring of participants who cease adherence to their assigned intervention, in trials of sustained treatment strategies. These methods depend on strong assumptions, which should be appropriate and justified if the answer to this question is 'Yes' or 'Probably yes'. It is possible that a paper reports an analysis based on such methods without reporting information on the deviations from intended intervention, but it would be hard to judge such an analysis to be appropriate in the absence of such information.<br><br>If an important non-protocol intervention was administered to all participants in one intervention group, adjustments cannot be made to overcome this.<br><br>Some examples of analysis strategies that would not be appropriate to estimate the effect of adhering to intervention are (i) 'Intention to treat (ITT) analysis', (ii) 'per protocol analysis', (iii) 'as-treated analysis', (iv) 'analysis by treatment received'. | |
| **Risk-of-bias judgement** | See Table 7, Table 8 and Figure 3 | Low / High / Some concerns |
| Optional: What is the predicted direction of bias due to deviations from intended interventions? | If the likely direction of bias can be predicted, it is helpful to state this. The direction might be characterized either as being towards (or away from) the null, or as being in favour of one of the interventions. | NA / Favours experimental / Favours comparator / Towards null /Away from null / Unpredictable |

**Table 7. Reaching risk-of-bias judgements for bias due to deviations from intended interventions (*effect of adhering to intervention*)**

| | |
|---|---|
| Low risk of bias | (i.1) Participants, carers and people delivering the interventions were unaware of intervention groups during the trial<br>OR<br>(i.2.1) Participants, carers or people delivering the interventions were aware of intervention groups<br>AND<br>(i.2.2) [If applicable] The important non-protocol interventions were balanced across intervention groups<br>AND<br>(ii) [If applicable] Failures in implementing the intervention could not have affected the outcome<br>AND<br>(iii) [If applicable] Study participants adhered to the assigned intervention regimen |
| Some concerns | (i.1.1) Participants, carers and people delivering the interventions were unaware of intervention groups during the trial<br>AND<br>(i.1.2.1) [If applicable] Failures in implementing the intervention could have affected the outcome<br>OR<br>(i.1.2.2) [If applicable] Study participants did not adhere to the assigned intervention regimen<br>OR<br>(i.2.1) Participants, carers or people delivering the interventions were aware of intervention groups<br>AND<br>(i.2.2) [If applicable] The important non-protocol interventions were balanced across intervention groups<br>AND<br>(i.2.3.1) [If applicable] Failures in implementing the intervention could have affected the outcome<br>OR<br>(i.2.3.2) [If applicable] Study participants did not adhere to the assigned intervention regimen<br>OR<br>(i.3.1) Participants, carers or people delivering the interventions were aware of intervention groups<br>AND<br>(i.3.2) [If applicable] The important non-protocol interventions were not balanced across intervention groups<br>AND<br>(ii) An appropriate analysis was used to estimate the effect of adhering to intervention |
| High risk of bias | (i.1.1) Participants, carers and people delivering the interventions were unaware of intervention groups during the trial<br>AND<br>(i.1.2.1) [If applicable] Failures in implementing the intervention could have affected the outcome<br>OR<br>(i.1.2.2) [If applicable] Study participants did not adhere to the assigned intervention regimen<br>OR |

(i.2.1) Participants, carers or people delivering the interventions were aware of intervention groups
AND
(i.2.2) [If applicable] The important non-protocol interventions were balanced across intervention groups
AND
(i.2.3.1) [If applicable] Failures in implementing the intervention could have affected the outcome
OR
(i.2.3.2) [If applicable] Study participants did not adhere to the assigned intervention regimen

OR

(i.3.1) Participants, carers or people delivering the interventions were aware of intervention groups
AND
(i.3.2) [If applicable] The important non-protocol interventions were not balanced across intervention groups

AND
(ii) An appropriate analysis was not used to estimate the effect of adhering to intervention

**Table 8. Mapping of signalling questions to suggested risk-of-bias judgements for bias due to deviations from intended interventions (*effect adhering to intervention*).** This is only a suggested decision tree: all default judgements can be overridden by assessors.

| Signalling question | | | | | | Domain level judgement |
|---|---|---|---|---|---|---|
| **2.1 Participant aware?** | **2.2 Personnel aware?** | **2.3 Balanced non-protocol-ints?** | **2.4 Failure in implementation affects outcome?** | **2.5 Non-adherence?** | **2.6 Appropriate analysis?** | **Default risk of bias** |
| Both 2.1 & 2.2 N/PN | | NA | N/PN or NA | N/PN or NA | NA | Low |
| Either 2.1 or 2.2 Y/PY/NI | | Y/PY or NA | N/PN or NA | N/PN or NA | NA | Low |
| Both 2.1 & 2.2 N/PN | | NA | Y/PY/NI | N/PN or NA | Y/PY | Some concerns |
| Both 2.1 & 2.2 N/PN | | NA | N/PN or NA | Y/PY /NI | Y/PY | Some concerns |
| Either 2.1 or 2.2 Y/PY/NI | | Y/PY or NA | Y/PY/NI | N/PN or NA | Y/PY | Some concerns |
| Either 2.1 or 2.2 Y/PY/NI | | Y/PY or NA | N/PN or NA | Y/PY /NI | Y/PY | Some concerns |
| Either 2.1 or 2.2 Y/PY/NI | | N/PN/NI | Any response | | Y/PY | Some concerns |
| Both 2.1 & 2.2 N/PN | | NA | Y/PY/NI | N/PN or NA | N/PN/NI | High |
| Both 2.1 & 2.2 N/PN | | NA | N/PN or NA | Y/PY /NI | N/PN/NI | High |
| Either 2.1 or 2.2 Y/PY/NI | | Y/PY or NA | Y/PY/NI | N/PN or NA | N/PN/NI | High |
| Either 2.1 or 2.2 Y/PY/NI | | Y/PY or NA | N/PN or NA | Y/PY/NI | N/PN/NI | High |
| Either 2.1 or 2.2 Y/PY/NI | | N/PN/NI | Any response | | N/PN/NI | High |

Y/PY = 'Yes' or 'Probably yes'; N/PN = 'No' or 'Probably no'; NI = 'No information'; NA = 'Not applicable'

**Figure 3. Algorithm for suggested judgement of risk of bias due to deviations from the intended interventions (*effect adhering to intervention*).** This is only a suggested decision tree: all default judgements can be overridden by assessors.

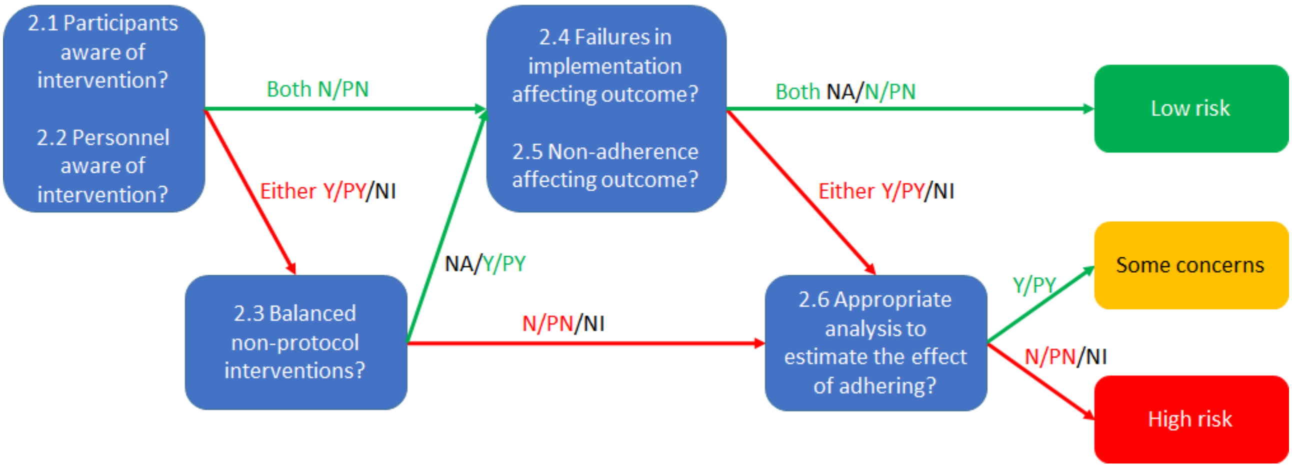

### 6 Detailed guidance: bias due to missing outcome data

#### 6.1 Background

Randomization provides a fair comparison between two or more intervention groups by balancing, on average, the distribution of known and unknown prognostic factors at baseline between the intervention groups. Missing measurements of the outcome, for example due to dropout during the study, may lead to bias in the intervention effect estimate.

Possible reasons for missing outcome data include (83):

- participants withdraw from the study or cannot be located ('loss to follow-up' or 'dropout');
- participants do not attend a study visit at which outcomes should have been measured;
- participants attend a study visit but do not provide relevant data;
- data or records are lost or are unavailable for other reasons; and
- participants can no longer experience the outcome, for example because they have died.

This domain addresses risk of bias due to missing outcome data, including biases introduced by procedures used to impute, or otherwise account for, the missing outcome data.

Some participants may be excluded from an analysis for reasons other than missing outcome data. In particular, a naïve 'per protocol' analysis is restricted to participants who received the intended intervention (see section 1.3.1). Potential bias introduced by such analyses, or by other exclusions of eligible participants for whom outcome data are available, is addressed in the domain 'Bias due to deviations from intended interventions' (see section 5), in which the final signalling questions examine whether the analysis approach was appropriate. **This is a notable change from the previous Cochrane RoB tool for randomized trials**, in which the domain addressing bias due to incomplete outcome data addressed both genuinely missing data and data deliberately excluded by the trial investigators.

##### *6.1.1 Missing outcome data when estimating the effect of assignment to intervention*

The effect of assignment to intervention should be estimated using an intention-to-treat (ITT) analysis (8). As noted in section 1.3.1, the principles underlying ITT analyses are (9, 10):

(1) analyse participants in the intervention groups to which they were randomized, regardless of the intervention they actually received; and
(2) include all randomized participants in the analysis, which requires measuring all participants' outcomes.

While the first and second principles can always be followed for participants for whom outcome data are available, measuring outcome data on all participants is frequently difficult or impossible to achieve in practice. Therefore, it can often only be followed by making assumptions about the missing values of the outcome.

Even when an analysis is described as ITT, it may exclude participants with missing outcome data, and so be at risk of bias. Such analyses are sometimes described as 'modified intention-to-treat' (mITT) analyses, but others use this term in different ways (84). Therefore, assessments of risk of bias due to missing outcome data should be based on the issues addressed in the signalling questions for this domain, and not on the way that trial investigators described the analysis.

##### *6.1.2 Missing outcome data when estimating the effect of adhering to intervention*

As noted above, the potential for bias arising from exclusion of eligible trial participants who did not adhere to their assigned intervention is addressed in the domain 'Bias due to deviations from intended interventions' (see section 5). Such analyses may be additionally biased if participants are excluded due to missing outcome data. Appropriate methods to estimate the effect of adhering to intervention (for example, instrumental variable analyses to estimate the effect of a single intervention that is administered at baseline (5)) may nonetheless produce biased estimates if participants are excluded due to missing outcome data.

The circumstances in which missing outcome data lead to bias are similar regardless of the effect of interest, so there is a single set of signalling questions for this domain.

##### *6.1.3 When do missing outcome data lead to bias?*

Statistical analyses excluding participants with missing outcome data are examples of 'complete-case' analyses (analyses restricted to individuals in whom there were no missing values of included variables). To understand when missing outcome data lead to bias in such analyses, we need to consider:

(1) The **true value of the outcome** in participants with missing outcome data. This is the value of the outcome that should have been measured but was not;
(2) The **missingness mechanism**, which is the process that led to outcome data being missing.

Whether missing outcome data lead to bias in complete-case analyses depends on whether the missingness mechanism is related to the true value of the outcome. In the presence of such a relationship data are often described as 'missing not at random' (MNAR) (85). Equivalently, we can consider whether the measured (non-missing) outcomes differ systematically from the missing outcomes (the true values in participants with missing outcome data). For example, consider a trial of cognitive behavioural therapy compared with usual care for depression. If participants who are more depressed are less likely to return for follow-up, then whether the depression outcome is missing depends on its true value, which implies that the measured depression outcomes will differ systematically from the true values of the missing depression outcomes.

Below, we summarize situations in which missing outcome data do and do not lead to bias in the estimated intervention effect of intervention in a complete-case analysis:

(1) When missingness in the outcome is unrelated to its true value, within each intervention group, missing outcome data **will not lead to bias**. In this situation, the complete cases are representative of those with missing data. For example, missing outcome data would not lead to bias if the missingness occurred by chance, because an automatic measuring device failed.
(2) When missingness in the outcome depends on both the intervention group and the true value of the outcome, missing outcome data **will lead to bias**. For example, the results of a complete-case analysis will be biased in a placebo-controlled trial of an antidepressant drug for symptoms of depression, in which (1) participants with continuing symptoms of depression are more likely to be lost to follow up, and (2) side effects of the drug cause participants assigned to that drug to drop out of the study.
(3) When missingness in the outcome is related to its true value and, additionally, the effect of the experimental intervention differs from that of the comparator intervention, missing outcome data **will lead to bias except in the special case described below**. For example, in the trial of an antidepressant drug described above, its estimated effect on symptoms of depression will be biased if (1) participants with continuing symptoms of depression are more likely to be lost to follow up, and (2) the drug affects symptoms of depression, compared with placebo.
    - The special case is that when the outcome variable is dichotomous, and the intervention effect estimate is an odds ratio, missing outcome data **will not lead to bias** even if missingness depends on the true value of the outcome, providing that missingness is not also related to intervention group assignment. For example, the odds ratio from a trial comparing the effect of an experimental and comparator intervention on all-cause mortality will not be biased by missing outcome data, even if outcome data are more likely to be missing in participants who died, and mortality was less likely in the experimental than comparator intervention group. This is because the proportional reduction in the risk of death applies to both the intervention and control groups, so cancels out in the calculation of the odds ratio. This exception does not apply if the outcome is a dichotomous variable derived from a numerical variable (for example, a classification of hypertension based on measured blood pressure): odds ratios will still be biased in this situation.

    Because many dichotomous outcomes are derived from numerical variables, and because it is difficult to distinguish situations in which outcome data do and do not depend on intervention group assignment, the signalling questions for this domain do not distinguish odds ratios from other measures of intervention effect. Further we do not distinguish between situations in which the effect of the experimental intervention does and does not differ from that of the comparator intervention, because it is usually not possible to exclude a difference (absence of evidence is not evidence of absence).

(4) There are further exceptions, which are not listed because they are hard to identify in practice, and likely to be rare.

If the intervention effect differs between participant subgroups ('effect modification'), then the considerations above apply separately within the subgroups. If missingness in the outcome differs between the subgroups, this will change the overall intervention effect estimate, because the subgroup proportions analysed will differ from those originally randomized. This issue is not considered further, because differences in the distribution of effect modifiers between trials are usually considered to be a source of heterogeneity rather than bias.

###### *6.1.3.1 Analyses that adjust for participant characteristics at baseline*

Some trial analyses adjust for characteristics of participants at baseline, and so exclude participants in whom data on baseline characteristics are missing. Exclusions because of missing data on baseline characteristics only lead to bias if, additionally, missingness in the outcome depends on its true value (providing there is no effect modification by the baseline characteristic). Therefore, they are not considered separately. An exception is if baseline characteristics are collected retrospectively, and the outcome causes missingness in the baseline characteristics: this has the same implications for bias as when missingness in the outcome depends on its the true value, discussed above. For example, in a trial of a new treatment for acute stroke, where the outcome was 24-hour mortality, baseline characteristics such as duration of symptoms could not be collected in those who died before recovering consciousness.

It may be possible to reduce or remove bias due to missing outcome data by accounting for participant characteristics at baseline. For example, suppose that the outcome variable is blood pressure, and that both older people and those in the experimental intervention group were more likely to drop out of the trial. This would lead to bias in the estimated effect of intervention on blood pressure, because older people tend to have higher blood pressure. The bias would be removed in an analysis adjusting for age at baseline, if this fully accounts for the relation between missingness in the outcome and its true value.

###### *6.1.3.2 When is the amount of missing outcome data small enough to exclude bias?*

**Unfortunately, there is no sensible threshold for 'small enough' in relation to the proportion of missing outcome data.**

In situations where missing outcome data lead to bias, the extent of bias will increase as the amount of missing outcome data increases. There is a tradition of regarding a proportion of less than 5% missing outcome data as "small" (with corresponding implications for risk of bias), and over 20% as "large". However, the potential impact of missing data on estimated intervention effects depends on the proportion of participants with missing data, the type of outcome and (for dichotomous outcome) the risk of the event. For example, consider a study of 1000 participants in the intervention group where the observed mortality is 2% for the 900 participants with outcome data (18 deaths). Even though the proportion of data missing is only 10%, if the mortality rate in the 100 missing participants is 20% (20 deaths), the overall true mortality of the intervention group would be nearly double (3.8% vs. 2%) that estimated from the observed data.

##### ***6.1.4 How can we identify evidence of bias due to missing outcome data?***

It is not possible to examine directly whether the chance that the outcome is missing depends on its true value: judgements of risk of bias will depend on the circumstances of the trial. Therefore, we can only be sure that there is no bias due to missing outcome data when: (1) the outcome is measured in all participants; (2) the number of participants with missing outcome data is sufficiently small that their outcomes could have made no important difference to the estimated effect of intervention; or (3) sensitivity analyses (conducted by either the trial investigators or the review authors) confirm that plausible values of the missing outcome data could make no important difference to the estimated intervention effect.

However, indirect evidence that missing outcome data are likely to cause bias can come from examining: (1) differences between the proportion of missing outcome data in the experimental and comparator intervention groups and (2) reasons that outcome data are missing.

###### *6.1.4.1 Differing proportions of missing outcome data*

If the effects of the experimental and comparator interventions on the outcome are different, and missingness in the outcome depends on its true value, then the proportion of participants with missing data is likely to differ between the intervention groups. Therefore, differing proportions of missing outcome data in the experimental and comparator intervention groups provide evidence of potential bias.

It is possible that missing outcome data do not lead to bias, even when the proportion of missing outcome data differs between intervention groups. This will only be the case if the chance of the outcome being missing is not related to its true value: for example if engagement with the trial was greater in the experimental than comparator intervention group but engagement, and hence missing outcome data, was not related to the outcome. If the proportion of missing outcome data differs between the intervention groups, then the risk of bias is higher, because the trial results will be sensitive to missingness in the outcome being related to its true value.

###### *6.1.4.2 Examining reasons for missing outcome data*

Trial reports may provide reasons why participants have missing data. For example, trials of haloperidol to treat dementia reported various reasons such as 'lack of efficacy', 'adverse experience', 'positive response', 'withdrawal of consent' and 'patient ran away', and 'patient sleeping' (86). It is likely that some of these (for example, 'lack of efficacy' and 'positive response') are related to the true values of the missing outcome data. Therefore, these reasons increase the risk of bias if the effects of the experimental and comparator interventions differ, or if the reasons are related to intervention group (for example, 'adverse experience').

The situation most likely to lead to bias is when reasons for missing outcome data differ between the intervention groups: for example if participants who became seriously unwell withdrew from the comparator group while participants who recovered withdrew from the experimental intervention group.

In practice, our ability to assess risk of bias will be limited by the extent to which trial investigators collected and reported reasons that outcome data were missing.

##### *6.1.5 Time-to-event data*

Many trial analyses use methods for 'time-to-event' data, in which the outcome is a dichotomous variable that indicates whether the outcome event was observed in each participant. The follow up time ends either when the outcome event occurs or when observation stops for other reasons. Follow-up times for participants in whom the outcome event was not observed before observation stopped are said to be 'censored'. Intervention effects in analyses of time-to-event data are typically estimated as rate ratios or hazard ratios. Results of time-to-event analyses will be unbiased only if censoring is 'non-informative', which means that censoring times for censored participants are unrelated to the (subsequent) times at which outcome events occur. For example, if all participants are followed until a specified date after which follow up ends, then censoring can be assumed to be non-informative.

Informative censoring implies that the chance that the outcome is not observed depends on its true value. For example, there would be informative censoring if participants who were lost to follow up were more likely to die than participants who were retained in care. In the presence of informative censoring, a time-to-event analysis will be biased if:

- the chance that the follow up is censored also depends on the intervention group (for example, if censoring is more likely because participants in the experimental intervention group are lost to follow up because of severe side effects); and
- the effect of the experimental and comparator interventions on the outcome differs.

Either differences in rates of censoring or differing reasons for censoring may provide evidence that censoring was informative.

**A particular risk of bias arises when participants' follow up is censored if they stop or change their assigned intervention**, for example because of drug toxicity or, in cancer trials, when participants switch to second-line chemotherapy. Participants censored during trial follow-up, for example because they withdrew from the study, should be regarded as having missing outcome data, even though some of their follow up is included in analyses. Note CONSORT flow diagrams may show such participants as included in analyses in.

##### *6.1.6 Statistical methods for handling missing outcome data*

Trial investigators may present statistical analyses (in addition to or instead of complete case analyses) that attempt to address the potential for bias caused by missing outcome data (83, 87-89). The most common approaches are:

(1) single imputation (i.e., generate a complete dataset by filling in the missing values of the outcome);

(2) multiple imputation (generate multiple complete datasets based on a predictive distribution for the outcome variable);
(3) methods that weight participants with measured outcome data in order to adjust for the missing data (90); and
(4) methods that do not require a complete data set such as likelihood-based methods, moment-based methods, and semiparametric models for survival data (83, 91).

Imputation approaches replace missing values by one or more new values. In single imputation, only one estimate is filled in. Commonly used approaches include 'last observation carried forward' (LOCF) and 'baseline observation carried forward' (BOCF). Each of these is unlikely to remove the bias that occurs when missingness in the outcome depends on its true value, unless there is no change in the outcome after the last time it was measured. Further, they generally improve precision artificially (so that the confidence interval for the intervention effect estimate is too narrow), since they do not reflect uncertainty about the missing outcomes. Therefore, intervention effect estimates based on these methods should be considered as at low risk of bias only when there is clear justification (86, 92).

In multiple imputation, multiple values of the missing outcomes are drawn at random from a predictive distribution, forming multiple distinct filled-in datasets (93, 94). These multiple datasets are analysed to produce a single summary estimate and confidence interval that reflect the uncertainty associated with missing data (unlike single imputation methods). However, multiple imputation methods will not remove or reduce the bias that occurs when missingness in the outcome depends on its true value, unless such missingness can be explained by measured variables. In particular, imputing missing outcome data based only on intervention group will give results that are near-identical to those from a complete-case analysis, and does not reduce or remove bias when outcome data are MNAR. If the imputation model (the model used to predict the missing values of the outcome) is not correctly specified, multiple imputation will not remove, and can even increase, bias. Multiple imputed estimates should be considered as at low risk of bias only when there is justification for the assumption that missingness in the outcome does not depend on its true value other than through measured variables included in the imputation model, and where there is justification for the variables included and the form of the imputation model.

It may be possible to reduce bias associated with missing outcome data when values of the outcome are measured repeatedly over time and these measurements are used to predict the missing outcome data (95). Even when such an approach is used, review authors should consider carefully whether loss to follow up is plausibly related to the outcome trajectory after the last recorded measurement.

Bias because of loss to follow up can also be addressed by modelling its probability over time and using the probability of loss to follow up to give more weight to participants who resemble those lost. The aim is to conduct a weighted analysis in which characteristics of participants after baseline are unrelated to loss to follow up: this is an example of a semiparametric approach to dealing with missing data (see approach (3) above). As with imputation-based approaches, such analyses will not remove bias if missingness in the outcome depends on its true value, even after accounting for the variables used to derive the weights. Weighting will only remove bias if the model for the probability of loss to follow up is correctly specified.

##### *6.1.7 Sensitivity analyses*

Sensitivity analyses can be performed to assess the potential impact of missing outcome data, based on assumptions about the relationship between missingness in the outcome and its true value. They may be conducted by trial investigators or by review authors. However, they are only helpful in judging risk of bias if they address the potential relationship between missingness in the outcome and its true value. The methods summarized in section 6.1.6 can be extended to conduct such sensitivity analyses (96, 97).

Several methods are available for review authors to assess the robustness of a result from a randomized trial in the presence of missing data (98). For dichotomous outcomes, Higgins and colleagues propose a strategy involving different assumptions about how the risk of the event among the missing participants differs from the risk of the event among the observed participants, taking account of uncertainty introduced by the assumptions (86). Akl and colleagues propose a suite of simple imputation methods, including a similar approach to that of Higgins and colleagues based on relative risks of the event in missing versus observed participants (99). Similar ideas can be applied to continuous outcome data (100-102). Particular care is required to avoid double counting events, since it can be unclear whether reported numbers of events in trial reports apply to the full randomized sample or only to those who did not drop out (103).

Although there is a tradition of implementing 'worst case' and 'best case' analyses clarifying the extreme boundaries of what is theoretically possible, such analyses may not be informative for the most plausible scenarios (86).

#### 6.2 Empirical evidence of bias due to missing outcome data

Empirical research has investigated the adequacy with which missing data are addressed in reports of trials. A study that included 71 trial reports from four general medical journals, concluded that missing data are common and often inadequately handled in the statistical analysis (104).

Concerns over bias resulting from missing outcome data are driven mainly by theoretical considerations. Several empirical studies have looked at whether various aspects of missing data are associated with the magnitude of effect estimates (18, 34, 74, 79, 105-107). In a systematic review of meta-epidemiological studies, missing data were associated with overestimation of effect estimates in some studies, but underestimation or no difference in others(33). Many of the studies do not differentiate between missing outcome data and participants being excluded from the analysis (see also section 5.2). In one study, having a dropout rate >20% (versus ≤20%) was not associated with different effect estimates on average (74).

#### 6.3 Using this domain of the tool

(1) Risk of bias will be low if outcome data are available for all, or nearly all, randomized participants. The meaning of 'nearly all' in this context is that the number of participants with missing outcome data is so small that their outcomes, whatever they were, could have made no important difference to the estimated effect of intervention. If this is the case then no further signalling questions need be answered. Absence of information about the extent of missing outcome data (for example, when no CONSORT flow diagram was provided in the trial report) will usually lead to a judgement of high risk of bias for this domain.

(2) Risk of bias will be low if sensitivity analyses, conducted by either the trial investigators or the review authors (see section 6.1.7), confirm that the finding is robust to plausible values of the missing outcome data. Such analyses are likely to be particularly useful when the amount of missing data is sufficiently large for the potential impact on the estimated effect of intervention to be substantial. If sensitivity analyses confirm that the result is robust, then the result may be regarded as at low risk of bias and no further signalling questions need be answered.

(3) As explained in section 6.1.3, missing outcome data can only lead to bias if the chance that the outcome is missing depends on its true value. It may be possible to exclude this based on reported reasons for missing outcome data (for example, if outcome data are only missing because of failure of a measuring instrument or closure of a centre in a multicentre trial). However, if it is possible that **missingness in the outcome could depend on its true value** then review investigators will need to consider the proportions of and reasons for missing outcome data.

(4) A difference between the experimental and comparator intervention groups in the proportions of missing outcome data may indicate a risk of bias (see section 6.1.4). For time-to-event-data, review authors should consider whether rates of censoring (loss to follow-up) differ between the intervention groups.

(5) Either reasons for missing outcome data reported by trial investigators, or the circumstances of the trial, may lead review authors to conclude that it is **likely that missingness in the outcome depended on its true value**.

(6) Differing reasons for missing outcome data in the experimental and comparator intervention groups may lead to substantial bias. For example, in a trial of an experimental intervention aimed at smoking cessation there would be serious bias if some comparator intervention participants left the study due to a lack of enthusiasm at receiving nothing novel (and continued to smoke) while some experimental intervention participants left the study due to successful cessation of smoking.

#### 6.4 Signalling questions and criteria for judging risk of bias

Signalling questions for this domain are provided in Box 8. Criteria for reaching risk-of-bias judgements are given in Table 11, and an algorithm for implementing these is provided in Table 12 and Figure 4.

**Box 8. The RoB 2 tool (part 5): Risk of bias due to missing outcome data**

| Signalling questions | Elaboration | Response options |
|---|---|---|
| **3.1 Were data for this outcome available for all, or nearly all, participants randomized?** | The appropriate study population for an analysis of the intention to treat effect is all randomized participants.<br>"Nearly all" should be interpreted as that the number of participants with missing outcome data is sufficiently small that their outcomes, whatever they were, could have made no important difference to the estimated effect of intervention.<br>For continuous outcomes, availability of data from 95% of the participants will often be sufficient. For dichotomous outcomes, the proportion required is directly linked to the risk of the event. If the observed number of events is much greater than the number of participants with missing outcome data, the bias would necessarily be small.<br>Only answer 'No information' if the trial report provides no information about the extent of missing outcome data. This situation will usually lead to a judgement that there is a high risk of bias due to missing outcome data.<br>Note that imputed data should be regarded as missing data, and not considered as 'outcome data' in the context of this question. | Y/PY/PN/N/NI |
| **3.2 If N/PN/NI to 3.1: Is there evidence that the result was not biased by missing outcome data?** | Evidence that the result was not biased by missing outcome data may come from: (1) analysis methods that correct for bias; or (2) sensitivity analyses showing that results are little changed under a range of plausible assumptions about the relationship between missingness in the outcome and its true value. However, imputing the outcome variable, either through methods such as 'last-observation-carried-forward' or via multiple imputation based only on intervention group, should not be assumed to correct for bias due to missing outcome data. | NA/Y/PY/PN/N |
| **3.3 If N/PN to 3.2: Could missingness in the outcome depend on its true value?** | If loss to follow up, or withdrawal from the study, could be related to participants' health status, then it is possible that missingness in the outcome was influenced by its true value. However, if all missing outcome data occurred for documented reasons that are unrelated to the outcome then the risk of bias due to missing outcome data will be low (for example, failure of a measuring device or interruptions to routine data collection).<br>In time-to-event analyses, participants censored during trial follow-up, for example because they withdrew from the study, should be regarded as having missing outcome data, even though some of their follow up is included in the analysis. Note that such participants may be shown as included in analyses in CONSORT flow diagrams. | NA/Y/PY/PN/N/NI |
| **3.4 If Y/PY/NI to 3.3: Is it likely that missingness in the outcome depended on its true value?** | This question distinguishes between situations in which (i) missingness in the outcome could depend on its true value (assessed as 'Some concerns') from those in which (ii) it is likely that missingness in the outcome depended on its true value (assessed as 'High risk of bias'). Five reasons for answering 'Yes' are:<br>1. Differences between intervention groups in the proportions of missing outcome data. If there is a difference between the effects of the experimental and comparator interventions on the outcome, and the missingness in the outcome is influenced by its true value, then the proportions of missing outcome data are likely to differ between intervention groups. Such a difference suggests a risk of bias due to missing outcome data, because the trial result will be sensitive to missingness in the outcome being related to its true value. For time-to-event-data, the analogue is that rates of censoring (loss to follow-up) differ between the intervention groups.<br>2. Reported reasons for missing outcome data provide evidence that missingness in the outcome depends on its true value; | NA/Y/PY/PN/N/NI |

| | | |
|---|---|---|
| | 3. Reported reasons for missing outcome data differ between the intervention groups;<br>4. The circumstances of the trial make it likely that missingness in the outcome depends on its true value. For example, in trials of interventions to treat schizophrenia it is widely understood that continuing symptoms make drop out more likely.<br>5. In time-to-event analyses, participants' follow up is censored when they stop or change their assigned intervention, for example because of drug toxicity or, in cancer trials, when participants switch to second-line chemotherapy.<br>Answer 'No' if the analysis accounted for participant characteristics that are likely to explain the relationship between missingness in the outcome and its true value. | |
| **Risk-of-bias judgement** | See Table 9, Table 10 and Figure 4. | Low / High / Some concerns |
| Optional: What is the predicted direction of bias due to missing outcome data? | If the likely direction of bias can be predicted, it is helpful to state this. The direction might be characterized either as being towards (or away from) the null, or as being in favour of one of the interventions. | NA / Favours experimental / Favours comparator / Towards null /Away from null / Unpredictable |

**Table 9. Reaching risk-of-bias judgements for bias due to missing outcome data**

| | |
|---|---|
| Low risk of bias | (i) Outcome data were available for all, or nearly all, randomized participants<br>OR<br>(ii) There is evidence that the result was not biased by missing outcome data<br>OR<br>(iii) Missingness in the outcome could not depend on its true value |
| Some concerns | (i) Outcome data were not available for all, or nearly all, randomized participants<br>AND<br>(ii) There is not evidence that the result was not biased by missing outcome data<br>AND<br>(iii) Missingness in the outcome could depend on its true value<br>AND<br>(iv) It is not likely that missingness in the outcome depended on its true value |
| High risk of bias | (i) Outcome data were not available for all, or nearly all, randomized participants<br>AND<br>(ii) There is not evidence that the result was not biased by missing outcome data<br>AND<br>(iii) Missingness in the outcome could depend on its true value<br>AND<br>(iv) It is likely that missingness in the outcome depended on its true value. |

**Table 10. Mapping of signalling questions to suggested risk-of-bias judgements for bias due to missing outcome data.** This is only a suggested decision tree: all default judgements can be overridden by assessors.

| Signalling question | | | | Domain-level judgement |
|---|---|---|---|---|
| 3.1 | 3.2 | 3.3 | 3.4 | Default risk of bias |
| Complete data? | Evidence of no bias? | Could depend on true? | Likely depend on true? | |
| Y/PY | NA | NA | NA | Low |
| N/PN/NI | Y/PY | NA | NA | Low |
| N/PN/NI | N/PN | N/PN | NA | Low |
| N/PN/NI | N/PN | Y/PY/NI | N/PN | Some concerns |
| N/PN/NI | N/PN | Y/PY/NI | Y/PY/NI | High |

Y/PY = 'Yes' or 'Probably yes'; N/PN = 'No' or 'Probably no'; NI = 'No information'; NA = 'Not applicable'

**Figure 4. Algorithm for suggested judgement of risk of bias for bias due to missing outcome data.** This is only a suggested decision tree: all default judgements can be overridden by assessors.

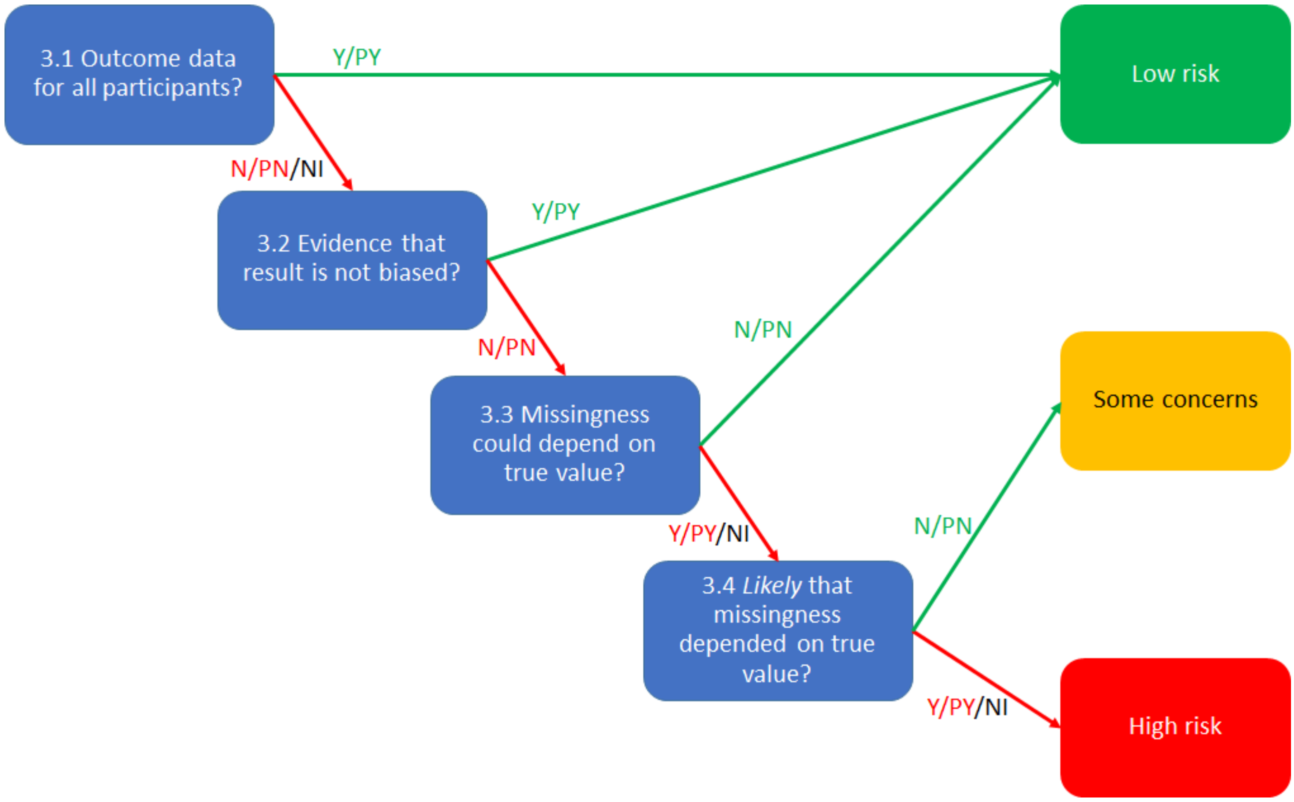

### 7 Detailed guidance: bias in measurement of the outcome

#### 7.1 Background

Errors in measuring of participants' outcome variables arise when the measured values do not equal the true or underlying values. Such errors can bias estimates of intervention effect from a randomized trial. These errors are often referred to as **measurement error** (for continuous outcomes), **misclassification** (for dichotomous or categorical outcomes) or **under-ascertainment/over-ascertainment** (for events). Errors in measurement may be **differential** or **non-differential** in relation to intervention assignment.

- Differential measurement errors are related to intervention assignment. Such errors are systematically different between experimental and comparator intervention groups, and are less likely when outcome assessors are blinded to intervention assignment.
- Non-differential measurement errors are unrelated to intervention assignment.

This domain relates primarily to differential errors. Non-differential errors may be addressed if they are considered to be important but are not addressed in detail, for reasons explained in Box 9.

**Box 9. Implications of non-differential measurement error for intervention effect estimates**

Errors in measurement or classification of an outcome that are unrelated to intervention assignment are known as 'non-differential' errors. They will generally not cause bias in intervention effect estimates of mean differences for continuous outcomes but are likely to attenuate (i.e. produce bias towards the null in) intervention effect estimates such as odds ratios, risk ratios or hazard ratios when the outcome is dichotomous or categorical. There are situations in which non-differential measurement error can bias effect estimates away from the null, but these are usually considered unlikely to occur in randomized trials.

The consequences of non-differential measurement error in an outcome depend on the intervention effect measure and the nature of the measurement error. For example:

- There will be no bias due to non-differential errors if the effect measure is a difference measure (such as a mean difference) and the measurement error acts additively on the outcome. For instance, if a blood pressure measuring device used in both intervention groups systematically produces a measurement that is 10 mmHg too high (an additive error), then this error will apply equally to mean values in the experimental and comparator intervention groups, and on computation of the difference between the means, the errors will 'cancel out'.
- There will be bias due to non-differential errors if the effect measure is a difference measure and both (1) the measurement error acts multiplicatively on the outcome and (2) the intervention effect is non-zero. For instance, if the blood pressure measuring device systematically produces a measurement that is 120% of the truth (a multiplicative error), then:
  - o in the absence of an intervention effect the mean difference will be unbiased (taking the correct value of zero); but
  - o in the presence of an intervention effect, the difference in means will be over-estimated by 20% (e.g. true means of 100 and 110 mmHg (difference 10 mmHg) would be measured as 120 and 132 mmHg (difference 12 mmHg));
  - o if blood pressure measurements are used to classify participants as hypertensive or normotensive then estimates of risk ratios or odds ratios will be biased in the presence of an intervention effect.

  Note that errors in continuous measurements are typically assumed to be additive rather than multiplicative.

- For dichotomous outcomes, the effects of non-differential measurement error are different for different effect measures (i.e. odds ratios, risk ratios and risk differences). As for continuous outcomes, the key consideration is whether the error acts in a manner that is 'congruent' with the effect measure. A detailed discussion is provided by Rothman and colleagues (108).

The first signalling question in this domain asks whether the method of measuring the outcome was inappropriate, and can be used to highlight situations in which non-differential measurement errors are particularly important. Addressing non-differential measurement error in more detail would add substantial complexity. Furthermore, a meaningful assessment would require primary reports of trials to distinguish explicitly between the outcome and the way that it is measured, and to consider the relationship between the true value and the measured value so that judgements can be made about whether errors are 'congruent' with the chosen intervention effect measures. For example, hypertension is usually diagnosed based on one or more blood pressure measurements, each of which is subject to measurement error. However, a trial result based on whether participants were diagnosed with hypertension at the end of follow is not usually considered inherently biased due to non-differential measurement error.

Consideration of risk of bias in this domain depends on:

(1) whether the method of measuring the outcome is appropriate;
(2) whether measurement or ascertainment of the outcome differs, or could differ, between intervention groups;
(3) who is the outcome assessor;
(4) whether the outcome assessor is blinded to intervention assignment; and
(5) whether the assessment of outcome is likely to be influenced by knowledge of intervention received.

(1) Outcomes in randomized trials should be assessed appropriately. For example, a portable blood glucose machine used by participants in a trial comparing insulin intervention with placebo may not reliably measure levels below 3.1 mmol. The machine would then be unable to detect differences in rates of severe hypoglycaemia, with consequent under-representation of the true incidence of this adverse effect. Such a measurement method would be inappropriate for this outcome. Alternatively, a measurement instrument may have been demonstrated to have such poor validity that it does not adequately measure the outcome variable.

(2) Outcomes should be measured or ascertained using a method that is comparable across intervention groups. This is usually the case for pre-specified outcomes. However, problems may arise with passive collection of outcome data, as is often the case for unexpected adverse effects. For example:

- In a placebo-controlled trial, severe headaches might occur more frequently in participants assigned to a new drug than in those assigned to placebo. These headaches might lead to more MRI scans being done in the experimental intervention group, and therefore to more diagnoses of symptomless brain tumours, even though the drug does not increase the incidence of brain tumours. This would lead to bias if the outcome is defined as presence of a brain tumour (although not if the outcome is defined as *diagnosis* of a brain tumour).
- Clemens et al identified the potential for what they called "diagnostic testing bias" in trials of the protective effect of BCG vaccine against tuberculosis (109). BCG vaccination usually leads to an easily identified scar, and there is potential for bias if tuberculosis is identified only using passive follow up, and either participants are less likely to seek care or assessors are less likely to order a radiograph if a scar is present. A systematic review found evidence that estimated protection was lower in trials assessed as at higher risk of such bias (110).

Such bias was described by Sackett: "an innocent exposure may become suspect if, rather than causing a disease, it causes a sign or symptom which precipitates a search for the disease" (111).

Even for a pre-specified outcome measure, the nature of the intervention may lead to methods of measuring the outcome that are not comparable across intervention groups. For example, an intervention involving additional visits to a healthcare provider may lead to additional opportunities for outcome events to be identified, compared with the comparator intervention.

(3) The outcome assessor can be:

- the **participant** when the outcome is a participant-reported outcome such as pain, quality of life, or self-completed questionnaire evaluating depression, anxiety or function;
- the **intervention provider** when the outcome is the result of a clinical examination, the occurrence of a clinical event or a therapeutic decision such as a decision to offer a surgical intervention or to discharge the patient; or
- an **outcome assessor** who is an observer not directly involved in the intervention provided to the participant, such as an adjudication committee, a biologist performing an automated test, or a health professional recording outcomes for inclusion in health records or disease registries.

(4) Blinding of outcome assessors is often possible (and often done) even when blinding of participants and personnel during the trial is not feasible. However, it is particularly difficult for participant-reported outcomes: for example, in a trial comparing surgery with medical management when the outcome is a participant's pain at 3 months, it is impossible to blind the assessor (the participant). Inability to blind outcome assessors does not mean that the resulting potential for bias can be ignored: review authors must always assess the risk of bias due to error in measuring the outcome.

(5) For trials in which outcome assessors are not blinded, whether the assessment of outcome is likely to be influenced by knowledge of the intervention received will depend on the observers' preconceptions and on the degree of judgement involved in assessing an outcome. The latter depends on the type of outcome, as some outcomes have no or little room for judgement (e.g. all-cause mortality) and other outcome have considerable room for judgement (e.g. assessment of depression scores). We distinguish five different type of outcomes as follows.

**1 Participant-reported outcomes**

Participant-reported outcomes are any reports coming directly from participants about how they function or feel in relation to a health condition and its therapy, without interpretation of the participant's responses by a clinician, or anyone else. Participant-reported outcomes include any outcome evaluation obtained directly from participants through interviews, self-completed questionnaires, diaries or other data collection tools such as hand-held devices and web-based forms (112). Examples include pain, nausea and health-related quality of life.

The outcome assessor here is **the participant,** even if a blinded interviewer is questioning the participant and completing a questionnaire on their behalf. The interviewer is not considered to be the outcome assessor in a strict sense but rather a facilitator of the measurement.

For participant-reported outcomes, the assessment of outcome is **potentially influenced** by knowledge of intervention received, leading to a judgement of at least 'Some concerns'. Review authors will need to judge whether it is likely that participants' reporting of the outcome was influenced by knowledge of intervention received, in which case risk of bias is considered to be high. For example, a severe or unexpected adverse effect recorded some time after the start of the intervention may be considered unlikely to be influenced by knowledge of the intervention received. On the other hand, level of pain reported at the end of a course of acupuncture, in a study comparing acupuncture with no treatment, is likely to be affected by knowledge of the intervention received.

**2 Observer-reported outcomes not involving judgement**

These are outcomes reported by an external observer (e.g. an intervention provider, independent researcher, or physician not involved in the care provided to participants such as a radiologist) that do not involve any judgement from the observer. Examples include all-cause mortality or the result of an automated test.

The outcome assessor here is **the observer**. For observer-reported outcomes not involving judgement the assessment of outcome is usually **not likely to be influenced** by knowledge of intervention received.

**3 Observer-reported outcomes involving some judgement**

These are outcomes reported by an external observer (e.g. an intervention provider) that involve some judgement, such as is involved in a clinical examination. Examples include tests involving assessment of a radiograph, clinical examination and clinical events other than death (e.g. myocardial infarction) that require judgements based on medical records.

The outcome assessor here is **the observer**. If the observer is aware of the intervention received then assessment of the outcome is **potentially influenced** by this knowledge, leading to a judgement of at least 'Some concerns'. Review authors will need to judge whether it is likely that assessment of the outcome was influenced by knowledge of intervention received, in which case risk of bias is considered to be high.

**4 Outcomes that reflect decisions made by the intervention provider**

These are outcomes that reflect a decision made by the intervention provider. The recording of this decision does not involve any judgement. However, the decision itself can be influenced by knowledge of intervention received. For example, in a trial comparing the impact of laparoscopic versus small-incision cholecystectomy on hospital stay, it was essential to keep the carers blinded to the intervention received to make sure their decision to discharge participants was influenced only by the clinical evaluation of the participants. In general, examples of intervention provider decision outcomes include hospitalization, stopping treatment, referral to a different ward, performing a caesarean section, stopping ventilation and discharge of the participant.

The outcome assessor here is **the care provider making the decision**. The assessment of outcome is usually **likely to be influenced** by knowledge of intervention received, if the care provider is aware of this. This is particularly important when preferences, expectations or hunches regarding the effect of the experimental intervention are strong.

###### 5 Composite outcomes

A composite outcome combines multiple end points into a single outcome. Typically, participants who have experienced any of a specified set of endpoints are considered to have experienced the composite outcome. Examples include major adverse cardiac and cerebrovascular events (MACCE). Composite endpoints can also be constructed from continuous outcome measures.

Assessment of risk of bias for composite outcomes should take into account the frequency or contribution of each component of the composite outcome and take into account the risk of bias due to the most influential components.

#### 7.2 Empirical evidence of bias in measurement of the outcome

Empirical evidence of bias in measurement of the outcome largely comes from studies exploring whether reporting of “double blinding” is associated with intervention effects. These studies are summarized in section 5.2. Lack of blinding of outcome assessors in randomized trials has been associated with more exaggerated estimated intervention odds ratios, by 34% on average (113).

#### 7.3 Using this domain of the tool

The first question in this domain is a screening question to identify rare instances when the method of measuring the outcome was inappropriate. This is unlikely to be the case for pre-specified outcomes in randomized trials, but may be the case for some adverse effects, particularly if there are no systems in place to define the adverse outcome or to identify its occurrence. This question **does not aim to assess whether the choice of outcome being evaluated was sensible** (e.g. because it is an appropriate surrogate or proxy for the main outcome of interest).

The second question addresses whether measurement or ascertainment of the outcome measurement could have differed between the intervention groups. Methods are likely to be comparable in most randomized trials. The question aims to identify situations in which there were systematic differences between the groups, for example because the experimental intervention involved more encounters with healthcare professionals and so led to more opportunities to identify the outcome than in the comparator group, or because of ‘diagnostic detection bias’ in the context of passively collected outcomes (see section 7.1).

The subsequent questions address blinding and its potential implications for assessments of outcomes. It is important to determine whether outcome assessments were made blinded to intervention assignment. If blinding was successfully implemented, then the risk of bias due to differential measurement error is low. Note that outcomes that are assessed using an interaction between a participant and a data collector (for example, when data collectors ask questions or help participants to complete a questionnaire) then if either the participant is blinded and the data collector is not, or the data collector is blinded and the participant is not, then the outcome assessors should be considered to be aware of intervention received unless convincing evidence is available to the contrary.

The importance of lack of blinding of the outcome assessor will depend on the extent to which the assessment can be influenced by knowledge of the intervention assignment. For outcomes such as all-cause mortality, assessment of the outcome is unlikely to have been influenced, but for subjective outcomes such as ‘clinical impression of improvement’, knowledge of the intervention received could be highly influential.

When the outcome assessor is not blinded and the outcome **could have** been influenced by knowledge of intervention received, review authors should assess whether **it is likely that** such influence occurred. In doing so, they should account for both conscious and subconscious influences on the outcome assessor (both 'interests' and 'expectations'). These could vary according to:

- the comparator (higher risk of bias if the comparator is no treatment or usual care than when the comparator is another active intervention, because in the latter case participants may not have a prior belief that one of the active interventions is more beneficial than the other),
- involvement of the outcome assessor in participants' care (lower risk of bias if the outcome assessor is an independent researcher, e.g. a surgeon, who may have a vested interest in demonstrating that their care was effective).
- influence of other actors. For example, for participant-reported outcomes recorded through interview, risk of bias might be lower if the person interviewing the participant is an independent researcher compared with when the interviewer is the care provider and involved in the administration of the intervention (such as surgeons interviewing patients).

Timing of outcome assessment could also influence the likelihood that assessment of the outcome was influenced by knowledge of intervention received. For example, in a trial comparing 12 weeks of supervised exercise with shockwave treatment (114), if the assessment is 1 year after randomization then knowledge of intervention received might not matter very much. This may particularly be the case if many other treatments have been used by participants in the interim, so that participants' recall is unlikely to be influenced by what they received initially.

#### 7.4 Signalling questions and criteria for judging risk of bias

Signalling questions for this domain are provided in Box 10. Criteria for reaching risk-of-bias judgements are given in Table 11 and an algorithm for implementing these is provided in Table 12 and Figure 5.

**Box 10. The RoB 2 tool (part 6): Risk of bias in measurement of the outcome**

| Signalling questions | Elaboration | Response options |
|---|---|---|
| **4.1 Was the method of measuring the outcome inappropriate?** | This question aims to identify methods of outcome measurement (data collection) that are unsuitable for the outcome they are intended to evaluate. The question *does not* aim to assess whether the choice of outcome being evaluated was sensible (e.g. because it is a surrogate or proxy for the main outcome of interest). In most circumstances, for pre-specified outcomes, the answer to this question will be 'No' or 'Probably no'.<br>Answer 'Yes' or 'Probably yes' if the method of measuring the outcome is inappropriate, for example because:<br>(1) it is unlikely to be sensitive to plausible intervention effects (e.g. important ranges of outcome values fall outside levels that are detectable using the measurement method); or<br>(2) the measurement instrument has been demonstrated to have poor validity. | Y/PY/PN/N/NI |
| **4.2 Could measurement or ascertainment of the outcome have differed between intervention groups?** | Comparable methods of outcome measurement (data collection) involve the same measurement methods and thresholds, used at comparable time points. Differences between intervention groups may arise because of 'diagnostic detection bias' in the context of passive collection of outcome data, or if an intervention involves additional visits to a healthcare provider, leading to additional opportunities for outcome events to be identified. | Y/PY/PN/N/NI |
| **4.3 If N/PN/NI to 4.1 and 4.2: Were outcome assessors aware of the intervention received by study participants?** | Answer 'No' if outcome assessors were blinded to intervention status. For participant-reported outcomes, the outcome assessor is the study participant. | NA/Y/PY/PN/N/NI |
| **4.4 If Y/PY/NI to 4.3: Could assessment of the outcome have been influenced by knowledge of intervention received?** | Knowledge of the assigned intervention could influence participant-reported outcomes (such as level of pain), observer-reported outcomes involving some judgement, and intervention provider decision outcomes. They are unlikely to influence observer-reported outcomes that do not involve judgement, for example all-cause mortality. | NA/Y/PY/PN/N/NI |
| **4.5 If Y/PY/NI to 4.4: Is it likely that assessment of the outcome was influenced by knowledge of intervention received?** | This question distinguishes between situations in which (i) knowledge of intervention status could have influenced outcome assessment but there is no reason to believe that it did (assessed as 'Some concerns') from those in which (ii) knowledge of intervention status was likely to influence outcome assessment (assessed as 'High'). When there are strong levels of belief in either beneficial or harmful effects of the intervention, it is more likely that the outcome was influenced by knowledge of the intervention received. Examples may include patient-reported symptoms in trials of homeopathy, or assessments of recovery of function by a physiotherapist who delivered the intervention. | NA/Y/PY/PN/N/NI |
| **Risk-of-bias judgement** | See Table 11, Table 12 and Figure 5. | Low / High / Some concerns |

| | | |
|---|---|---|
| Optional: What is the predicted direction of bias in measurement of the outcome? | If the likely direction of bias can be predicted, it is helpful to state this. The direction might be characterized either as being towards (or away from) the null, or as being in favour of one of the interventions. | NA / Favours experimental / Favours comparator / Towards null /Away from null / Unpredictable |

**Table 11. Reaching risk-of-bias judgements for bias in measurement of the outcome**

| | |
|---|---|
| Low risk of bias | (i) The method of measuring the outcome was not inappropriate<br>AND<br>(ii) The measurement or ascertainment of the outcome did not differ between intervention groups<br>AND<br>(iii.1) The outcome assessors were unaware of the intervention received by study participants<br>OR<br>(iii.2) The assessment of the outcome could not have been influenced by knowledge of the intervention received |
| Some concerns | (i.1) The method of measuring the outcome was not inappropriate<br>AND<br>(i.2) The measurement or ascertainment of the outcome did not differ between intervention groups<br>AND<br>(i.3) The assessment of the outcome <u>could</u> have been influenced by knowledge of the intervention received<br>AND<br>(i.4) It is unlikely that assessment of the outcome was influenced by knowledge of intervention received<br>OR<br>(ii.1) The method of measuring the outcome was not inappropriate<br>AND<br>(ii.2) There is no information on whether the measurement or ascertainment of the outcome could have differed between intervention groups<br>AND<br>(ii.3.1) The outcome assessors were unaware of the intervention received by study participants<br>OR<br>(ii.3.2) The assessment of the outcome could not have been influenced by knowledge of the intervention received |
| High risk of bias | (i) The method of measuring the outcome was <u>inappropriate</u><br>OR<br>(ii) The measurement or ascertainment of the outcome <u>could</u> have differed between intervention groups<br>OR<br>(iii) It is <u>likely</u> that assessment of the outcome was influenced by knowledge of the intervention received |

**Table 12. Mapping of signalling questions to suggested risk-of-bias judgements for bias in measurement of the outcome.** This is only a suggested decision tree: all default judgements can be overridden by assessors.

| Signalling question | | | | | Domain level judgement |
|---|---|---|---|---|---|
| **4.1** | **4.2** | **4.3** | **4.4** | **4.5** | **Default risk of bias** |
| **Inappropriate?** | **Differed between groups?** | **Aware?** | **Could be influenced?** | **Likely to be influenced?** | |
| N/PN/NI | N/PN | N/PN | NA | NA | Low |
| N/PN/NI | N/PN | Y/PY/NI | N/PN | NA | Low |
| N/PN/NI | N/PN | Y/PY/NI | Y/PY/NI | N/PN | Some concerns |
| N/PN/NI | N/PN | Y/PY/NI | Y/PY/NI | Y/PY/NI | High risk |
| N/PN/NI | NI | N/PN | NA | NA | Some concerns |
| N/PN/NI | NI | Y/PY/NI | N/PN | NA | Some concerns |
| N/PN/NI | NI | Y/PY/NI | Y/PY/NI | N/PN | Some concerns |
| N/PN/NI | NI | Y/PY/NI | Y/PY/NI | Y/PY/NI | High risk |
| Y/PY | Any response | Any response | Any response | Any response | High risk |
| Any response | Y/PY | Any response | Any response | Any response | High risk |

Y/PY = 'Yes' or 'Probably yes'; N/PN = 'No' or 'Probably no'; NI = 'No information'; NA = 'Not applicable'

**Figure 5. Algorithm for suggested judgment of risk of bias in measurement of the outcome.** This is only a suggested decision tree: all default judgements can be overridden by assessors.

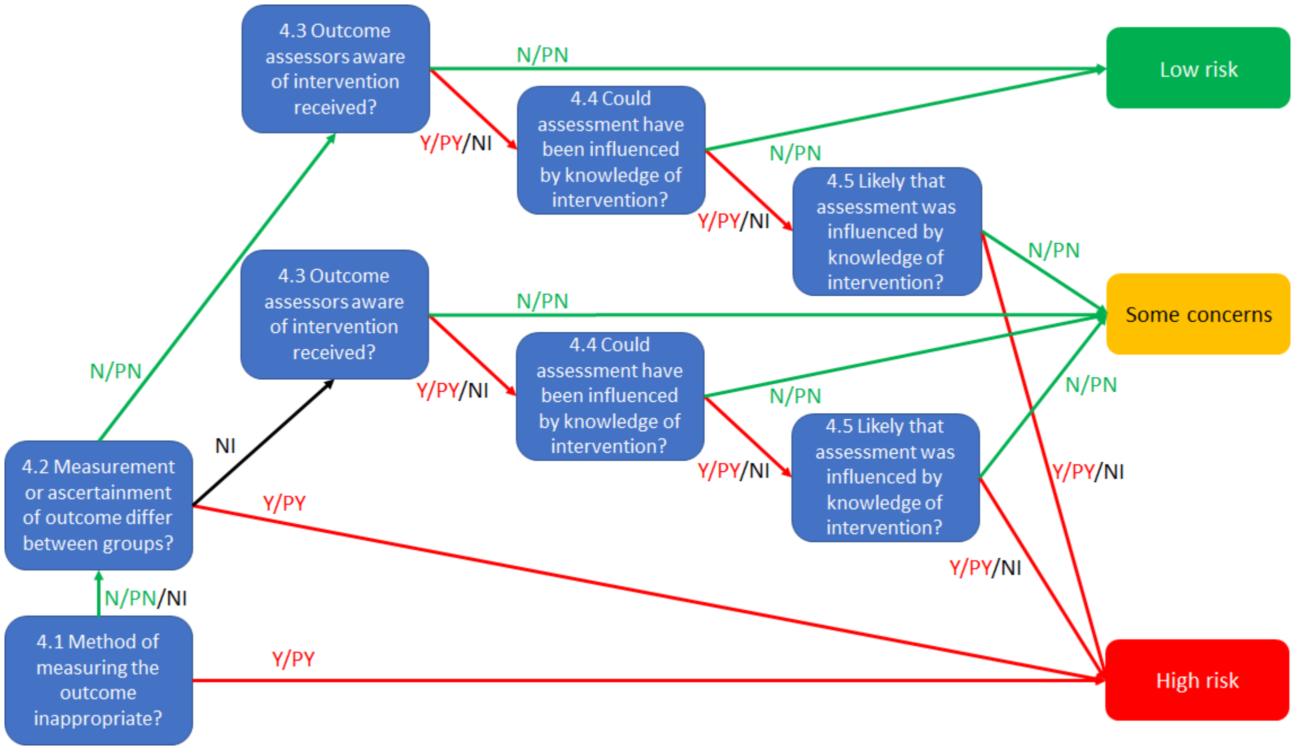

### 8 Detailed guidance: bias in selection of the reported result

#### 8.1 Background

This domain addresses bias that arises because the reported result is selected (based on its direction, magnitude or statistical significance) from among multiple intervention effect estimates that were calculated by the trial investigators. We call this **bias in selection of the reported result**. Consideration of risk of bias requires distinction between:

- An **outcome domain.** This is a state or endpoint of interest, irrespective of how it is measured (e.g. severity of depression);
- An **outcome measurement**. This is a specific way in which an outcome domain is measured (e.g. measurement of depression using the Hamilton rating scale 6 weeks after starting intervention); and
- An **outcome analysis**. This is a specific result obtained by analysing one or more outcome measurements (e.g. the difference in mean change in Hamilton rating scale scores from baseline to 6 weeks between experimental and comparator groups).

**This domain does not address bias due to selective non-reporting** (or incomplete reporting) of outcome domains that were measured and analysed by the trial investigators (115). For example, deaths of trial participants may be recorded by the trialists, but the reports of the trial might contain no mortality data, or state only that the intervention effect estimate for mortality was not statistically significant. Such bias puts the result of a synthesis at risk because results are omitted based on their direction, magnitude or statistical significance. It should therefore be addressed at the review level, as part of an integrated assessment of the risk of reporting bias (116).

An example of the distinction between an outcome domain, measurement and analysis is shown in Figure 6, along with an illustration of how risk of bias in selection of the reported result differs from risk of bias due to missing results. In this hypothetical example, the two outcome domains of depression and anxiety are of interest. Depression is measured using two scales: the Hamilton Depression Rating Scale (HAM-D) and a modified version (mHAM-D). These outcomes are analysed using both final values and change values (final minus baseline values). The reported effect estimate for depression (the difference in mean mHAM-D change values between experimental and comparator groups) was selected by the trialists because it was most favourable to the experimental intervention. The available result from the trial for depression is therefore at risk of bias due to selection of the reported result, addressed by this RoB 2 domain. In contrast, anxiety was measured and analysed by the trialists, but no data were reported because the effect estimate was not favourable to the experimental intervention. Such non-reporting will cause bias in a meta-analysis of results for anxiety but does not, by itself, put reported effect estimates for depression or other outcomes from the same trial at risk of bias.

The separation of selection in reporting a result for an outcome from selective non-reporting of an outcome domain **is a notable change from version 1 of the Cochrane RoB tool for randomized trials**.

**Figure 6. Examples of bias in selection of the reported result and outcome non-reporting bias**

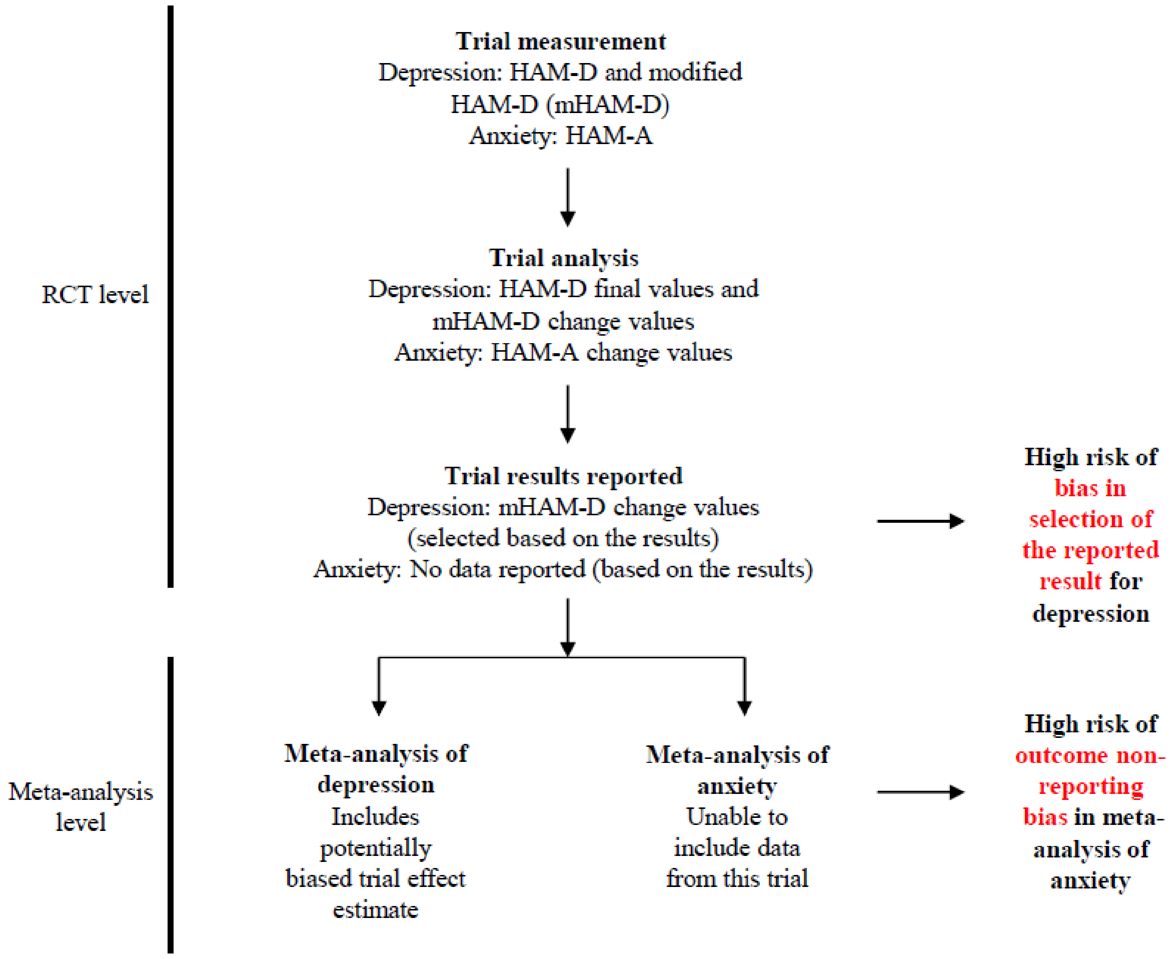


##### *8.1.1 The role of core outcome sets*

Recognition of the serious implications of selective non-reporting of outcomes by trial investigators, and substantial variability between trials in the same clinical area in the choice of outcomes measured, led to initiatives to develop **core outcome sets** (117). These are recommended lists of a small number of essential outcome domains that should be measured in all trials in a specified clinical setting. They are usually derived through a formal consensus process by eliciting the perceived importance of various outcomes from clinicians and patients (118).

Widespread adoption of core outcome sets should reduce the occurrence of selective non-reporting of outcomes. Their impact on selection of the reported result is, however, likely to be more limited, since they have not consistently included recommendations on how the core outcomes should be defined and measured. Some core outcome sets recommend the instrument that trialists should use to measure important outcomes.(119, 120) If such a recommendation was not followed in a trial designed after the core outcome measurement set was published, review authors may consider it plausible that selective reporting is present.

##### *8.1.2 Selective reporting of a result contributing to the synthesis*

This domain considers both (i) **selective reporting of a particular outcome measurement** from multiple measurements assessed within an outcome domain; and (ii) **selective reporting of a particular analysis** from multiple analyses of a specific outcome measurement. Either type of selective reporting will lead to bias if selection is based on the direction, magnitude or statistical significance of the effect estimate.

**Selective reporting of a particular outcome measurement** occurs when the reported effect estimate was selected (based on the results) from among effect estimates for multiple outcome within an outcome domain that would have been of interest to the review author. Examples include:

- reporting only one or a subset of time points at which the outcome was measured (for example, reporting only the effect at 3 weeks after baseline despite having also measured the outcome at 6 and 8 weeks);
- reporting only one or a subset of measurement instruments (e.g. pain scales);
- reporting only one or a subset of outcome assessors (e.g. patient-rated but not clinician-rated scales); and
- reporting only one or a subset of subscales of a measurement instrument.

**Selective reporting of an analysis** occurs when reported results are selected (based on the results) from intervention effects estimated in multiple ways that would have been of interest to the review author. For example:

- reporting only one of (i) the unadjusted intervention effect estimate and (ii) the effect adjusted for the baseline value of the outcome measurement ;
- reporting only one or a subset of multiple analyses adjusting for different sets of prognostic factors; and
- reporting only one or a subset of intervention effect estimates for multiple composite outcomes (for example, cardiovascular disease events defined as different combinations of death, coronary heart disease, myocardial infarction and stroke).

Bias in selection of the reported result typically arises from a desire for findings to be sufficiently noteworthy to merit publication, and this could be the case if previous evidence (or a prior hypothesis) is either supported or contradicted. Bias of this kind can arise for both harms and benefits, although the motivations (and direction of bias) underlying selection of effect estimates for harms and benefits may differ. For example, in trials comparing an experimental intervention with placebo, trialists who have a preconception or vested interest in showing that the experimental intervention is beneficial and safe may be inclined to be selective in reporting efficacy estimates that are statistically significant and favourable to the experimental intervention, along with harm estimates that are not significantly different between groups. In contrast, other trialists may selectively report harm estimates that are statistically significant and unfavourable to the experimental intervention if they believe that publicizing the existence of a harm will increase their chances of publishing in a high impact journal. Such motivations are not always easy to decipher; for example, in trials comparing active interventions, different trialists may have different preconceptions about the efficacy and safety of the different interventions.

It may be important to consider selection of the reported result in relation to the data sought by the review authors (e.g. for inclusion in a meta-analysis). Suppose a hierarchy of outcome measures and analysis methods has been pre-specified in a systematic review protocol such that, for example, change values would be taken in preference to final values, and modified HAM-D would be taken in preference to unmodified HAM-D. Considering again the scenario presented in Figure 1, in this instance the data reported are exactly the data that were sought by the review authors, despite there being evidence of potential bias in the process of selecting the reported result from the trial. Inclusion of the word “eligible” in the signalling question allows review authors to answer the question in the context of what data they would have selected from the study had all results been fully reported. In this example, the review author might consider that despite evidence of selective reporting, the planned meta-analysis is unaffected by the problem.

#### 8.2 Empirical evidence of bias in selection of the reported result

In a systematic review comparing different sections within trial reports, and comparing trial reports with their protocols, discrepancies were often found in definitions of composite outcomes, handling of missing data, unadjusted versus adjusted analyses, and subgroup analyses (121). Such discrepancies are suggestive of bias in selection of the reported result, although may be due to other reasons such as requests from journal editors or peer reviewers.

#### 8.3 Using this domain of the tool

##### *8.3.1 Importance of seeking the analysis intentions of a trial*

We strongly encourage review authors to attempt to retrieve the pre-specified analysis intentions for each trial. Doing so allows for the identification of any outcome measures or analyses that have been omitted from, or added to, the trial report subsequent to unblinded outcome data becoming available.

Analysis intentions may be documented in a variety of sources, including the trial registry entry (e.g. ClinicalTrials.gov record), trial protocol or design paper (which may be published in a journal or available via the trial funder's website). The statistical analysis plan (SAP) often provides the most details about whether a pre-specified plan was finalized before unblinded outcome data were available, but may not be published. If the researchers' pre-specified intentions are available in sufficient detail, then planned outcome measurements and analyses can be compared with those presented in the published report(s).

When comparing analysis intentions with the source(s) reporting the result being assessed, the dates of such documents must be considered carefully. There should be a 'date-stamp' confirming that the analysis intentions were finalized before unblinded outcome data were available to the trial investigators (other than staff who provided confidential data to a monitoring committee). Outcome data may sometimes be made available in a blinded format, such that analysts are unaware which intervention group is which, for the purposes of refining the analysis strategy. This should generally be considered acceptable unless the data provided are likely to include variables that clearly identify the interventions.

Amendments or updates to analysis intentions should also be retrieved and compared with the original intentions. These are usually date-stamped in trial registries or journal publications, so review authors should be able to determine when changes were made.

Review authors should ideally ask the study authors to supply the study protocol and full statistical analysis plan if these are not publicly available. In addition, if outcome measures and analyses mentioned in an article or protocol are not reported, study authors could be asked to clarify whether those outcome measures were in fact analysed and, if so, to supply the data.

Trial protocols should describe how unexpected adverse outcomes (that potentially reflect unanticipated harms) will be collected and analysed. However, results based on spontaneously-reported adverse outcomes may lead to concerns that these were selected based on the finding being noteworthy.

##### *8.3.2 Inferring selective reporting when analysis intentions are unavailable*

For some trials, the analysis intentions will not be readily available. Nevertheless,,it is still possible to assess the risk of bias in selection of the reported result. For example, outcome measures and analyses listed in the methods section of an article can be compared with those that are reported. Furthermore, outcome measures and analyses can be compared across different sources describing the trial and its results. In addition, the following questions may help review authors to infer selective reporting:

(1) Are subscales aggregated in an unusual manner?

(2) Is there a discrepancy between different reports in the designation of the primary and secondary outcomes or specific outcomes?

(3) Is there any suggestion that multiple adjusted analyses were carried out but only one (or a subset) was reported? Were one or more adjusted analyses performed but none reported?

(4) Have the researchers categorized continuous outcome measures in an unusual way? Are different cut-points for creating categories reported across multiple publications relating to the same study?

(5) Is there is a discrepancy between different reports in the sample of participants analysed?

(6) Has an unusual composite outcome been reported? Could different definitions of a composite outcome have been considered, for example by grouping different combinations of unanticipated adverse events under a category of "major adverse event" or "minor adverse event"?

It is important to recognize that some differences between analysis intentions and publication may be due to legitimate changes to the protocol. For example, planned subgroup analyses or planned cut points for continuous outcome measures may need to be modified because the distribution of data differed from what was anticipated, resulting in subgroups with no data or very uneven spread. Although such changes should be reported in publications, few trialists do so (121). Further, trialists may amend their analysis intentions before conducting any analyses, yet not update the publicly available trial registry record or protocol. Therefore, contact with authors to seek clarification for any discrepancies identified will be necessary.

Insufficient detail in some documents may limit review authors' ability to determine whether the data that produced the result being assessed were analysed in accordance with a pre-specified plan. For example, the statistical analysis plan may not be date-stamped, or trialists may state in the trial registry record only that they will measure pain, without specifying the measurement scale, time point or metric that will be used. Review authors should indicate in the free text box for responses to RoB 2 signalling questions when they made inferences based on incomplete information, and provide the basis for these.

#### 8.4 Signalling questions and criteria for judging risk of bias

Signalling questions for this domain are provided in Box 11. Criteria for reaching risk-of-bias judgements are given in Table 13, and an algorithm for implementing these is provided in Table 14 and

Figure 7.

#### Box 11. The RoB 2 tool (part 7): Risk of bias in selection of the reported result

| Signalling questions | Elaboration | Response options |
|---|---|---|
| **5.1 Were the data that produced this result analysed in accordance with a pre-specified analysis plan that was finalized before unblinded outcome data were available for analysis?** | If the researchers' pre-specified intentions are available in sufficient detail, then planned outcome measurements and analyses can be compared with those presented in the published report(s). To avoid the possibility of selection of the reported result, finalization of the analysis intentions must precede availability of unblinded outcome data to the trial investigators.<br>Changes to analysis plans that were made before unblinded outcome data were available, or that were clearly unrelated to the results (e.g. due to a broken machine making data collection impossible) do not raise concerns about bias in selection of the reported result. | Y/PY/PN/N/NI |
| **Is the numerical result being assessed likely to have been selected, on the basis of the results, from...** | | |
| **5.2. ... multiple eligible outcome measurements (e.g. scales, definitions, time points) within the outcome domain?** | A particular outcome domain (i.e. a true state or endpoint of interest) may be **measured** in multiple ways. For example, the domain pain may be measured using multiple scales (e.g. a visual analogue scale and the McGill Pain Questionnaire), each at multiple time points (e.g. 3, 6 and 12 weeks post-treatment). If multiple measurements were made, but only one or a subset is reported on the basis of the results (e.g. statistical significance), there is a high risk of bias in the fully reported result. Attention should be restricted to outcome measurements that are eligible for consideration by the RoB 2 tool user. For example, if only a result using a specific measurement scale is eligible for inclusion in a meta-analysis (e.g. Hamilton Depression Rating Scale), and this is reported by the trial, then there would not be an issue of selection even if this result was reported (on the basis of the results) in preference to the result from a different measurement scale (e.g. Beck Depression Inventory).<br>Answer 'Yes' or 'Probably yes' if:<br>There is clear evidence (usually through examination of a trial protocol or statistical analysis plan) that a domain was measured in multiple eligible ways, but data for only one or a subset of measures is fully reported (without justification), and the fully reported result is likely to have been selected on the basis of the results. Selection on the basis of the results can arise from a desire for findings to be newsworthy, sufficiently noteworthy to merit publication, or to confirm a prior hypothesis. For example, trialists who have a preconception, or vested interest in showing, that an experimental intervention is beneficial may be inclined to report outcome measurements selectively that are favourable to the experimental intervention.<br>Answer 'No' or 'Probably no' if:<br>There is clear evidence (usually through examination of a trial protocol or statistical analysis plan) that all eligible reported results for the outcome domain correspond to all intended outcome measurements. | Y/PY/PN/N/NI |

| | | |
|---|---|---|
| | or<br>There is only one possible way in which the outcome domain can be measured (hence there is no opportunity to select from multiple measures).<br>or<br>Outcome measurements are inconsistent across different reports on the same trial, but the trialists have provided the reason for the inconsistency and it is not related to the nature of the results.<br>Answer 'No information' if:<br>Analysis intentions are not available, or the analysis intentions are not reported in sufficient detail to enable an assessment, and there is more than one way in which the outcome domain could have been measured. | |
| **5.3 ... multiple eligible analyses of the data?** | A particular outcome measurement may be analysed in multiple ways. Examples include: unadjusted and adjusted models; final value vs change from baseline vs analysis of covariance; transformations of variables; different definitions of composite outcomes (e.g. 'major adverse event'); conversion of continuously scaled outcome to categorical data with different cut-points; different sets of covariates for adjustment; and different strategies for dealing with missing data. Application of multiple methods generates multiple effect estimates for a specific outcome measurement. If multiple estimates are generated but only one or a subset is reported on the basis of the results (e.g. statistical significance), there is a high risk of bias in the fully reported result. Attention should be restricted to analyses that are eligible for consideration by the RoB 2 tool user. For example, if only the result from an analysis of post-intervention values is eligible for inclusion in a meta-analysis (e.g. at 12 weeks after randomization), and this is reported by the trial, then there would not be an issue of selection even if this result was reported (on the basis of the results) in preference to the result from an analysis of changes from baseline.<br>Answer 'Yes' or 'Probably yes' if:<br>There is clear evidence (usually through examination of a trial protocol or statistical analysis plan) that a measurement was analysed in multiple eligible ways, but data for only one or a subset of analyses is fully reported (without justification), and the fully reported result is likely to have been selected on the basis of the results. Selection on the basis of the results arises from a desire for findings to be newsworthy, sufficiently noteworthy to merit publication, or to confirm a prior hypothesis. For example, trialists who have a preconception or vested interest in showing that an experimental intervention is beneficial may be inclined to selectively report analyses that are favourable to the experimental intervention.<br>Answer 'No' or 'Probably no' if:<br>There is clear evidence (usually through examination of a trial protocol or statistical analysis plan) that all eligible reported results for the outcome measurement correspond to all intended analyses.<br>or | Y/PY/PN/N/NI |

| | | |
|---|---|---|
| | There is only one possible way in which the outcome measurement can be analysed (hence there is no opportunity to select from multiple analyses).<br>or<br>Analyses are inconsistent across different reports on the same trial, but the trialists have provided the reason for the inconsistency and it is not related to the nature of the results.<br>Answer 'No information' if:<br>Analysis intentions are not available, or the analysis intentions are not reported in sufficient detail to enable an assessment, and there is more than one way in which the outcome measurement could have been analysed. | |
| **Risk-of-bias judgement** | See Table 13, Table 14 and Figure 7. | Low / High / Some concerns |
| Optional: What is the predicted direction of bias due to selection of the reported result? | If the likely direction of bias can be predicted, it is helpful to state this. The direction might be characterized either as being towards (or away from) the null, or as being in favour of one of the interventions. | NA / Favours experimental / Favours comparator / Towards null /Away from null / Unpredictable |

**Table 13. Reaching risk-of-bias judgements for bias in selection of the reported result**

| | |
|---|---|
| Low risk of bias | (i) The data were analysed in accordance with a pre-specified plan that was finalised before unblinded outcome data were available for analysis<br><br>AND<br><br>(ii) The result being assessed is unlikely to have been selected, on the basis of the results, from multiple eligible outcome measurements (e.g. scales, definitions, time points) within the outcome domain<br><br>AND<br><br>(iii) Reported outcome data are unlikely to have been selected, on the basis of the results, from multiple eligible analyses of the data |
| Some concerns | (i.1) The data were not analysed in accordance with a pre-specified plan that was finalised before unblinded outcome data were available for analysis<br><br>AND<br><br>(i.2) The result being assessed is unlikely to have been selected, on the basis of the results, from multiple eligible outcome measurements (e.g. scales, definitions, time points) within the outcome domain<br><br>AND<br><br>(i.3) The result being assessed is unlikely to have been selected, on the basis of the results, from multiple eligible analyses of the data<br><br>OR<br><br>(ii) There is no information on whether the result being assessed is likely to have been selected, on the basis of the results, from multiple eligible outcome measurements (e.g. scales, definitions, time points) within the outcome domain AND from multiple eligible analyses of the data |
| High risk of bias | i) The result being assessed is likely to have been selected, on the basis of the results, from multiple eligible outcome measurements (e.g. scales, definitions, time points) within the outcome domain<br><br>OR<br><br>(ii) The result being assessed is likely to have been selected, on the basis of the results, from multiple eligible analyses of the data |

**Table 14. Mapping of signalling questions to suggested risk-of-bias judgements for bias in selection of the reported result.**

| Signalling question | | | Domain level judgement |
|---|---|---|---|
| **5.1** | **5.2** | **5.3** | **Default risk of bias** |
| **In accordance with plan?** | **Selected from multiple outcomes?** | **Selected from multiple analyses?** | |
| Y/PY | N/PN | N/PN | Low |
| N/PN/NI | N/PN | N/PN | Some concerns |
| Any answer | N/PN | NI | Some concerns |
| Any answer | NI | N/PN | Some concerns |
| Any answer | NI | NI | Some concerns |
| Any answer | Either 5.2 or 5.3 Y/PY | | High |

Y/PY = 'Yes' or 'Probably yes'; N/PN = 'No' or 'Probably no'; NI = 'No information'

**Figure 7. Algorithm for suggested judgment of risk of bias in selection of the reported result.** This is only a suggested decision tree: all default judgements can be overridden by assessors.

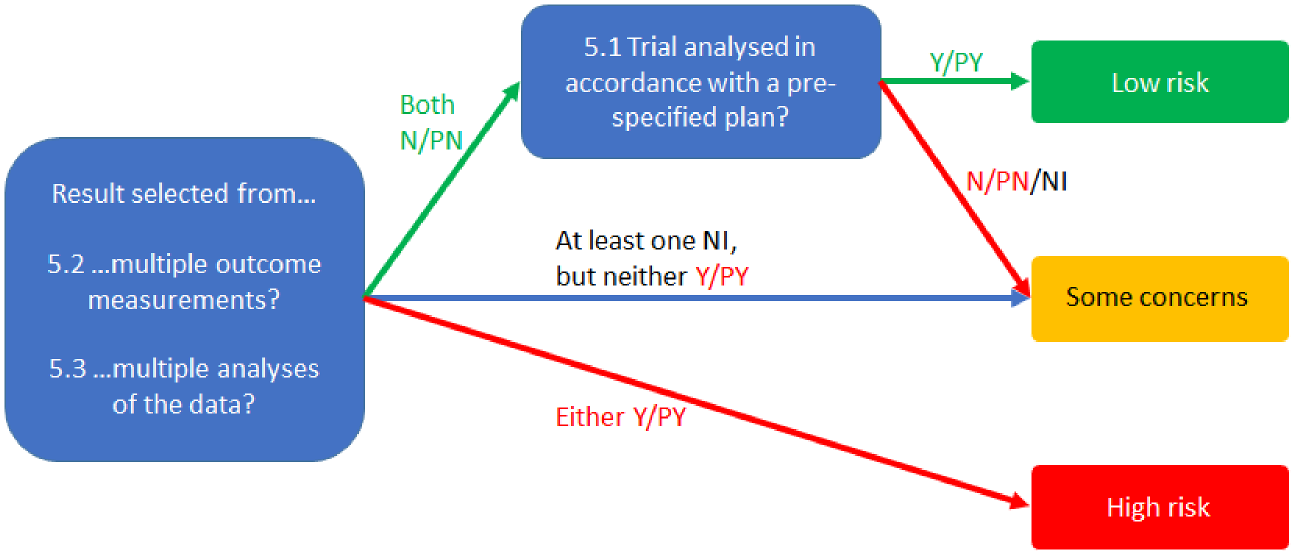

### 9 Acknowledgements

The development of the RoB 2 tool was supported by the MRC Network of Hubs for Trials Methodology Research (MR/L004933/2- N61), with the support of the host MRC ConDuCT-II Hub (Collaboration and innovation for Difficult and Complex randomised controlled Trials In Invasive procedures - MR/K025643/1), by MRC research grant MR/M025209/1, and by a grant from The Cochrane Collaboration.

### 10 Contributors

*Core group*: Julian Higgins, Jonathan Sterne, Jelena Savović, Matthew Page, Roy Elbers

*Contributors to bias domain development*: Natalie Blencowe, Isabelle Boutron, Christopher Cates, Rachel Churchill, Mark Corbett, Nicky Cullum, Jonathan Emberson, Sally Hopewell, Asbjørn Hróbjartsson, Sharea Ijaz, Peter Jüni, Jamie Kirkham, Toby Lasserson, Tianjing Li, Barney Reeves, Sasha Shepperd, Ian Shrier, Lesley Stewart, Kate Tilling, Ian White, Penny Whiting

*Other contributors*: Henning Keinke Andersen, Mike Clarke, Jon Deeks, Miguel Hernán, Daniela Junqueira, Yoon Loke, Geraldine MacDonald, Richard Morris, Mona Nasser, Nishith Patel, Jani Ruotsalainen, Holger Schünemann, Jayne Tierney, Sunita Vohra, Liliane Zorzela
